# Population-scale burden analysis of rare damaging coding variants identifies novel risk genes for Alzheimer’s disease and related dementias and Parkinson’s disease and related disorders

**DOI:** 10.64898/2026.03.03.26347540

**Authors:** Yann Le Guen, Andrés Peña-Tauber, Rafael Catoia Pulgrossi, Junyoung Park, Holden Orias, Michael D. Greicius

**Affiliations:** Quantitative Sciences Unit, Division of Computational Medicine, Department of Medicine Stanford University, Stanford, CA; Department of Neurology and Neurological Sciences, Stanford University, Stanford, CA

## Abstract

Alzheimer’s disease and related dementias (ADRD)^1^ and Parkinson’s disease and related disorders (PDRD)^2^ have substantial genetic contributions, yet the role of rare damaging coding variants remains incompletely characterized at population scale^3–6^. We performed gene-burden testing of loss-of-function and deleterious missense variants in biobanks and disease-focused sequencing cohorts, using calibrated proxy-phenotypes and diagnosed-case sensitivity analyses to increase power for late-onset disease^7^. At study-wide significance, we confirmed rare-variant burden in established ADRD genes (*TREM2, ABCA7, SORL1, PSEN1, GRN, ATP8B4, ABCA1, ADAM10*) and PDRD genes (*GBA1, LRRK2*). We also identified a novel ADRD burden association at *IMPA2*, and novel PDRD associations at *ANKRD27*, and *CCL7*. The strongest signal was observed for *ANKRD27*, where damaging variants clustered within domains mediating interactions with Rab GTPases and retromer components. Our results demonstrate the value of population-scale sequencing to identify rare coding risk genes for neurodegenerative diseases.

---

Genome-wide association studies (GWAS) have extensively characterized the common variant genetic architecture of Alzheimer’s disease and related dementias (ADRD) and Parkinson’s disease and related disorders (PDRD), identifying dozens of loci across diverse biological pathways^1,2^. Increasing sample sizes, improved imputation panels, and inclusion of diverse populations have further refined association signals and enabled identification of ancestry-specific risk variants^8–12^. In parallel, sequencing-based studies have demonstrated that rare, damaging coding variants contribute substantially to disease risk, often with larger effect sizes than common variants. Rare loss-of-function (LoF) and deleterious missense variants in genes such as *TREM2, SORL1* and *ABCA7* for ADRD and *GBA1*, *LRRK2, RAB32* for PDRD have established rare coding variation as an important component of neurodegenerative disease genetics^3–6,13,14^. However, rare variant discovery remains limited by sample size, sequencing heterogeneity across cohorts, and the requirement for sufficient numbers of variant carriers for gene-based burden testing^3^. Population-scale sequencing in large biobanks, combined with proxy phenotypes, provides an opportunity to increase statistical power while minimizing sequencing heterogeneity^7,13,15^. Furthermore, whole-genome sequencing reduces biases associated with heterogeneous exome capture kits and nonuniform exon coverage across cohorts.

We performed gene-based burden testing of rare damaging coding variants in ADRD and PDRD using whole-genome sequencing data from population studies including the UK Biobank (UKB, N = 484,909)^16^ and All Of Us (AoU, N = 222,274)^17^, disease-focused sequencing cohorts including the Alzheimer’s Disease Sequencing Project^18^ (ADSP, N = 41,483) and the Accelerating Medicine Partnership in Parkinson’s Disease and Related Disorders (AMP-PDRD, N = 10,011)^19^, and summary statistics from a previous study of AD (Holstege et al. 2022, N = 21,345)^3^. Variants were restricted to rare (minor allele frequency [MAF] < 1%) protein-coding LoF and *in silico* predicted deleterious missense variants, grouped into four nested sets (LoF; LoF+REVEL≥0.75; LoF+REVEL≥0.50; LoF+REVEL≥0.25), consistent with prior studies^3,20,21^. Gene-based tests were performed with REGENIE^22^ and meta-analyzed across cohorts using inverse-variance– weighted fixed-effects models (**Supplementary Notes 1 and 2**). We required cumulative minor allele count (cMAC) > 10 per gene within each set; 18,298 unique genes were included.

For ADRD, the primary meta-analysis, which combined AD-directly diagnosed and proxy-defined cases and is hereafter referred to as the “AD-combined-proxy” analysis, recovered study-wide associations in established or previously implicated ADRD/dementia genes, including *TREM2, ABCA7, SORL1, PSEN1, GRN, ATP8B4* and *ADAM10* (**Fig. 1**; **Table 1; Supplementary Fig. 1-4**). A novel study-wide association was identified in the all-ancestry primary analysis *IMPA2* (LoF+REVEL≥0.75; all-ancestry, OR=3.614 [2.171-6.014]; P=7.67×10^−7^). Secondary European-ancestry analyses identified associations at *SYNE1* and *SP140L*; both showing consistent burden across cohorts (*SYNE1,* LoF+REVEL≥0.50; OR=1.215 [1.114-1.325]; P=1.30×10^−6^; I²=0; and *SP140L* LoF+REVEL≥0.75; OR=1.995 [1.497-2.658]; P=3.54×10^−6^; I²=0) (**Supplementary Table 1**). Near-threshold signals in primary and secondary AD-combined-proxy analyses included *CHRNA4, ABCA1, SHARPIN, APOE, PMM2, MME* and *CCL7*, spanning lipid biology, immune regulation, amyloid-β clearance and glycosylation (**Supplementary Tables 1-2**). Single-variant meta-analysis (“lollipop”) plots provide a compact view of allelic architecture in established genes (**Supplementary Fig. 5, Supplementary Table 3**), with *TREM2* burden signal dominated by known AD variant R47H^1^, whereas *ABCA7* showed a multi-allelic pattern driven by LoF variants. Among genes not already highlighted *TBK1, RIN3, CLU, RBM12, ZNF256* all reached p < ×10^−5^ in the AD-direct diagnosis sensitivity analyses (**Supplementary Fig. 6–7; Supplementary Table 4**).

**Figure 1.**
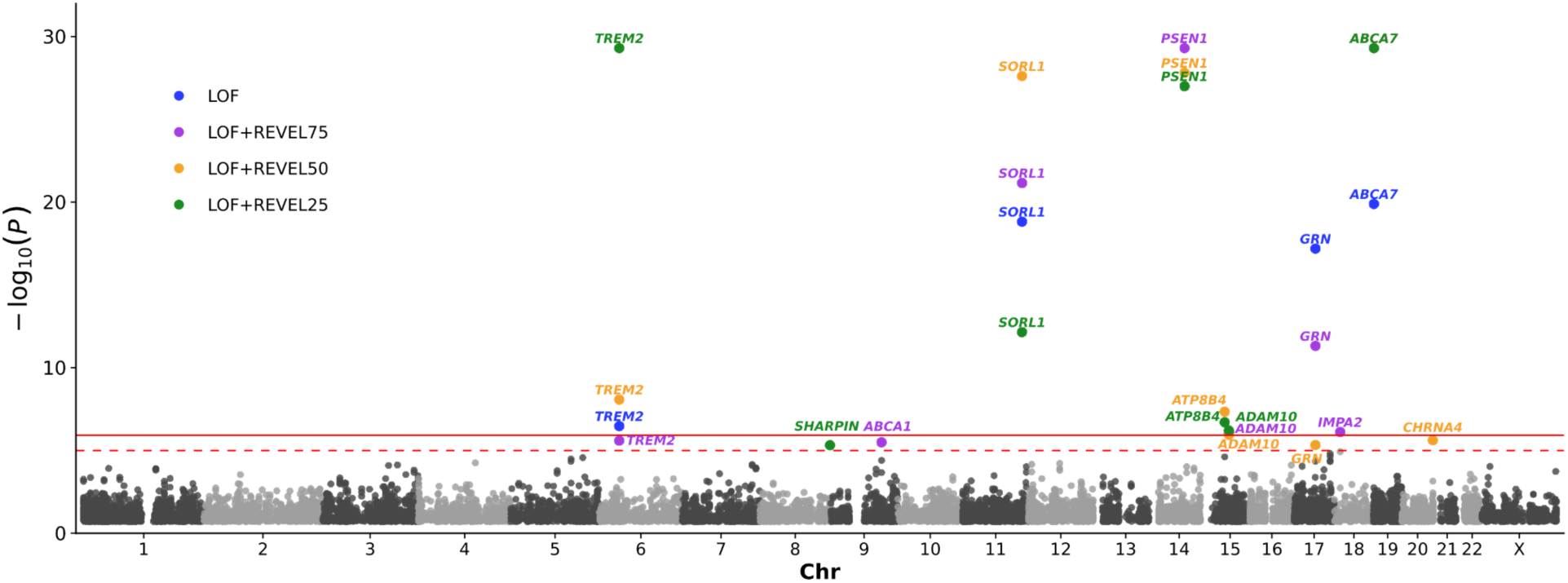
Gene-based rare-variant burden associations with the AD-combined-proxy in the all-ancestry meta-analysis. P-values capped at p = 5×10^-30^. The solid horizontal line denotes the study-wide significance threshold for ADRD (P<1.20 ×10^-6^), and the dashed line denotes the suggestive threshold (P<10^-5^).

**Table 1.**
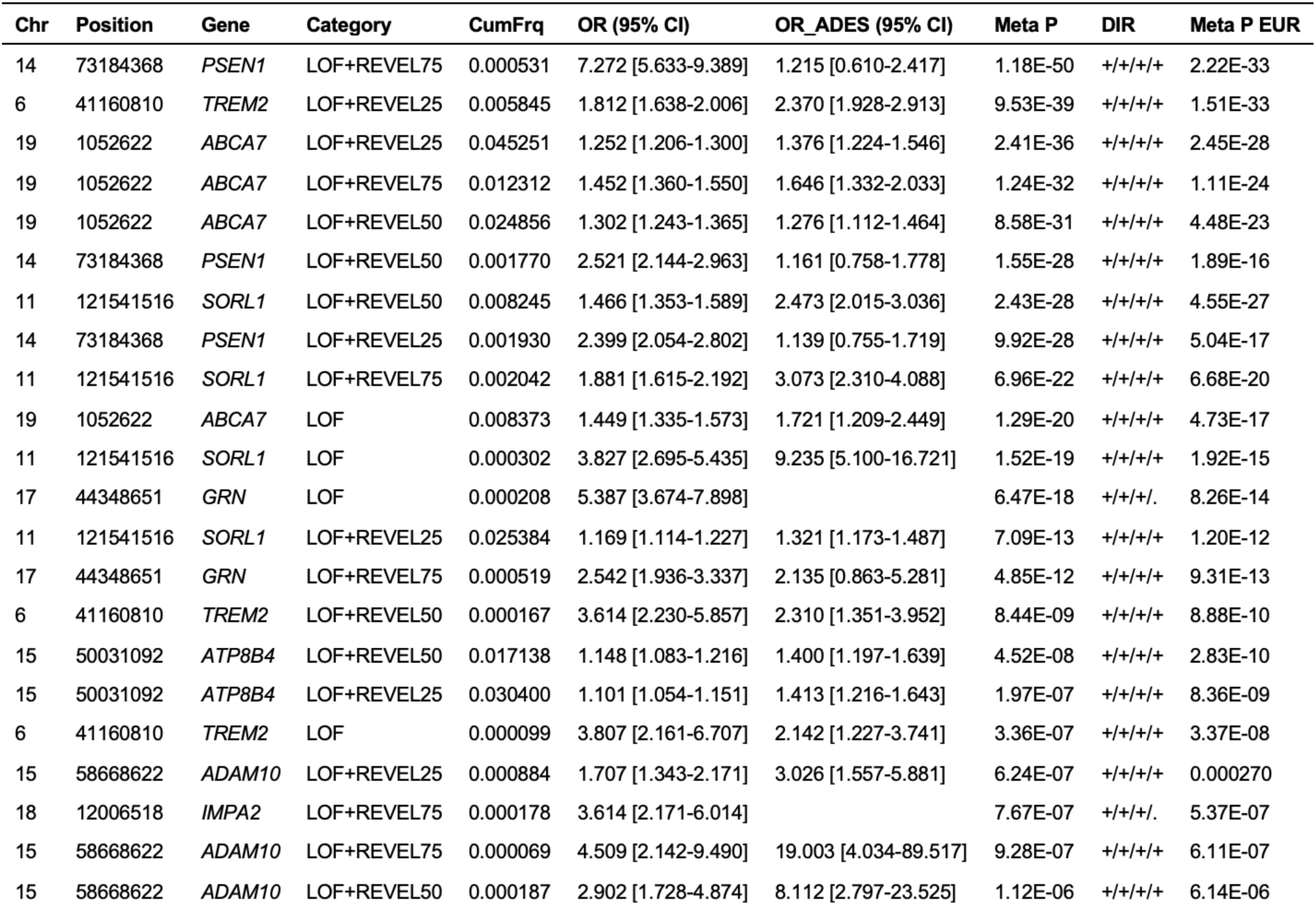
Summary of study-wide significant gene-based burden associations in the primary all-ancestry AD-combined-proxy analysis. Gene–mask associations reaching the prespecified study-wide significance threshold (P<1.20×10^−6^) in the primary all-ancestry meta-analysis are shown. Chr and Position, chromosome and GRCh38 genomic position; Category, qualifying-variant mask; CumFrq, cumulative frequency of qualifying variants; OR (95% CI), pooled odds ratio and 95% confidence interval from UKB, AoU and ADSP; OR_ADES (95% CI), corresponding estimate from ADES stage 1; Meta P, final meta-analysis P-value including ADES when available; DIR, direction of effects in UKB/AoU/ADSP/ADES; Meta P EUR, corresponding European-ancestry meta-analysis P-value.

In *GRN*, the association was strongest for LoF variants, consistent with haploinsufficiency and the known role of *GRN* in frontotemporal dementia.^23^ *GRN* LoF burden is strongly associated under both broad all-dementia and narrower AD direct diagnosis in biobanks (**Supplementary Fig. 8**). Within the directly diagnosed UKB strata, *TBK1* and *VCP*, two ALS/FTD-spectrum genes, showed descriptively larger point estimates for AD than all-cause dementia (mostly ADRD). ADSP likewise showed positive point estimates for rare damaging variants in *GRN*, *TBK1*, and *VCP*, with associations reaching P<0.05 for *GRN* and for *TBK1* at the LoF+REVEL25 mask, despite specialist-driven ascertainment and biomarker or neuropathological confirmation in only a subset of participants (**Supplementary Fig. 8**). Conversely, ADES stage 1 excluded pathogenic-variant carriers in *PSEN1*, *PSEN2*, *APP*, and unspecified “other genes causative for Mendelian dementia diseases,” as well as individuals with clinical information suggestive of non-AD dementia. Because the complete excluded-gene list was not reported, whether carriers of pathogenic variants in *GRN*, *TBK1*, *VCP*, or other FTD/ALS-spectrum genes were removed cannot be confirmed. Such exclusion, based genomics information, would attenuate signals from Mendelian non-AD dementia genes in ADES, such as for *GRN* LoFs. Nevertheless, for less established pathogenic categories, ADES showed positive point estimates descriptively larger than ADSP and in the same range as biobanks all-cause dementia direct diagnosis (*VCP* LoF+REVEL50, *GRN* LoF+REVEL75 burdens **Supplementary Fig. 8**). Together, these observations support the conclusion that both broad dementia and clinically diagnosed AD labels capture etiologically heterogeneous syndromes and support interpreting *GRN*, *TBK1*, and *VCP* as indicators of diagnostic or etiologic heterogeneity rather than AD-specific mechanisms.

For PDRD, the primary meta-analysis, corresponding to the PD-combined-proxy in all-ancestry, recapitulated established rare-variant associations at *GBA1* and *LRRK2* (**Fig. 2**; **Table 2, Supplementary Fig. 9-12**), with allelic patterns consistent with known risk alleles (**Supplementary Fig. 13, Supplementary Table 5**). The strongest novel PD-combined-proxy association was *ANKRD27* (LoF+REVEL≥0.25; OR=1.305 [1.184-1.438]; P=7.31×10^−8^), with consistent direction of effect across cohorts and minimal heterogeneity in the most significant category (**Fig. 3a**). In *ANKRD27* single-variant meta-analysis (**Fig. 3b**), the lead variant was R21C (∼0.5% in Europeans), with additional contributing missense alleles largely clustering within regions implicated in interactions with Rab GTPases and retromer/WASH pathway components (**Fig. 3b**). Of note, the lead variant (R21C) shows largely independent support in the GP2 clinical-only (non-biobank) imputed European-ancestry case–control meta-analysis (MAF=0.0114; OR=1.71; p=4.0×10^-5^).^24^ The second novel study-wide significant PD combined-proxy was *CCL7* (LoF+REVEL≥0.50; OR=14.025 [5.168-38.058]; P=2.16×10^−7^). Suggestive PD-combine-proxy include *USP19, BNIP3, SOBP, KANSL3, KRBA2, AGFG1* in the all-ancestry analysis (**Supplementary Table 6**), and additionally *ZSCAN25, SPIR2, ATAD2B, HOXA13* in the European ancestry secondary analysis (**Supplementary Table 7**), motivating replication in independent sequencing datasets and orthogonal functional prioritization. Directly diagnosed PD analyses were considered identified *LRRK2* and *GBA1* as study-wide significance; suggestive signals were observed at *PANX2* (**Supplementary Fig. 14-15**; **Supplementary Table 8**).

**Figure 2.**
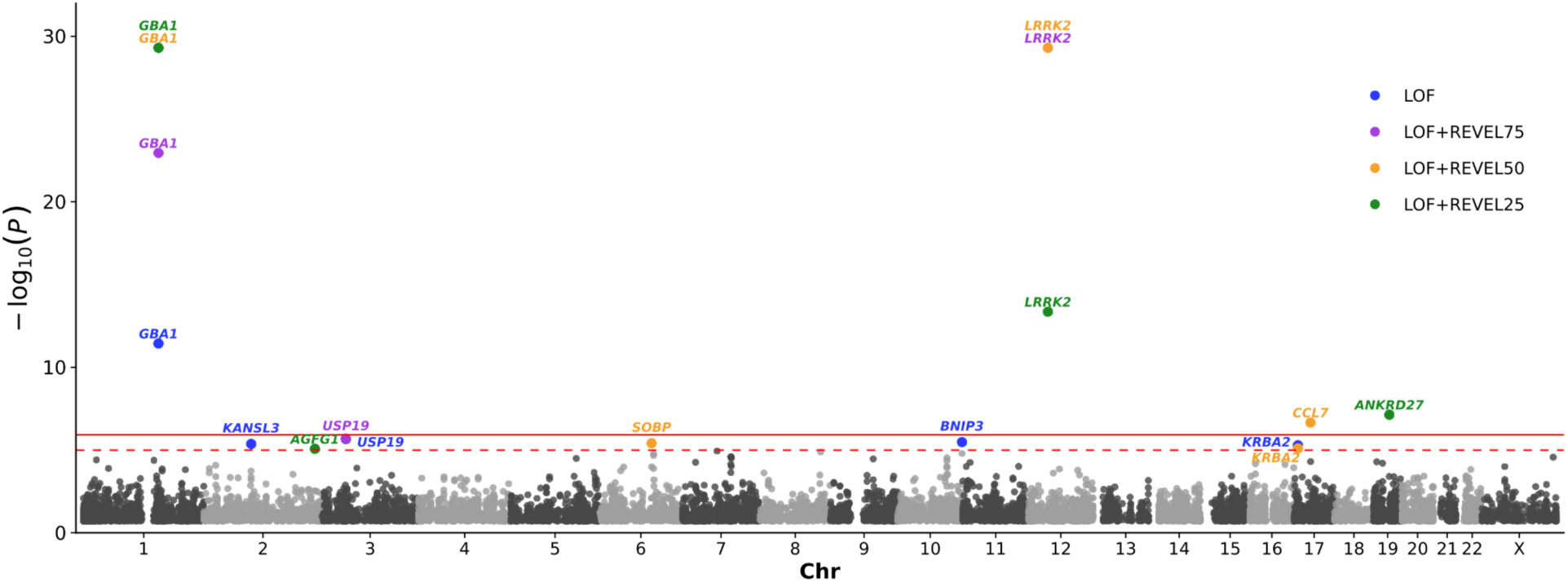
Gene-based rare-variant burden associations with the PD-combined-proxy in the all-ancestry meta-analysis. P-values capped at p = 5×10^-30^). The solid horizontal line denotes the study-wide significance threshold for PDRD (P<1.20×10^-6^), and the dashed line denotes the suggestive threshold (P<10^-5^).

**Figure 3.**
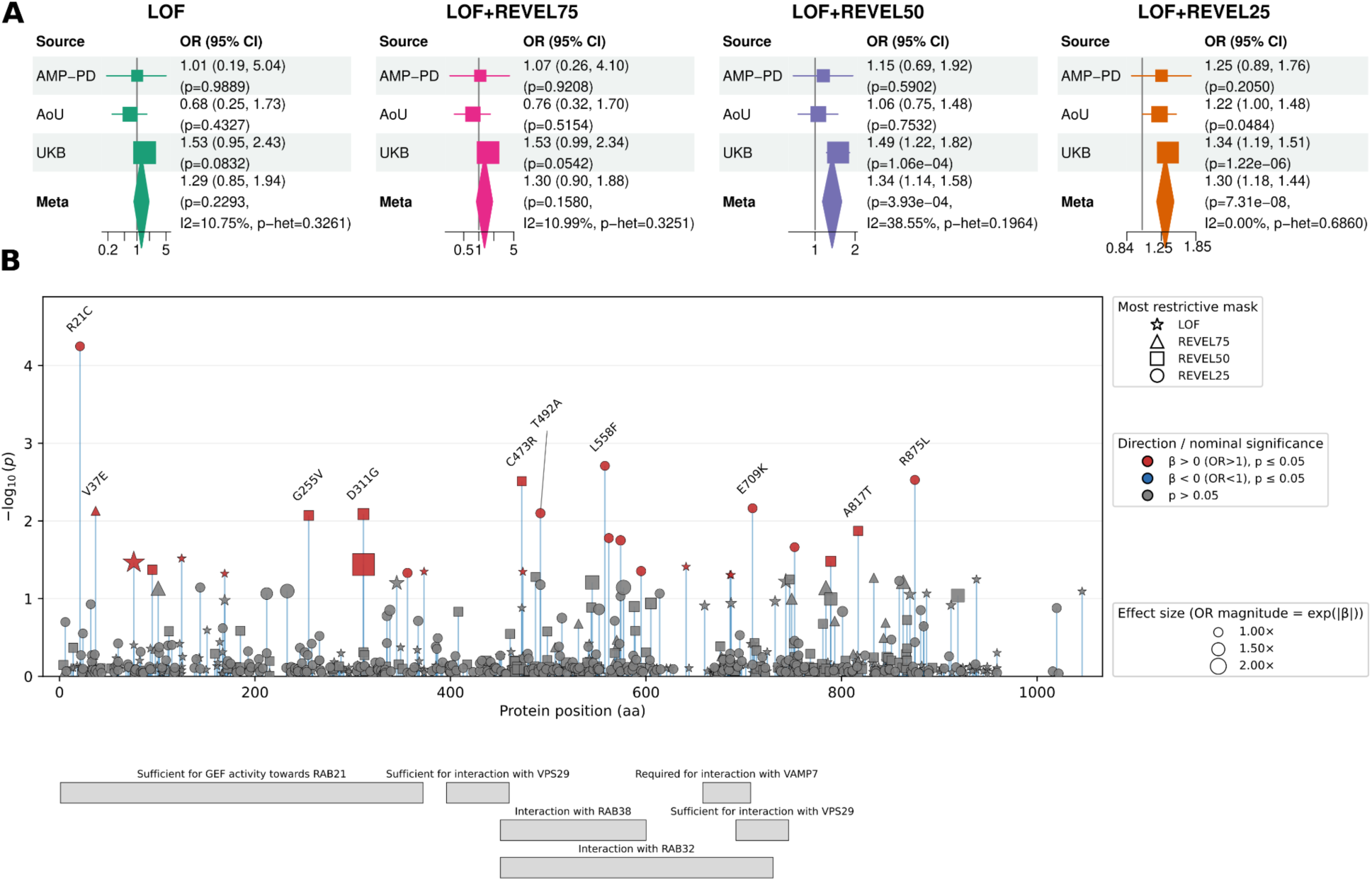
*ANKRD27* association with Parkinson’s disease and contributing variants. (a) Forest plots for the *ANKRD27* burden sets in the in the primary all-ancestry PD-combined-proxy meta-analysis, showing odds ratios (ORs) and 95% confidence intervals across contributing cohorts (UKB, AoU, AMP-PDRD) and the fixed-effect meta-analysis. P values and heterogeneity metrics are shown for the meta-analysis. (b) Lollipop plot of *ANKRD27* single-variant meta-analysis results, with variants positioned by protein coordinate (aa) and –log_10_(P); point size scales with absolute effect size, and top 10 variants are labeled; domain and feature annotations are derived from UniProt protein features.

**Table 2.**
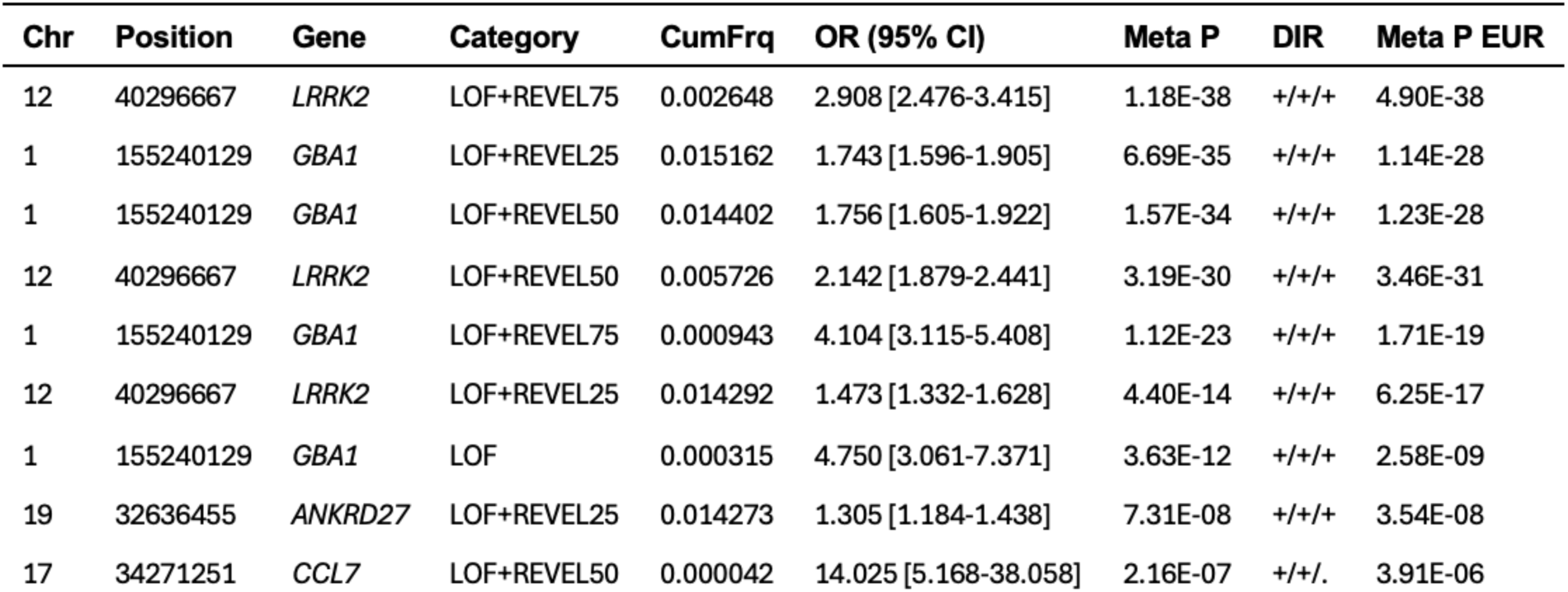
Summary of study-wide significant gene-based burden associations in the primary all-ancestry PD-combined-proxy analysis. Gene–mask associations reaching the prespecified study-wide significance threshold (P<1.20×10^−6^) in the primary all-ancestry meta-analysis are shown. Chr and Position, chromosome and GRCh38 genomic position; Category, qualifying-variant mask; CumFrq, cumulative frequency of qualifying variants; OR (95% CI), pooled odds ratio and 95% confidence interval from UKB, AoU and AMP-PD; Meta P, final meta-analysis P-value; DIR, direction of effects in UKB/AoU/AMP-PD; Meta P EUR, corresponding European-ancestry meta-analysis P-value.

In AD-combined-proxy analyses, the retained associations highlight both established AD mechanisms and additional metabolic, synaptic, immune and regulatory nodes that warrant follow-up. The novel study-wide primary association at *IMPA2* implicates inositol recycling and phosphoinositide-dependent membrane dynamics, consistent with reports of altered myo-inositol monophosphatase activity in AD brain and broader links between inositol-related pathways and neurodegeneration^25,26^. *IMPA2* has also been implicated as a pharmacological target of lithium in brain^27^, and recent work showing that dietary lithium deficiency accelerates amyloid-β and phospho-tau pathology in AD mouse models provides disease-relevant context for this pathway^28^. Although *ADAM10* did not reach study-wide significance in the previous largest rare-variant burden analysis^3^, its association has strong prior family-based and functional support: *ADAM10* encodes the principal α-secretase involved in non-amyloidogenic APP processing, and rare *ADAM10* variants have been shown to attenuate α-secretase activity and segregate with late-onset AD in families^29^. The European-ancestry secondary analysis identified a suggestive association at *SYNE1*, encoding a nuclear-envelope spectrin-repeat protein involved in nuclear-cytoskeletal coupling and neuronal maintenance, consistent with the complex neurodegenerative phenotypes produced by damaging *SYNE1* variation^30,31^. Among suggestive signals in the combined-proxy analyses, the protective *APOE* burden reinforces the ability of WGS-based burden testing to resolve directionally distinct rare-allele architectures, in this case driven by the known protective variants R269G and V254E^11^. *SHARPIN* and *MME*, together with *CLU* and *RIN3* in AD-direct sensitivity analyses, are best interpreted as rare-coding support for established AD GWAS loci^1^. *CLU* implicates clusterin-mediated amyloid-β handling, lipid transport and complement biology^32^; *RIN3* implicates Rab-dependent endosomal trafficking^33^ and RIN3–BIN1–RAB5 biology^34^; and *MME* encodes neprilysin, a major amyloid-β-degrading enzyme, making its association biologically coherent with impaired amyloid clearance^35^. *PMM2* highlights protein glycosylation, a process repeatedly implicated in AD and supported by integrative analyses nominating the *KIAA0319–PMM2* axis^36,37^, whereas *CHRNA4* implicates α4β2 nicotinic receptor signaling, which is reduced in AD brain and linked to synaptic and cognitive dysfunction^38,39^. The suggestive *SP140L* and *CCL7* signals extend the immune component, respectively nominating interferon-responsive transcriptional restriction^40^ and chemokine-mediated immune-cell recruitment^41,42^, although direct AD-specific evidence remains limited. The suggestive *TBK1* signal, discussed above, is best interpreted through its established role as an ALS/FTD haploinsufficiency gene.^43^

In PD-combined-proxy analyses, the study-wide associations at *ANKRD27* and *CCL7* extend human genetic support for endosomal recycling and immune recruitment, whereas suggestive signals nominate mitochondrial quality control, proteostasis and transcriptional regulation. *ANKRD27* encodes VARP, an endosomal trafficking effector with documented links to retromer-dependent recycling and Rab GTPase biology, providing a mechanistic bridge from the observed multi-allelic burden signal to vesicular recycling pathways implicated in PDRD^44–46^. This interpretation is strengthened by convergent rare-variant evidence at *RAB32* in familial Parkinson’s disease^14^. *CCL7* implicates chemokine signaling and immune cell recruitment, consistent with broader literature connecting chemokine networks and neuroinflammation to PD-related neurodegeneration^41,42^. Among suggestive signals in the combined-proxy analyses, *USP19* extends the proteostasis theme: genetic inactivation of *USP19* modifies α-synuclein ubiquitination and reduces Lewy body-like aggregate accumulation in vivo, while biochemical studies place USP19 within ER-associated degradation and chaperone-linked protein-quality-control networks^47,48^*. BNIP3* implicates mitochondrial stress and receptor-mediated mitophagy, a pathway closely related to PINK1/Parkin-mediated mitochondrial quality control^49^. *KANSL3*, a component of the NSL/KAT8 complex required for KAT8-dependent mitochondrial transcription and respiration^50^, nominates chromatin–mitochondrial coupling^51^, although direct PD-specific evidence remains limited. By contrast, *ZSCAN25, KRBA2, SOBP, AGFG1, SPIRE2, ATAD2B* and *HOXA13* have little direct support from current PD biology and should be treated as statistical prioritization signals pending independent replication and orthogonal functional validation. In the PD-direct sensitivity analysis, the suggestive *PANX2* signal nominates a neurally expressed, ATP-permeable pannexin channel that localizes to early endosomal vesicles, linking the association to pannexin-dependent ATP and endosomal signaling^52^.

To facilitate translational interpretation and variant exploration, we developed an interactive query resource enabling gene-level and variant-level interrogation of burden signals, carrier counts, set-specific effect estimates (including whether LoF enrichment is risk-increasing, protective, or null), and cohort-stratified results (https://qsushiny.shinyapps.io/adpd_burden/). This resource is intended to support translational investigators and clinicians, and functional genomics studies as whole-genome sequencing becomes increasingly routine in research and clinical settings.

In summary, by combining population-scale WGS from large biobanks with disease-focused sequencing cohorts and harmonized gene-based tests, we confirm rare-variant enrichment in established ADRD and PDRD genes x Proxy phenotypes can introduce heterogeneity, attenuate effect estimates and preclude precise inference on age at onset, whereas clinically diagnosed AD and PD labels remain imperfect proxies for etiologic pathology. Across 26 AD/ADRD and 10 PD/PDRD exact gene/mask pairs, study-wide significance in either the all-ancestry AD/PD combined-proxy or direct-diagnosis analysis, effect directions were preserved across all corresponding phenotype comparisons. Absolute agreement was close between broad and narrow combined-proxy definitions (ADRD versus AD: Lin’s CCC=0.987, Deming slope=1.057; PDRD versus PD: CCC=0.990, slope=0.996), but more variable between the AD/PD combined-proxy and direct analyses (AD: CCC=0.857, slope=1.262; PD: CCC=0.893, slope=0.815; **Supplementary Table 9**). Gene-based burden tests also do not resolve variant-specific mechanisms, motivating functional follow-up. Despite these limitations, large-scale WGS burden analyses combined with a queryable data portal provide a resource for prioritizing rare coding risk genes and generating mechanistic hypotheses backed by human genetic evidence.

## Data Availability

All data produced are available online on their respective Platform UK Biobank, All Of Us, NIAGADS, AMP-PD.
Summary statistics have been deposited on zenodo
doi: 10.5281/zenodo.22702628

https://zenodo.org/records/22702628

## Online Methods

### Cohorts and phenotypes definition

#### UK Biobank (UKB)

Whole-genome sequencing (WGS) data were available for 490,640 UKB participants as described previously.^16^ We defined last known age as the maximum age observed across assessment age (field 21003), age at death (field 40007), and ages derived from first in-patient diagnosis dates (fields 41280 for ICD-10 and 41281 for ICD-9), computed using birth month and year and the corresponding diagnosis date fields. Sex was obtained from field 31. *APOE* ε2 and ε4 allele dosages were determined from rs7412 and rs429358 using WGS genotypes and, when missing, from whole-exome sequencing data. Genetic principal components were obtained from field 22009 (40 PCs). Family history was captured at assessment visits via self-report of first-degree relatives affected by “Alzheimer’s disease/dementia” or “Parkinson’s disease” (fields 20110 [mother], 20107 [father], and 20111 [siblings]). In release v20, maternal history counts were 51,659 for ADRD and 7,526 for PDRD; paternal history counts were 27,382 for ADRD and 10,819 for PDRD; and sibling history counts were 4,605 for ADRD and 3,164 for PDRD. UKB also provides algorithmically defined outcomes, including all-cause dementia (field 42018; N = 11,616) with confirmed Alzheimer’s disease (field 42020; N = 5,209), and parkinsonism (field 42030; N = 5,231) with confirmed Parkinson’s disease (field 42032; N = 4,737). For the primary UKB analyses, proxy-cases were defined as participants with either a AD (respectively PD) diagnosis (field 42020, respectively field 42032) and/or a first-degree family history of the corresponding condition; family history–based proxy phenotypes have been shown to increase statistical power in population cohorts.^7,53^ Analyses were conducted in an all-ancestry cohort (N = 484,909) and in a British ancestry subset defined by field 22006 (N = 406,872). For AD-combined-proxy, the all-ancestry cohort comprised 79,275 proxy cases and 400,648 controls, and the British subset comprised 68,885 proxy cases and 333,752 controls. For PD-combined-proxy, the all-ancestry cohort comprised 24,541 proxy cases and 459,924 controls, and the British subset comprised 21,130 proxy cases and 385,364 controls. Participant demographics are provided in **Supplementary Table 10**. Covariates included genetic sex, last known age, defined as age at the most recent EHR-recorded event or last UKB visit, and the 40 genetic principal components provided by UKB. ADRD analyses additionally adjusted for *APOE* ε2 and ε4 allele dosages; participants missing *APOE* genotypes at rs429358 and rs7412 were excluded. We additionally performed diagnosed-only sensitivity analyses in UKB using the corresponding algorithmically defined outcomes. For ADRD, diagnosed-only cases were defined using all-cause dementia (ADRD-direct), with confirmed Alzheimer’s disease used as a narrower AD sensitivity definition (AD-direct). For PDRD, diagnosed-only cases were defined using parkinsonism (PDRD-direct), with confirmed Parkinson’s disease used as a narrower PD sensitivity definition (PD-direct). Family-history-only proxy cases were not treated as cases in diagnosed-only analyses, and cleaned controls were required to have no diagnosis and no first-degree family history of the corresponding condition. **Supplementary Tables 11-12** provide exact case-control counts per analytical strata in UK Biobank and all cohorts.

#### All Of Us (AoU)

The All of Us (AoU) Research Program is a population-based cohort in the United States with linked whole-genome sequencing (WGS), electronic health record (EHR), and survey data.^17^ We used the v8 WGS release (N = 414,830). Participants were eligible for phenotype assignment if they passed AoU genetic quality control, had reported sex at birth, and had sufficient EHR or personal and family health history survey information to classify them as a case or control. Availability of phenotype information was distinguished from evidence of disease: interpretable negative survey responses supported control classification, whereas a qualifying EHR diagnosis code or positive personal or family history supported case classification. Missing or uninterpretable survey responses were not treated as negative responses. Participants without qualifying evidence for case status and without sufficient survey information for control assignment were excluded; absence of a relevant EHR code alone was not sufficient to establish control status. Direct cases were identified using the phenotype-specific EHR diagnosis-code sets and qualifying, phenotype-specific self-reported personal history (**Supplementary Tables 14–15**). For the primary AD-combined-proxy and PD-combined-proxy analyses, direct cases were restricted to participants meeting the AD and PD definitions, respectively. The combined-proxy analyses additionally included participants reporting an affected parent or sibling who did not meet the corresponding direct-case definition. Family-history information retained the scope of the original survey questions concerning dementia and Parkinson’s disease; it was not reinterpreted as confirmation of a specific neuropathological diagnosis. The broader ADRD and PDRD definitions were retained for diagnostic-sensitivity analyses. Controls were required to have no qualifying EHR diagnosis or self-reported personal history within the corresponding broad disease family and interpretable survey information indicating no affected parent or sibling. In the narrower combined-proxy analyses, participants meeting the broader ADRD or PDRD definition who met neither the corresponding narrow direct-case criteria nor the qualifying family-history criteria were excluded rather than reassigned as controls. In the direct-diagnosis analyses, family-history-only participants were excluded from both the case and control groups, retaining the corresponding clean-control definition. Before downstream filtering, N = 3,966 participants had an ADRD clinical code and N = 2,396 had a PDRD clinical code. Among participants reporting personal or first-degree family history, N = 28,736 reported dementia (self: 552; parent: 27,005; sibling: 2,448) and N = 7,555 reported Parkinson’s disease (self: 913; parent: 5,637; sibling: 1,362). Personal, parental and sibling history categories were not mutually exclusive. Analyses were conducted separately in the all-ancestry sample and a European-ancestry subset defined using the AoU ancestry inference pipeline.^17^ Final analytic sample sizes differed by phenotype. The AD-combined-proxy analysis included 29,097 cases and 190,229 controls in the all-ancestry sample, and 24,144 cases and 132,167 controls in the European-ancestry subset. The PD-combined-proxy analysis included 9,006 cases and 212,990 controls in the all-ancestry sample, and 7,582 cases and 150,370 controls in the European-ancestry subset. Combined-proxy case counts include both direct cases and family-history-only cases. Exact case-control counts across phenotype definitions are provided in **Supplementary Tables 11–12**, and participant demographics are provided in **Supplementary Table 13**. Covariates included reported sex at birth, last known age, defined as age at the most recent EHR-recorded event across 12 clinical tables, and the first 16 genetic principal components provided by AoU. AD and ADRD analyses additionally adjusted for *APOE* ε2 and ε4 allele dosages, determined from rs429358 and rs7412; participants missing the genotype information required to assign these dosages were excluded from those analyses.

### Alzheimer’s Disease Sequencing Project (ADSP)

We analyzed WGS data from the ADSP release 5, comprising 58,507 genomes from 67 contributing cohorts hosted at NIAGADS.^18,54^ Sample duplicates and relatedness were assessed using KING v2.3.^55^ and duplicate individuals were removed. AD cases were defined as participants with an adjudicated diagnosis of Alzheimer’s disease dementia according to ADSP/source-cohort criteria, supported by clinical assessment and, where available, AD biomarkers and/or neuropathological confirmation. Biomarker positivity alone, in the absence of a clinical diagnosis of AD dementia or neuropathological AD, was not sufficient to define an AD case. Participants with mild cognitive impairment, non-AD dementia diagnoses, or discordant biomarker/pathology information were excluded. Genetic ancestry was assigned using SNPweights with 1000 Genomes reference populations.^56^ Using an ancestry proportion threshold ≥50%, participants were grouped into five super-populations (South Asian, East Asian, Admixed American/Amerindian, African, and European), with remaining individuals classified as admixed. Primary analyses were performed in the combined all-ancestry cohort and within the European subset. Genetic principal components were computed separately within these two strata using PC-AiR.^57^ The all-ancestry cohort included 13,268 cases and 28,215 controls, and the European subset included 8,328 cases and 14,315 controls. Participant demographics and cohorts description are provided in **Supplementary Table 16-17**. Covariates included 10 PCs in the all-ancestry analysis and 5 PCs in the European-ancestry analysis, as well as sex and *APOE* ε2 and ε4 allele dosages. Age was not included as a covariate in ADSP because the available age variable is not directly comparable between cases and controls: for cases, age corresponds to the earliest known age at onset or age at examination, whereas for controls, age corresponds to the latest known age at examination or death. Adjustment for this non-comparable age variable could therefore introduce bias and reduce statistical power^58^. Additionally, following a reviewer’s request, we excluded all samples from Cardiff and Erasmus Rucphen Family (ERF), as we could not determine whether these were included in ADES.

### ADES/Holstege et al. - Stage 1

We used publicly available gene-based burden summary statistics from the stage 1 analysis reported by Holstege et al.,^3^ which included 12,652 cases and 8,693 controls aggregated across multiple whole-exome sequencing cohorts, including a subset of ADSP released via dbGaP (phs000572.v7.p4). Because individual-level data for the full stage 1 dataset are not publicly available, we used summary statistics. To avoid sample overlap between ADSP WGS release 5 and dbGaP WES releases, we identified duplicate individuals using KING^55^ and excluded 1,612 overlapping participants from the ADSP WGS analysis.

### Accelerating Medicine Partnership in Parkinson’s Disease and Related Disorders (AMP-PDRD)

The AMP-PDRD is a harmonized resource integrating multiple longitudinal PD and related disorder cohorts with uniformly processed genomic and clinical data^19^. We used AMP-PDRD release 4.0, comprising 10,910 participants, with phenotype breakdown reported in **Supplementary Table 18**. Diagnosis was obtained from the PD medical history module; when multiple diagnoses were present, the diagnosis recorded at the most recent visit was used. PDRD cases were defined as participants with a diagnosis of Parkinson’s disease, idiopathic PD, parkinsonism, dementia with Lewy bodies (DLB), Lewy body dementia (LBD), prodromal non-motor PD, and prodromal motor PD (N = 6,316). For participants with available parental history data (N = 8,119), we additionally classified proxy cases as individuals reporting one or more affected parents (N = 1,136). Controls were defined as participants with “No PD nor other neurological disorder” and no reported parental history of PD (N = 3,533). Ancestries were defined using SNPWeights with 1000 Genomes reference populations^56^. Covariates included self-reported sex, age at first available clinical assessment, and the first 16 genetic principal components. Final analytic sample sizes differed by phenotype definition. For the primary PD-combined-proxy analysis, the final analytic sample included 3,850 cases and 3,533 controls in the all-ancestry cohort, and 3,799 cases and 3,496 controls in the European ancestry subset. Participant demographics and cohorts description are provided in **Supplementary Tables 19-20**.

#### Variants annotation

Variants were annotated using the Ensembl Variant Effect Predictor (VEP v.114) using GRCh38 reference genome and Ensembl cache resources^59^. Functional annotation incorporated multiple prediction and constraint resources via VEP plugins and custom tracks, including LOFTEE loss-of-function classification^20^, and REVEL missense pathogenicity prediction^21^, and population allele frequencies and filters from gnomAD v4.1^60^. Multiallelic variants were decomposed into biallelic variants prior to annotation. Variants were restricted to protein-coding genes. Rare variants were defined as minor allele frequency (MAF) < 1% within cohort and reference populations. Variants were grouped into four hierarchical deleteriousness sets based on functional annotation and REVEL prediction score: Loss-of-function (LOF; HIGH impact consequence with LOFTEE high-confidence classification^20^); LOF or missense variants with REVEL ≥ 0.75; LOF or missense variants with REVEL ≥ 0.50, LOF or missense variants with REVEL ≥ 0.25. Variants with missing REVEL scores were not assigned a default value and were excluded from REVEL-thresholded missense masks. We only retain the LOF category for these 2,398 genes with no additional variants in the LOF+REVEL masks. Set assignment and generation of REGENIE annotation, set definition, and gene set files were performed using custom Python pipelines.

#### Variants Quality Control

Across all cohorts, variants were restricted to gnomAD v4.1 FILTER = PASS, when available, and cohort-specific genotype quality filters, including Hardy–Weinberg equilibrium (p > 10^-8^) and genotype missingness (<10%). Multiallelic variants were split into separate biallelic variants using bcftools.

UKB WGS data sequencing and quality control is described elsewhere^16^. Data were released in pgen/pvar on the UKB research analysis platform; variants with a PASS flag were considered for analysis, while others were excluded.

AllOfUs WGS data sequencing and quality control is described elsewhere^17^. Data were released in pgen/pvar format solely for three smaller callsets, including ACAF (Allele Count/Allele Frequency thresholds, MAF > 1% or Allele Count > 100), ClinVar, and Exome. For our purpose, we used the ACAF set for REGENIE step 1 and the Exome set for REGENIE step 2, described in the next section.

For ADSP WGS, we used centrally released QC-passed genotype data in PLINK2 pgen/pvar format and implemented additional burden-specific variant QC. Sample duplication and relatedness were evaluated using KING, and sample inclusion additionally considered sex concordance, excess heterozygosity, and ancestry concordance with clinical race/ethnicity. Because most rare variants have only a small number of carriers, we did not define new sample-level contamination, depth, or missingness filters from the rare-variant burden set itself. Instead, we implemented carrier-aware variant-level QC focused on the genotypes directly contributing to rare-variant burden masks. For each rare variant considered for burden analysis (1,342,742 total variants, including 283,239 predicted loss-of-function variants), we extracted genotype quality (GQ), allelic depth (AD) for reference and alternate alleles, and read depth (DP) across alternate-allele carriers. For heterozygous carriers, we computed variant allelic fraction as VAF = AD_ALT_ / DP. For each variant, we summarized carrier genotype support using both mean and median values for genotype quality, alternate allele depth, and variant allele fraction (GQ_mean_, GQ_median, AD_ALT_mean_, AD_ALT_median_, VAF_mean_, VAF_median_). Empirical filtering thresholds were derived without reference to phenotype or association statistics. For each metric, we estimated the empirical density distribution and identified the lower-quality tail; the local density minimum separating this tail from the primary high-quality distribution was used as the filtering threshold. The final thresholds were GQ_mean_ <76.22, GQ_median_ <64.03, AD_ALT_mean_ <7.28 reads, AD_ALT_median_ <7.28 reads, VAF_mean_ <0.238, and VAF_median_ <0.238. Variants were excluded if any of these carrier-level support summaries fell below the corresponding threshold. **Supplementary Fig. 16-17** shows the carrier-metric distributions, selected thresholds, and variant-exclusion counts. We additionally excluded ADSP variants overlapping short tandem repeat, low-complexity, low-copy repeat, or segmental-duplication regions using GRCh38 region masks. To evaluate batch-like missingness differences between cases and controls, we also performed variant-level differential missingness tests within ADSP and excluded variants showing differential missingness at Fisher exact test P < 10^-5^, with an absolute case-control missingness difference >0.001, in either the full ADSP analysis set or the EUR subset. The combined ADSP exclusion list from repetitive/segmental-duplication region overlap and differential missingness contained N = 70,194 variants, and the final ADSP burden-test variant set after all filters contained N = 1,142,266 variants.

AMP-PDRD WGS was downloaded in pgen/pvar format with pre-processed quality control flags^19^. In brief, quality control was based on contamination, coverage, Ti/Tv outliers, missingness, sample duplication, sex concordance, excess heterogeneity, and ancestry concordance with clinical race/ethnicity. Samples and variants failing any filter were excluded.

#### Statistical Analyses

Rare variant association testing was performed using REGENIE v4.1 two-step whole-genome regression^22^. Step 1 ridge regression models were trained using ancestry/subset-specific LD-pruned common variants selected with PLINK2. Variants were restricted to MAF ≥1% and pruned using --indep-pairwise 1000 100 0.1, corresponding to an LD threshold of r² = 0.1. Prior to Step 1 model fitting, long-range LD regions, low-complexity regions, variants failing genotype quality control, and variants with Hardy–Weinberg equilibrium P <10^−8^ were excluded. Predictions from Step 1 were used as offsets in Step 2 association testing. Step 2 gene-based tests were performed using binary logistic models. Association models were fit separately within each cohort and analysis stratum. Covariates were cohort-specific, as described above: biobank analyses included age, sex, genetic principal components, and *APOE* ε2/ε4 dosages for ADRD analyses; AMP-PDRD analyses included age at first available clinical assessment, sex, and genetic principal components; and ADSP analyses included sex, APOE ε2/ε4 dosages, sequencing-platform indicators, and genetic principal components. For binary traits, Firth correction with approximate inference was applied to reduce bias from case-control imbalance and sparse carrier counts. Gene-based rare variant tests were performed using burden tests restricted to variants with allele frequency <1%. Tests were performed independently for each nested deleteriousness set: LOF, LOF+REVEL ≥0.75, LOF+REVEL ≥0.50, and LOF+REVEL ≥0.25. Gene/mask results were retained for downstream meta-analysis only when the cumulative minor allele count across contributing cohorts was >10. For non-redundant reporting of gene-level results, when multiple nested masks within the same gene reached a reporting threshold, the most significant gene/mask association was prioritized unless otherwise specified.

#### Meta-Analyses

Gene-level results were meta-analyzed separately for the all-ancestry cohorts and European-ancestry subsets. The primary discovery analyses were the all-ancestry AD-combined-proxy and PD-combined-proxy meta-analyses. For ADRD, the REGENIE component included UKB, AoU, and ADSP, with additional statistical evidence incorporated from the publicly available ADES stage 1 summary statistics from Holstege et al.^3^, as described below. For PDRD, the primary meta-analysis included UKB, AoU, and AMP-PDRD. In the population-biobank cohorts, the combined-proxy phenotypes included participants with a diagnosed, self-reported, or clinically ascertained phenotype and unaffected participants reporting an affected first-degree relative. Family-history-only cases did not have a personal diagnosis, and clean controls had neither a personal diagnosis nor a reported affected first-degree relative.

We additionally performed secondary direct-diagnosis-based meta-analyses restricted to diagnosed, self-reported, or clinically ascertained cases. The meta-analyses included AD, ADRD, PD and PDRD under combined-proxy and direct-diagnosis definitions, each evaluated in all-ancestry and European-ancestry strata. The broader ADRD/PDRD analyses were retained only as diagnostic-sensitivity comparisons. The all-ancestry AD-combined and PD-combined-proxy analyses were prespecified as the primary discovery analyses. European-ancestry and direct-diagnosis-based analyses were considered ancestry- and phenotype-sensitivity analyses, respectively.

For binary-trait REGENIE results requiring Firth correction, we used standard errors compatible with the reported likelihood-ratio association evidence. For previously generated Firth-corrected results without --firth-se, we reconstructed an equivalent normal-scale standard error from the coefficient and two-sided likelihood-ratio *P* value as

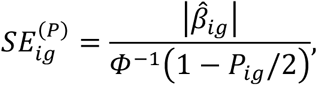

where *i* indexes the cohort and *g* indexes the gene/deleteriousness-mask pair. For results not requiring Firth correction, the ordinary REGENIE standard error was retained. This procedure ensured that the coefficient, standard error, and association evidence supplied to the meta-analysis were internally aligned. Further details are provided in **Supplementary Note 2**.

Before meta-analysis, combined-proxy REGENIE effects were placed on a common approximate directly diagnosed scale using a cohort-, phenotype-, and ancestry-specific common-control mixture model. For each combined-proxy analysis, we calculated

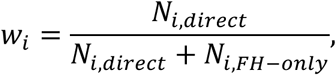

where *N_i_*_,*direct*_ is the number of diagnosed, self-reported, or clinically ascertained cases and *N_i_*_,*FH*−*only*_ is the number of unaffected family-history-only cases. Let *β_M_*_,*i*_ denote the observed combined-proxy log-odds effect, *β_D_*_,*i*_ the corresponding effect on the directly diagnosed scale, and *κ* the effective attenuation of the family-history-only effect relative to the directly diagnosed effect. Under the common-control mixture model,

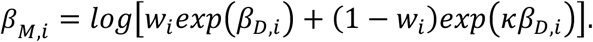

The central analysis used *κ* = 0.5. Rather than applying a uniform factor-of-two correction or its first-order mixture approximation, we obtained the adjusted coefficient by exact nonlinear inversion of this equation. The adjusted standard error was calculated using the delta method and the derivative of the forward mixture transformation. Directly diagnosed cohorts required no adjustment and were represented by *w_i_* = 1. We evaluated the limitations of the fixed κ=0.5 attenuation assumption using nuclear-family simulations that varied family-history reporting sensitivity and specificity and quantified the resulting attenuation and adjustment error (**Supplementary Note 1; Supplementary Fig. 18**). The mathematical derivation and interpretation of this adjustment are provided in **Supplementary Note 1**. Cohort- and subset-specific case and control counts and *w_i_* values are reported in **Supplementary Tables 11-12**.

Only cohorts analyzed using the compatible REGENIE maximum-allele, binary-trait burden framework were included in the pooled effect-size meta-analysis. For ADRD, these cohorts were UKB, AoU, and ADSP; for PDRD, they were UKB, AoU, and AMP-PDRD. Adjusted coefficients and standard errors were combined using inverse-variance-weighted fixed-effect models. For gene/mask pair *g*, the cohort-specific weight was

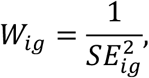

and the pooled REGENIE coefficient and standard error were

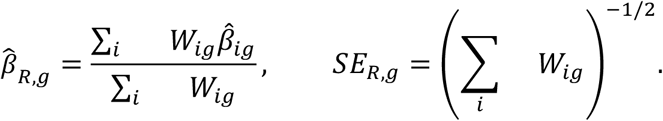

Two-sided *P* values were derived from 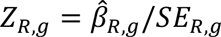. Pooled odds ratios and 95% confidence intervals were calculated as 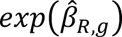 and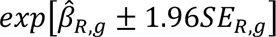, respectively. Between-cohort heterogeneity was evaluated among the compatible REGENIE estimates using Cochran’s *Q* statistic, the corresponding heterogeneity *P* value, and *I*^2^.

The ADES stage 1 results were not included as an additional coefficient in the inverse-variance effect-size meta-analysis. ADES used an additive summed-dosage ordinal logistic model in which the outcome was ordered as controls, late-onset Alzheimer’s disease, and early-onset Alzheimer’s disease, whereas the REGENIE cohorts used binary outcomes and maximum-allele burden masks. Consequently, the ADES coefficient, skewed for early-onset genes, and the pooled REGENIE coefficient do not have the same numerical interpretation or measurement unit and were not treated as directly commensurate log-odds estimates.

Instead, ADES was incorporated into the ADRD analyses through a weighted signed-*Z* evidence combination. The pooled REGENIE and ADES likelihood-ratio *P* values were converted to signed standardized statistics using the direction of their respective coefficients:

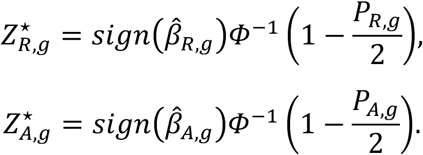

The two statistics were combined using fixed information weights based on the effective sample size,

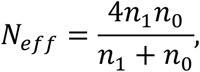

where *n*_1_ and *n*_0_ denote the numbers of cases and controls, respectively. Effective sample sizes were summed across the independent REGENIE cohorts, and the early- and late-onset AD cases were combined solely for calculation of the ADES information weight. With weights *u_R_* = 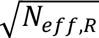 and 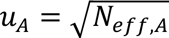, the ADES-inclusive statistic was

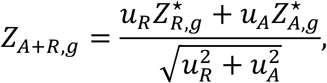

with two-sided *P_A_*_+*R*,g_ = 2*Φ*(−%*Z_A_*_+*R*,g_%). These weights were determined solely from phenotype counts and were independent of the observed gene-level association results. The ADES-inclusive statistic was calculated for every eligible gene/mask pair rather than only for gene/mask pairs selected from the REGENIE analysis.

The ADES-inclusive statistic provides a combined test of gene-level association but does not define an ADES-plus-REGENIE pooled odds ratio. Accordingly, the pooled REGENIE odds ratio was used as the principal meta-analytic effect-size estimate, while the ADES ordinal odds ratio was reported separately. Cochran’s *Q*, *I*^2^, and heterogeneity *P* values were calculated only among the compatible REGENIE cohort estimates and did not treat the ADES ordinal coefficient as an additional binary-trait log-odds estimate. For ADRD signals interpreted as shared risk associations, we additionally required concordant effect directions between the pooled REGENIE and ADES results. Additional details of the REGENIE meta-analysis and ADES evidence-combination procedure are provided in **Supplementary Note 2**.

To account for the strong correlation among the four nested deleteriousness masks, we estimated the effective number of independent tests using the Li and Ji eigenvalue method applied to the correlation structure of the gene-level statistics.^61^ The resulting study-wide significance threshold for the primary all-ancestry analysis was *P* < 1.20 × 10^−6^.

We used this threshold as a conservative study-wide benchmark for the primary and sensitivity analyses. The all-ancestry AD-combined-proxy and PD-combined-proxy analyses were used for primary discovery claims. European-ancestry analyses were secondary, AD-direct and PD-direct analyses were nested phenotype-sensitivity analyses, and all broader ADRD/PDRD analyses were diagnostic-sensitivity comparisons. We defined *P* < 10^−5^ as a suggestive threshold, motivated by the approximate unadjusted *P*-value range corresponding to the false-discovery-rate threshold used in the previous large AD rare-variant burden analysis.^3^ Gene/mask associations with 1.20 × 10^−6^ ≤ *P* < 10^−5^ were therefore described as suggestive or near the study-wide threshold.

We classified genes as established or previously implicated based on prior Mendelian, family-based, rare-variant or gene-implicating GWAS evidence for the relevant disease family. Novel associations required study-wide significance in a primary all-ancestry AD-combined-proxy or PD-combined-proxy analysis and no such prior evidence. *ADAM10* and *GRN* were not considered novel; secondary and suggestive findings were labeled separately.

To evaluate calibration and robustness, we generated quantile-quantile plots and calculated genomic inflation factors for each cohort-level analysis and each meta-analysis (**Supplementary Figs. 1 and 9**). We also compared combined-proxy-based and direct-diagnosis-based pooled effect estimates for gene/mask pairs reaching *P* < 1.20 × 10^−6^ in either analysis, separately for ADRD and PDRD and within the all-ancestry and European-ancestry strata. Because the ADES-inclusive procedure produces a combined significance test rather than a pooled effect estimate, ADRD combined-proxy versus direct-diagnosis effect-size comparisons were based on the pooled REGENIE coefficients and odds ratios, with the corresponding ADES estimates presented separately.

## Funding and acknowledgments

This work was supported by the National Institute of Health and National Institute of Aging grants AG072290 (MDG), AG066515 (MDG). UK Biobank WGS data were analyzed under Application Number 45420. We gratefully acknowledge *All of Us* participants for their contributions, without whom this research would not have been possible. We also thank the National Institutes of Health’s [*All of Us* Research Program](https://allofus.nih.gov/) for making available the participant data examined in this study.

## AMP-PD Acknowledgment Statement

Data used in the preparation of this article were obtained from the Accelerating Medicine Partnership® (AMP®) Parkinson’s Disease (AMP PD) Knowledge Platform. For up-to-date information on the study, visit https://www.amp-pd.org. The AMP® PD program is a public-private partnership managed by the Foundation for the National Institutes of Health and funded by the National Institute of Neurological Disorders and Stroke (NINDS) in partnership with the Aligning Science Across Parkinson’s (ASAP) initiative; Celgene Corporation, a subsidiary of Bristol-Myers Squibb Company; GlaxoSmithKline plc (GSK); The Michael J. Fox Foundation for Parkinson’s Research ; Pfizer Inc.; AbbVie Inc.; Sanofi US Services Inc.; and Verily Life Sciences. ACCELERATING MEDICINES PARTNERSHIP and AMP are registered service marks of the U.S. Department of Health and Human Services. Clinical data and biosamples used in preparation of this article were obtained from the (i) Michael J. Fox Foundation for Parkinson’s Research (MJFF) and National Institutes of Neurological Disorders and Stroke (NINDS) BioFIND study, (ii) Harvard Biomarkers Study (HBS) and the Stephen & Denise Adams Center for Parkinson’s Disease Research of Yale School of Medicine (CPDR-Y), (iii) National Institute on Aging (NIA) International Lewy Body Dementia Genetics Consortium Genome Sequencing in Lewy Body Dementia Case-control Cohort (LBD), (iv) MJFF LRRK2 Cohort Consortium (LCC), (v) NINDS Parkinson’s Disease Biomarkers Program (PDBP), (vi) MJFF Parkinson’s Progression Markers Initiative (PPMI), and (vii) NINDS Study of Isradipine as a Disease-modifying Agent in Subjects With Early Parkinson Disease, Phase 3 (STEADY-PD3) and (viii) the NINDS Study of Urate Elevation in Parkinson’s Disease, Phase 3 (SURE-PD3). BioFIND is sponsored by The Michael J. Fox Foundation for Parkinson’s Research (MJFF) with support from the National Institute for Neurological Disorders and Stroke (NINDS). The BioFIND Investigators have not participated in reviewing the data analysis or content of the manuscript. For up-to-date information on the study, visit michaeljfox.org/news/biofind. Genome sequence data for the Lewy body dementia case-control cohort were generated at the Intramural Research Program of the U.S. National Institutes of Health. The study was supported in part by the National Institute on Aging (program #: 1ZIAAG000935) and the National Institute of Neurological Disorders and Stroke (program #: 1ZIANS003154). The Harvard Biomarker Study (HBS) is a collaboration of HBS investigators [full list of HBS investigators found at https://www.bwhparkinsoncenter.org/biobank/ and funded through philanthropy and NIH and Non-NIH funding sources. The Stephen & Denise Adams Center for Parkinson’s Disease Research of Yale School of Medicine is funded through philanthropy and NIH and non-NIH funding sources. The HBS and CPDR-Y Investigators have not participated in reviewing the data analysis or content of the manuscript. Data used in preparation of this article were obtained from The Michael J. Fox Foundation sponsored LRRK2 Cohort Consortium (LCC). The LCC Investigators have not participated in reviewing the data analysis or content of the manuscript. For up-to-date information on the study, visit https://www.michaeljfox.org/biospecimens). PPMI is sponsored by The Michael J. Fox Foundation for Parkinson’s Research and supported by a consortium of scientific partners: The PPMI investigators have not participated in reviewing the data analysis or content of the manuscript. For up-to-date information on the study, visit www.ppmi-info.org. The Parkinson’s Disease Biomarker Program (PDBP) consortium is supported by the National Institute of Neurological Disorders and Stroke (NINDS) at the National Institutes of Health. A full list of PDBP investigators can be found at https://pdbp.ninds.nih.gov/policy. The PDBP investigators have not participated in reviewing the data analysis or content of the manuscript. The Study of Isradipine as a Disease-modifying Agent in Subjects With Early Parkinson Disease, Phase 3 (STEADY-PD3) is funded by the National Institute of Neurological Disorders and Stroke (NINDS) at the National Institutes of Health with support from The Michael J. Fox Foundation and the Parkinson Study Group. For additional study information, visit https://clinicaltrials.gov/ct2/show/study/NCT02168842. The STEADY-PD3 investigators have not participated in reviewing the data analysis or content of the manuscript. The Study of Urate Elevation in Parkinson’s Disease, Phase 3 (SURE-PD3) is funded by the National Institute of Neurological Disorders and Stroke (NINDS) at the National Institutes of Health with support from The Michael J. Fox Foundation and the Parkinson Study Group. For additional study information, visit https://clinicaltrials.gov/ct2/show/NCT02642393. The SURE-PD3 investigators have not participated in reviewing the data analysis or content of the manuscript.

## ADSP Acknowledgement Statement

The Alzheimer’s Disease Sequencing Project (ADSP) is comprised of two Alzheimer’s Disease (AD) genetics consortia and three National Human Genome Research Institute (NHGRI) funded Large Scale Sequencing and Analysis Centers (LSAC). The two AD genetics consortia are the Alzheimer’s Disease Genetics Consortium (ADGC) funded by NIA (U01 AG032984), and the Cohorts for Heart and Aging Research in Genomic Epidemiology (CHARGE) funded by NIA (R01 AG033193), the National Heart, Lung, and Blood Institute (NHLBI), other National Institute of Health (NIH) institutes and other foreign governmental and non-governmental organizations. The Discovery Phase analysis of sequence data is supported through UF1AG047133 (to Drs. Schellenberg, Farrer, Pericak-Vance, Mayeux, and Haines); U01AG049505 to Dr. Seshadri; U01AG049506 to Dr. Boerwinkle; U01AG049507 to Dr. Wijsman; and U01AG049508 to Dr. Goate and the Discovery Extension Phase analysis is supported through U01AG052411 to Dr. Goate, U01AG052410 to Dr. Pericak-Vance and U01 AG052409 to Drs. Seshadri and Fornage.

Sequencing for the Follow Up Study (FUS) is supported through U01AG057659 (to Drs. PericakVance, Mayeux, and Vardarajan) and U01AG062943 (to Drs. Pericak-Vance and Mayeux). Data generation and harmonization in the Follow-up Phase is supported by U54AG052427 (to Drs. Schellenberg and Wang). The FUS Phase analysis of sequence data is supported through U01AG058589 (to Drs. Destefano, Boerwinkle, De Jager, Fornage, Seshadri, and Wijsman), U01AG058654 (to Drs. Haines, Bush, Farrer, Martin, and Pericak-Vance), U01AG058635 (to Dr. Goate), RF1AG058066 (to Drs. Haines, Pericak-Vance, and Scott), RF1AG057519 (to Drs. Farrer and Jun), R01AG048927 (to Dr. Farrer), and RF1AG054074 (to Drs. Pericak-Vance and Beecham).

The ADGC cohorts include: Adult Changes in Thought (ACT) (U01 AG006781, U19 AG066567), the Alzheimer’s Disease Research Centers (ADRC) (P30 AG062429, P30 AG066468, P30 AG062421, P30 AG066509, P30 AG066514, P30 AG066530, P30 AG066507, P30 AG066444, P30 AG066518, P30 AG066512, P30 AG066462, P30 AG072979, P30 AG072972, P30 AG072976, P30 AG072975, P30 AG072978, P30 AG072977, P30 AG066519, P30 AG062677, P30 AG079280, P30 AG062422, P30 AG066511, P30 AG072946, P30 AG062715, P30 AG072973, P30 AG066506, P30 AG066508, P30 AG066515, P30 AG072947, P30 AG072931, P30 AG066546, P20 AG068024, P20 AG068053, P20 AG068077, P20 AG068082, P30 AG072958, P30 AG072959), the Chicago Health and Aging Project (CHAP) (R01 AG11101, RC4 AG039085, K23 AG030944), Indiana Memory and Aging Study (IMAS) (R01 AG019771), Indianapolis Ibadan (R01 AG009956, P30 AG010133), the Memory and Aging Project (MAP) ( R01 AG17917), Mayo Clinic (MAYO) (R01 AG032990, U01 AG046139, R01 NS080820, RF1 AG051504, P50 AG016574), Mayo Parkinson’s Disease controls (NS039764, NS071674, 5RC2HG005605), University of Miami (R01 AG027944, R01 AG028786, R01 AG019085, IIRG09133827, A2011048), the Multi-Institutional Research in Alzheimer’s Genetic Epidemiology Study (MIRAGE) (R01 AG09029, R01 AG025259), the National Centralized Repository for Alzheimer’s Disease and Related Dementias (NCRAD) (U24 AG021886), the National Institute on Aging Late Onset Alzheimer’s Disease Family Study (NIA-LOAD) (U24 AG056270), the Religious Orders Study (ROS) (P30 AG10161, R01 AG15819), the Texas Alzheimer’s Research and Care Consortium (TARCC) (funded by the Darrell K Royal Texas Alzheimer’s Initiative), Vanderbilt University/Case Western Reserve University (VAN/CWRU) (R01 AG019757, R01 AG021547, R01 AG027944, R01 AG028786, P01 NS026630, and Alzheimer’s Association), the Washington Heights-Inwood Columbia Aging Project (WHICAP) (RF1 AG054023), the University of Washington Families (VA Research Merit Grant, NIA: P50AG005136, R01AG041797, NINDS: R01NS069719), the Columbia University Hispanic Estudio Familiar de Influencia Genetica de Alzheimer (EFIGA) (RF1 AG015473), the University of Toronto (UT) (funded by Wellcome Trust, Medical Research Council, Canadian Institutes of Health Research), and Genetic Differences (GD) (R01 AG007584). The CHARGE cohorts are supported in part by National Heart, Lung, and Blood Institute (NHLBI) infrastructure grant HL105756 (Psaty), RC2HL102419 (Boerwinkle) and the neurology working group is supported by the National Institute on Aging (NIA) R01 grant AG033193.

The CHARGE cohorts participating in the ADSP include the following: Austrian Stroke Prevention Study (ASPS), ASPS-Family study, and the Prospective Dementia Registry-Austria (ASPS/PRODEM-Aus), the Atherosclerosis Risk in Communities (ARIC) Study, the Cardiovascular Health Study (CHS), the Erasmus Rucphen Family Study (ERF), the Framingham Heart Study (FHS), and the Rotterdam Study (RS). ASPS is funded by the Austrian Science Fond (FWF) grant number P20545-P05 and P13180 and the Medical University of Graz. The ASPS-Fam is funded by the Austrian Science Fund (FWF) project I904), the EU Joint Programme – Neurodegenerative Disease Research (JPND) in frame of the BRIDGET project (Austria, Ministry of Science) and the Medical University of Graz and the Steiermärkische Krankenanstalten Gesellschaft. PRODEM-Austria is supported by the Austrian Research Promotion agency (FFG) (Project No. 827462) and by the Austrian National Bank (Anniversary Fund, project 15435. ARIC research is carried out as a collaborative study supported by NHLBI contracts (HHSN268201100005C, HHSN268201100006C, HHSN268201100007C, HHSN268201100008C, HHSN268201100009C, HHSN268201100010C, HHSN268201100011C, and HHSN268201100012C). Neurocognitive data in ARIC is collected by U01 2U01HL096812, 2U01HL096814, 2U01HL096899, 2U01HL096902, 2U01HL096917 from the NIH (NHLBI, NINDS, NIA and NIDCD), and with previous brain MRI examinations funded by R01-HL70825 from the NHLBI. CHS research was supported by contracts HHSN268201200036C, HHSN268200800007C, N01HC55222, N01HC85079, N01HC85080, N01HC85081, N01HC85082, N01HC85083, N01HC85086, and grants U01HL080295 and U01HL130114 from the NHLBI with additional contribution from the National Institute of Neurological Disorders and Stroke (NINDS). Additional support was provided by R01AG023629, R01AG15928, and R01AG20098 from the NIA. FHS research is supported by NHLBI contracts N01-HC-25195 and HHSN268201500001I. This study was also supported by additional grants from the NIA (R01s AG054076, AG049607 and AG033040 and NINDS (R01 NS017950). The ERF study as a part of EUROSPAN (European Special Populations Research Network) was supported by European Commission FP6 STRP grant number 018947 (LSHG-CT-2006-01947) and also received funding from the European Community’s Seventh Framework Programme (FP7/2007-2013)/grant agreement HEALTH-F4-2007-201413 by the European Commission under the programme “Quality of Life and Management of the Living Resources” of 5th Framework Programme (no. QLG2-CT-2002-01254). High-throughput analysis of the ERF data was supported by a joint grant from the Netherlands Organization for Scientific Research and the Russian Foundation for Basic Research (NWO-RFBR 047.017.043). The Rotterdam Study is funded by Erasmus Medical Center and Erasmus University, Rotterdam, the Netherlands Organization for Health Research and Development (ZonMw), the Research Institute for Diseases in the Elderly (RIDE), the Ministry of Education, Culture and Science, the Ministry for Health, Welfare and Sports, the European Commission (DG XII), and the municipality of Rotterdam. Genetic data sets are also supported by the Netherlands Organization of Scientific Research NWO Investments (175.010.2005.011, 911-03-012), the Genetic Laboratory of the Department of Internal Medicine, Erasmus MC, the Research Institute for Diseases in the Elderly (014-93-015; RIDE2), and the Netherlands Genomics Initiative (NGI)/Netherlands Organization for Scientific Research (NWO) Netherlands Consortium for Healthy Aging (NCHA), project 050-060-810. All studies are grateful to their participants, faculty and staff. The content of these manuscripts is solely the responsibility of the authors and does not necessarily represent the official views of the National Institutes of Health or the U.S. Department of Health and Human Services.

The FUS cohorts include: the Alzheimer’s Disease Research Centers (ADRC) (P30 AG062429, P30 AG066468, P30 AG062421, P30 AG066509, P30 AG066514, P30 AG066530, P30 AG066507, P30 AG066444, P30 AG066518, P30 AG066512, P30 AG066462, P30 AG072979, P30 AG072972, P30 AG072976, P30 AG072975, P30 AG072978, P30 AG072977, P30 AG066519, P30 AG062677, P30 AG079280, P30 AG062422, P30 AG066511, P30 AG072946, P30 AG062715, P30 AG072973, P30 AG066506, P30 AG066508, P30 AG066515, P30 AG072947, P30 AG072931, P30 AG066546, P20 AG068024, P20 AG068053, P20 AG068077, P20 AG068082, P30 AG072958, P30 AG072959), Alzheimer’s Disease Neuroimaging Initiative (ADNI) (U19AG024904), Amish Protective Variant Study (RF1AG058066), Cache County Study (R01AG11380, R01AG031272, R01AG21136, RF1AG054052), Case Western Reserve University Brain Bank (CWRUBB) (P50AG008012), Case Western Reserve University Rapid Decline (CWRURD) (RF1AG058267, NU38CK000480), CubanAmerican Alzheimer’s Disease Initiative (CuAADI) (3U01AG052410), Estudio Familiar de Influencia Genetica en Alzheimer (EFIGA) (5R37AG015473, RF1AG015473, R56AG051876), Genetic and Environmental Risk Factors for Alzheimer Disease Among African Americans Study (GenerAAtions) (2R01AG09029, R01AG025259, 2R01AG048927), Gwangju Alzheimer and Related Dementias Study (GARD) (U01AG062602), Hillblom Aging Network (2014-A-004-NET, R01AG032289, R01AG048234), Hussman Institute for Human Genomics Brain Bank (HIHGBB) (R01AG027944, Alzheimer’s Association “Identification of Rare Variants in Alzheimer Disease”), Ibadan Study of Aging (IBADAN) (5R01AG009956), Longevity Genes Project (LGP) and LonGenity (R01AG042188, R01AG044829, R01AG046949, R01AG057909, R01AG061155, P30AG038072), Mexican Health and Aging Study (MHAS) (R01AG018016), Multi-Institutional Research in Alzheimer’s Genetic Epidemiology (MIRAGE) (2R01AG09029, R01AG025259, 2R01AG048927), Northern Manhattan Study (NOMAS) (R01NS29993), Peru Alzheimer’s Disease Initiative (PeADI) (RF1AG054074), Puerto Rican 1066 (PR1066) (Wellcome Trust (GR066133/GR080002), European Research Council (340755)), Puerto Rican Alzheimer Disease Initiative (PRADI) (RF1AG054074), Reasons for Geographic and Racial Differences in Stroke (REGARDS) (U01NS041588), Research in African American Alzheimer Disease Initiative (REAAADI) (U01AG052410), the Religious Orders Study (ROS) (P30 AG10161, P30 AG72975, R01 AG15819, R01 AG42210), the RUSH Memory and Aging Project (MAP) (R01 AG017917, R01 AG42210Stanford Extreme Phenotypes in AD (R01AG060747), University of Miami Brain Endowment Bank (MBB), University of Miami/Case Western/North Carolina A&T African American (UM/CASE/NCAT) (U01AG052410, R01AG028786), Wisconsin Registry for Alzheimer’s Prevention (WRAP) (R01AG027161 and R01AG054047), Mexico-Southern California Autosomal Dominant Alzheimer’s Disease Consortium (R01AG069013), Center for Cognitive Neuroscience and Aging (R01AG047649), and the A4 Study (R01AG063689, U19AG010483 and U24AG057437).

The four LSACs are: the Human Genome Sequencing Center at the Baylor College of Medicine (U54 HG003273), the Broad Institute Genome Center (U54HG003067), The American Genome Center at the Uniformed Services University of the Health Sciences (U01AG057659), and the Washington University Genome Institute (U54HG003079). Genotyping and sequencing for the ADSP FUS is also conducted at John P. Hussman Institute for Human Genomics (HIHG) Center for Genome Technology (CGT).

Biological samples and associated phenotypic data used in primary data analyses were stored at Study Investigators institutions, and at the National Centralized Repository for Alzheimer’s Disease and Related Dementias (NCRAD, U24AG021886) at Indiana University funded by NIA. Associated Phenotypic Data used in primary and secondary data analyses were provided by Study Investigators, the NIA funded Alzheimer’s Disease Centers (ADCs), and the National Alzheimer’s Coordinating Center (NACC, U24AG072122) and the National Institute on Aging Genetics of Alzheimer’s Disease Data Storage Site (NIAGADS, U24AG041689) at the University of Pennsylvania, funded by NIA. Harmonized phenotypes were provided by the ADSP Phenotype Harmonization Consortium (ADSP-PHC), funded by NIA (U24 AG074855, U01 AG068057 and R01 AG059716) and Ultrascale Machine Learning to Empower Discovery in Alzheimer’s Disease Biobanks (AI4AD, U01 AG068057). This research was supported in part by the Intramural Research Program of the National Institutes of health, National Library of Medicine. Contributors to the Genetic Analysis Data included Study Investigators on projects that were individually funded by NIA, and other NIH institutes, and by private U.S. organizations, or foreign governmental or nongovernmental organizations.

The ADSP Phenotype Harmonization Consortium (ADSP-PHC) is funded by NIA (U24 AG074855, U01 AG068057 and R01 AG059716). The harmonized cohorts within the ADSP-PHC include: the Anti-Amyloid Treatment in Asymptomatic Alzheimer’s study (A4 Study), a secondary prevention trial in preclinical Alzheimer’s disease, aiming to slow cognitive decline associated with brain amyloid accumulation in clinically normal older individuals. The A4 Study is funded by a public-private-philanthropic partnership, including funding from the National Institutes of Health-National Institute on Aging, Eli Lilly and Company, Alzheimer’s Association, Accelerating Medicines Partnership, GHR Foundation, an anonymous foundation and additional private donors, with in-kind support from Avid and Cogstate. The companion observational Longitudinal Evaluation of Amyloid Risk and Neurodegeneration (LEARN) Study is funded by the Alzheimer’s Association and GHR Foundation. The A4 and LEARN Studies are led by Dr. Reisa Sperling at Brigham and Women’s Hospital, Harvard Medical School and Dr. Paul Aisen at the Alzheimer’s Therapeutic Research Institute (ATRI), University of Southern California. The A4 and LEARN Studies are coordinated by ATRI at the University of Southern California, and the data are made available through the Laboratory for Neuro Imaging at the University of Southern California. The participants screening for the A4 Study provided permission to share their de-identified data in order to advance the quest to find a successful treatment for Alzheimer’s disease. We would like to acknowledge the dedication of all the participants, the site personnel, and all of the partnership team members who continue to make the A4 and LEARN Studies possible. The complete A4 Study Team list is available on: a4study.org/a4-study-team.; the Adult Changes in Thought study (ACT), U01 AG006781, U19 AG066567; Alzheimer’s Disease Neuroimaging Initiative (ADNI): Data collection and sharing for this project was funded by the Alzheimer’s Disease Neuroimaging Initiative (ADNI) (National Institutes of Health Grant U01 AG024904) and DOD ADNI (Department of Defense award number W81XWH-12-2-0012). ADNI is funded by the National Institute on Aging, the National Institute of Biomedical Imaging and Bioengineering, and through generous contributions from the following: AbbVie, Alzheimer’s Association; Alzheimer’s Drug Discovery Foundation; Araclon Biotech; BioClinica, Inc.; Biogen; Bristol-Myers Squibb Company; CereSpir, Inc.; Cogstate; Eisai Inc.; Elan Pharmaceuticals, Inc.; Eli Lilly and Company; EuroImmun; F. Hoffmann-La Roche Ltd and its affiliated company Genentech, Inc.; Fujirebio; GE Healthcare; IXICO Ltd.;Janssen Alzheimer Immunotherapy Research & Development, LLC.; Johnson & Johnson Pharmaceutical Research & Development LLC.; Lumosity; Lundbeck; Merck & Co., Inc.;Meso Scale Diagnostics, LLC.; NeuroRx Research; Neurotrack Technologies; Novartis Pharmaceuticals Corporation; Pfizer Inc.; Piramal Imaging; Servier; Takeda Pharmaceutical Company; and Transition Therapeutics. The Canadian Institutes of Health Research is providing funds to support ADNI clinical sites in Canada. Private sector contributions are facilitated by the Foundation for the National Institutes of Health (www.fnih.org). The grantee organization is the Northern California Institute for Research and Education, and the study is coordinated by the Alzheimer’s Therapeutic Research Institute at the University of Southern California. ADNI data are disseminated by the Laboratory for Neuro Imaging at the University of Southern California; Estudio Familiar de Influencia Genetica en Alzheimer (EFIGA): 5R37AG015473, RF1AG015473, R56AG051876; the Health & Aging Brain Study – Health Disparities (HABS-HD), supported by the National Institute on Aging of the National Institutes of Health under Award Numbers R01AG054073, R01AG058533, R01AG070862, P41EB015922, and U19AG078109; the Korean Brain Aging Study for the Early Diagnosis and Prediction of Alzheimer’s disease (KBASE), which was supported by a grant from Ministry of Science, ICT and Future Planning (Grant No: NRF-2014M3C7A1046042); Memory & Aging Project at Knight Alzheimer’s Disease Research Center (MAP at Knight ADRC): The Memory and Aging Project at the Knight-ADRC (Knight-ADRC). This work was supported by the National Institutes of Health (NIH) grants R01AG064614, R01AG044546, RF1AG053303, RF1AG058501, U01AG058922 and R01AG064877 to Carlos Cruchaga. The recruitment and clinical characterization of research participants at Washington University was supported by NIH grants P30AG066444, P01AG03991, and P01AG026276. Data collection and sharing for this project was supported by NIH grants RF1AG054080, P30AG066462, R01AG064614 and U01AG052410. We thank the contributors who collected samples used in this study, as well as patients and their families, whose help and participation made this work possible. This work was supported by access to equipment made possible by the Hope Center for Neurological Disorders, the Neurogenomics and Informatics Center (NGI: https://neurogenomics.wustl.edu/) and the Departments of Neurology and Psychiatry at Washington University School of Medicine; National Alzheimer’s Coordinating Center (NACC): The NACC database is funded by NIA/NIH Grant U24 AG072122. SCAN is a multi-institutional project that was funded as a U24 grant (AG067418) by the National Institute on Aging in May 2020. Data collected by SCAN and shared by NACC are contributed by the NIA-funded ADRCs as follows: P30 AG062429 (PI James Brewer, MD, PhD), P30 AG066468 (PI Oscar Lopez, MD), P30 AG062421 (PI Bradley Hyman, MD, PhD), P30 AG066509 (PI Thomas Grabowski, MD), P30 AG066514 (PI Mary Sano, PhD), P30 AG066530 (PI Helena Chui, MD), P30 AG066507 (PI Marilyn Albert, PhD), P30 AG066444 (PI John Morris, MD), P30 AG066518 (PI Jeffrey Kaye, MD), P30 AG066512 (PI Thomas Wisniewski, MD), P30 AG066462 (PI Scott Small, MD), P30 AG072979 (PI David Wolk, MD), P30 AG072972 (PI Charles DeCarli, MD), P30 AG072976 (PI Andrew Saykin, PsyD), P30 AG072975 (PI David Bennett, MD), P30 AG072978 (PI Neil Kowall, MD), P30 AG072977 (PI Robert Vassar, PhD), P30 AG066519 (PI Frank LaFerla, PhD), P30 AG062677 (PI Ronald Petersen, MD, PhD), P30 AG079280 (PI Eric Reiman, MD), P30 AG062422 (PI Gil Rabinovici, MD), P30 AG066511 (PI Allan Levey, MD, PhD), P30 AG072946 (PI Linda Van Eldik, PhD), P30 AG062715 (PI Sanjay Asthana, MD, FRCP), P30 AG072973 (PI Russell Swerdlow, MD), P30 AG066506 (PI Todd Golde, MD, PhD), P30 AG066508 (PI Stephen Strittmatter, MD, PhD), P30 AG066515 (PI Victor Henderson, MD, MS), P30 AG072947 (PI Suzanne Craft, PhD), P30 AG072931 (PI Henry Paulson, MD, PhD), P30 AG066546 (PI Sudha Seshadri, MD), P20 AG068024 (PI Erik Roberson, MD, PhD), P20 AG068053 (PI Justin Miller, PhD), P20 AG068077 (PI Gary Rosenberg, MD), P20 AG068082 (PI Angela Jefferson, PhD), P30 AG072958 (PI Heather Whitson, MD), P30 AG072959 (PI James Leverenz, MD); National Institute on Aging Alzheimer’s Disease Family Based Study (NIA-AD FBS): U24 AG056270; Religious Orders Study (ROS): P30AG10161,R01AG15819, R01AG42210; Memory and Aging Project (MAP -Rush): R01AG017917, R01AG42210; Minority Aging Research Study (MARS): R01AG22018, R01AG42210; the Texas Alzheimer’s Research and Care Consortium (TARCC), funded by the Darrell K Royal Texas Alzheimer’s Initiative, directed by the Texas Council on Alzheimer’s Disease and Related Disorders; Washington Heights/Inwood Columbia Aging Project (WHICAP): RF1 AG054023;and Wisconsin Registry for Alzheimer’s Prevention (WRAP): R01AG027161 and R01AG054047. Additional acknowledgments include the National Institute on Aging Genetics of Alzheimer’s Disease Data Storage Site (NIAGADS, U24AG041689) at the University of Pennsylvania, funded by NIA.

## Data availability

Gene-level burden test summary statistics were deposited at doi: 10.5281/zenodo.22702628 and are publicly available.

Data used in preparation of this manuscript can be obtained upon application at:

- UK Biobank (https://biobank.ndph.ox.ac.uk/showcase/)
- AllOfUs (https://workbench.researchallofus.org/)
- NIAGADS DSS (https://www.niagads.org/)
- AMP-PD (https://amp-pdrd.org/data)
- Holstege et al., summary statistics (Zenodo: https://doi.org/10.5281/zenodo.6818051)

## References

1. Bellenguez, C. et al. New insights into the genetic etiology of Alzheimer’s disease and related dementias. Nat. Genet. 54, 412–436 (2022).

2. Nalls, M. A. et al. Identification of novel risk loci, causal insights, and heritable risk for Parkinson’s disease: a meta-analysis of genome-wide association studies. Lancet Neurol. 18, 1091–1102 (2019).

3. Holstege, H. et al. Exome sequencing identifies rare damaging variants in ATP8B4 and ABCA1 as risk factors for Alzheimer’s disease. Nat. Genet. 54, 1786–1794 (2022).

4. Bis, J. C. et al. Whole exome sequencing study identifies novel rare and common Alzheimer’s-Associated variants involved in immune response and transcriptional regulation. Mol. Psychiatry 25, 1859–1875 (2020).

5. Makarious, M. B. et al. Large-scale rare variant burden testing in Parkinson’s disease. Brain 146, 4622–4632 (2023).

6. Fan, Y., Hu, Z., Yan, Q., Wan, J. & Liu, J. Whole-exome sequencing and burden analysis identify six novel candidate risk genes and expand the genetic landscape of Parkinson’s disease. Npj Park. Dis. 11, 347 (2025).

7. Liu, J. Z., Erlich, Y. & Pickrell, J. K. Case-control association mapping by proxy using family history of disease. Nat. Genet. 49, 325–331 (2017).

8. Loesch, D. P. et al. Characterizing the Genetic Architecture of Parkinson’s Disease in Latinos. Ann. Neurol. 90, 353–365 (2021).

9. Lake, J. et al. Multi-ancestry meta-analysis and fine-mapping in Alzheimer’s disease. Mol. Psychiatry 28, 3121–3132 (2023).

10. Le Guen, Y. et al. Association of African Ancestry–Specific APOE Missense Variant R145C With Risk of Alzheimer Disease. JAMA 329, 551–560 (2023).

11. Le Guen, Y. et al. Association of Rare APOE Missense Variants V236E and R251G With Risk of Alzheimer Disease. JAMA Neurol. 79, 652–663 (2022).

12. Kim, J. J. et al. Multi-ancestry genome-wide association meta-analysis of Parkinson’s disease. Nat. Genet. 56, 27–36 (2024).

13. Wightman, D. P., Savage, J. E., de Leeuw, C. A., Jansen, I. E. & Posthuma, D. Rare variant aggregation in 148,508 exomes identifies genes associated with proxy dementia. Sci. Rep. 13, 2179 (2023).

14. Hop, P. J. et al. Systematic rare variant analyses identify RAB32 as a susceptibility gene for familial Parkinson’s disease. Nat. Genet. 56, 1371–1376 (2024).

15. Lee, S., Abecasis, G. R., Boehnke, M. & Lin, X. Rare-variant association analysis: Study designs and statistical tests. Am. J. Hum. Genet. 95, 5–23 (2014).

16. Carss, K. et al. Whole-genome sequencing of 490,640 UK Biobank participants. Nature 645, 692–701 (2025).

17. Bick, A. G. et al. Genomic data in the All of Us Research Program. Nature 627, 340–346 (2024).

18. Beecham, G. W. et al. The Alzheimer’s Disease Sequencing Project: Study design and sample selection. Neurol. Genet. 3, e194 (2017).

19. Iwaki, H. et al. Accelerating Medicines Partnership: Parkinson’s Disease. Genetic Resource. Mov. Disord. Off. J. Mov. Disord. Soc. 36, 1795–1804 (2021).

20. Karczewski, K. J. et al. The mutational constraint spectrum quantified from variation in 141,456 humans. Nature 581, 434–443 (2020).

21. Ioannidis, N. M. et al. REVEL: An Ensemble Method for Predicting the Pathogenicity of Rare Missense Variants. Am. J. Hum. Genet. 99, 877–885 (2016).

22. Mbatchou, J. et al. Computationally efficient whole-genome regression for quantitative and binary traits. Nat. Genet. 53, 1097–1103 (2021).

23. Beber, B. C. & Chaves, M. L. F. Evaluation of patients with behavioral and cognitive complaints: misdiagnosis in frontotemporal dementia and Alzheimer’s disease. Dement. Neuropsychol. 7, 60–65 (2013).

24. Leonard, H. L. Novel Parkinson’s Disease Genetic Risk Factors Within and Across European Populations. medRxiv 2025.03.14.24319455 (2025) doi:10.1101/2025.03.14.24319455.

25. Shimohama, S., Tanino, H., Sumida, Y., Tsuda, J. & Fujimoto, S. Alteration of myo-inositol monophosphatase in Alzheimer’s disease brains. Neurosci. Lett. 245, 159–162 (1998).

26. Miller, B. L. et al. Alzheimer disease: depiction of increased cerebral myo-inositol with proton MR spectroscopy. Radiology 187, 433–437 (1993).

27. Ohnishi, T. et al. Spatial expression patterns and biochemical properties distinguish a second myo-inositol monophosphatase IMPA2 from IMPA1. J. Biol. Chem. 282, 637–646 (2007).

28. Aron, L. et al. Lithium deficiency and the onset of Alzheimer’s disease. Nature 645, 712–721 (2025).

29. Kim, M. et al. Potential late-onset Alzheimer’s disease-associated mutations in the ADAM10 gene attenuate α-secretase activity. Hum. Mol. Genet. 18, 3987–3996 (2009).

30. Synofzik, M. et al. SYNE1 ataxia is a common recessive ataxia with major non-cerebellar features: a large multi-centre study. Brain 139, 1378–1393 (2016).

31. Naruse, H. et al. Juvenile amyotrophic lateral sclerosis with complex phenotypes associated with novel SYNE1 mutations. Amyotroph. Lateral Scler. Front. Degener. 22, 576– 578 (2021).

32. Foster, E. M., Dangla-Valls, A., Lovestone, S., Ribe, E. M. & Buckley, N. J. Clusterin in Alzheimer’s Disease: Mechanisms, Genetics, and Lessons From Other Pathologies. Front. Neurosci. 13, 164 (2019).

33. Shen, R. et al. Upregulation of RIN3 induces endosomal dysfunction in Alzheimer’s disease. Transl. Neurodegener. 9, 26 (2020).

34. Maaser-Hecker, A. K. et al. RIN3 mutations impairing binding of the Alzheimer’s disease–associated protein BIN1 lead to RAB5 hyperactivation and endosomal pathology. Sci. Adv. 12, eadx2127 (2026).

35. Marr, R. A. et al. Neprilysin gene transfer reduces human amyloid pathology in transgenic mice. J. Neurosci. Off. J. Soc. Neurosci. 23, 1992–1996 (2003).

36. Wen, P., Han, C., Zhao, H., Yao, S. & Chen, H. KIAA0319 modulates Alzheimer’s disease risk through PMM2 regulation: Evidence from integrated pQTL-mediation and transcriptomic analyses. J. Alzheimers Dis. Rep. 9, 25424823251384245 (2025).

37. Regan, P., McClean, P. L., Smyth, T. & Doherty, M. Early Stage Glycosylation Biomarkers in Alzheimer’s Disease. Medicines 6, 92 (2019).

38. Wevers, A. et al. Expression of nicotinic acetylcholine receptors in Alzheimer’s disease: postmortem investigations and experimental approaches. Behav. Brain Res. 113, 207– 215 (2000).

39. Sabri, O. et al. Cognitive correlates of α4β2 nicotinic acetylcholine receptors in mild Alzheimer’s dementia. Brain 141, 1840–1854 (2018).

40. Cable, J. M., Wongwiwat, W., Grabowski, J. C., White, R. E. & Luftig, M. A. Sp140L functions as a herpesvirus restriction factor suppressing viral transcription and activating interferon-stimulated genes. Proc. Natl. Acad. Sci. 122, e2426339122 (2025).

41. Spiteri, A. G., Wishart, C. L., Pamphlett, R., Locatelli, G. & King, N. J. C. Microglia and monocytes in inflammatory CNS disease: integrating phenotype and function. Acta Neuropathol. (Berl.) 143, 179–224 (2022).

42. Liu, J.-Q., Chu, S.-F., Zhou, X., Zhang, D.-Y. & Chen, N.-H. Role of chemokines in Parkinson’s disease. Brain Res. Bull. 152, 11–18 (2019).

43. Freischmidt, A. et al. Haploinsufficiency of TBK1 causes familial ALS and fronto-temporal dementia. Nat. Neurosci. 18, 631–636 (2015).

44. Fukuda, M. Multiple Roles of VARP in Endosomal Trafficking: Rabs, Retromer Components and R-SNARE VAMP7 Meet on VARP. Traffic 17, 709–719 (2016).

45. Hesketh, G. G. et al. VARP Is Recruited on to Endosomes by Direct Interaction with Retromer, Where Together They Function in Export to the Cell Surface. Dev. Cell 29, 591–606 (2014).

46. McGough, I. J. et al. Retromer Binding to FAM21 and the WASH Complex Is Perturbed by the Parkinson Disease-Linked VPS35(D620N) Mutation. Curr. Biol. 24, 1670– 1676 (2014).

47. Schorova, L. et al. USP19 deubiquitinase inactivation regulates α-synuclein ubiquitination and inhibits accumulation of Lewy body-like aggregates in mice. Npj Park. Dis. 9, 157 (2023).

48. Characterization of the Deubiquitinating Activity of USP19 and Its Role in Endoplasmic Reticulum-associated Degradation. J. Biol. Chem. 289, 3510–3517 (2014).

49. Zhang, T. et al. BNIP3 Protein Suppresses PINK1 Kinase Proteolytic Cleavage to Promote Mitophagy. J. Biol. Chem. 291, 21616–21629 (2016).

50. Hicks, A. R. et al. The non-specific lethal complex regulates genes and pathways genetically linked to Parkinson’s disease. Brain J. Neurol. 146, 4974–4987 (2023).

51. Chatterjee, A. et al. MOF Acetyl Transferase Regulates Transcription and Respiration in Mitochondria. Cell 167, 722–738.e23 (2016).

52. Boassa, D., Nguyen, P., Hu, J., Ellisman, M. H. & Sosinsky, G. E. Pannexin2 oligomers localize in the membranes of endosomal vesicles in mammalian cells while Pannexin1 channels traffic to the plasma membrane. Front. Cell. Neurosci. 8, (2015).

53. Jansen, I. E. et al. Genome-wide meta-analysis identifies new loci and functional pathways influencing Alzheimer’s disease risk. Nat. Genet. 51, 404–413 (2019).

54. Kuzma, A. et al. NIAGADS: The NIA Genetics of Alzheimer’s Disease Data Storage Site. Alzheimers Dement. 12, 1200–1203 (2016).

55. Manichaikul, A. et al. Robust relationship inference in genome-wide association studies. Bioinformatics 26, 2867–2873 (2010).

56. Chen, C. Y. et al. Improved ancestry inference using weights from external reference panels. Bioinformatics 29, 1399–1406 (2013).

57. Conomos, M. P., Miller, M. B. & Thornton, T. A. Robust Inference of Population Structure for Ancestry Prediction and Correction of Stratification in the Presence of Relatedness. Genet. Epidemiol. 39, 276–293 (2015).

58. Le Guen, Y. et al. A novel age-informed approach for genetic association analysis in Alzheimer’s disease. Alzheimers Res. Ther. 13, 72 (2021).

59. McLaren, W. et al. The Ensembl Variant Effect Predictor. Genome Biol. 17, 122 (2016).

60. Chen, S. et al. A genomic mutational constraint map using variation in 76,156 human genomes. Nature 625, 92–100 (2024).

61. Li, J. & Ji, L. Adjusting multiple testing in multilocus analyses using the eigenvalues of a correlation matrix. Heredity 95, 221–227 (2005).

