## Supplementary_Figures_and_Tables for "Population-scale burden analysis of rare damaging coding variants identifies novel risk genes for Alzheimer’s disease and related dementias and Parkinson’s disease and related disorders"

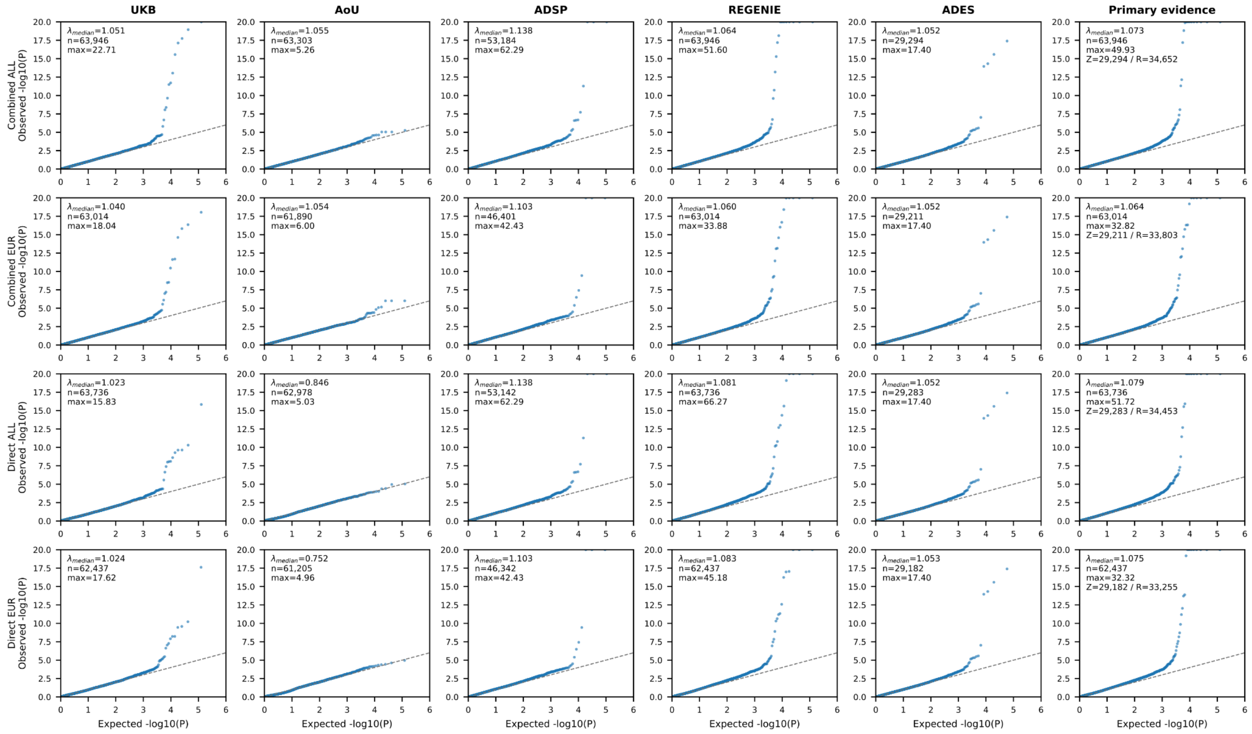

**Supplementary Fig. 1 | Quantile–quantile plots for ADRD gene-burden analyses across cohorts and meta-analyses.** Quantile–quantile plots of observed versus expected $-\log_{10}(P)$ values for gene-based burden tests in ADRD, shown separately for combined-proxy and directly-diagnosed analyses in the all-ancestry and European-ancestry strata. Columns correspond to UK Biobank (UKB), All of Us (AoU), ADSP, and the corresponding REGENIE meta-analysis, ADES stage 1 summary statistics (Holstege et al.), and the REGENIE+ADES meta-analysis; rows correspond to AD-combined-proxy (all-ancestry and European ancestry) and AD-directly-diagnosed (all-ancestry and European ancestry) analyses. Each point represents one gene/category burden test. The dashed line indicates the null expectation. Insets report the genomic inflation factor ($\lambda_{GC}$), the number of tested gene/category pairs (n), and the maximum observed $-\log_{10}(P)$ value in each analysis. Overall, cohort-level and meta-analysis results showed good calibration, with enrichment in the tail reflecting true association signals.

**Supplementary Fig. 2 | Cohort-specific effects for AD-combined-proxy gene-based burden study-wide significant and near threshold associations (multi-page pdf).** Forest plots showing cohort-specific odds ratios (ORs) and 95% confidence intervals. For each gene, results are shown for all set (LoF; LoF+REVEL≥0.75; LoF+REVEL≥0.50; LoF+REVEL≥0.25) in the all-ancestry and/or European-ancestry meta-analyses, as applicable. Cohorts include UKB, AoU, ADSP, and ADES summary statistics were used; the final row shows the inverse-variance–weighted fixed-effects meta-analysis of REGENIE summary statistics, with overall p corresponding to the meta-analysis with ADES. For combined-proxy-phenotype cohorts (UKB, AoU), effect estimates were rescaled prior to meta-analysis (**Supplementary Note 1**). Between-cohort heterogeneity is summarized by $I^2$ and $P_{het}$.

**Supplementary Fig. 3 | Sensitivity forest plots comparing all-cause dementia (mostly ADRD) and AD biobank definitions in the all-ancestry analysis (multi-page pdf).** Forest plots showing odds ratios (ORs) and 95% confidence intervals for selected ADRD gene/category pairs in the all-ancestry stratum. Gene/category pairs were selected if they reached $P<1\times10^{-5}$ in the AD-combined-proxy meta-analysis or in the AD-directly-diagnosed meta-analysis. Hereafter, ADRD corresponds to analyses including all-cause dementia direct diagnosis in UKB/AoU, as opposed to solely AD. Broader ADRD-only estimates were displayed for the same selected pairs but did not define the inclusion set. For each gene/category pair, rows show fixed-effects meta-analysis estimates for ADRD combined-proxy, AD combined-proxy, ADRD directly-diagnosed and AD directly-diagnosed definitions, followed by source-specific estimates from ADES, ADSP, AoU and UKB where available. Proxy estimates correspond to combined diagnosed/self-reported and first-degree family-history phenotypes; directly-diagnosed estimates correspond to diagnosed, self-reported or clinically ascertained cases only. For proxy-phenotype cohorts, effect estimates were rescaled by the cohort- and phenotype-specific exact inversion before meta-analysis (**Supplementary Note 1**).

**Supplementary Fig. 4 | Sensitivity forest plots comparing all-cause dementia (mostly ADRD) and AD biobank definitions in the European-ancestry analysis (multi-page pdf).** Forest plots showing ORs and 95% confidence intervals for selected ADRD gene/category pairs in the European-ancestry stratum. Gene/category pairs were selected if they reached $P<1\times10^{-5}$ in the AD-combined-proxy meta-analysis or in the AD-directly-diagnosed meta-analysis; hereafter ADRD corresponds to analyses including all-cause dementia direct diagnosis in UKB/AoU, as opposed to solely AD. Broader ADRD-only estimates were displayed for the same selected pairs but did not define the inclusion set. Rows show fixed-effects meta-analysis estimates for ADRD combined-proxy, AD combined-proxy, ADRD directly-diagnosed and AD directly-diagnosed definitions, followed by source-specific estimates from ADES, ADSP, AoU and UKB where available. These plots assess whether AD associations were preserved under broader ADRD definitions and diagnosed/self/clinical-only case ascertainment. For proxy-phenotype cohorts, effect estimates were rescaled by the cohort- and phenotype-specific exact inversion before meta-analysis (**Supplementary Note 1**).

**Supplementary Fig. 5 | Single-variant associations in selected ADRD genes under the combined-proxy definition.** Results are shown separately for the all-ancestry (ALL) and European-ancestry (EUR) analyses, with ALL followed by EUR for each gene. Variants are positioned by protein coordinate, and the vertical axis shows $-\log_{10}(P)$. Shapes indicate the most restrictive functional category; red and blue indicate positive and negative effects, respectively, at $P < 0.001$, and gray indicates $P >= 0.001$. Marker size represents effect magnitude, exp(abs(beta)), on the displayed scale. Selected variants with $P < 0.05$ are annotated; labels may be omitted to avoid overlap. Variant annotations, odds ratios, 95% confidence intervals and P values are provided in **Supplementary Table 3**.

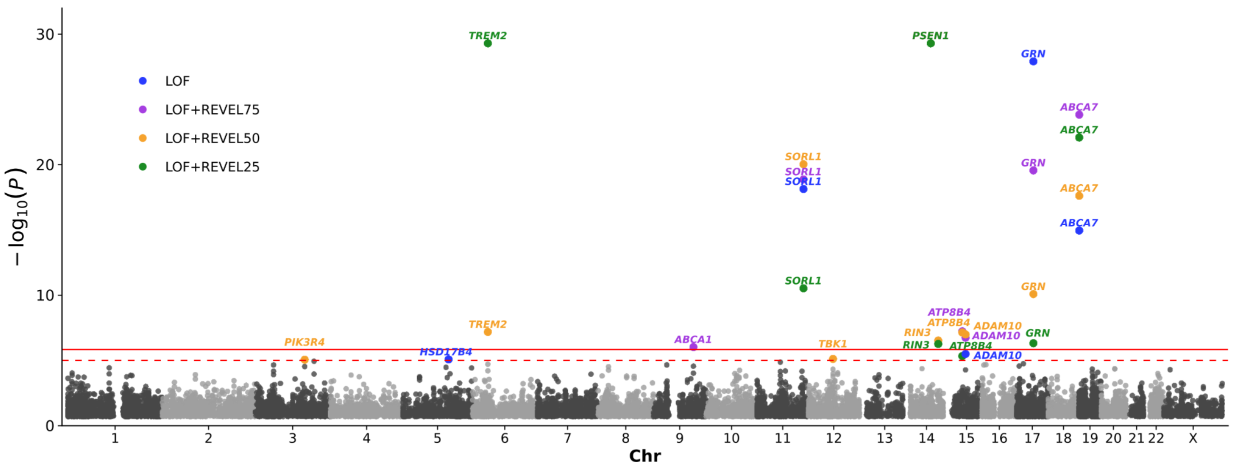

**Supplementary Fig. 6 | Gene-based rare-variant burden associations with directly-diagnosed AD meta-analysis.** For each gene, the most significant association across the all-ancestry and European ancestry meta-analyses is shown (p-values capped at p = $5\times10^{-30}$). The solid horizontal line denotes the study-wide significance threshold for AD ($P<1.20\times10^{-6}$), and the dashed line denotes the suggestive threshold ($P<1\times10^{-5}$).

**Supplementary Fig. 7 | Cohort-specific effects for AD-direct gene-based burden study-wide significant and near threshold associations (multi-page pdf).** Forest plots showing cohort-specific odds ratios (ORs) and 95% confidence intervals. For each gene, results are shown for all set (LoF; LoF+REVEL≥0.75; LoF+REVEL≥0.50; LoF+REVEL≥0.25) in the all-ancestry and/or European-ancestry meta-analyses, as applicable. Cohorts include UKB, AoU, ADSP, and ADES summary statistics were used; the final row shows the inverse-variance–weighted fixed-effects meta-analysis of REGENIE summary statistics, with overall p corresponding to the meta-analysis with ADES. Between-cohort heterogeneity is summarized by $I^2$ and P_het.

**Supplementary Fig. 8 | All-cause dementia and AD diagnostic-sensitivity analyses for FTD/ALS-spectrum genes (multi-page pdf).** Forest plots showing odds ratios (ORs) and 95% confidence intervals for rare damaging variant burden in *GRN, TBK1* and *VCP* across four deleteriousness masks: LoF, LoF+REVEL≥0.75, LoF+REVEL≥0.50 and LoF+REVEL≥0.25. Results are shown separately for all-ancestry and European-ancestry analyses. For each gene/mask, rows show fixed-effects meta-analysis estimates for broad ADRD combined-proxy, narrower AD combined-proxy, broad ADRD directly-diagnosed and narrower AD directly-diagnosed definitions, followed by source-specific estimates from ADES, ADSP, AoU and UKB where available. Proxy estimates correspond to combined diagnosed/self-reported and first-degree family-history phenotypes; directly-diagnosed estimates correspond to diagnosed, self-reported or clinically ascertained cases only. For combined-proxy-phenotype cohorts, effect estimates were rescaled using cohort- and phenotype-specific mixture scaling. The persistence of *GRN, TBK1* and *VCP* burden under narrower AD directly-diagnosed definitions illustrates that clinically diagnosed AD labels, like broad ADRD labels, can include etiologically heterogeneous dementia syndromes.

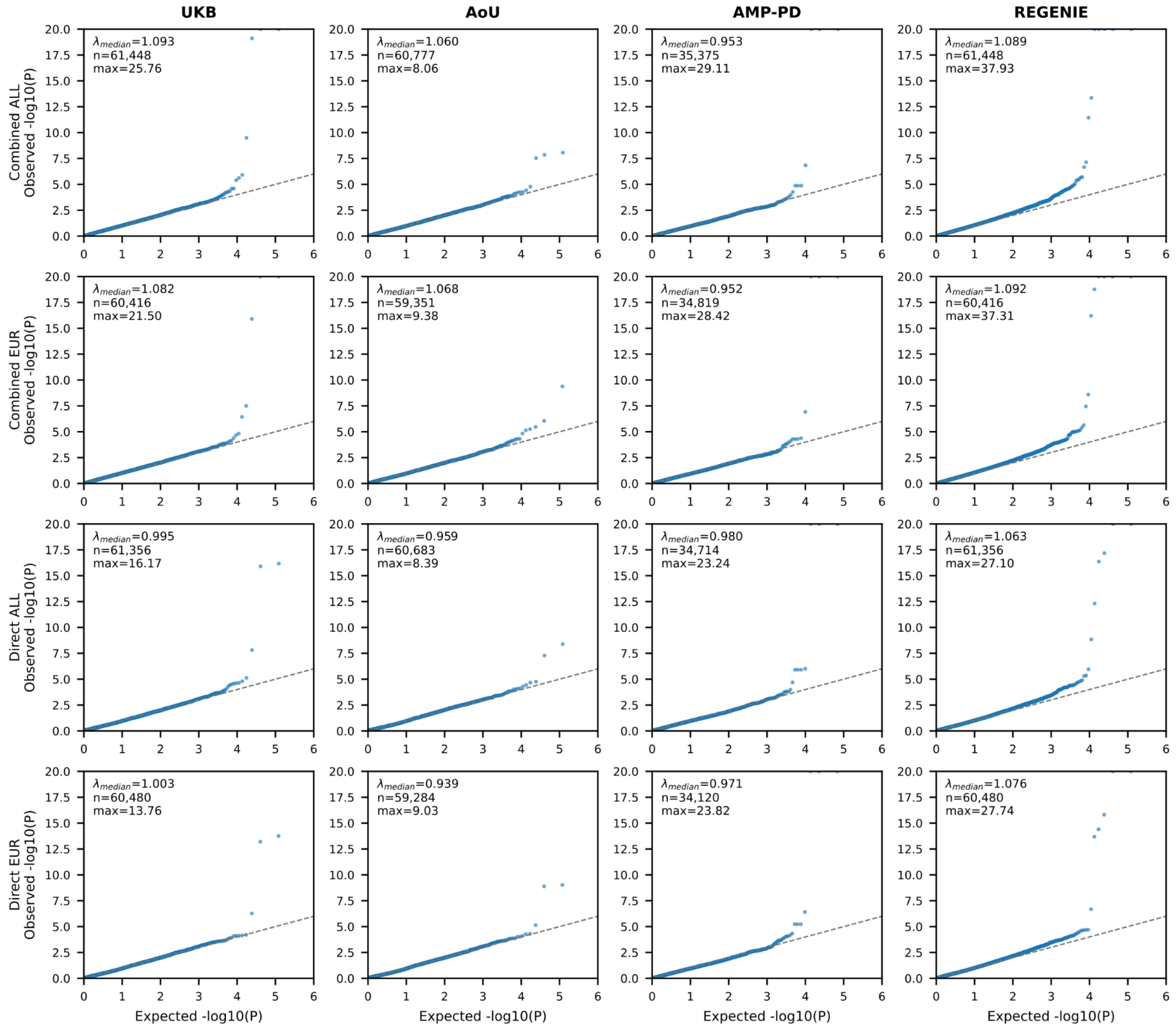

**Supplementary Fig. 9 | Quantile–quantile plots for PDRD gene-burden analyses across cohorts and meta-analyses.** Quantile–quantile plots of observed versus expected $-\log_{10}(P)$ values for gene-based burden tests in PDRD, shown separately for combined-proxy and directly-diagnosed analyses in the all-ancestry and European-ancestry strata. Columns correspond to UK Biobank (UKB), All of Us (AoU), AMP-PD, and the corresponding fixed-effects meta-analysis; rows correspond to PD-combined-proxy (all-ancestry and European ancestry) and PD-directly-diagnosed (all-ancestry and European ancestry) analyses. Each point represents one gene/category burden test. The dashed line indicates the null expectation. Insets report the genomic inflation factor ($\lambda_{GC}$), the number of tested gene/category pairs (n), and the maximum observed $-\log_{10}(P)$ value in each analysis. Most cohort-level analyses were well calibrated, whereas the directly-diagnosed-only PDRD meta-analysis showed modest inflation relative to the corresponding cohort-level analyses.

**Supplementary Fig. 10 | Cohort-specific effects for PD-combined-proxy gene-based burden study-wide significant and near threshold associations (multi-page pdf).** Forest plots showing cohort-specific ORs and 95% confidence intervals. For each gene, results are shown for the all set in the all-ancestry and/or European-ancestry meta-analyses, as applicable. Cohorts include UKB, AoU, and AMP-PDRD; the final row shows the inverse-variance–weighted fixed-effects meta-analysis. For combined-proxy-phenotype cohorts (UKB, AoU, AMP-PDRD), effect estimates were rescaled prior to meta-analysis (**Supplementary Note 1**). Between-cohort heterogeneity is summarized by $I^2$ and P_het.

**Supplementary Fig. 11 | Sensitivity forest plots comparing parkinsonism (mostly PDRD) and PD biobank definitions in the all-ancestry analysis (multi-page pdf).** Forest plots showing ORs and 95% confidence intervals for selected PDRD gene/category pairs in the all-ancestry stratum. Gene/category pairs were selected if they reached $P<1\times10^{-5}$ in the narrow PD-combined-proxy meta-analysis or in the PD-directly-diagnosed meta-analysis; broad PDRD-only estimates were displayed for the same selected pairs but did not define the inclusion set. For each gene/category pair, rows show fixed-effects meta-analysis estimates for PDRD combined-proxy, PD combined-proxy, PDRD directly-diagnosed and PD directly-diagnosed definitions, followed by source-specific estimates from AMP-PDRD, AoU and UKB where available. Proxy estimates correspond to combined diagnosed/self-reported/clinical and first-degree family-history phenotypes; directly-diagnosed estimates correspond to diagnosed, self-reported or clinically ascertained cases only. For combined-proxy-phenotype cohorts, effect estimates were rescaled by the cohort- and phenotype-specific mixture scale factor before meta-analysis.

**Supplementary Fig. 12 | Sensitivity forest plots comparing parkinsonism (mostly PDRD) and PD definitions in the European-ancestry analysis (multi-page pdf).** Forest plots showing ORs and 95% confidence intervals for selected PDRD gene/category pairs in the European-ancestry stratum. Gene/category pairs were selected if they reached $P<1\times10^{-5}$ in the PD-combined-proxy meta-analysis or in the PD-directly-diagnosed meta-analysis; broader PDRD-only estimates were displayed for the same selected pairs but did not define the inclusion set. Rows show fixed-effects meta-analysis estimates for PDRD combined-proxy, PD combined-proxy, PDRD directly-diagnosed and PD directly-diagnosed definitions, followed by source-specific estimates from AMP-PDRD, AoU and UKB where available. For combined-proxy-phenotype cohorts, effect estimates were rescaled by the cohort- and phenotype-specific mixture scale factor before meta-analysis.

**Supplementary Fig. 13 | Single-variant associations in selected PDRD genes under the combined-proxy definition.** Results are shown separately for the all-ancestry (ALL) and European-ancestry (EUR) analyses, with ALL followed by EUR for each gene. Variants are positioned by protein coordinate, and the vertical axis shows $-\log_{10}(P)$. Shapes indicate the most restrictive functional category; red and blue indicate positive and negative effects, respectively, at $P < 0.001$, and gray indicates $P >= 0.001$. Marker size represents effect magnitude, exp(abs(beta)), on the displayed scale. Selected variants with $P < 0.05$ are annotated; labels may be omitted to avoid overlap. Variant annotations, odds ratios, 95% confidence intervals and P values are provided in Supplementary Table 5.

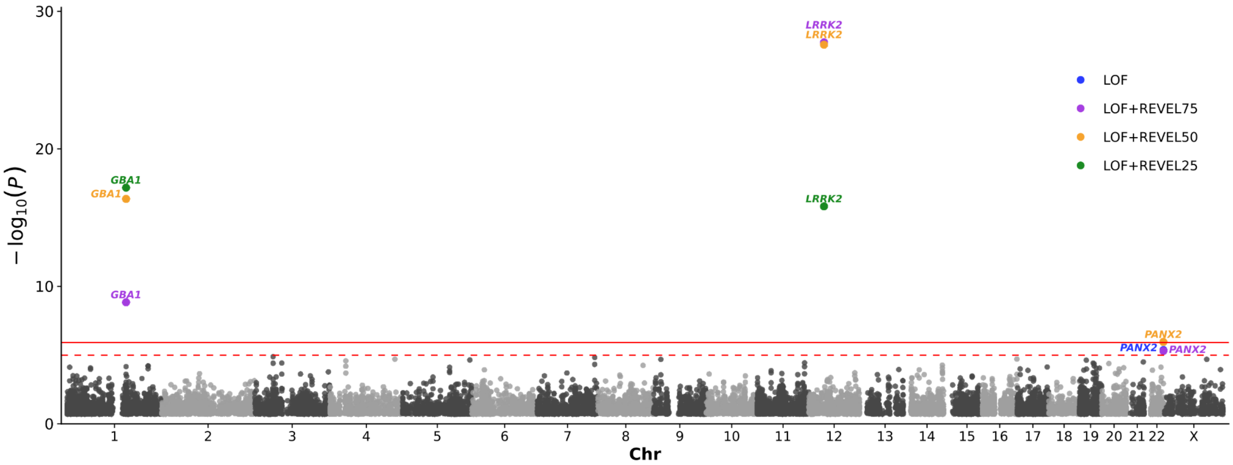

**Supplementary Fig. 14 | Gene-based rare-variant burden associations with PD-directly-diagnosed meta-analysis.** For each gene, the most significant association across the all-ancestry and European ancestry meta-analyses is shown (p-values capped at $p = 5 \times 10^{-30}$). The solid horizontal line denotes the study-wide significance threshold for PD ($P<1.20 \times 10^{-6}$), and the dashed line denotes the suggestive threshold ($P<1 \times 10^{-5}$). Exact cohort-specific and meta-analysis P values and effect estimates for significant genes are provided in **Supplementary Fig. 15** (forest plots).

**Supplementary Fig. 15 | Cohort-specific effects for PD-directly-diagnosed gene-based burden study-wide significant associations (multi-page pdf).** Forest plots showing cohort-specific ORs and 95% confidence intervals. For each gene, results are shown for the all set in the all-ancestry and/or European-ancestry meta-analyses, as applicable. Cohorts include UKB, AoU, and AMP-PDRD; the final row shows the inverse-variance–weighted fixed-effects meta-analysis. Between-cohort heterogeneity is summarized by $I^2$ and $P_{het}$.

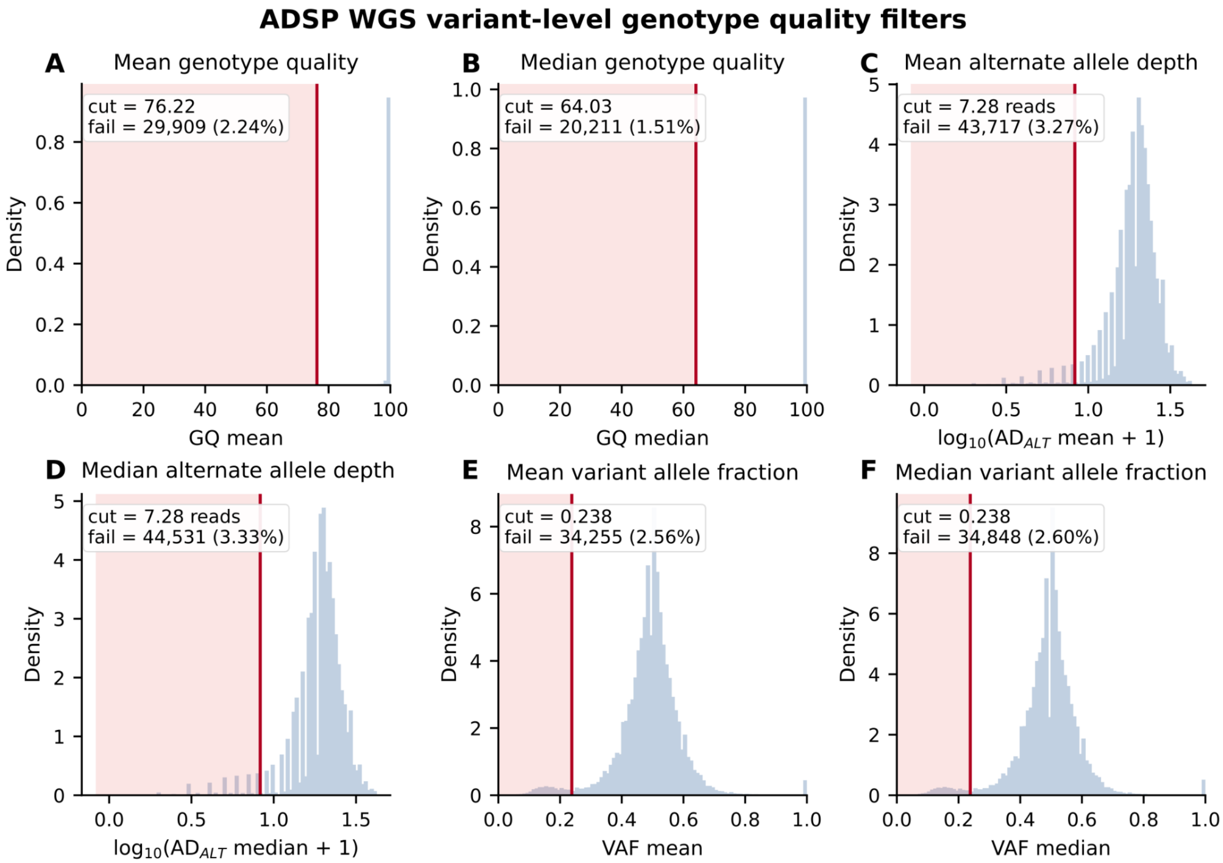

**Supplementary Fig. 16 | ADSP WGS carrier-level variant-quality filters.** Distributions of carrier-level genotype-support metrics used for additional phenotype-blind variant filtering in ADSP WGS. For each rare coding variant considered in burden testing, genotype quality (GQ), alternate-allele depth ($AD_{ALT}$) and variant allele fraction (VAF) were summarized across alternate-allele carriers using the mean and median. Red vertical lines indicate the empirically selected exclusion thresholds: mean GQ <76.22 (A), median GQ <64.03 (B), mean $AD_{ALT}$ <7.28 reads (C), median $AD_{ALT}$ <7.28 reads (D), mean VAF <0.238 (E) and median VAF <0.238 (F). Red shading denotes the excluded region for each metric. Insets report the number and percentage of variants failing each individual criterion; counts are not mutually exclusive across panels. $AD_{ALT}$ panels are plotted on the $\log_{10}(AD_{ALT}+1)$ scale for visualization.

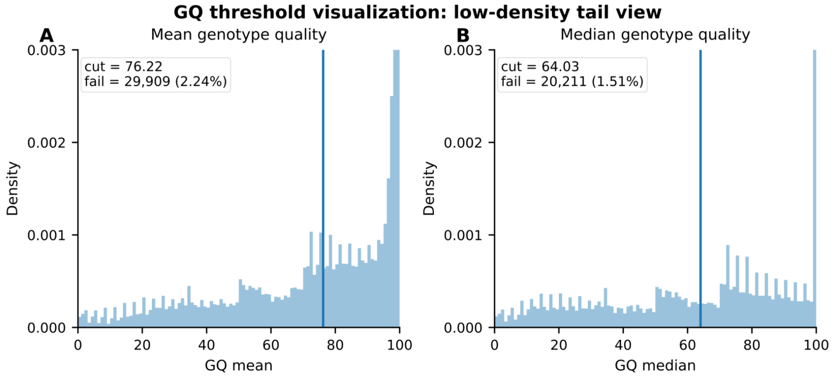

**Supplementary Fig. 17 | Low-density tail view of ADSP genotype-quality filters.** Truncated y-axis view of the mean and median genotype-quality distributions shown in Supplementary Fig. above. This visualization highlights the low-density tail used to define empirical GQ exclusion thresholds while avoiding compression by the high-density mode of well-supported variants. Red vertical lines indicate the final thresholds for mean GQ (<76.22) and median GQ (<64.03), and red shading denotes variants excluded by each metric.

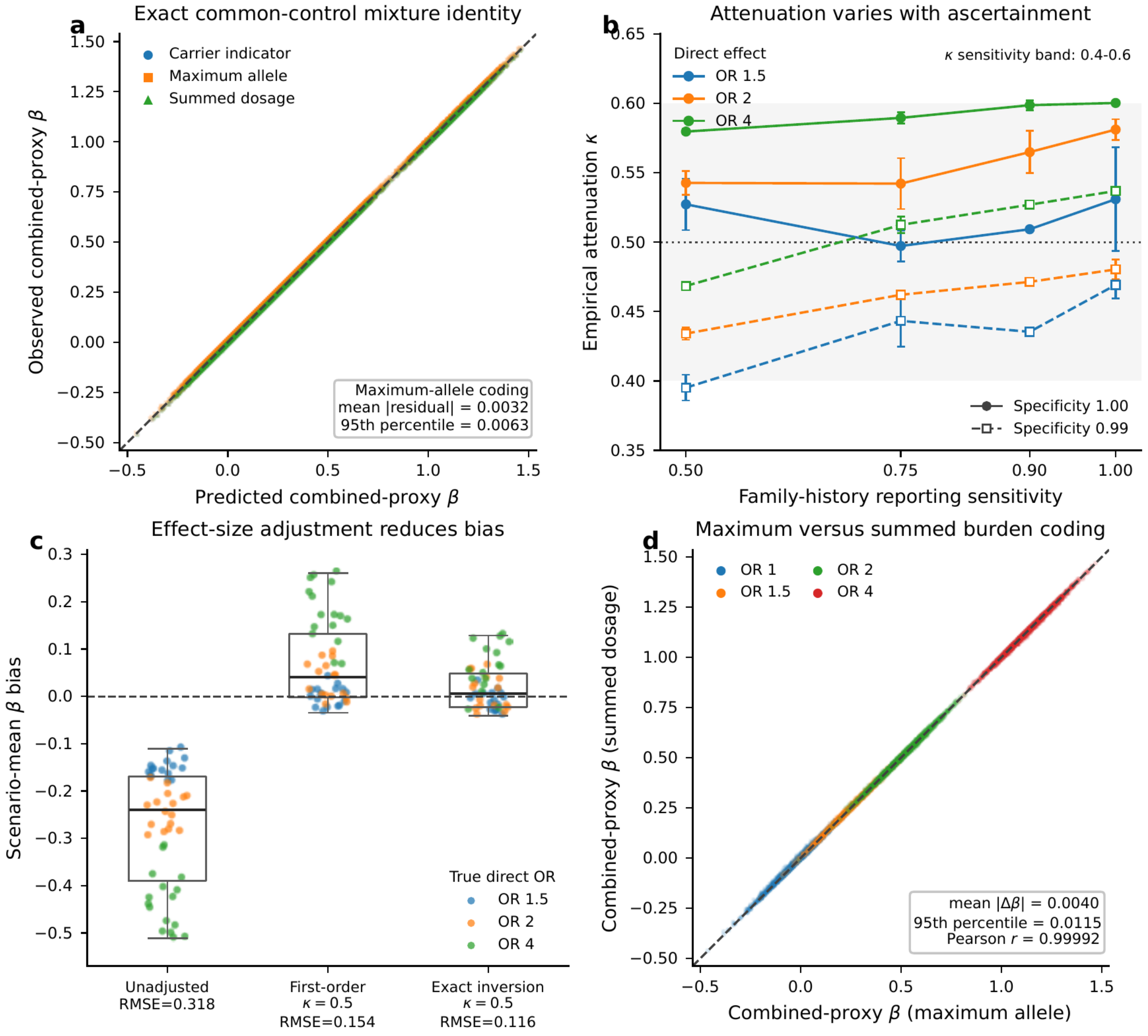

**Supplementary Fig. 18 | Simulation validation of the proxy mixture model.** The figure shows the close agreement between the predicted and fitted combined-proxy effect, the dependence of empirical attenuation on reporting sensitivity and specificity, the reduction in effect-size bias after exact inversion, and the close, but not identical, behavior of maximum-allele and summed-dosage burden coding under the simulated rare-variant architecture. Simulation is described in **Supplementary Note 1**.

**Supplementary Table 1. Summary of AD-combined-proxy gene-based burden secondary analysis in European ancestry.** For each gene meeting suggestive significance in the European ancestry stratum, the best-performing deleteriousness set (LoF; LoF+REVEL≥0.75; LoF+REVEL≥0.50; LoF+REVEL≥0.25) is reported, together with effect estimates (OR [95% CI]) and significance (Meta P). Meta P ALL corresponds to the p-value in the primary analysis in all-ancestry. Between-cohort heterogeneity is summarized by $I^2$, and direction of effect across cohorts is shown.

| Chr | Position | Gene | Category | CumFrq | OR (95% CI) | OR_ADES (95% CI) | Meta P | I2 | DIR | Meta P ALL |
|---|---|---|---|---|---|---|---|---|---|---|
| 6 | 41160810 | *TREM2* | LOF+REVEL25 | 0.004829 | 1.767 [1.587-1.967] | 2.370 [1.928-2.913] | 1.51E-33 | 71.23 | +/+/+/+ | 9.53E-39 |
| 14 | 73184368 | *PSEN1* | LOF+REVEL75 | 0.000500 | 6.487 [4.812-8.745] | 1.215 [0.610-2.417] | 2.22E-33 | 96.08 | +/+/+/+ | 1.18E-50 |
| 19 | 1052622 | *ABCA7* | LOF+REVEL25 | 0.038317 | 1.254 [1.200-1.310] | 1.376 [1.224-1.546] | 2.45E-28 | 60.98 | +/+/+/+ | 2.41E-36 |
| 11 | 121541516 | *SORL1* | LOF+REVEL50 | 0.008228 | 1.513 [1.381-1.656] | 2.473 [2.015-3.036] | 4.55E-27 | 66.14 | +/+/+/+ | 2.43E-28 |
| 17 | 44348651 | *GRN* | LOF | 0.000231 | 4.870 [3.214-7.380] | | 8.26E-14 | 73.38 | +/+/+/. | 6.47E-18 |
| 15 | 50031092 | *ATP8B4* | LOF+REVEL50 | 0.016615 | 1.204 [1.127-1.288] | 1.400 [1.197-1.639] | 2.83E-10 | 0.00 | +/+/+/+ | 4.52E-08 |
| 18 | 12006518 | *IMPA2* | LOF+REVEL75 | 0.000177 | 4.282 [2.425-7.562] | | 5.37E-07 | 0.00 | +/+/+/. | 7.67E-07 |
| 15 | 58668622 | *ADAM10* | LOF+REVEL75 | 0.000066 | 5.128 [2.288-11.496] | 19.003 [4.034-89.517] | 6.11E-07 | 0.00 | +/+/+/+ | 9.28E-07 |
| 8 | 144101259 | *SHARPIN* | LOF+REVEL25 | 0.004647 | 1.368 [1.212-1.544] | 0.912 [0.436-1.908] | 1.16E-06 | 25.77 | +/+/+/- | 4.80E-06 |
| 6 | 152377569 | *SYNE1* | LOF+REVEL50 | 0.009569 | 1.215 [1.114-1.325] | 1.281 [1.051-1.562] | 1.30E-06 | 0.00 | +/+/+/+ | 0.00051131 |
| 19 | 44907441 | *APOE* | LOF+REVEL25 | 0.005211 | 0.755 [0.673-0.846] | | 1.50E-06 | 0.00 | -/-/-/. | 0.05115377 |
| 2 | 230364973 | *SP140L* | LOF+REVEL75 | 0.000851 | 1.995 [1.497-2.658] | 1.134 [0.452-2.842] | 3.54E-06 | 0.00 | +/+/+/+ | 0.00380986 |
| 16 | 8825953 | *PMM2* | LOF+REVEL25 | 0.011039 | 1.212 [1.116-1.315] | 1.013 [0.839-1.224] | 7.23E-06 | 72.51 | +/+/+/+ | 0.00051216 |
| 17 | 34271251 | *CCL7* | LOF | 0.000013 | 82.677 [11.796-579.451] | | 8.83E-06 | 40.83 | +/+/./. | 0.00284817 |

**Supplementary Table 2. Summary of AD-combined-proxy gene-based burden near-threshold associations in the primary-analysis.** Gene–mask associations reaching the prespecified near-threshold significance threshold ($P<1\times10^{-5}$) in the primary all-ancestry meta-analysis are shown. **Table 1** provides results at $P<1.20\times10^{-6}$. Chr and Position, chromosome and GRCh38 genomic position; Category, qualifying-variant mask; CumFrq, cumulative frequency of qualifying variants; OR (95% CI), pooled odds ratio and 95% confidence interval from UKB, AoU and ADSP; OR_ADES (95% CI), corresponding estimate from ADES stage 1; Meta P, final meta-analysis P-value including ADES when available; DIR, direction of effects in UKB/AoU/ADSP/ADES; Meta P EUR, corresponding European-ancestry meta-analysis P-value. Between-cohort heterogeneity is summarized by $I^2$, and direction of effect across cohorts is shown.

| Chr | Position | Gene | Category | CumFrq | OR (95% CI) | OR_ADES (95% CI) | Meta P | I2 | DIR | Meta P EUR |
|---|---|---|---|---|---|---|---|---|---|---|
| 20 | 63354177 | *CHRNA4* | LOF+REVEL50 | 0.003442 | 0.742 [0.647-0.850] | 0.679 [0.492-0.936] | 2.38E-06 | 28.70 | -/-/-/- | 1.07E-05 |
| 6 | 41160810 | *TREM2* | LOF+REVEL75 | 0.000105 | 3.367 [1.918-5.910] | 2.122 [1.218-3.697] | 2.54E-06 | 60.67 | +/+/+/+ | 3.80E-07 |
| 9 | 104861824 | *ABCA1* | LOF+REVEL75 | 0.007647 | 1.180 [1.082-1.287] | 1.661 [1.334-2.068] | 3.21E-06 | 49.19 | +/-/+/+ | 2.38E-05 |
| 17 | 44348651 | *GRN* | LOF+REVEL50 | 0.001196 | 1.576 [1.294-1.921] | 1.332 [0.664-2.672] | 4.65E-06 | 32.74 | +/+/+/+ | 3.26E-06 |
| 8 | 144101259 | *SHARPIN* | LOF+REVEL25 | 0.005137 | 1.295 [1.164-1.441] | 0.912 [0.436-1.908] | 4.80E-06 | 70.71 | +/+/+/- | 1.16E-06 |

**Supplementary Table 3. Variant annotations and association results for the ADRD combined-proxy lollipop analyses.** The table reports gene, GRCh38 variant identifier (chr:position:REF), protein and coding changes, consequence, functional category and REVEL score, together with odds ratios (95% confidence intervals) and P values for ALL and EUR. ALT is the effect allele. Variants with $P < 0.05$ in at least one plotted ancestry are included; NA indicates unavailable information. ALL and EUR overlap. P values are reported from the final association tests and are not inferred from the displayed confidence intervals.

**Supplementary Table 4. Summary of AD-directly-diagnosed gene-based burden suggestive significant associations.** For each gene meeting study-wide significance in at least one ancestry stratum, the best-performing deleteriousness set (LoF; LoF+REVEL≥0.75; LoF+REVEL≥0.50; LoF+REVEL≥0.25) is reported, together with effect estimates (OR [95% CI]) and P-value (P) for the corresponding ancestry (Anc), p-value in the other ancestry is also reporter (P other Anc). Between-cohort heterogeneity is summarized by $I^2$, and direction of effect across cohorts is shown.

| Chr | Position | Gene | Category | CumFrq | OR (95% CI) | OR ADES (95% CI) | P | I2 | DIR | Anc | P other |
|---|---|---|---|---|---|---|---|---|---|---|---|
| 14 | 73184368 | *PSEN1* | LOF+REVEL75 | 0.00053776 | 12.774 [9.571-17.050] | 1.215 [0.610-2.417] | 1.91E-52 | 86 | +/+/+/+ | ALL | 4.78E-33 |
| 6 | 41160810 | *TREM2* | LOF+REVEL25 | 0.00577685 | 2.114 [1.825-2.449] | 2.370 [1.928-2.913] | 3.17E-37 | 37 | +/+/+/+ | ALL | 1.05E-31 |
| 19 | 1052622 | *ABCA7* | LOF+REVEL75 | 0.01223082 | 1.542 [1.405-1.693] | 1.646 [1.332-2.033] | 2.43E-24 | 57 | +/+/+/+ | ALL | 2.40E-21 |
| 11 | 121541516 | *SORL1* | LOF+REVEL50 | 0.00805159 | 1.483 [1.317-1.669] | 2.473 [2.015-3.036] | 1.62E-23 | 71 | +/+/+/+ | ALL | 2.57E-23 |
| 17 | 44348651 | *GRN* | LOF | 0.0001863 | 5.983 [3.713-9.641] | | 2.00E-13 | 80 | +/-/+/. | ALL | 6.51E-12 |
| 15 | 50031092 | *ATP8B4* | LOF+REVEL50 | 0.01643114 | 1.271 [1.142-1.415] | 1.400 [1.197-1.639] | 1.95E-09 | 54 | +/+/+/+ | EUR | 1.09E-06 |
| 9 | 104861824 | *ABCA1* | LOF+REVEL75 | 0.00760697 | 1.286 [1.125-1.470] | 1.661 [1.334-2.068] | 5.00E-08 | 0 | +/+/+/+ | ALL | 3.10E-06 |
| 15 | 58668622 | *ADAM10* | LOF+REVEL50 | 0.00018004 | 3.945 [1.879-8.280] | 8.112 [2.797-23.525] | 1.30E-07 | 0 | +/+/+/+ | ALL | 1.46E-07 |
| 12 | 64476323 | *TBK1* | LOF+REVEL50 | 0.00026536 | 4.297 [2.448-7.544] | | 3.83E-07 | 0 | +/+/+/. | ALL | 4.57E-06 |
| 14 | 92598828 | *RIN3* | LOF+REVEL50 | 0.00769151 | 1.313 [1.153-1.496] | 1.256 [1.039-1.519] | 2.18E-06 | 30 | +/+/+/+ | ALL | 0.00018154 |
| 8 | 27604873 | *CLU* | LOF+REVEL50 | 8.30E-05 | 6.300 [2.075-19.124] | 7.855 [2.374-25.992] | 2.40E-06 | 0 | +/+/+/+ | EUR | 1.59E-05 |
| 20 | 35657446 | *RBM12* | LOF | 5.02E-05 | 13.030 [4.215-40.276] | | 8.24E-06 | 0 | +/+/./. | ALL | 3.18E-05 |
| 19 | 57945625 | *ZNF256* | LOF+REVEL25 | 0.00027709 | 5.505 [2.600-11.653] | | 8.30E-06 | 74 | +/+/./. | ALL | 1.78E-05 |
| 3 | 155102551 | *MME* | LOF+REVEL75 | 0.00221498 | 1.679 [1.283-2.197] | 1.684 [1.089-2.604] | 9.25E-06 | 0 | +/+/+/+ | EUR | 0.00013436 |

**Supplementary Table 5. Variant annotations and association results for the PDRD combined-proxy lollipop analyses.** The table reports gene, GRCh38 variant identifier (chr:position:REF), protein and coding changes, consequence, functional category and REVEL score, together with odds ratios (95% confidence intervals) and P values for ALL and EUR. ALT is the effect allele. Variants with $P < 0.05$ in at least one plotted ancestry are included; NA indicates unavailable information. ALL and EUR overlap. P values are reported from the final association tests and are not inferred from the displayed confidence intervals.

**Supplementary Table 6. Summary of PD-combined-proxy gene-based burden near-threshold associations in the primary-analysis.** Gene–mask associations reaching the prespecified near-threshold significance threshold ($P<1\times10^{-5}$) in the primary all-ancestry meta-analysis are shown. **Table 2** provides results at $P<1.20\times10^{-6}$. Chr and Position, chromosome and GRCh38 genomic position; Category, qualifying-variant mask; CumFrq, cumulative frequency of qualifying variants; OR (95% CI), pooled odds ratio and 95% confidence interval from UKB, AoU and AMP-PD meta-analysis; DIR, direction of effects in UKB/AoU/AMP-PD; Meta P EUR, corresponding European-ancestry meta-analysis P-value. Between-cohort heterogeneity is summarized by $I^2$, and direction of effect across cohorts is shown.

| Chr | Position | Gene | Category | CumFrq | OR (95% CI) | Meta P | I2 | DIR | Meta P EUR |
|---|---|---|---|---|---|---|---|---|---|
| 3 | 49113401 | *USP19* | LOF+REVEL75 | 0.000095 | 7.053 [3.151-15.785] | 2.01E-06 | 16.15 | +/+/. | 9.64E-06 |
| 3 | 49113401 | *USP19* | LOF | 0.000069 | 8.498 [3.506-20.597] | 2.17E-06 | 18.32 | +/+/. | 5.17E-05 |
| 10 | 131975518 | *BNIP3* | LOF | 0.000091 | 6.844 [3.044-15.384] | 3.26E-06 | 0.00 | +/+/. | 2.09E-05 |
| 6 | 107574358 | *SOBP* | LOF+REVEL50 | 0.000072 | 8.281 [3.377-20.306] | 3.86E-06 | 0.00 | +/+/. | 0.00092786 |
| 2 | 96617186 | *KANSL3* | LOF | 0.000132 | 9.698 [3.682-25.543] | 4.26E-06 | 0.00 | +/+/. | 0.00270788 |
| 17 | 8367162 | *KRBA2* | LOF | 0.000013 | 42.167 [8.438-210.718] | 5.16E-06 | 0.00 | +/+/. | |
| 17 | 8367162 | *KRBA2* | LOF+REVEL50 | 0.000013 | 38.178 [7.723-188.722] | 7.92E-06 | 0.00 | +/+/. | |
| 2 | 227515179 | *AGFG1* | LOF+REVEL25 | 0.000307 | 3.424 [1.992-5.886] | 8.43E-06 | 34.76 | +/+/+ | 0.00078233 |

**Supplementary Table 7. Summary of PD-combined-proxy gene-based burden secondary analysis in European ancestry.** For each gene meeting suggestive significance in the European ancestry stratum, the best-performing deleteriousness set (LoF; LoF+REVEL≥0.75; LoF+REVEL≥0.50; LoF+REVEL≥0.25) is reported, together with effect estimates (OR [95% CI]) and significance (Meta P). Meta P ALL corresponds to the p-value in the primary analysis in all-ancestry. Between-cohort heterogeneity is summarized by $I^2$, and direction of effect across cohorts is shown.

| Chr | Position | Gene | Category | CumFrq | OR (95% CI) | Meta P | I2 | DIR | Meta P ALL |
|---|---|---|---|---|---|---|---|---|---|
| 12 | 40296667 | *LRRK2* | LOF+REVEL75 | 0.002547 | 3.048 [2.573-3.611] | 4.90E-38 | 76.91 | +/+/+ | 1.18E-38 |
| 1 | 155240129 | *GBA1* | LOF+REVEL25 | 0.012732 | 1.801 [1.623-1.998] | 1.14E-28 | 46.92 | +/+/+ | 6.69E-35 |
| 19 | 32636455 | *ANKRD27* | LOF+REVEL25 | 0.014976 | 1.339 [1.207-1.485] | 3.54E-08 | 0.00 | +/+/+ | 7.31E-08 |
| 7 | 99624456 | *ZSCAN25* | LOF | 0.000105 | 6.818 [3.076-15.114] | 2.28E-06 | 0.00 | +/+/. | 2.53E-05 |
| 17 | 34271251 | *CCL7* | LOF+REVEL50 | 0.000037 | 15.975 [4.926-51.809] | 3.91E-06 | 44.29 | +/-/. | 2.16E-07 |
| 8 | 125010086 | *SQLE* | LOF+REVEL50 | 0.000677 | 2.345 [1.618-3.399] | 6.69E-06 | 0.00 | +/+/+ | 1.30E-05 |
| 16 | 89848129 | *SPIRE2* | LOF+REVEL50 | 0.001689 | 1.819 [1.399-2.365] | 7.90E-06 | 0.00 | +/+/+ | 0.00011565 |
| 2 | 23845563 | *ATAD2B* | LOF | 0.000037 | 13.417 [4.275-42.102] | 8.58E-06 | 0.00 | +/+/. | 8.22E-05 |
| 3 | 49113401 | *USP19* | LOF+REVEL75 | 0.000091 | 7.352 [3.038-17.790] | 9.64E-06 | 54.05 | +/+/. | 2.01E-06 |
| 7 | 27197974 | *HOXA13* | LOF | 0.000016 | 34.556 [7.178-166.344] | 9.94E-06 | 49.36 | +/-/. | 5.48E-05 |

**Supplementary Table 8. Summary of PD-directly-diagnosed gene-based burden suggestive significant associations.** For each gene meeting study-wide significance in at least one ancestry stratum, the best-performing deleteriousness set (LoF; LoF+REVEL≥0.75; LoF+REVEL≥0.50; LoF+REVEL≥0.25) is reported, together with effect estimates (OR [95% CI]) and P-value (P) for the corresponding ancestry (Anc), p-value in the other ancestry is also reporter (P other anc). Between-cohort heterogeneity is summarized by $I^2$, and direction of effect across cohorts is shown.

| Chr | Position | Gene | Category | CumFrq | OR (95% CI) | P | I2 | DIR | Anc | P other anc |
|---|---|---|---|---|---|---|---|---|---|---|
| 12 | 40296667 | *LRRK2* | LOF+REVEL75 | 0.002335 | 3.007 [2.474-3.655] | 1.80E-28 | 77.22 | +/+/+ | EUR | 8.02E-28 |
| 1 | 155240129 | *GBA1* | LOF+REVEL25 | 0.014974 | 1.627 [1.457-1.818] | 6.63E-18 | 83.58 | +/+/+ | ALL | 3.94E-15 |
| 22 | 50174774 | *PANX2* | LOF+REVEL50 | 0.000372 | 4.354 [2.409-7.868] | 1.10E-06 | 0.00 | +/+/+ | ALL | 0.00668125 |

**Supplementary Table 9. Absolute agreement of pooled REGENIE effect estimates across phenotype definitions**

For each prespecified pairwise comparison of broad versus narrow disease definitions and combined versus directly diagnosed ascertainment, the table reports the ancestry stratum, number of complete exact gene/mask pairs, Lin's concordance correlation coefficient with bootstrap 95% confidence interval, Deming-regression slope and intercept with bootstrap 95% confidence intervals, Bland-Altman mean beta difference and 95% limits of agreement, median absolute beta difference, direction-consistency count and fraction, and Pearson correlation as a descriptive measure of linear association. Differences are oriented as the second phenotype coefficient minus the first. The comparison universe was defined from exact gene/mask pairs meeting P <= 1.20 x $10^{-6}$ in the all-ancestry AD/PD -combined-proxy or AD/PD -direct-diagnosis analysis for the corresponding disease family. The same selected pairs were evaluated in the all-ancestry and European-ancestry strata. Confidence intervals were estimated using 2,000 bootstrap resamples.

| Ancestry | Comparison, X versus Y | Lin CCC (95% CI) | Deming slope (95% CI) | Deming intercept (95% CI) | Mean Delta beta [95% limits of agreement] | Median \|Delta beta\| | Direction consistent | Pearson r |
|---|---|---|---|---|---|---|---|---|
| ALL | ADRD combined vs AD combined | 0.987 (0.977-0.994) | 1.057 (0.990-1.122) | 0.005 (-0.030-0.039) | 0.050 [-0.089, 0.188] | 0.055 | 26/26 | 0.993 |
| ALL | ADRD combined vs ADRD direct | 0.896 (0.817-0.942) | 1.265 (1.091-1.415) | -0.118 (-0.224--0.036) | 0.091 [-0.407, 0.590] | 0.047 | 26/26 | 0.928 |
| ALL | ADRD combined vs AD direct | 0.796 (0.655-0.884) | 1.358 (1.065-1.642) | -0.111 (-0.321-0.035) | 0.171 [-0.524, 0.866] | 0.109 | 26/26 | 0.857 |
| ALL | AD combined vs ADRD direct | 0.923 (0.843-0.962) | 1.190 (1.033-1.320) | -0.118 (-0.232--0.041) | 0.042 [-0.415, 0.498] | 0.102 | 26/26 | 0.938 |
| ALL | AD combined vs AD direct | 0.857 (0.741-0.923) | 1.262 (1.060-1.428) | -0.098 (-0.247-0.008) | 0.122 [-0.487, 0.730] | 0.110 | 26/26 | 0.893 |
| ALL | ADRD direct vs AD direct | 0.958 (0.925-0.974) | 1.047 (0.910-1.150) | 0.038 (-0.036-0.119) | 0.080 [-0.266, 0.426] | 0.118 | 26/26 | 0.966 |
| EUR | ADRD combined vs AD combined | 0.988 (0.977-0.995) | 1.051 (0.987-1.118) | 0.006 (-0.026-0.039) | 0.048 [-0.095, 0.190] | 0.038 | 26/26 | 0.992 |
| EUR | ADRD combined vs ADRD direct | 0.901 (0.838-0.939) | 1.241 (1.074-1.384) | -0.066 (-0.135-0.009) | 0.131 [-0.335, 0.596] | 0.062 | 26/26 | 0.941 |
| EUR | ADRD combined vs AD direct | 0.778 (0.635-0.876) | 1.320 (1.073-1.593) | -0.007 (-0.139-0.112) | 0.253 [-0.419, 0.926] | 0.191 | 26/26 | 0.869 |
| EUR | AD combined vs ADRD direct | 0.923 (0.860-0.959) | 1.178 (1.026-1.322) | -0.070 (-0.151-0.006) | 0.083 [-0.363, 0.529] | 0.120 | 26/26 | 0.942 |
| EUR | AD combined vs AD direct | 0.837 (0.723-0.910) | 1.239 (1.042-1.412) | -0.001 (-0.088-0.095) | 0.206 [-0.391, 0.803] | 0.174 | 26/26 | 0.898 |
| EUR | ADRD direct vs AD direct | 0.939 (0.896-0.962) | 1.041 (0.896-1.167) | 0.084 (0.019-0.175) | 0.123 [-0.271, 0.517] | 0.179 | 26/26 | 0.956 |
| ALL | PDRD combined vs PD combined | 0.990 (0.917-0.997) | 0.996 (0.909-1.138) | 0.066 (-0.016-0.156) | 0.063 [-0.095, 0.220] | 0.066 | 10/10 | 0.994 |
| ALL | PDRD combined vs PDRD direct | 0.914 (0.686-0.959) | 0.814 (0.723-1.397) | 0.074 (-0.218-0.238) | -0.102 [-0.598, 0.394] | 0.112 | 10/10 | 0.943 |
| ALL | PDRD combined vs PD direct | 0.870 (0.498-0.954) | 0.807 (0.641-1.833) | 0.176 (-0.349-0.427) | -0.006 [-0.660, 0.648] | 0.201 | 10/10 | 0.886 |
| ALL | PD combined vs PDRD direct | 0.902 (0.688-0.942) | 0.818 (0.721-1.253) | 0.019 (-0.219-0.162) | -0.165 [-0.642, 0.313] | 0.201 | 10/10 | 0.947 |
| ALL | PD combined vs PD direct | 0.893 (0.599-0.956) | 0.815 (0.691-1.547) | 0.117 (-0.320-0.326) | -0.069 [-0.648, 0.510] | 0.087 | 10/10 | 0.914 |
| ALL | PDRD direct vs PD direct | 0.965 (0.830-0.992) | 1.003 (0.887-1.229) | 0.093 (-0.041-0.237) | 0.096 [-0.151, 0.342] | 0.067 | 10/10 | 0.977 |
| EUR | PDRD combined vs PD combined | 0.988 (0.883-0.997) | 0.972 (0.833-1.135) | 0.080 (-0.027-0.195) | 0.053 [-0.152, 0.258] | 0.079 | 10/10 | 0.991 |
| EUR | PDRD combined vs PDRD direct | 0.874 (0.525-0.924) | 0.690 (0.539-0.922) | 0.164 (-0.005-0.392) | -0.133 [-0.728, 0.462] | 0.102 | 10/10 | 0.948 |
| EUR | PDRD combined vs PD direct | 0.834 (0.254-0.925) | 0.693 (0.324-1.529) | 0.260 (-0.213-0.662) | -0.035 [-0.777, 0.707] | 0.232 | 10/10 | 0.879 |
| EUR | PD combined vs PDRD direct | 0.873 (0.567-0.912) | 0.713 (0.587-0.902) | 0.105 (-0.028-0.316) | -0.186 [-0.721, 0.349] | 0.222 | 10/10 | 0.957 |
| EUR | PD combined vs PD direct | 0.876 (0.447-0.936) | 0.726 (0.515-1.267) | 0.190 (-0.145-0.479) | -0.088 [-0.700, 0.524] | 0.101 | 10/10 | 0.923 |
| EUR | PDRD direct vs PD direct | 0.952 (0.713-0.988) | 1.029 (0.838-1.358) | 0.074 (-0.098-0.257) | 0.098 [-0.171, 0.367] | 0.106 | 10/10 | 0.968 |

**Supplementary Table 10.** Participant characteristics for AD- and PD- combined-proxy analyses in the UK Biobank whole-genome sequencing.

| | | | N | Age [mean (std)] | Female [N (%)] | *APOE* [N (%)] | | | | | |
|---|---|---|---|---|---|---|---|---|---|---|---|
| | | | | | | ε2/ε2 | ε2/ε3 | ε2/ε4 | ε3/ε3 | ε3/ε4 | ε4/ε4 |
| All-ancestries | AD | Controls | 400,648 | 64.9 (10.4) | 184,328 (46.0) | 2,755 (0.7) | 51,065 (12.7) | 9,491 (2.4) | 242,298 (60.5) | 87,297 (21.8) | 7,742 (1.9) |
| | | Combined-proxy Cases | 79,275 | 67.7 (9.0) | 34,805 (43.9) | 334 (0.4) | 7,511 (9.5) | 2,431 (3.1) | 40,353 (50.9) | 25,156 (31.7) | 3,490 (4.4) |
| | PD | Controls | 459,924 | 65.3 (10.2) | 210,403 (45.7) | 2,945 (0.6) | 55,993 (12.2) | 11,427 (2.5) | 270,504 (58.8) | 108,121 (23.5) | 10,934 (2.4) |
| | | Combined-proxy Cases | 24,541 | 68.7 (9.3) | 11,304 (46.1) | 164 (0.7) | 2,989 (12.2) | 639 (2.6) | 14,286 (58.2) | 5,869 (23.9) | 594 (2.4) |
| European ancestry | AD | Controls | 333,752 | 65.4 (10.2) | 154,067 (46.2) | 2,364 (0.7) | 43,260 (13.0) | 8,030 (2.4) | 200,091 (60.0) | 73,528 (22.0) | 6,479 (1.9) |
| | | Combined-proxy Cases | 68,885 | 67.9 (8.9) | 30,435 (44.2) | 294 (0.4) | 6,528 (9.5) | 2,153 (3.1) | 34,735 (50.4) | 22,109 (32.1) | 3,066 (4.5) |
| | PD | Controls | 385,364 | 65.8 (10.1) | 176,876 (45.9) | 2,526 (0.7) | 47,571 (12.3) | 9,739 (2.5) | 224,405 (58.2) | 91,847 (23.8) | 9,276 (2.4) |
| | | Combined-proxy Cases | 21,130 | 69.0 (9.1) | 9,815 (46.5) | 151 (0.7) | 2,571 (12.2) | 571 (2.7) | 12,195 (57.7) | 5,119 (24.2) | 523 (2.5) |

**Supplementary Table 11. Directly diagnosed, family-history proxy and control counts (All-ancestry analyses).**

| Cohort / analysis | N directly diagnosed | N proxy | N controls |
|---|---|---|---|
| ADES | 12,652 | 0 | 8,693 |
| ADSP | 13,268 | 0 | 28,215 |
| UKB AD direct | 4,993 | 0 | 400,648 |
| UKB ADRD direct | 11,099 | 0 | 400,648 |
| UKB AD combined | 4,993 | 74,282 | 400,648 |
| UKB ADRD combined | 11,099 | 73,162 | 400,648 |
| AoU AD direct | 1,404 | 0 | 190,229 |
| AoU ADRD direct | 4,277 | 0 | 190,229 |
| AoU AD combined | 1,404 | 27,693 | 190,229 |
| AoU ADRD combined | 4,277 | 27,693 | 190,229 |
| AMP-PD PD direct | 3,317 | 0 | 3,533 |
| AMP-PD PD combined | 3,317 | 533 | 3,533 |
| AMP-PD PDRD direct | 5,945 | 0 | 3,533 |
| AMP-PD PDRD combined | 5,945 | 533 | 3,533 |
| UKB PD direct | 4,546 | 0 | 459,924 |
| UKB PDRD direct | 5,019 | 0 | 459,924 |
| UKB PD combined | 4,546 | 19,995 | 459,924 |
| UKB PDRD combined | 5,019 | 19,966 | 459,924 |
| AoU PD direct | 2,466 | 0 | 212,990 |
| AoU PDRD direct | 2,744 | 0 | 212,990 |
| AoU PD combined | 2,466 | 6,540 | 212,990 |
| AoU PDRD combined | 2,744 | 6,540 | 212,990 |

**Supplementary Table 12. Directly diagnosed, family-history proxy and control counts (European-ancestry analyses).**

| Cohort / analysis | N directly diagnosed | N proxy | N controls |
|---|---|---|---|
| ADES | 12,652 | 0 | 8,693 |
| ADSP | 8,328 | 0 | 14,315 |
| UKB AD direct | 4,380 | 0 | 333,752 |
| UKB ADRD direct | 9,610 | 0 | 333,752 |
| UKB AD combined | 4,380 | 64,505 | 333,752 |
| UKB ADRD combined | 9,610 | 63,510 | 333,752 |
| AoU AD direct | 968 | 0 | 132,167 |
| AoU ADRD direct | 2,758 | 0 | 132,167 |
| AoU AD combined | 968 | 23,176 | 132,167 |
| AoU ADRD combined | 2,758 | 23,176 | 132,167 |
| AMP-PD PD direct | 3,267 | 0 | 3,496 |
| AMP-PD PD combined | 3,267 | 532 | 3,496 |
| AMP-PD PDRD direct | 5,893 | 0 | 3,496 |
| AMP-PD PDRD combined | 5,893 | 532 | 3,496 |
| UKB PD direct | 3,942 | 0 | 385,364 |
| UKB PDRD direct | 4,346 | 0 | 385,364 |
| UKB PD combined | 3,942 | 17,188 | 385,364 |
| UKB PDRD combined | 4,346 | 17,162 | 385,364 |
| AoU PD direct | 2,015 | 0 | 150,370 |
| AoU PDRD direct | 2,221 | 0 | 150,370 |
| AoU PD combined | 2,015 | 5,567 | 150,370 |
| AoU PDRD combined | 2,221 | 5,567 | 150,370 |

**Supplementary Table 13.** Participant characteristics for AD- and PD- combined-proxy analyses in the AllOfUs whole-genome sequencing.

| | | | N | Age [mean (std)] | Female [N (%)] | *APOE* [N (%)] | | | | | |
|---|---|---|---|---|---|---|---|---|---|---|---|
| | | | | | | ε2/ε2 | ε2/ε3 | ε2/ε4 | ε3/ε3 | ε3/ε4 | ε4/ε4 |
| All-ancestries | ADRD | Controls | 190,229 | 54.7 (17.4) | 122,473 (64.4) | 1,226 (0.6) | 23,179 (12.2) | 4,064 (2.1) | 118,378 (62.2) | 39,838 (20.9) | 3,544 (1.9) |
| | | Proxy-Cases | 31,970 | 66.8 (11.3) | 20,340 (63.6) | 168 (0.5) | 3,237 (10.1) | 847 (2.6) | 18,121 (56.7) | 8,608 (26.9) | 989 (3.1) |
| | PDRD | Controls | 212,990 | 56.0 (17.3) | 137,384 (64.5) | 1,332 (0.6) | 25,354 (11.9) | 4,708 (2.2) | 130,690 (61.4) | 46,456 (21.8) | 4,377 (2.1) |
| | | Proxy-Cases | 9,284 | 66.3 (12.4) | 5,479 (59.0) | 62 (0.7) | 1,062 (11.4) | 203 (2.2) | 5,809 (62.6) | 1,990 (21.4) | 156 (1.7) |
| European ancestry | ADRD | Controls | 150,370 | 58.7 (16.9) | 94,648 (62.9) | 932 (0.6) | 18,748 (12.5) | 3,226 (2.1) | 92,540 (61.5) | 32,085 (21.3) | 2,783 (1.9) |
| | | Proxy-Cases | 7,788 | 67.2 (12.0) | 4,534 (58.2) | 57 (0.7) | 925 (11.9) | 176 (2.3) | 4,863 (62.4) | 1,650 (21.2) | 116 (1.5) |
| | PDRD | Controls | 132,167 | 57.5 (17.2) | 83,036 (62.8) | 856 (0.6) | 16,991 (12.9) | 2,737 (2.1) | 82,645 (62.5) | 26,805 (20.3) | 2,133 (1.6) |
| | | Proxy-Cases | 25,934 | 67.7 (10.7) | 16,110 (62.1) | 133 (0.5) | 2,682 (10.3) | 665 (2.6) | 14,758 (56.9) | 6,930 (26.7) | 766 (3.0) |

**Supplementary Table 14**. EHR codes included in the ADRD and AD phenotypes for AllOfUs.

| Standard concept name | ADRD | AD | Participant count |
|---|---|---|---|
| Dementia | x | | 2441 |
| Dementia associated with another disease | x | | 1371 |
| Alzheimer's disease | x | x | 881 |
| Vascular dementia without behavioral disturbance | x | | 556 |
| Dementia with behavioral disturbance | x | | 534 |
| Primary degenerative dementia of the Alzheimer type, senile onset | x | x | 247 |
| Mild dementia | x | | 160 |
| Primary degenerative dementia of the Alzheimer type, presenile onset | x | x | 130 |
| Uncomplicated senile dementia | x | | 114 |
| Vascular dementia with behavioral disturbance | x | | 106 |
| Senile dementia of the Lewy body type | x | | 90 |
| Multi-infarct dementia, uncomplicated | x | | 78 |
| Uncomplicated presenile dementia | x | | 71 |
| Moderate dementia | x | | 61 |
| Dementia associated with alcoholism | x | | 48 |
| Huntington's chorea | x | | 42 |
| Vascular dementia | x | | 42 |
| Presenile dementia with depression | x | | 29 |
| Severe dementia | x | | 29 |
| Primary degenerative dementia of the Alzheimer type, senile onset, uncomplicated | x | x | 14 |
| Multi-infarct dementia with depression | x | | 11 |
| General paresis - neurosyphilis | x | | 10 |
| Senile dementia with depression | x | | 9 |
| Senile and presenile organic psychotic conditions | x | | 9 |
| Senile dementia with delusion | x | | 7 |
| Senile dementia with delirium | x | | 6 |
| Dementia of the Alzheimer type with behavioral disturbance | x | x | 6 |
| Drug-induced dementia | x | | 5 |
| Multi-infarct dementia with delirium | x | | 5 |
| Mixed dementia | x | | 4 |
| Presenile dementia with delirium | x | | 4 |
| Multi-infarct dementia with delusions | x | | 4 |
| Primary degenerative dementia of the Alzheimer type, presenile onset, uncomplicated | x | x | 4 |
| Presenile dementia with delusions | x | | 3 |
| Psychoactive substance-induced organic dementia | x | | 3 |
| Multi-infarct dementia | x | | 3 |
| Dementia of frontal lobe type | x | | 3 |
| Presenile dementia | x | | 3 |
| Post-traumatic dementia with behavioral change | x | | 3 |
| Dementia due to Huntington chorea | x | | 1 |
| Progressive aphasia in Alzheimer's disease | x | x | 1 |
| Early onset Alzheimer's disease with behavioral disturbance | x | x | 1 |
| Dementia associated with AIDS | x | | 1 |
| Behavioral disturbance co-occurrent and due to late onset Alzheimer dementia | x | x | 1 |
| Hallucinations co-occurrent and due to late onset dementia | x | | 1 |
| Mixed cortical and subcortical vascular dementia | x | | 1 |
| Senile dementia | x | | 1 |

**Supplementary Table 15.** EHR codes included in the PDRD and PD phenotypes for AllOfUs.

| Standard concept name | PDRD | PD | Participant count |
|---|---|---|---|
| Parkinson's disease | x | x | 2072 |
| Secondary parkinsonism | x | | 234 |
| Parkinsonism due to drug | x | | 128 |
| Senile dementia of the Lewy body type | x | | 90 |
| Diffuse Lewy body disease | x | | 70 |
| Neuroleptic-induced parkinsonism | x | | 55 |
| Vascular parkinsonism | x | | 42 |
| Parkinsonism | x | | 36 |
| Neuroleptic malignant syndrome | x | | 25 |
| Corticobasal degeneration | x | | 16 |
| Striatonigral degeneration | x | | 8 |
| Progressive supranuclear palsy | x | | 4 |
| Lewy body dementia with behavioral disturbance | x | | 4 |
| Atypical Parkinsonism | x | | 2 |
| Postencephalitic parkinsonism | x | | 2 |
| Symptomatic parkinsonism | x | | 1 |
| Young onset Parkinson disease | x | | 1 |
| Orthostatic hypotension co-occurrent and due to Parkinson's disease | x | | 1 |

**Supplementary Table 16.** Participant characteristics for Alzheimer's disease analyses in the ADSP whole-genome sequencing. In ADSP the age for AD cases correspond to the earliest known age-at-onset (or age-at-examination), while age for controls corresponds to the latest known age (or death). Thus, cases are on average younger than controls, and age was not adjusted for in ADSP analyses.

| | | N | Age [mean (std) [non-missing % ]] | Female [N (%)] | *APOE* [N (%)] | | | | | |
|---|---|---|---|---|---|---|---|---|---|---|
| | | | | | ε2/ε2 | ε2/ε3 | ε2/ε4 | ε3/ε3 | ε3/ε4 | ε4/ε4 |
| All | Controls | 28215 | 74.3 (9.1) [98.2] | 17,490 (62.0) | 141 (0.5) | 2,816 (10.0) | 604 (2.1) | 16,929 (60.0) | 6,932 (24.6) | 792 (2.8) |
| All | AD Cases | 13268 | 72.8 (10.4) [97.5] | 8,093 (61.0) | 28 (0.2) | 635 (4.8) | 288 (2.2) | 5,411 (40.8) | 5,243 (39.5) | 1,663 (12.5) |
| European | Controls | 14315 | 76.9 (8.6) [97.9] | 8,660 (60.5) | 64 (0.4) | 1,453 (10.2) | 302 (2.1) | 8,249 (57.6) | 3,770 (26.3) | 477 (3.3) |
| European | AD Cases | 8328 | 72.1 (11.1) [96.5] | 4,791 (57.5) | 14 (0.2) | 316 (3.8) | 170 (2.0) | 3,206 (38.5) | 3,444 (41.4) | 1,178 (14.1) |

**Supplementary Table 17. Cohorts contributing to the ADSP whole-genome sequencing dataset.** The 67 contributing cohorts are listed; additional cohort descriptions are available through NIAGADS DSS under accession ng00067, release v20.

A4 Study (A4)

Adult Changes in Thought (ACT)

Alzheimer's Disease Neuroimaging Initiative (ADNI)

Amish Protective Variant Study

Arizona APOE Cohort Study

ASPrin in Reducing Events in the Elderly (ASPREE) & ASPREE-XT

Atherosclerosis Risk in Communities (ARIC)

Brain Amyloid Cognitive Normal Elders Study (BACNE)

Cache County Study (CCS)

Cardiff EOAD

Cardiovascular Health Study (CHS)

Case Western Reserve University (CWRU) Autopsy

Case Western Reserve University (CWRU) Rapid Decline

Center for Cognitive Neuroscience and Aging (CNSA)

Chicago Health and Aging Project (CHAP)

Corticobasal Degeneration (CBD)

Cuban American Alzheimer’s Disease Initiative (CuAADI)

Erasmus Rucphen Family (ERF)

Estudio Familiar de Influencia Genetica en Alzheimer (EFIGA)

Framingham Heart Study (FHS)

Genetic and Environmental Risk Factors for Alzheimer’s Disease Among African Americans (GenerAAtions)

Genetic Differences (GenDiff)

Gwangju Alzheimer's and Related Dementia (GARD)

Harmonized Diagnostic Assessment of Dementia for the Longitudinal Aging Study of India (LASI-DAD)

Health and Aging Brain Study - Health Disparities (HABS-HD)

Healthy Elderly Active Longevity (HEAL) Cohort “Wellderly”

Heart SCORE

Hillblom Aging Network (HAN)

Human Connectome Project

Indianapolis-Ibadan (IIAA/IIBD)

Knight Alzheimer’s Disease Research Center (KGAD)

Korean Brain Aging Study for the Early Diagnosis and Prediction of AD (KBASE)

LonGenity

Longevity Genes Project (LGP)

Mayo Clinic (MAYO)

Mexican Health and Aging Study (MHAS)

Mexico-Southern California Autosomal Dominant Alzheimer's Disease Consortium

Minority Aging Research Study (MARS)

Mount Sinai Brain Bank (MSBB)

Multi-Institutional Research in Alzheimer's Genetic Epidemiology (MIRAGE)

National Centralized Repository for Alzheimer’s Disease and Related Dementias Family (NCRAD Family)

National Institute of Aging Alzheimer’s Disease Family Based Study (NIA AD-FBS)

National Institute of Mental Health (NIMH)

NIA Alzheimer's Disease Research Centers (ADRC)

Northern Manhattan Study (NOMAS)

Peru Alzheimer’s Disease Initiative (PeADI)

Progressive Supranuclear Palsy (PSP)

Progressive Supranuclear Palsy at the University of California, Los Angeles (PSP UCLA)

Puerto Rican 10/66 Study (PR1066)

Puerto Rican Alzheimer’s Disease Initiative (PRADI)

Religious Orders Study/Memory and Aging Project (ROSMAP)

Research in African-American Alzheimer’s Disease Initiative (REAAADI)

Resource for Early-onset Alzheimer’s Disease Research (READR)

Rotterdam Study (RS)

Stanford Extreme Phenotypes in AD (StEP AD)

Texas Alzheimer’s Research and Care Consortium (TARCC)

The Ginkgo Evaluation of Memory Study

The Monongahela-Youghiogheny Healthy Aging Team (MYHAT)

The University of Alabama Birmingham Alzheimer's Disease Research Center (UAB_ADRC)

University of Miami (MIA)

University of Miami Brain Bank (MBB)

University of Pittsburgh (PITT)

University of Toronto (TOR)

University of Washington Families (RAS)

Vanderbilt University (VAN)

Washington Heights and Inwood Community Aging project (WHICAP)

Wisconsin Registry for Alzheimer’s Prevention (WRAP)

**Supplementary Table 18.** Phenotype breakdown for AMP-PDRD. In the diagnosis column, gray color indicates non-PDRD diagnoses; light blue indicates PDRD diagnoses; dark blue indicates PD and PDRD diagnoses; and green indicates healthy control diagnoses.

| Diagnosis | Parental PD history | PDRD proxy | PDRD proband | PD proxy | PD proband | N |
|---|---|---|---|---|---|---|
| Alzheimer's Disease | FALSE | N/A | N/A | N/A | N/A | 1 |
| Corticobasal Degeneration | FALSE | N/A | N/A | N/A | N/A | 5 |
| Essential Tremor | FALSE | N/A | N/A | N/A | N/A | 28 |
| Fahr's Syndrome | FALSE | N/A | N/A | N/A | N/A | 1 |
| Juvenile Autosomal Recessive Parkinsonism | FALSE | N/A | N/A | N/A | N/A | 1 |
| Multiple System Atrophy | FALSE | N/A | N/A | N/A | N/A | 60 |
| Neuroleptic-Induced Parkinsonism | FALSE | N/A | N/A | N/A | N/A | 2 |
| Olivopontocerebellar Atrophy | FALSE | N/A | N/A | N/A | N/A | 1 |
| Other Neurological Disorder(s) | FALSE | N/A | N/A | N/A | N/A | 15 |
| Possible Alzheimer's Disease | FALSE | N/A | N/A | N/A | N/A | 3 |
| Progressive Supranuclear Palsy | FALSE | N/A | N/A | N/A | N/A | 55 |
| Essential Tremor | TRUE | N/A | N/A | N/A | N/A | 8 |
| Multiple System Atrophy | TRUE | N/A | N/A | N/A | N/A | 1 |
| Other Neurological Disorder(s) | TRUE | N/A | N/A | N/A | N/A | 11 |
| Progressive Supranuclear Palsy | TRUE | N/A | N/A | N/A | N/A | 1 |
| LBD | N/A | 1 | 1 | N/A | N/A | 93 |
| Dementia With Lewy Bodies | FALSE | 1 | 1 | N/A | N/A | 11 |
| LBD | FALSE | 1 | 1 | N/A | N/A | 2506 |
| Dementia With Lewy Bodies | TRUE | 1 | 1 | N/A | N/A | 3 |
| LBD | TRUE | 1 | 1 | N/A | N/A | 15 |
| Idiopathic PD | N/A | 1 | 1 | 1 | 1 | 9 |
| Parkinson's Disease | N/A | 1 | 1 | 1 | 1 | 571 |
| Parkinsonism | N/A | 1 | 1 | 1 | 1 | 1 |
| Idiopathic PD | FALSE | 1 | 1 | 1 | 1 | 762 |
| Parkinson's Disease | FALSE | 1 | 1 | 1 | 1 | 1502 |
| Parkinsonism | FALSE | 1 | 1 | 1 | 1 | 1 |
| Prodromal motor PD | FALSE | 1 | 1 | 1 | 1 | 18 |
| Prodromal non-motor PD | FALSE | 1 | 1 | 1 | 1 | 32 |
| Idiopathic PD | TRUE | 1 | 1 | 1 | 1 | 232 |
| Parkinson's Disease | TRUE | 1 | 1 | 1 | 1 | 168 |
| Parkinsonism | TRUE | 1 | 1 | 1 | 1 | 1 |
| Prodromal motor PD | TRUE | 1 | 1 | 1 | 1 | 18 |
| Prodromal non-motor PD | TRUE | 1 | 1 | 1 | 1 | 2 |
| No PD Nor Other Neurological Disorder | N/A | 0 | 0 | 0 | 0 | 1998 |
| No PD Nor Other Neurological Disorder | FALSE | 0 | 0 | 0 | 0 | 1535 |
| No PD Nor Other Neurological Disorder | TRUE | 1 | N/A | 1 | N/A | 533 |

**Supplementary Table 19.** Participant characteristics for Parkinson's disease (PD-combined-proxy) analyses in the AMP-PD whole-genome sequencing. Age refers to the age at baseline visit.

| | | N | Age [mean (std)] | Female [N (%)] | *APOE* [N (%)] | | | | | |
|---|---|---|---|---|---|---|---|---|---|---|
| | | | | | ε2/ε2 | ε2/ε3 | ε2/ε4 | ε3/ε3 | ε3/ε4 | ε4/ε4 |
| All | Controls | 3533 | 68.5 (13.7) | 1668 (47.2) | 13 (0.4) | 393 (11.1) | 50 (1.4) | 2263 (64.1) | 758 (21.5) | 56 (1.6) |
| All | PD Proxy-cases | 3850 | 63.3 (10.6) | 2236 (58.1) | 23 (0.6) | 475 (12.3) | 64 (1.7) | 2437 (63.3) | 797 (20.7) | 54 (1.4) |
| European | Controls | 3496 | 68.6 (13.7) | 1658 (47.4) | 13 (0.4) | 386 (11.0) | 49 (1.4) | 2240 (64.1) | 752 (21.5) | 56 (1.6) |
| European | PD Proxy-cases | 3799 | 63.3 (10.6) | 2210 (58.2) | 20 (0.5) | 469 (12.3) | 64 (1.7) | 2409 (63.4) | 786 (20.7) | 51 (1.3) |

**Supplementary Table 20. Cohorts contributing to the AMP-PDRD whole-genome sequencing dataset.**

| |
|---|
| BioFIND |
| Harvard Biomarkers Discovery |
| Lewy Body Dementia Study |
| LRRK2 Cohort Consortium |
| Parkinson's Disease Biomarkers Program |
| Parkinson's Progression Markers Initiative |
| STEADY-PD Phase 3 |
| SURE-PD Phase 3 |
