## Supplementary_Fig13 for "Population-scale burden analysis of rare damaging coding variants identifies novel risk genes for Alzheimer’s disease and related dementias and Parkinson’s disease and related disorders"

GBA1: PDRD combined-proxy single-variant associations (ALL)

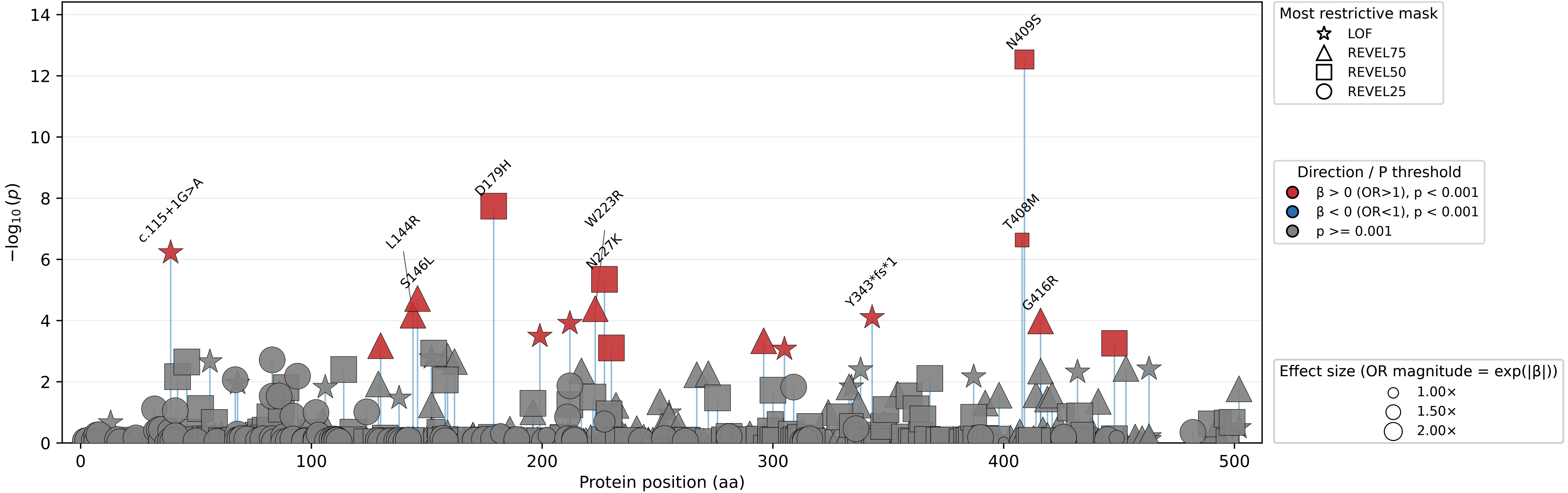

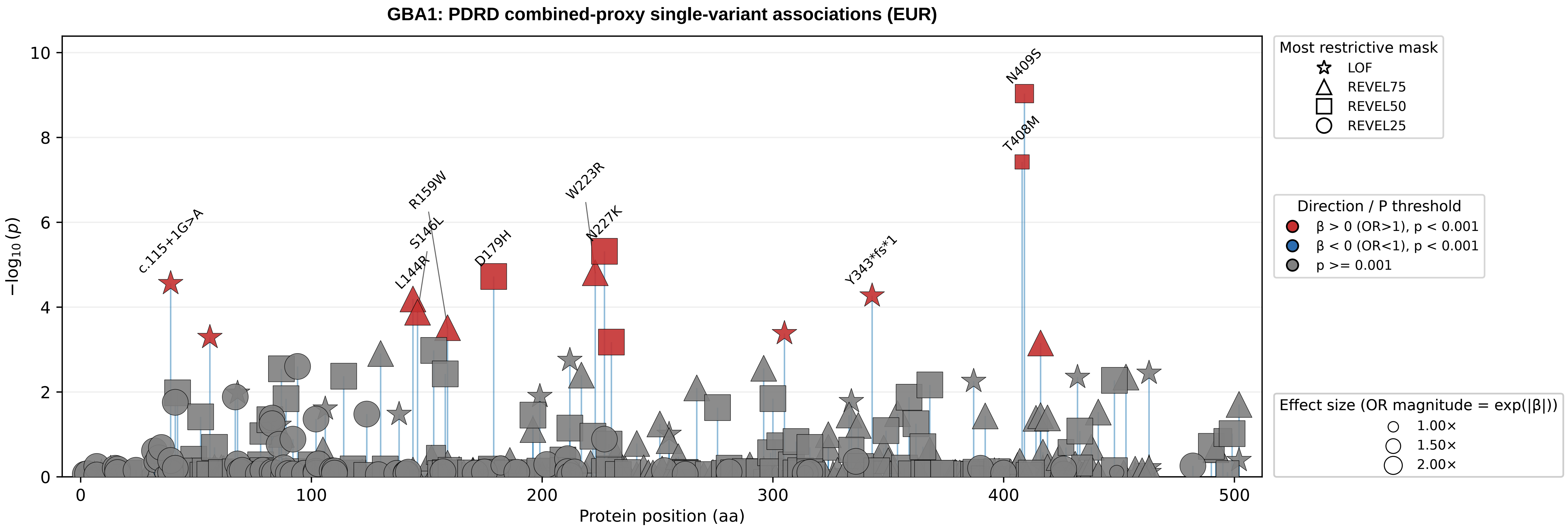

GBA1: PDRD combined-proxy single-variant associations (EUR)
N409S
T408M
W223R
N227K
R159W
S146L
L144R
D179H
c.115+1G>A
Y343*fs*1
−log10(p)
Protein position (aa)
0
2
4
6
8
10
100
200
300
400
500
Most restrictive mask
LOF
REVEL75
REVEL50
REVEL25
Direction / P threshold
β > 0 (OR>1), p < 0.001
β < 0 (OR<1), p < 0.001
p >= 0.001
Effect size (OR magnitude = exp(|β|))
1.00×
1.50×
2.00×

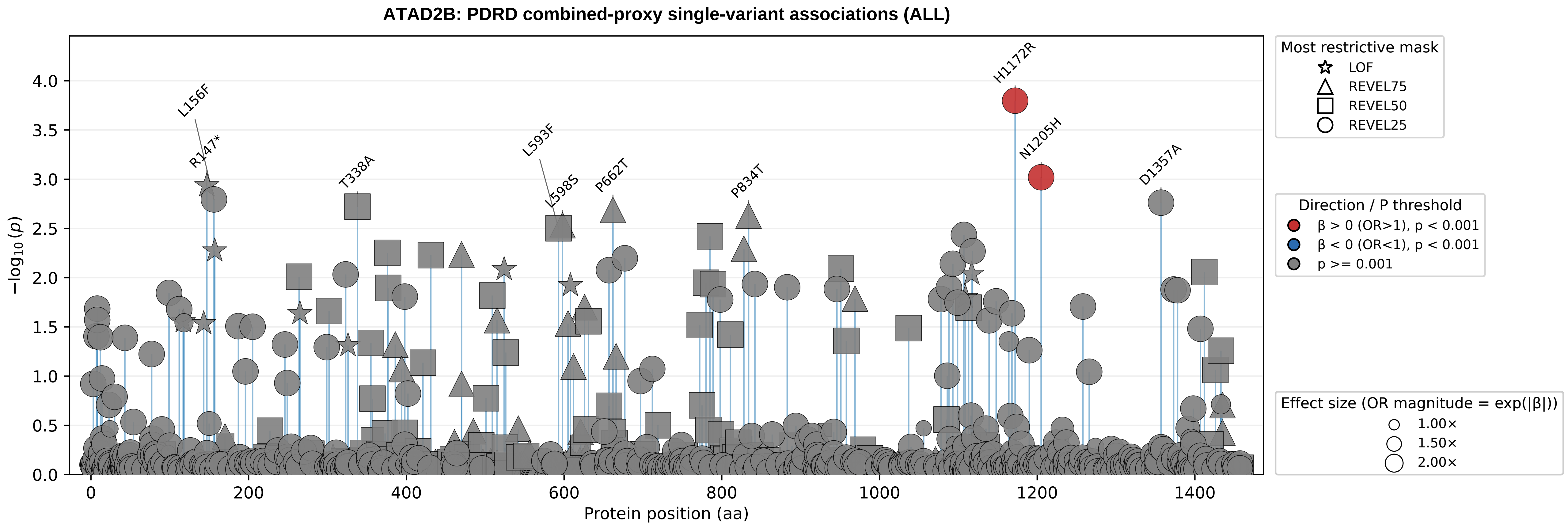
ATAD2B: PDRD combined-proxy single-variant associations (ALL)
−log10 (p)
Protein position (aa)
L156F
R147*
T338A
L593F
L598S
P662T
P834T
H1172R
N1205H
D1357A
Most restrictive mask
LOF
REVEL75
REVEL50
REVEL25
Direction / P threshold
β > 0 (OR>1), p < 0.001
β < 0 (OR<1), p < 0.001
p >= 0.001
Effect size (OR magnitude = exp(|β|))
1.00×
1.50×
2.00×

### ATAD2B: PDRD combined-proxy single-variant associations (EUR)

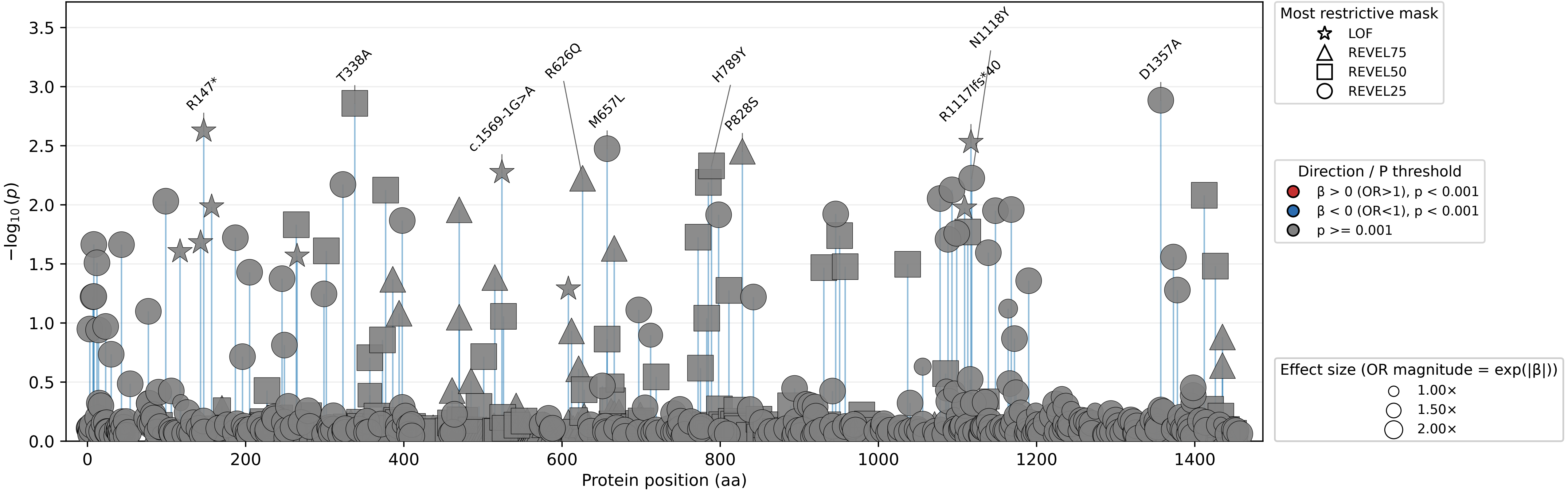

**KANSL3: PDRD combined-proxy single-variant associations (ALL)**

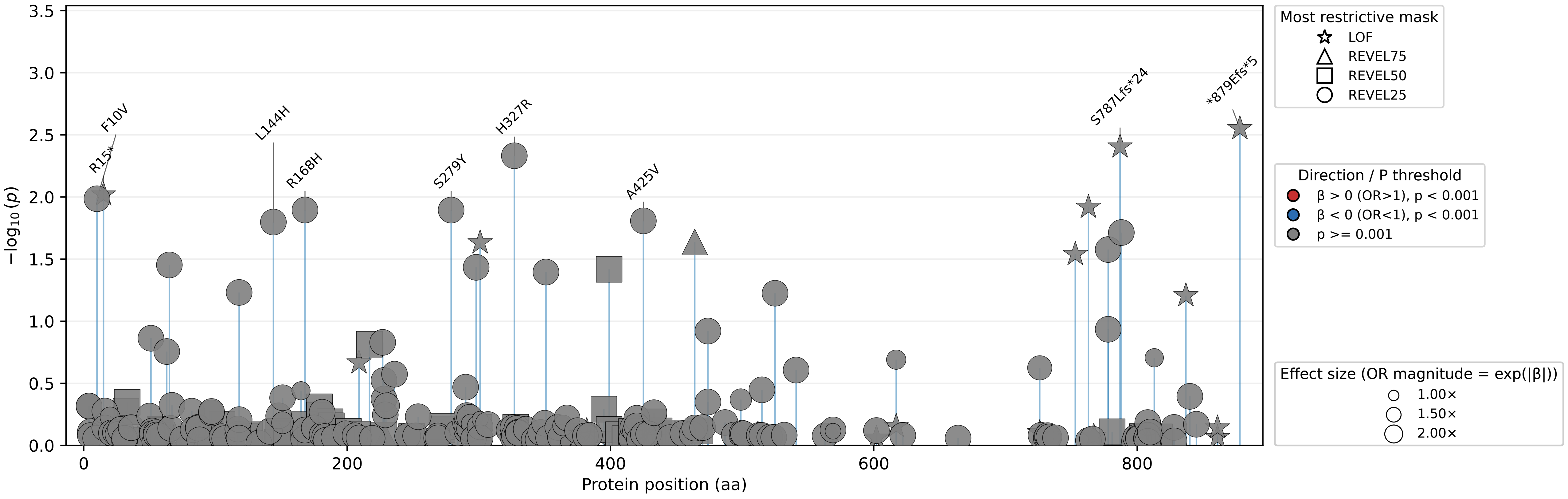

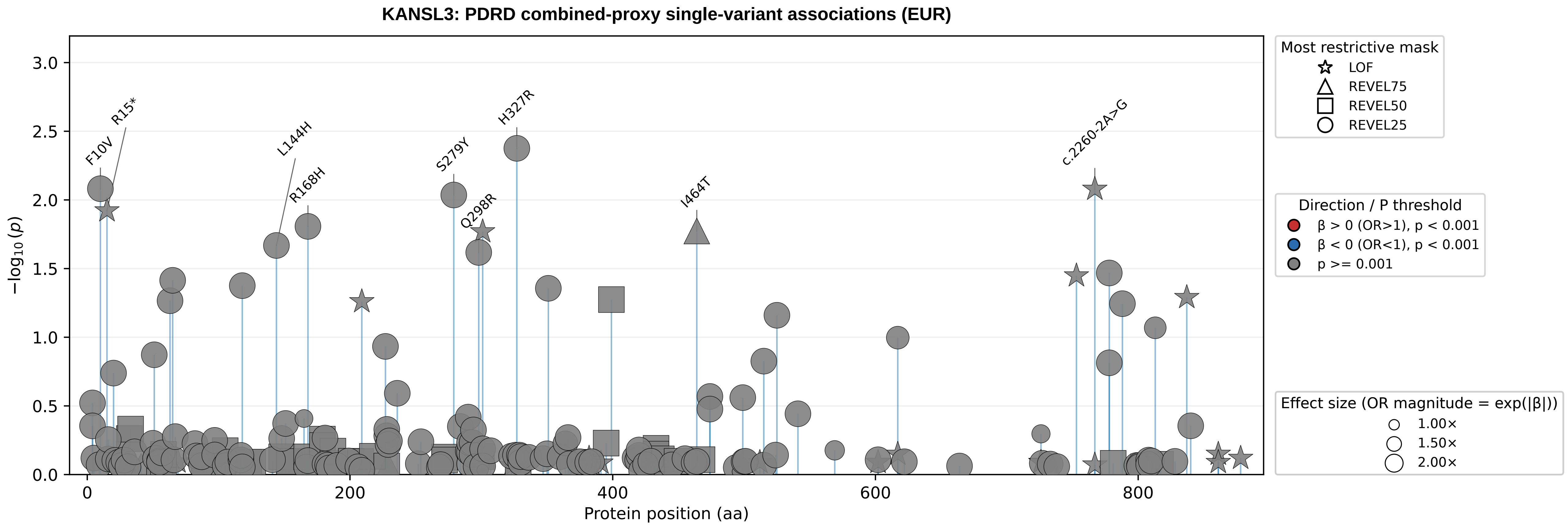

KANSL3: PDRD combined-proxy single-variant associations (EUR)
−log10(p)
Protein position (aa)
F10V
R15*
L144H
R168H
S279Y
Q298R
H327R
I464T
c.2260-2A>G
Most restrictive mask
LOF
REVEL75
REVEL50
REVEL25
Direction / P threshold
β > 0 (OR>1), p < 0.001
β < 0 (OR<1), p < 0.001
p >= 0.001
Effect size (OR magnitude = exp(|β|))
1.00×
1.50×
2.00×

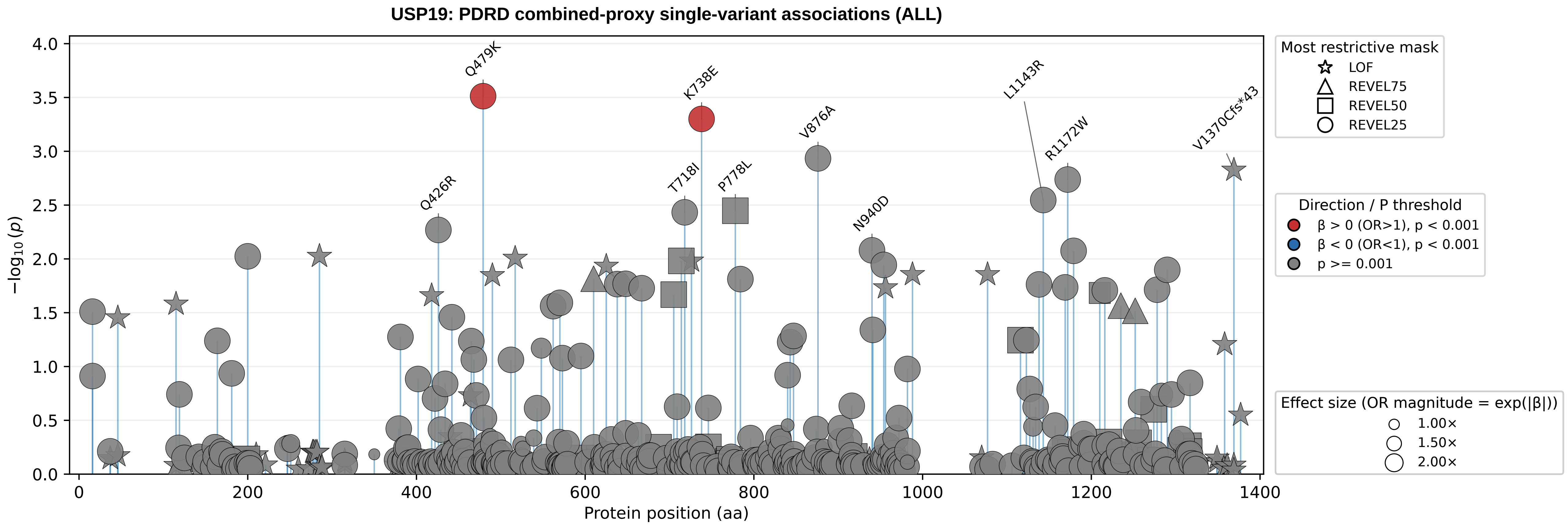

USP19: PDRD combined-proxy single-variant associations (ALL)
−log10(p)
Protein position (aa)
Q479K
K738E
V876A
L1143R
R1172W
V1370Cfs*43
Q426R
T718I
P778L
N940D
Most restrictive mask
LOF
REVEL75
REVEL50
REVEL25
Direction / P threshold
β > 0 (OR>1), p < 0.001
β < 0 (OR<1), p < 0.001
p >= 0.001
Effect size (OR magnitude = exp(|β|))
1.00×
1.50×
2.00×

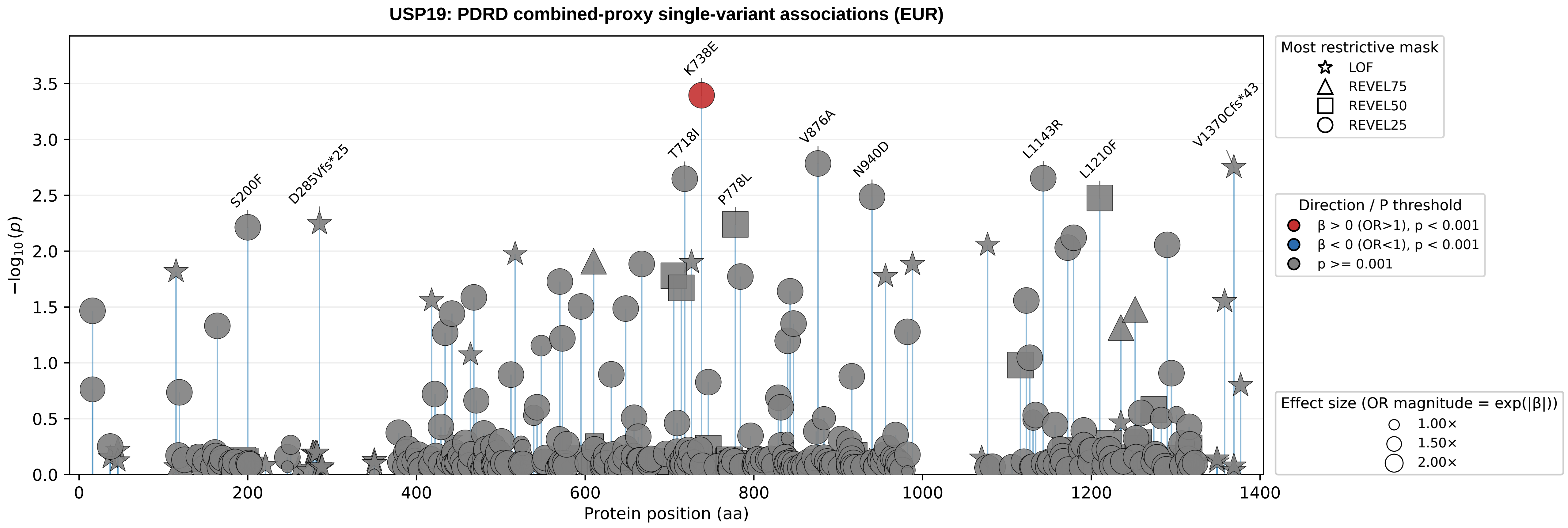

USP19: PDRD combined-proxy single-variant associations (EUR)
−log10(p)
Protein position (aa)
K738E
T718I
P778L
V876A
N940D
L1143R
L1210F
V1370Cfs*43
S200F
D285Vfs*25
Most restrictive mask
LOF
REVEL75
REVEL50
REVEL25
Direction / P threshold
β > 0 (OR>1), p < 0.001
β < 0 (OR<1), p < 0.001
p >= 0.001
Effect size (OR magnitude = exp(|β|))
1.00×
1.50×
2.00×

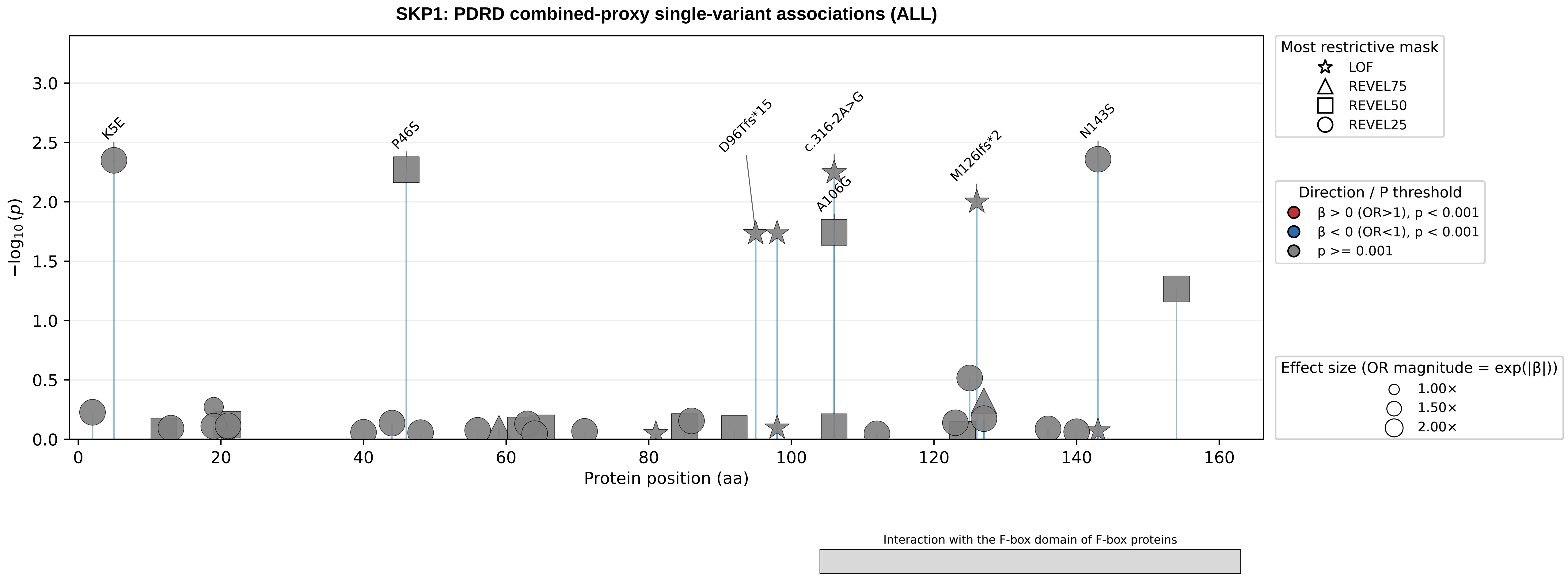

SKP1: PDRD combined-proxy single-variant associations (ALL)
−log10(p)
3.0
2.5
2.0
1.5
1.0
0.5
0.0
0
20
40
60
80
100
120
140
160
Protein position (aa)
K5E
P46S
D96Tfs*15
c.316-2A>G
A106G
M126Ifs*2
N143S
Most restrictive mask
LOF
REVEL75
REVEL50
REVEL25
Direction / P threshold
β > 0 (OR>1), p < 0.001
β < 0 (OR<1), p < 0.001
p >= 0.001
Effect size (OR magnitude = exp(|β|))
1.00×
1.50×
2.00×
Interaction with the F-box domain of F-box proteins

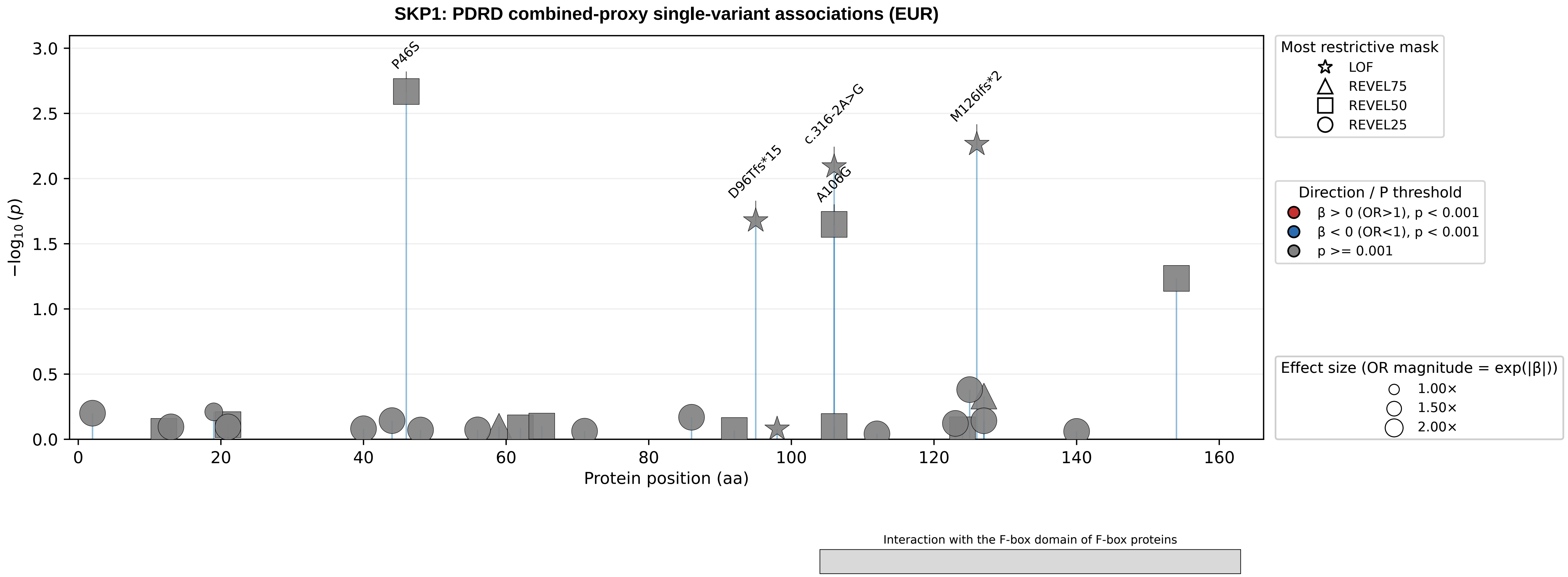

SKP1: PDRD combined-proxy single-variant associations (EUR)
P46S
D96Tfs*15
c.316-2A>G
A106G
M126Ifs*2
−log10(p)
3.0
2.5
2.0
1.5
1.0
0.5
0.0
0
20
40
60
80
100
120
140
160
Protein position (aa)
Most restrictive mask
LOF
REVEL75
REVEL50
REVEL25
Direction / P threshold
β > 0 (OR>1), p < 0.001
β < 0 (OR<1), p < 0.001
p >= 0.001
Effect size (OR magnitude = exp(|β|))
1.00×
1.50×
2.00×
Interaction with the F-box domain of F-box proteins

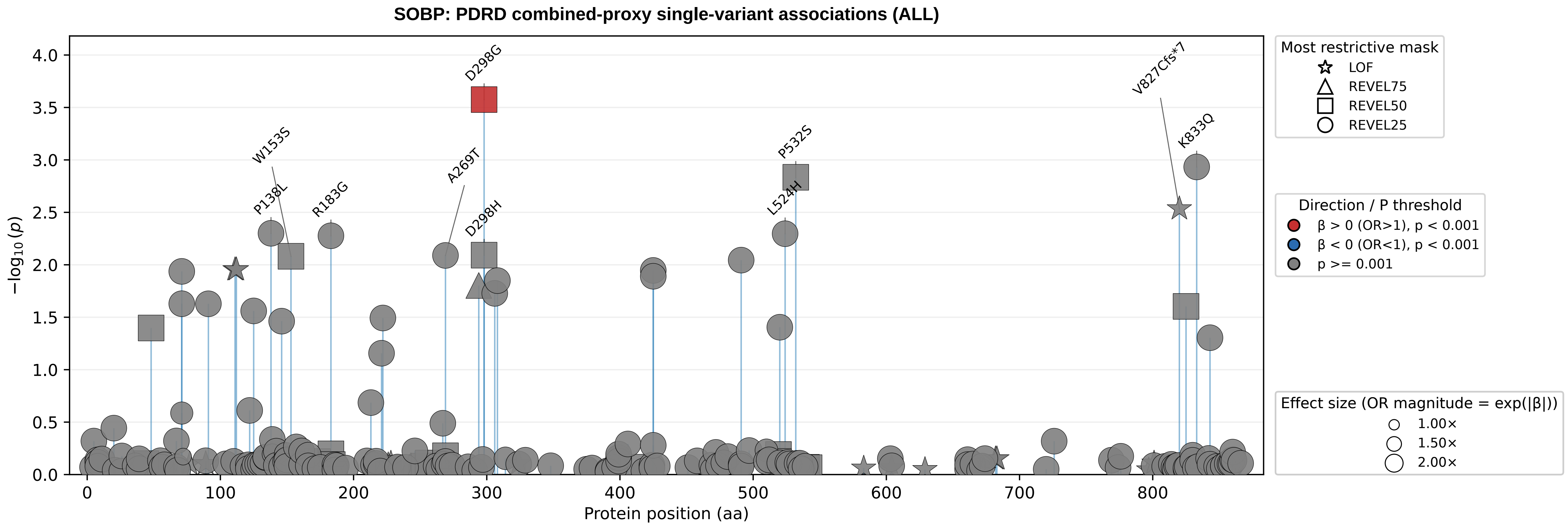
SOBP: PDRD combined-proxy single-variant associations (ALL)
−log10(p)
Protein position (aa)
D298G
W153S
A269T
P532S
P138L
R183G
D298H
L524H
V827Cfs*7
K833Q
Most restrictive mask
LOF
REVEL75
REVEL50
REVEL25
Direction / P threshold
β > 0 (OR>1), p < 0.001
β < 0 (OR<1), p < 0.001
p >= 0.001
Effect size (OR magnitude = exp(|β|))
1.00×
1.50×
2.00×

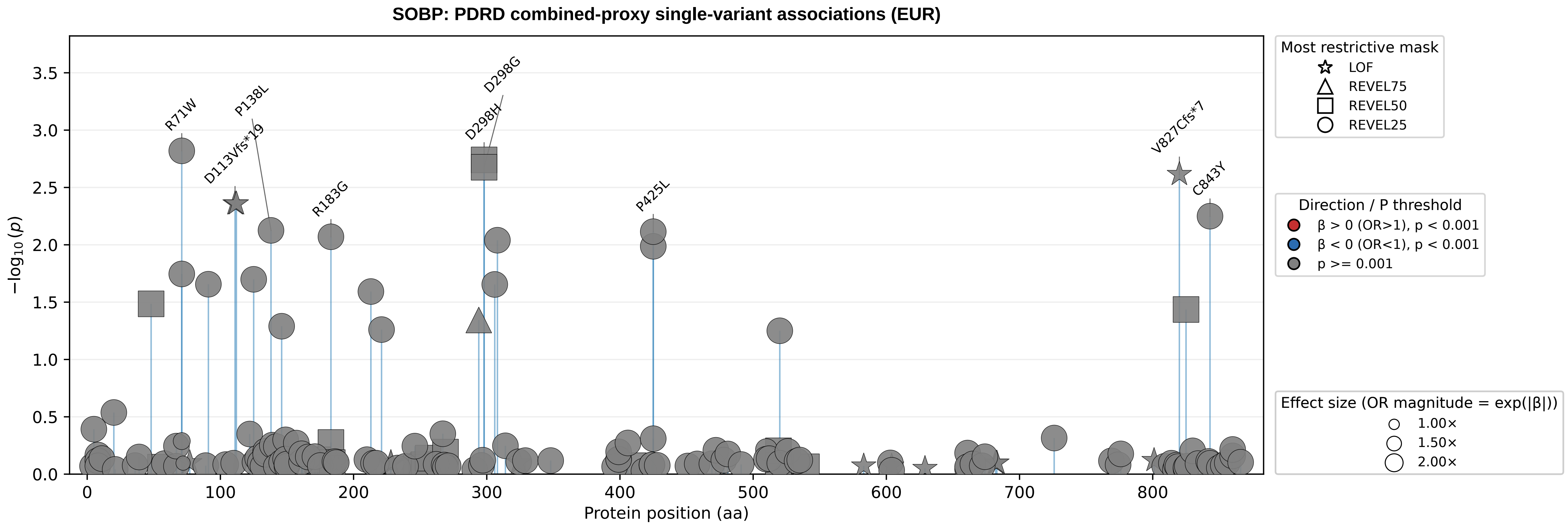

SOBP: PDRD combined-proxy single-variant associations (EUR)
−log10(p)
Protein position (aa)
R71W
P138L
D113Vfs*19
R183G
D298G
D298H
P425L
V827Cfs*7
C843Y
Most restrictive mask
LOF
REVEL75
REVEL50
REVEL25
Direction / P threshold
β > 0 (OR>1), p < 0.001
β < 0 (OR<1), p < 0.001
p >= 0.001
Effect size (OR magnitude = exp(|β|))
1.00×
1.50×
2.00×

### TRAF3IP2: PDRD combined-proxy single-variant associations (ALL)

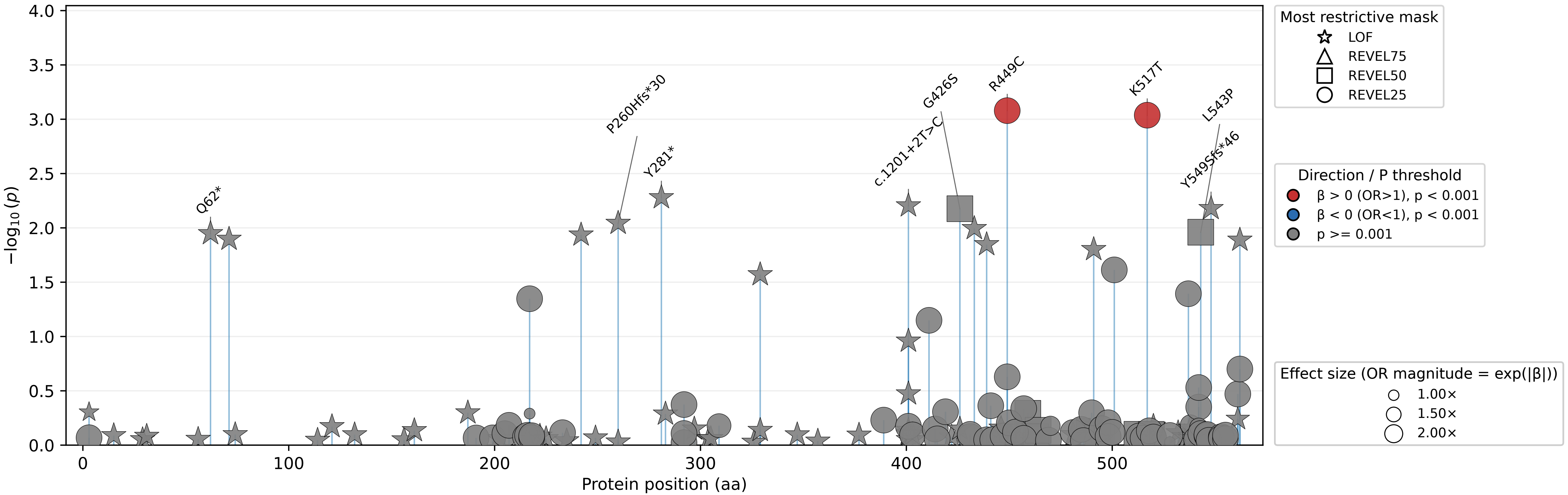

**TRAF3IP2: PDRD combined-proxy single-variant associations (EUR)**

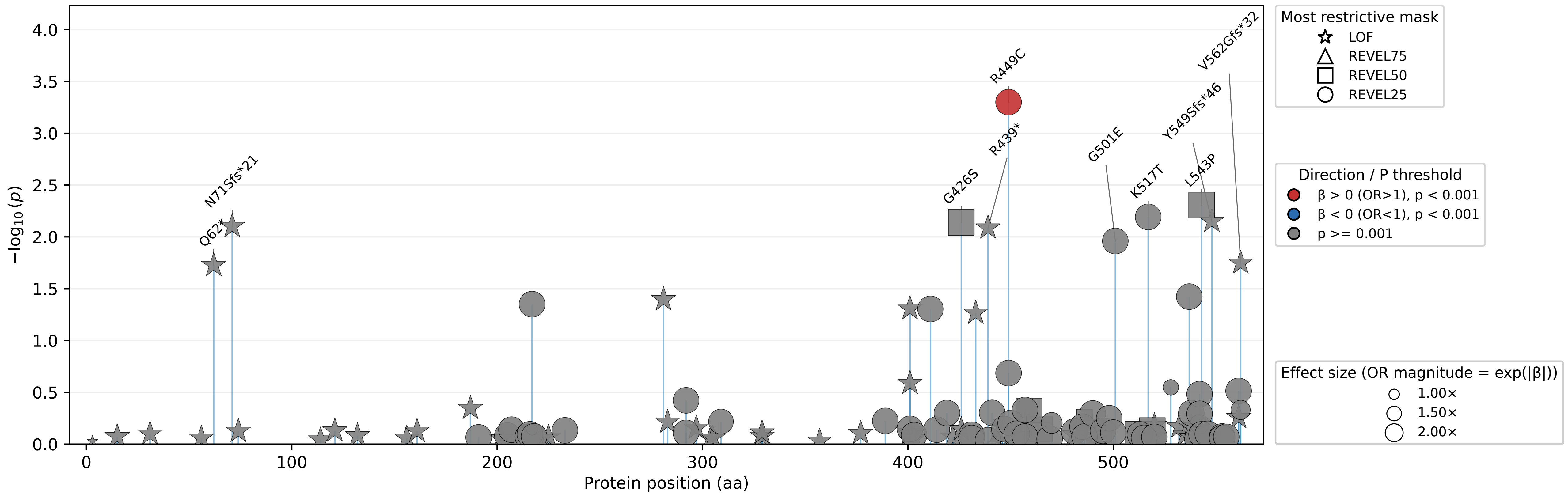

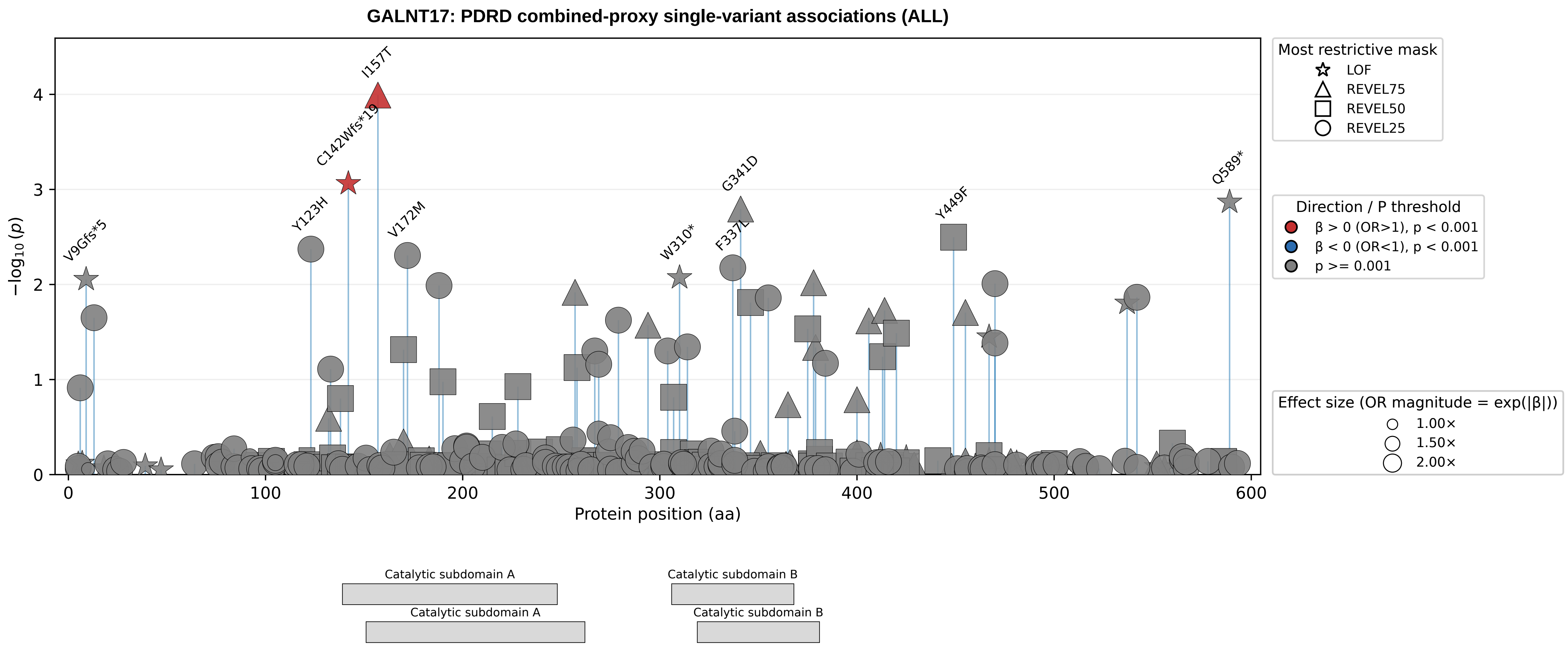

GALNT17: PDRD combined-proxy single-variant associations (ALL)
−log10(p)
Protein position (aa)
I157T
C142Wfs*19
G341D
Q589*
Y123H
V172M
F337L
Y449F
V9Gfs*5
W310*
Most restrictive mask
LOF
REVEL75
REVEL50
REVEL25
Direction / P threshold
β > 0 (OR>1), p < 0.001
β < 0 (OR<1), p < 0.001
p >= 0.001
Effect size (OR magnitude = exp(|β|))
1.00×
1.50×
2.00×
Catalytic subdomain A
Catalytic subdomain B
Catalytic subdomain A
Catalytic subdomain B

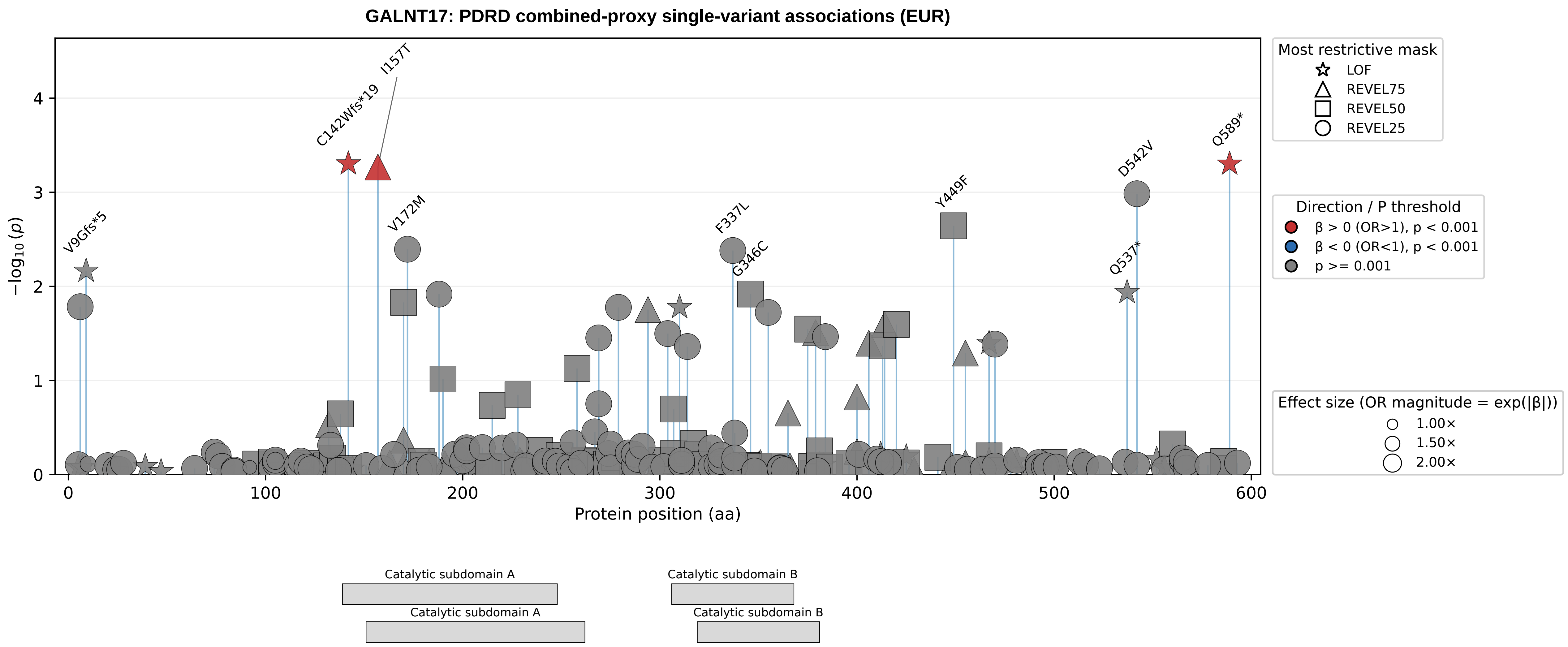
GALNT17: PDRD combined-proxy single-variant associations (EUR)
−log10(p)
Protein position (aa)
0
1
2
3
4
0
100
200
300
400
500
600
V9Gfs*5
C142Wfs*19
I157T
V172M
F337L
G346C
Y449F
D542V
Q537*
Q589*
Catalytic subdomain A
Catalytic subdomain A
Catalytic subdomain B
Catalytic subdomain B
Most restrictive mask
LOF
REVEL75
REVEL50
REVEL25
Direction / P threshold
β > 0 (OR>1), p < 0.001
β < 0 (OR<1), p < 0.001
p >= 0.001
Effect size (OR magnitude = exp(|β|))
1.00×
1.50×
2.00×

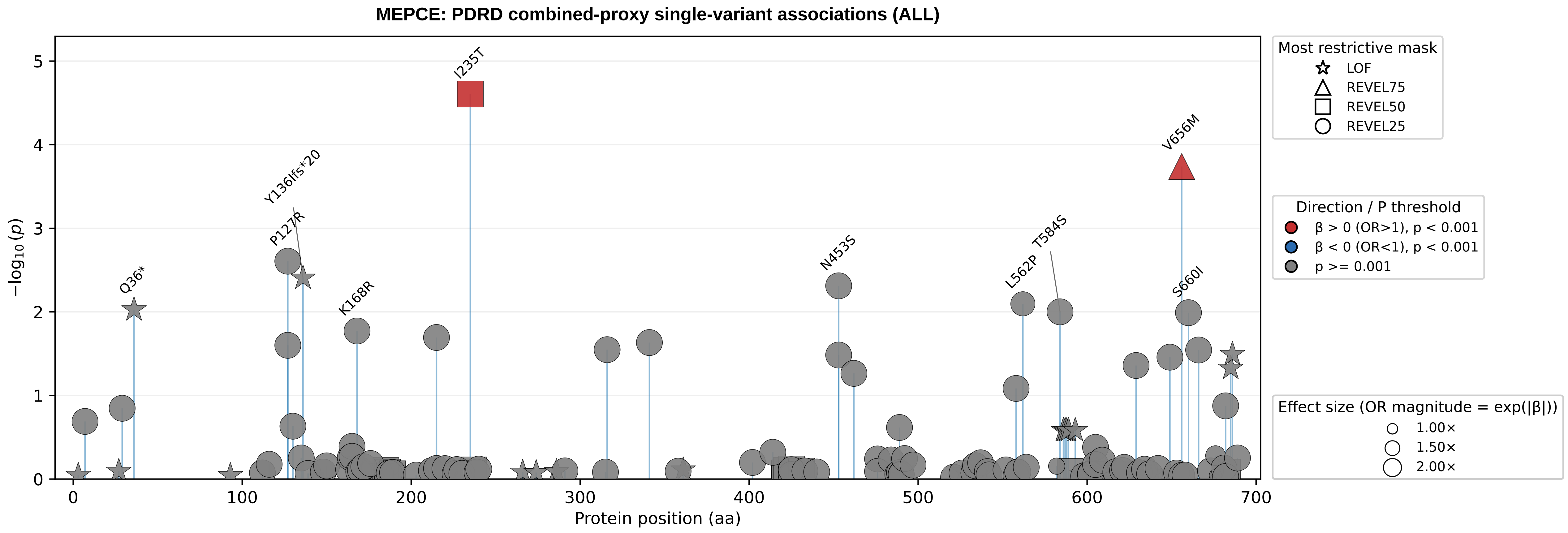

MEPCE: PDRD combined-proxy single-variant associations (ALL)
I235T
V656M
Y136Ifs*20
P127R
Q36*
K168R
N453S
L562P
T584S
S660I
−log10(p)
Protein position (aa)
Most restrictive mask
LOF
REVEL75
REVEL50
REVEL25
Direction / P threshold
β > 0 (OR>1), p < 0.001
β < 0 (OR<1), p < 0.001
p >= 0.001
Effect size (OR magnitude = exp(|β|))
1.00×
1.50×
2.00×

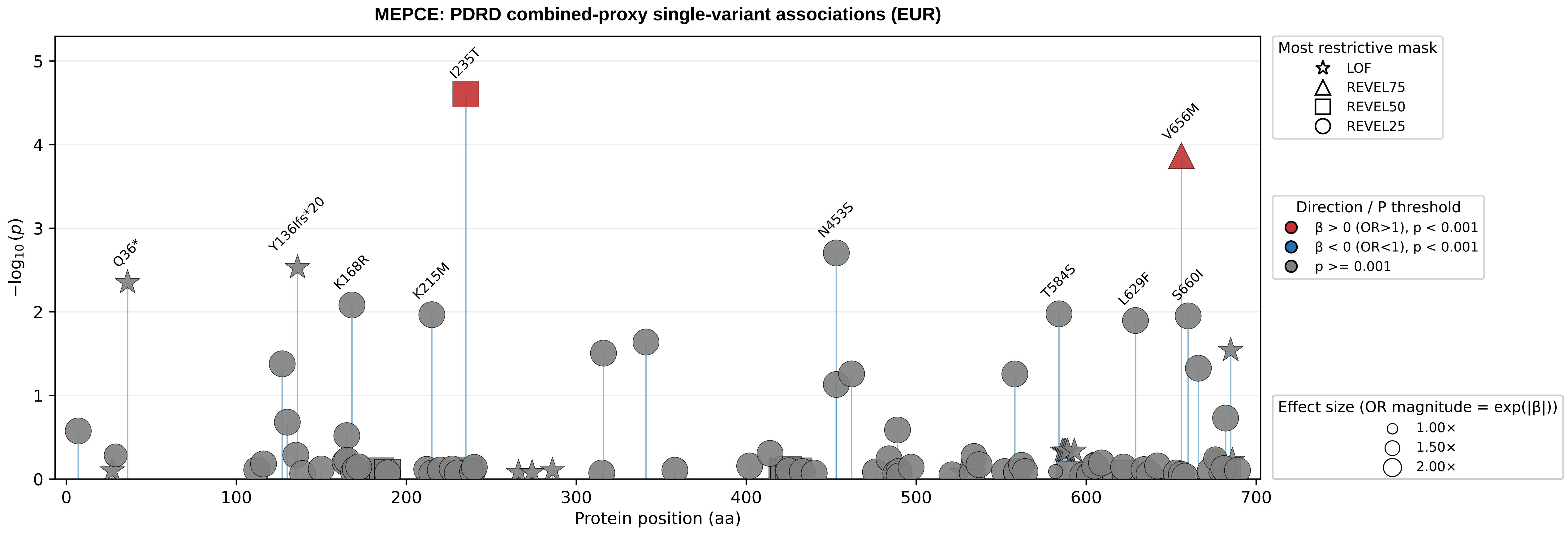

MEPCE: PDRD combined-proxy single-variant associations (EUR)
−log10(p)
Protein position (aa)
I235T
V656M
Q36*
Y136Ifs*20
K168R
K215M
N453S
T584S
L629F
S660I
Most restrictive mask
LOF
REVEL75
REVEL50
REVEL25
Direction / P threshold
β > 0 (OR>1), p < 0.001
β < 0 (OR<1), p < 0.001
p >= 0.001
Effect size (OR magnitude = exp(|β|))
1.00×
1.50×
2.00×

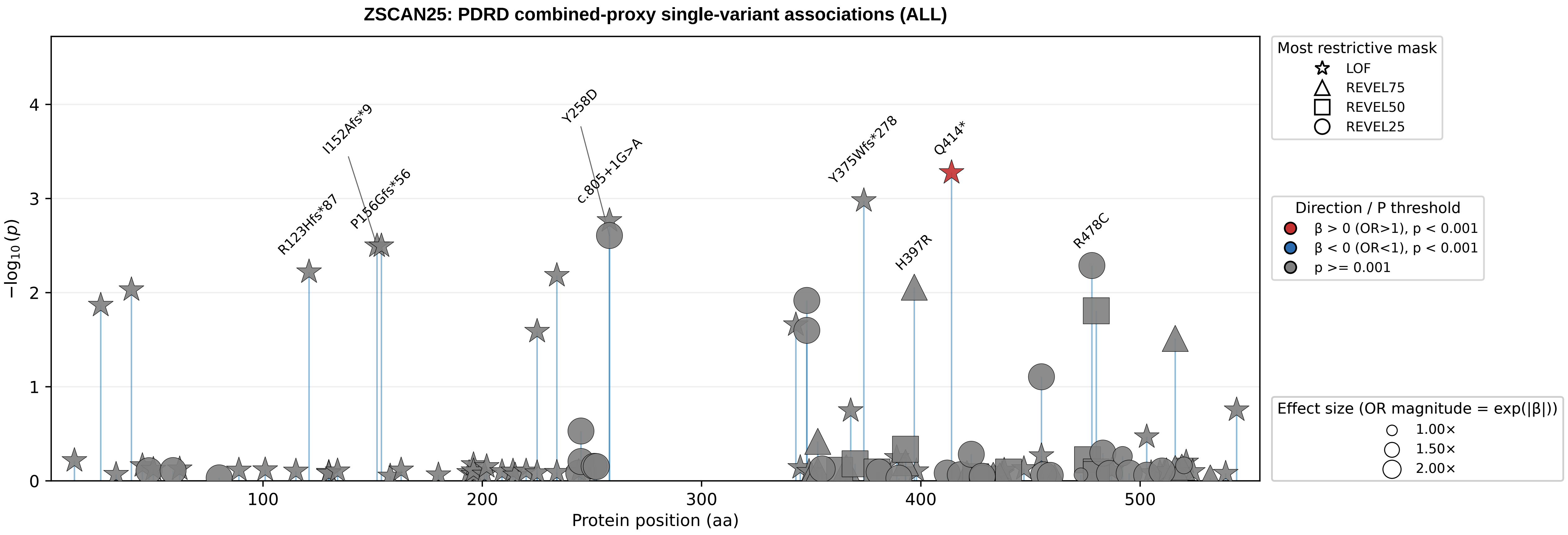
ZSCAN25: PDRD combined-proxy single-variant associations (ALL)
I152Afs*9
Y258D
c.805+1G>A
Y375Wfs*278
Q414*
R123Hfs*87
P156Gfs*56
H397R
R478C
−log10(p)
Protein position (aa)
0
1
2
3
4
100
200
300
400
500
Most restrictive mask
LOF
REVEL75
REVEL50
REVEL25
Direction / P threshold
β > 0 (OR>1), p < 0.001
β < 0 (OR<1), p < 0.001
p >= 0.001
Effect size (OR magnitude = exp(|β|))
1.00×
1.50×
2.00×

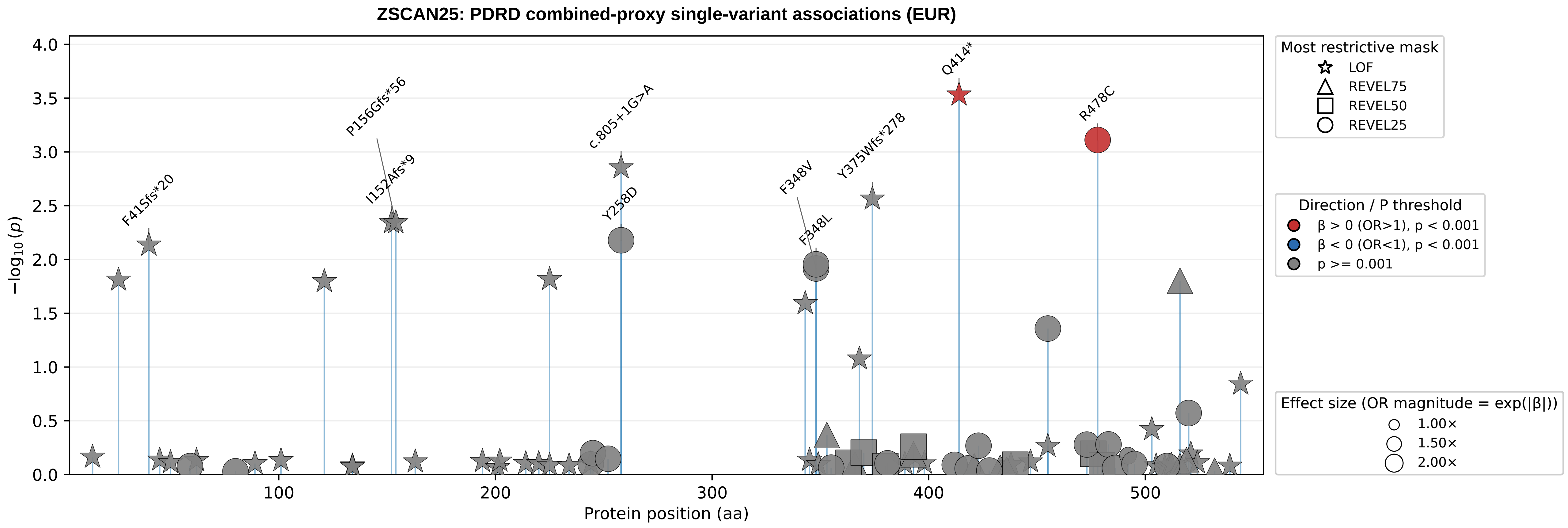
ZSCAN25: PDRD combined-proxy single-variant associations (EUR)
−log10 (p)
Protein position (aa)
F41Sfs*20
P156Gfs*56
I152Afs*9
c.805+1G>A
Y258D
F348V
F348L
Y375Wfs*278
Q414*
R478C
Most restrictive mask
LOF
REVEL75
REVEL50
REVEL25
Direction / P threshold
β > 0 (OR>1), p < 0.001
β < 0 (OR<1), p < 0.001
p >= 0.001
Effect size (OR magnitude = exp(|β|))
1.00×
1.50×
2.00×

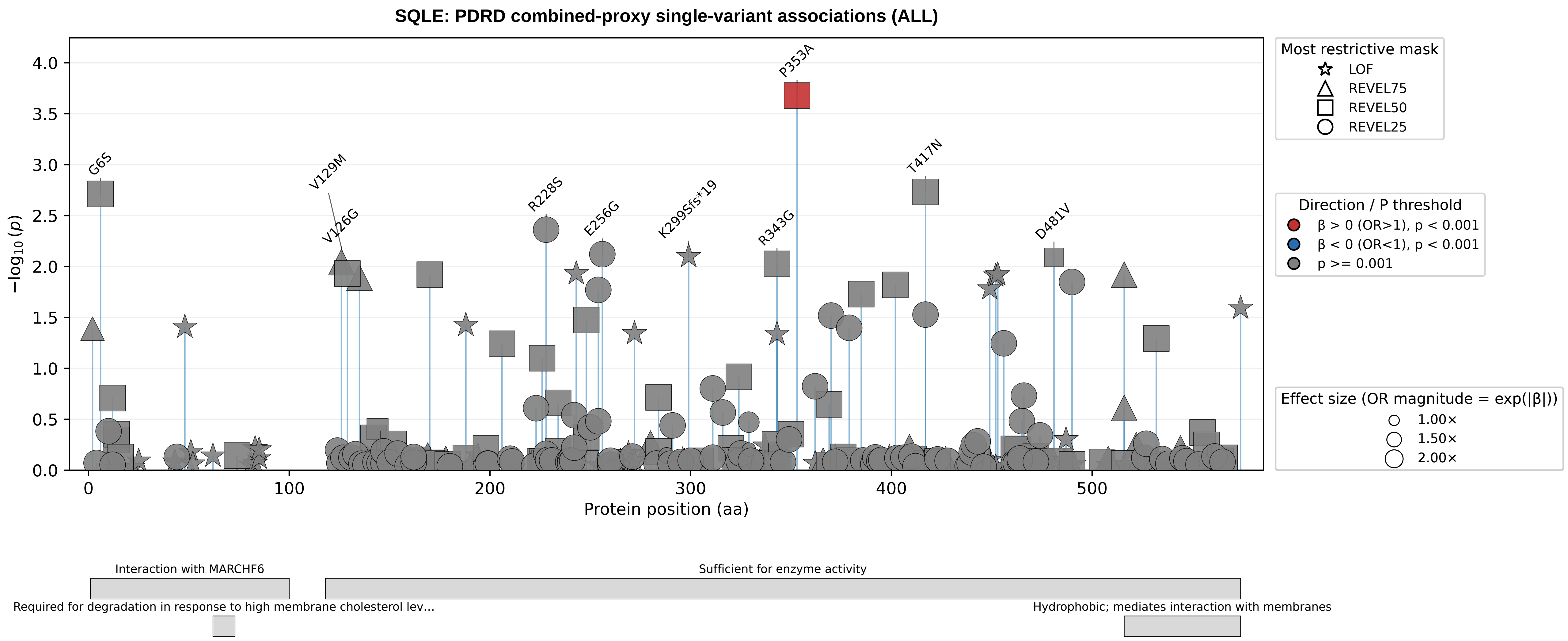

SQLE: PDRD combined-proxy single-variant associations (ALL)
−log10(p)
Protein position (aa)
G6S
V129M
V126G
R228S
E256G
K299Sfs*19
R343G
P353A
T417N
D481V
Most restrictive mask
LOF
REVEL75
REVEL50
REVEL25
Direction / P threshold
β > 0 (OR>1), p < 0.001
β < 0 (OR<1), p < 0.001
p >= 0.001
Effect size (OR magnitude = exp(|β|))
1.00×
1.50×
2.00×
Interaction with MARCHF6
Sufficient for enzyme activity
Required for degradation in response to high membrane cholesterol lev...
Hydrophobic; mediates interaction with membranes

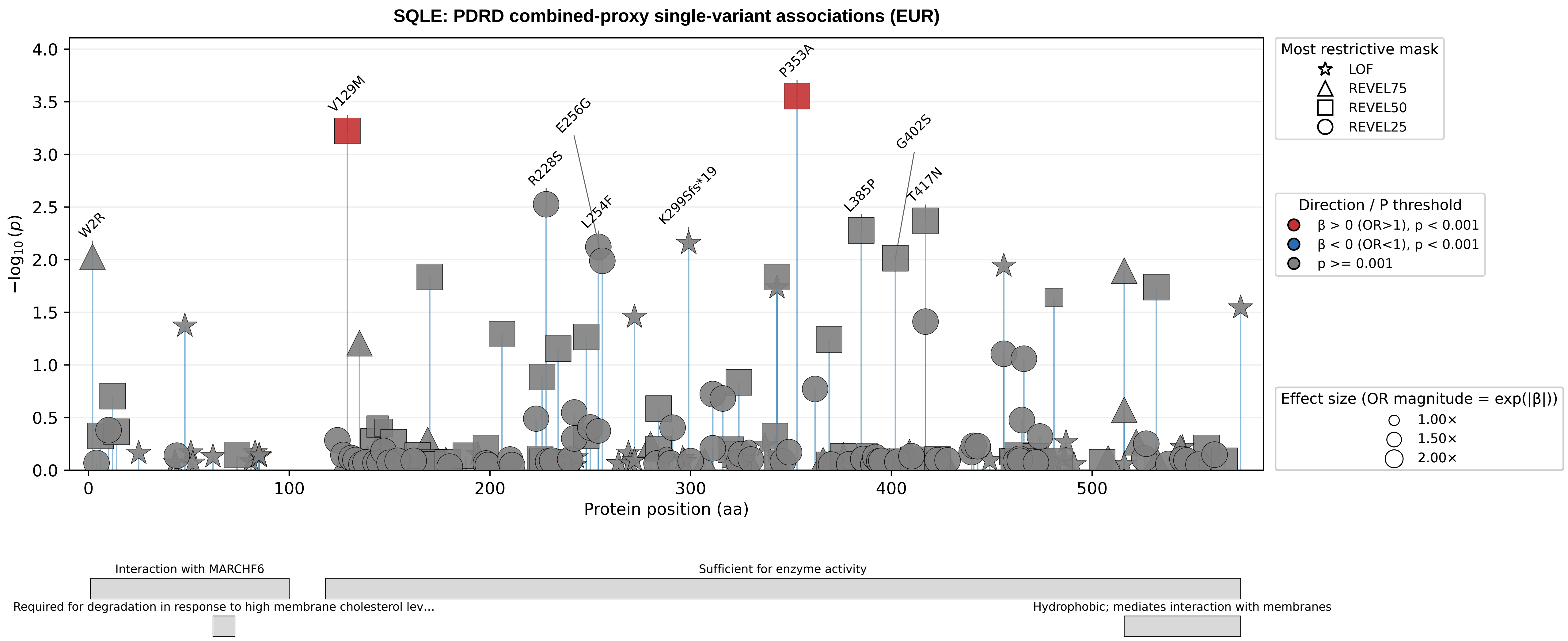

SQLE: PDRD combined-proxy single-variant associations (EUR)
−log10(p)
Protein position (aa)
W2R
V129M
R228S
E256G
L254F
K299Sfs*19
P353A
L385P
G402S
T417N
Most restrictive mask
LOF
REVEL75
REVEL50
REVEL25
Direction / P threshold
β > 0 (OR>1), p < 0.001
β < 0 (OR<1), p < 0.001
p >= 0.001
Effect size (OR magnitude = exp(|β|))
1.00×
1.50×
2.00×
Interaction with MARCHF6
Sufficient for enzyme activity
Required for degradation in response to high membrane cholesterol lev...
Hydrophobic; mediates interaction with membranes

**BNIP3: PDRD combined-proxy single-variant associations (ALL)**

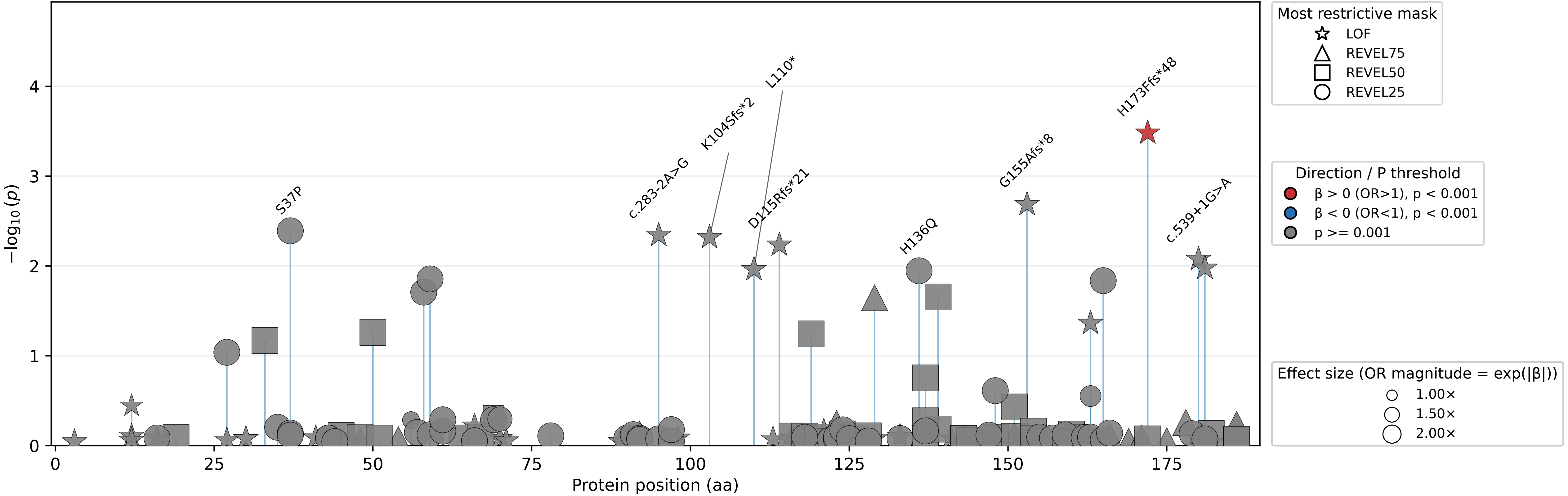

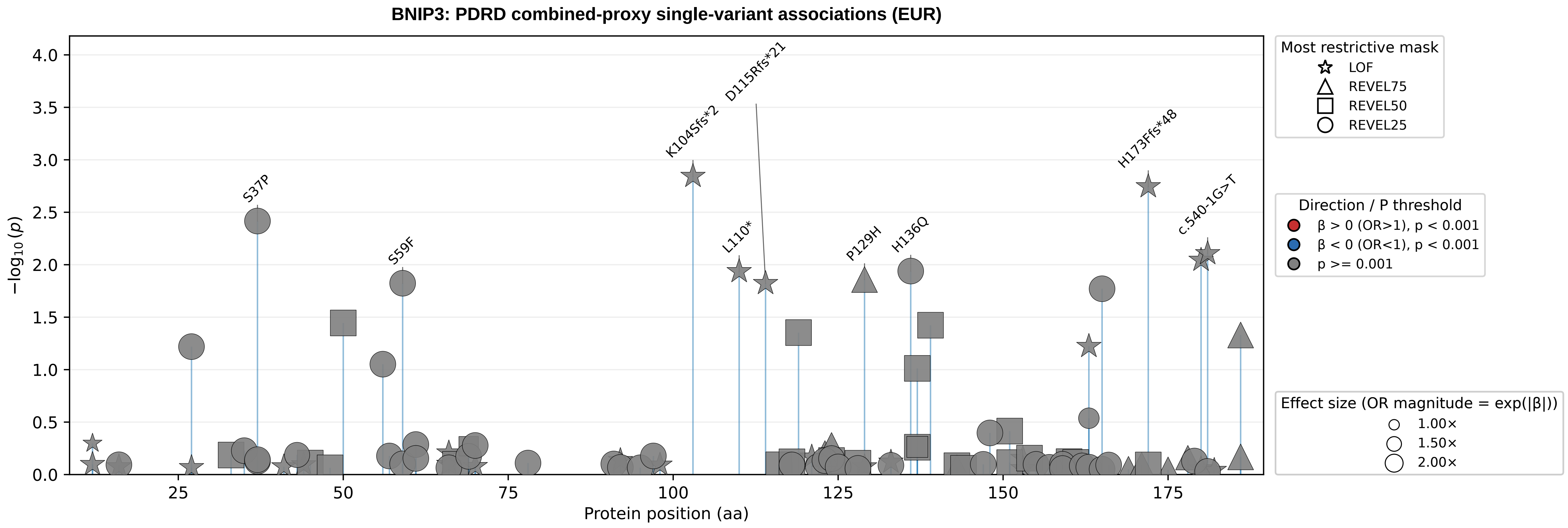
BNIP3: PDRD combined-proxy single-variant associations (EUR)
−log10(p)
Protein position (aa)
S37P
S59F
K104Sfs*2
D115Rfs*21
L110*
P129H
H136Q
H173Ffs*48
c.540-1G>T
Most restrictive mask
LOF
REVEL75
REVEL50
REVEL25
Direction / P threshold
β > 0 (OR>1), p < 0.001
β < 0 (OR<1), p < 0.001
p >= 0.001
Effect size (OR magnitude = exp(|β|))
1.00×
1.50×
2.00×

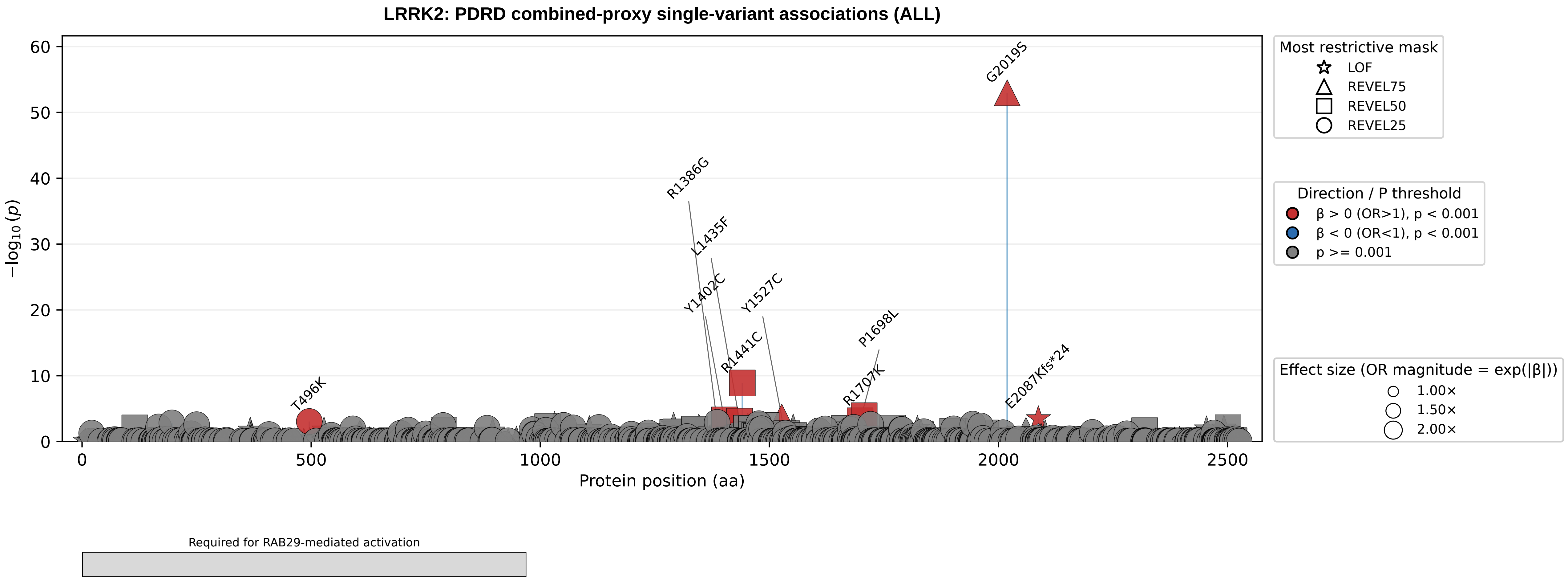

LRRK2: PDRD combined-proxy single-variant associations (ALL)
−log10(p)
0
10
20
30
40
50
60
Protein position (aa)
0
500
1000
1500
2000
2500
G2019S
R1386G
L1435F
Y1402C
Y1527C
R1441C
P1698L
R1707K
E2087Kfs*24
T496K
Most restrictive mask
LOF
REVEL75
REVEL50
REVEL25
Direction / P threshold
β > 0 (OR>1), p < 0.001
β < 0 (OR<1), p < 0.001
p >= 0.001
Effect size (OR magnitude = exp(|β|))
1.00×
1.50×
2.00×
Required for RAB29-mediated activation

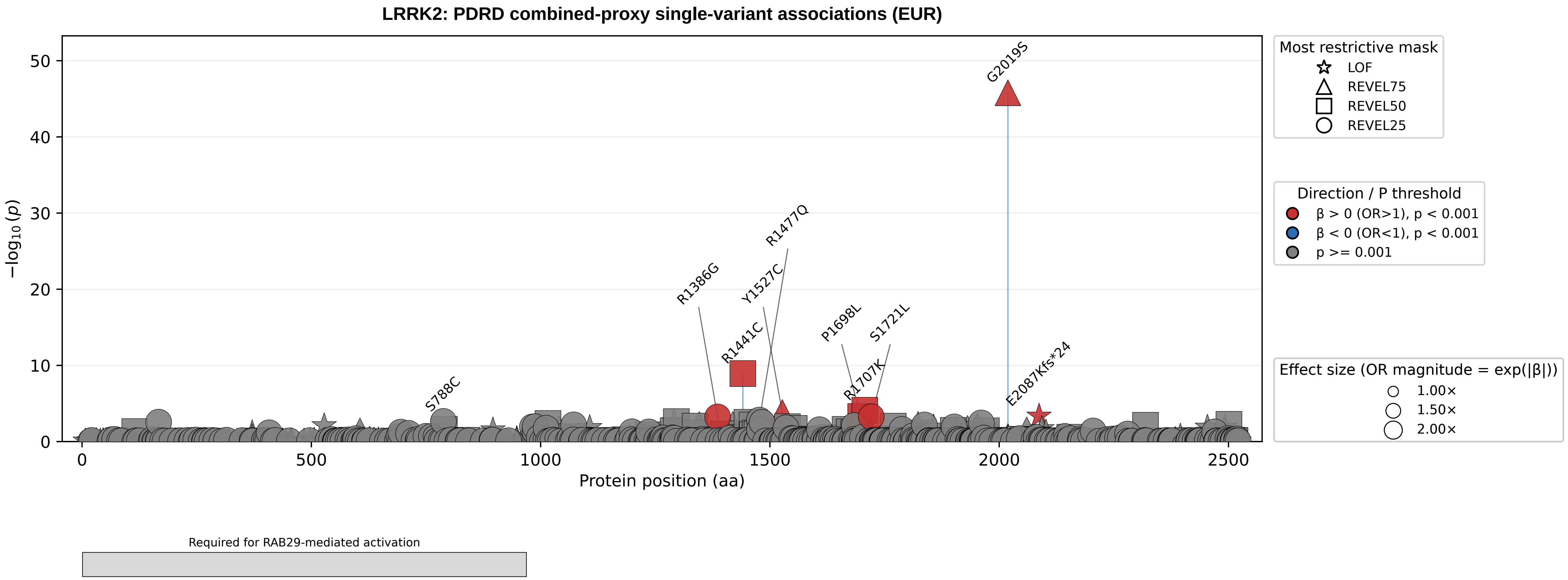

LRRK2: PDRD combined-proxy single-variant associations (EUR)
−log10(p)
Protein position (aa)
G2019S
R1477Q
R1386G
Y1527C
R1441C
P1698L
S1721L
R1707K
E2087Kfs*24
S788C
Most restrictive mask
LOF
REVEL75
REVEL50
REVEL25
Direction / P threshold
β > 0 (OR>1), p < 0.001
β < 0 (OR<1), p < 0.001
p >= 0.001
Effect size (OR magnitude = exp(|β|))
1.00×
1.50×
2.00×
Required for RAB29-mediated activation

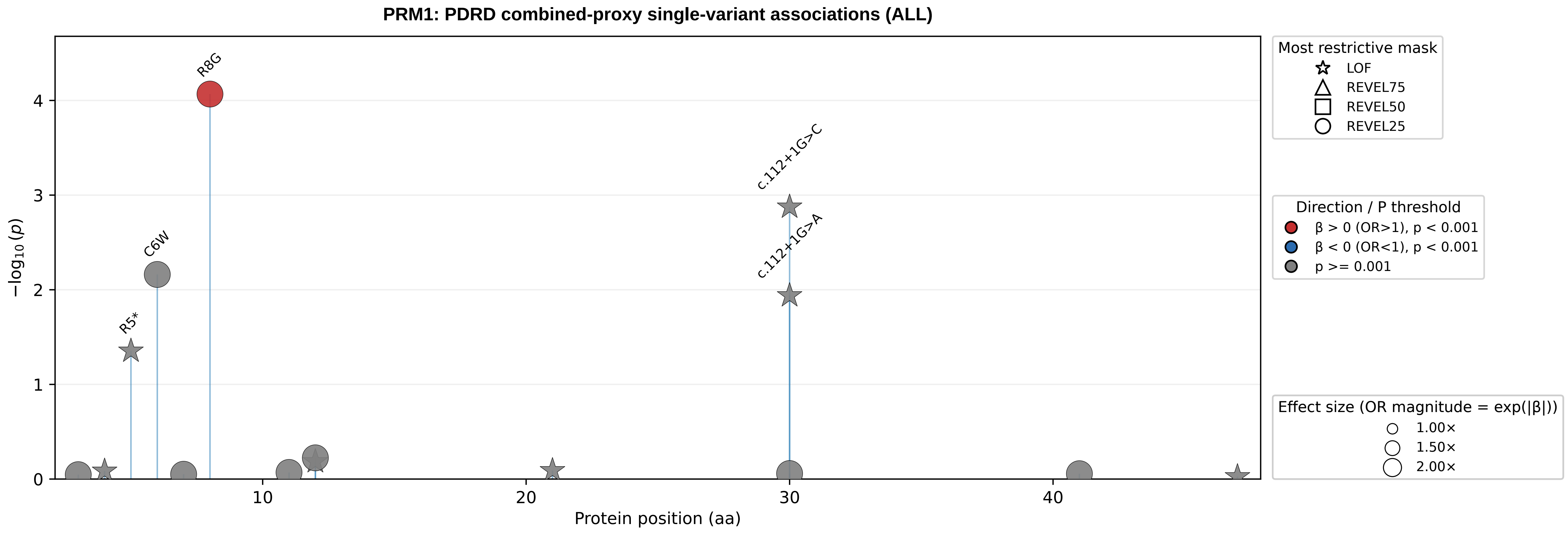

PRM1: PDRD combined-proxy single-variant associations (ALL)
R8G
C6W
R5*
c.112+1G>C
c.112+1G>A
−log10(p)
Protein position (aa)
0
1
2
3
4
10
20
30
40
Most restrictive mask
LOF
REVEL75
REVEL50
REVEL25
Direction / P threshold
β > 0 (OR>1), p < 0.001
β < 0 (OR<1), p < 0.001
p >= 0.001
Effect size (OR magnitude = exp(|β|))
1.00×
1.50×
2.00×

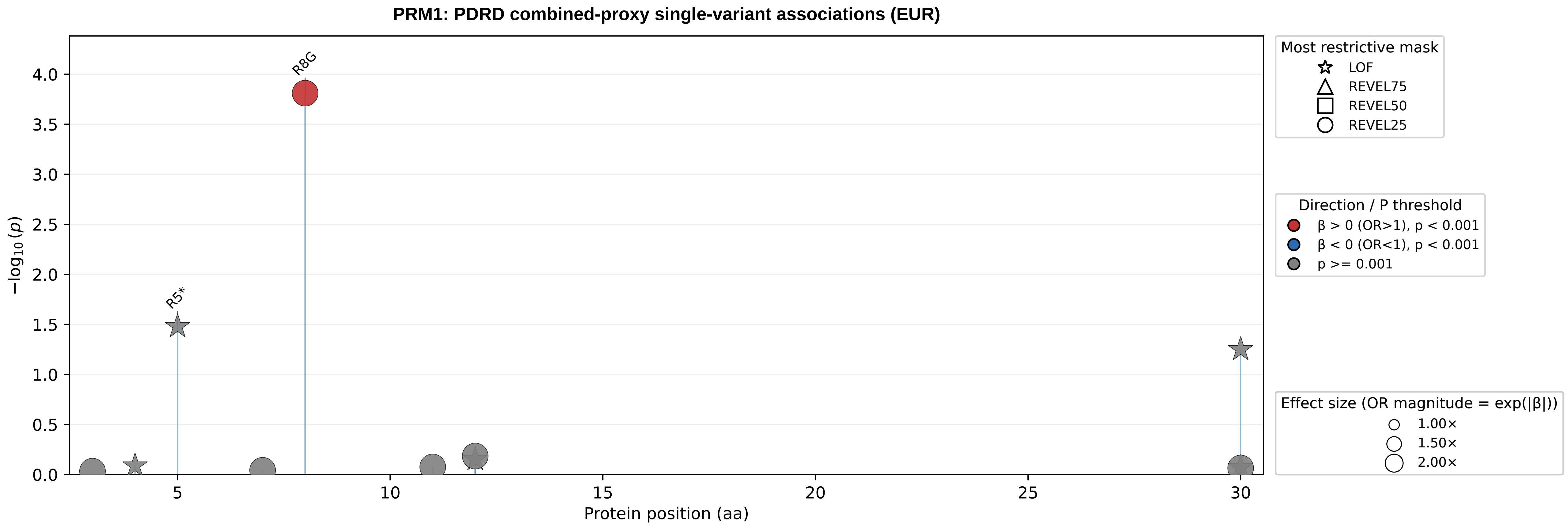

PRM1: PDRD combined-proxy single-variant associations (EUR)
R8G
R5*
−log10(p)
4.0
3.5
3.0
2.5
2.0
1.5
1.0
0.5
0.0
5
10
15
20
25
30
Protein position (aa)
Most restrictive mask
LOF
REVEL75
REVEL50
REVEL25
Direction / P threshold
β > 0 (OR>1), p < 0.001
β < 0 (OR<1), p < 0.001
p >= 0.001
Effect size (OR magnitude = exp(|β|))
1.00×
1.50×
2.00×

**CCL7: PDRD combined-proxy single-variant associations (ALL)**

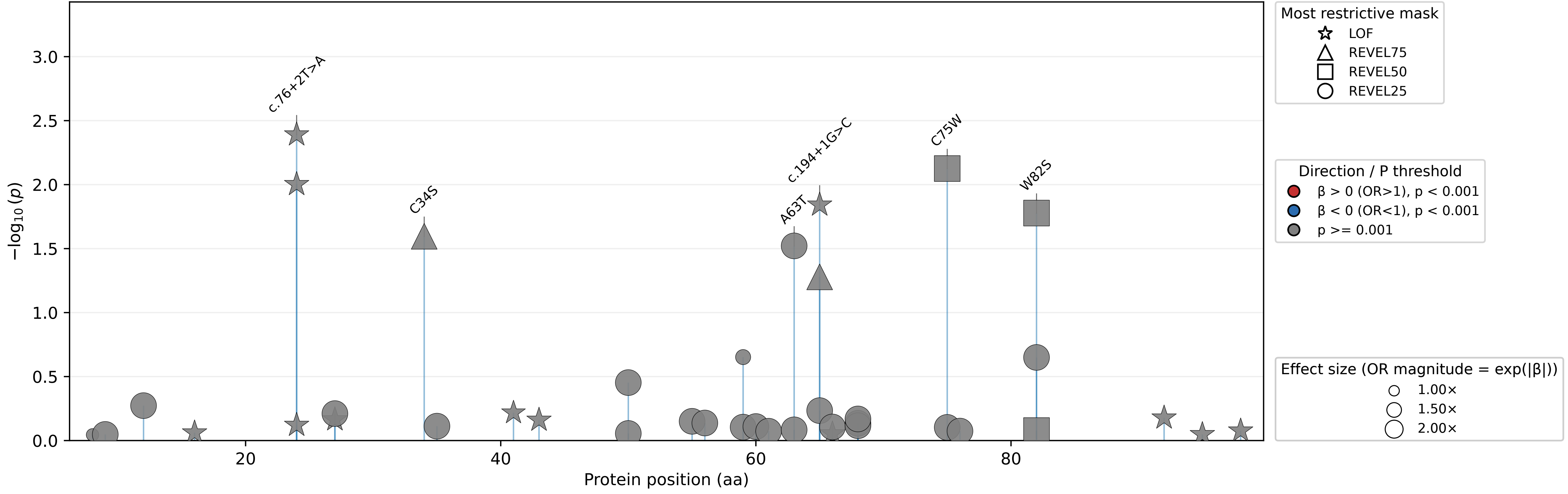

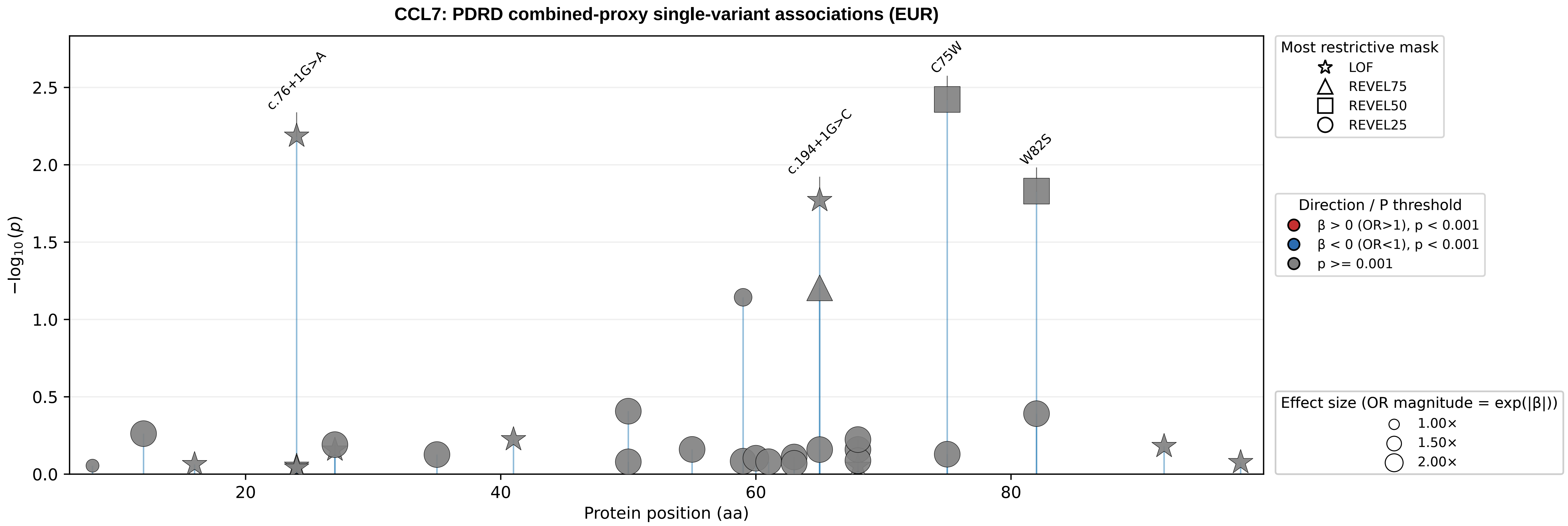
CCL7: PDRD combined-proxy single-variant associations (EUR)
−log10(p)
Protein position (aa)
0.0
0.5
1.0
1.5
2.0
2.5
20
40
60
80
c.76+1G>A
c.194+1G>C
C75W
W82S
Most restrictive mask
LOF
REVEL75
REVEL50
REVEL25
Direction / P threshold
β > 0 (OR>1), p < 0.001
β < 0 (OR<1), p < 0.001
p >= 0.001
Effect size (OR magnitude = exp(|β|))
1.00×
1.50×
2.00×

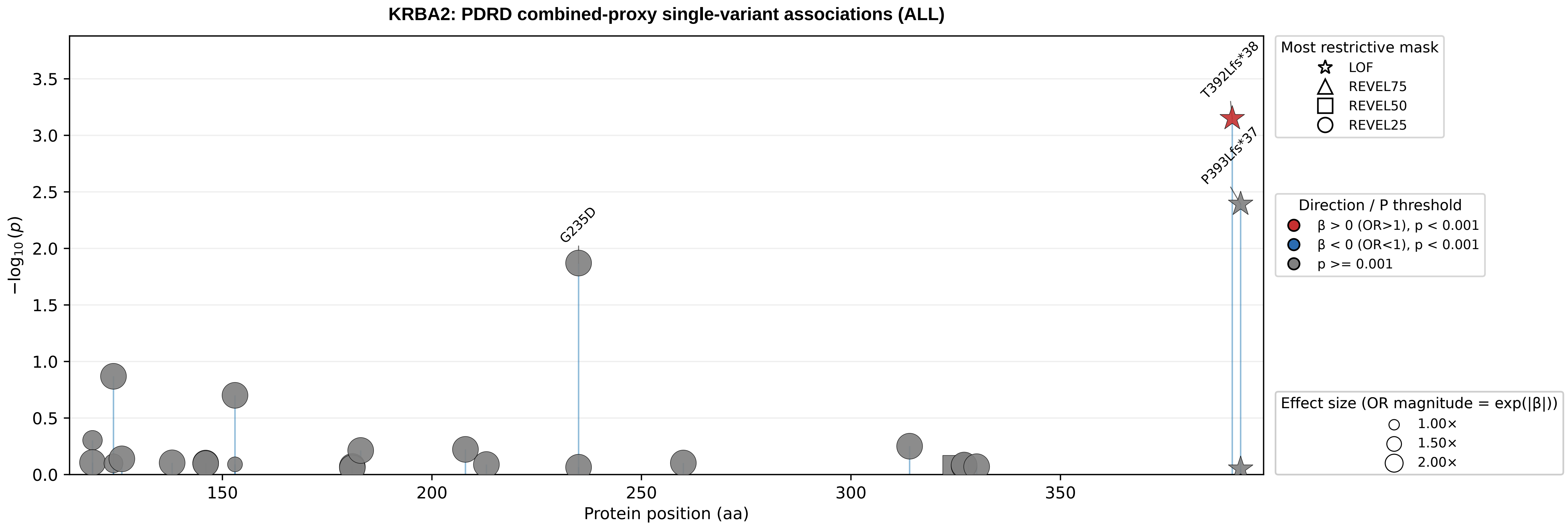
KRBA2: PDRD combined-proxy single-variant associations (ALL)
T392Lfs*38
P393Lfs*37
G235D
−log10(p)
Protein position (aa)
0.0
0.5
1.0
1.5
2.0
2.5
3.0
3.5
150
200
250
300
350
Most restrictive mask
LOF
REVEL75
REVEL50
REVEL25
Direction / P threshold
β > 0 (OR>1), p < 0.001
β < 0 (OR<1), p < 0.001
p >= 0.001
Effect size (OR magnitude = exp(|β|))
1.00×
1.50×
2.00×

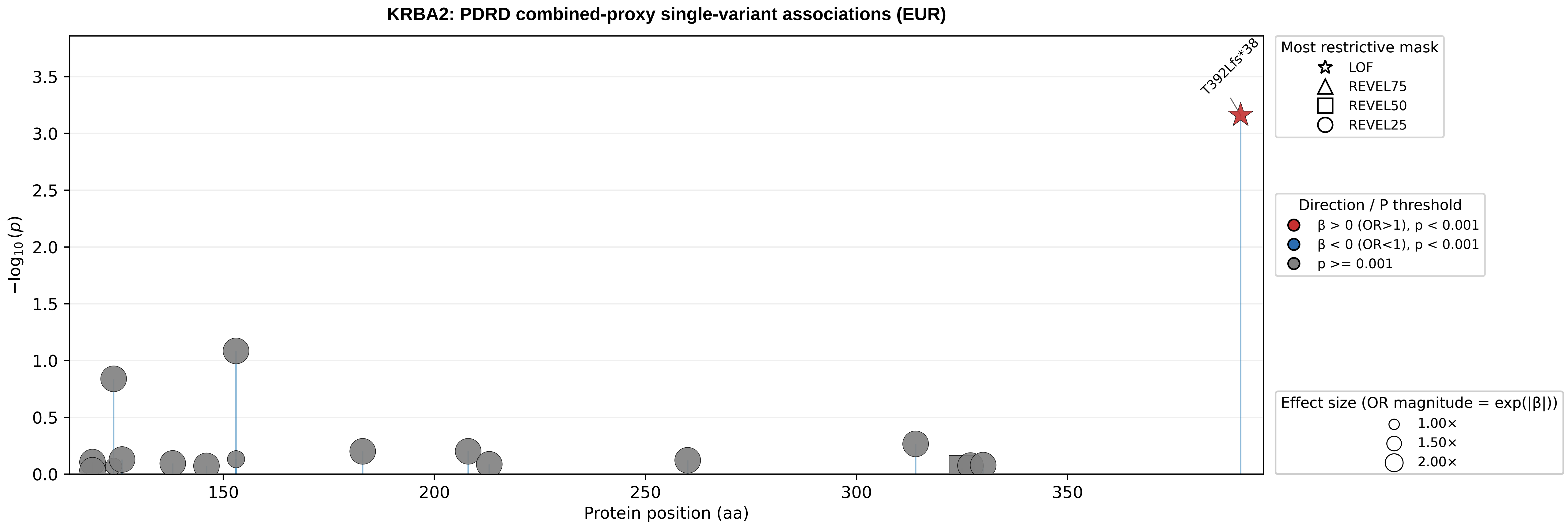
KRBA2: PDRD combined-proxy single-variant associations (EUR)
T392Lfs*38
−log10(p)
Protein position (aa)
0.0
0.5
1.0
1.5
2.0
2.5
3.0
3.5
150
200
250
300
350
Most restrictive mask
LOF
REVEL75
REVEL50
REVEL25
Direction / P threshold
β > 0 (OR>1), p < 0.001
β < 0 (OR<1), p < 0.001
p >= 0.001
Effect size (OR magnitude = exp(|β|))
1.00×
1.50×
2.00×

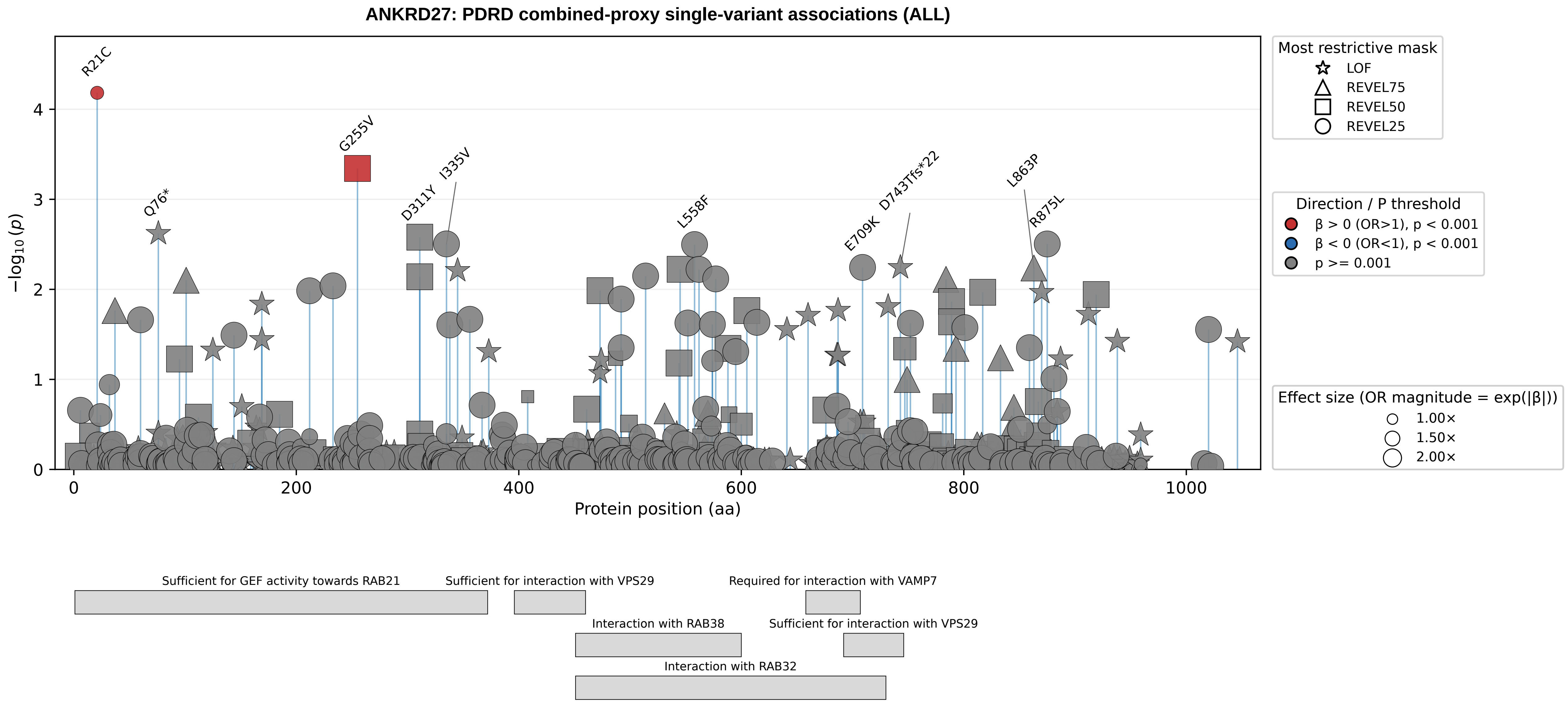

ANKRD27: PDRD combined-proxy single-variant associations (ALL)
R21C
G255V
D311Y
I335V
Q76*
L558F
E709K
D743Tfs*22
L863P
R875L
$-\log_{10}(p)$
Protein position (aa)
0
1
2
3
4
0
200
400
600
800
1000
Most restrictive mask
LOF
REVEL75
REVEL50
REVEL25
Direction / P threshold
β > 0 (OR>1), p < 0.001
β < 0 (OR<1), p < 0.001
p >= 0.001
Effect size (OR magnitude = exp(|β|))
1.00×
1.50×
2.00×
Sufficient for GEF activity towards RAB21
Sufficient for interaction with VPS29
Required for interaction with VAMP7
Interaction with RAB38
Sufficient for interaction with VPS29
Interaction with RAB32

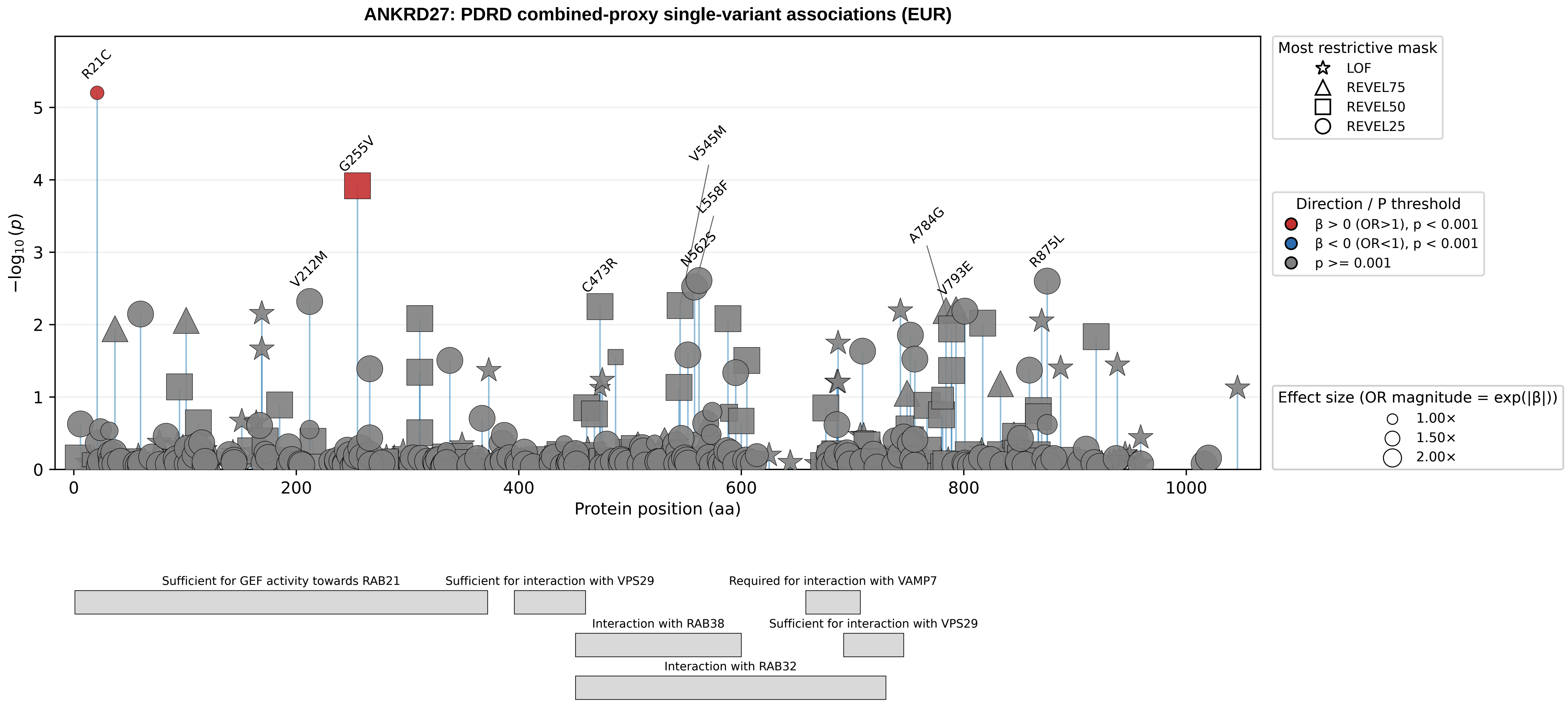
ANKRD27: PDRD combined-proxy single-variant associations (EUR)
R21C
G255V
V545M
L558F
N562S
V212M
C473R
A784G
V793E
R875L
−log10 (p)
Protein position (aa)
Most restrictive mask
LOF
REVEL75
REVEL50
REVEL25
Direction / P threshold
β > 0 (OR>1), p < 0.001
β < 0 (OR<1), p < 0.001
p >= 0.001
Effect size (OR magnitude = exp(|β|))
1.00×
1.50×
2.00×
Sufficient for GEF activity towards RAB21
Sufficient for interaction with VPS29
Required for interaction with VAMP7
Interaction with RAB38
Sufficient for interaction with VPS29
Interaction with RAB32
