## Supplementary_Fig5 for "Population-scale burden analysis of rare damaging coding variants identifies novel risk genes for Alzheimer’s disease and related dementias and Parkinson’s disease and related disorders"

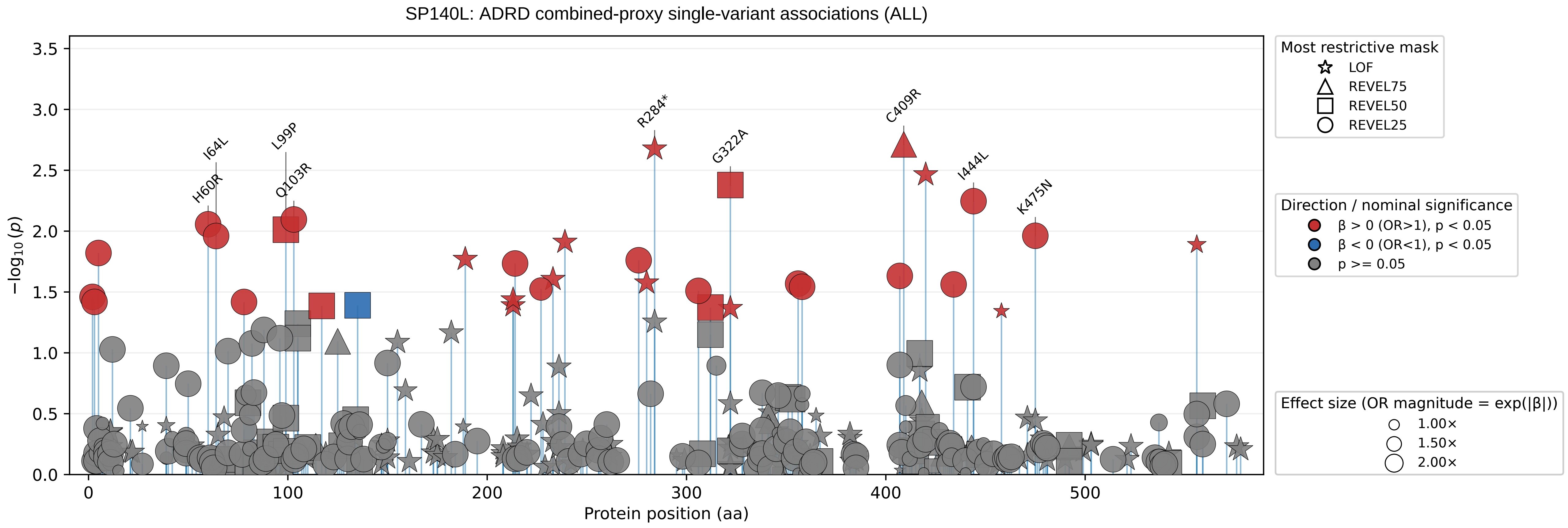
SP140L: ADRD combined-proxy single-variant associations (ALL)
−log10 (p)
Protein position (aa)
3.5
3.0
2.5
2.0
1.5
1.0
0.5
0.0
0
100
200
300
400
500
I64L
H60R
L99P
Q103R
R284*
G322A
C409R
I444L
K475N
Most restrictive mask
LOF
REVEL75
REVEL50
REVEL25
Direction / nominal significance
β > 0 (OR>1), p < 0.05
β < 0 (OR<1), p < 0.05
p >= 0.05
Effect size (OR magnitude = exp(|β|))
1.00×
1.50×
2.00×

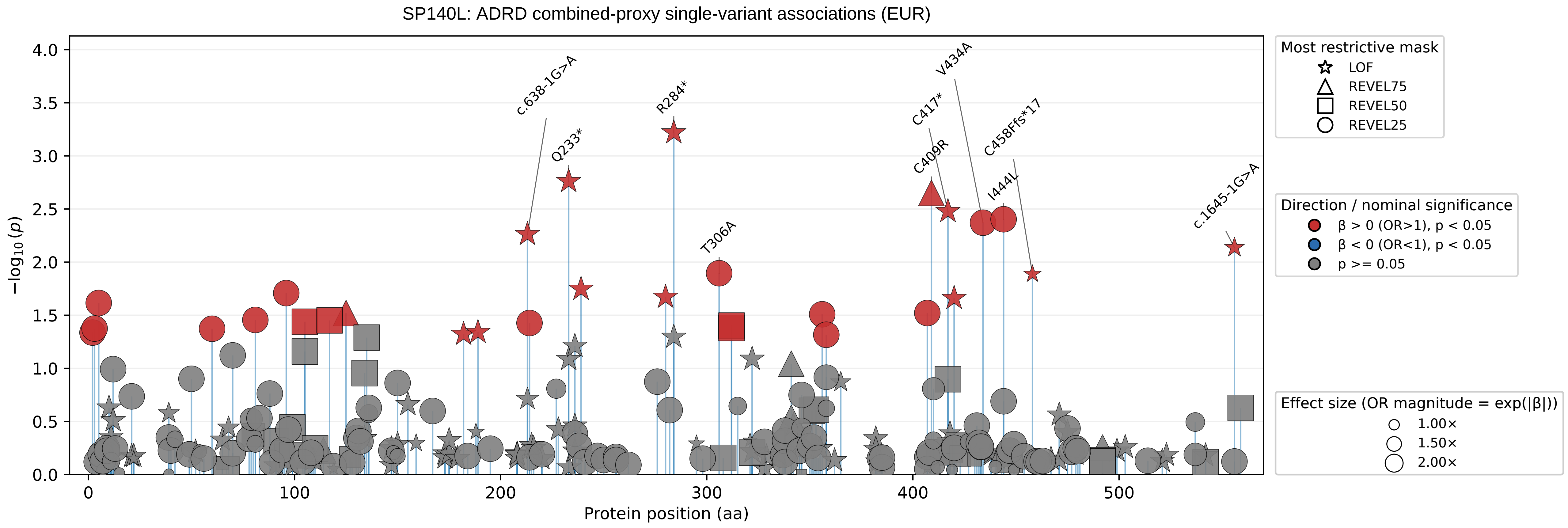

SP140L: ADRD combined-proxy single-variant associations (EUR)
−log10 (p)
Protein position (aa)
c.638-1G>A
Q233*
R284*
T306A
C417*
C409R
V434A
C458Ffs*17
I444L
c.1645-1G>A
Most restrictive mask
LOF
REVEL75
REVEL50
REVEL25
Direction / nominal significance
β > 0 (OR>1), p < 0.05
β < 0 (OR<1), p < 0.05
p >= 0.05
Effect size (OR magnitude = exp(|β|))
1.00×
1.50×
2.00×

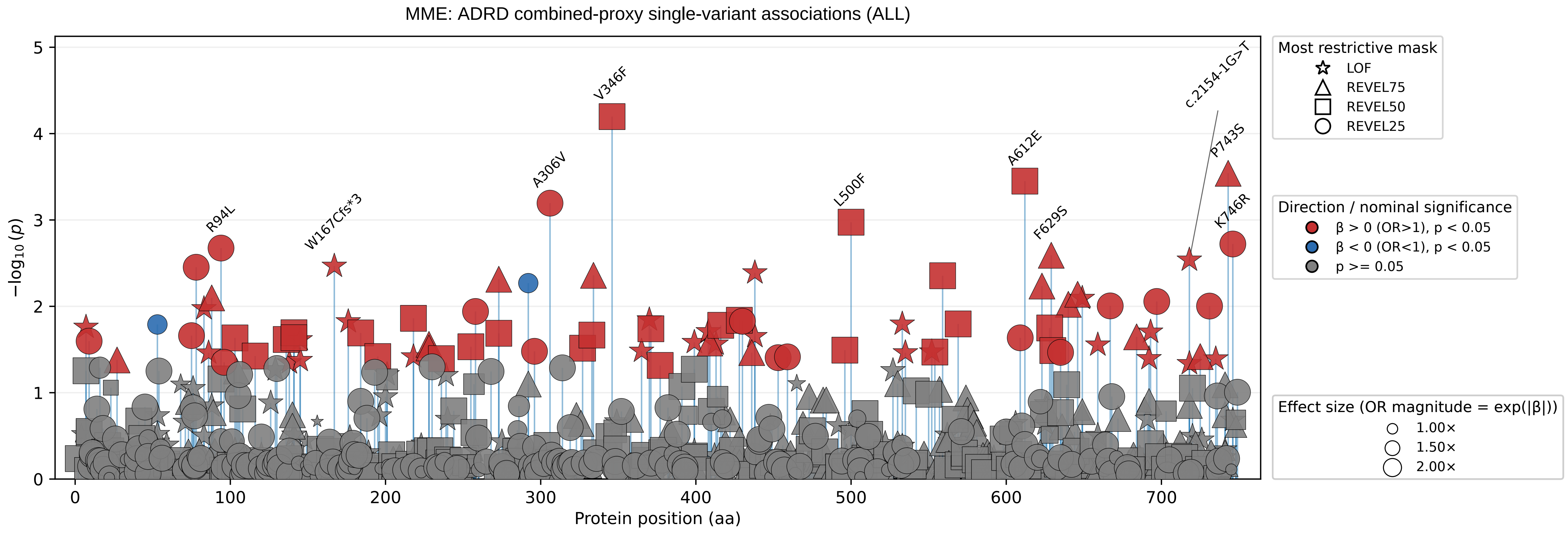

MME: ADRD combined-proxy single-variant associations (ALL)
−log10(p)
Protein position (aa)
V346F
c.2154-1G>T
A612E
P743S
A306V
L500F
K746R
R94L
W167Cfs*3
F629S
Most restrictive mask
LOF
REVEL75
REVEL50
REVEL25
Direction / nominal significance
β > 0 (OR>1), p < 0.05
β < 0 (OR<1), p < 0.05
p >= 0.05
Effect size (OR magnitude = exp(|β|))
1.00×
1.50×
2.00×

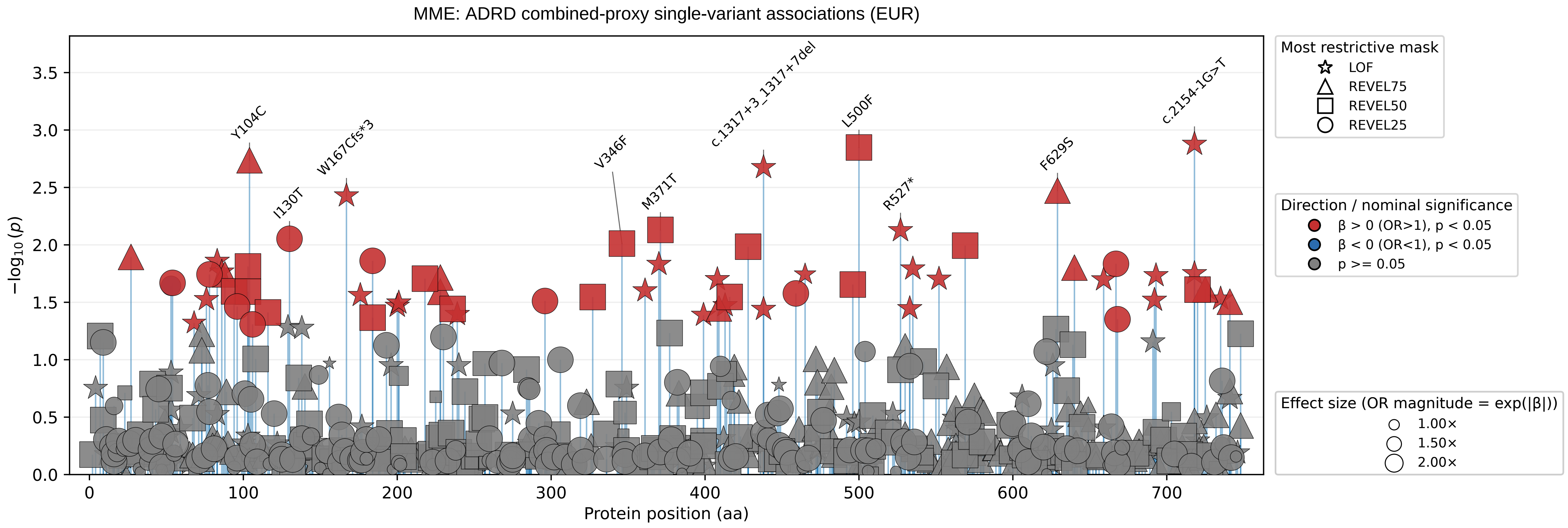

MME: ADRD combined-proxy single-variant associations (EUR)
−log10(p)
Protein position (aa)
Y104C
I130T
W167Cfs*3
V346F
M371T
c.1317+3_1317+7del
L500F
R527*
F629S
c.2154-1G>T
Most restrictive mask
LOF
REVEL75
REVEL50
REVEL25
Direction / nominal significance
β > 0 (OR>1), p < 0.05
β < 0 (OR<1), p < 0.05
p >= 0.05
Effect size (OR magnitude = exp(|β|))
1.00×
1.50×
2.00×

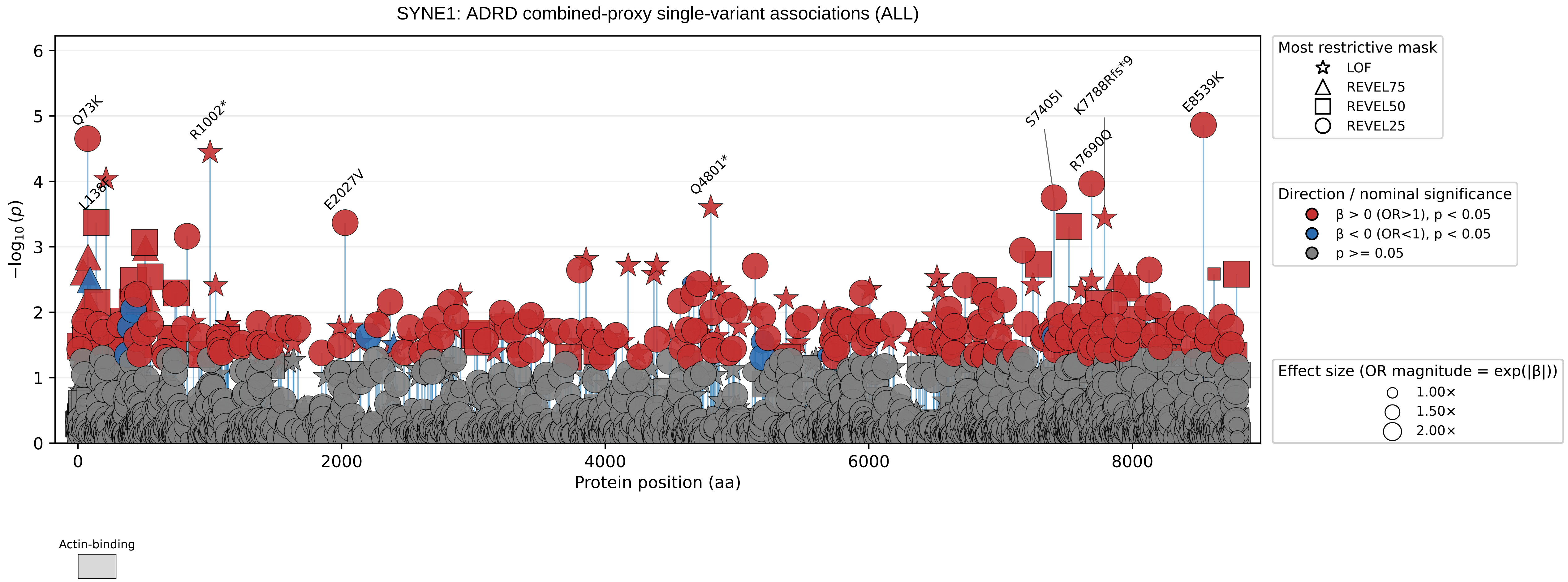
SYNE1: ADRD combined-proxy single-variant associations (ALL)
−log10(p)
Protein position (aa)
0
1
2
3
4
5
6
0
2000
4000
6000
8000
Q73K
L138F
R1002*
E2027V
Q4801*
S7405I
K7788Rfs*9
R7690Q
E8539K
Actin-binding
Most restrictive mask
LOF
REVEL75
REVEL50
REVEL25
Direction / nominal significance
β > 0 (OR>1), p < 0.05
β < 0 (OR<1), p < 0.05
p >= 0.05
Effect size (OR magnitude = exp(|β|))
1.00×
1.50×
2.00×

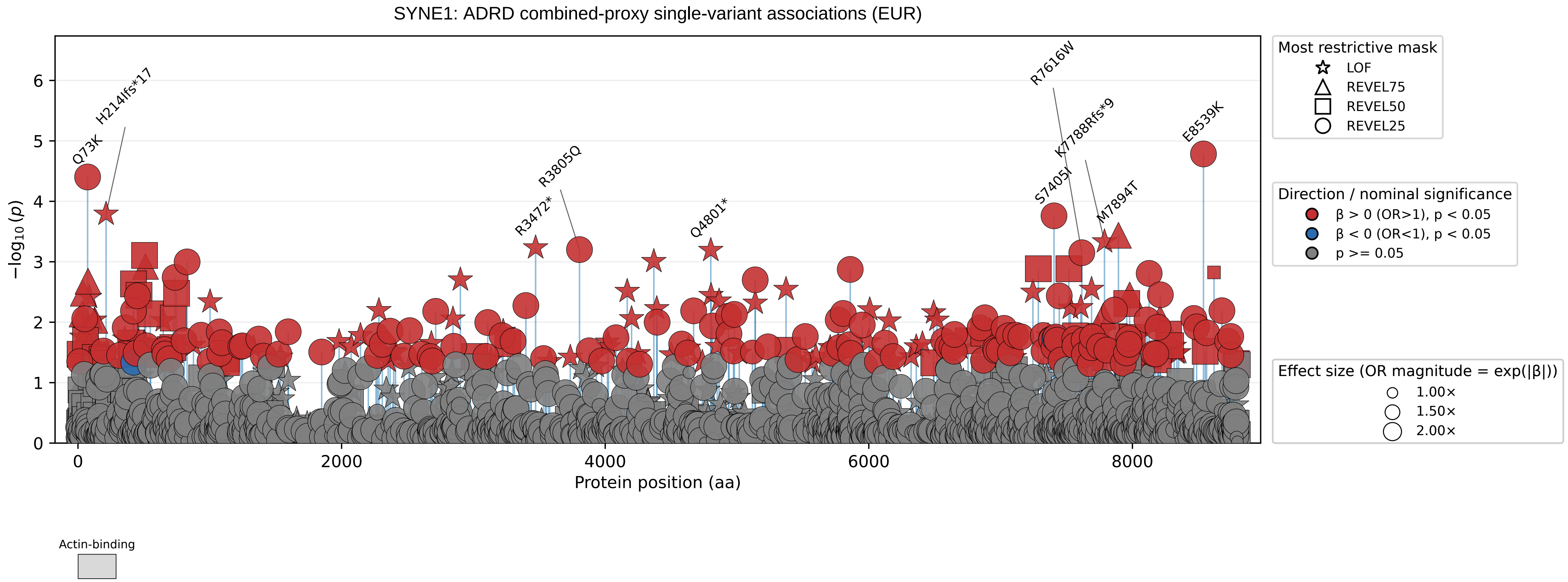
SYNE1: ADRD combined-proxy single-variant associations (EUR)
$-\log_{10}(p)$
Protein position (aa)
0
1
2
3
4
5
6
0
2000
4000
6000
8000
Q73K
H214Ifs*17
R3472*
R3805Q
Q4801*
S7405I
R7616W
K7788Rfs*9
M7894T
E8539K
Most restrictive mask
LOF
REVEL75
REVEL50
REVEL25
Direction / nominal significance
β > 0 (OR>1), p < 0.05
β < 0 (OR<1), p < 0.05
p >= 0.05
Effect size (OR magnitude = exp(|β|))
1.00×
1.50×
2.00×
Actin-binding

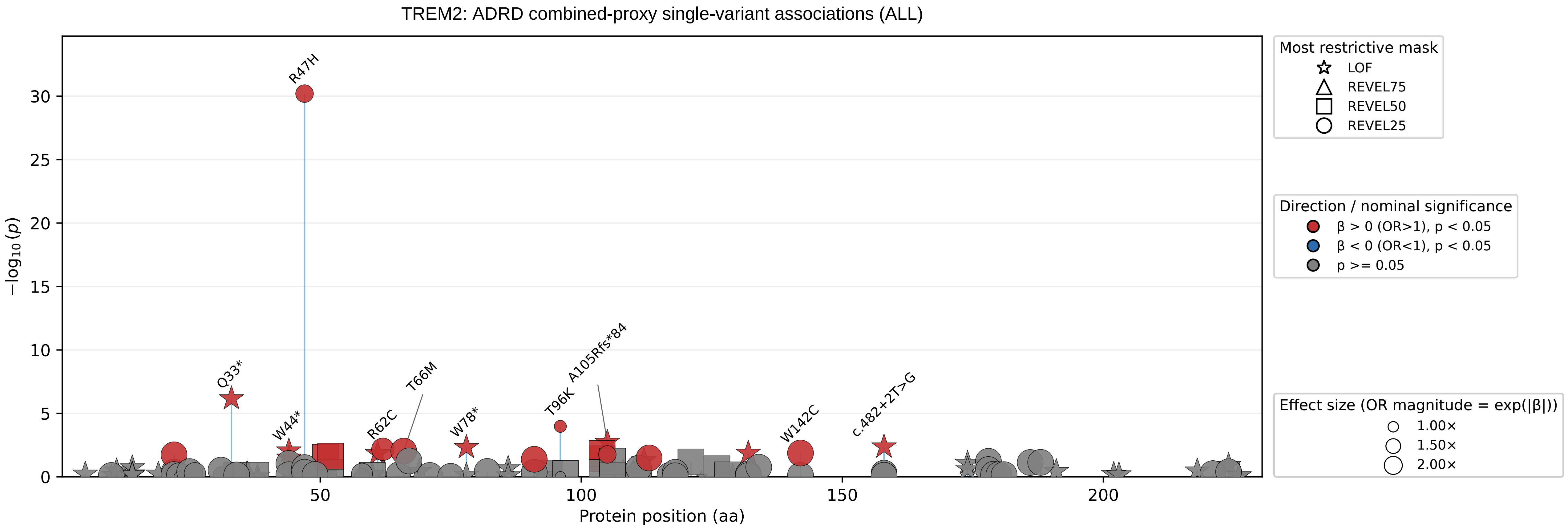

TREM2: ADRD combined-proxy single-variant associations (ALL)
−log10(p)
Protein position (aa)
0
5
10
15
20
25
30
50
100
150
200
R47H
Q33*
W44*
R62C
T66M
W78*
T96K
A105Rfs*84
W142C
c.482+2T>G
Most restrictive mask
LOF
REVEL75
REVEL50
REVEL25
Direction / nominal significance
β > 0 (OR>1), p < 0.05
β < 0 (OR<1), p < 0.05
p >= 0.05
Effect size (OR magnitude = exp(|β|))
1.00×
1.50×
2.00×

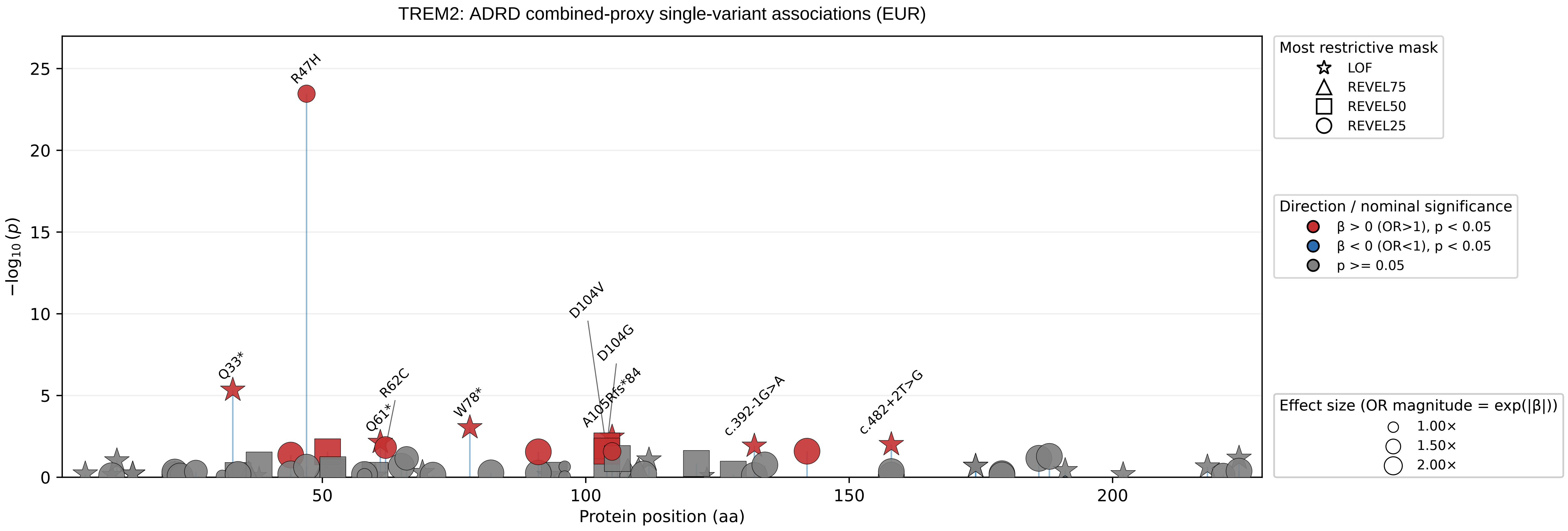
TREM2: ADRD combined-proxy single-variant associations (EUR)
R47H
Q33*
R62C
Q61*
W78*
D104V
D104G
A105Rfs*84
c.392-1G>A
c.482+2T>G
−log10(p)
Protein position (aa)
0
5
10
15
20
25
50
100
150
200
Most restrictive mask
LOF
REVEL75
REVEL50
REVEL25
Direction / nominal significance
β > 0 (OR>1), p < 0.05
β < 0 (OR<1), p < 0.05
p >= 0.05
Effect size (OR magnitude = exp(|β|))
1.00×
1.50×
2.00×

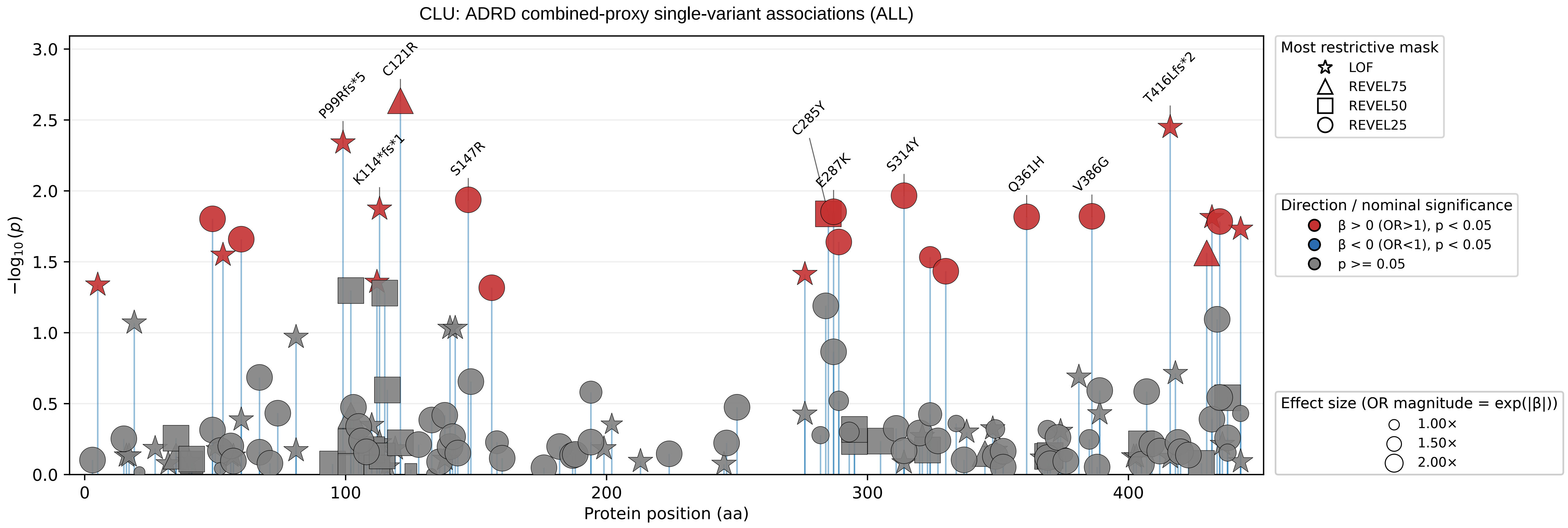

CLU: ADRD combined-proxy single-variant associations (ALL)
−log10(p)
Protein position (aa)
P99Rfs*5
C121R
K114*fs*1
S147R
C285Y
E287K
S314Y
Q361H
V386G
T416Lfs*2
Most restrictive mask
LOF
REVEL75
REVEL50
REVEL25
Direction / nominal significance
β > 0 (OR>1), p < 0.05
β < 0 (OR<1), p < 0.05
p >= 0.05
Effect size (OR magnitude = exp(|β|))
1.00×
1.50×
2.00×

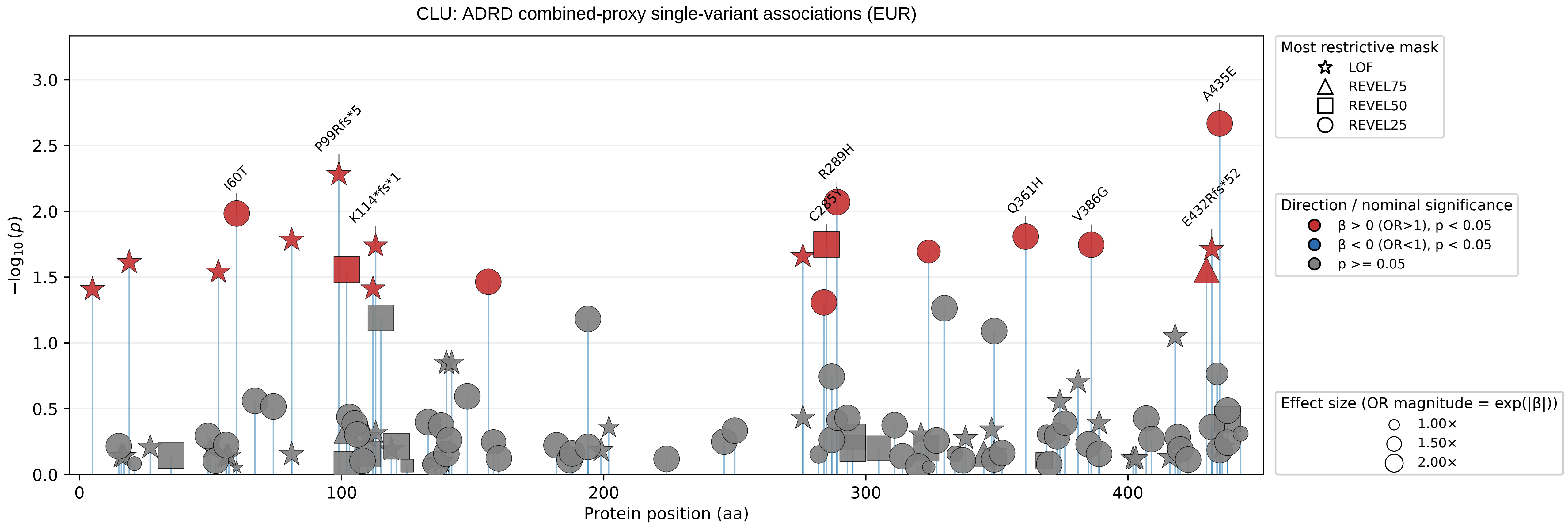

CLU: ADRD combined-proxy single-variant associations (EUR)
−log10(p)
Protein position (aa)
I60T
P99Rfs*5
K114*fs*1
R289H
C285Y
Q361H
V386G
A435E
E432Rfs*52
Most restrictive mask
LOF
REVEL75
REVEL50
REVEL25
Direction / nominal significance
β > 0 (OR>1), p < 0.05
β < 0 (OR<1), p < 0.05
p >= 0.05
Effect size (OR magnitude = exp(|β|))
1.00×
1.50×
2.00×

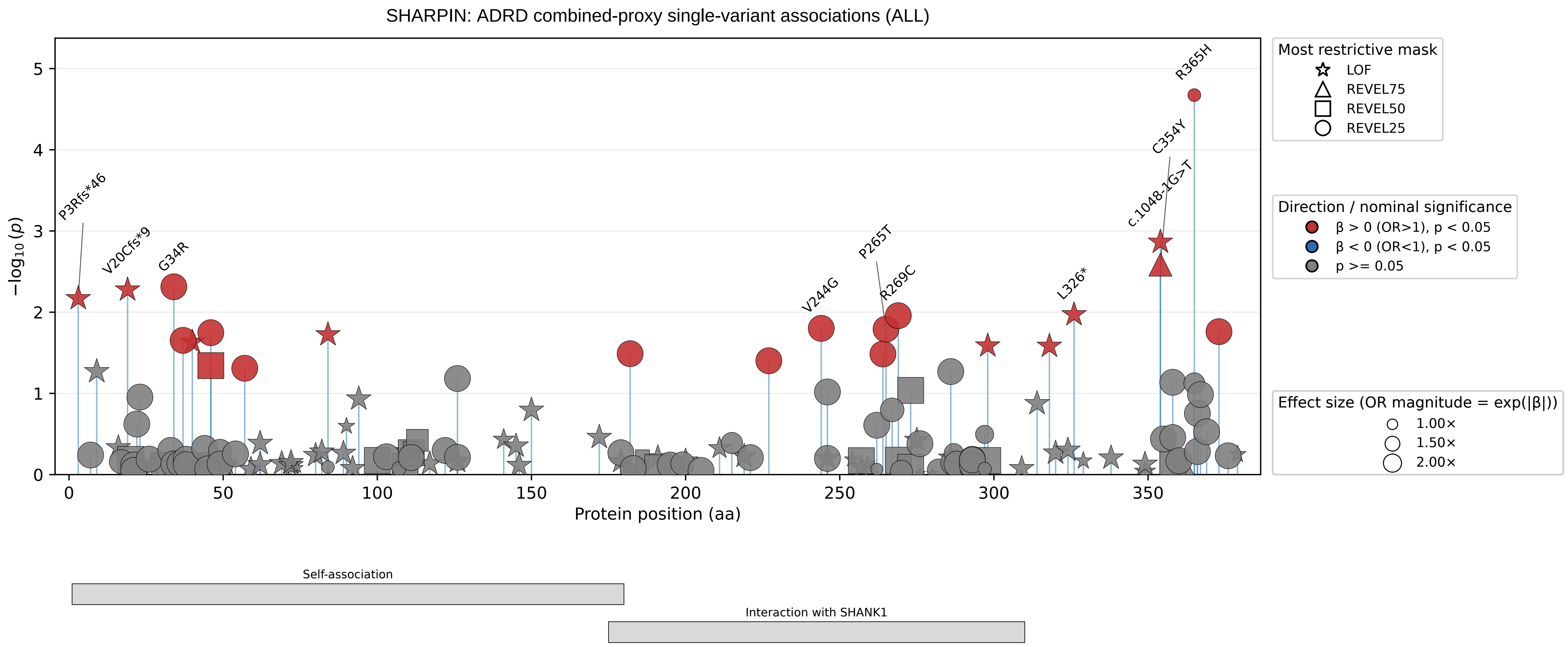

SHARPIN: ADRD combined-proxy single-variant associations (ALL)
−log₁₀(p)
Protein position (aa)
P3Rfs*46
V20Cfs*9
G34R
V244G
P265T
R269C
L326*
c.1048-1G>T
C354Y
R365H
Most restrictive mask
LOF
REVEL75
REVEL50
REVEL25
Direction / nominal significance
β > 0 (OR>1), p < 0.05
β < 0 (OR<1), p < 0.05
p >= 0.05
Effect size (OR magnitude = exp(|β|))
1.00×
1.50×
2.00×
Self-association
Interaction with SHANK1

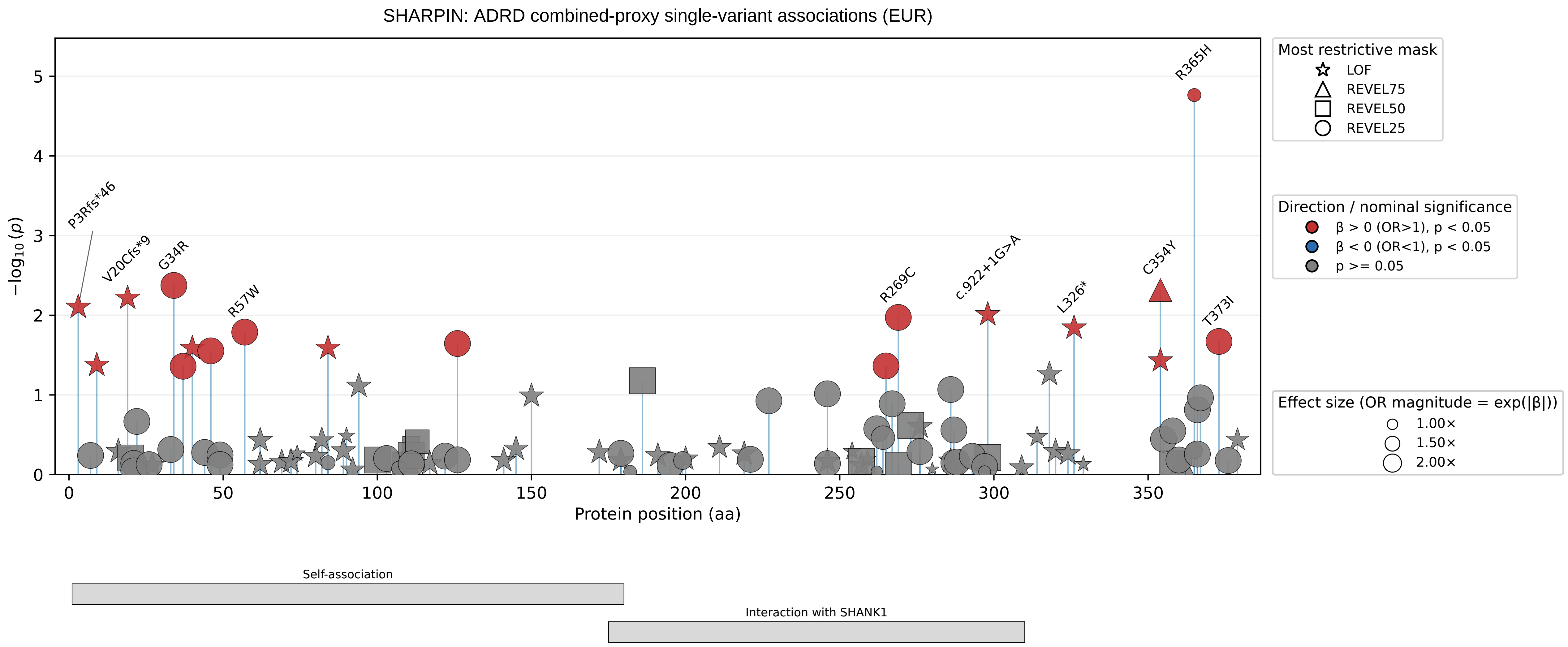

SHARPIN: ADRD combined-proxy single-variant associations (EUR)
−log10(p)
Protein position (aa)
P3Rfs*46
V20Cfs*9
G34R
R57W
R269C
c.922+1G>A
L326*
C354Y
R365H
T373I
Most restrictive mask
LOF
REVEL75
REVEL50
REVEL25
Direction / nominal significance
β > 0 (OR>1), p < 0.05
β < 0 (OR<1), p < 0.05
p >= 0.05
Effect size (OR magnitude = exp(|β|))
1.00×
1.50×
2.00×
Self-association
Interaction with SHANK1

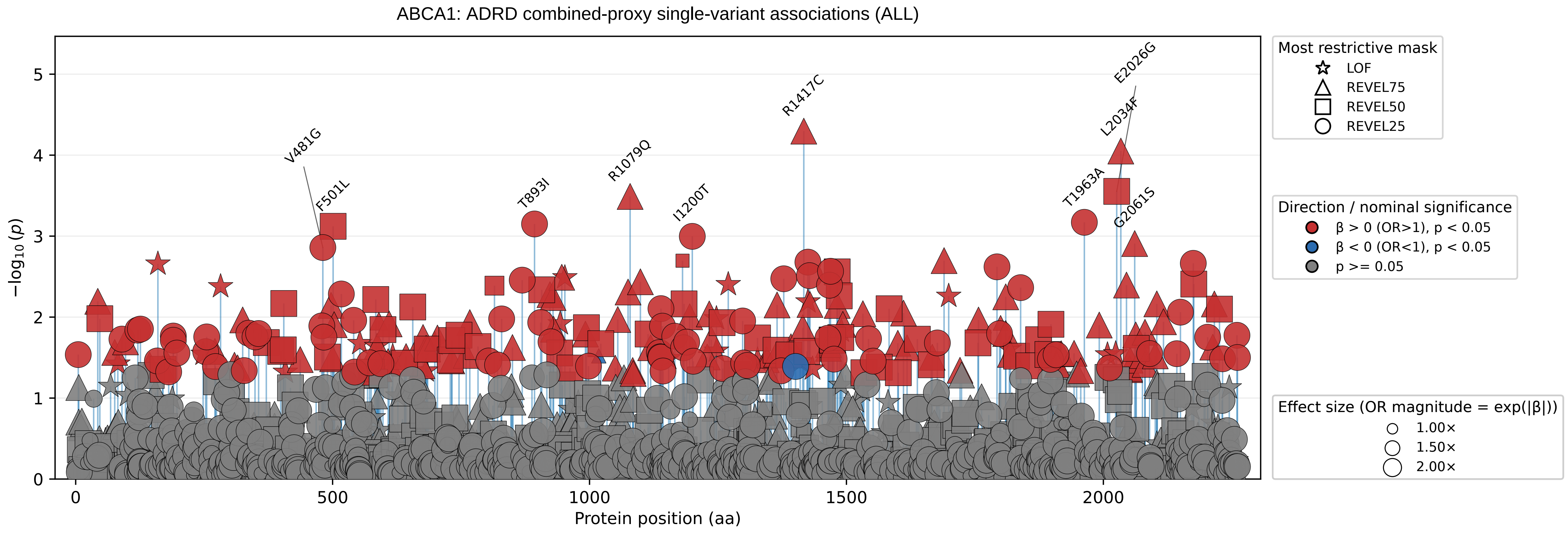

ABCA1: ADRD combined-proxy single-variant associations (ALL)
−log10(p)
Protein position (aa)
V481G
F501L
T893I
R1079Q
I1200T
R1417C
T1963A
L2034F
E2026G
G2061S
Most restrictive mask
LOF
REVEL75
REVEL50
REVEL25
Direction / nominal significance
β > 0 (OR>1), p < 0.05
β < 0 (OR<1), p < 0.05
p >= 0.05
Effect size (OR magnitude = exp(|β|))
1.00×
1.50×
2.00×

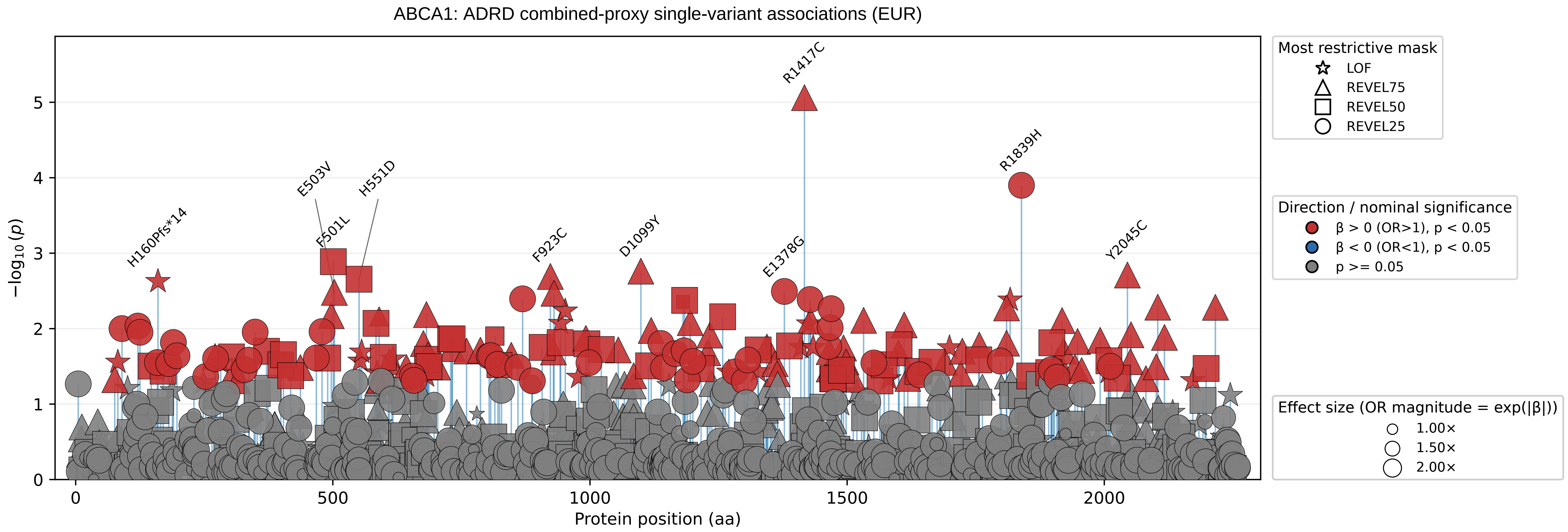

ABCA1: ADRD combined-proxy single-variant associations (EUR)
−log10 (p)
Protein position (aa)
0
1
2
3
4
5
0
500
1000
1500
2000
H160Pfs*14
E503V
F501L
H551D
F923C
D1099Y
R1417C
E1378G
R1839H
Y2045C
Most restrictive mask
LOF
REVEL75
REVEL50
REVEL25
Direction / nominal significance
β > 0 (OR>1), p < 0.05
β < 0 (OR<1), p < 0.05
p >= 0.05
Effect size (OR magnitude = exp(|β|))
1.00×
1.50×
2.00×

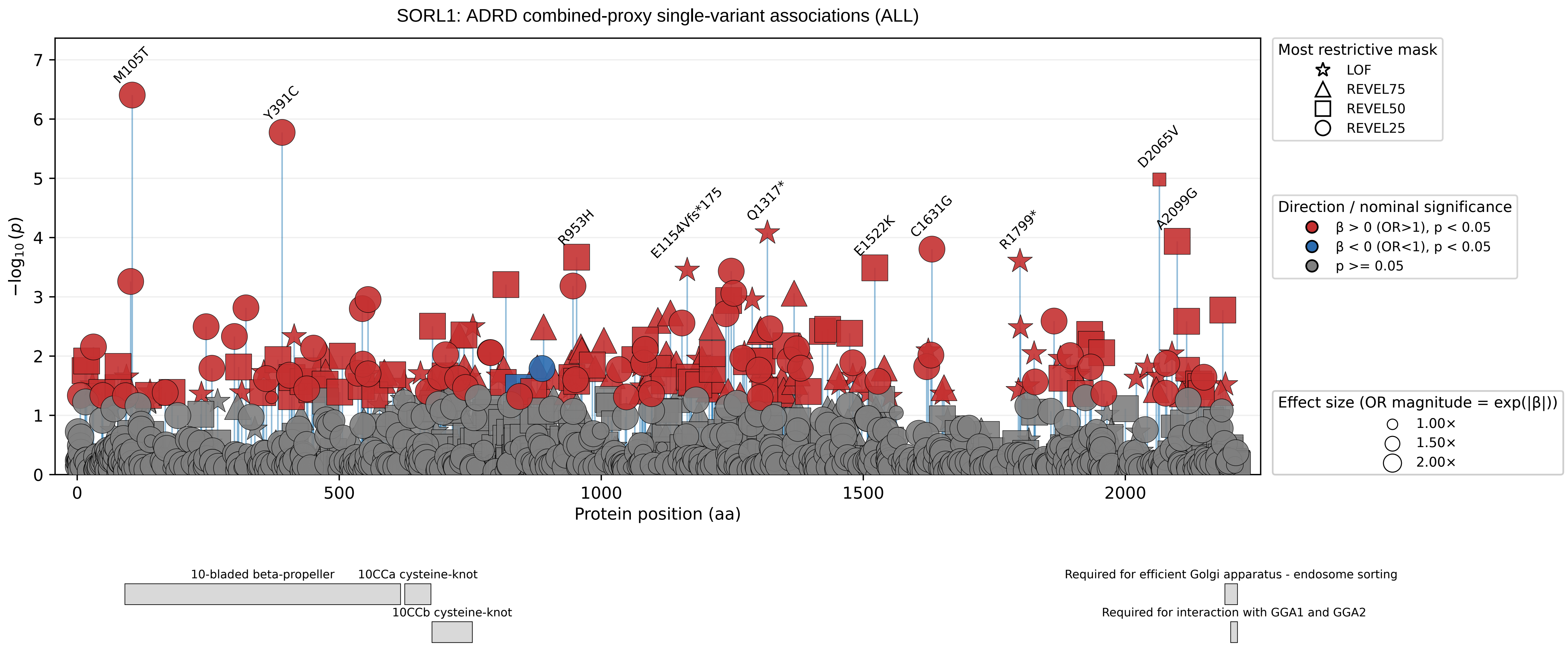

SORL1: ADRD combined-proxy single-variant associations (ALL)
−log10(p)
Protein position (aa)
M105T
Y391C
R953H
E1154Vfs*175
Q1317*
E1522K
C1631G
R1799*
D2065V
A2099G
Most restrictive mask
LOF
REVEL75
REVEL50
REVEL25
Direction / nominal significance
β > 0 (OR>1), p < 0.05
β < 0 (OR<1), p < 0.05
p >= 0.05
Effect size (OR magnitude = exp(|β|))
1.00×
1.50×
2.00×
10-bladed beta-propeller
10CCa cysteine-knot
10CCb cysteine-knot
Required for efficient Golgi apparatus - endosome sorting
Required for interaction with GGA1 and GGA2

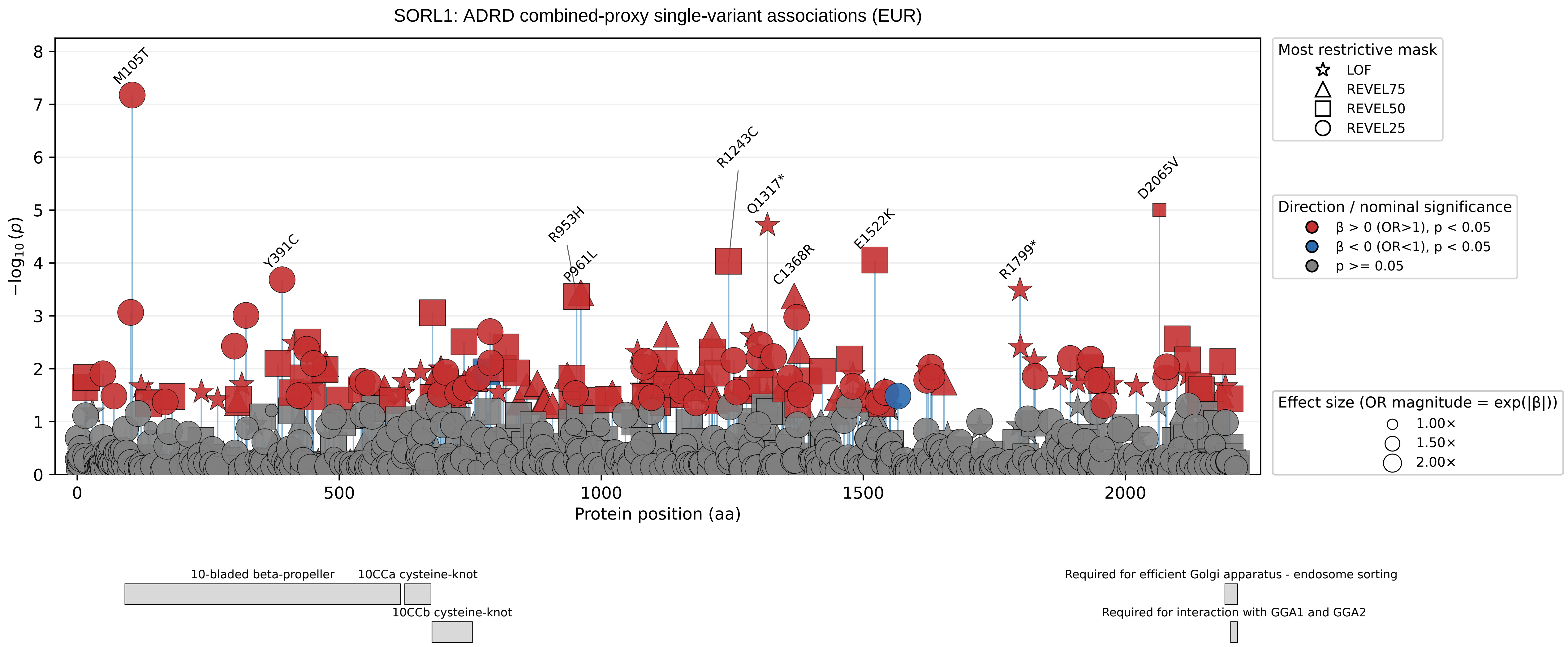

SORL1: ADRD combined-proxy single-variant associations (EUR)
−log10(p)
Protein position (aa)
0
500
1000
1500
2000
M105T
Y391C
R953H
P961L
R1243C
Q1317*
C1368R
E1522K
R1799*
D2065V
Most restrictive mask
LOF
REVEL75
REVEL50
REVEL25
Direction / nominal significance
β > 0 (OR>1), p < 0.05
β < 0 (OR<1), p < 0.05
p >= 0.05
Effect size (OR magnitude = exp(|β|))
1.00×
1.50×
2.00×
10-bladed beta-propeller
10CCa cysteine-knot
10CCb cysteine-knot
Required for efficient Golgi apparatus - endosome sorting
Required for interaction with GGA1 and GGA2

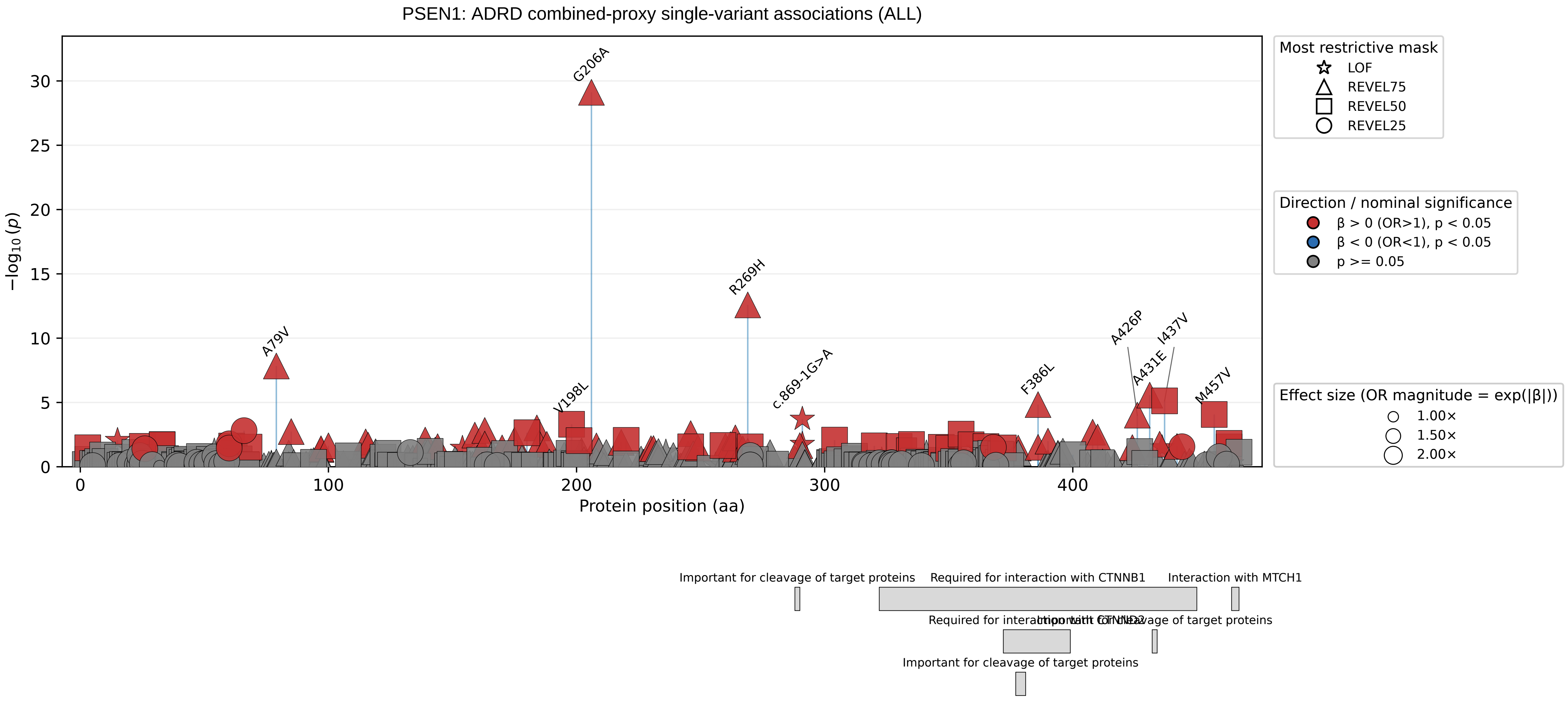
PSEN1: ADRD combined-proxy single-variant associations (ALL)
−log10 (p)
Protein position (aa)
0
5
10
15
20
25
30
0
100
200
300
400
G206A
R269H
A79V
V198L
c.869-1G>A
F386L
A426P
I437V
A431E
M457V
Most restrictive mask
LOF
REVEL75
REVEL50
REVEL25
Direction / nominal significance
β > 0 (OR>1), p < 0.05
β < 0 (OR<1), p < 0.05
p >= 0.05
Effect size (OR magnitude = exp(|β|))
1.00×
1.50×
2.00×
Important for cleavage of target proteins
Required for interaction with CTNNB1
Interaction with MTCH1
Important for cleavage of target proteins
Important for cleavage of target proteins

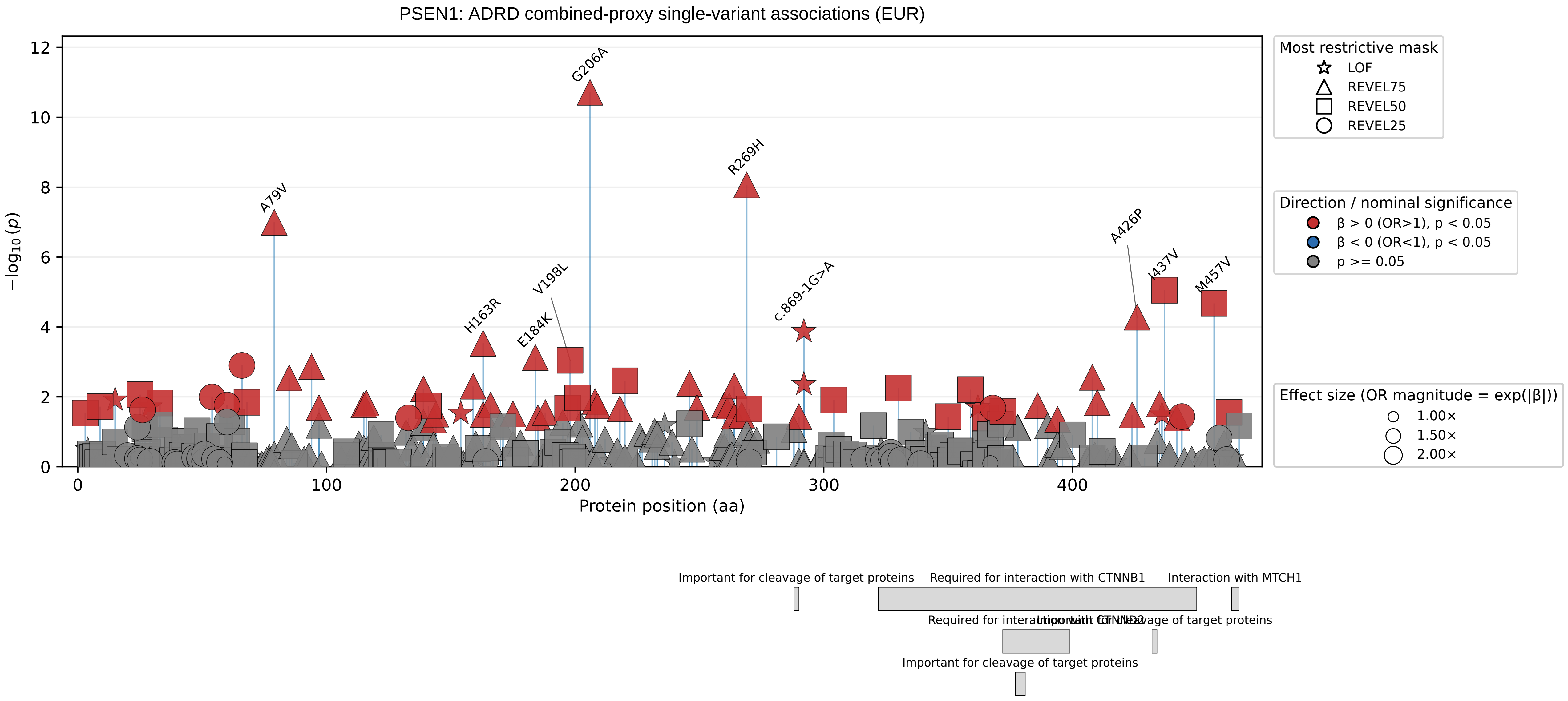
PSEN1: ADRD combined-proxy single-variant associations (EUR)
−log10 (p)
Protein position (aa)
0
2
4
6
8
10
12
0
100
200
300
400
G206A
R269H
A79V
H163R
E184K
V198L
c.869-1G>A
A426P
I437V
M457V
Most restrictive mask
LOF
REVEL75
REVEL50
REVEL25
Direction / nominal significance
β > 0 (OR>1), p < 0.05
β < 0 (OR<1), p < 0.05
p >= 0.05
Effect size (OR magnitude = exp(|β|))
1.00×
1.50×
2.00×
Important for cleavage of target proteins
Required for interaction with CTNNB1
Interaction with MTCH1
Required for interaction with CTNND2
Important for cleavage of target proteins
Important for cleavage of target proteins

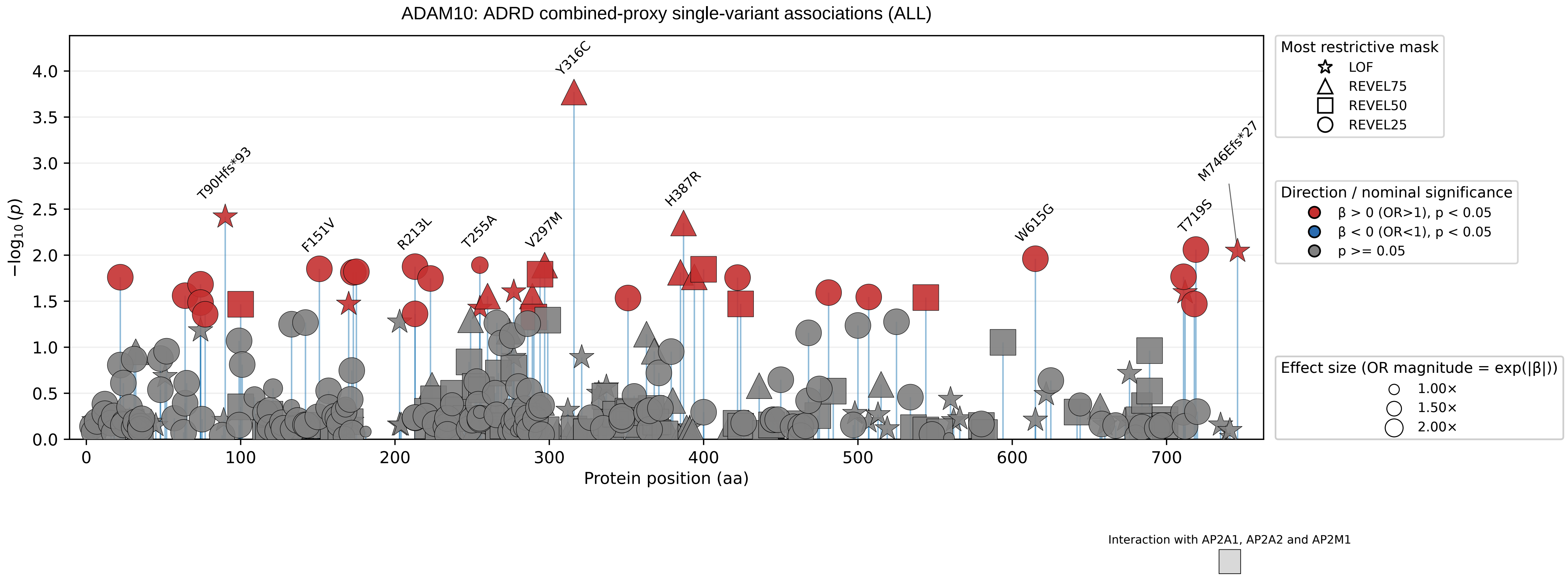
ADAM10: ADRD combined-proxy single-variant associations (ALL)
−log10 (p)
Protein position (aa)
0
100
200
300
400
500
600
700
0.0
0.5
1.0
1.5
2.0
2.5
3.0
3.5
4.0
Y316C
T90Hfs*93
F151V
R213L
T255A
V297M
H387R
W615G
T719S
M746Efs*27
Most restrictive mask
LOF
REVEL75
REVEL50
REVEL25
Direction / nominal significance
β > 0 (OR>1), p < 0.05
β < 0 (OR<1), p < 0.05
p >= 0.05
Effect size (OR magnitude = exp(|β|))
1.00×
1.50×
2.00×
Interaction with AP2A1, AP2A2 and AP2M1

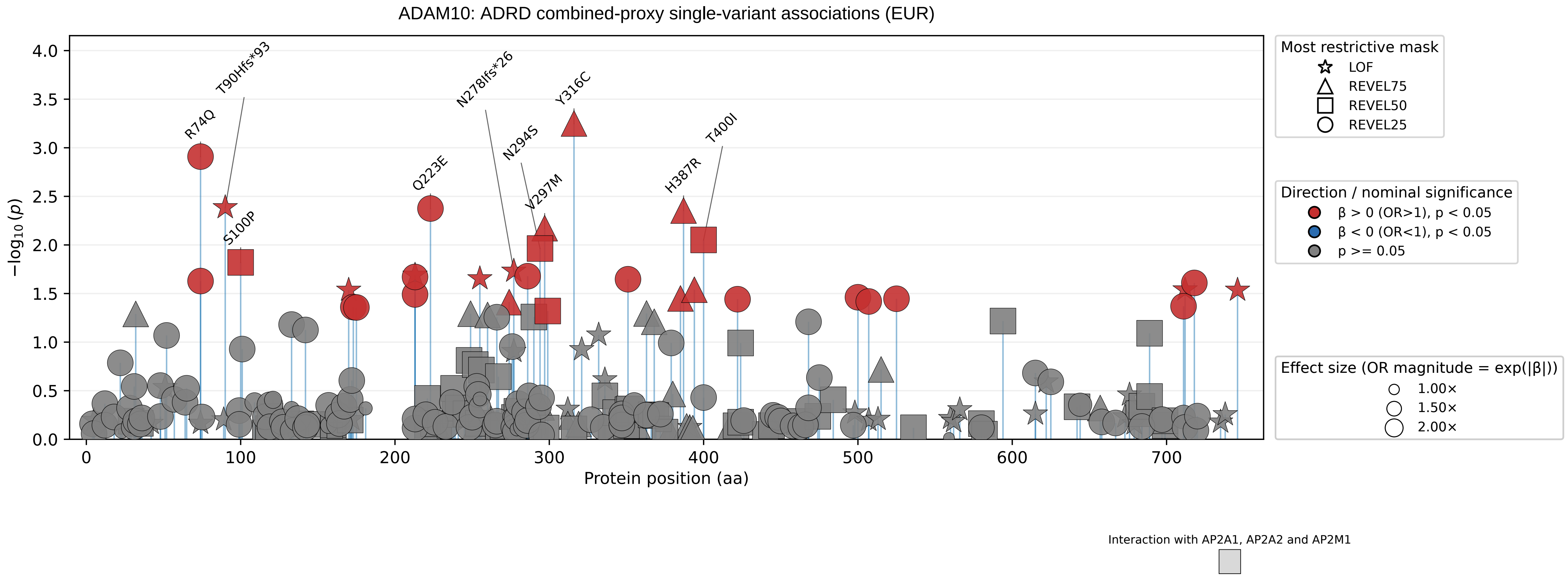

ADAM10: ADRD combined-proxy single-variant associations (EUR)
−log10 (p)
Protein position (aa)
R74Q
T90Hfs*93
S100P
Q223E
N278Ifs*26
N294S
V297M
Y316C
H387R
T400I
Most restrictive mask
LOF
REVEL75
REVEL50
REVEL25
Direction / nominal significance
β > 0 (OR>1), p < 0.05
β < 0 (OR<1), p < 0.05
p >= 0.05
Effect size (OR magnitude = exp(|β|))
1.00×
1.50×
2.00×
Interaction with AP2A1, AP2A2 and AP2M1

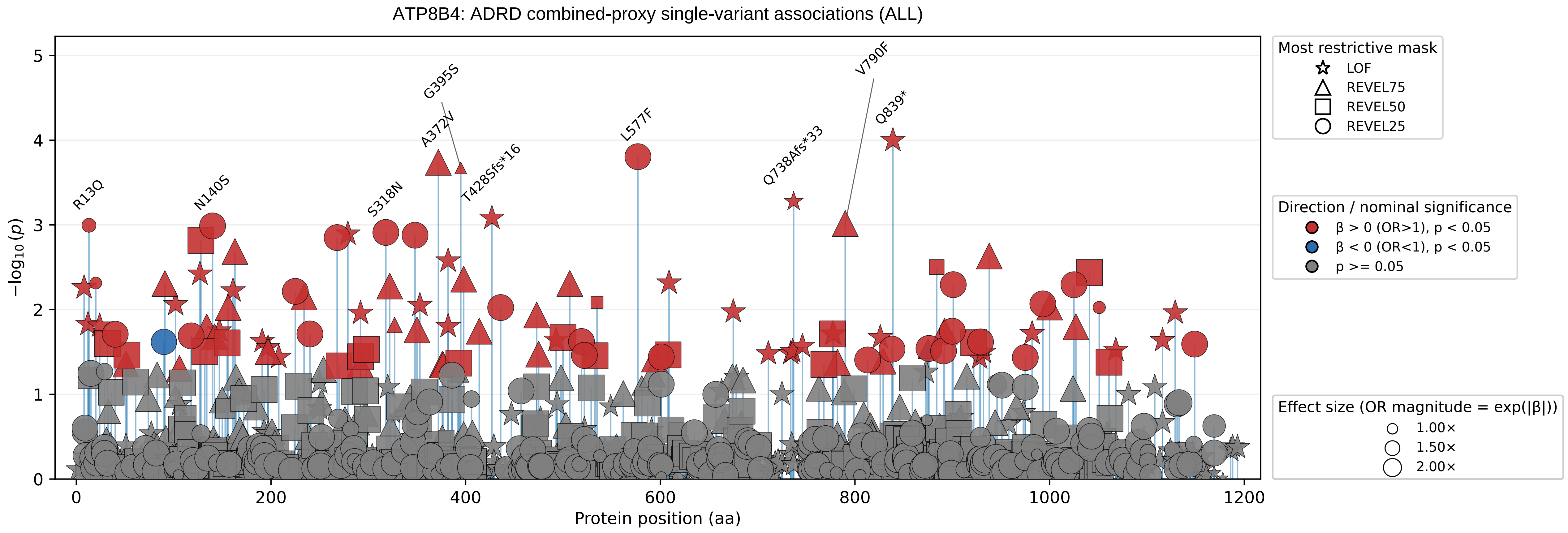

ATP8B4: ADRD combined-proxy single-variant associations (ALL)
R13Q
N140S
S318N
G395S
A372V
T428Sfs*16
L577F
Q738Afs*33
V790F
Q839*
−log10(p)
0
1
2
3
4
5
0
200
400
600
800
1000
1200
Protein position (aa)
Most restrictive mask
LOF
REVEL75
REVEL50
REVEL25
Direction / nominal significance
β > 0 (OR>1), p < 0.05
β < 0 (OR<1), p < 0.05
p >= 0.05
Effect size (OR magnitude = exp(|β|))
1.00×
1.50×
2.00×

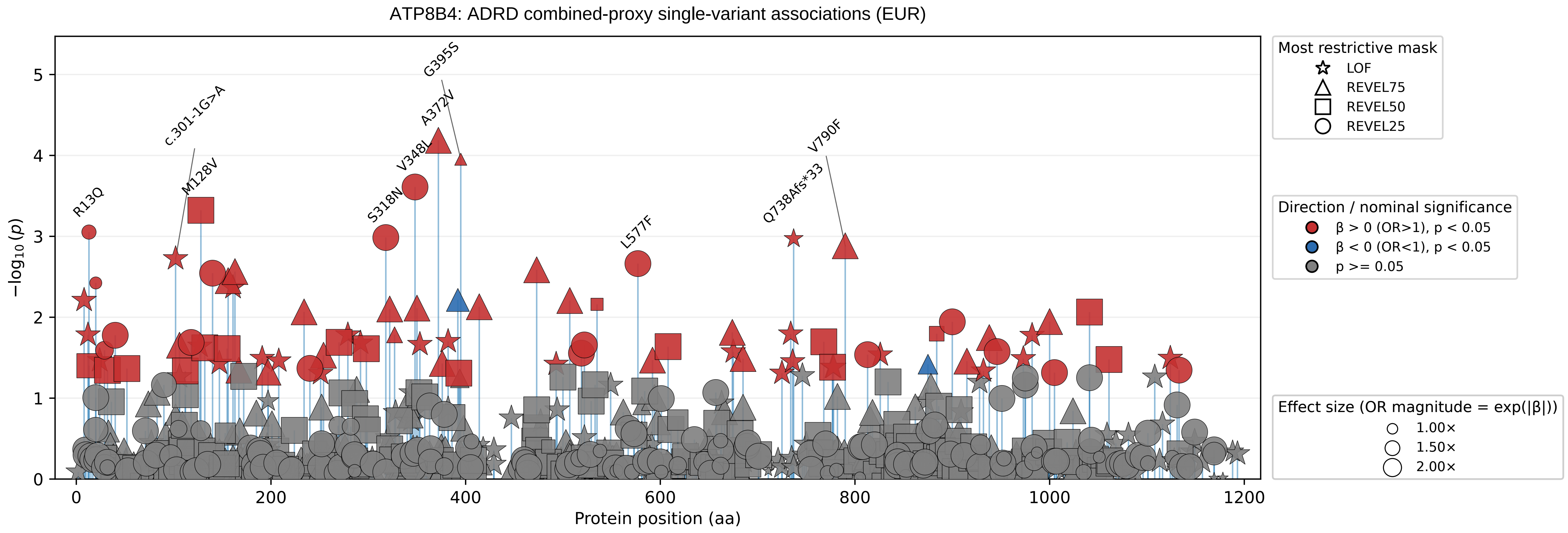

ATP8B4: ADRD combined-proxy single-variant associations (EUR)
−log10(p)
Protein position (aa)
R13Q
c.301-1G>A
M128V
G395S
A372V
V348L
S318N
L577F
V790F
Q738Afs*33
Most restrictive mask
LOF
REVEL75
REVEL50
REVEL25
Direction / nominal significance
β > 0 (OR>1), p < 0.05
β < 0 (OR<1), p < 0.05
p >= 0.05
Effect size (OR magnitude = exp(|β|))
1.00×
1.50×
2.00×

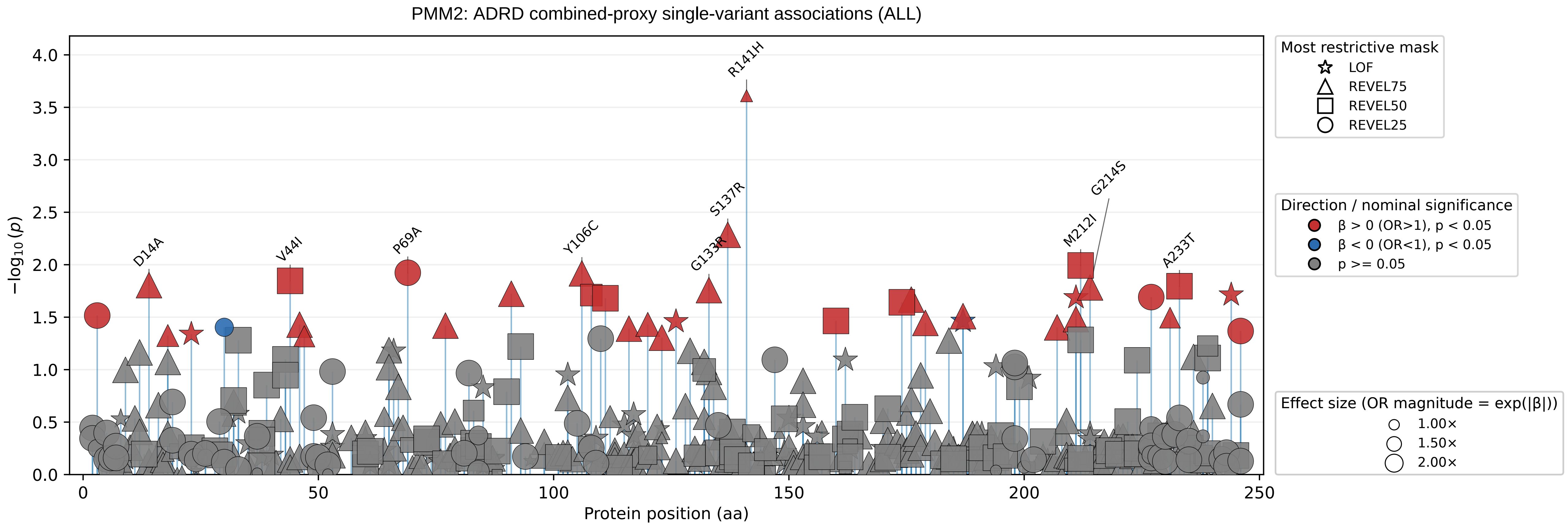

PMM2: ADRD combined-proxy single-variant associations (ALL)
−log10 (p)
Protein position (aa)
0.0
0.5
1.0
1.5
2.0
2.5
3.0
3.5
4.0
0
50
100
150
200
250
R141H
S137R
G133R
Y106C
P69A
V44I
D14A
G214S
M212I
A233T
Most restrictive mask
LOF
REVEL75
REVEL50
REVEL25
Direction / nominal significance
β > 0 (OR>1), p < 0.05
β < 0 (OR<1), p < 0.05
p >= 0.05
Effect size (OR magnitude = exp(|β|))
1.00×
1.50×
2.00×

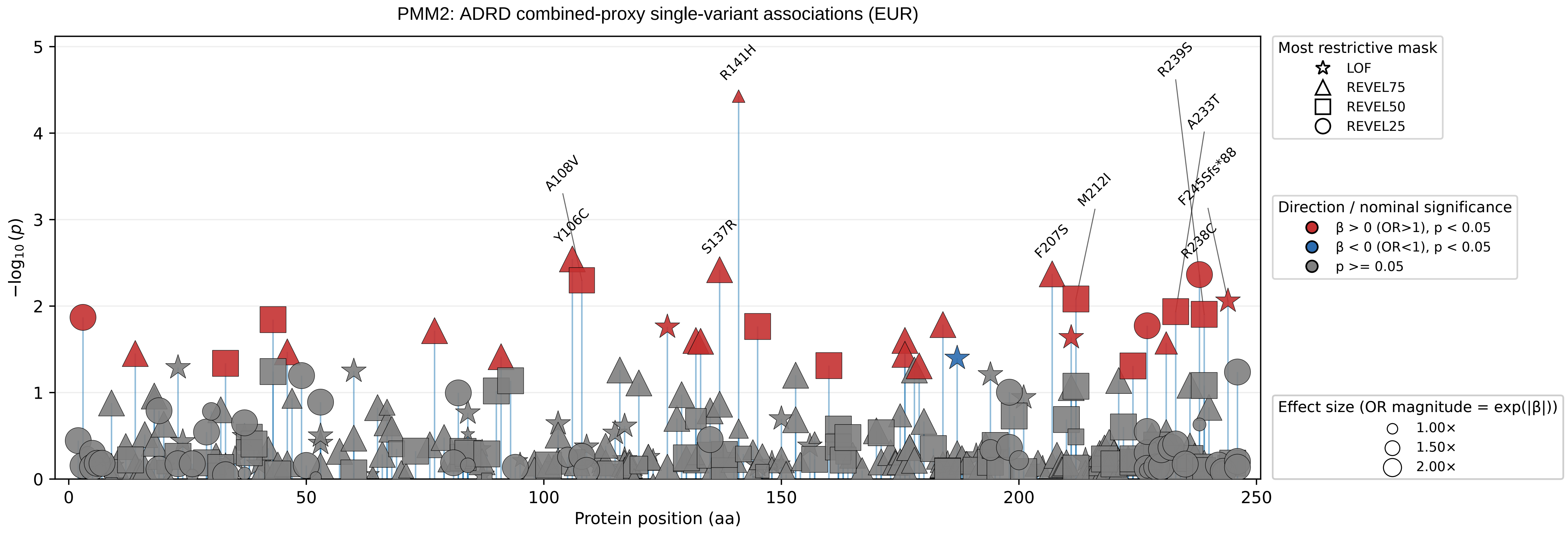

PMM2: ADRD combined-proxy single-variant associations (EUR)
−log10 (p)
Protein position (aa)
R141H
A108V
Y106C
S137R
F207S
M212I
R239S
A233T
F245Sfs*88
R238C
Most restrictive mask
LOF
REVEL75
REVEL50
REVEL25
Direction / nominal significance
β > 0 (OR>1), p < 0.05
β < 0 (OR<1), p < 0.05
p >= 0.05
Effect size (OR magnitude = exp(|β|))
1.00×
1.50×
2.00×

GRN: ADRD combined-proxy single-variant associations (ALL)

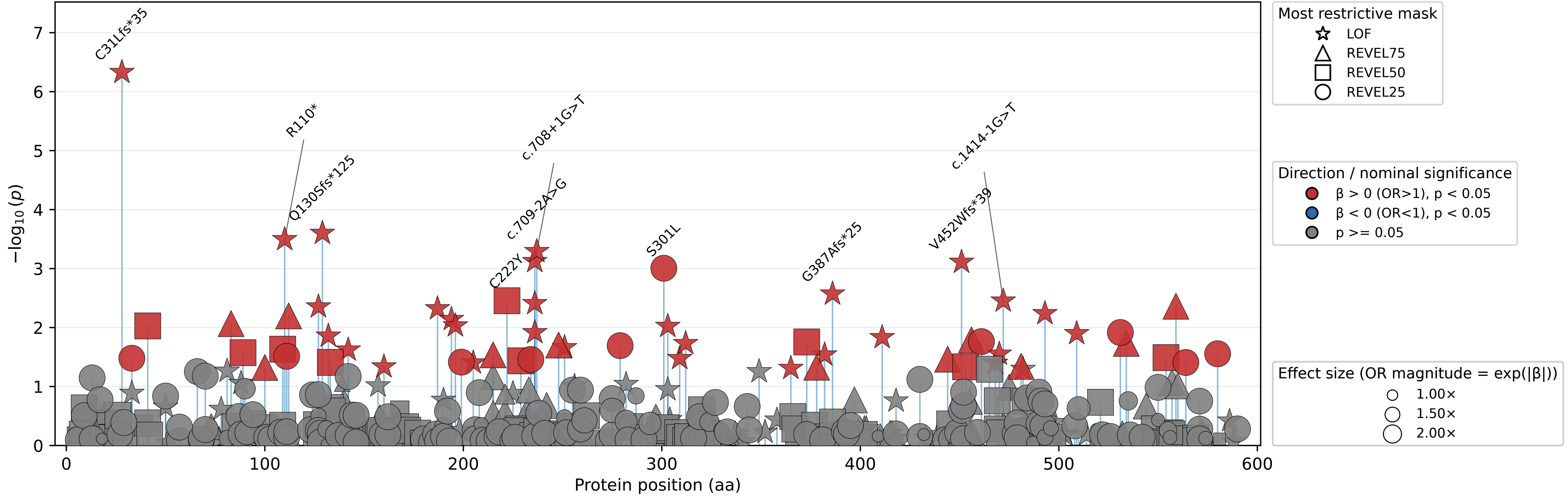

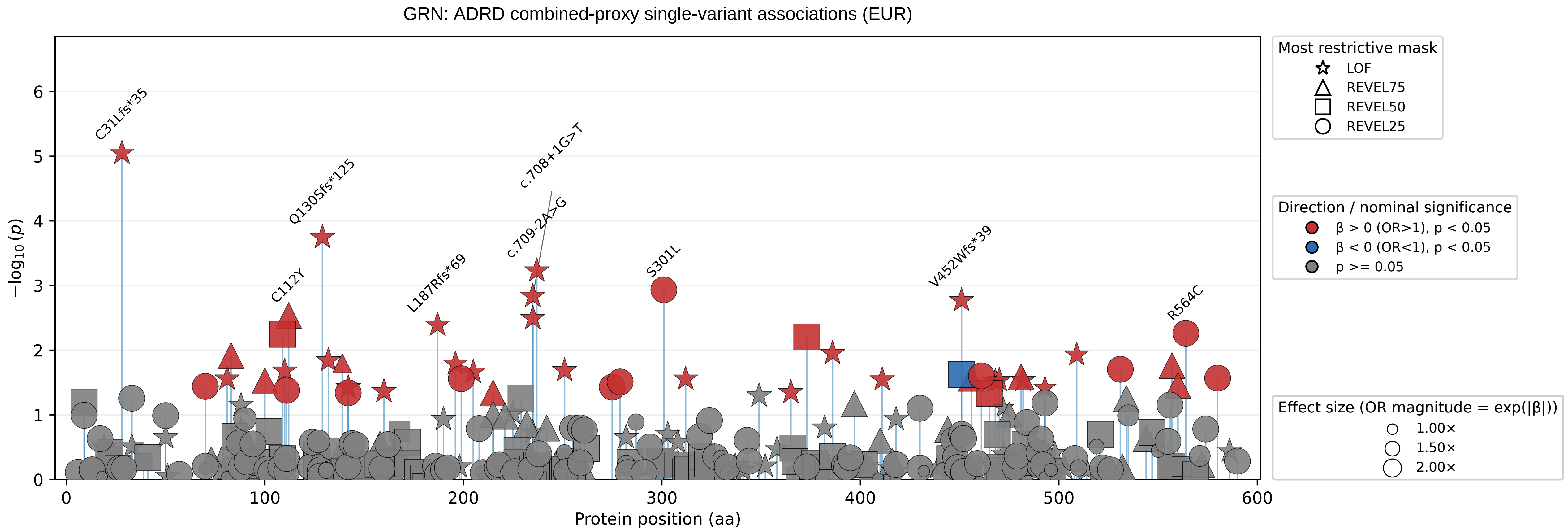
GRN: ADRD combined-proxy single-variant associations (EUR)
C31Lfs*35
Q130Sfs*125
c.708+1G>T
c.709-2A>G
C112Y
L187Rfs*69
S301L
V452Wfs*39
R564C
−log10(p)
Protein position (aa)
Most restrictive mask
LOF
REVEL75
REVEL50
REVEL25
Direction / nominal significance
β > 0 (OR>1), p < 0.05
β < 0 (OR<1), p < 0.05
p >= 0.05
Effect size (OR magnitude = exp(|β|))
1.00×
1.50×
2.00×

IMPA2: ADRD combined-proxy single-variant associations (ALL)

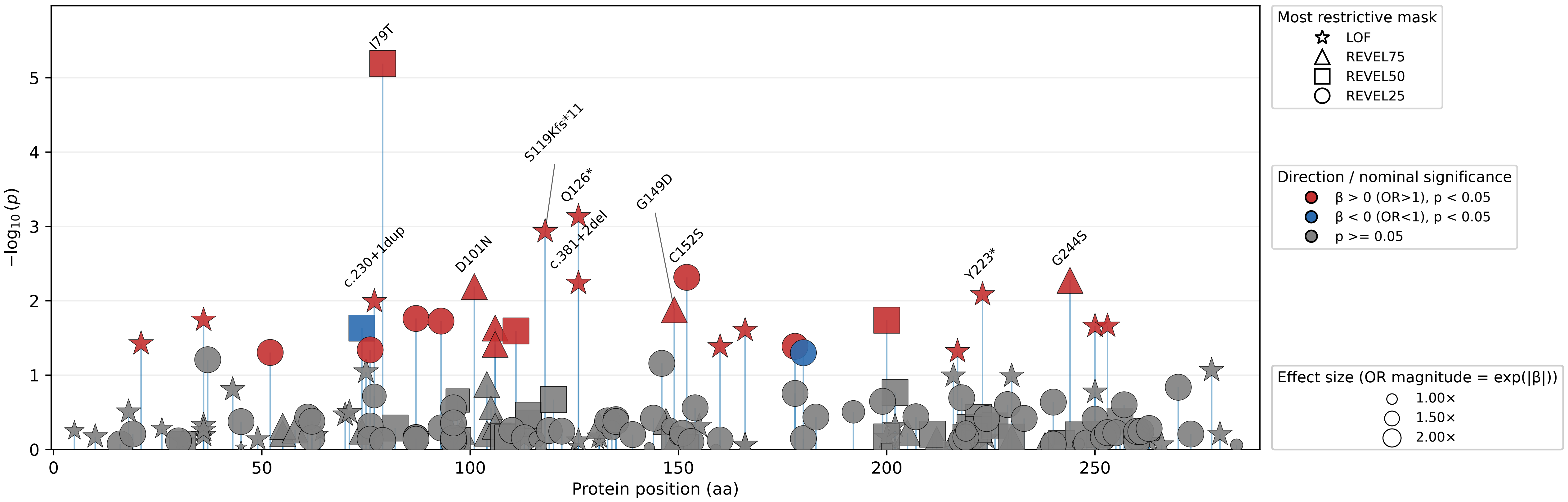

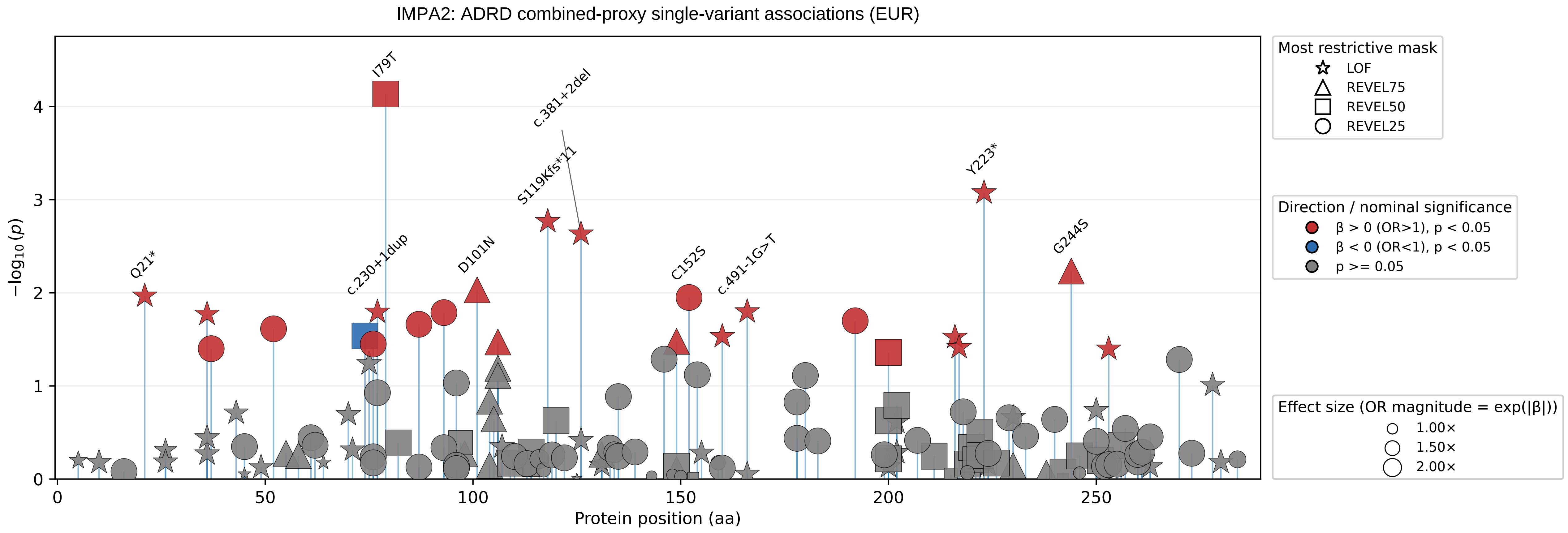

IMPA2: ADRD combined-proxy single-variant associations (EUR)
−log10(p)
Protein position (aa)
0
1
2
3
4
0
50
100
150
200
250
I79T
c.381+2del
S119Kfs*11
Y223*
Q21*
c.230+1dup
D101N
C152S
c.491-1G>T
G244S
Most restrictive mask
LOF
REVEL75
REVEL50
REVEL25
Direction / nominal significance
β > 0 (OR>1), p < 0.05
β < 0 (OR<1), p < 0.05
p >= 0.05
Effect size (OR magnitude = exp(|β|))
1.00×
1.50×
2.00×

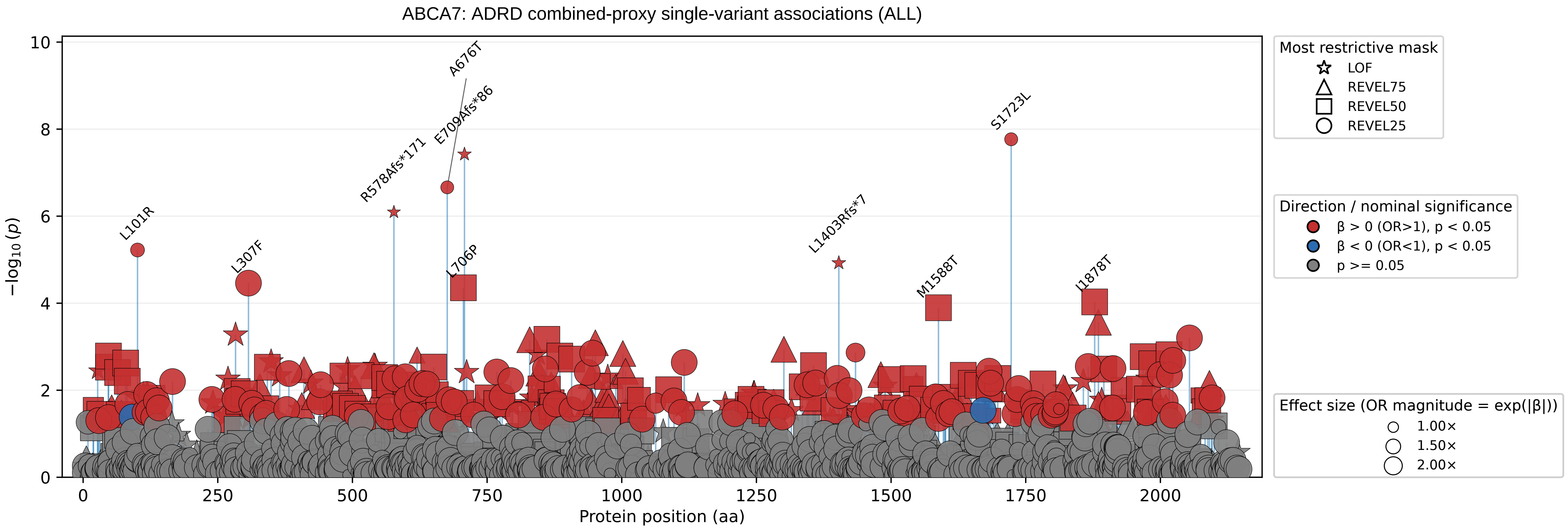
ABCA7: ADRD combined-proxy single-variant associations (ALL)
−log10(p)
Protein position (aa)
0
2
4
6
8
10
0
250
500
750
1000
1250
1500
1750
2000
L101R
L307F
R578Afs*171
A676T
E709Afs*86
L706P
L1403Rfs*7
M1588T
S1723L
I1878T
Most restrictive mask
LOF
REVEL75
REVEL50
REVEL25
Direction / nominal significance
β > 0 (OR>1), p < 0.05
β < 0 (OR<1), p < 0.05
p >= 0.05
Effect size (OR magnitude = exp(|β|))
1.00×
1.50×
2.00×

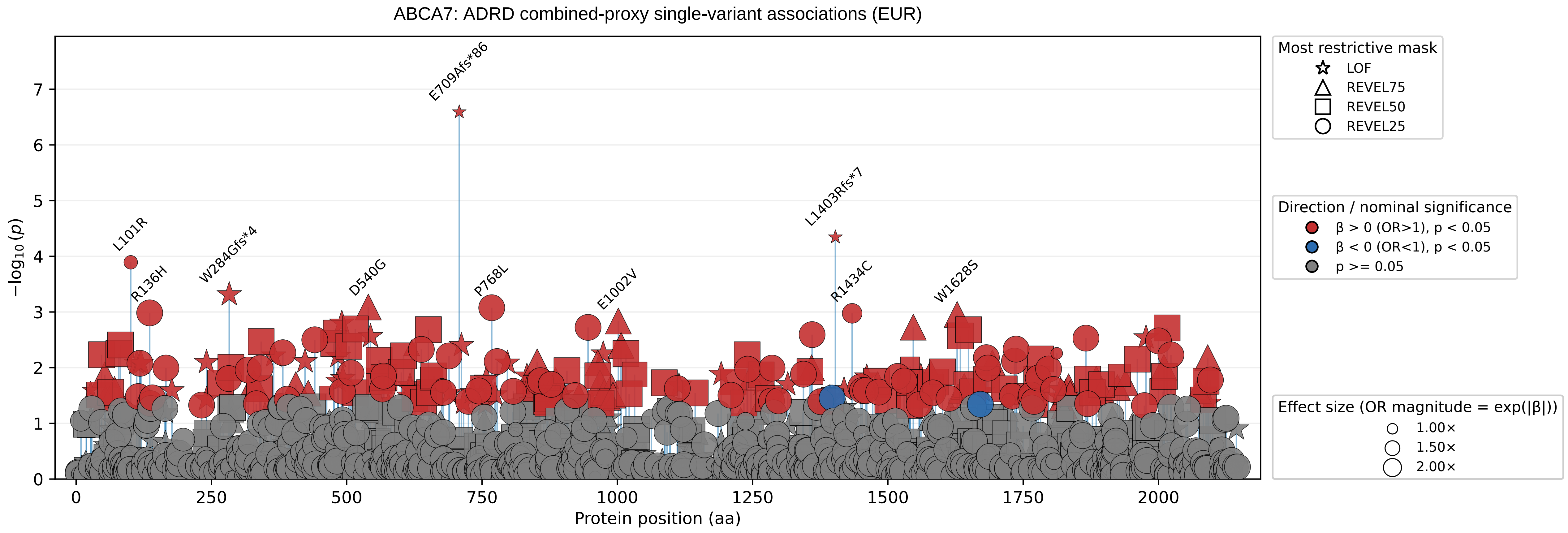

ABCA7: ADRD combined-proxy single-variant associations (EUR)
−log10(p)
Protein position (aa)
0
1
2
3
4
5
6
7
0
250
500
750
1000
1250
1500
1750
2000
E709Afs*86
L1403Rfs*7
L101R
R136H
W284Gfs*4
D540G
P768L
E1002V
R1434C
W1628S
Most restrictive mask
LOF
REVEL75
REVEL50
REVEL25
Direction / nominal significance
β > 0 (OR>1), p < 0.05
β < 0 (OR<1), p < 0.05
p >= 0.05
Effect size (OR magnitude = exp(|β|))
1.00×
1.50×
2.00×

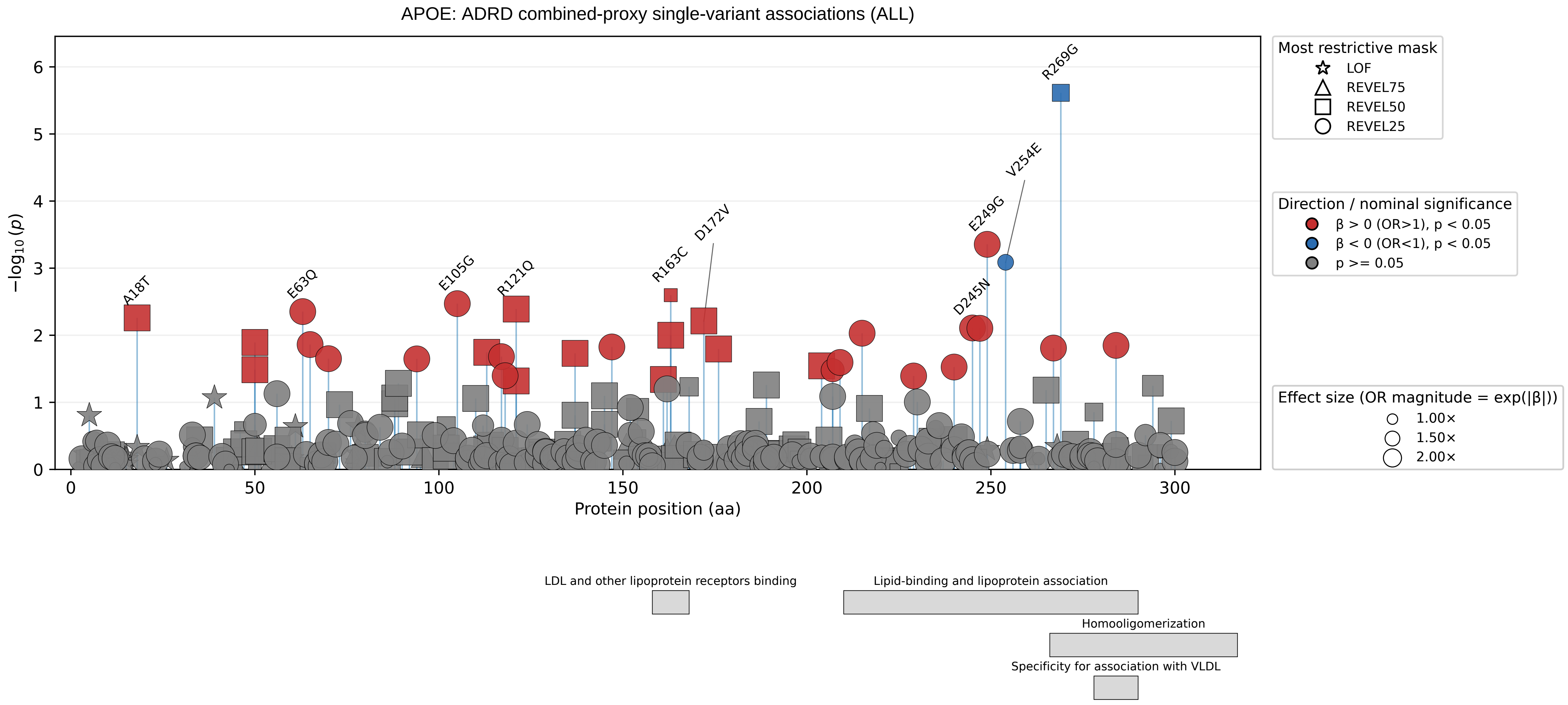
APOE: ADRD combined-proxy single-variant associations (ALL)
−log10 (p)
Protein position (aa)
A18T
E63Q
E105G
R121Q
R163C
D172V
D245N
E249G
V254E
R269G
Most restrictive mask
LOF
REVEL75
REVEL50
REVEL25
Direction / nominal significance
β > 0 (OR>1), p < 0.05
β < 0 (OR<1), p < 0.05
p >= 0.05
Effect size (OR magnitude = exp(|β|))
1.00×
1.50×
2.00×
LDL and other lipoprotein receptors binding
Lipid-binding and lipoprotein association
Homooligomerization
Specificity for association with VLDL

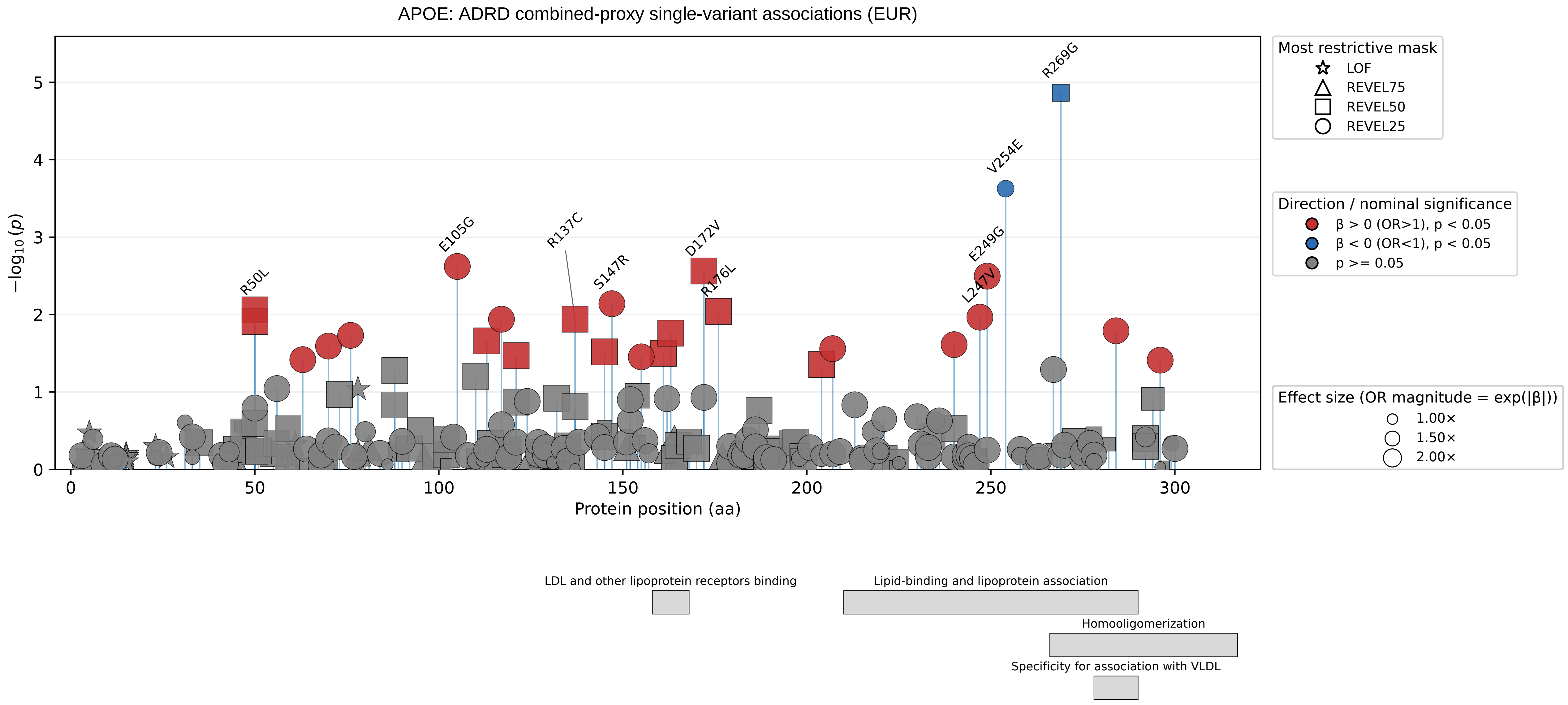
APOE: ADRD combined-proxy single-variant associations (EUR)
R269G
V254E
E249G
L247V
D172V
R176L
S147R
R137C
E105G
R50L
−log10(p)
Protein position (aa)
Most restrictive mask
LOF
REVEL75
REVEL50
REVEL25
Direction / nominal significance
β > 0 (OR>1), p < 0.05
β < 0 (OR<1), p < 0.05
p >= 0.05
Effect size (OR magnitude = exp(|β|))
1.00×
1.50×
2.00×
LDL and other lipoprotein receptors binding
Lipid-binding and lipoprotein association
Homooligomerization
Specificity for association with VLDL

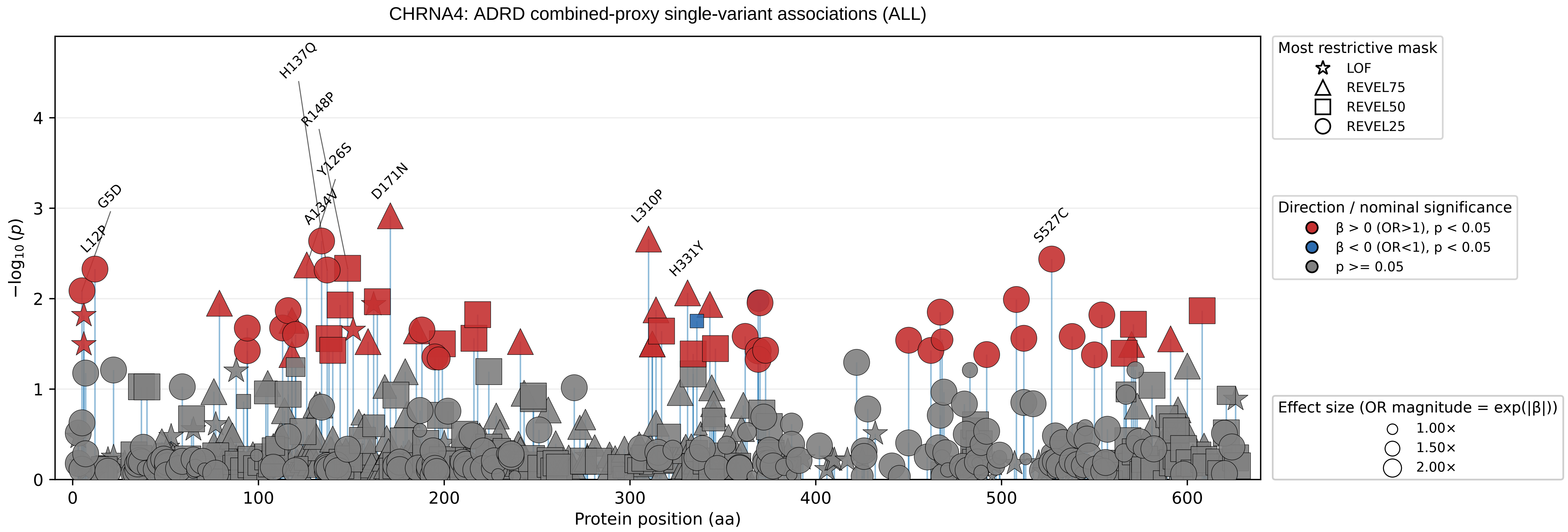

CHRNA4: ADRD combined-proxy single-variant associations (ALL)
−log10 (p)
Protein position (aa)
0
1
2
3
4
0
100
200
300
400
500
600
H137Q
R148P
Y126S
D171N
G5D
A134V
L12P
L310P
H331Y
S527C
Most restrictive mask
LOF
REVEL75
REVEL50
REVEL25
Direction / nominal significance
β > 0 (OR>1), p < 0.05
β < 0 (OR<1), p < 0.05
p >= 0.05
Effect size (OR magnitude = exp(|β|))
1.00×
1.50×
2.00×

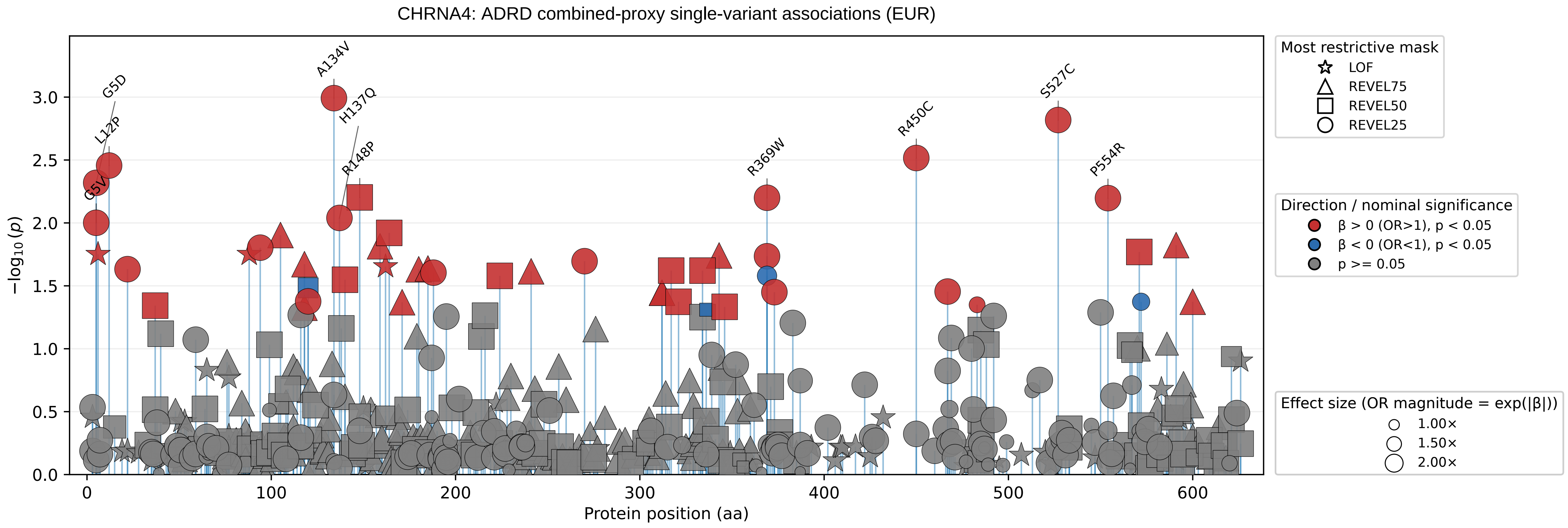
CHRNA4: ADRD combined-proxy single-variant associations (EUR)
−log10(p)
Protein position (aa)
G5D
L12P
G5V
A134V
H137Q
R148P
R369W
R450C
S527C
P554R
Most restrictive mask
LOF
REVEL75
REVEL50
REVEL25
Direction / nominal significance
β > 0 (OR>1), p < 0.05
β < 0 (OR<1), p < 0.05
p >= 0.05
Effect size (OR magnitude = exp(|β|))
1.00×
1.50×
2.00×
