## Supplementary_Note1 for "Population-scale burden analysis of rare damaging coding variants identifies novel risk genes for Alzheimer’s disease and related dementias and Parkinson’s disease and related disorders"

### Supplementary Note 1

#### Mathematical derivation and simulation evaluation of effect-size adjustment for combined proxy phenotypes

Population-scale burden analysis of rare damaging coding variants

#### Overview

The combined-proxy phenotypes used in the population cohorts include two mutually exclusive case strata: participants with a personal diagnosis, self-reported diagnosis or clinical ascertainment, and participants without a personal diagnosis who report an affected first-degree relative. Both case strata are compared with the same clean control group. This note derives the exact relationship between the effect estimated for the combined-proxy phenotype and the effects in the directly diagnosed and family-history-only strata, evaluates the commonly used first-order attenuation approximation, and describes a simulation study designed to assess the approximation under rare-variant gene-burden settings.

Throughout this note, the term *directly diagnosed* encompasses diagnosed, self-reported or clinically ascertained cases, as appropriate for each cohort.

#### Definition of the phenotype strata

For a given cohort, phenotype and ancestry stratum, let

$$D = \{\text{directly diagnosed/self-reported/clinical cases}\},$$

$$F = \{\text{family-history-only cases}\},$$

$$C = \{\text{clean controls}\},$$

and

$$M = D \cup F$$

denote the combined-proxy case group.

The strata $D$, $F$ and $C$ are mutually exclusive. Family-history-only cases are participants who are not directly affected but report at least one affected first-degree relative. Clean controls have neither a personal diagnosis nor a reported affected first-degree relative. Therefore, the directly diagnosed and family-history-only comparisons use the same control group $C$.

Let $G$ denote the gene-level burden score. For a binary carrier model,

$$G = \begin{cases} 1, & \text{at least one qualifying damaging allele is present,} \\ 0, & \text{otherwise.} \end{cases}$$

REGENIE's maximum-allele mask and a summed-dosage mask may take values greater than one. However, for sufficiently rare qualifying variants, nearly all nonzero burden scores equal one, and the binary-carrier derivation provides a close approximation.

### Exact common-control mixture identity

Consider the directly diagnosed-versus-control and family-history-only-versus-control odds at burden value $G = g$:

$$\frac{\Pr(D \mid G = g)}{\Pr(C \mid G = g)} = a_D \exp(\beta_D g),$$

and

$$\frac{\Pr(F \mid G = g)}{\Pr(C \mid G = g)} = a_F \exp(\beta_F g),$$

where $a_D$ and $a_F$ are the corresponding baseline case-to-control odds at $G = 0$, and $\beta_D$ and $\beta_F$ are the burden log-odds effects in the two case strata.

Because $D$ and $F$ are mutually exclusive and use the same control stratum,

$$\begin{aligned}\frac{\Pr(M \mid G = g)}{\Pr(C \mid G = g)} &= \frac{\Pr(D \mid G = g) + \Pr(F \mid G = g)}{\Pr(C \mid G = g)} \\ &= a_D \exp(\beta_D g) + a_F \exp(\beta_F g).\end{aligned}$$

At $G = 0$, the combined case-to-control odds are

$$a_D + a_F.$$

At $G = 1$, the combined case-to-control odds are

$$a_D \exp(\beta_D) + a_F \exp(\beta_F).$$

Define

$$w_0 = \frac{a_D}{a_D + a_F} = \frac{\Pr(D \mid G = 0)}{\Pr(D \mid G = 0) + \Pr(F \mid G = 0)}.$$

Thus, $w_0$ is the directly diagnosed fraction among combined-proxy cases with burden score zero. The exact combined-proxy odds ratio for a binary burden score is

$$\text{OR}_M = w_0 \exp(\beta_D) + (1 - w_0) \exp(\beta_F),$$

and the exact combined-proxy log-odds effect is

$$\beta_M = \log \left[ w_0 \exp(\beta_D) + (1 - w_0) \exp(\beta_F) \right]. \tag{1}$$

Equation (1) explicitly accounts for the common control group. The control probability appears in both component odds and cancels when the two mutually exclusive case strata are added.

For a general burden value $g$,

$$\log\left[\frac{\Pr(M \mid G=g)}{\Pr(C \mid G=g)}\right] = \log\left[a_D\exp(\beta_D g) + a_F\exp(\beta_F g)\right].$$

This expression is not generally linear in $g$. Equation (1) is therefore an exact logistic-slope identity for a binary burden score. It is an approximation for maximum-allele or summed-dosage scores when values greater than one occur, although that approximation is expected to be close when multiple qualifying alleles in the same participant are uncommon.

### Relationship to the observed directly diagnosed fraction

The exact quantity $w_0$ is defined among combined cases with $G = 0$. In practice, the readily observed quantity is

$$w_{\text{diag}} = \frac{N_D}{N_D + N_F},$$

the overall directly diagnosed fraction among all combined-proxy cases.

Because qualifying rare-variant carriers constitute a small proportion of the cohort,

$$w_{\text{diag}} \approx w_0.$$

In the simulations described below, the mean absolute difference between $w_{\text{diag}}$ and $w_0$ was 0.00109, the 95th percentile was 0.00374, and the maximum difference was 0.00605. We therefore use the observed $w_{\text{diag}}$ in cohort-level applications and refer to it as $w$ below.

### Family-history attenuation parameter

Let the family-history-only effect be related to the directly diagnosed effect through

$$\beta_F = \kappa\beta_D,$$

where $\kappa$ is the effective family-history attenuation parameter. Substituting this relationship into Equation (1) gives

$$\beta_M = \log\left[w\exp(\beta_D) + (1-w)\exp(\kappa\beta_D)\right]. \tag{2}$$

The value $\kappa = 0.5$ is a useful central approximation motivated by the approximately one-half genetic sharing between first-degree relatives. However, $\kappa = 0.5$ is not an exact identity for an "at least one affected first-degree relative" phenotype.

The effective value of $\kappa$ can differ from 0.5 because the family-history phenotype may include multiple relatives, uses a nonlinear "at least one affected relative" rule, conditions family-history-only cases on the proband being unaffected, and compares them with controls selected to have neither personal nor reported family disease. Reporting sensitivity and specificity also affect the observed attenuation.

For $r$ queried relatives with unaffected true status and per-relative reporting specificity $s$, the probability of at least one false-positive family-history report is

$$1 - s^r.$$

For example, with three queried relatives and specificity $s = 0.99$,

$$1 - 0.99^3 = 0.0297.$$

Thus, a per-relative specificity of 99% corresponds to an approximately 3% family-level false-positive probability when three relatives are considered.

#### First-order approximation

Expanding Equation (2) around $\beta_D = 0$ gives

$$\begin{aligned}\beta_M = {} & [w + (1-w)\kappa]\,\beta_D \\ & + \frac{1}{2} w(1-w)(1-\kappa)^2 \beta_D^2 \\ & + \frac{1}{6} w(1-w)(1-2w)(1-\kappa)^3 \beta_D^3 \\ & + O(\beta_D^4).\end{aligned} \tag{3}$$

Retaining only the first-order term gives

$$\beta_M \approx [w + (1-w)\kappa]\,\beta_D,$$

and hence

$$\beta_D \approx \frac{\beta_M}{w + (1-w)\kappa}. \tag{4}$$

For $\kappa = 0.5$,

$$w + (1-w)\kappa = \frac{1+w}{2},$$

so Equation (4) becomes

$$\beta_D \approx \frac{2}{1+w}\beta_M. \tag{5}$$

Equation (5) is therefore the first-order, small-effect approximation to Equation (2). It is not an exact transformation for arbitrary effect sizes.

The leading omitted term in Equation (3) is

$$\frac{1}{2} w(1-w)(1-\kappa)^2 \beta_D^2,$$

which is nonnegative. For a positive risk effect, the exact combined-proxy log-odds effect is therefore larger than its first-order linear approximation, and applying Equation (4) tends to overestimate a positive directly diagnosed effect when $\kappa$ is correctly specified.

#### Exact inversion

For a specified value of $\kappa$, the adjusted directly diagnosed-scale effect is defined as the unique solution of

$$\log\left[w \exp(\beta_D) + (1-w)\exp(\kappa\beta_D)\right] - \beta_M = 0. \tag{6}$$

The left-hand side is monotone in $\beta_D$ for $\kappa \geq 0$, so Equation (6) can be solved numerically without ambiguity.

##### Closed-form solution for $\kappa = 0.5$

When $\kappa = 0.5$,

$$\exp(\beta_M) = w \exp(\beta_D) + (1 - w) \exp(\beta_D/2).$$

Let

$$t = \exp(\beta_D/2).$$

Then

$$wt^2 + (1 - w)t - \exp(\beta_M) = 0.$$

The positive root is

$$t = \frac{\sqrt{(1 - w)^2 + 4w \exp(\beta_M)} - (1 - w)}{2w},$$

and therefore

$$\beta_D = 2 \log \left[ \frac{\sqrt{(1 - w)^2 + 4w \exp(\beta_M)} - (1 - w)}{2w} \right]. \tag{7}$$

A numerically stable equivalent form is

$$\beta_D = 2 \log \left[ \frac{2 \exp(\beta_M)}{\sqrt{(1 - w)^2 + 4w \exp(\beta_M)} + (1 - w)} \right]. \tag{8}$$

The limiting cases are

$$w = 0 : \qquad \beta_D = 2\beta_M,$$

and

$$w = 1 : \qquad \beta_D = \beta_M.$$

For other positive values of $\kappa$, Equation (6) can be solved numerically.

#### Standard errors and confidence intervals

Define

$$f(\beta_D) = \log \left[ w \exp(\beta_D) + (1 - w) \exp(\kappa\beta_D) \right].$$

Its derivative is

$$f'(\beta_D) = \frac{w \exp(\beta_D) + \kappa(1 - w) \exp(\kappa\beta_D)}{w \exp(\beta_D) + (1 - w) \exp(\kappa\beta_D)}. \tag{9}$$

If $\widehat{\beta}_D = f^{-1}(\widehat{\beta}_M)$, the delta-method standard error is

$$\mathrm{SE}(\widehat{\beta}_D) \approx \frac{\mathrm{SE}(\widehat{\beta}_M)}{f'(\widehat{\beta}_D)}. \tag{10}$$

At the null,

$$f'(0) = w + (1-w)\kappa.$$

Thus, Equation (10) reduces at the null to the same factor used in the first-order adjustment. For $\kappa = 0.5$,

$$f'(0) = \frac{1+w}{2}.$$

Cohort-specific confidence limits may alternatively be transformed directly. If

$$L_M = \widehat{\beta}_M - 1.96\,\mathrm{SE}(\widehat{\beta}_M)$$

and

$$U_M = \widehat{\beta}_M + 1.96\,\mathrm{SE}(\widehat{\beta}_M),$$

then an asymmetric transformed interval is

$$\left[f^{-1}(L_M),\, f^{-1}(U_M)\right]. \tag{11}$$

The transformation is monotone and satisfies $f(0) = 0$. It therefore does not change the direction of effect or the cohort-level null hypothesis. Original cohort-level association $P$ values remain valid tests of no association; the transformation is used to place effect estimates on an approximate directly diagnosed scale for comparison and meta-analysis.

### Simulation design

We simulated 100,000 independent nuclear families per replicate. Each family contained a proband, two parents and one sibling. Twenty independent rare variants were generated with heterogeneous allele frequencies and rescaled to produce a target gene-level carrier frequency of either 0.5% or 1%.

Founder haplotypes were generated under Hardy-Weinberg equilibrium. Proband and sibling genotypes were generated through Mendelian transmission from the parental haplotypes. Individuals could therefore carry zero, one or multiple qualifying alleles.

Three gene-level burden encodings were evaluated:

$$G_{\text{carrier}} = I\left(\sum_j G_j > 0\right),$$

$$G_{\max} = \max_j G_j,$$

and

$$G_{\text{sum}} = \sum_j G_j,$$

where $G_j \in \{0, 1, 2\}$ is the alternate-allele count at variant $j$.

Disease status was generated independently for each family member under

$$\text{logit } \Pr(Y = 1 \mid G_{\text{sum}}) = \text{logit}(0.05) + \log(\text{OR}_D)G_{\text{sum}},$$

with

$$\text{OR}_D \in \{1, 1.5, 2, 4\}.$$

The summed-dosage score was used as the data-generating score so that the simulation explicitly allowed more than one qualifying allele to contribute to disease risk. Carrier, maximum-allele and summed-dosage analyses were then fit to the same simulated datasets.

For each relative, a reported family-history value was generated from the relative's true disease status using

$$\text{Se}_{FH} = \Pr(\text{reported affected} \mid \text{truly affected})$$

with values

$$\text{Se}_{FH} \in \{0.50, 0.75, 0.90, 1.00\},$$

and

$$\text{Sp}_{FH} = \Pr(\text{reported unaffected} \mid \text{truly unaffected})$$

with values

$$\text{Sp}_{FH} \in \{0.99, 1.00\}.$$

The proband was classified as a directly diagnosed case if personally affected, a family-history-only case if unaffected with at least one reported affected first-degree relative, and a clean control if unaffected with no reported affected relative.

For each phenotype and burden encoding, Firth-corrected logistic regression was fit with an intercept and burden score. The simulation comprised

$$4 \text{ odds ratios} \times 4 \text{ sensitivities} \times 2 \text{ specificities} \times 2 \text{ carrier frequencies} = 64$$

scenarios. Each scenario was evaluated in 200 replicates, giving 12,800 simulated family datasets, 38,400 score-specific analysis combinations and 115,200 fitted phenotype models.

The primary simulation summaries were:

1. agreement between the fitted combined-proxy effect and Equation (1);
2. the empirical attenuation ratio;
3. bias of the unadjusted combined-proxy estimate;
4. bias after the first-order $\kappa = 0.5$ correction;
5. bias after exact inversion with $\kappa = 0.5$;
6. bias after exact inversion using the scenario-specific empirical attenuation;
7. the frequency and effect of burden scores greater than one; and
8. null rejection and confidence-interval coverage.

For non-null scenarios, bias was evaluated relative to the fitted directly diagnosed effect within the same simulated dataset:

$$\text{Bias} = \widehat{\beta}_{\text{adjusted}} - \widehat{\beta}_D.$$

This choice isolates error caused by proxy mixing and adjustment from finite-sample bias in the directly diagnosed model itself.

### Simulation results

#### Convergence and burden-score distributions

All 115,200 Firth models converged.

Observed gene-level carrier frequencies closely matched the target values. Under summed-dosage coding, the proportion of carriers with more than one qualifying allele averaged approximately 0.23% for the 0.5% carrier-frequency scenarios and 0.46% for the 1% carrier-frequency scenarios. Across scenario means, the range was approximately 0.20%-0.49% of carriers.

Because scores greater than one were uncommon, maximum-allele and summed-dosage estimates were similar in this simulation. For the combined-proxy effect,

$$\text{mean}\left|\widehat{\beta}_{\text{sum}} - \widehat{\beta}_{\text{max}}\right| = 0.00403,$$

the 95th percentile absolute difference was 0.01145, and the maximum absolute difference was 0.03648. This supports the use of the binary-carrier derivation as a close approximation under the simulated rare-variant architecture, while not implying that maximum and summed dosage are mathematically identical.

#### Exact common-control identity

Across the 12,800 maximum-allele analyses, the absolute residual

$$\left|\widehat{\beta}_M - \log\left[w_0 \exp(\widehat{\beta}_D) + (1 - w_0)\exp(\widehat{\beta}_F)\right]\right|$$

had mean 0.00323, median 0.00279, 95th percentile 0.00631, and maximum 0.01223. For non-null maximum-allele scenarios, the mean absolute residual was 0.00281 and the maximum was 0.00805. Thus, Equation (1) accurately reproduced the fitted combined-proxy effect. Agreement was similarly close for carrier and summed-dosage coding Supplementary Fig. 18.

#### Direct-case proportions

Across the simulated reporting scenarios, the scenario-mean $w_0$ values ranged from 0.236 to 0.419. The overall directly diagnosed fraction $w_{\text{diag}}$ closely approximated $w_0$, with mean absolute difference 0.00109 and maximum difference 0.00605.

#### Null calibration

Under the null and maximum-allele coding, the empirical rejection rates at nominal $\alpha = 0.05$, pooled across null scenarios, were 6.1% for directly diagnosed analyses, 4.7% for family-history-only analyses, and 4.6% for combined-proxy analyses.

The simulation was primarily designed to evaluate effect-size calibration rather than reproduce every detail of REGENIE's likelihood-ratio inference. Nevertheless, the combined-proxy null rejection rate was close to nominal.

#### Empirical family-history attenuation

For each non-null scenario, the empirical attenuation was summarized as the ratio of the scenario-mean family-history-only and directly diagnosed coefficients:

$$\kappa_{\text{emp}} = \frac{\overline{\widehat{\beta}}_F}{\overline{\widehat{\beta}}_D}.$$

Using maximum-allele coding, the overall scenario-level range was

$$0.386 \le \kappa_{\text{emp}} \le 0.602.$$

**Empirical attenuation by directly diagnosed odds ratio and reporting specificity.** Ranges are across family-history sensitivities of 0.50-1.00 and carrier frequencies of 0.5%-1%.

| Directly diagnosed OR | $\kappa_{\text{emp}}$, specificity 0.99 | $\kappa_{\text{emp}}$, specificity 1.00 |
|---:|:---:|:---:|
| 1.5 | 0.386-0.479 | 0.486-0.568 |
| 2.0 | 0.430-0.487 | 0.524-0.588 |
| 4.0 | 0.467-0.539 | 0.578-0.602 |

Under perfect reporting specificity, empirical attenuation was approximately 0.49-0.60. With per-relative specificity of 0.99, it was approximately 0.39-0.54.

These results support $\kappa = 0.5$ as a reasonable central value.

#### Bias of the effect-size adjustments

Across all non-null maximum-allele scenarios, the unadjusted combined-proxy estimate was attenuated relative to the directly diagnosed estimate:

$$\text{mean bias} = -0.2768, \qquad \text{RMSE} = 0.3185.$$

The first-order $\kappa = 0.5$ correction reduced attenuation but had a positive mean bias:

$$\text{mean bias} = 0.0663, \qquad \text{RMSE} = 0.1536.$$

Exact inversion with $\kappa = 0.5$ further reduced the aggregate error:

$$\text{mean bias} = 0.0174, \qquad \text{RMSE} = 0.1159.$$

As a diagnostic, exact inversion using the scenario-specific empirical $\kappa$ gave

$$\text{mean bias} = -0.0039, \qquad \text{RMSE} = 0.0045.$$

The latter analysis is not proposed as the primary adjustment because the true attenuation is not known in empirical data. It demonstrates that the common-control mixture equation itself is accurate and that remaining error is primarily attributable to uncertainty in $\kappa$.

**Ranges of scenario-mean log-odds bias for maximum-allele coding.** Each range is across family-history sensitivity and carrier-frequency settings.

| Direct OR | Specificity | Unadjusted bias | First-order bias, $\kappa = 0.5$ | Exact-inverse bias, $\kappa = 0.5$ |
|---|---|---|---|---|
| 1.5 | 0.99 | -0.180 to -0.156 | -0.035 to -0.003 | -0.041 to -0.011 |
| 1.5 | 1.00 | -0.166 to -0.111 | 0.003 to 0.040 | -0.007 to 0.031 |
| 2.0 | 0.99 | -0.296 to -0.246 | -0.020 to 0.012 | -0.041 to -0.011 |
| 2.0 | 1.00 | -0.233 to -0.175 | 0.041 to 0.092 | 0.014 to 0.064 |
| 4.0 | 0.99 | -0.511 to -0.441 | 0.065 to 0.166 | -0.031 to 0.052 |
| 4.0 | 1.00 | -0.427 to -0.317 | 0.168 to 0.261 | 0.060 to 0.129 |

Exact inversion consistently removed the Taylor-linearization component of the error. Residual bias under fixed $\kappa = 0.5$ followed the direction expected from the empirical attenuation: the adjustment tended to underestimate the direct effect when the true effective attenuation was below 0.5 and overestimate it when the attenuation was above 0.5.

#### Error from the first-order approximation

The following table isolates the mathematical error caused by using Equation (4) rather than exact inversion. It assumes the observed simulation range

$$0.236 \leq w \leq 0.419.$$

The first numerical column fixes $\kappa = 0.5$. The second allows the correctly specified value of $\kappa$ to range from 0.4 to 0.6.

| True directly diagnosed OR | OR-scale overestimate from linear correction, $\kappa = 0.5$ | OR-scale overestimate, correctly specified $\kappa \in [0.4, 0.6]$ |
|---|---|---|
| 1.2 | 0.12%-0.14% | 0.07%-0.23% |
| 1.5 | 0.62%-0.72% | 0.35%-1.14% |
| 2.0 | 1.88%-2.12% | 1.05%-3.40% |
| 3.0 | 4.93%-5.47% | 2.73%-8.89% |
| 4.0 | 8.14%-8.90% | 4.47%-14.68% |

The first-order approximation is therefore close for modest effects but becomes increasingly inaccurate for large burden effects. Exact inversion avoids this approximation error.

#### Confidence-interval behavior

For the causal summed-dosage score, average 95% confidence-interval coverage after exact $\kappa = 0.5$ inversion was approximately 0.93-0.94 for OR 1.5 and 0.87-0.93 for OR 2. Coverage was lower for OR 4, approximately 0.30-0.56 depending on reporting specificity. Directly diagnosed model coverage was also only approximately 0.60-0.62 for OR 4.

Coverage was evaluated against the disease-generating log odds ratio. Because the directly diagnosed-versus-clean-control coefficient can differ from that generating parameter, these results do not distinguish differences in the target parameter from finite-sample interval behavior.

### Effect-size adjustment used in the revised analysis

For each combined-proxy cohort, phenotype and ancestry stratum, we calculated

$$w = \frac{N_{\text{direct}}}{N_{\text{direct}} + N_{\text{FH-only}}}.$$

The primary adjusted effect was obtained by solving

$$\widehat{\beta}_M = \log\left[w\exp(\widehat{\beta}_D) + (1-w)\exp(0.5\widehat{\beta}_D)\right].$$

For $\kappa = 0.5$, we used the closed-form solution in Equation (8). The corresponding standard error was calculated using Equation (10). Directly diagnosed cohorts required no adjustment and are represented by $w = 1$.

All reported empirical meta-analysis results use $\kappa = 0.5$ for combined-proxy effect adjustment. The simulation analyses characterize variation in effective family-history attenuation and the resulting error when a fixed attenuation parameter is used; they do not establish the robustness of individual empirical gene/mask associations to alternative $\kappa$ values. Adjusted effect estimates and confidence intervals are therefore interpreted conditional on the specified attenuation model.

### Conclusion

The common-control structure leads to the exact mixture identity in Equation (1) for a binary carrier score. The approximation factor

$$\frac{2}{1+w}$$

is the first-order form of this identity under the additional assumption $\kappa = 0.5$ and it is not exact for large effects.

The simulations confirmed that the exact common-control identity accurately reproduced the fitted combined-proxy effect and that the overall directly diagnosed proportion closely approximated the theoretically exact $w_0$. They also showed that the effective family-history attenuation varied approximately from 0.4 to 0.6 under the simulated family structure and reporting conditions.

Accordingly, the revised analysis uses exact nonlinear inversion with $\kappa = 0.5$ as the central effect-size adjustment. The adjusted effect should be interpreted as an estimate on the directly diagnosed scale conditional on the specified attenuation model, rather than as an assumption-free transformation.
