## Supplementary_Note2 for "Population-scale burden analysis of rare damaging coding variants identifies novel risk genes for Alzheimer’s disease and related dementias and Parkinson’s disease and related disorders"

### Supplementary Note 2

Meta-analysis of REGENIE gene-burden results and integration of ADES summary statistics

Population-scale burden analysis of rare damaging coding variants

#### Overview

Two distinct meta-analytic objectives are relevant to the ADRD analyses:

1. estimation of a pooled gene-burden effect across cohorts analyzed using compatible REGENIE binary-trait burden models; and
2. combination of the statistical evidence from the REGENIE analyses with the publicly available ADES results from Holstege et al.

These objectives require different methods. The REGENIE cohorts provide approximately commensurate binary-trait log-odds effects after proxy-effect adjustment and can therefore be combined through conventional inverse-variance fixed-effects meta-analysis. The ADES summary statistics were generated using an additive summed-dosage ordinal logistic model. Their coefficients do not have the same interpretation or measurement unit as the REGENIE maximum-mask binary-trait coefficients. ADES is therefore incorporated through a signed-$Z$ evidence combination rather than by pooling its beta and standard error with the REGENIE coefficients.

This framework produces a pooled REGENIE odds ratio for effect-size interpretation and an ADES-inclusive combined $P$ value for cross-study evidence of association. ADES-inclusive pooled odds ratio were not computed, thus for ADRD analyses the tables and forest-plots throughout the manuscript reports OR_meta corresponding to the odds ratio of meta-analysis of REGENIE summary statistics along side the ADES odds ratio OR_ADES.

#### REGENIE burden model

For each cohort, phenotype, ancestry stratum, gene and deleteriousness mask, REGENIE Step 2 provided a burden coefficient

$$\widehat{\beta}_{ig},$$

standard error

$$s_{ig},$$

and two-sided association $P$ value

$$p_{ig},$$

where $i$ indexes the cohort and $g$ indexes the gene/mask pair.

The primary REGENIE burden mask used the maximum alternate-allele count across qualifying variants:

$$G_{i,\max} = \max_j G_{ij}.$$

This represents a dominant-like carrier model for the usual situation in which qualifying rare variants are heterozygous and participants carry no more than one qualifying allele. It does not assume that carrying two different damaging variants necessarily doubles the log-odds effect of carrying one.

The simulation in Supplementary Note 1 showed that maximum-allele and summed-dosage estimates were close under carrier frequencies of 0.5%-1%, because only approximately 0.20%-0.49% of carriers had more than one qualifying allele across scenario means. Nevertheless, the encodings estimate different parameters and were not treated as mathematically interchangeable.

### Standard errors for Firth-corrected REGENIE results

For binary traits, REGENIE reports likelihood-ratio-test $P$ values after Firth correction. Unless the `--firth-se` option is used, the default standard error may be derived from the unpenalized Hessian and may not reproduce the reported likelihood-ratio $P$ value.

For Firth-corrected rows, we therefore used a $P$-aligned standard error. When generated directly by REGENIE with `--firth-se`, this value was used as reported. For previously generated results without `--firth-se`, the equivalent normal-scale standard error was reconstructed from the coefficient and two-sided likelihood-ratio $P$ value as

$$z_{ig}^{(P)} = \Phi^{-1}\left(1 - \frac{p_{ig}}{2}\right),$$

and

$$s_{ig}^{(P)} = \frac{\left|\widehat{\beta}_{ig}\right|}{z_{ig}^{(P)}}. \tag{1}$$

For rows not requiring Firth correction, the ordinary REGENIE standard error was retained. Extremely small $P$ values were evaluated from the reported $-\log_{10} P$ value using numerically stable tail-probability calculations.

This step ensures that the coefficient, standard error and likelihood-ratio evidence supplied to the meta-analysis are internally aligned.

### Adjustment of combined-proxy REGENIE effects

For a combined-proxy cohort, let

$$w_i = \frac{N_{i,\mathrm{direct}}}{N_{i,\mathrm{direct}} + N_{i,\mathrm{FH-only}}}$$

denote the directly diagnosed/self-reported/clinical fraction among combined-proxy cases.

As derived in Supplementary Note 1, the combined-proxy effect satisfies

$$\beta_{M,i} = \log\left[w_i \exp(\beta_{D,i}) + (1 - w_i)\exp(\kappa\beta_{D,i})\right]. \tag{2}$$

The central analysis used

$$\kappa = 0.5.$$

For each combined-proxy cohort, the adjusted coefficient $\widetilde{\beta}_{ig}$ was obtained by solving

$$\widehat{\beta}_{ig} = \log\left[w_i \exp(\widetilde{\beta}_{ig}) + (1 - w_i)\exp(0.5\widetilde{\beta}_{ig})\right]. \tag{3}$$

For $\kappa = 0.5$, the closed-form solution is

$$\widetilde{\beta}_{ig} = 2\log\left[\frac{2\exp(\widehat{\beta}_{ig})}{\sqrt{(1-w_i)^2 + 4w_i\exp(\widehat{\beta}_{ig})} + (1-w_i)}\right]. \tag{4}$$

The derivative of the forward transformation is

$$f_i'(\beta) = \frac{w_i\exp(\beta) + 0.5(1-w_i)\exp(0.5\beta)}{w_i\exp(\beta) + (1-w_i)\exp(0.5\beta)}. \tag{5}$$

The adjusted standard error was

$$\widetilde{s}_{ig} = \frac{s_{ig}^{(P)}}{f_i'(\widetilde{\beta}_{ig})}. \tag{6}$$

For directly diagnosed cohorts,

$$w_i = 1,$$

so

$$\widetilde{\beta}_{ig} = \widehat{\beta}_{ig}$$

and

$$\widetilde{s}_{ig} = s_{ig}^{(P)}.$$

### Fixed-effects meta-analysis of REGENIE cohorts

Only cohorts analyzed under the compatible REGENIE maximum-mask binary-trait framework were included in the pooled effect-size meta-analysis.

For ADRD, these cohorts were UKB, AoU and ADSP. For PDRD, these cohorts were UKB, AoU and AMP-PDRD.

For gene/mask pair $g$, define the inverse-variance weight

$$W_{ig} = \frac{1}{\widetilde{s}_{ig}^2}. \tag{7}$$

The fixed-effects pooled coefficient was

$$\widehat{\beta}_{R,g} = \frac{\sum_i W_{ig}\widetilde{\beta}_{ig}}{\sum_i W_{ig}}. \tag{8}$$

Its standard error was

$$\mathrm{SE}_{R,g} = \left(\sum_i W_{ig}\right)^{-1/2}. \tag{9}$$

The pooled test statistic and two-sided $P$ value were

$$Z_{R,g} = \frac{\widehat{\beta}_{R,g}}{\mathrm{SE}_{R,g}}, \tag{10}$$

and

$$P_{R,g} = 2\Phi\left(-\left|Z_{R,g}\right|\right). \tag{11}$$

The pooled odds ratio and 95% confidence interval were

$$\mathrm{OR}_{R,g} = \exp(\widehat{\beta}_{R,g}), \tag{12}$$

and

$$\mathrm{CI}_{95\%,R,g} = \exp\left[\widehat{\beta}_{R,g} \pm 1.96\,\mathrm{SE}_{R,g}\right]. \tag{13}$$

These are the effect estimates reported in the main tables and forest plots.

### Heterogeneity among REGENIE cohorts

Cochran's heterogeneity statistic was

$$Q_g = \sum_i W_{ig}\left(\widetilde{\beta}_{ig} - \widehat{\beta}_{R,g}\right)^2. \tag{14}$$

Under the fixed-effect null of homogeneous cohort effects,

$$Q_g \sim \chi^2_{K_g-1},$$

where $K_g$ is the number of contributing REGENIE cohorts.

The heterogeneity $P$ value was

$$P_{\mathrm{het},g} = \Pr\left(\chi^2_{K_g-1} \geq Q_g\right), \tag{15}$$

and

$$I_g^2 = \max\left[0,\ \frac{Q_g - (K_g - 1)}{Q_g}\right] \times 100\%. \tag{16}$$

Heterogeneity statistics were calculated only among the compatible REGENIE coefficients. They were not calculated by treating the ADES ordinal coefficient as an additional log-odds estimate.

### Characteristics of the ADES summary statistics

The publicly available ADES stage 1 summary statistics from Holstege et al. contain, for each gene and deleteriousness mask, the burden-test likelihood-ratio $P$ value $P_{A,g}$, the additive ordinal logistic coefficient $\widehat{\beta}_{A,g}$, its reported standard error $s_{A,g}$, and the summed minor-allele dosage $\mathrm{cMAC}_{A,g}$ across contributing variants and participants.

The ADES model ordered the outcome as control, late-onset AD and early-onset AD and used an additive summed-dosage burden score. Its coefficient can be interpreted as a proportional-odds parameter incorporating both AD-versus-control and early-onset-versus-later-onset information.

In contrast, the REGENIE coefficient is a binary-trait log-odds effect per unit of a maximum-allele burden mask. Thus,

$$\widehat{\beta}_{A,g}$$

and

$$\widehat{\beta}_{R,g}$$

do not estimate the same numerical parameter.

Accordingly, we did not calculate

$$\frac{W_R\widehat{\beta}_{R,g} + W_A\widehat{\beta}_{A,g}}{W_R + W_A}$$

or report an ADES-inclusive pooled odds ratio.

The two analyses nevertheless test a common gene-level null hypothesis:

$$H_{0,g} : \text{qualifying rare damaging variation in gene/mask } g \text{ is not associated with disease status.}$$

Their signed standardized evidence can therefore be combined without requiring the beta coefficients to have the same units.

### Conversion to signed $Z$ statistics

The REGENIE meta-analysis statistic was calculated from its two-sided $P$ value and pooled direction:

$$Z^*_{R,g} = \operatorname{sign}(\widehat{\beta}_{R,g})\Phi^{-1}\left(1 - \frac{P_{R,g}}{2}\right). \tag{17}$$

The ADES statistic was calculated analogously:

$$Z^*_{A,g} = \operatorname{sign}(\widehat{\beta}_{A,g})\Phi^{-1}\left(1 - \frac{P_{A,g}}{2}\right). \tag{18}$$

Using the likelihood-ratio $P$ values in Equations (17) and (18) avoids requiring the ADES ordinal beta/SE pair or the REGENIE Firth beta/SE pair to define exactly the same Wald statistic.

Positive values denote risk-increasing rare-variant burden, and negative values denote risk-decreasing burden.

### Information weights

For a binary case-control source with $n_1$ cases and $n_0$ controls, the effective sample size was defined as

$$N_{\text{eff}} = \frac{4n_1n_0}{n_1 + n_0} = \frac{4}{1/n_1 + 1/n_0}. \tag{19}$$

For the REGENIE meta-analysis,

$$N_{\text{eff},R} = \sum_{i \in R} N_{\text{eff},i}, \tag{20}$$

where the sum is across independent contributing REGENIE cohorts.

For ADES, early-onset and late-onset AD cases were combined for the sole purpose of defining the fixed information weight:

$$n_{1,A} = n_{\mathrm{EOAD}} + n_{\mathrm{LOAD}},$$

and

$$N_{\mathrm{eff},A} = \frac{4n_{1,A}n_{0,A}}{n_{1,A} + n_{0,A}}. \tag{21}$$

This weighting step does not transform the ADES ordinal coefficient into a binary coefficient. It supplies a phenotype-count-based weight to the standardized evidence combination.

The signed-$Z$ weights were

$$u_R = \sqrt{N_{\mathrm{eff},R}}$$

and

$$u_A = \sqrt{N_{\mathrm{eff},A}}. \tag{22}$$

Weights were determined solely from sample counts and were fixed independently of the observed gene-level association results.

### ADES-inclusive meta-analysis

The combined statistic was calculated as

$$Z_{A+R,g} = \frac{u_R Z^*_{R,g} + u_A Z^*_{A,g}}{\sqrt{u_R^2 + u_A^2}}. \tag{23}$$

Under the global null,

$$Z_{A+R,g} \sim N(0,1),$$

and the ADES-inclusive two-sided $P$ value was

$$P_{A+R,g} = 2\Phi\left(-\left|Z_{A+R,g}\right|\right). \tag{24}$$

Equation (24) is the ADES-inclusive significance value reported for each AD/ADRD gene/mask pair with eligible ADES evidence. When ADES evidence was unavailable, the pooled REGENIE significance value was retained. The ADES-inclusive result is an evidence-combination $P$ value and does not define a pooled ADES-plus-REGENIE odds ratio.

#### Study-wide significance

The final evidence statistic was computed for every eligible gene/mask pair, rather than restricting evidence combination to genes selected from the REGENIE analysis. This avoids inflation arising from that selection step. For AD, the final statistic was the ADES-inclusive statistic when eligible ADES evidence was available and the pooled REGENIE statistic otherwise. For PD, it was the pooled REGENIE statistic.

Let

$$N_{\text{genes}}^{\text{AD}} = 18{,}263$$

denote the number of genes with at least one retained mask in the primary all-ancestry AD-combined-proxy analysis, and let

$$m_{\text{eff}}^{A+R} \simeq 2.200834885$$

denote the effective number of tests across the four nested deleteriousness masks. The latter was estimated from the correlation structure of the final signed evidence statistics using the prespecified eigenvalue-based procedure, including REGENIE fallback statistics where ADES evidence was unavailable.

Under the prespecified genes-times-effective-masks correction, the estimated total effective number of gene/mask tests was

$$M_{\text{eff}}^{A+R} = N_{\text{genes}}^{\text{AD}} m_{\text{eff}}^{A+R} \simeq 40{,}193.85.$$

The AD-specific study-wide threshold was therefore

$$\alpha_{\text{SW}}^{A+R} = \frac{0.05}{M_{\text{eff}}^{A+R}} \simeq 1.24397148 \times 10^{-6}. \tag{29}$$

The analogous calculation for the primary all-ancestry PD-combined-proxy analysis used

$$N_{\text{genes}}^{\text{PD}} = 18{,}280, \qquad m_{\text{eff}}^{\text{PD}} \simeq 2.265816371,$$

yielding

$$M_{\text{eff}}^{\text{PD}} = N_{\text{genes}}^{\text{PD}} m_{\text{eff}}^{\text{PD}} \simeq 41{,}419.12$$

and

$$\alpha_{\text{SW}}^{\text{PD}} = \frac{0.05}{M_{\text{eff}}^{\text{PD}}} \simeq 1.20717186 \times 10^{-6}.$$

These gene counts refer to distinct genes with at least one retained mask in the corresponding primary analysis, not to the number of gene/mask rows or the union of genes across all analyses. The thresholds were recomputed from the final statistics for each primary analysis rather than carried over from earlier analyses.

For a common reporting threshold across the two primary disease analyses, we took the smaller of their separately computed thresholds,

$$\alpha_{\text{common}} = \min\left\{\alpha_{\text{SW}}^{A+R}, \alpha_{\text{SW}}^{\text{PD}}\right\} \simeq 1.20717186 \times 10^{-6},$$

and adopted the slightly more stringent cutoff

$$\alpha_{\text{SW}} = 1.20 \times 10^{-6}$$

by conservative downward rounding.

A gene/mask pair was classified as study-wide significant in a primary analysis when

$$P_{\text{overall},g} < \alpha_{\text{SW}}, \tag{30}$$

where $P_{\text{overall},g}$ denotes the final AD evidence $P$ value (ADES-inclusive when available, otherwise REGENIE-only) or the REGENIE-only PD meta-analysis $P$ value, as appropriate. For an AD association interpreted as shared risk across REGENIE and ADES, concordant effect directions were additionally required when ADES evidence was available. This directional requirement concerned the shared-risk interpretation and did not restrict the eligible test universe used for evidence combination or threshold estimation.

The primary discovery analyses were the all-ancestry AD-combined-proxy and PD-combined-proxy analyses. European-ancestry analyses were secondary ancestry analyses; AD-direct and PD-direct analyses were nested phenotype-sensitivity analyses; and all ADRD/PDRD analyses were diagnostic-sensitivity comparisons. The same numerical cutoff was shown as a benchmark in these secondary and sensitivity analyses, but signals arising only in those analyses were explicitly labeled as such and were not treated as additional primary discoveries or as having primary-family-wise error control. The correction was defined separately for the two primary disease families and was not a joint correction across all 16 analyses.

### Conclusion

Standard inverse-variance fixed-effects meta-analysis was used for the REGENIE cohorts after placing combined-proxy effects on a common approximate directly diagnosed scale and using Firth-compatible standard errors. The pooled REGENIE odds ratio is the principal effect-size estimate.

The ADES ordinal coefficient and the REGENIE binary-trait coefficient are not directly commensurate and are therefore not pooled through beta/SE inverse-variance weighting. Their statistical evidence is instead combined through signed standardized statistics. Under the shared global null, and assuming independence and appropriate null calibration of the source-specific signed statistics, this produces an appropriately calibrated ADES-inclusive significance test. When eligible ADES evidence is unavailable, the pooled REGENIE significance value is retained.

This separation preserves the REGENIE maximum-allele burden model, provides an interpretable pooled effect among compatible cohorts, uses the ADES results as large-scale external AD evidence, and avoids assigning an unjustified common odds-ratio interpretation to coefficients generated under different burden and outcome models.
