## Supplementary_Fig8 for "Population-scale burden analysis of rare damaging coding variants identifies novel risk genes for Alzheimer’s disease and related dementias and Parkinson’s disease and related disorders"

### AD primary/secondary definitions with ADRD sensitivity − GRN (ALL)

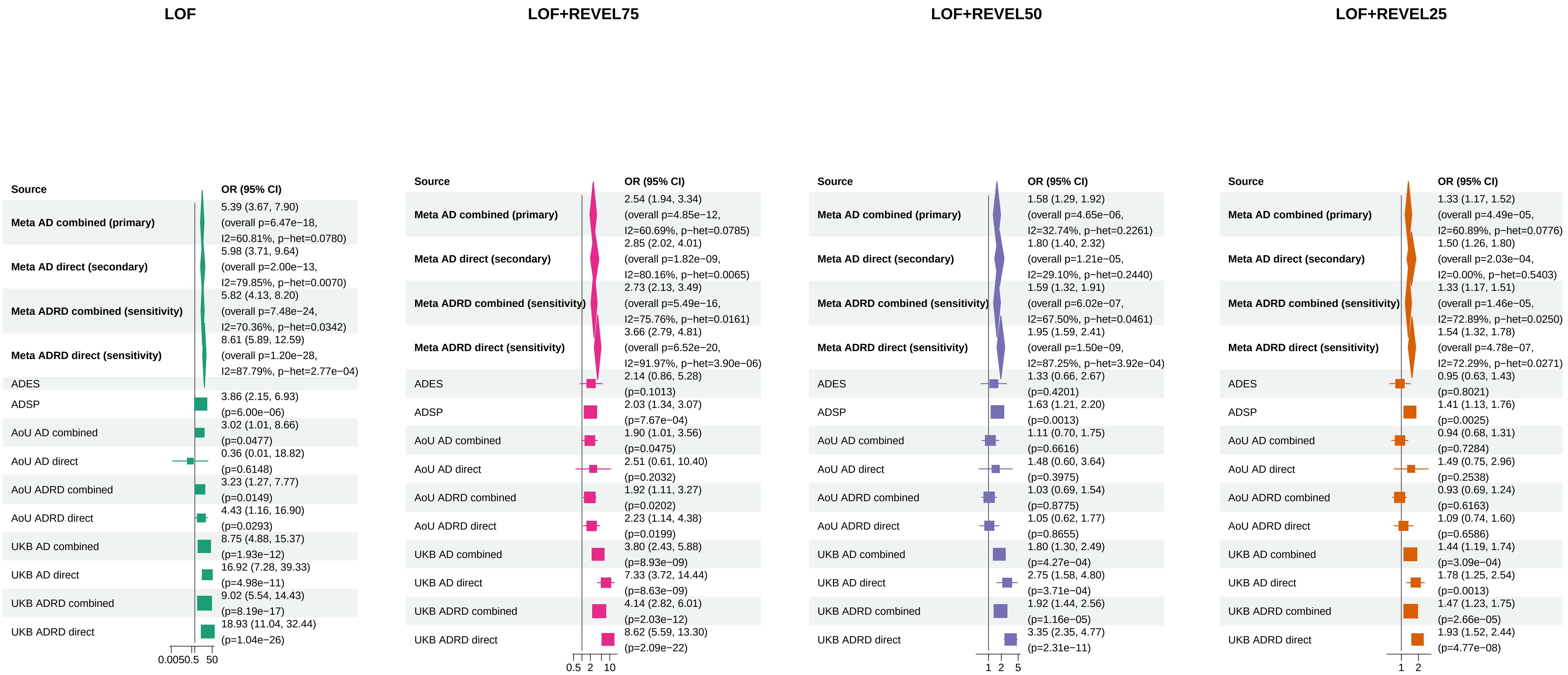


Meta OR/CI values are pooled REGENIE effects; overall P values use ADES+REGENIE signed−Z evidence when the exact ADES mask was available, otherwise REGENIE.

#### AD primary/secondary definitions with ADRD sensitivity − GRN (EUR)

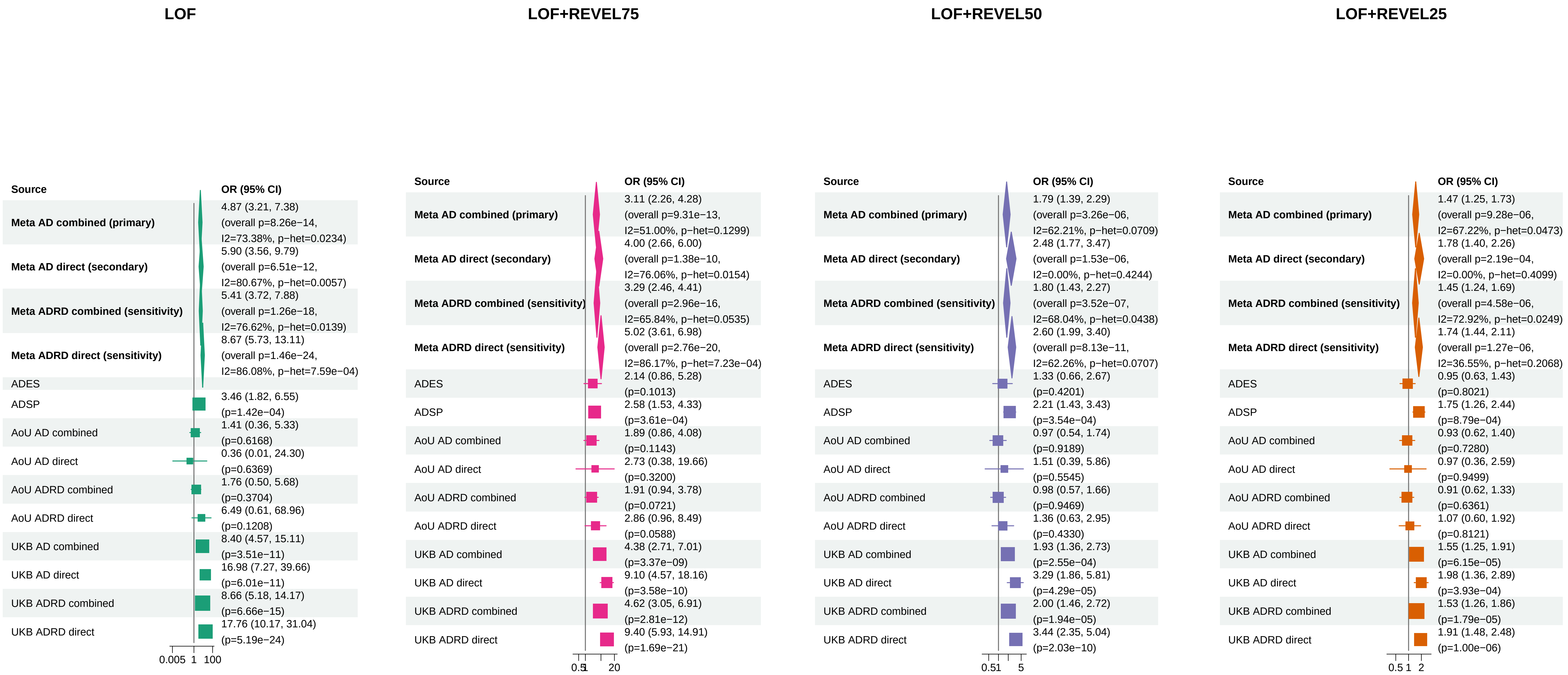


Meta OR/CI values are pooled REGENIE effects; overall P values use ADES+REGENIE signed−Z evidence when the exact ADES mask was available, otherwise REGENIE.

#### AD primary/secondary definitions with ADRD sensitivity − TBK1 (ALL)

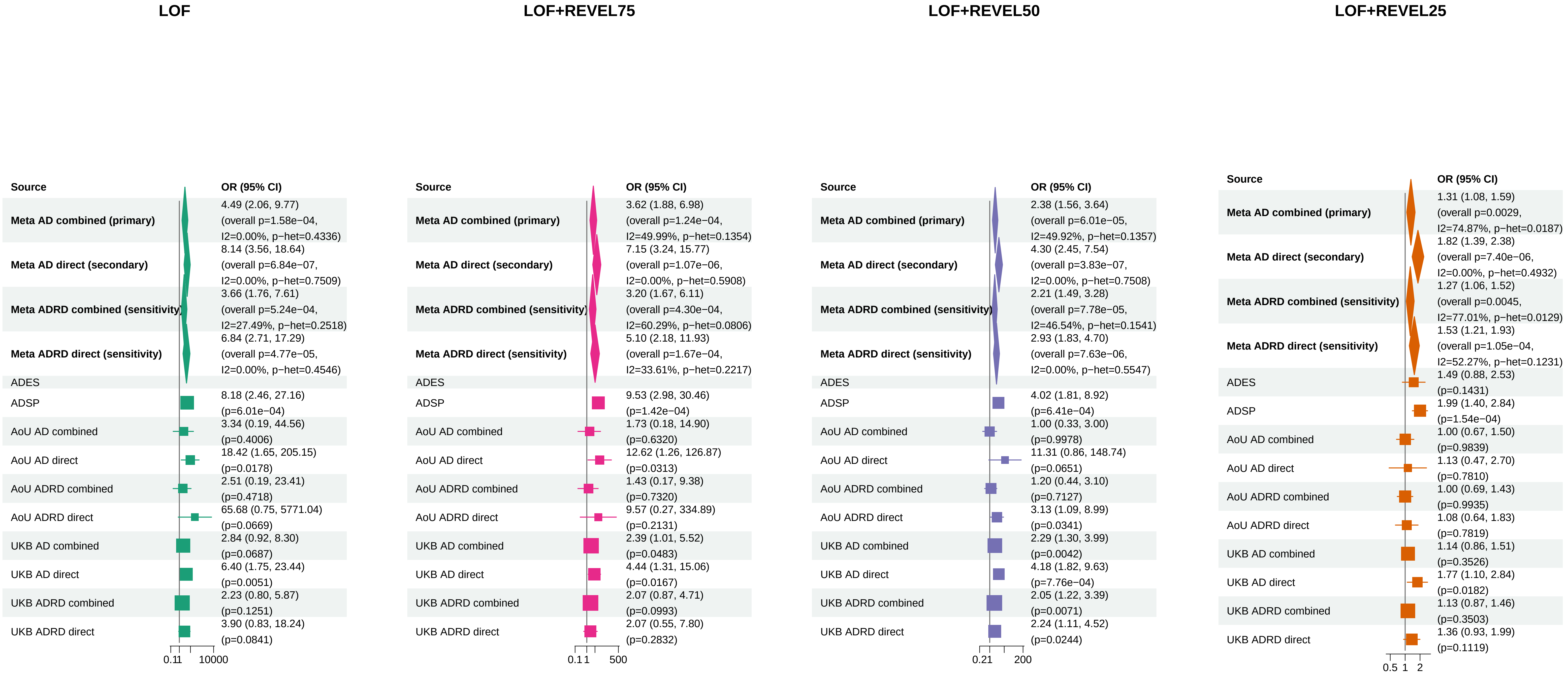


Meta OR/CI values are pooled REGENIE effects; overall P values use ADES+REGENIE signed−Z evidence when the exact ADES mask was available, otherwise REGENIE.

### AD primary/secondary definitions with ADRD sensitivity − TBK1 (EUR)

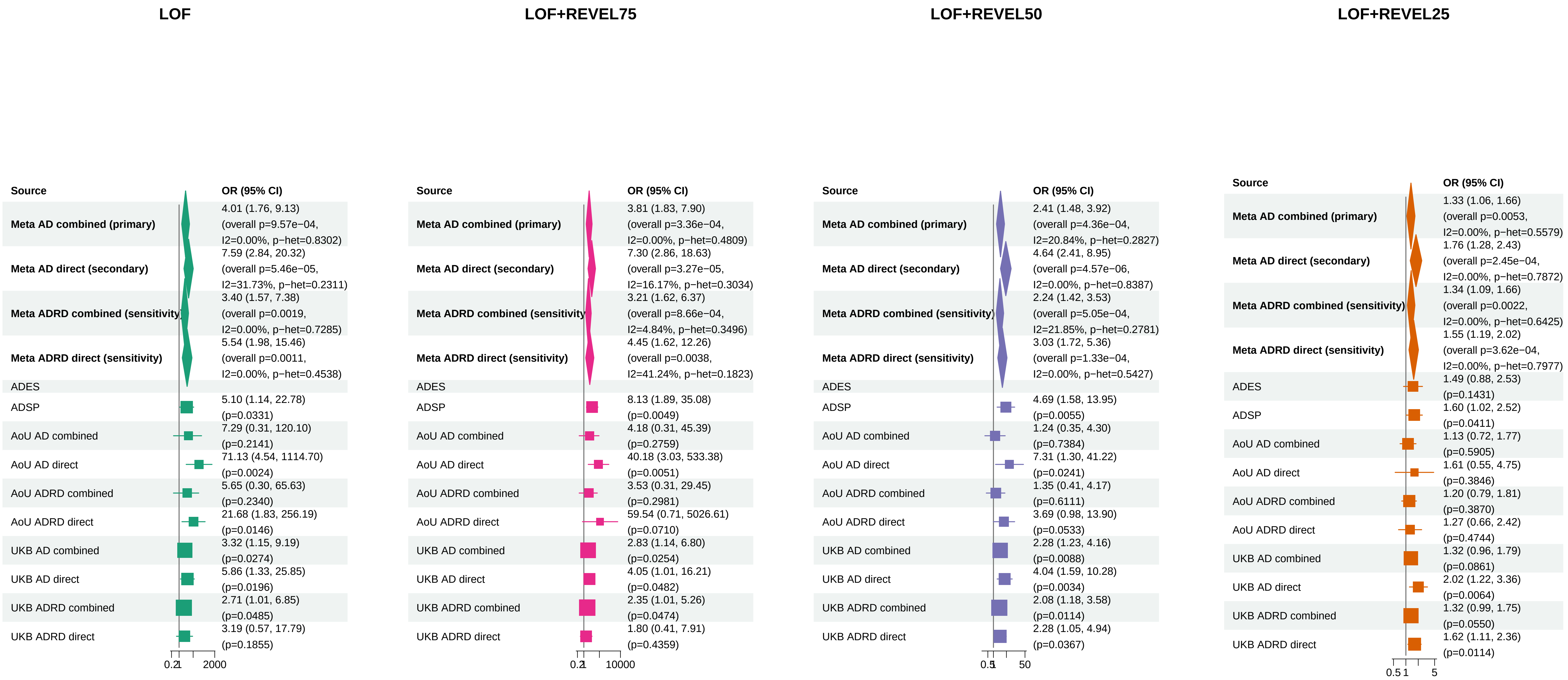


Meta OR/CI values are pooled REGENIE effects; overall P values use ADES+REGENIE signed−Z evidence when the exact ADES mask was available, otherwise REGENIE.

#### AD primary/secondary definitions with ADRD sensitivity − VCP (ALL)

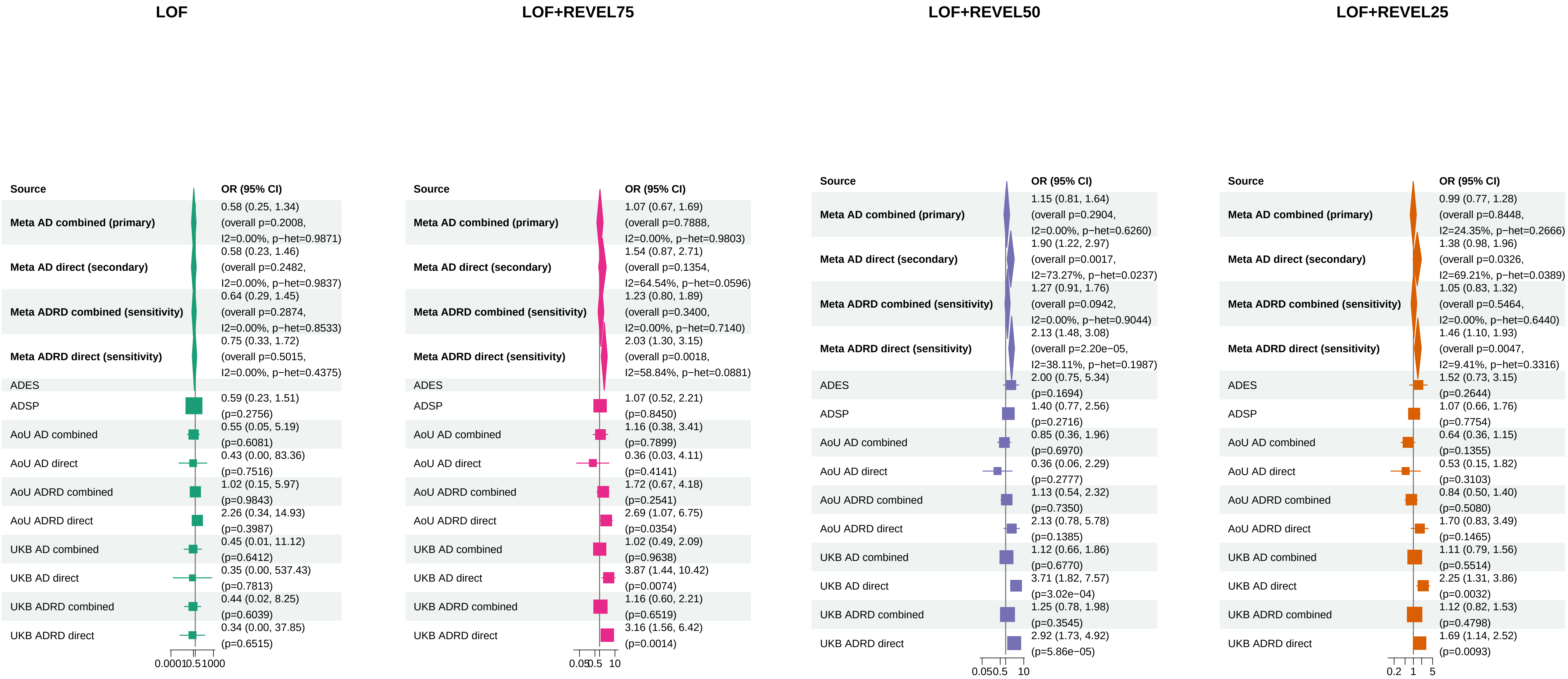


Meta OR/CI values are pooled REGENIE effects; overall P values use ADES+REGENIE signed−Z evidence when the exact ADES mask was available, otherwise REGENIE.

### AD primary/secondary definitions with ADRD sensitivity − VCP (EUR)

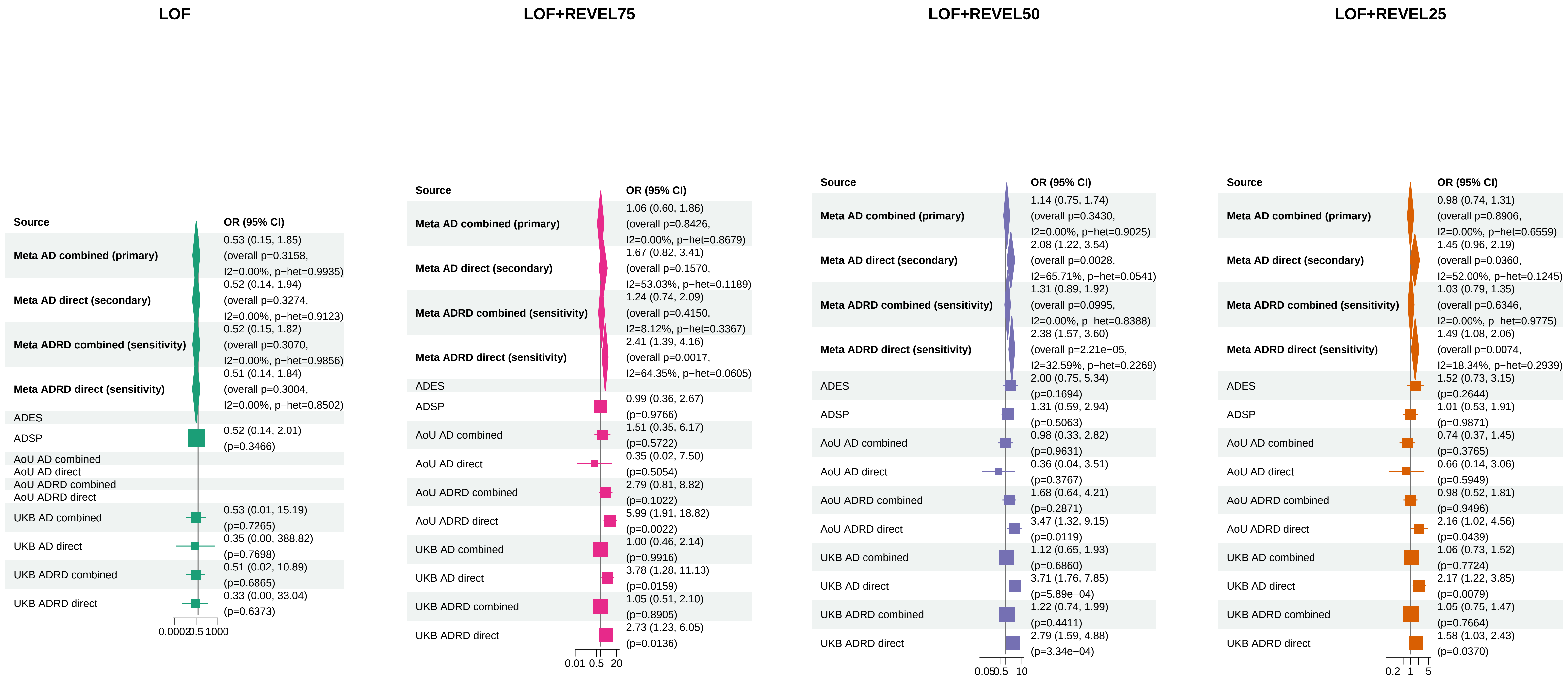


Meta OR/CI values are pooled REGENIE effects; overall P values use ADES+REGENIE signed−Z evidence when the exact ADES mask was available, otherwise REGENIE.
