## Supplementary_Fig12 for "Population-scale burden analysis of rare damaging coding variants identifies novel risk genes for Alzheimer’s disease and related dementias and Parkinson’s disease and related disorders"

### PD primary/secondary definitions with PDRD sensitivity − LRRK2 − LOF+REVEL75 (EUR)

| Source | OR (95% CI) |
| --- | --- |
| **Meta PD combined (primary)** | 3.05 (2.57, 3.61) (p=4.90e−38; I2=76.91%, p−het=0.0132) |
| **Meta PD direct (secondary)** | 3.01 (2.47, 3.65) (p=1.80e−28; I2=77.22%, p−het=0.0124) |
| **Meta PDRD combined (sensitivity)** | 2.51 (2.13, 2.95) (p=5.69e−28; I2=64.68%, p−het=0.0589) |
| **Meta PDRD direct (sensitivity)** | 2.24 (1.86, 2.70) (p=3.26e−17; I2=75.30%, p−het=0.0175) |
| AMP−PD PD combined | 3.52 (2.80, 4.43) (p=2.79e−26) |
| AMP−PD PD direct | 3.25 (2.56, 4.14) (p=8.76e−22) |
| AMP−PD PDRD combined | 2.49 (1.99, 3.10) (p=9.37e−16) |
| AMP−PD PDRD direct | 2.11 (1.67, 2.66) (p=2.48e−10) |
| AoU PD combined | 3.58 (2.43, 5.19) (p=4.16e−10) |
| AoU PD direct | 3.77 (2.46, 5.76) (p=9.39e−10) |
| AoU PDRD combined | 3.54 (2.43, 5.10) (p=2.29e−10) |
| AoU PDRD direct | 3.59 (2.38, 5.41) (p=1.10e−09) |
| UKB PD combined | 1.97 (1.40, 2.75) (p=1.10e−04) |
| UKB PD direct | 1.45 (0.85, 2.46) (p=0.1745) |
| UKB PDRD combined | 1.94 (1.39, 2.69) (p=1.10e−04) |
| UKB PDRD direct | 1.45 (0.88, 2.41) (p=0.1480) |

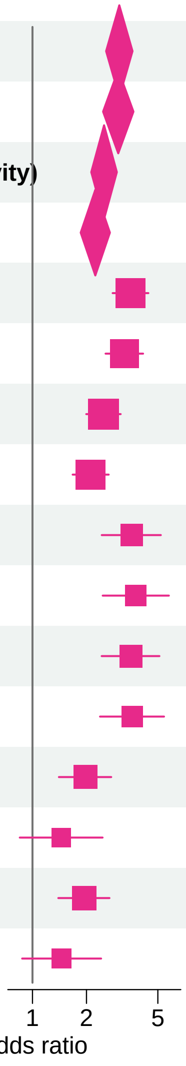


Meta OR/CI and P values are from the exact−kappa=0.5 pooled REGENIE fixed−effects analysis.

#### PD primary/secondary definitions with PDRD sensitivity − GBA1 − LOF+REVEL25 (EUR)

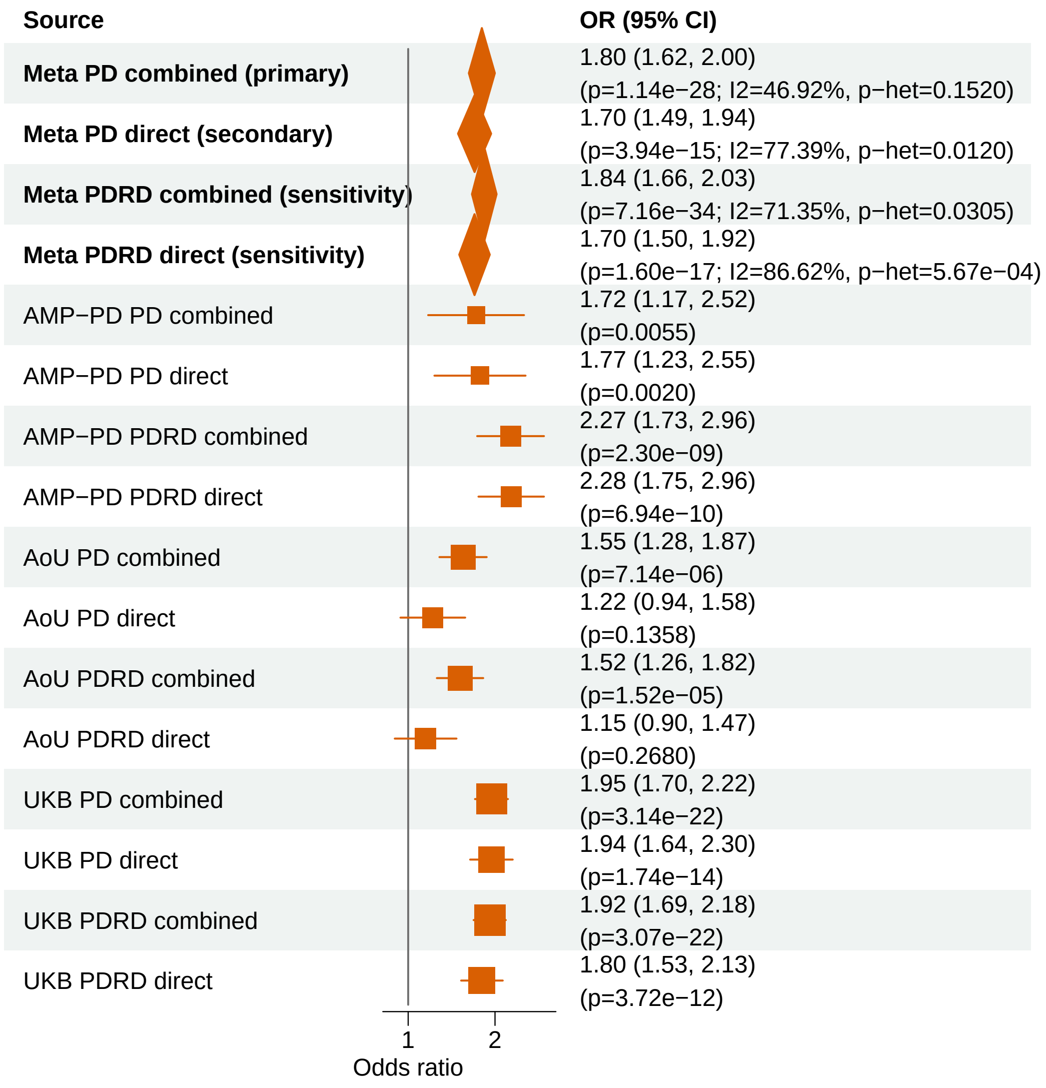


Meta OR/CI and P values are from the exact−kappa=0.5 pooled REGENIE fixed−effects analysis.

#### PD primary/secondary definitions with PDRD sensitivity − GBA1 − LOF+REVEL50 (EUR)

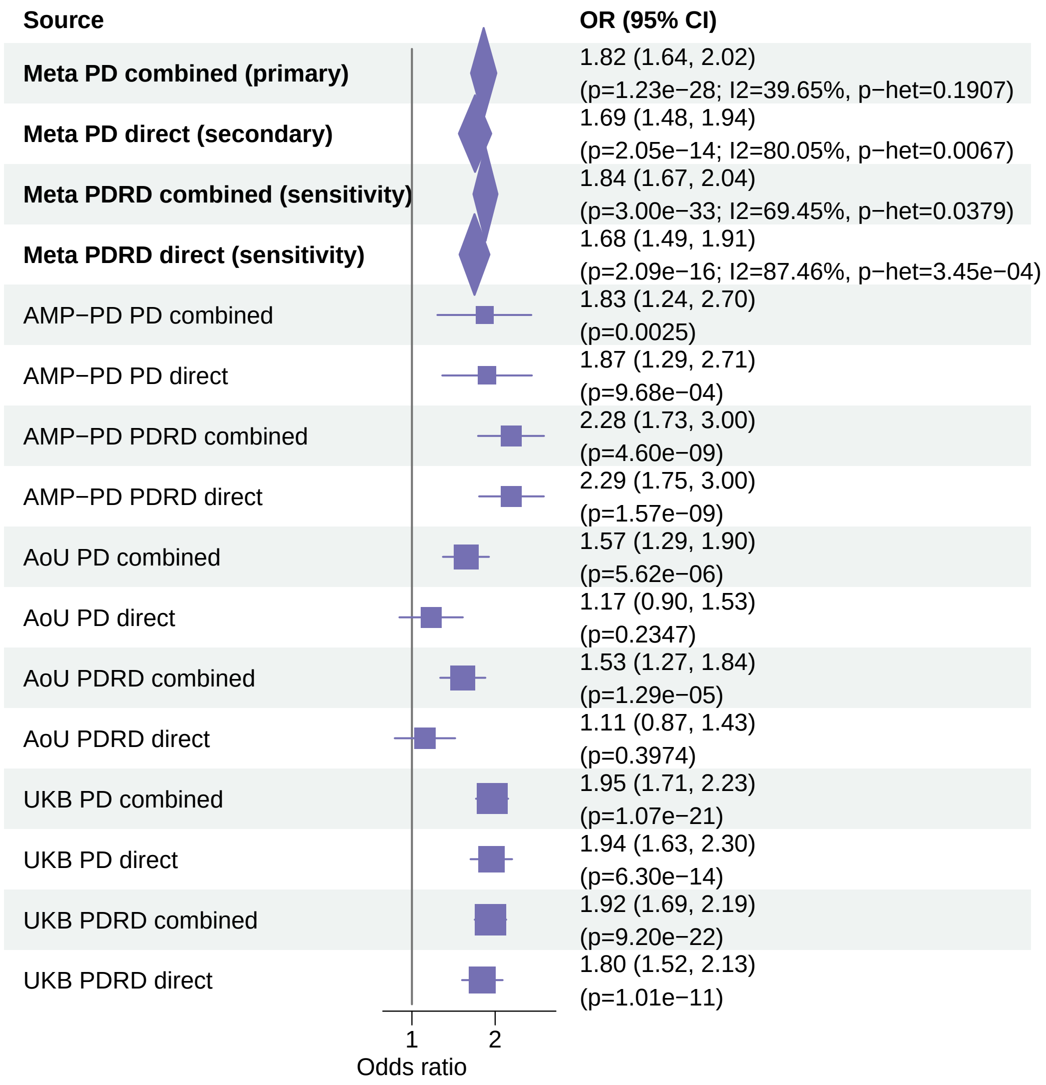


Meta OR/CI and P values are from the exact−kappa=0.5 pooled REGENIE fixed−effects analysis.

#### PD primary/secondary definitions with PDRD sensitivity − LRRK2 − LOF+REVEL50 (EUR)

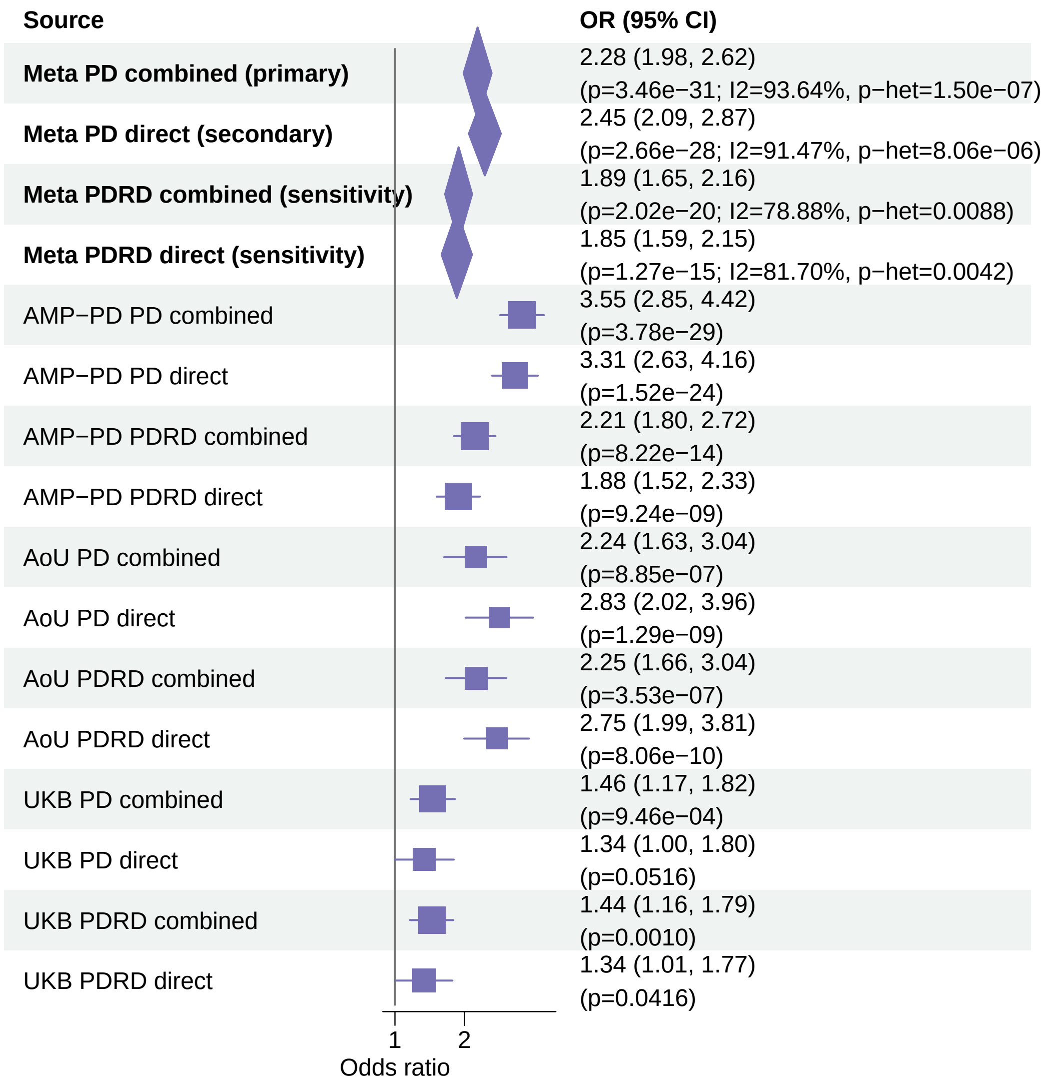


Meta OR/CI and P values are from the exact−kappa=0.5 pooled REGENIE fixed−effects analysis.

### PD primary/secondary definitions with PDRD sensitivity − GBA1 − LOF+REVEL75 (EUR)

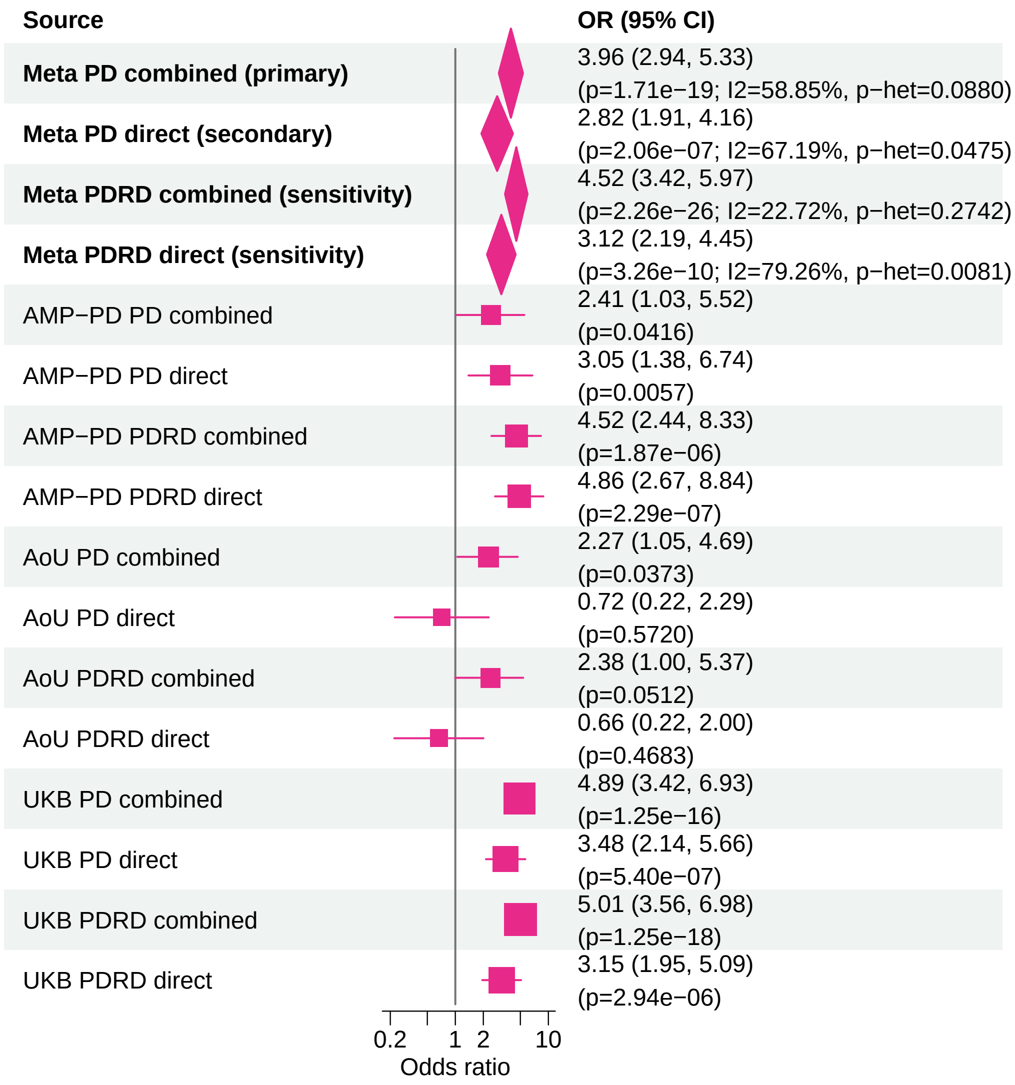


Meta OR/CI and P values are from the exact−kappa=0.5 pooled REGENIE fixed−effects analysis.

#### PD primary/secondary definitions with PDRD sensitivity − LRRK2 − LOF+REVEL25 (EUR)

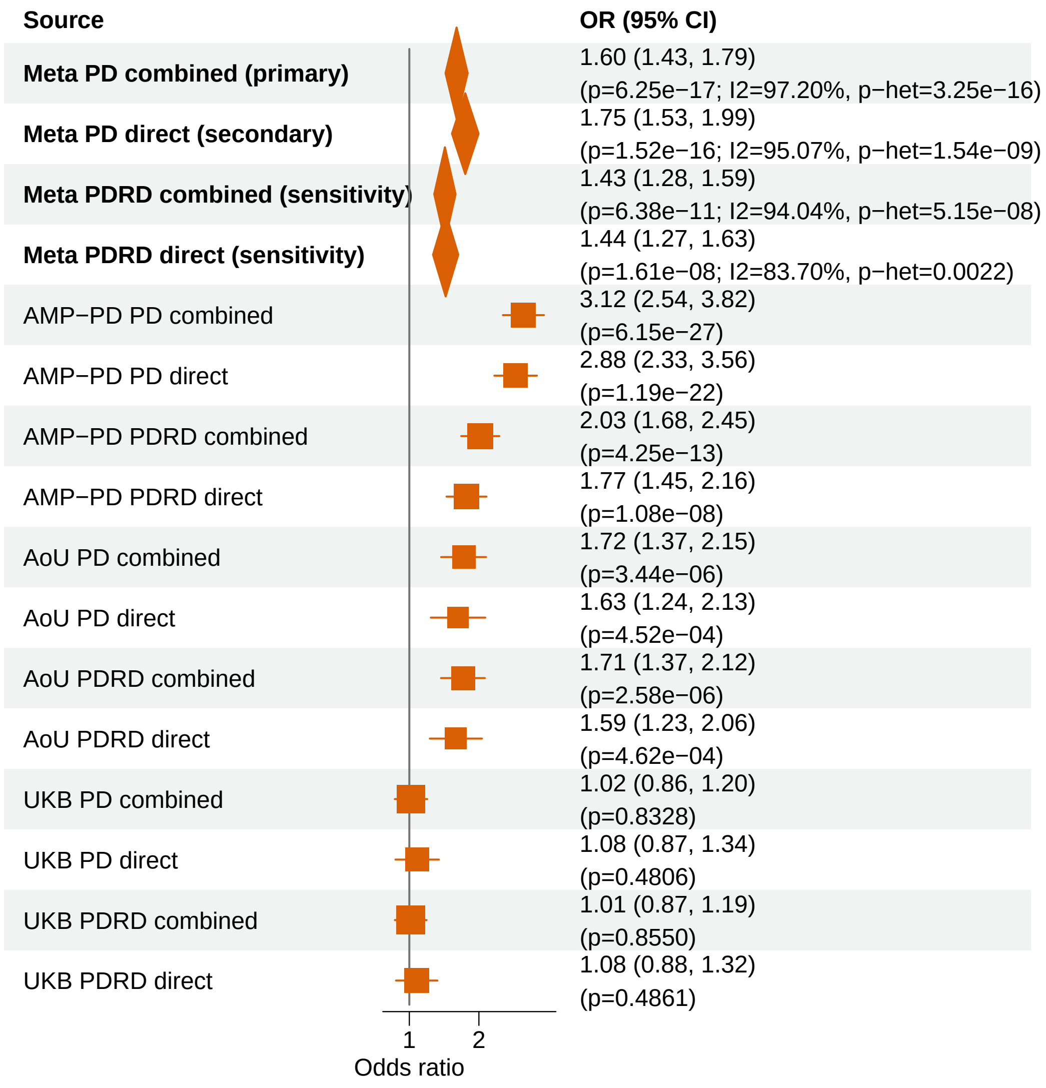


Meta OR/CI and P values are from the exact−kappa=0.5 pooled REGENIE fixed−effects analysis.

#### PD primary/secondary definitions with PDRD sensitivity − GBA1 − LOF (EUR)

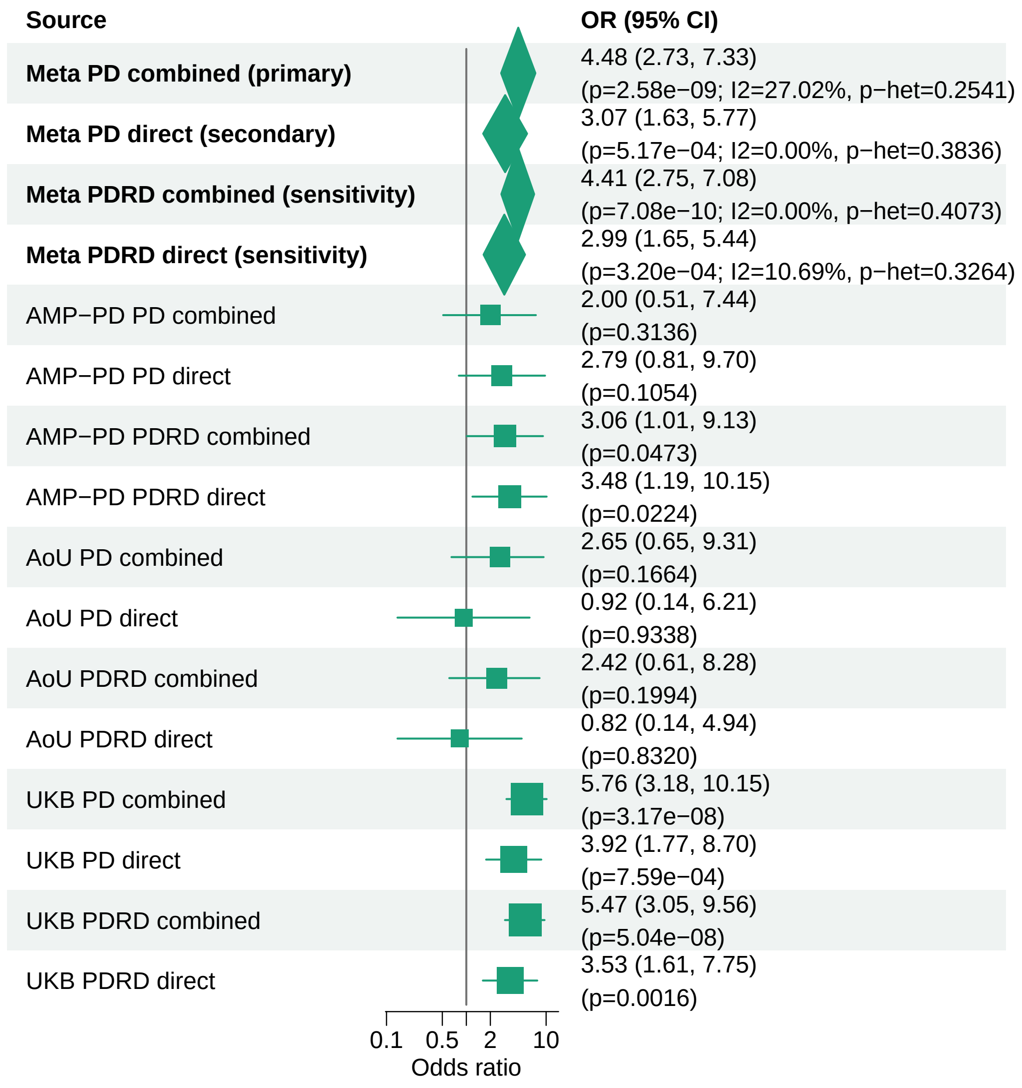


Meta OR/CI and P values are from the exact−kappa=0.5 pooled REGENIE fixed−effects analysis.

#### PD primary/secondary definitions with PDRD sensitivity − ANKRD27 − LOF+REVEL25 (EUR)

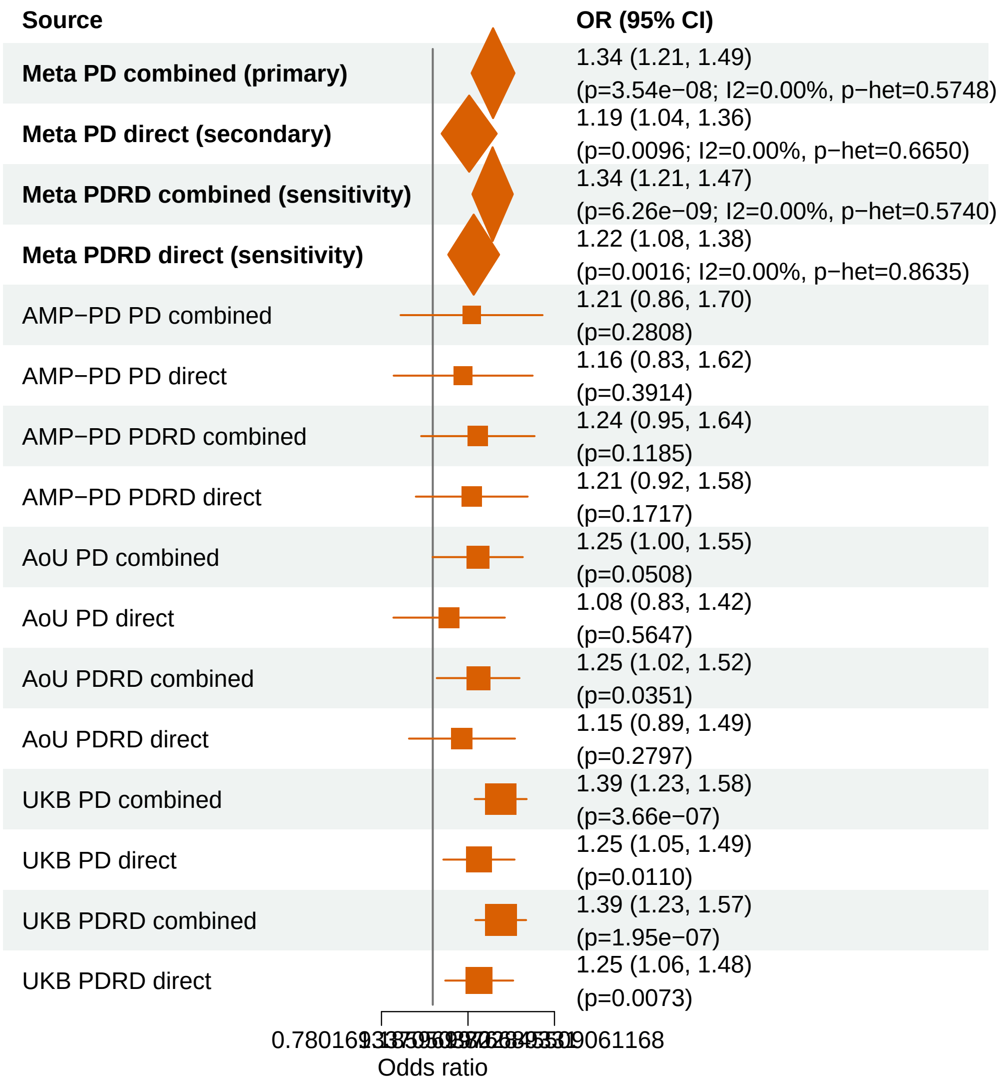


Meta OR/CI and P values are from the exact−kappa=0.5 pooled REGENIE fixed−effects analysis.

### PD primary/secondary definitions with PDRD sensitivity − CCL7 − LOF+REVEL50 (EUR)

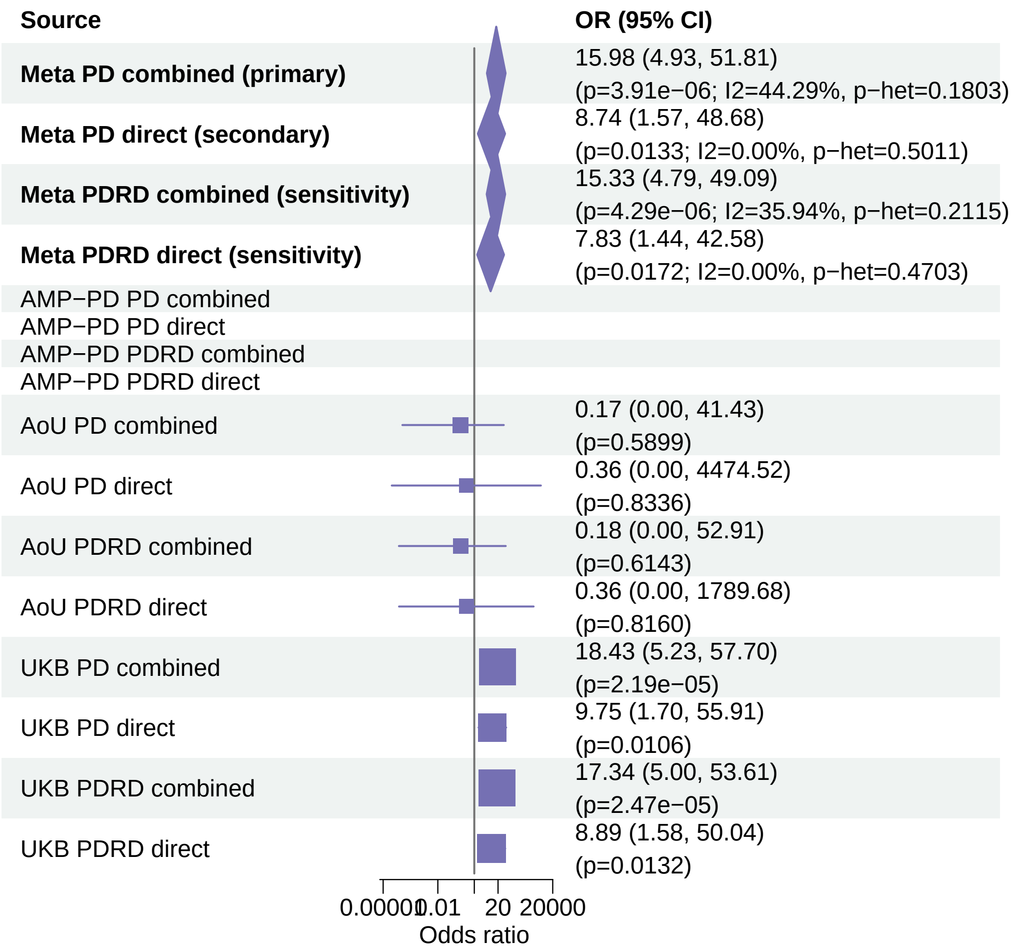


Meta OR/CI and P values are from the exact−kappa=0.5 pooled REGENIE fixed−effects analysis.

#### PD primary/secondary definitions with PDRD sensitivity − PANX2 − LOF+REVEL50 (EUR)

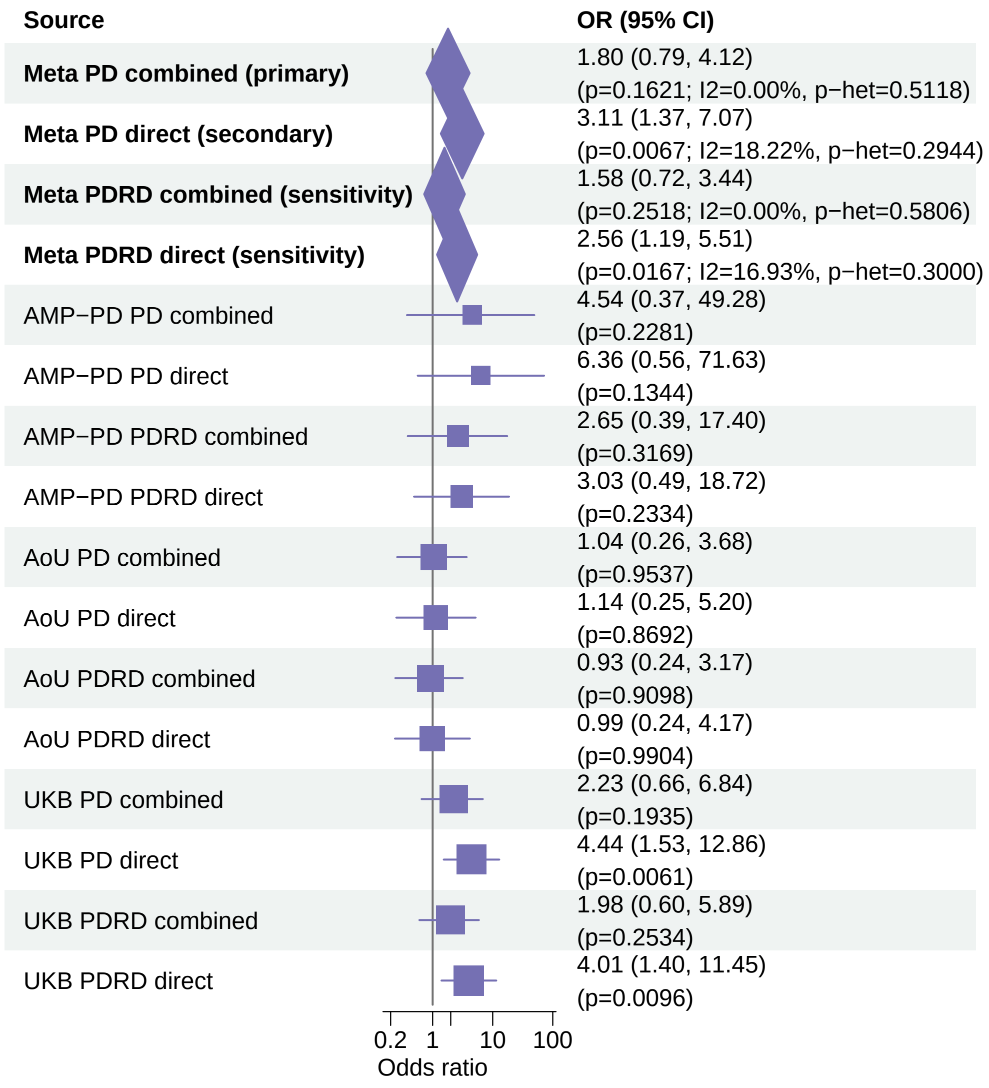


Meta OR/CI and P values are from the exact−kappa=0.5 pooled REGENIE fixed−effects analysis.

#### PD primary/secondary definitions with PDRD sensitivity − USP19 − LOF+REVEL75 (EUR)

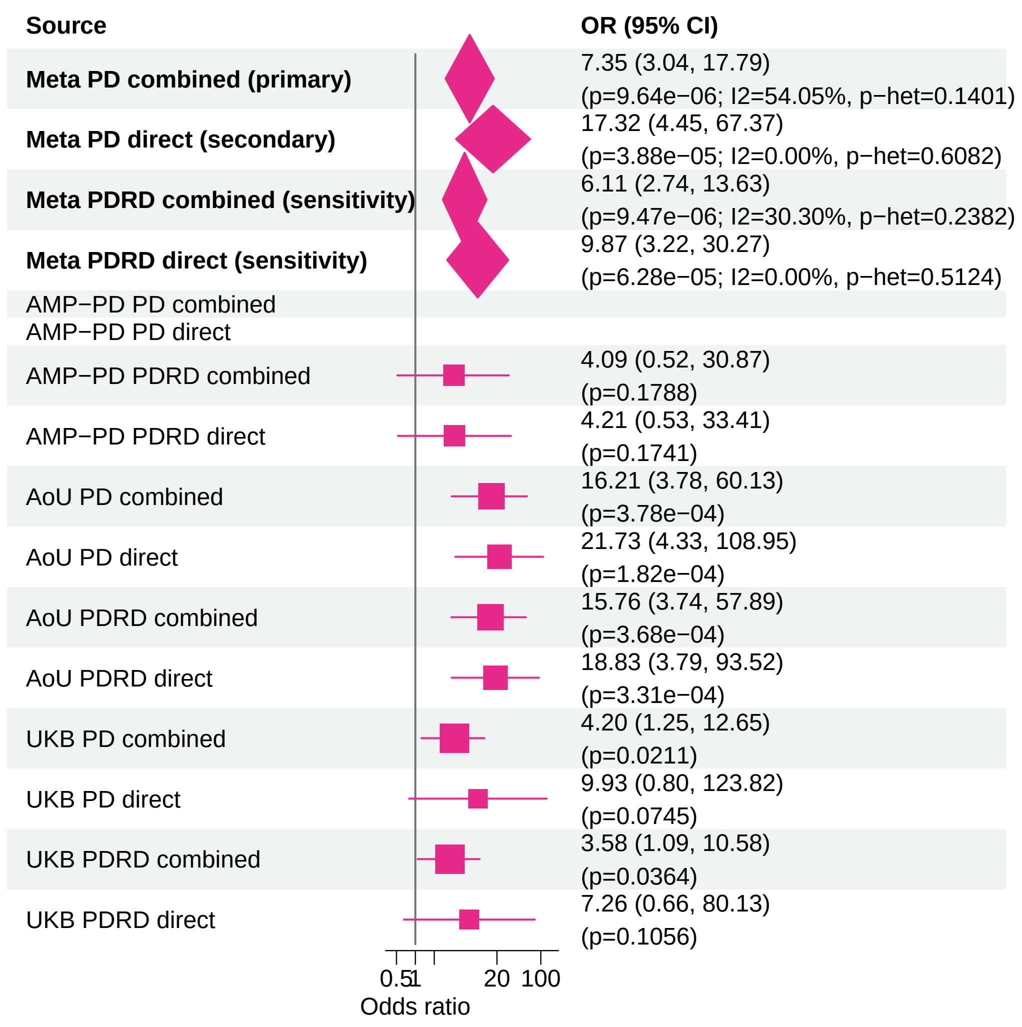


Meta OR/CI and P values are from the exact−kappa=0.5 pooled REGENIE fixed−effects analysis.

### PD primary/secondary definitions with PDRD sensitivity − USP19 − LOF (EUR)

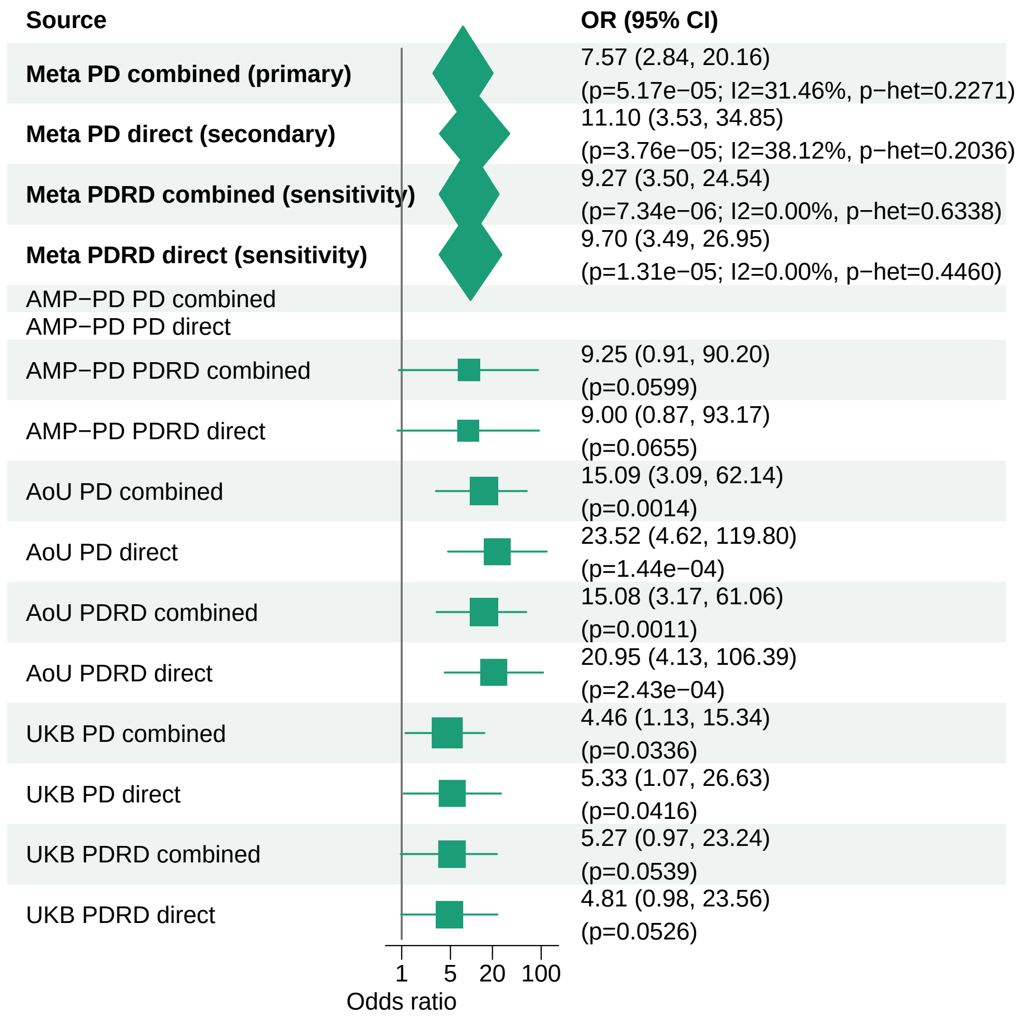


Meta OR/CI and P values are from the exact−kappa=0.5 pooled REGENIE fixed−effects analysis.

#### PD primary/secondary definitions with PDRD sensitivity − ZSCAN25 − LOF (EUR)

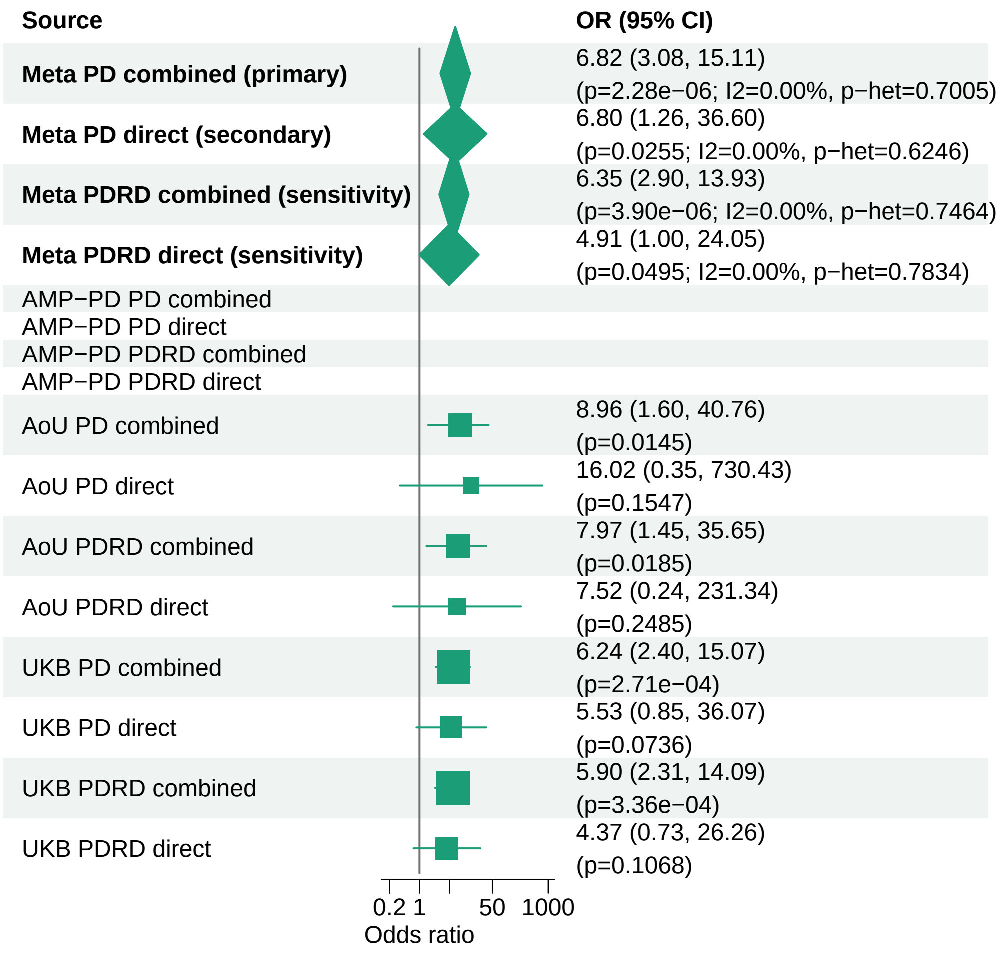


Meta OR/CI and P values are from the exact−kappa=0.5 pooled REGENIE fixed−effects analysis.

### PD primary/secondary definitions with PDRD sensitivity − BNIP3 − LOF (EUR)

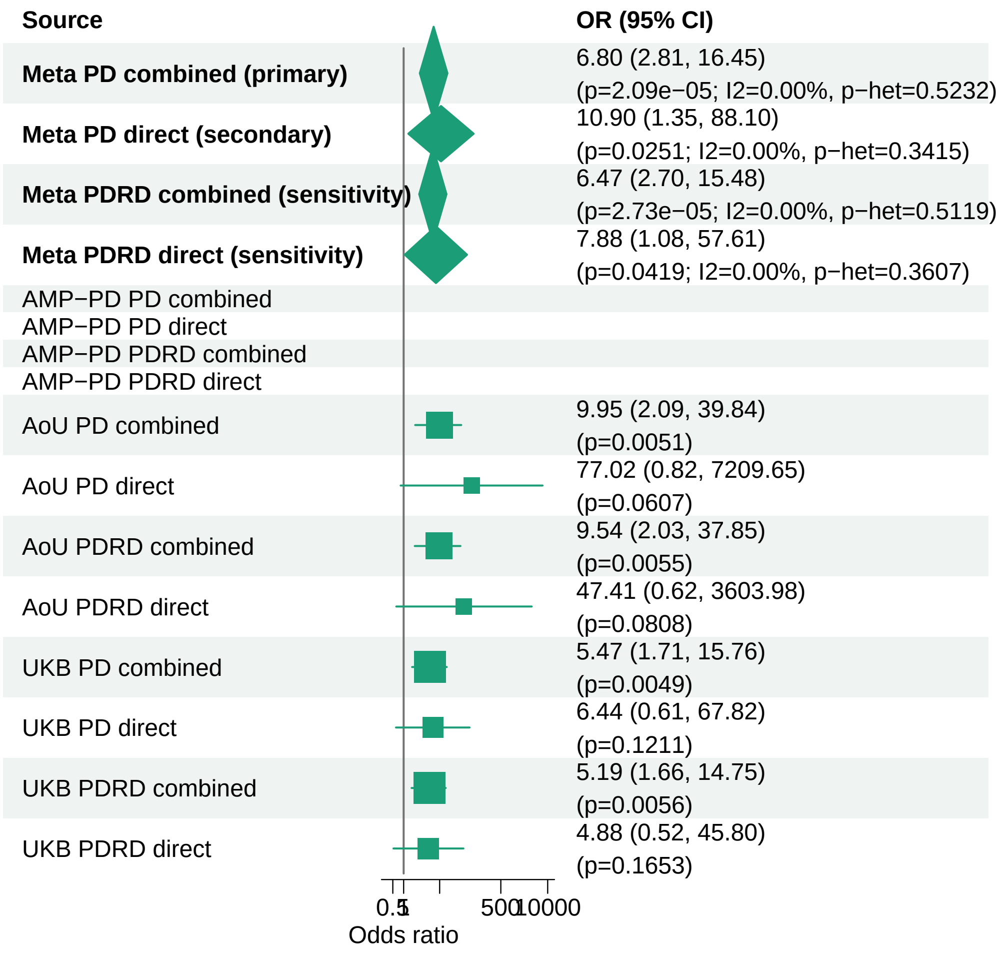


Meta OR/CI and P values are from the exact−kappa=0.5 pooled REGENIE fixed−effects analysis.

### PD primary/secondary definitions with PDRD sensitivity − SOBP − LOF+REVEL50 (EUR)

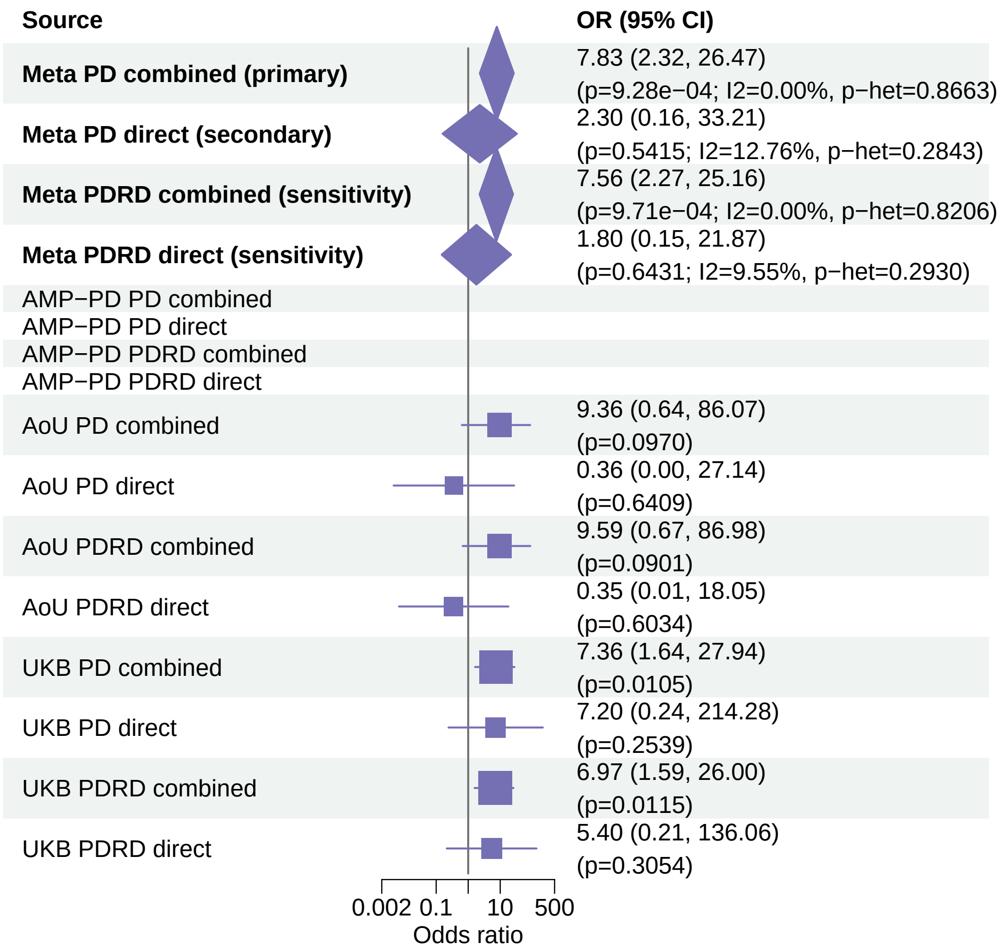


Meta OR/CI and P values are from the exact−kappa=0.5 pooled REGENIE fixed−effects analysis.

### PD primary/secondary definitions with PDRD sensitivity − KANSL3 − LOF (EUR)

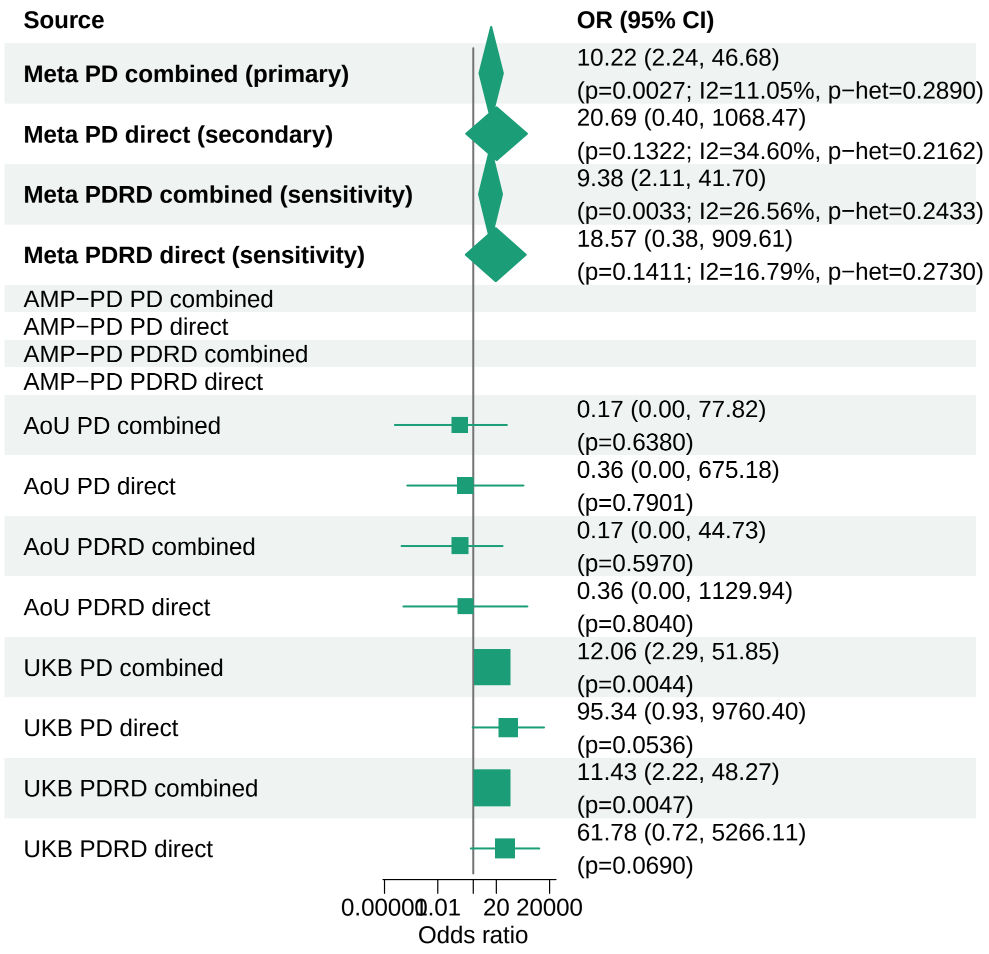


Meta OR/CI and P values are from the exact−kappa=0.5 pooled REGENIE fixed−effects analysis.

### PD primary/secondary definitions with PDRD sensitivity − PANX2 − LOF (EUR)

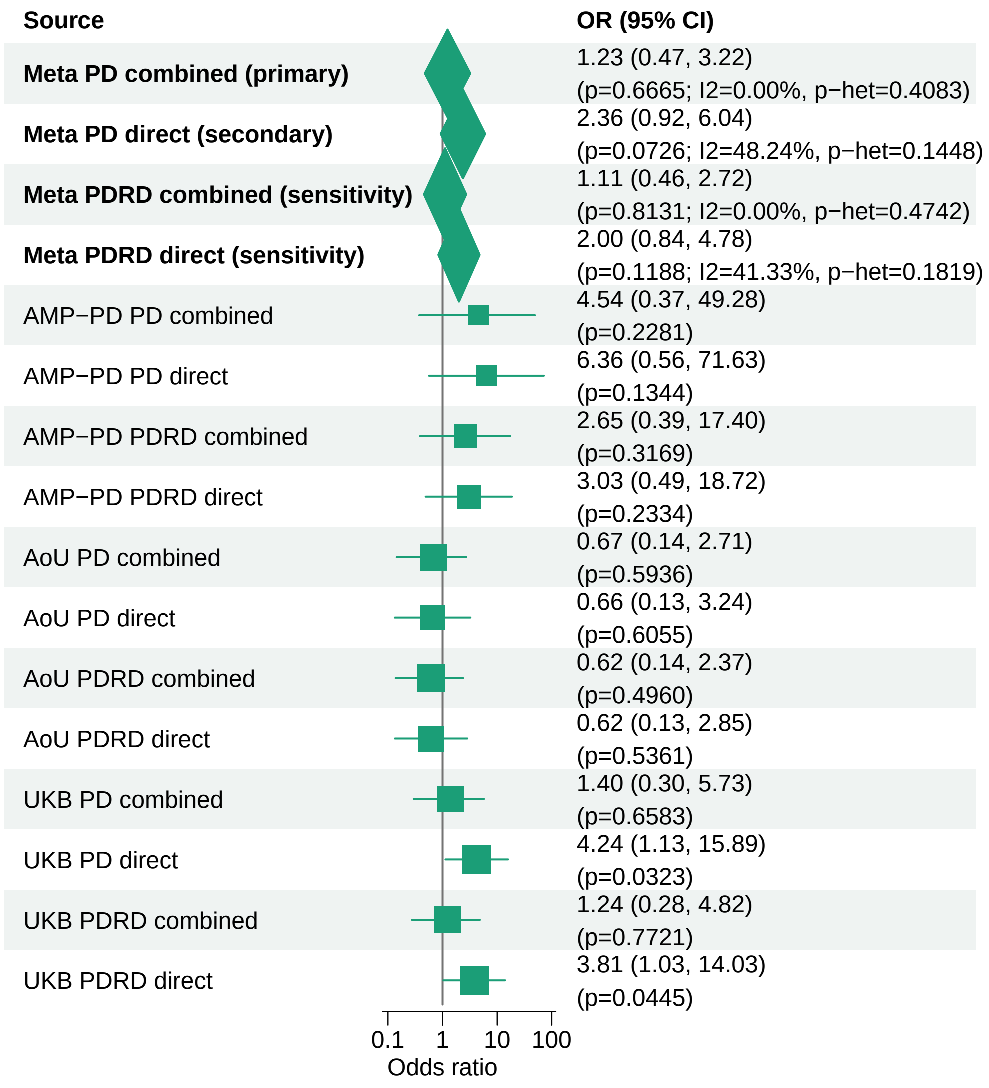


Meta OR/CI and P values are from the exact−kappa=0.5 pooled REGENIE fixed−effects analysis.

#### PD primary/secondary definitions with PDRD sensitivity − PANX2 − LOF+REVEL75 (EUR)

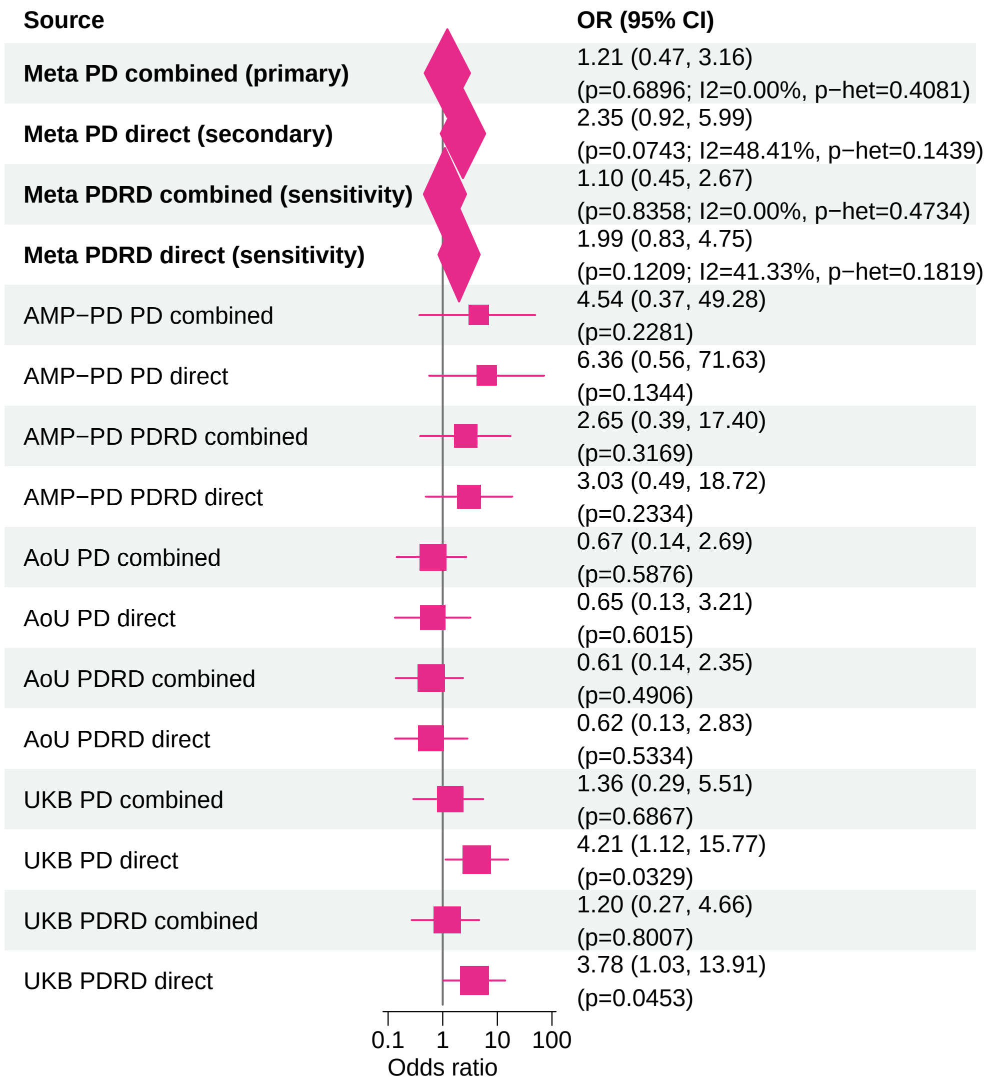


Meta OR/CI and P values are from the exact−kappa=0.5 pooled REGENIE fixed−effects analysis.

### PD primary/secondary definitions with PDRD sensitivity − KRBA2 − LOF (EUR)

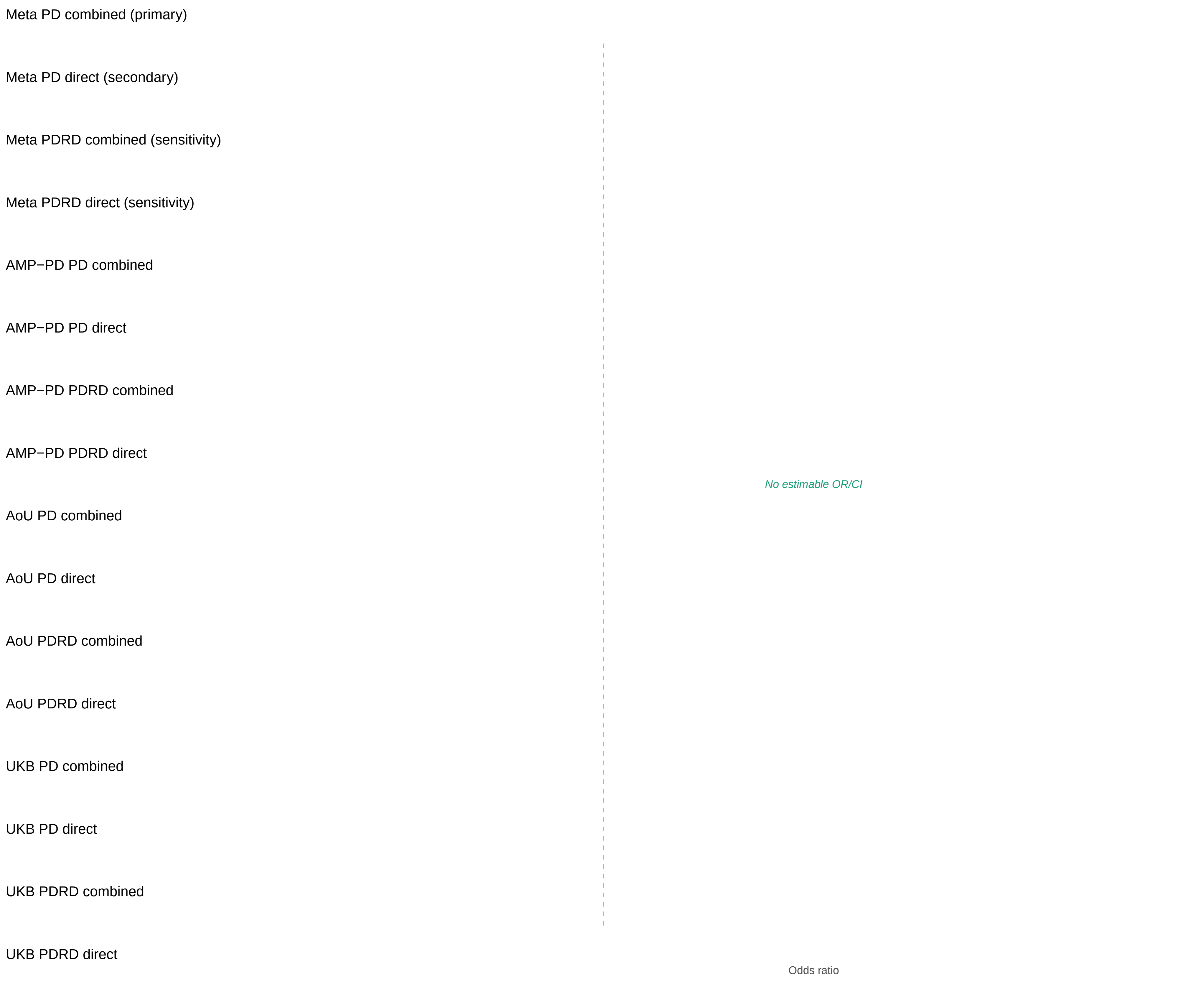


Meta OR/CI and P values are from the exact−kappa=0.5 pooled REGENIE fixed−effects analysis.

### PD primary/secondary definitions with PDRD sensitivity − SQLE − LOF+REVEL50 (EUR)

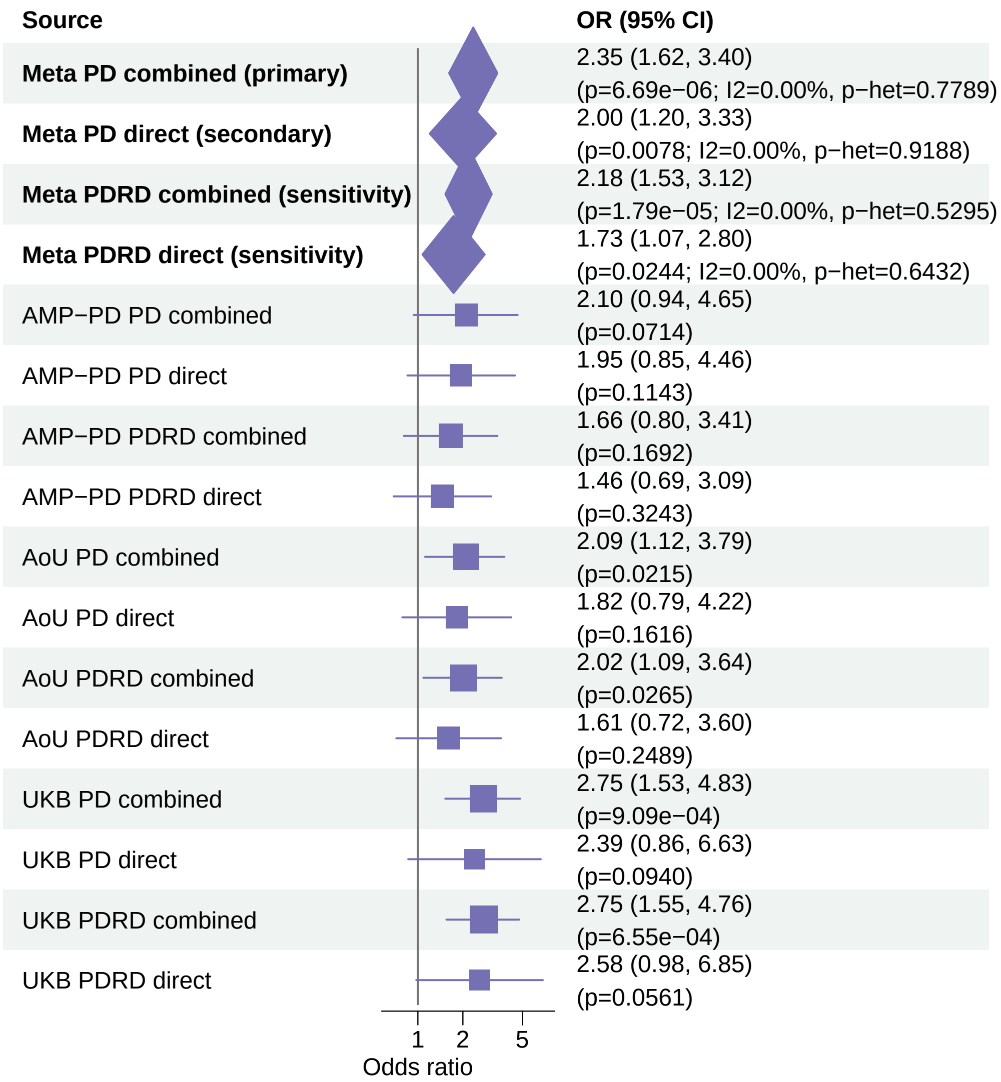


Meta OR/CI and P values are from the exact−kappa=0.5 pooled REGENIE fixed−effects analysis.

#### PD primary/secondary definitions with PDRD sensitivity − SPIRE2 − LOF+REVEL50 (EUR)

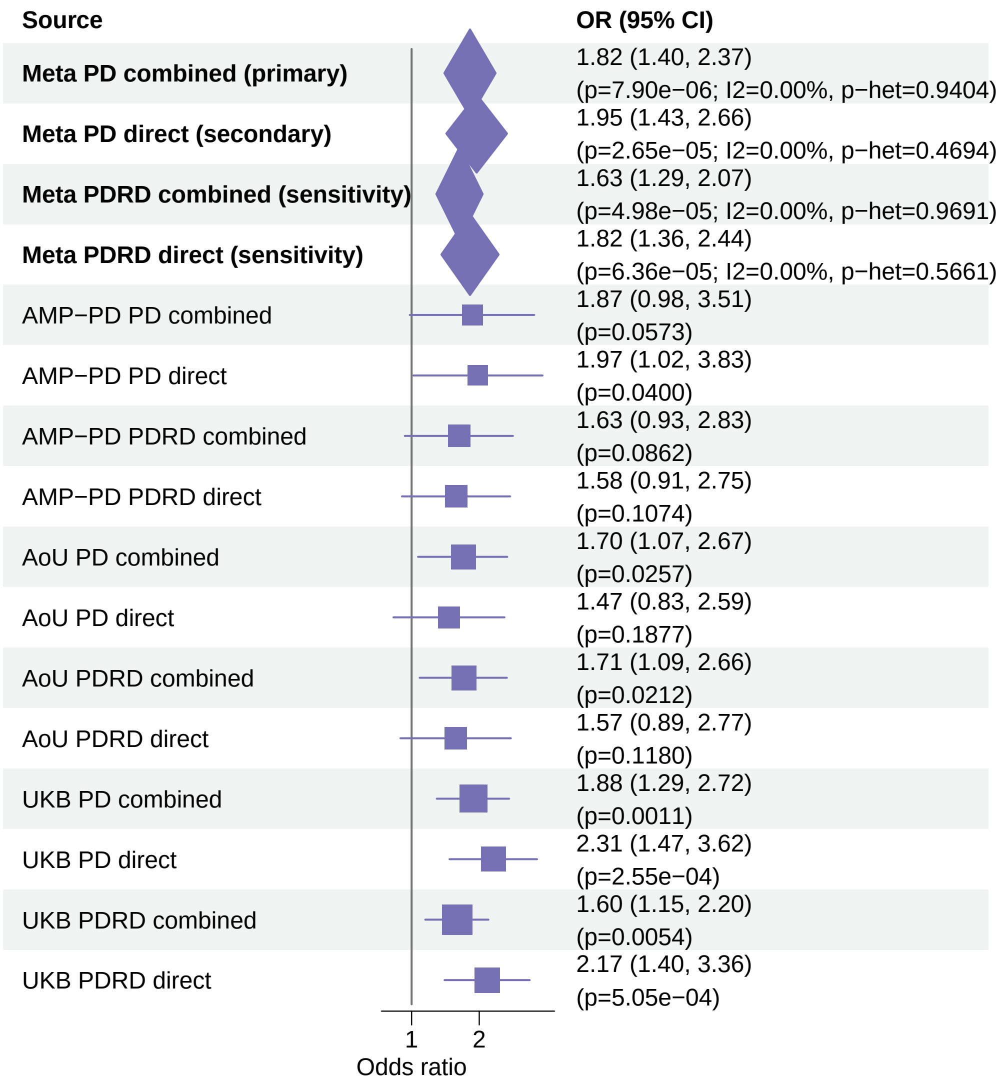


Meta OR/CI and P values are from the exact−kappa=0.5 pooled REGENIE fixed−effects analysis.

#### PD primary/secondary definitions with PDRD sensitivity − KRBA2 − LOF+REVEL50 (EUR)

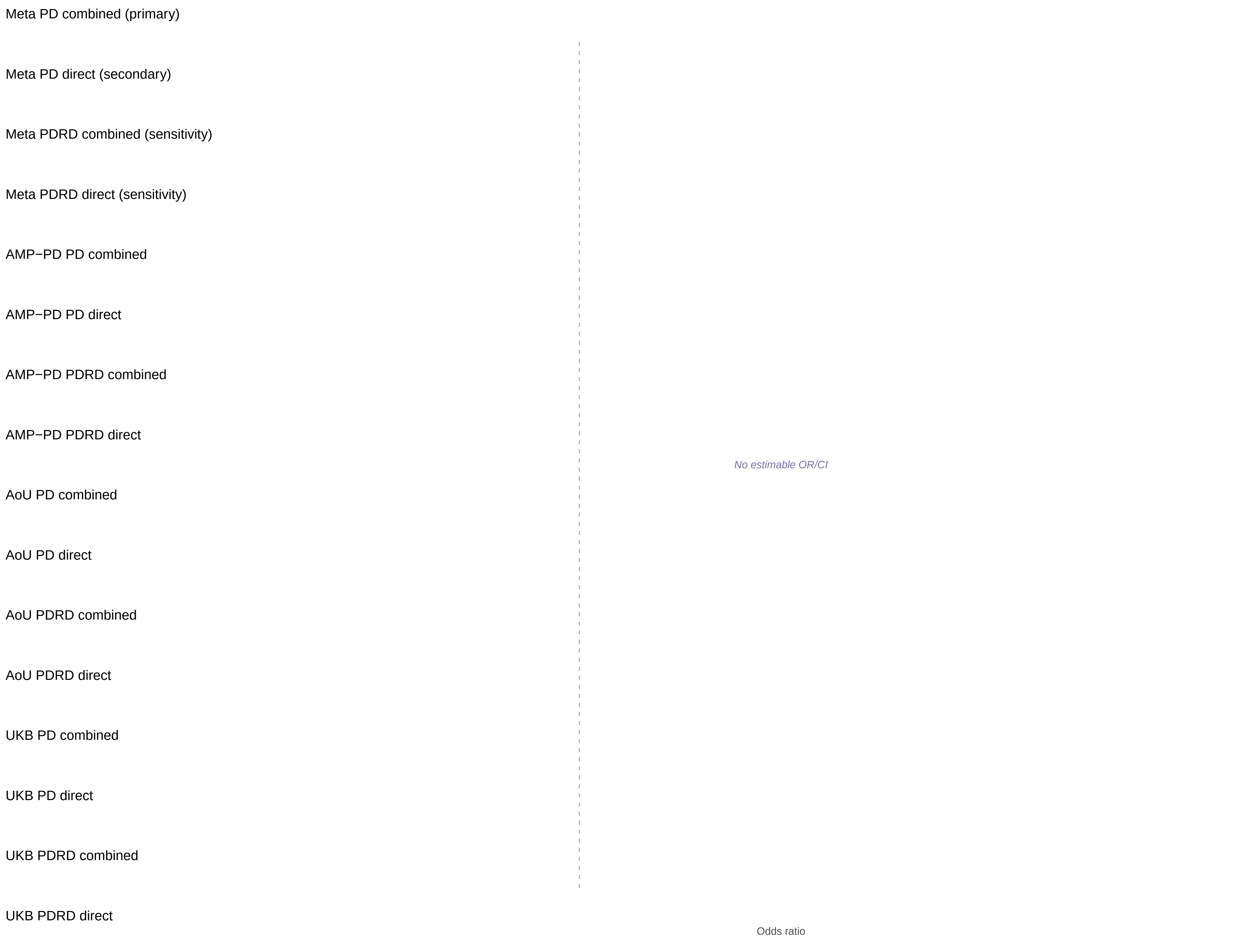


Meta OR/CI and P values are from the exact−kappa=0.5 pooled REGENIE fixed−effects analysis.

#### PD primary/secondary definitions with PDRD sensitivity − AGFG1 − LOF+REVEL25 (EUR)

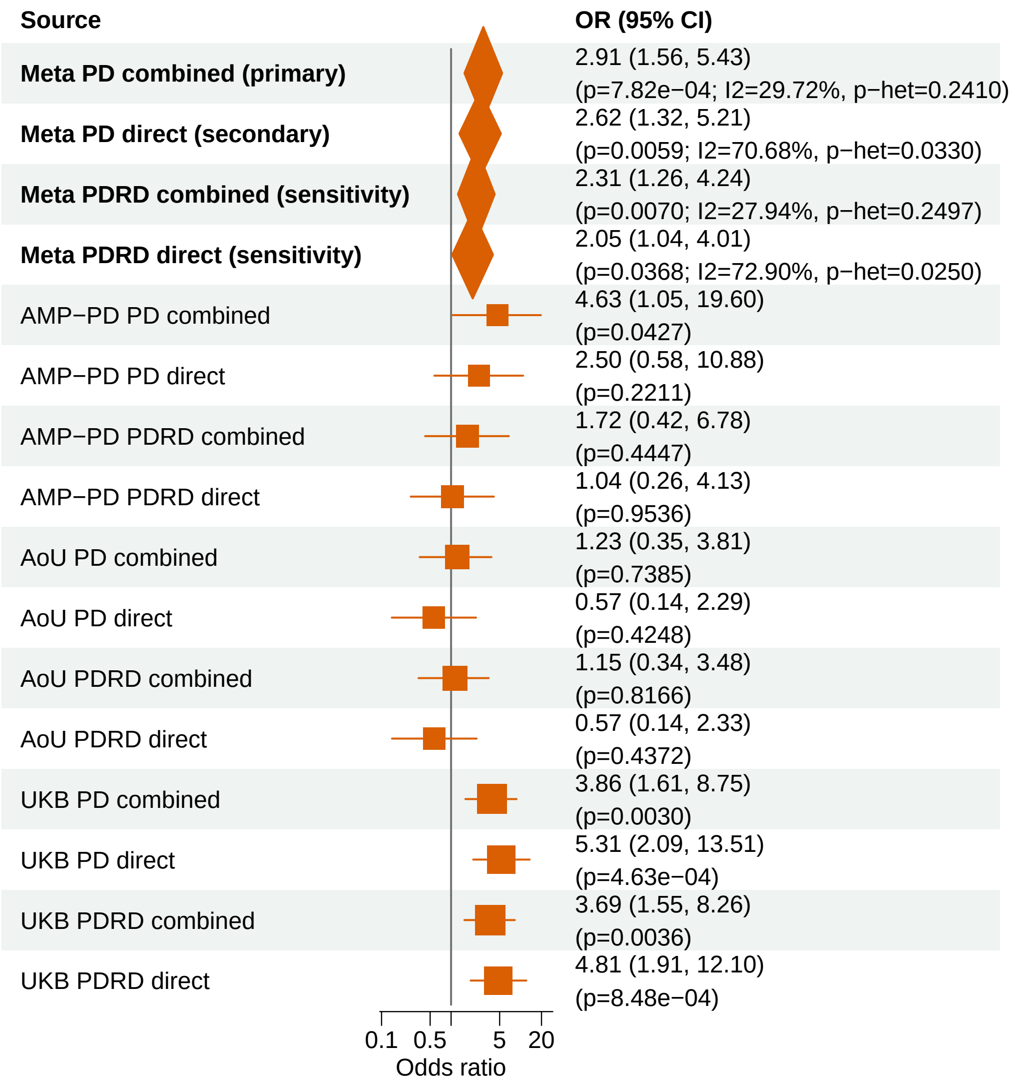


Meta OR/CI and P values are from the exact−kappa=0.5 pooled REGENIE fixed−effects analysis.

### PD primary/secondary definitions with PDRD sensitivity − ATAD2B − LOF (EUR)

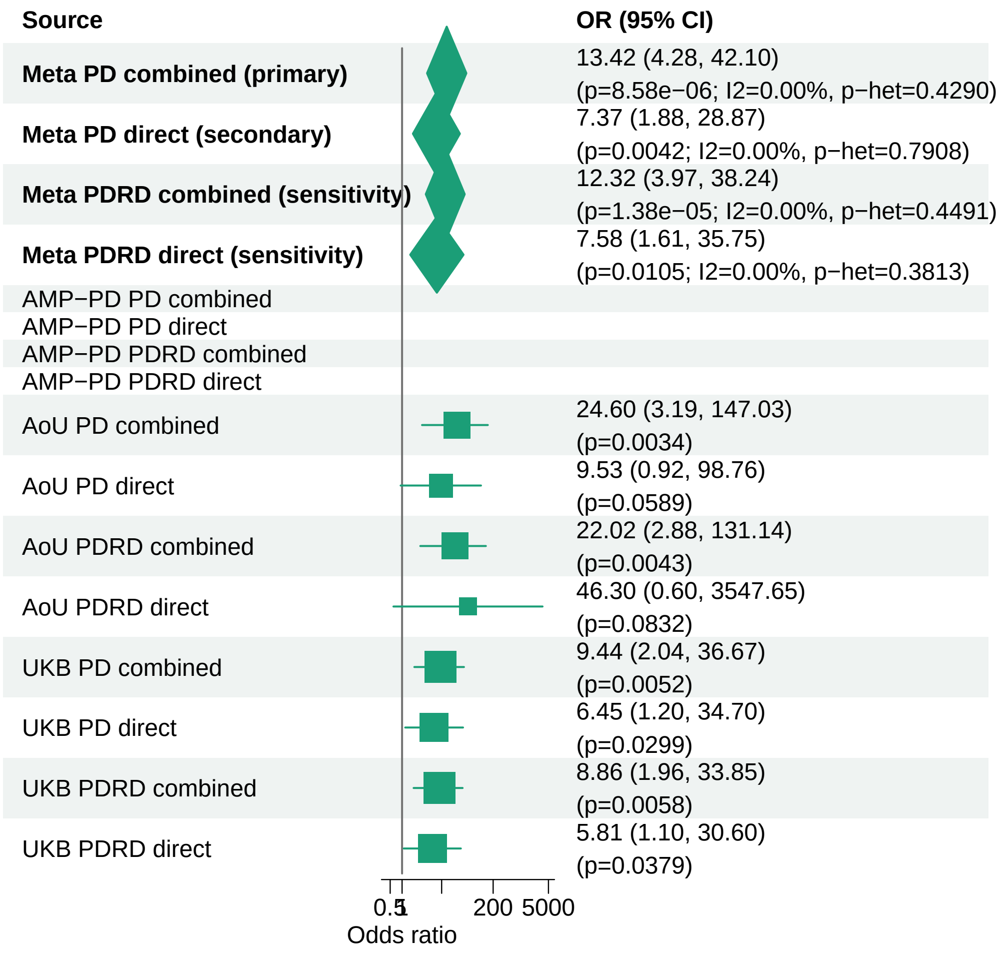


Meta OR/CI and P values are from the exact−kappa=0.5 pooled REGENIE fixed−effects analysis.

### PD primary/secondary definitions with PDRD sensitivity − HOXA13 − LOF (EUR)

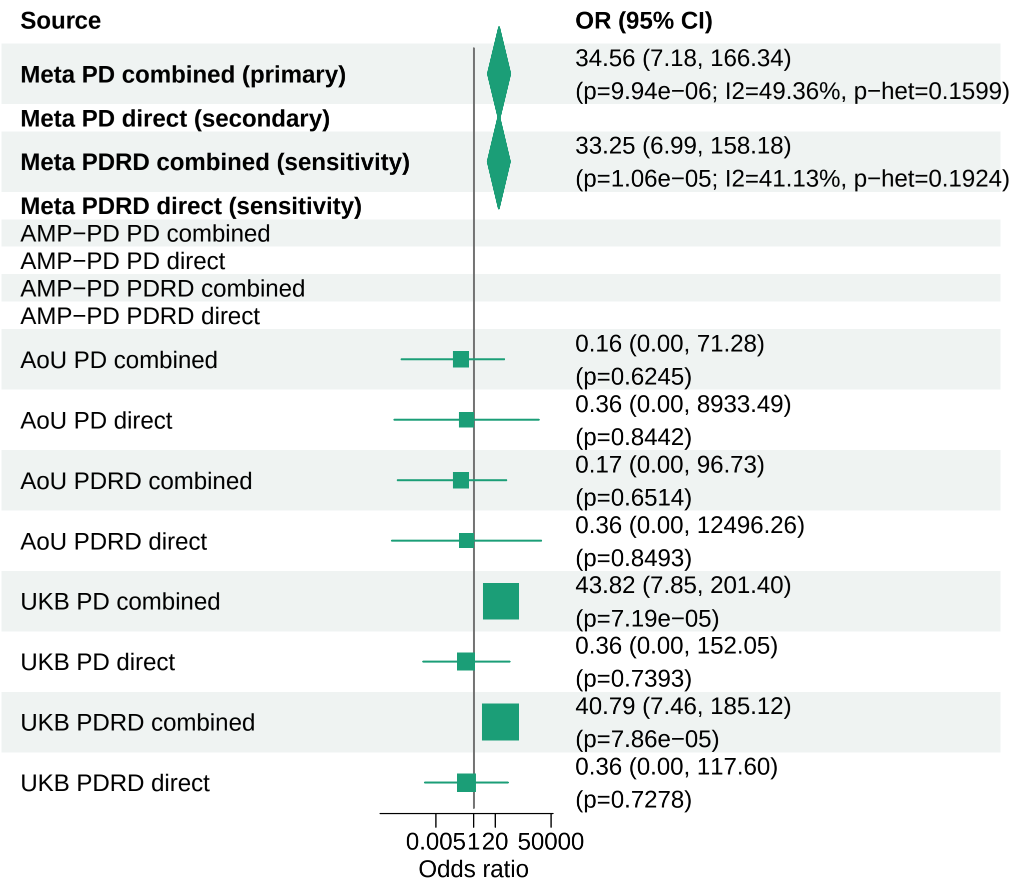


Meta OR/CI and P values are from the exact−kappa=0.5 pooled REGENIE fixed−effects analysis.
