## Supplementary_Fig11 for "Population-scale burden analysis of rare damaging coding variants identifies novel risk genes for Alzheimer’s disease and related dementias and Parkinson’s disease and related disorders"

### PD primary/secondary definitions with PDRD sensitivity − LRRK2 − LOF+REVEL75 (ALL)

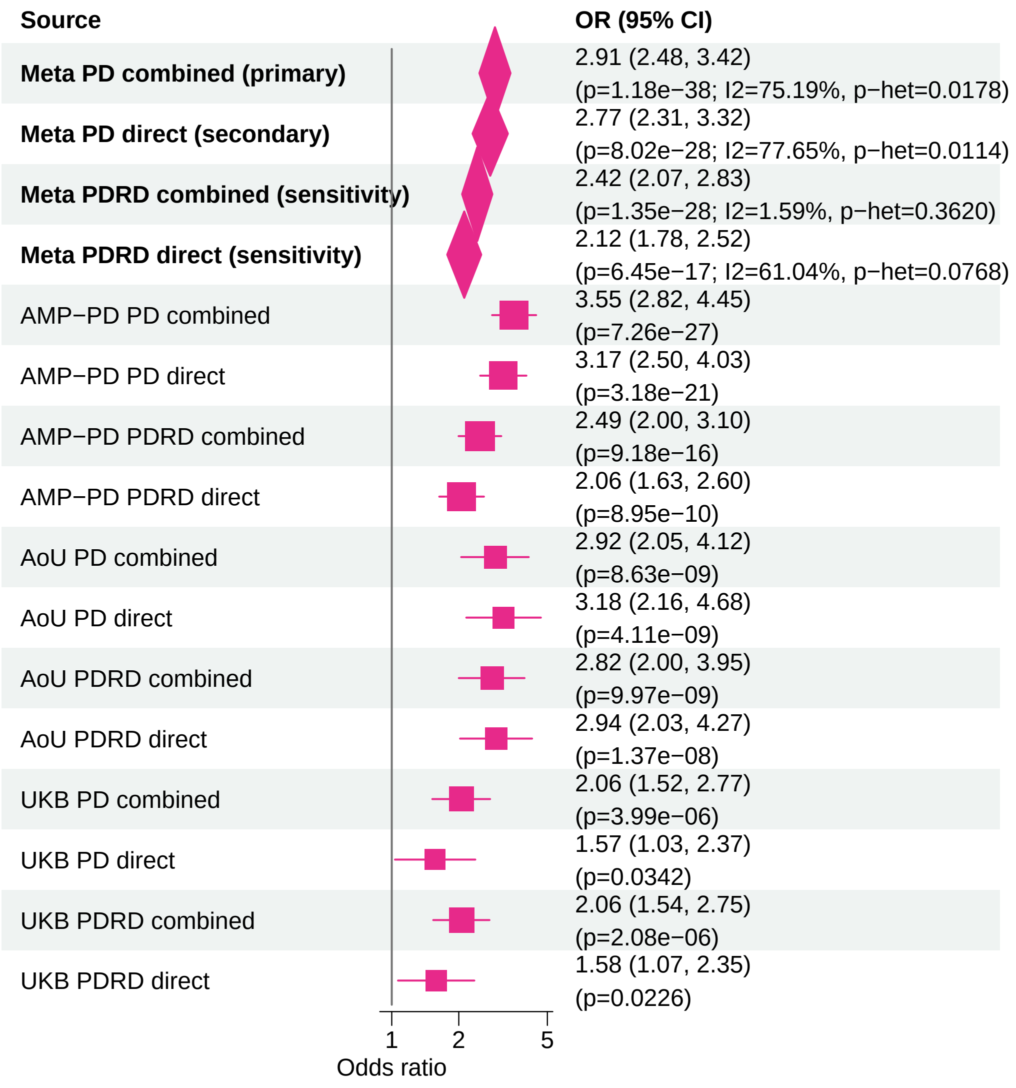


Meta OR/CI and P values are from the exact−kappa=0.5 pooled REGENIE fixed−effects analysis.

#### PD primary/secondary definitions with PDRD sensitivity − GBA1 − LOF+REVEL25 (ALL)

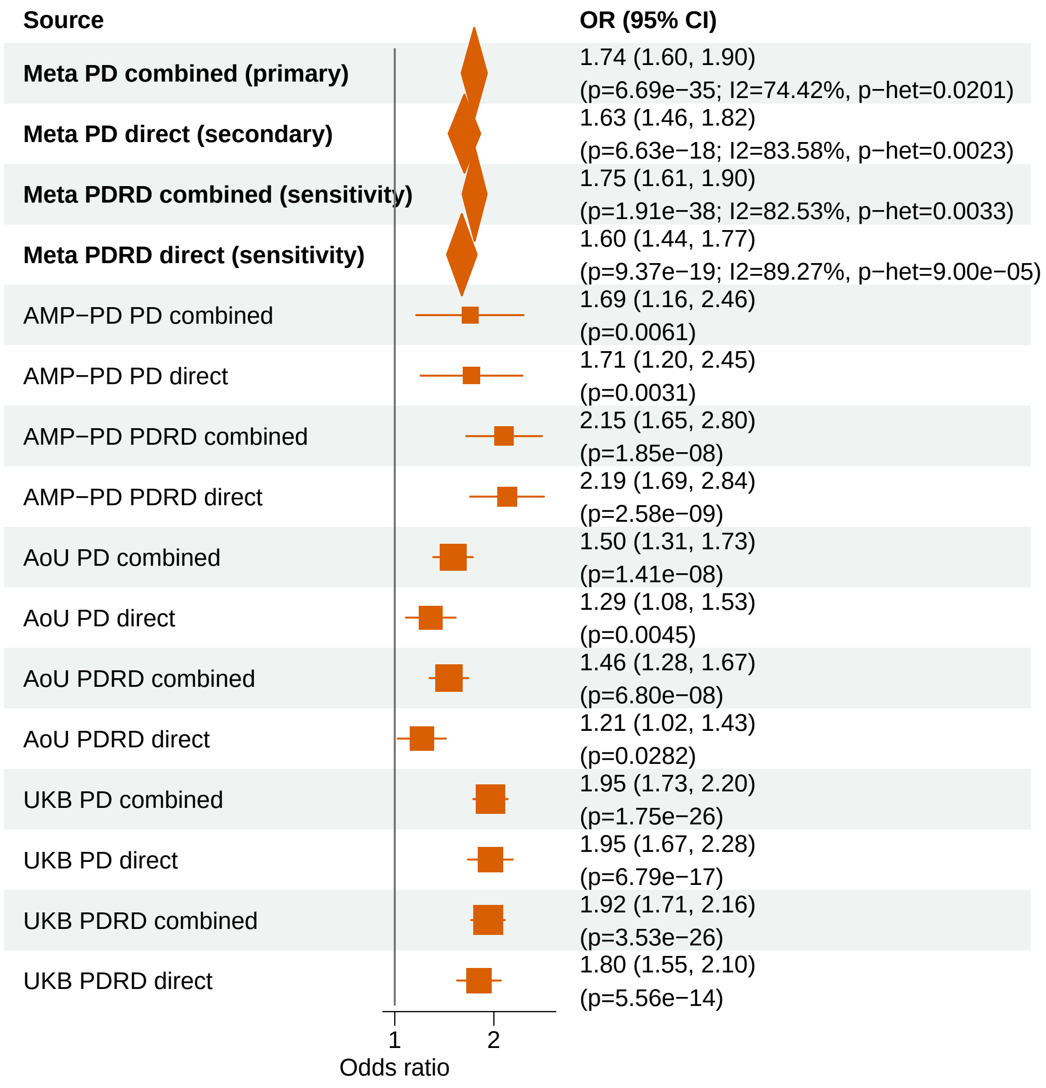


Meta OR/CI and P values are from the exact−kappa=0.5 pooled REGENIE fixed−effects analysis.

### PD primary/secondary definitions with PDRD sensitivity − GBA1 − LOF+REVEL50 (ALL)

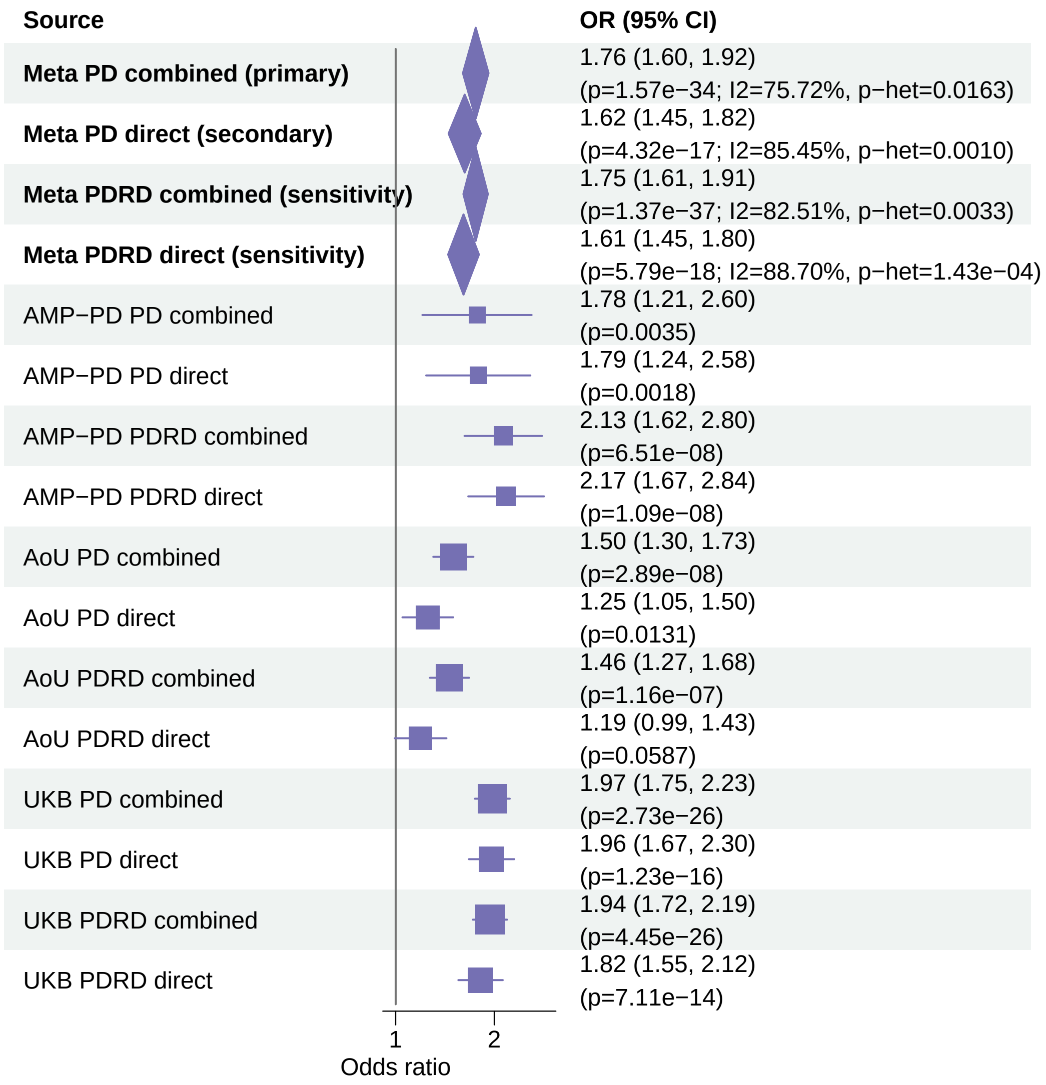


Meta OR/CI and P values are from the exact−kappa=0.5 pooled REGENIE fixed−effects analysis.

#### PD primary/secondary definitions with PDRD sensitivity − LRRK2 − LOF+REVEL50 (ALL)

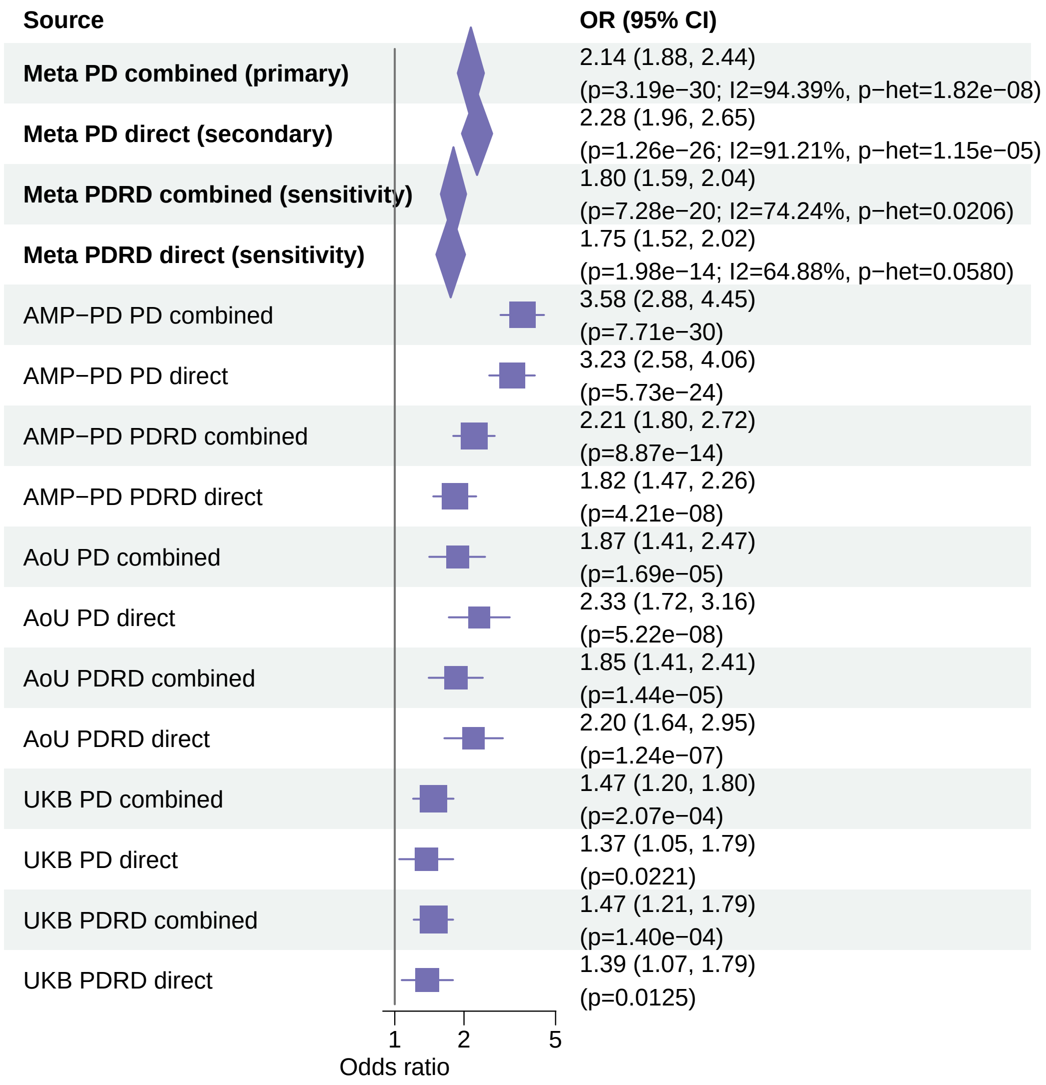


Meta OR/CI and P values are from the exact−kappa=0.5 pooled REGENIE fixed−effects analysis.

PD primary/secondary definitions with PDRD sensitivity − GBA1 − LOF+REVEL75 (ALL)

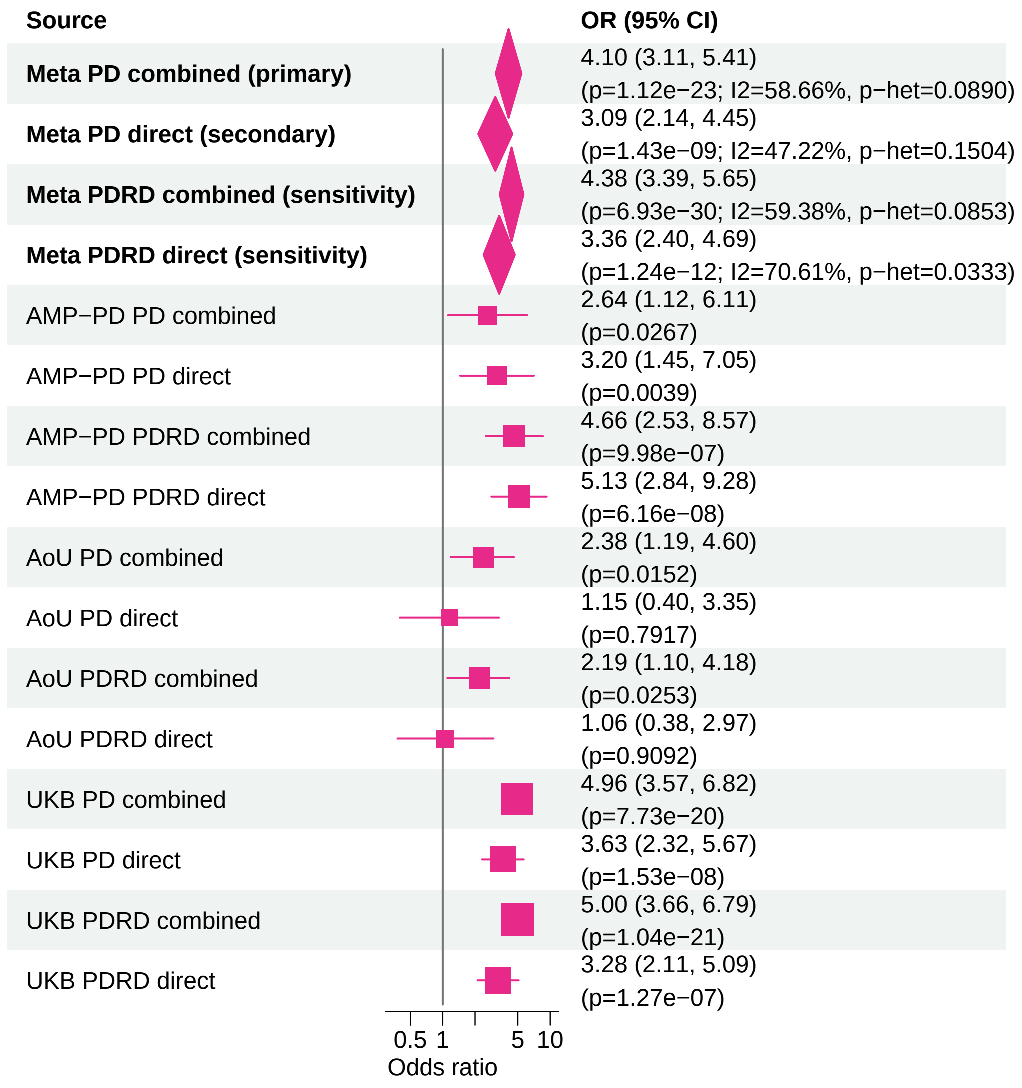


Meta OR/CI and P values are from the exact−kappa=0.5 pooled REGENIE fixed−effects analysis.

#### PD primary/secondary definitions with PDRD sensitivity − LRRK2 − LOF+REVEL25 (ALL)

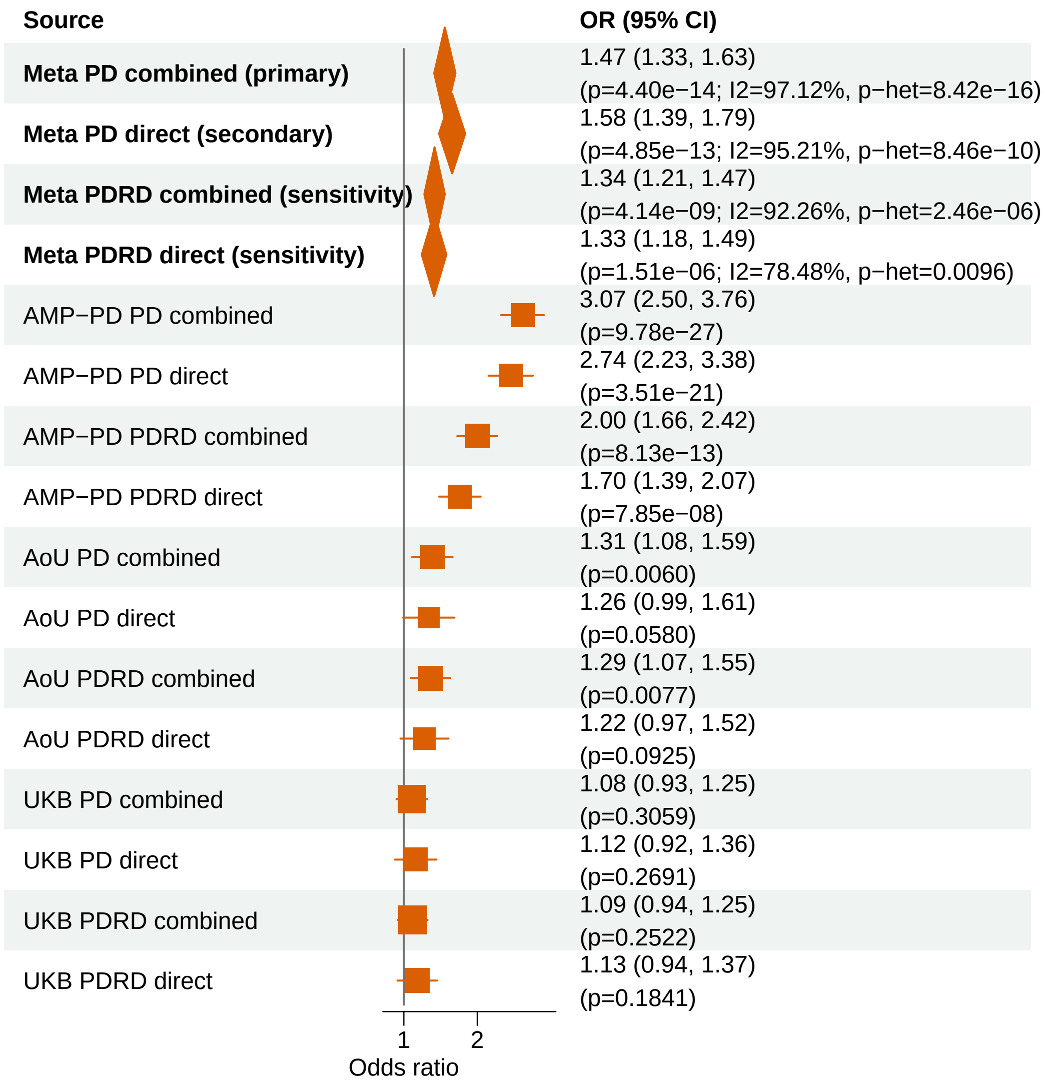


Meta OR/CI and P values are from the exact−kappa=0.5 pooled REGENIE fixed−effects analysis.

PD primary/secondary definitions with PDRD sensitivity − GBA1 − LOF (ALL)

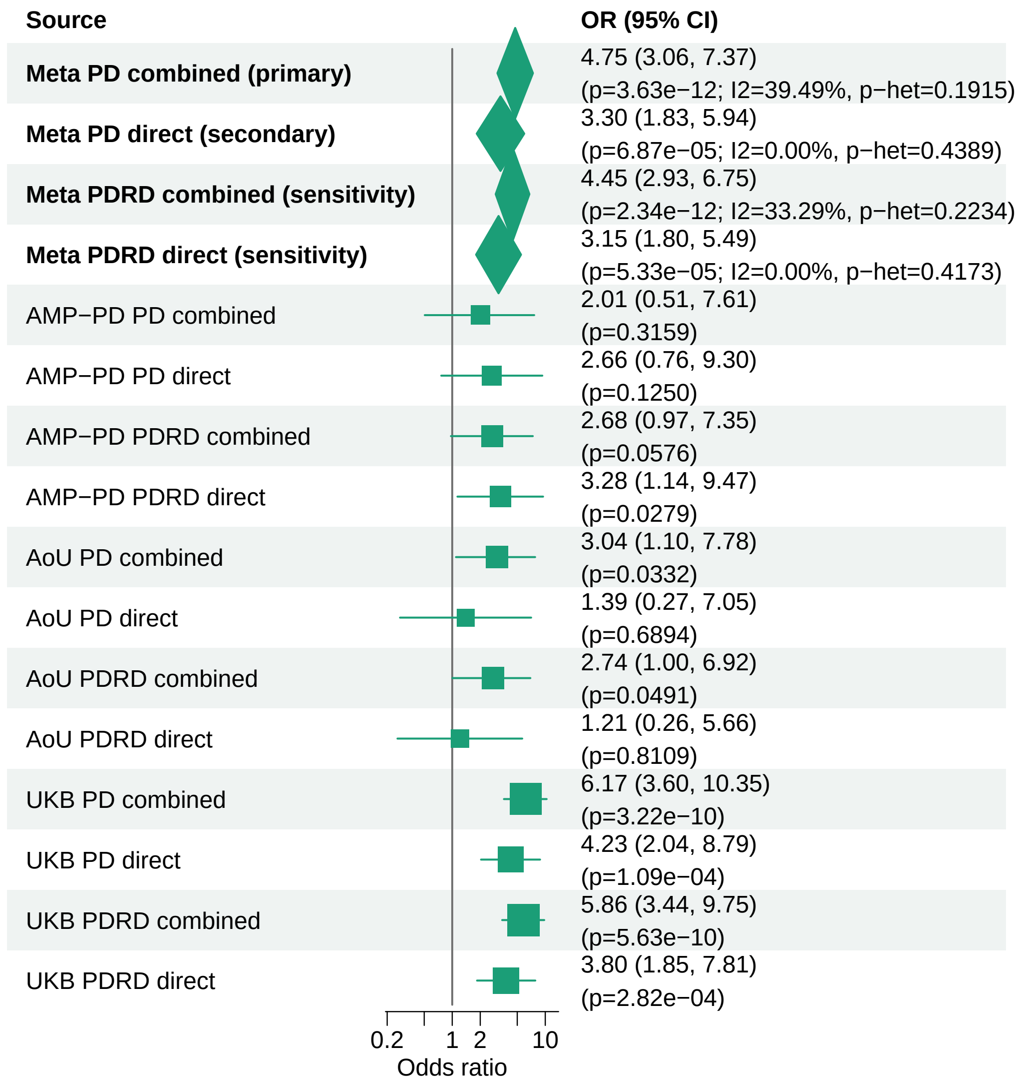


Meta OR/CI and P values are from the exact−kappa=0.5 pooled REGENIE fixed−effects analysis.

### PD primary/secondary definitions with PDRD sensitivity − ANKRD27 − LOF+REVEL25 (ALL)

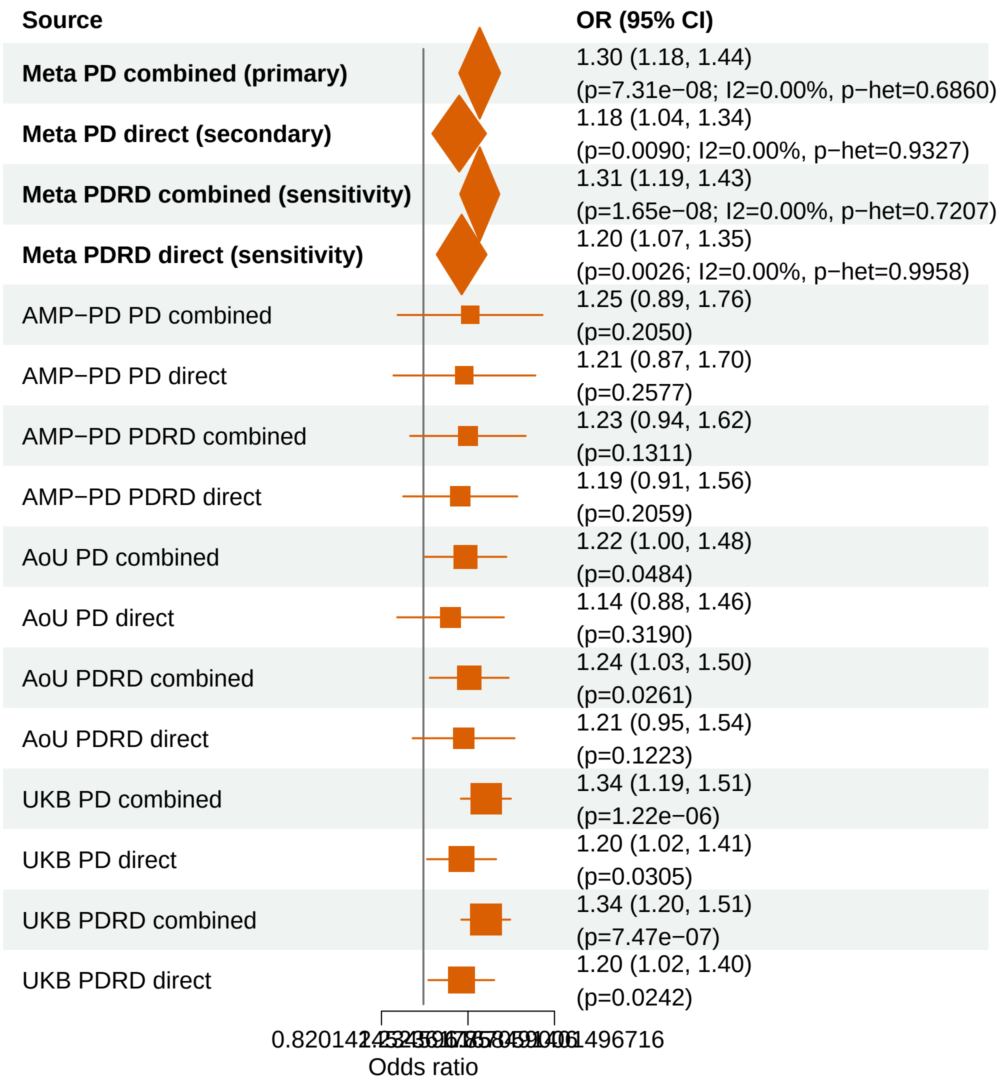


Meta OR/CI and P values are from the exact−kappa=0.5 pooled REGENIE fixed−effects analysis.

#### PD primary/secondary definitions with PDRD sensitivity − CCL7 − LOF+REVEL50 (ALL)

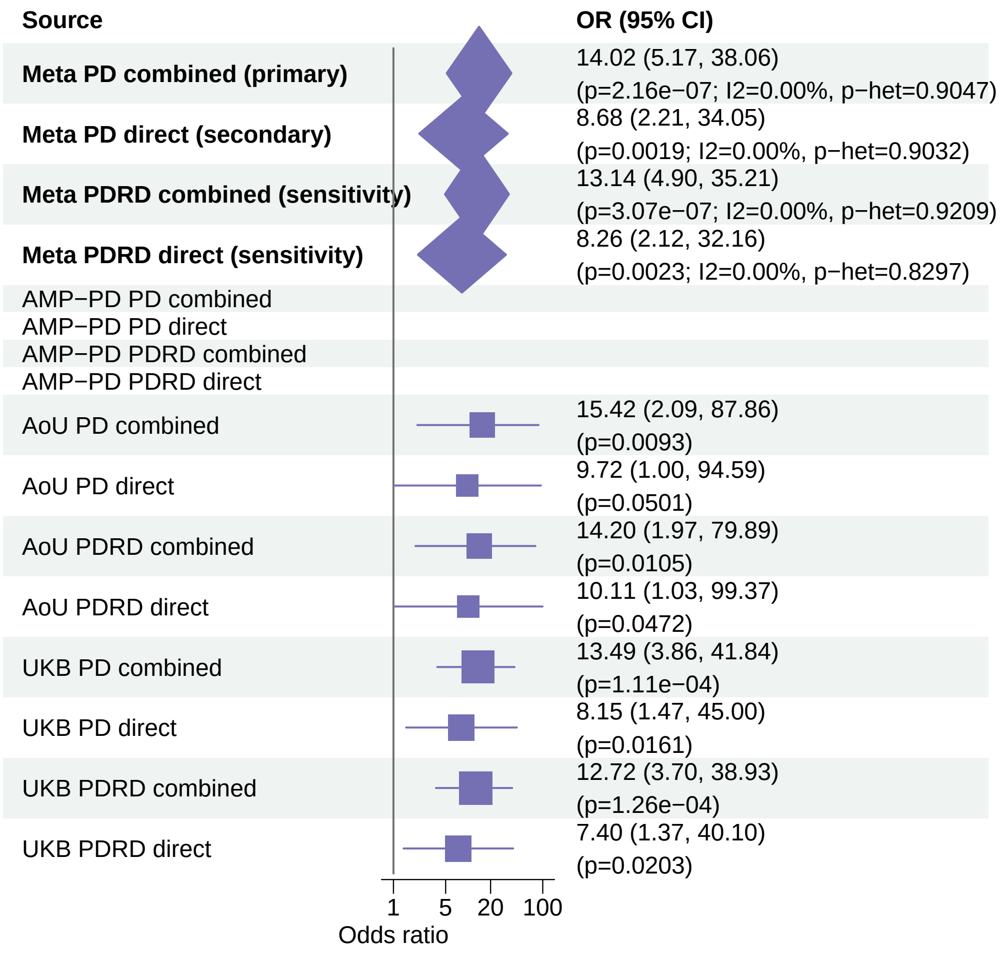


Meta OR/CI and P values are from the exact−kappa=0.5 pooled REGENIE fixed−effects analysis.

### PD primary/secondary definitions with PDRD sensitivity − PANX2 − LOF+REVEL50 (ALL)

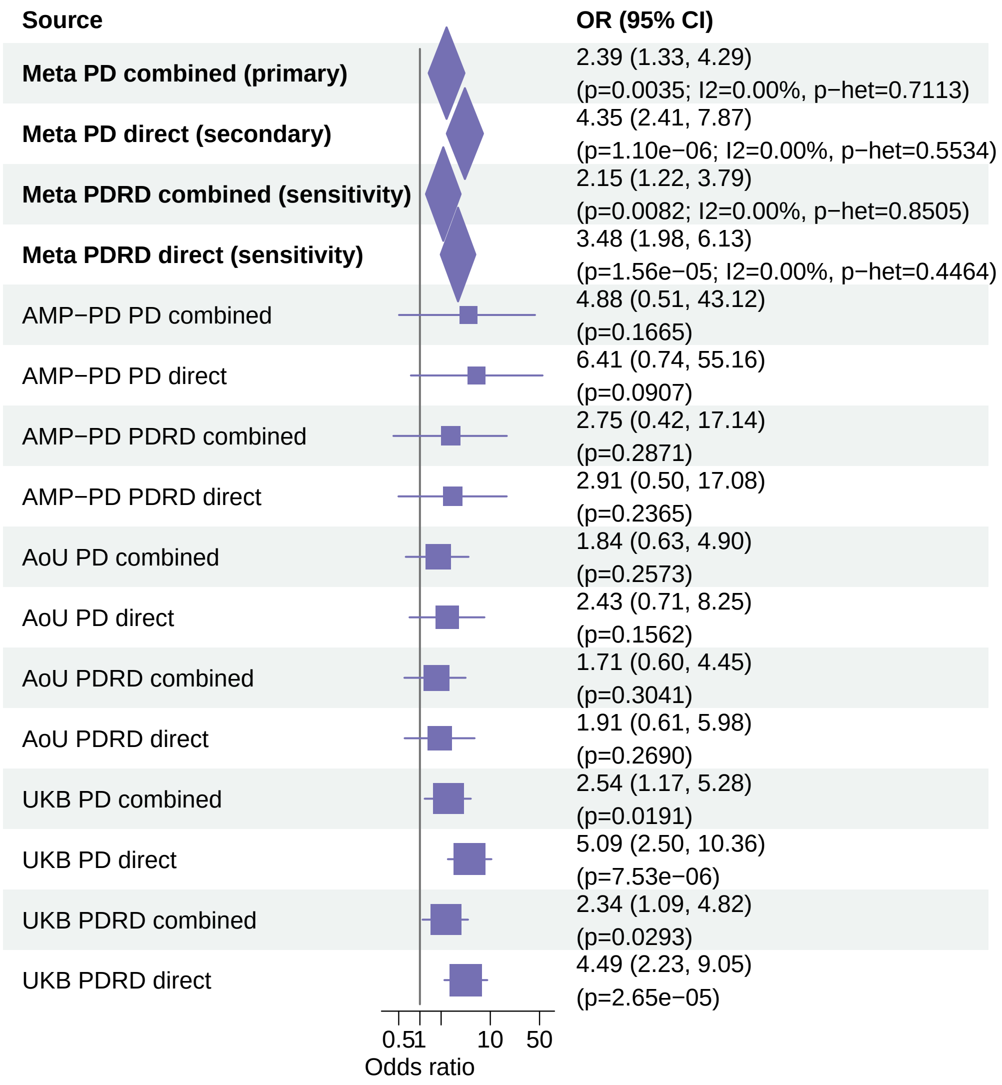


Meta OR/CI and P values are from the exact−kappa=0.5 pooled REGENIE fixed−effects analysis.

#### PD primary/secondary definitions with PDRD sensitivity − USP19 − LOF+REVEL75 (ALL)

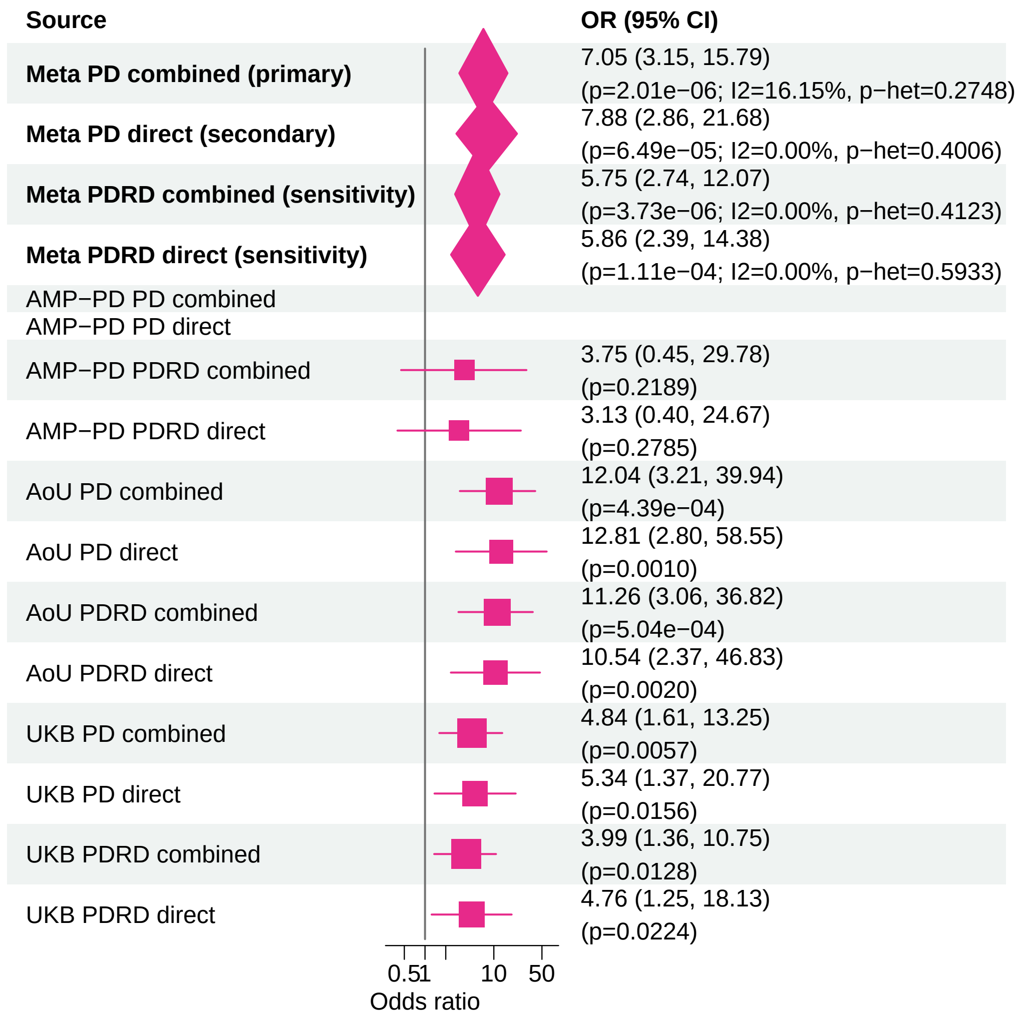


Meta OR/CI and P values are from the exact−kappa=0.5 pooled REGENIE fixed−effects analysis.

#### PD primary/secondary definitions with PDRD sensitivity − USP19 − LOF (ALL)

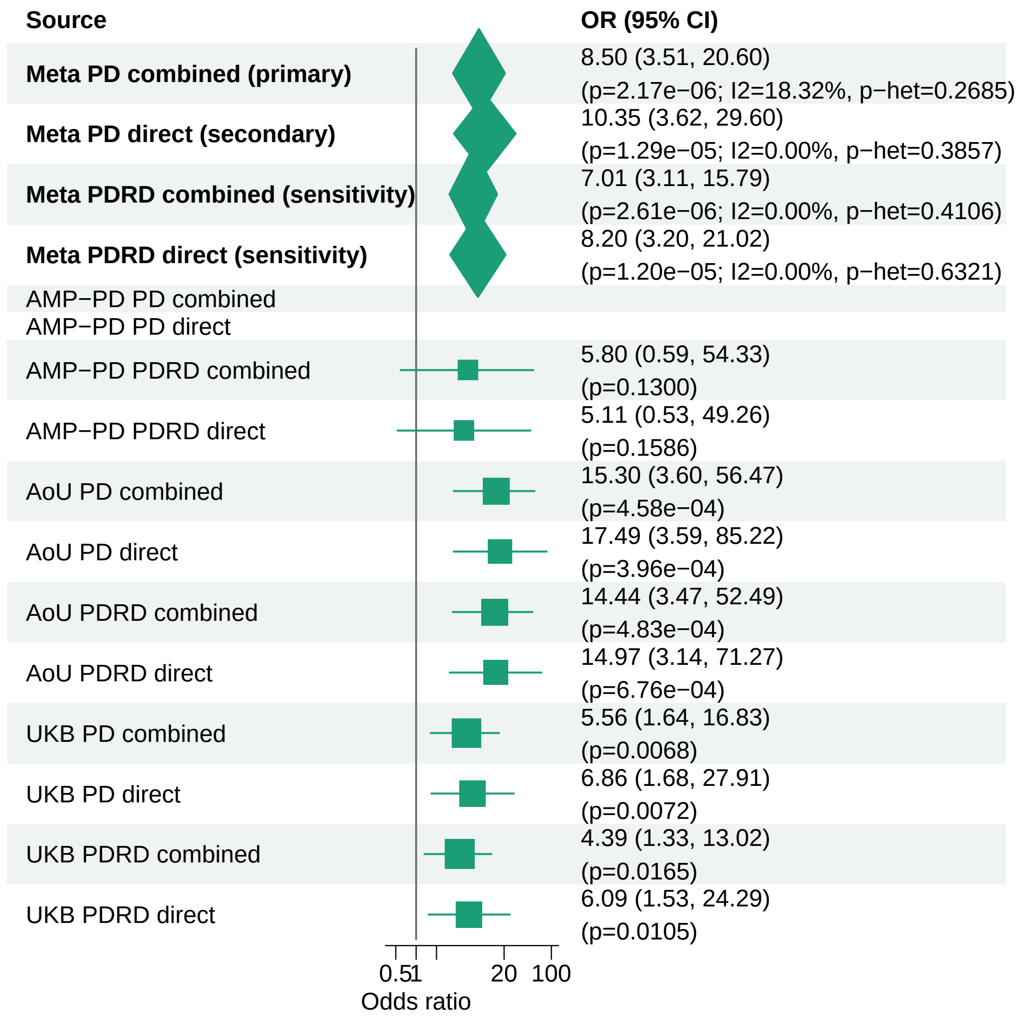


Meta OR/CI and P values are from the exact−kappa=0.5 pooled REGENIE fixed−effects analysis.

#### PD primary/secondary definitions with PDRD sensitivity − ZSCAN25 − LOF (ALL)

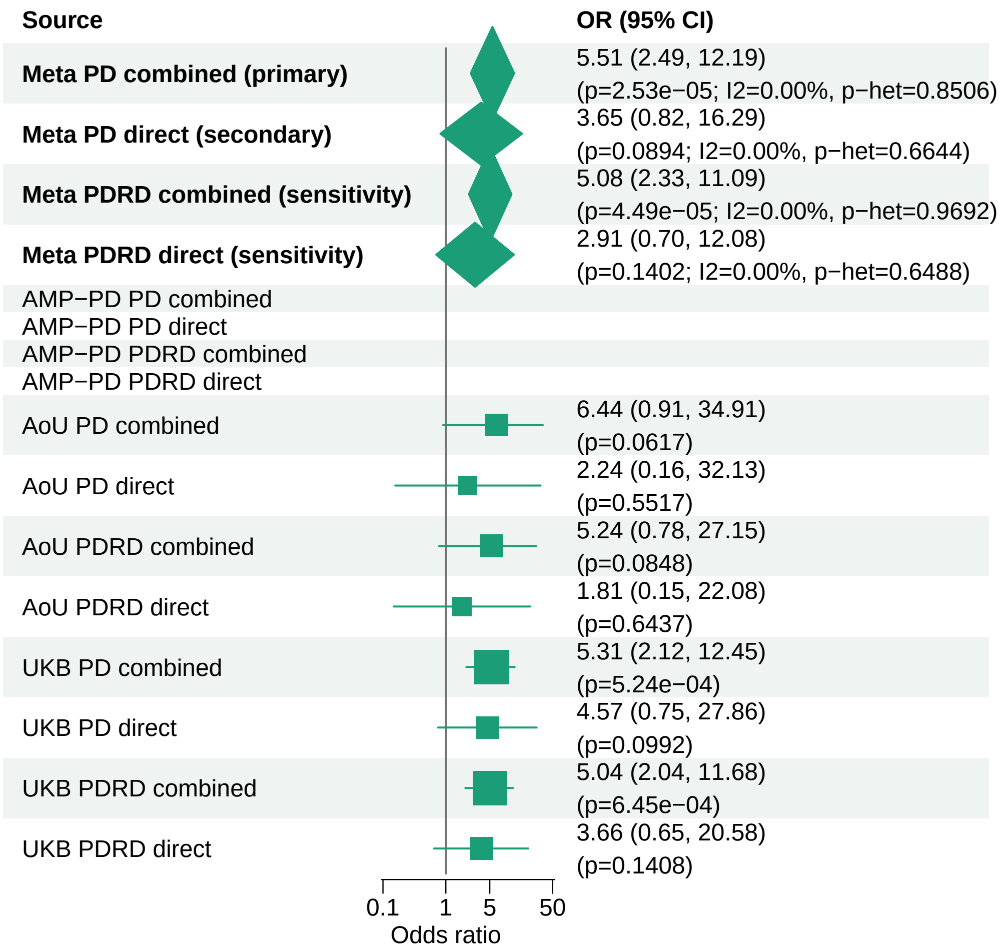


Meta OR/CI and P values are from the exact−kappa=0.5 pooled REGENIE fixed−effects analysis.

#### PD primary/secondary definitions with PDRD sensitivity − BNIP3 − LOF (ALL)

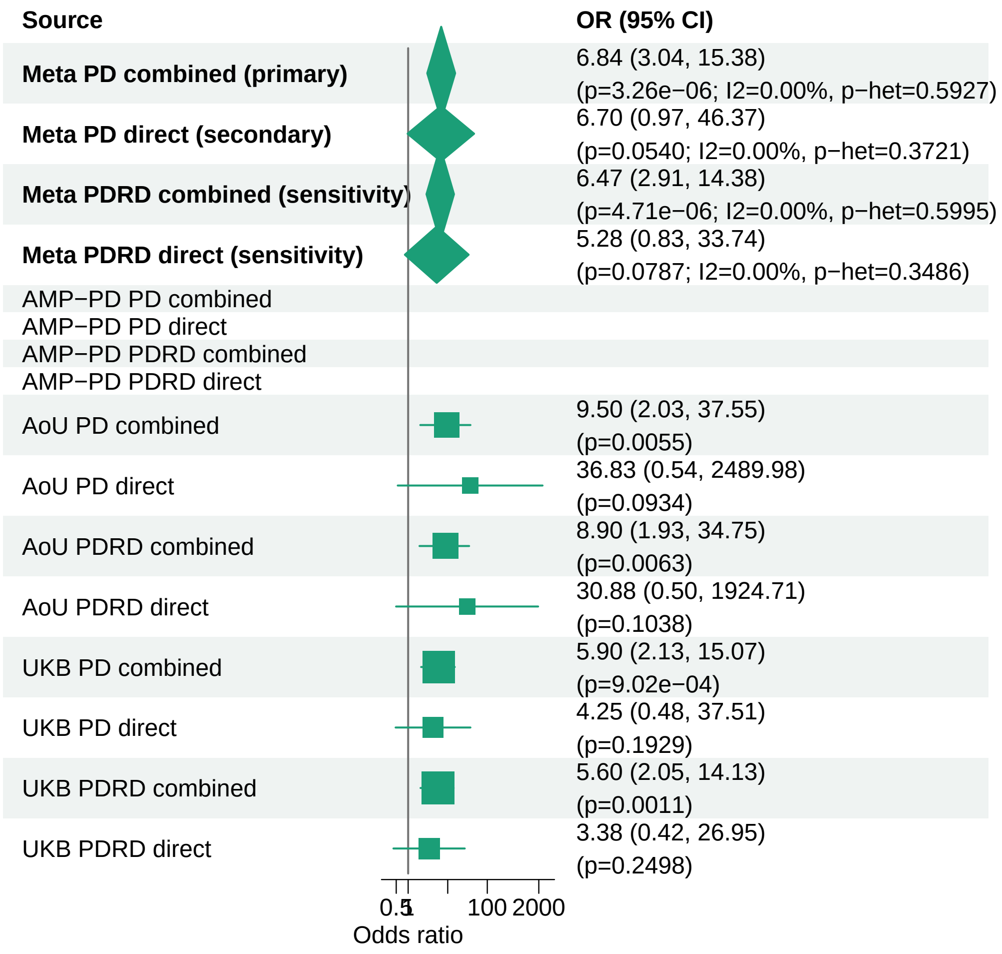


Meta OR/CI and P values are from the exact−kappa=0.5 pooled REGENIE fixed−effects analysis.

### PD primary/secondary definitions with PDRD sensitivity − SOBP − LOF+REVEL50 (ALL)

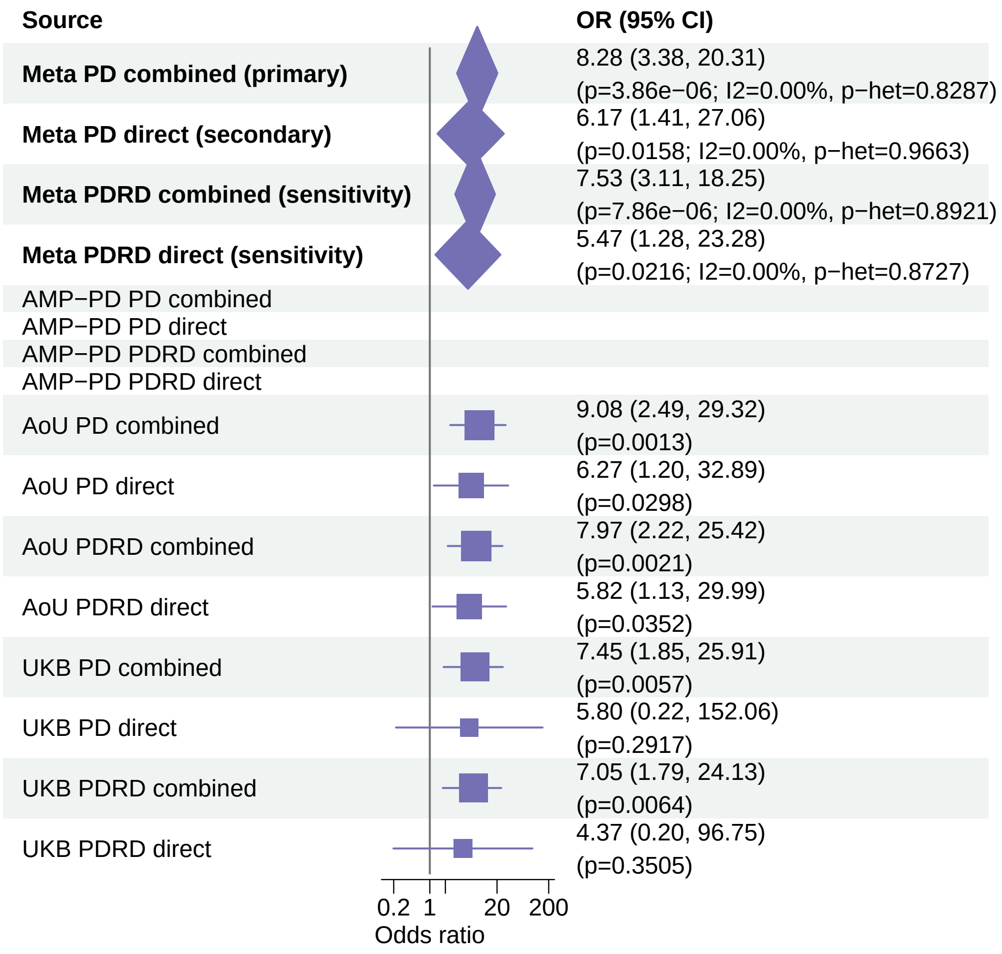


Meta OR/CI and P values are from the exact−kappa=0.5 pooled REGENIE fixed−effects analysis.

#### PD primary/secondary definitions with PDRD sensitivity − KANSL3 − LOF (ALL)

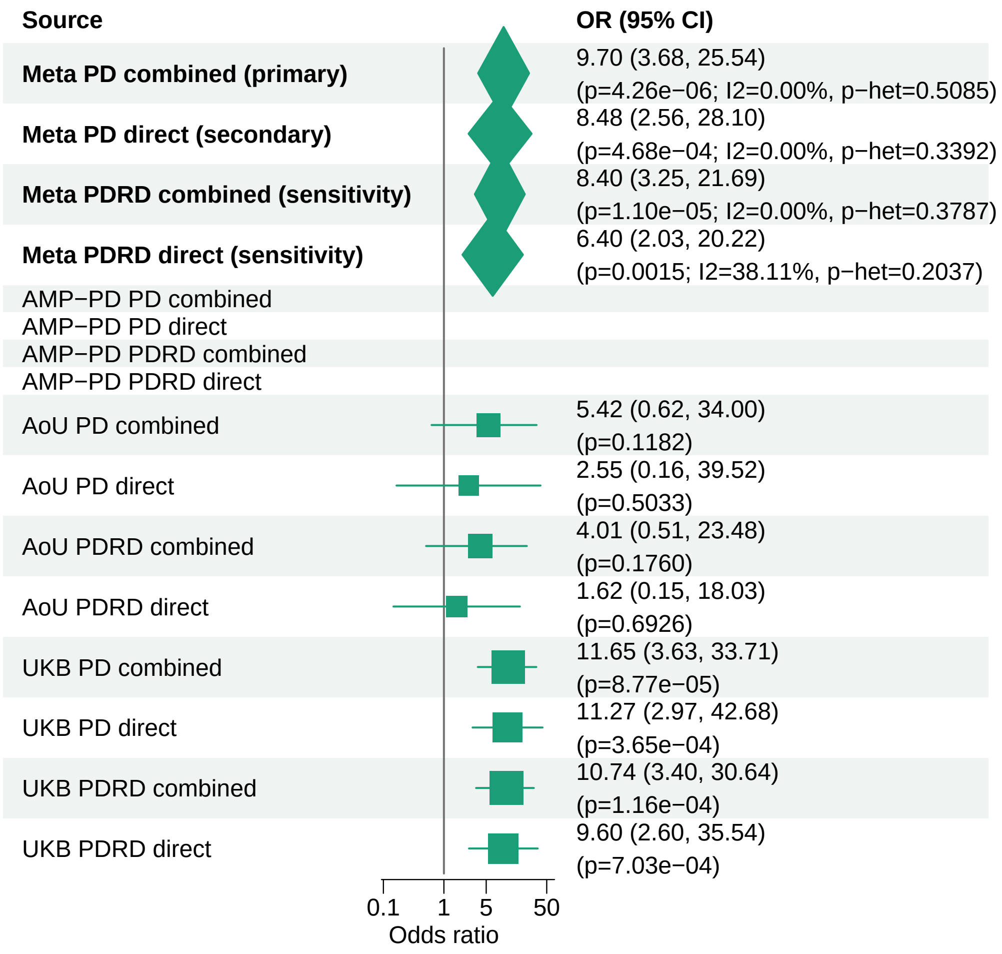


Meta OR/CI and P values are from the exact−kappa=0.5 pooled REGENIE fixed−effects analysis.

### PD primary/secondary definitions with PDRD sensitivity − PANX2 − LOF (ALL)

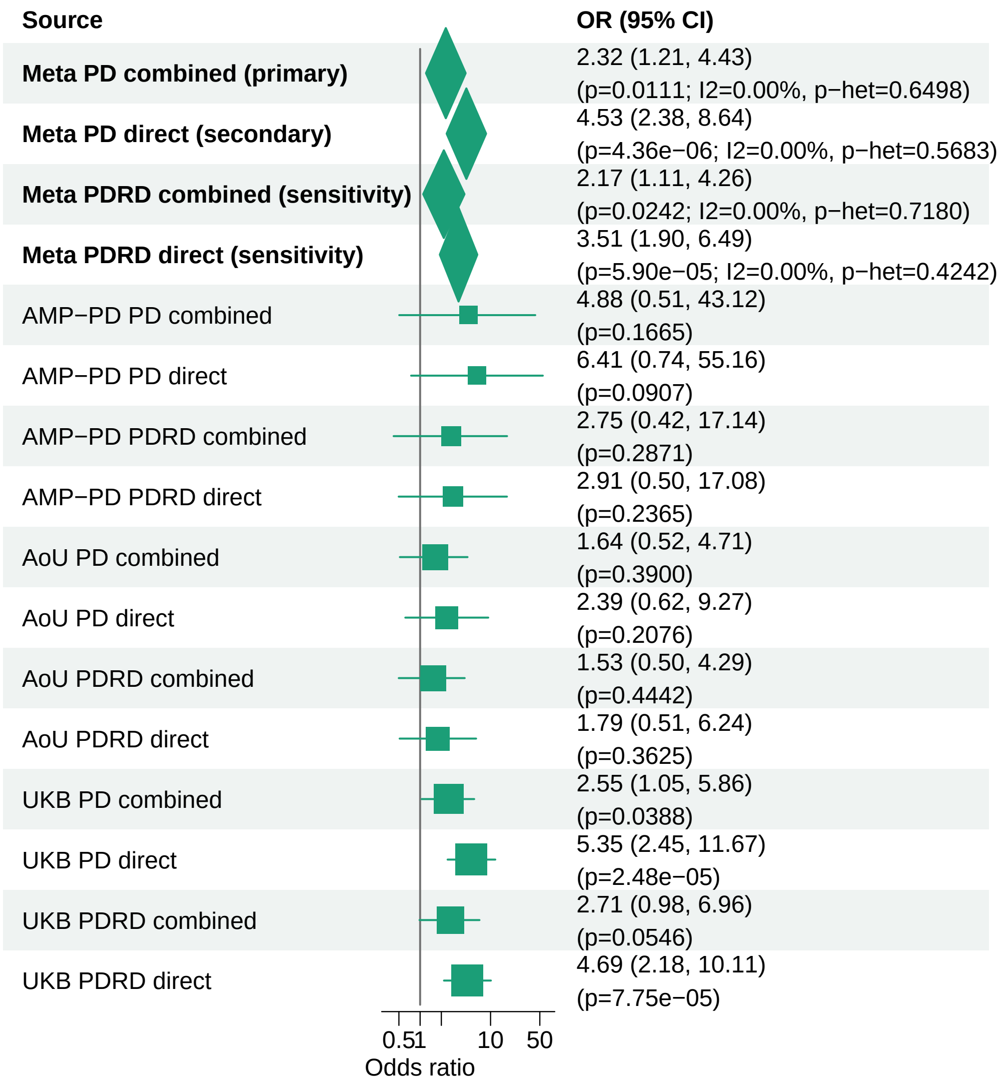


Meta OR/CI and P values are from the exact−kappa=0.5 pooled REGENIE fixed−effects analysis.

### PD primary/secondary definitions with PDRD sensitivity − PANX2 − LOF+REVEL75 (ALL)

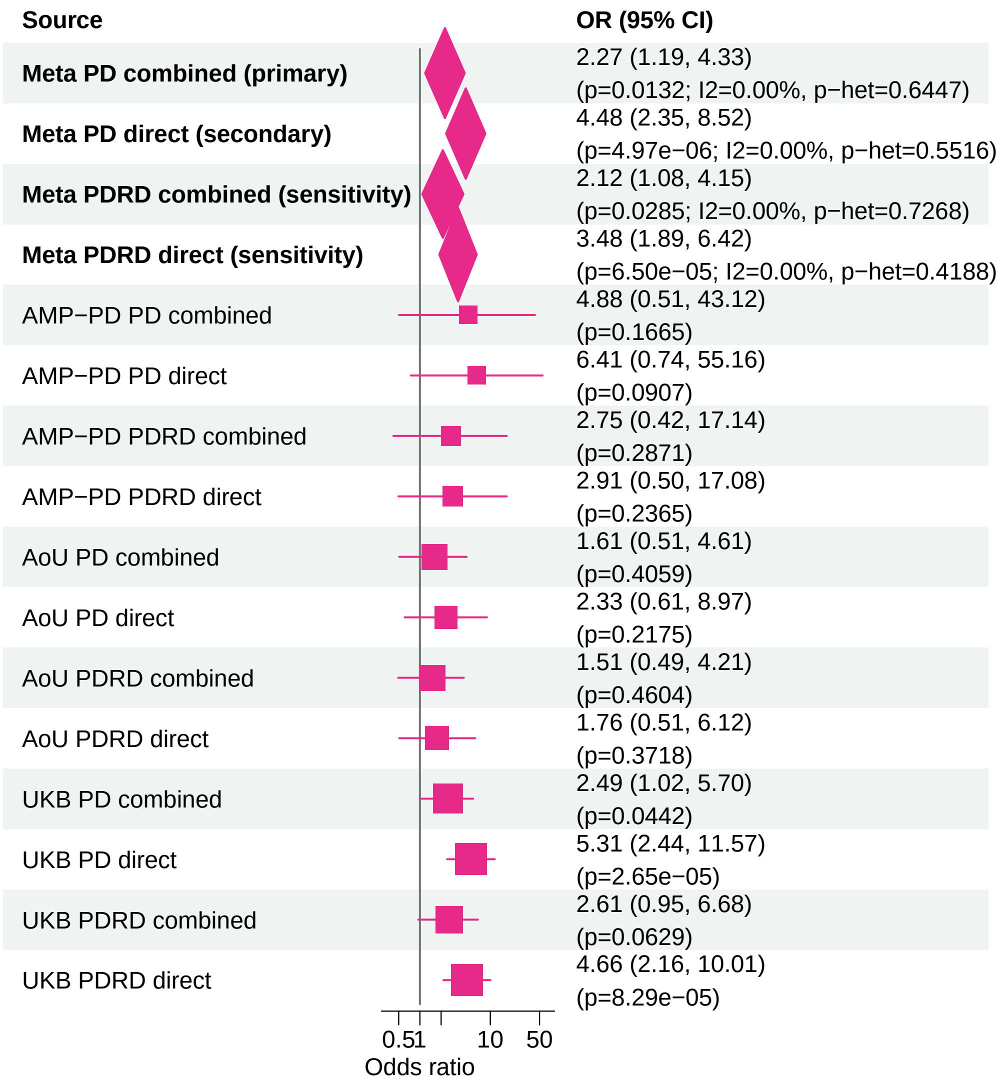


Meta OR/CI and P values are from the exact−kappa=0.5 pooled REGENIE fixed−effects analysis.

#### PD primary/secondary definitions with PDRD sensitivity − KRBA2 − LOF (ALL)

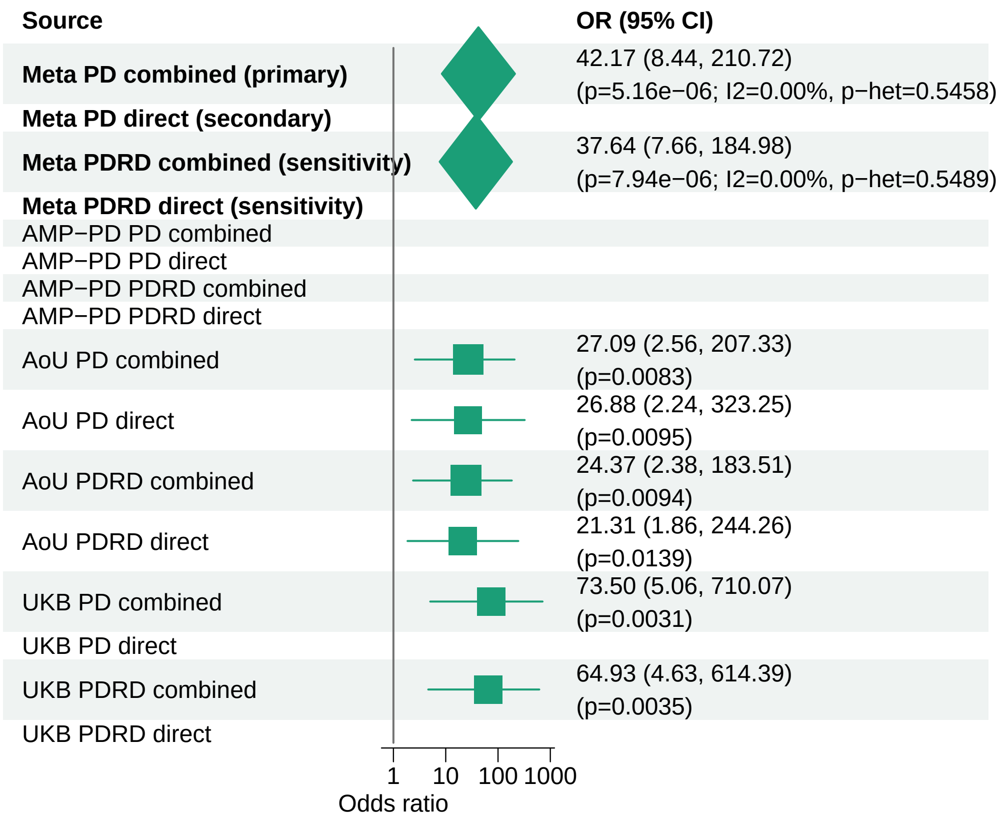


Meta OR/CI and P values are from the exact−kappa=0.5 pooled REGENIE fixed−effects analysis.

#### PD primary/secondary definitions with PDRD sensitivity − SQLE − LOF+REVEL50 (ALL)

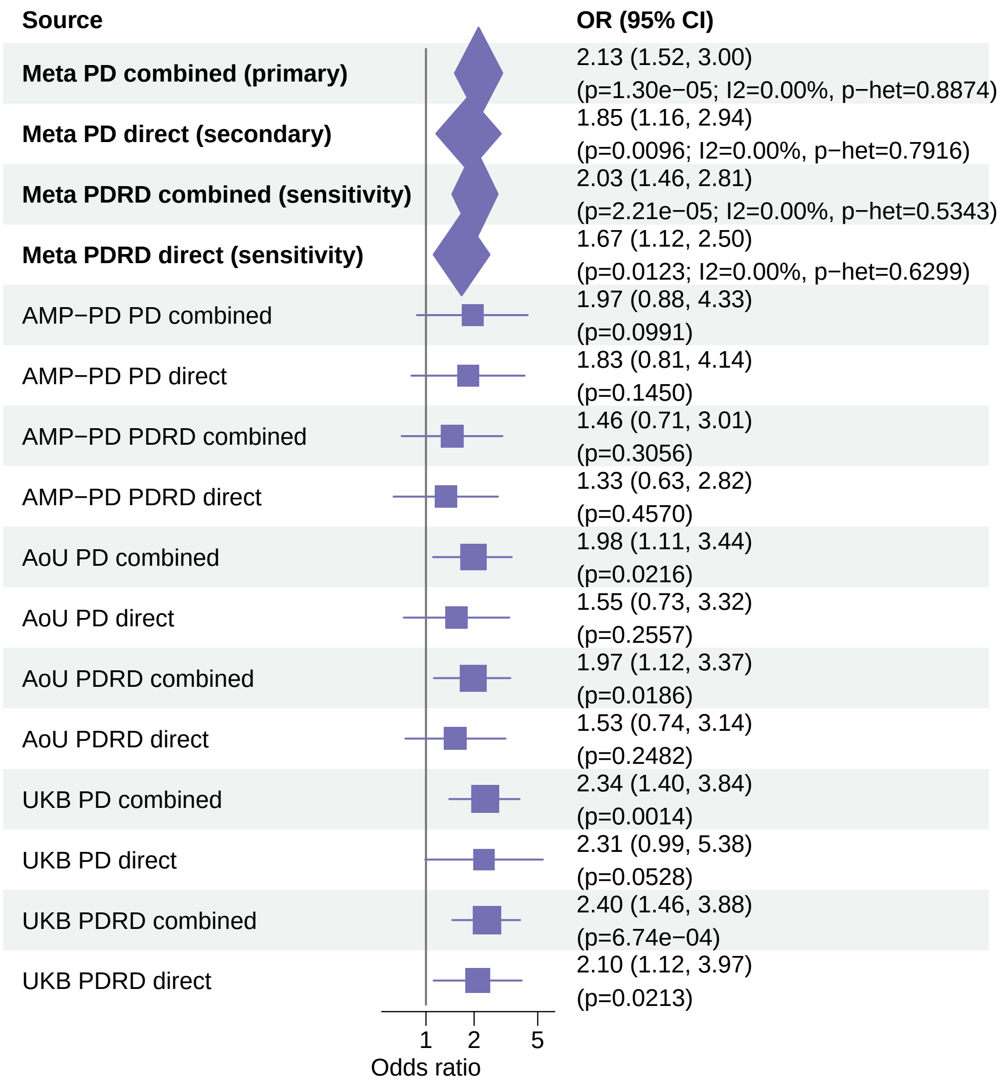


Meta OR/CI and P values are from the exact−kappa=0.5 pooled REGENIE fixed−effects analysis.

#### PD primary/secondary definitions with PDRD sensitivity − SPIRE2 − LOF+REVEL50 (ALL)

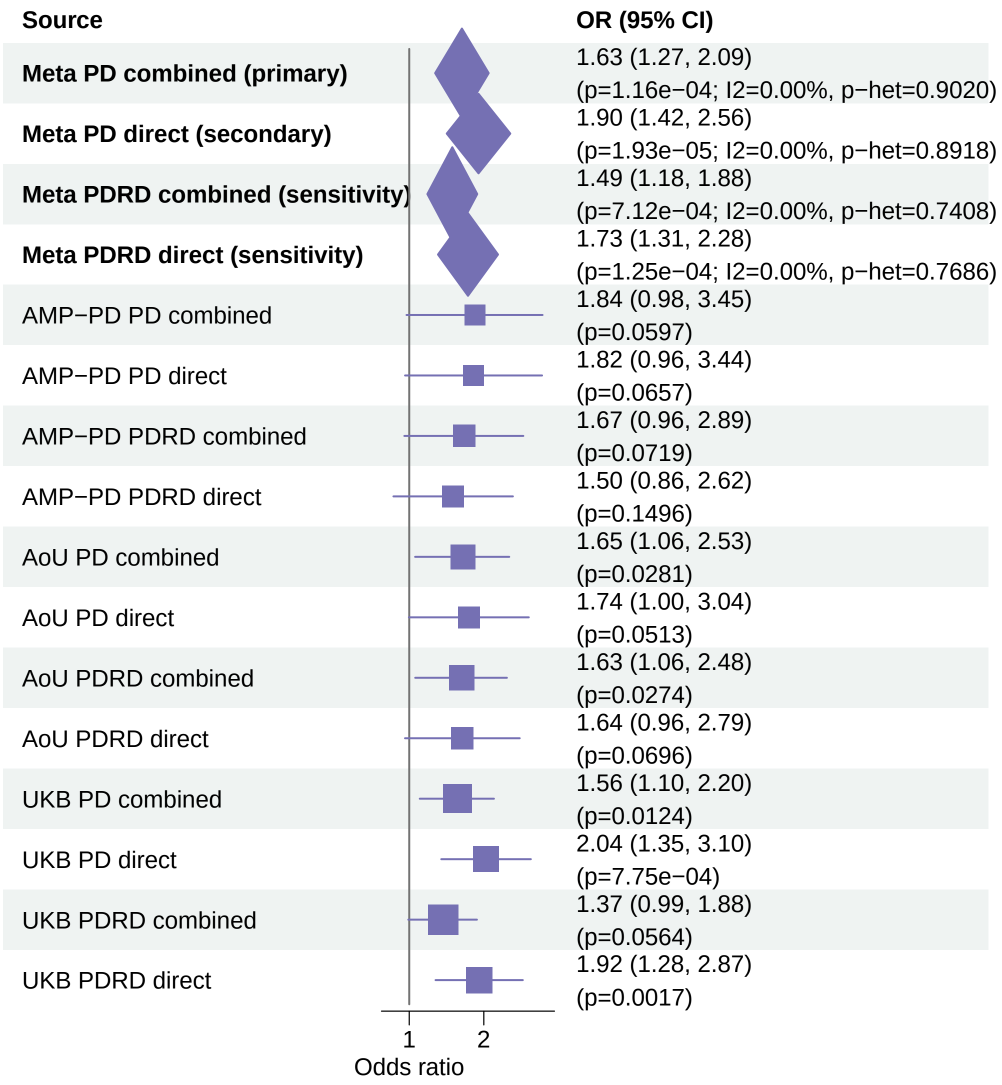


Meta OR/CI and P values are from the exact−kappa=0.5 pooled REGENIE fixed−effects analysis.

#### PD primary/secondary definitions with PDRD sensitivity − KRBA2 − LOF+REVEL50 (ALL)

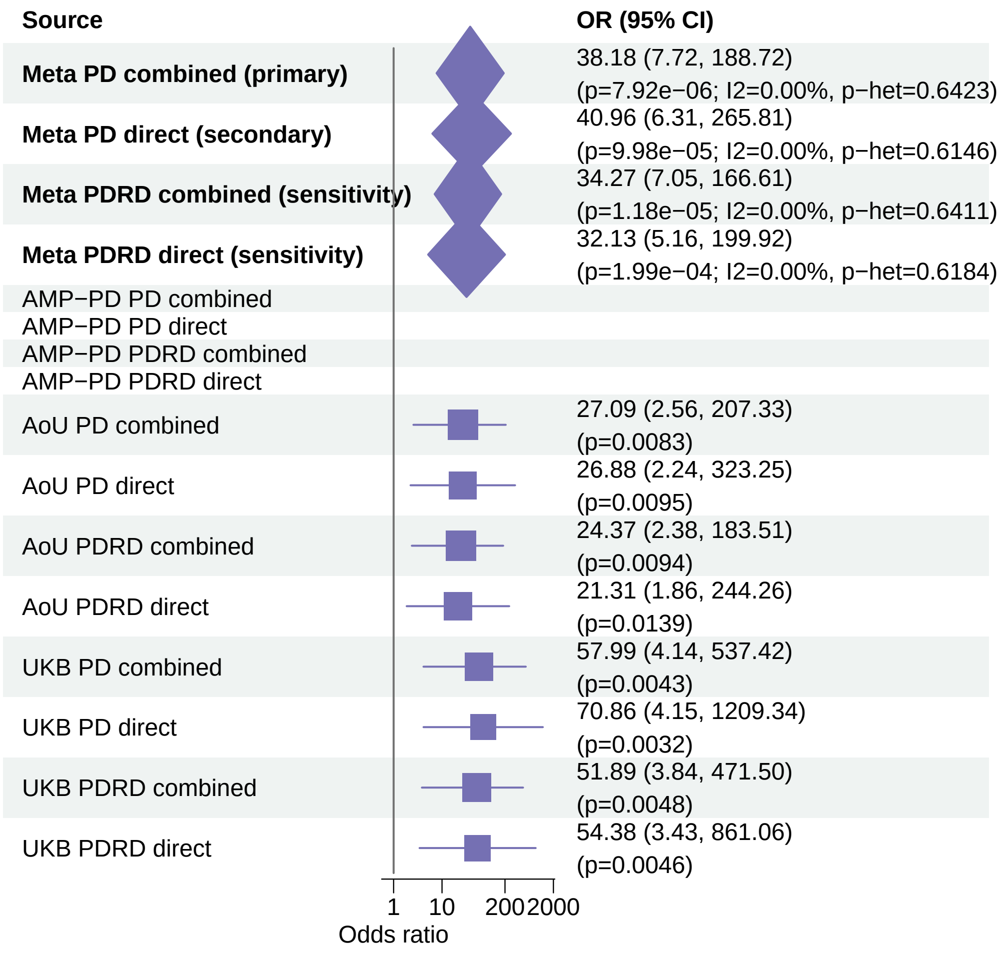


Meta OR/CI and P values are from the exact−kappa=0.5 pooled REGENIE fixed−effects analysis.

### PD primary/secondary definitions with PDRD sensitivity − AGFG1 − LOF+REVEL25 (ALL)

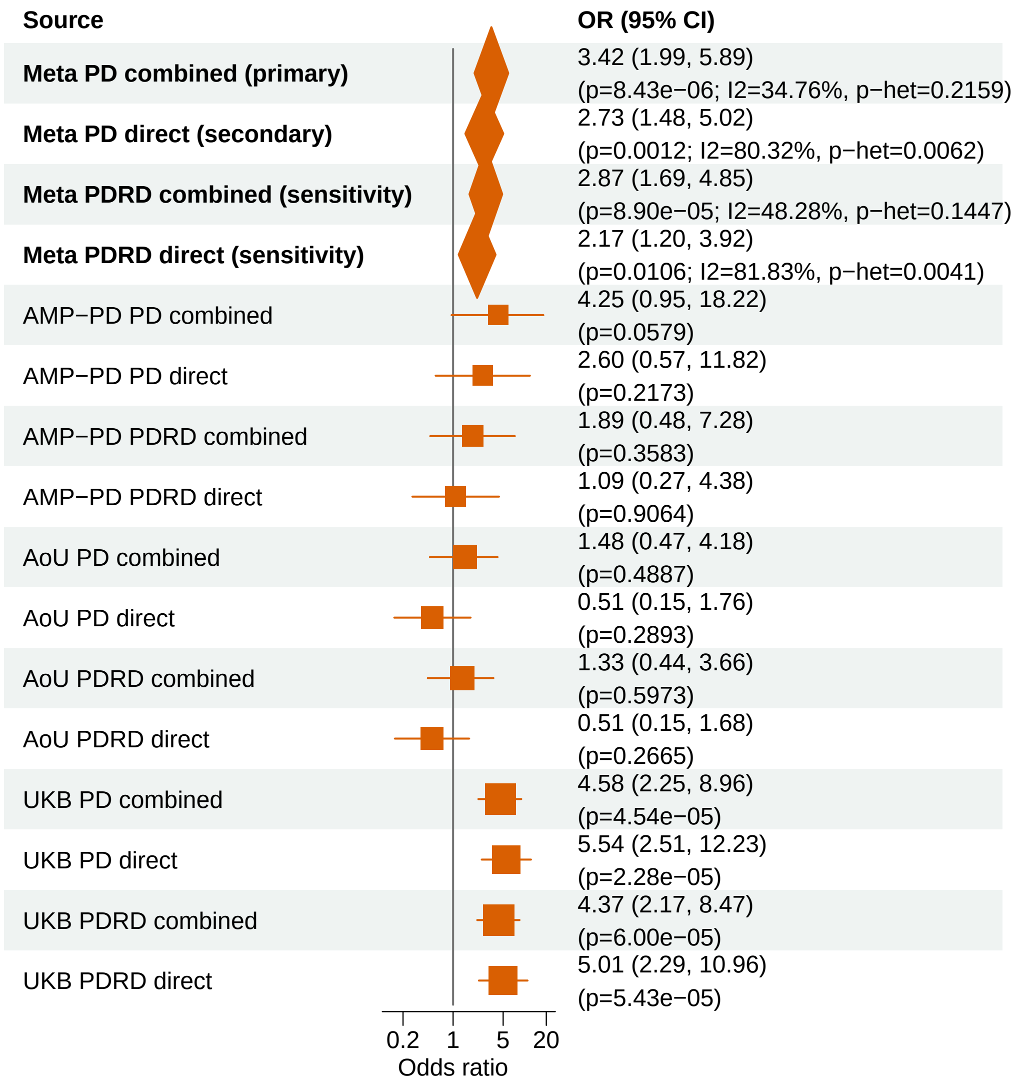


Meta OR/CI and P values are from the exact−kappa=0.5 pooled REGENIE fixed−effects analysis.

#### PD primary/secondary definitions with PDRD sensitivity − ATAD2B − LOF (ALL)

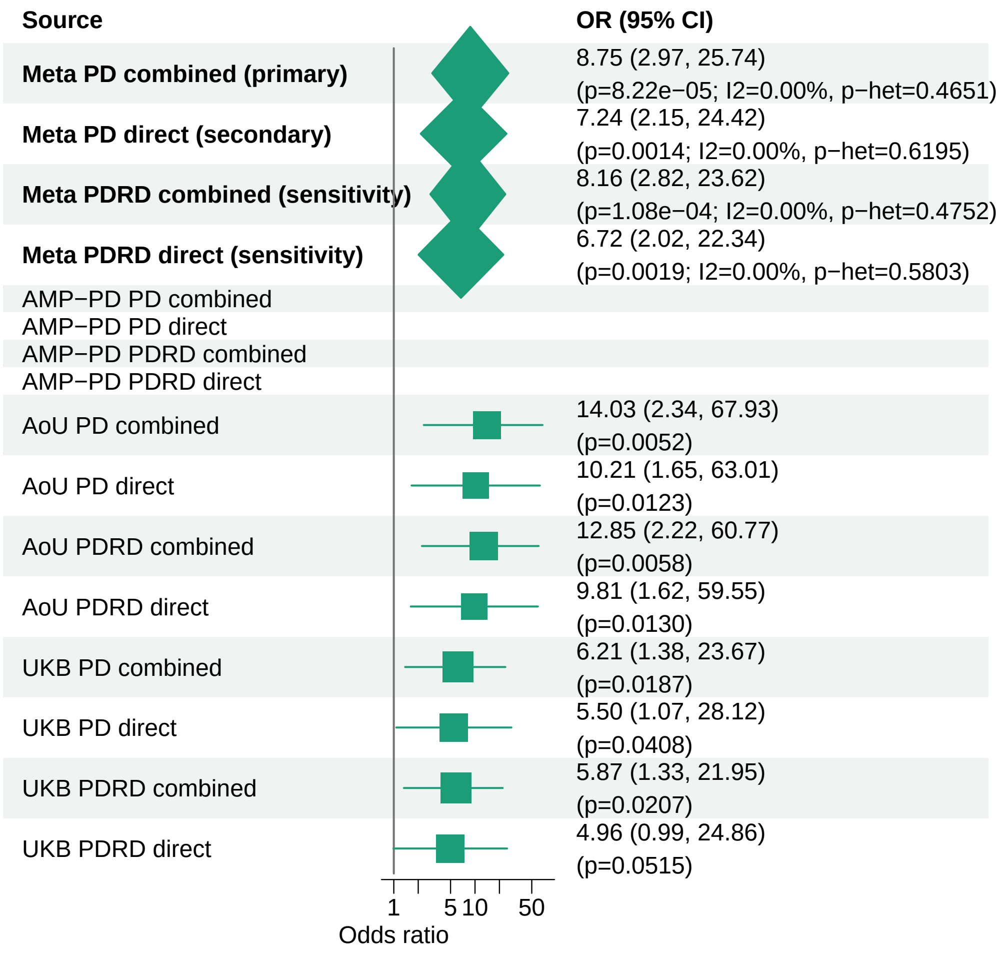


Meta OR/CI and P values are from the exact−kappa=0.5 pooled REGENIE fixed−effects analysis.

#### PD primary/secondary definitions with PDRD sensitivity − HOXA13 − LOF (ALL)

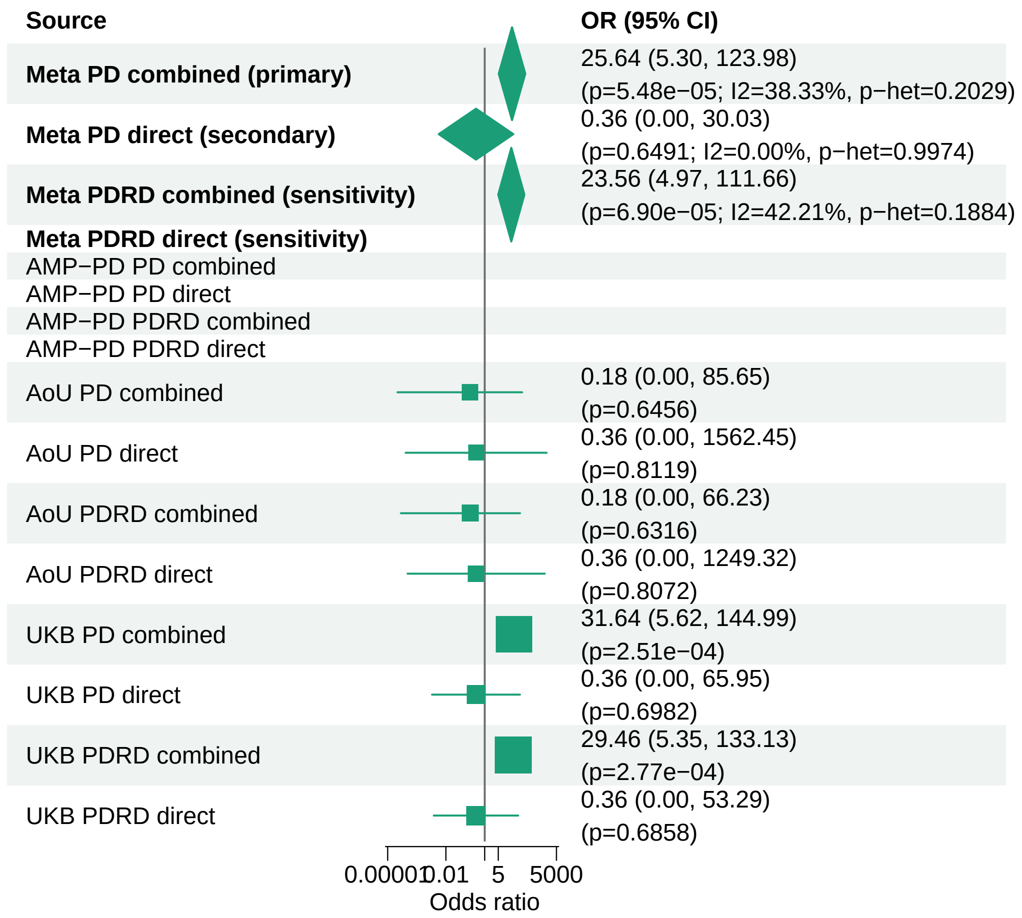


Meta OR/CI and P values are from the exact−kappa=0.5 pooled REGENIE fixed−effects analysis.
