## Supplementary_Fig4 for "Population-scale burden analysis of rare damaging coding variants identifies novel risk genes for Alzheimer’s disease and related dementias and Parkinson’s disease and related disorders"

### AD primary/secondary definitions with ADRD sensitivity − PSEN1 − LOF+REVEL75 (EUR)

| Source | OR (95% CI) |
|---|---|
| **Meta AD combined (primary)** | 6.49 (4.81, 8.74) (overall p=2.22e−33; I2=96.08%, p−het=8.20e−12) |
| **Meta AD direct (secondary)** | 12.43 (8.78, 17.59) (overall p=4.78e−33; I2=81.74%, p−het=0.0042) |
| **Meta ADRD combined (sensitivity)** | 4.93 (3.75, 6.48) (overall p=2.35e−29; I2=97.13%, p−het=7.17e−16) |
| **Meta ADRD direct (sensitivity)** | 8.92 (6.53, 12.17) (overall p=4.20e−35; I2=92.31%, p−het=2.25e−06) |
| ADES | 1.21 (0.61, 2.42) (p=0.5799) |
| ADSP | 17.89 (11.87, 26.97) (p=3.72e−43) |
| AoU AD combined | 3.55 (1.20, 10.10) (p=0.0220) |
| AoU AD direct | 14.39 (0.33, 617.92) (p=0.1646) |
| AoU ADRD combined | 3.18 (1.19, 8.05) (p=0.0218) |
| AoU ADRD direct | 3.73 (0.44, 31.71) (p=0.2275) |
| UKB AD combined | 1.86 (1.15, 2.98) (p=0.0117) |
| UKB AD direct | 4.81 (2.48, 9.32) (p=3.29e−06) |
| UKB ADRD combined | 1.60 (1.07, 2.36) (p=0.0220) |
| UKB ADRD direct | 3.47 (2.12, 5.65) (p=6.48e−07) |

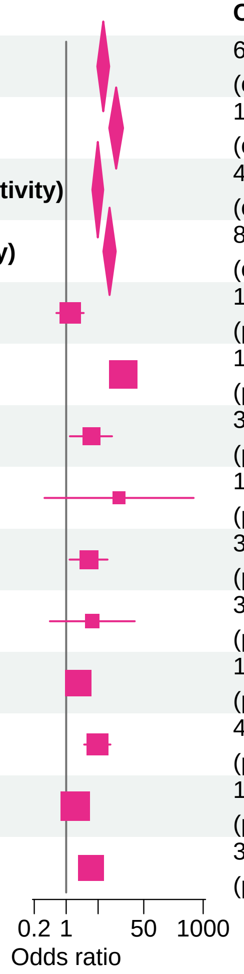


Meta OR/CI values are pooled REGENIE effects. Meta overall P values use ADES+REGENIE signed−Z evidence when the exact ADES mask was available; otherwise they use REGENIE.

#### AD primary/secondary definitions with ADRD sensitivity − TREM2 − LOF+REVEL25 (EUR)

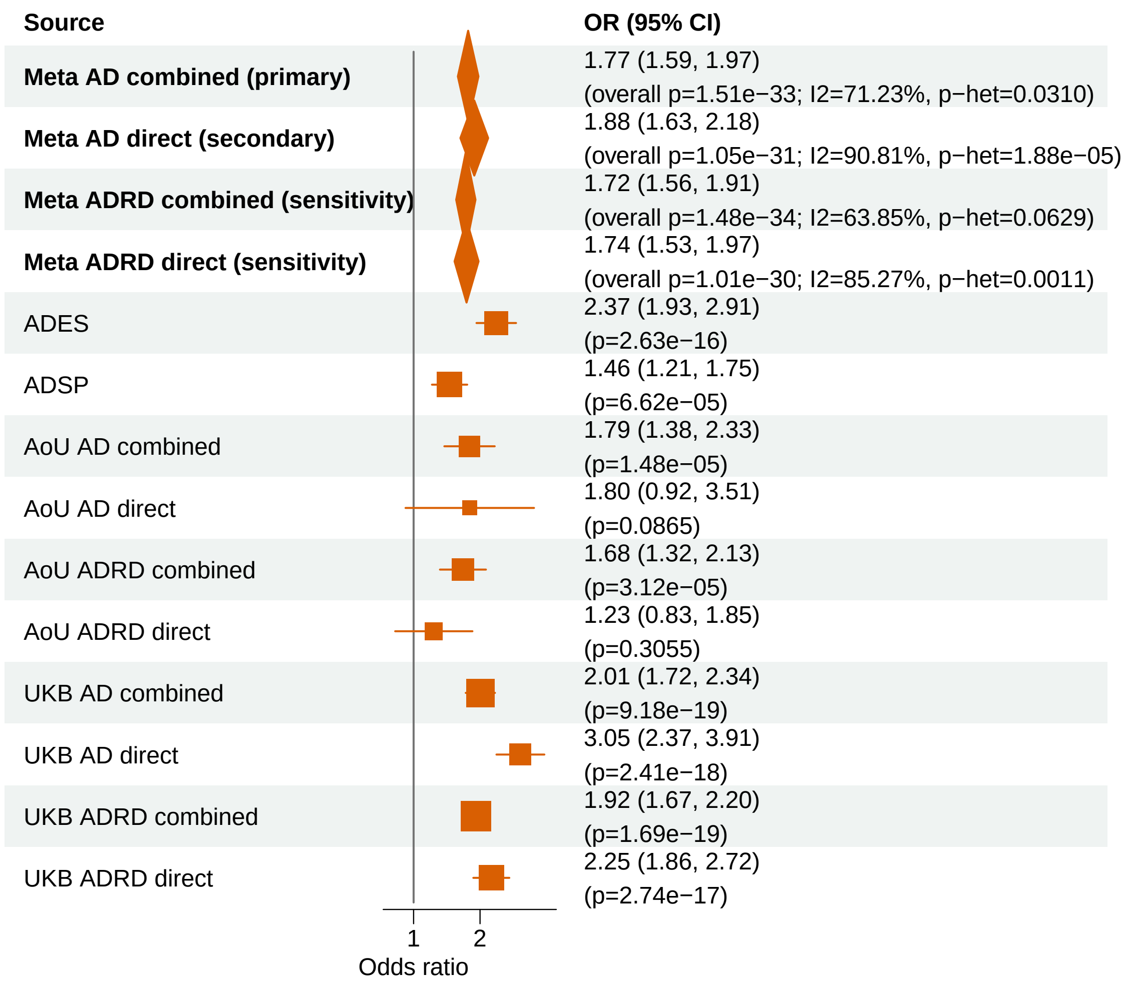


Meta OR/CI values are pooled REGENIE effects. Meta overall P values use ADES+REGENIE signed−Z evidence when the exact ADES mask was available; otherwise they use REGENIE.

#### AD primary/secondary definitions with ADRD sensitivity − ABCA7 − LOF+REVEL25 (EUR)

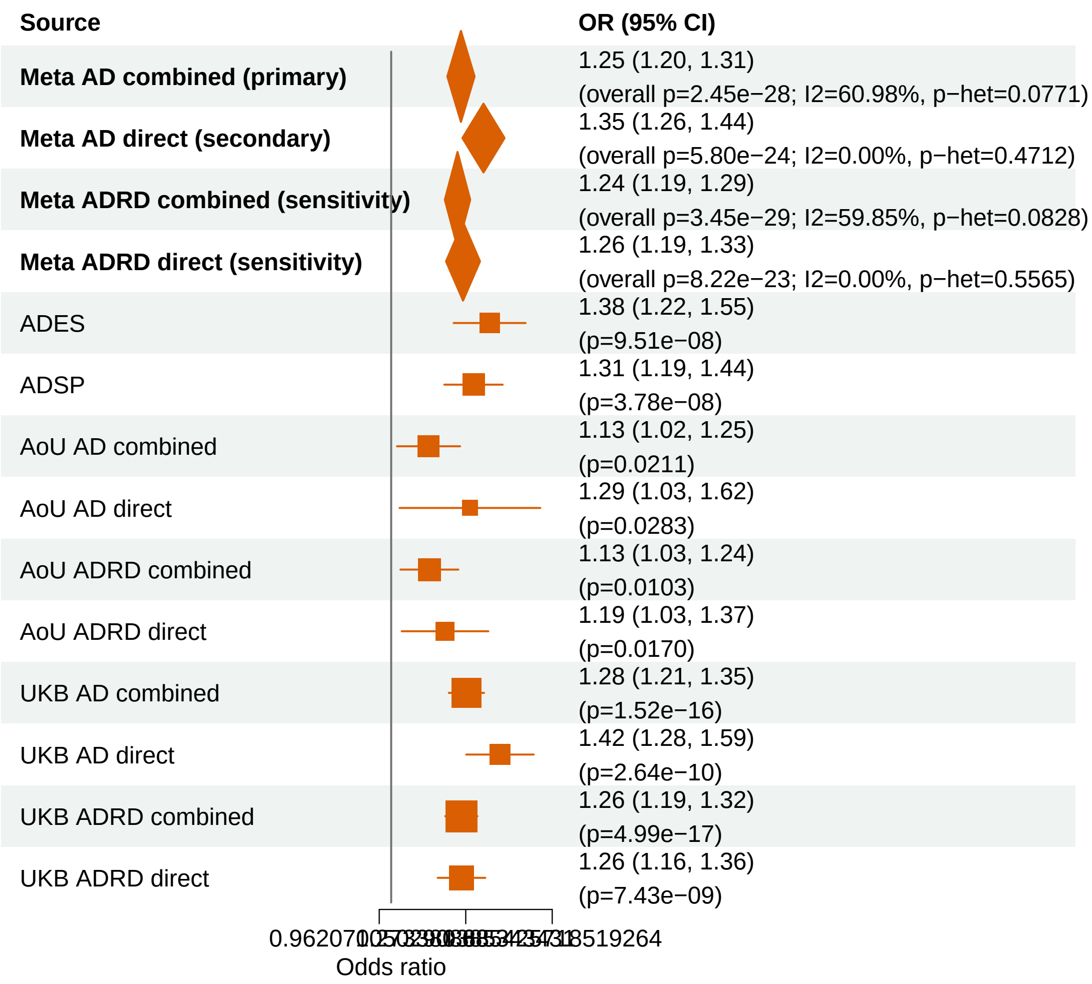


Meta OR/CI values are pooled REGENIE effects. Meta overall P values use ADES+REGENIE signed−Z evidence when the exact ADES mask was available; otherwise they use REGENIE.

#### AD primary/secondary definitions with ADRD sensitivity − PSEN1 − LOF+REVEL50 (EUR)

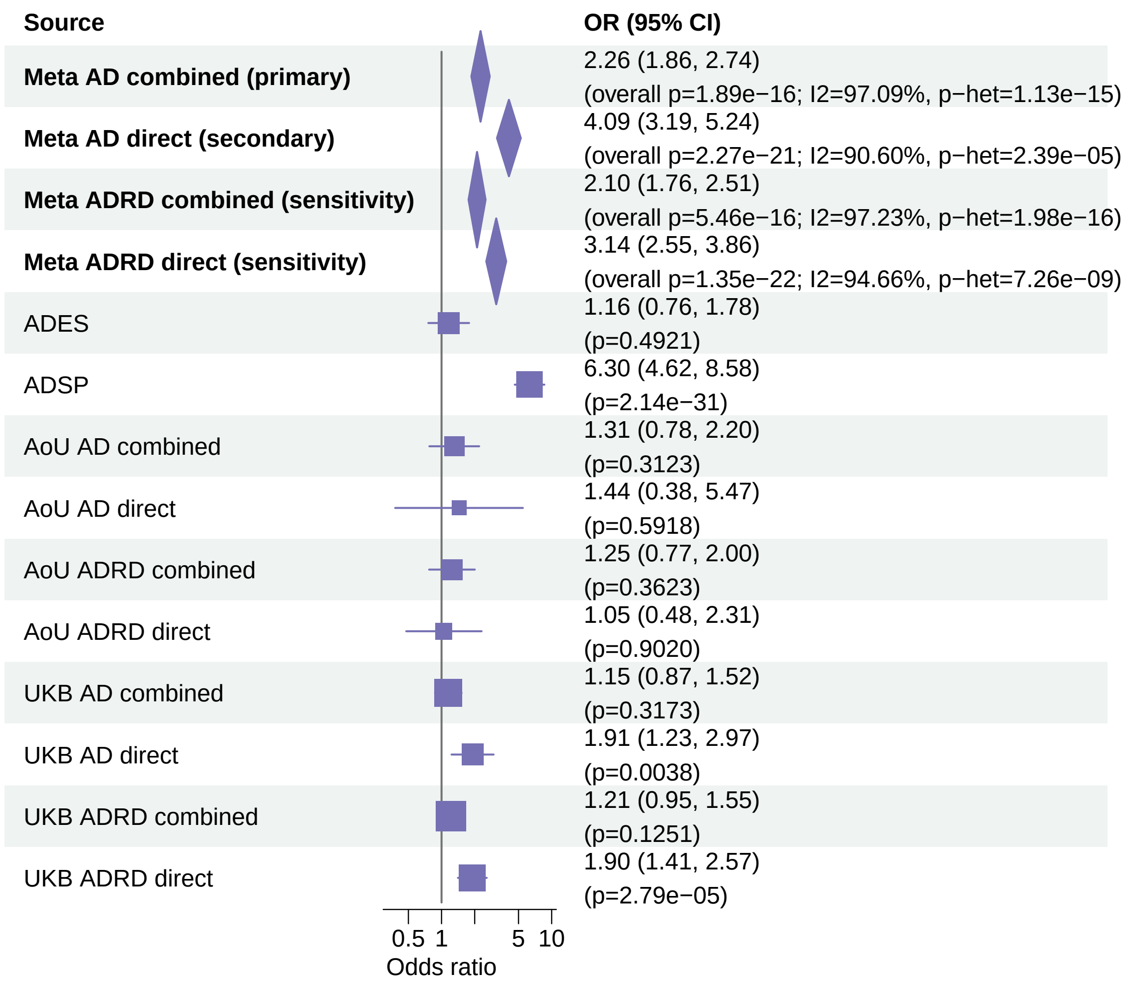


Meta OR/CI values are pooled REGENIE effects. Meta overall P values use ADES+REGENIE signed−Z evidence when the exact ADES mask was available; otherwise they use REGENIE.

### AD primary/secondary definitions with ADRD sensitivity − ABCA7 − LOF+REVEL75 (EUR)

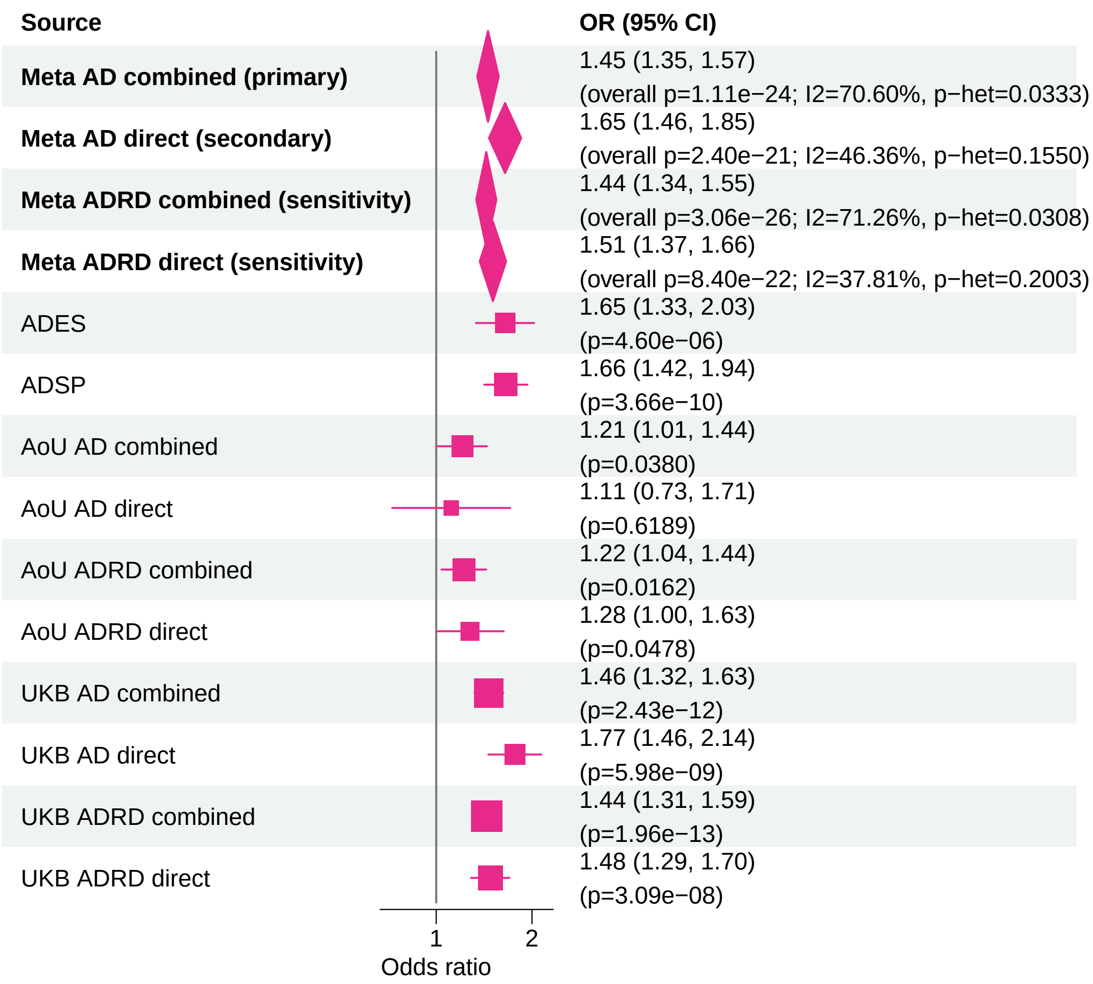


Meta OR/CI values are pooled REGENIE effects. Meta overall P values use ADES+REGENIE signed−Z evidence when the exact ADES mask was available; otherwise they use REGENIE.

#### AD primary/secondary definitions with ADRD sensitivity − PSEN1 − LOF+REVEL25 (EUR)

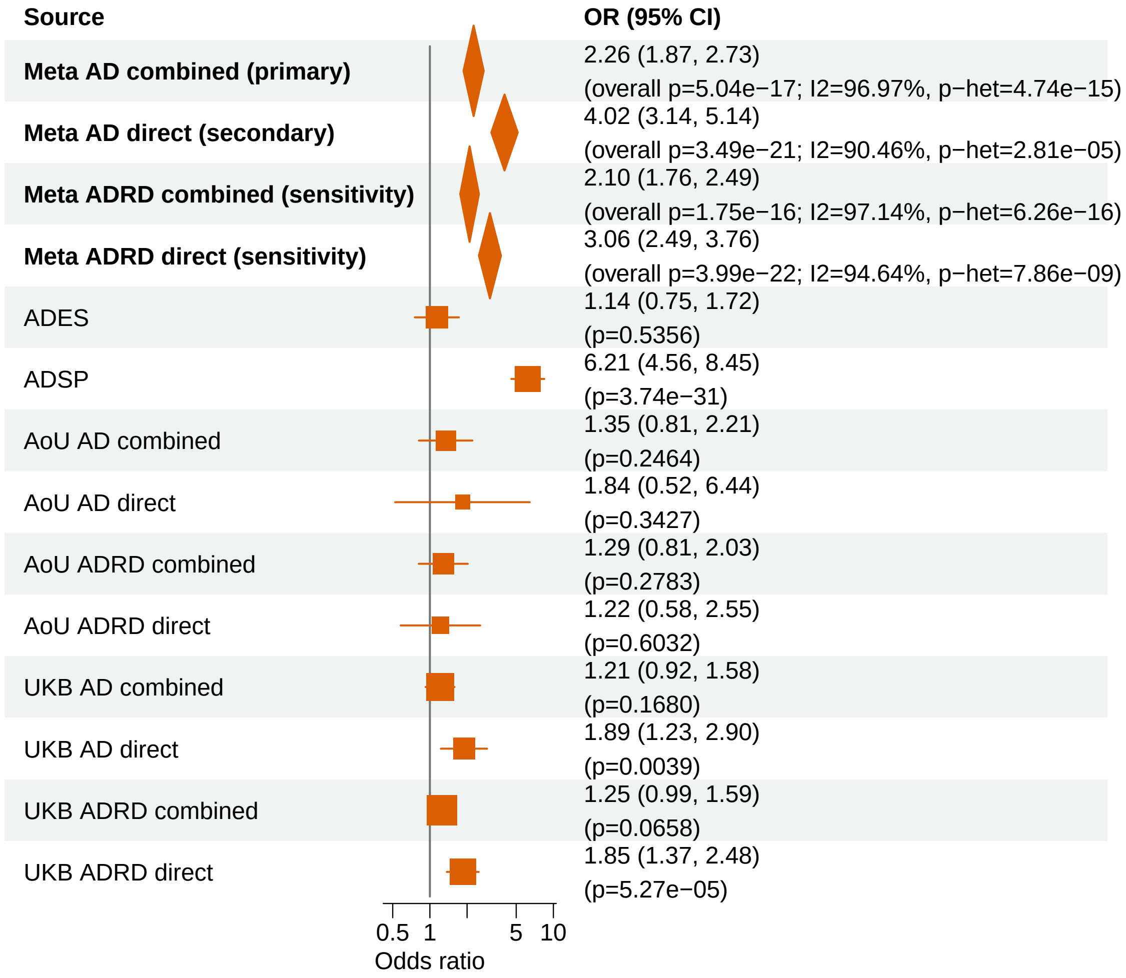


Meta OR/CI values are pooled REGENIE effects. Meta overall P values use ADES+REGENIE signed−Z evidence when the exact ADES mask was available; otherwise they use REGENIE.

#### AD primary/secondary definitions with ADRD sensitivity − ABCA7 − LOF+REVEL50 (EUR)

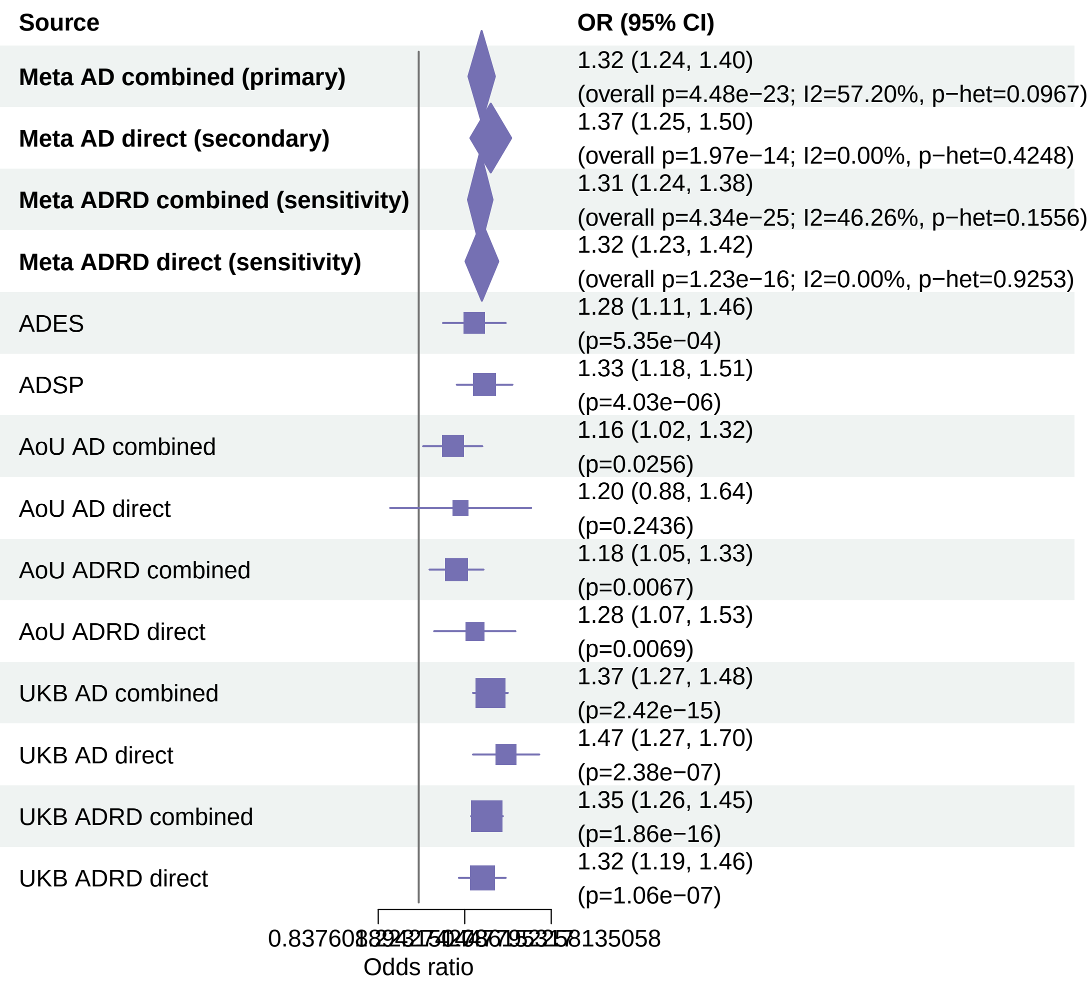


Meta OR/CI values are pooled REGENIE effects. Meta overall P values use ADES+REGENIE signed−Z evidence when the exact ADES mask was available; otherwise they use REGENIE.

#### AD primary/secondary definitions with ADRD sensitivity − SORL1 − LOF+REVEL50 (EUR)

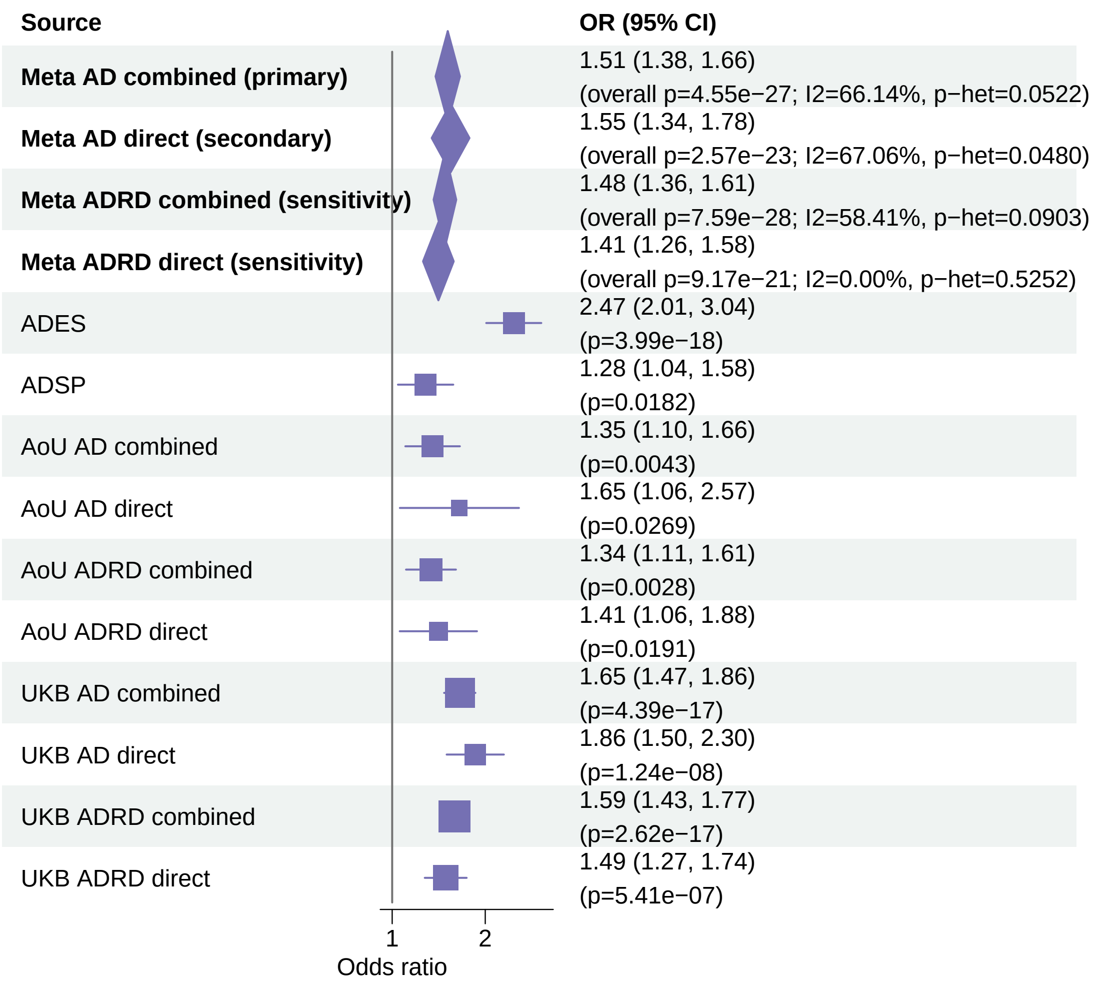


Meta OR/CI values are pooled REGENIE effects. Meta overall P values use ADES+REGENIE signed−Z evidence when the exact ADES mask was available; otherwise they use REGENIE.

#### AD primary/secondary definitions with ADRD sensitivity − SORL1 − LOF+REVEL75 (EUR)

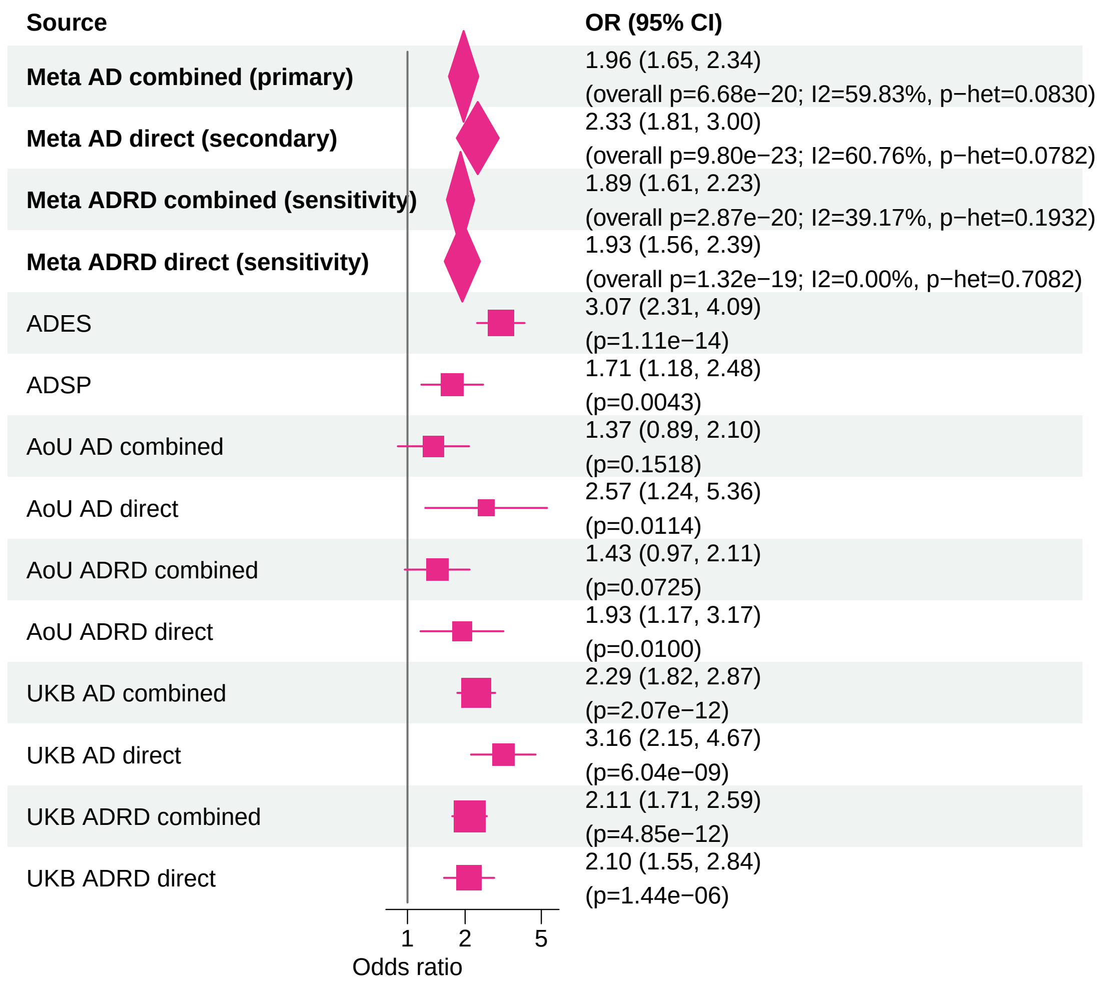


Meta OR/CI values are pooled REGENIE effects. Meta overall P values use ADES+REGENIE signed−Z evidence when the exact ADES mask was available; otherwise they use REGENIE.

#### AD primary/secondary definitions with ADRD sensitivity − SORL1 − LOF (EUR)

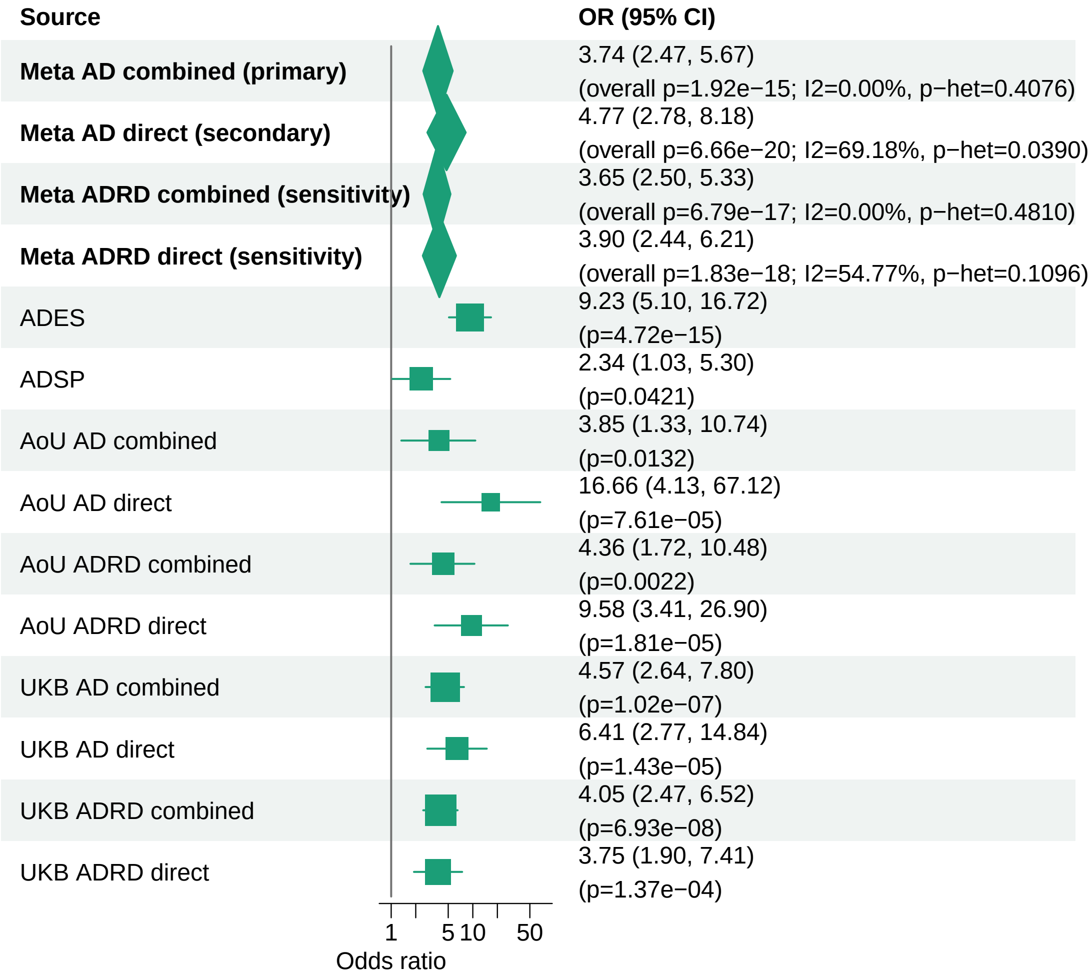


Meta OR/CI values are pooled REGENIE effects. Meta overall P values use ADES+REGENIE signed−Z evidence when the exact ADES mask was available; otherwise they use REGENIE.

### AD primary/secondary definitions with ADRD sensitivity − ABCA7 − LOF (EUR)

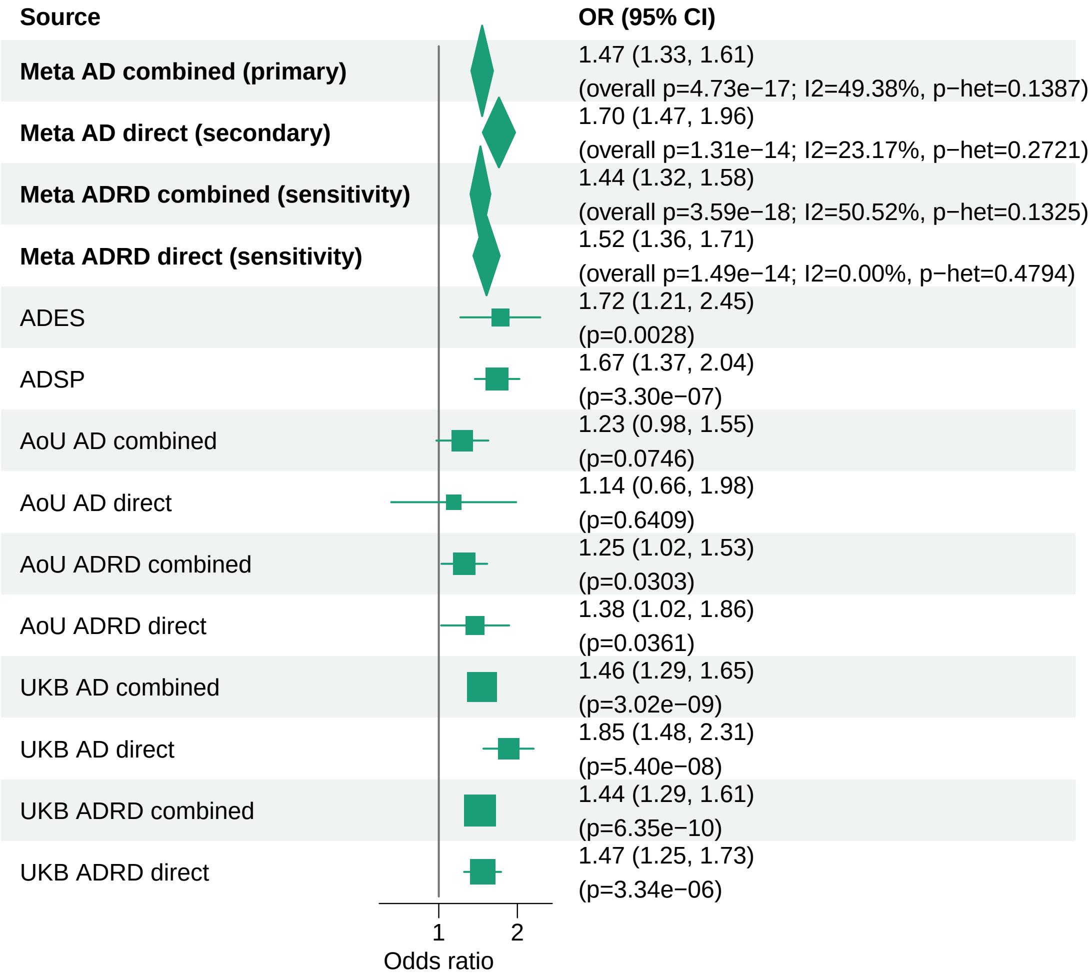


Meta OR/CI values are pooled REGENIE effects. Meta overall P values use ADES+REGENIE signed−Z evidence when the exact ADES mask was available; otherwise they use REGENIE.

#### AD primary/secondary definitions with ADRD sensitivity − GRN − LOF (EUR)

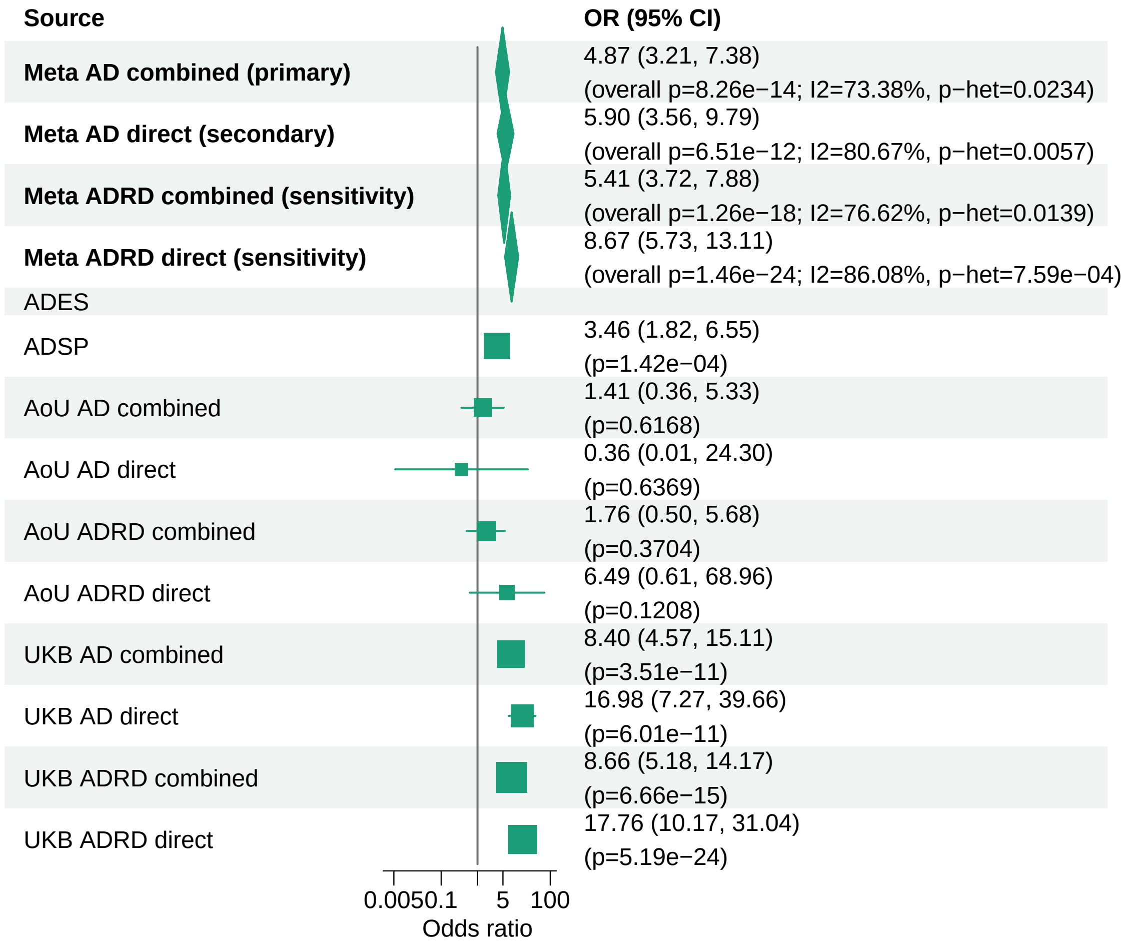


Meta OR/CI values are pooled REGENIE effects. Meta overall P values use ADES+REGENIE signed−Z evidence when the exact ADES mask was available; otherwise they use REGENIE.

#### AD primary/secondary definitions with ADRD sensitivity − SORL1 − LOF+REVEL25 (EUR)

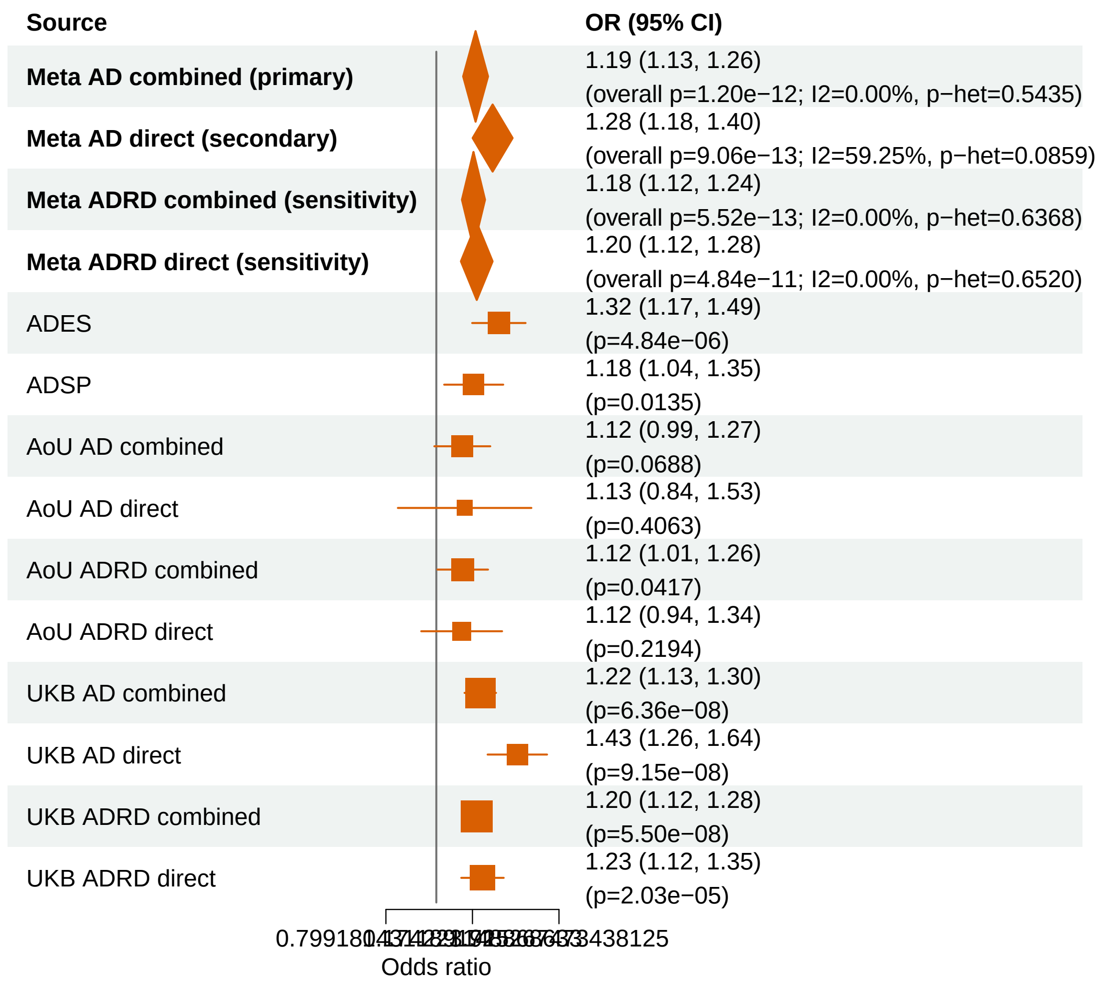


Meta OR/CI values are pooled REGENIE effects. Meta overall P values use ADES+REGENIE signed−Z evidence when the exact ADES mask was available; otherwise they use REGENIE.

#### AD primary/secondary definitions with ADRD sensitivity − GRN − LOF+REVEL75 (EUR)

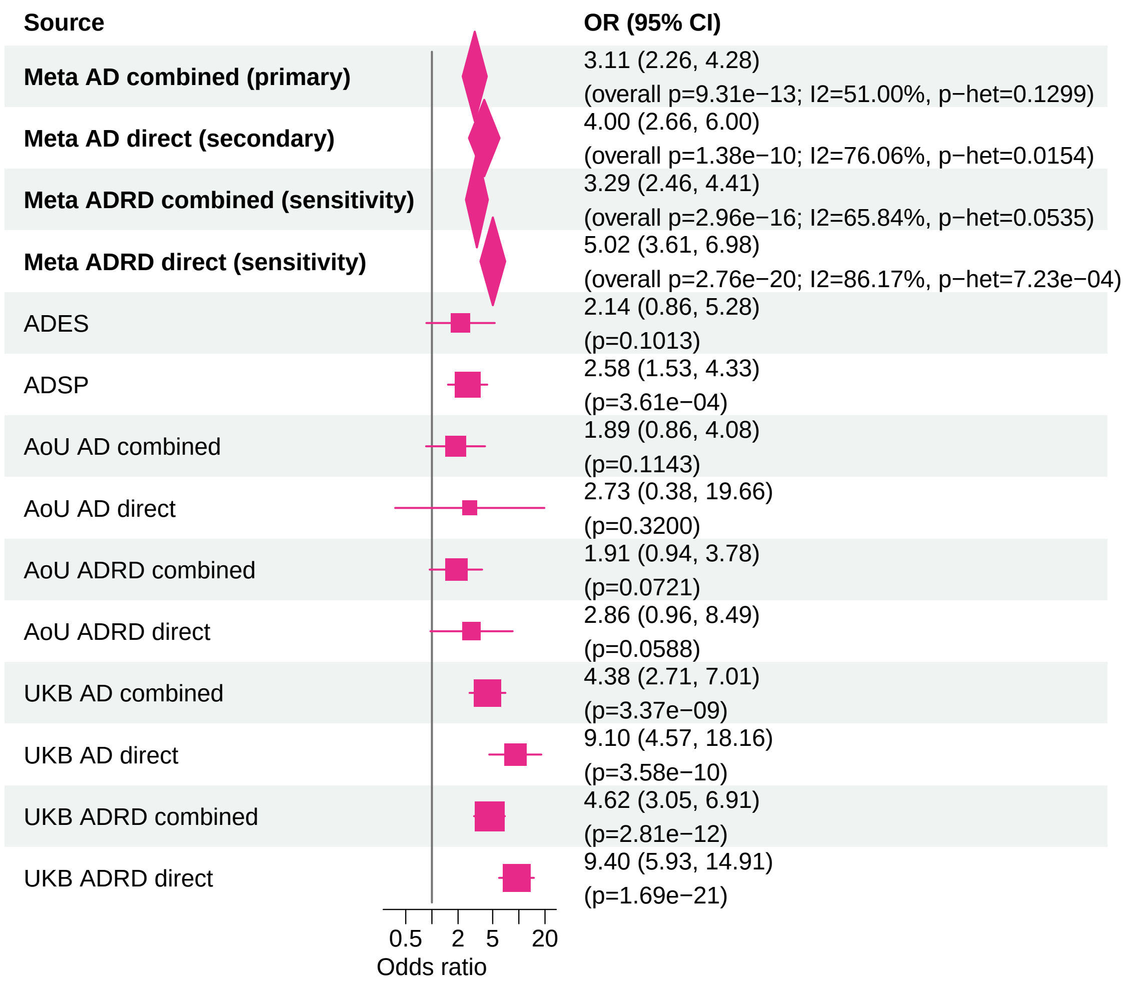


Meta OR/CI values are pooled REGENIE effects. Meta overall P values use ADES+REGENIE signed−Z evidence when the exact ADES mask was available; otherwise they use REGENIE.

### AD primary/secondary definitions with ADRD sensitivity − ATP8B4 − LOF+REVEL50 (EUR)

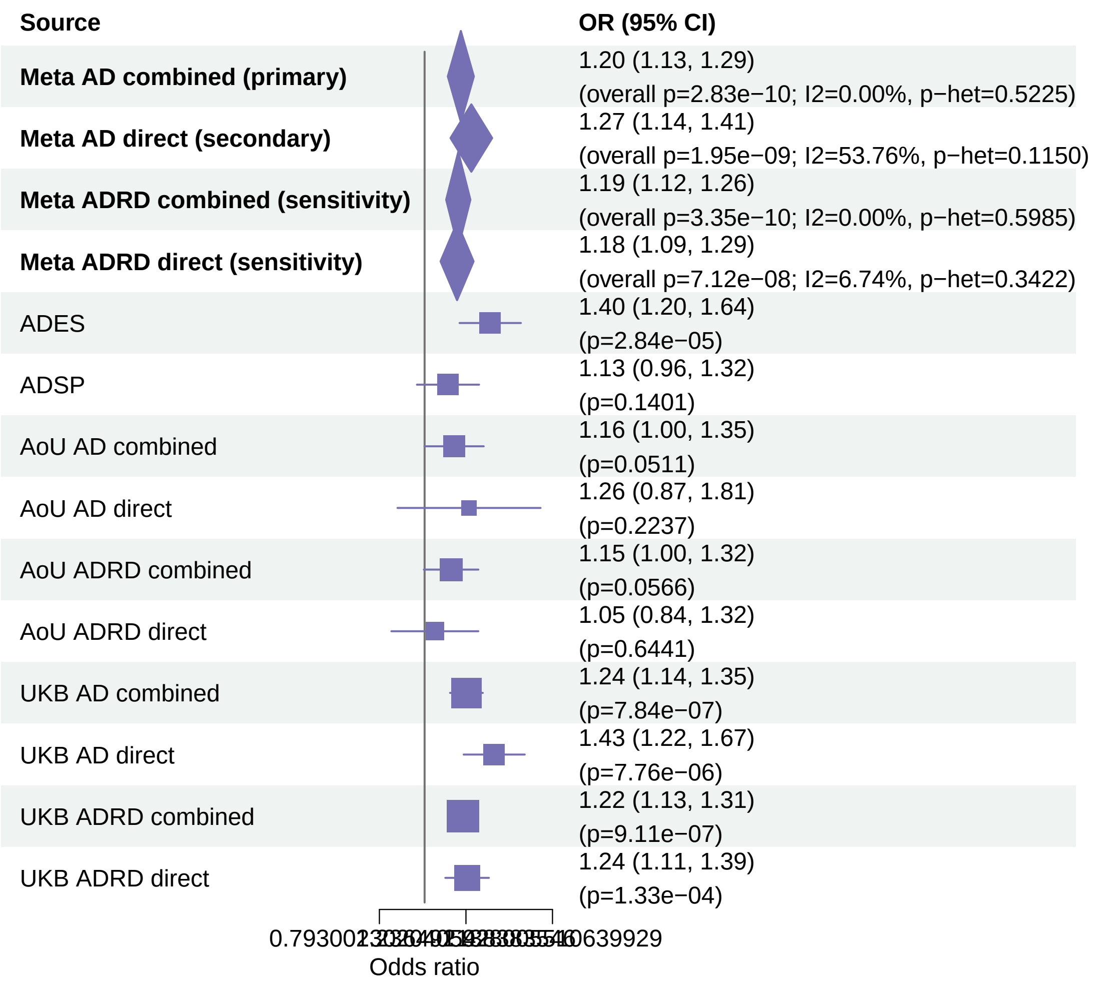


Meta OR/CI values are pooled REGENIE effects. Meta overall P values use ADES+REGENIE signed−Z evidence when the exact ADES mask was available; otherwise they use REGENIE.

#### AD primary/secondary definitions with ADRD sensitivity − TREM2 − LOF+REVEL50 (EUR)

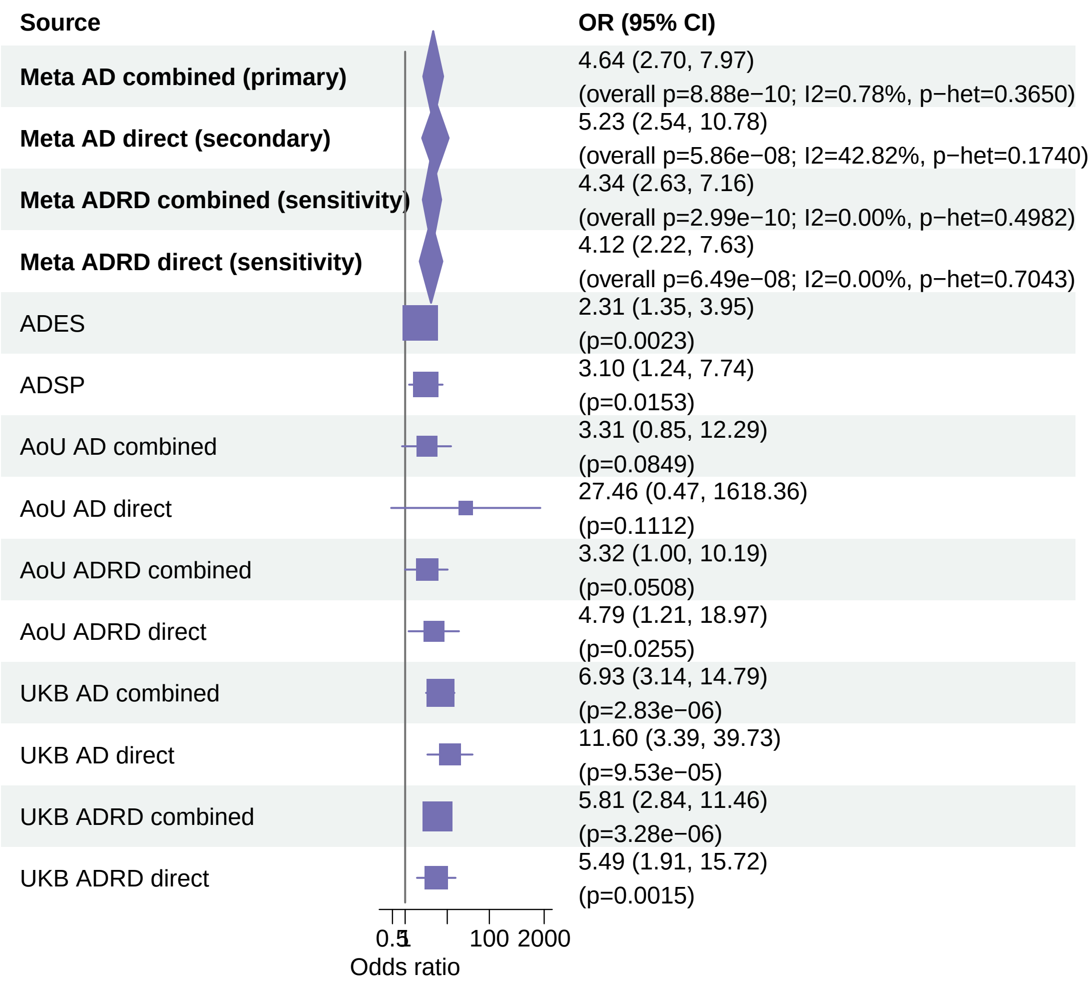


Meta OR/CI values are pooled REGENIE effects. Meta overall P values use ADES+REGENIE signed−Z evidence when the exact ADES mask was available; otherwise they use REGENIE.

### AD primary/secondary definitions with ADRD sensitivity − ATP8B4 − LOF+REVEL75 (EUR)

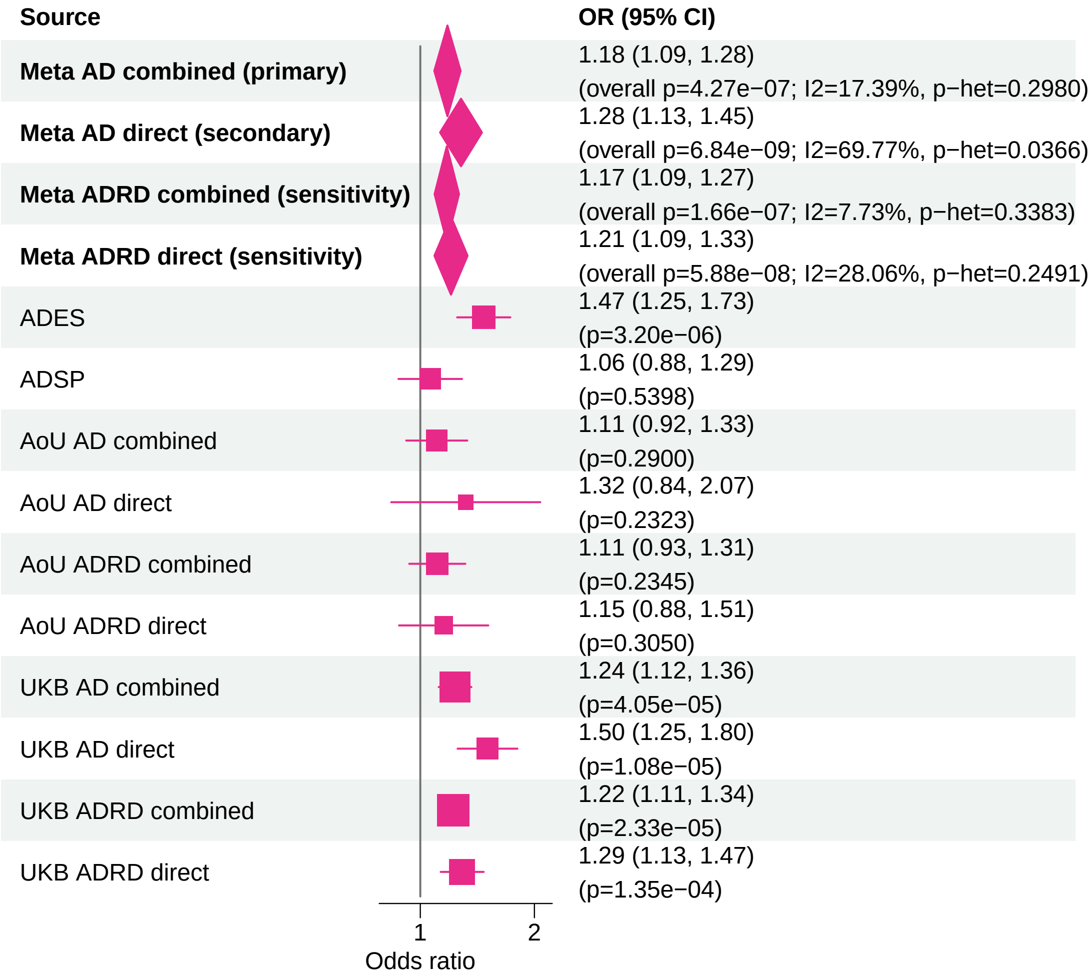


Meta OR/CI values are pooled REGENIE effects. Meta overall P values use ADES+REGENIE signed−Z evidence when the exact ADES mask was available; otherwise they use REGENIE.

#### AD primary/secondary definitions with ADRD sensitivity − ATP8B4 − LOF+REVEL25 (EUR)

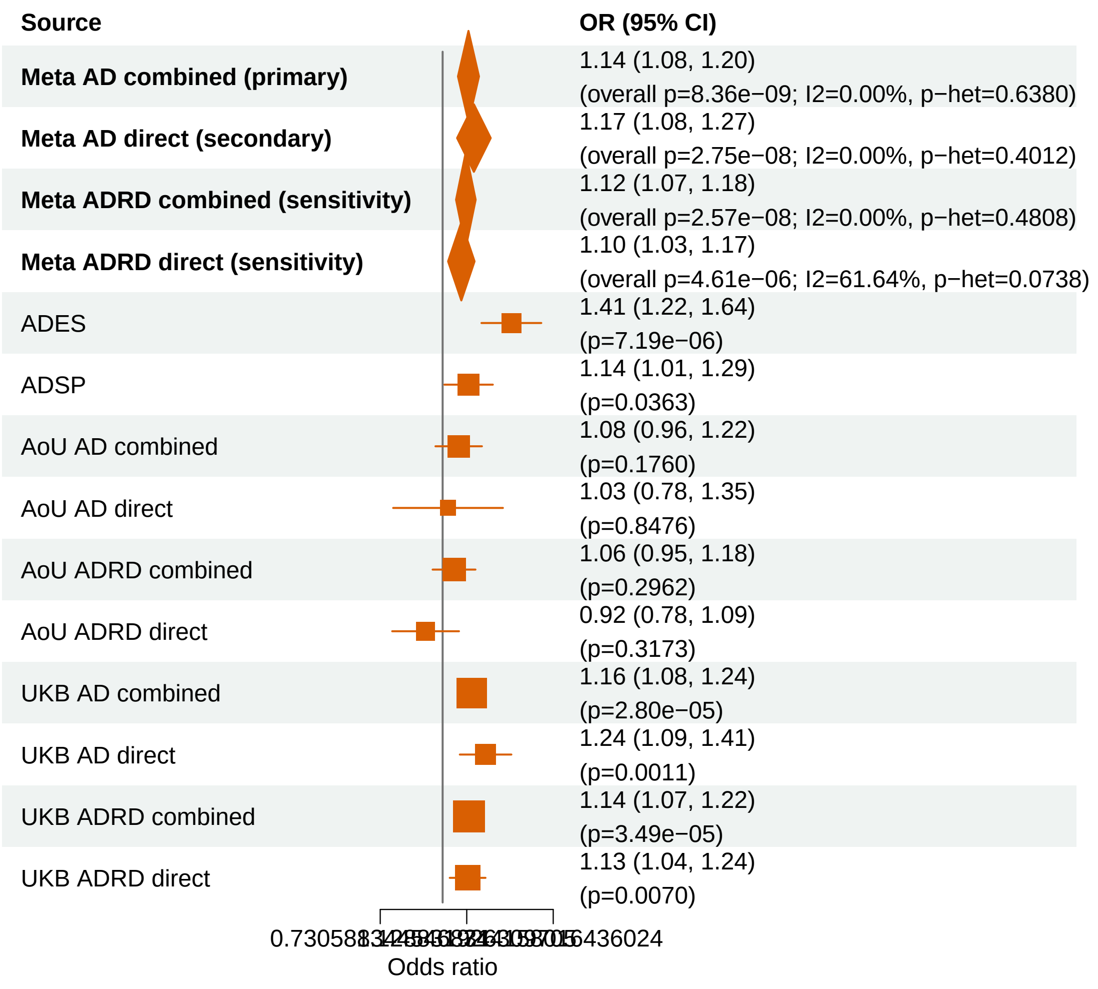


Meta OR/CI values are pooled REGENIE effects. Meta overall P values use ADES+REGENIE signed−Z evidence when the exact ADES mask was available; otherwise they use REGENIE.

#### AD primary/secondary definitions with ADRD sensitivity − TREM2 − LOF (EUR)

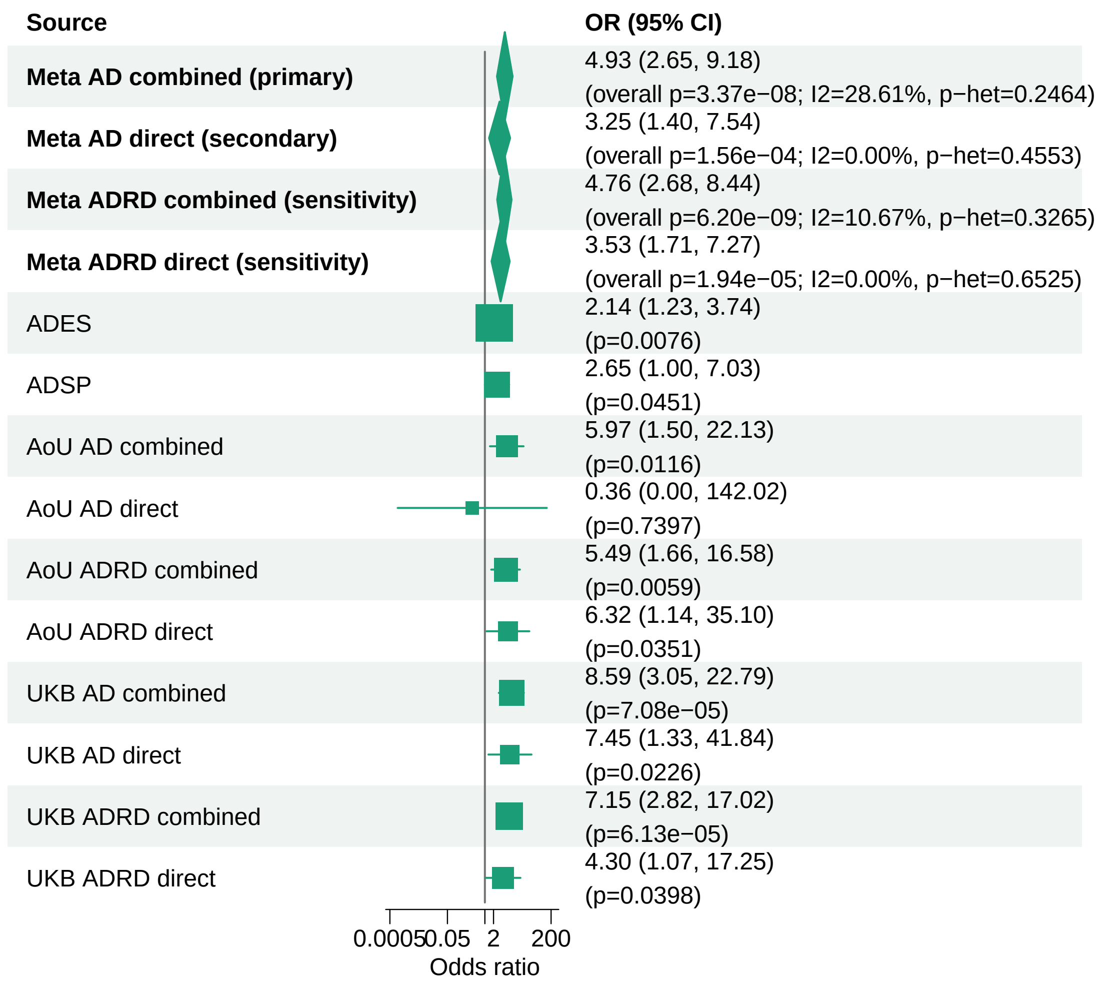


Meta OR/CI values are pooled REGENIE effects. Meta overall P values use ADES+REGENIE signed−Z evidence when the exact ADES mask was available; otherwise they use REGENIE.

#### AD primary/secondary definitions with ADRD sensitivity − ABCA1 − LOF+REVEL75 (EUR)

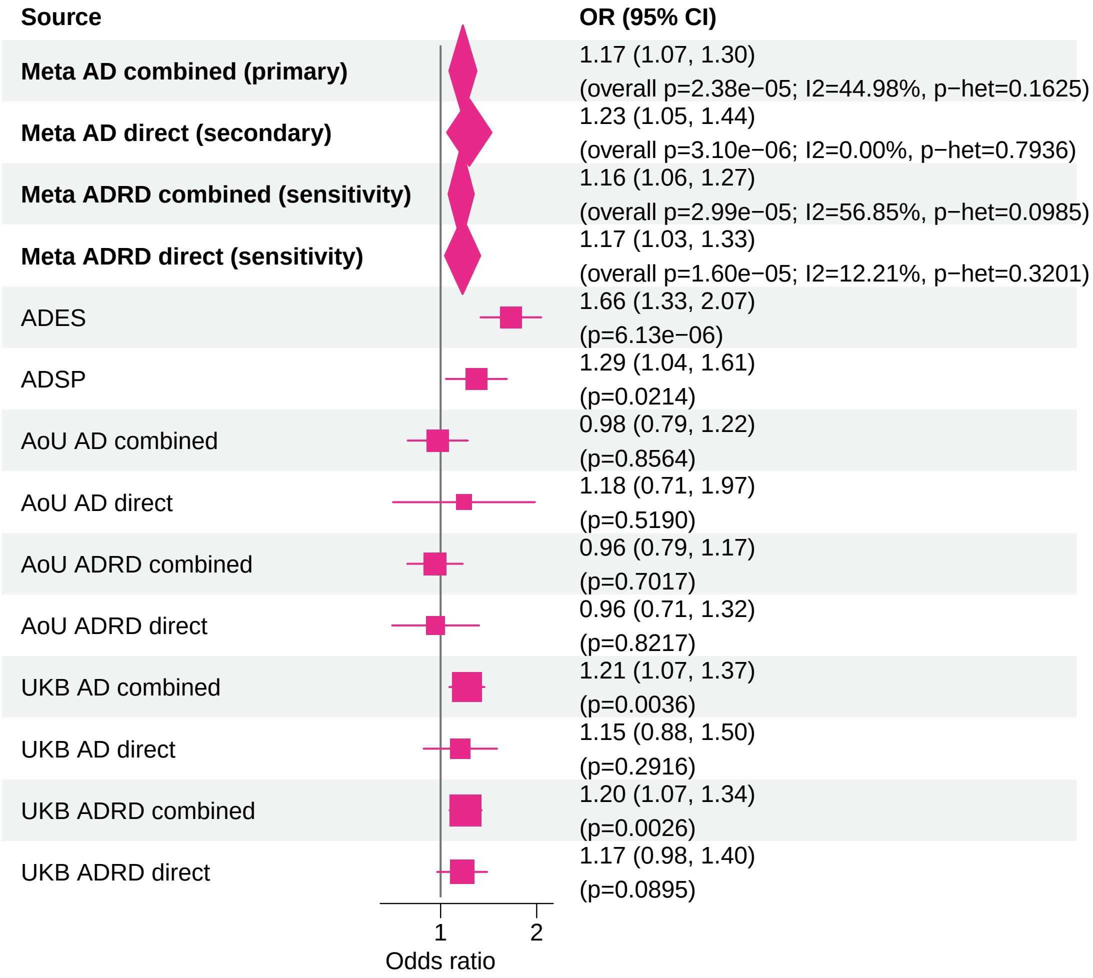


Meta OR/CI values are pooled REGENIE effects. Meta overall P values use ADES+REGENIE signed−Z evidence when the exact ADES mask was available; otherwise they use REGENIE.

#### AD primary/secondary definitions with ADRD sensitivity − ADAM10 − LOF+REVEL50 (EUR)

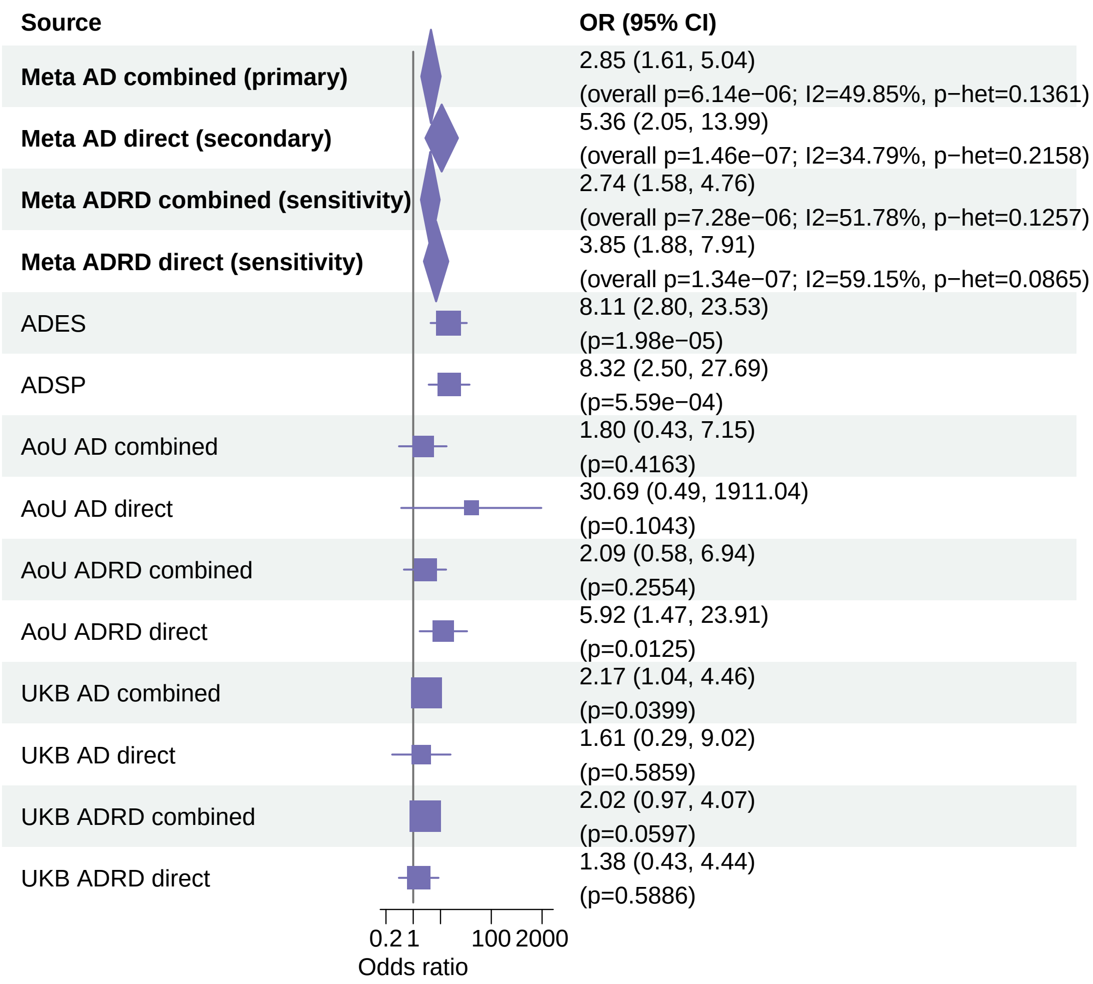


Meta OR/CI values are pooled REGENIE effects. Meta overall P values use ADES+REGENIE signed−Z evidence when the exact ADES mask was available; otherwise they use REGENIE.

#### AD primary/secondary definitions with ADRD sensitivity − TREM2 − LOF+REVEL75 (EUR)

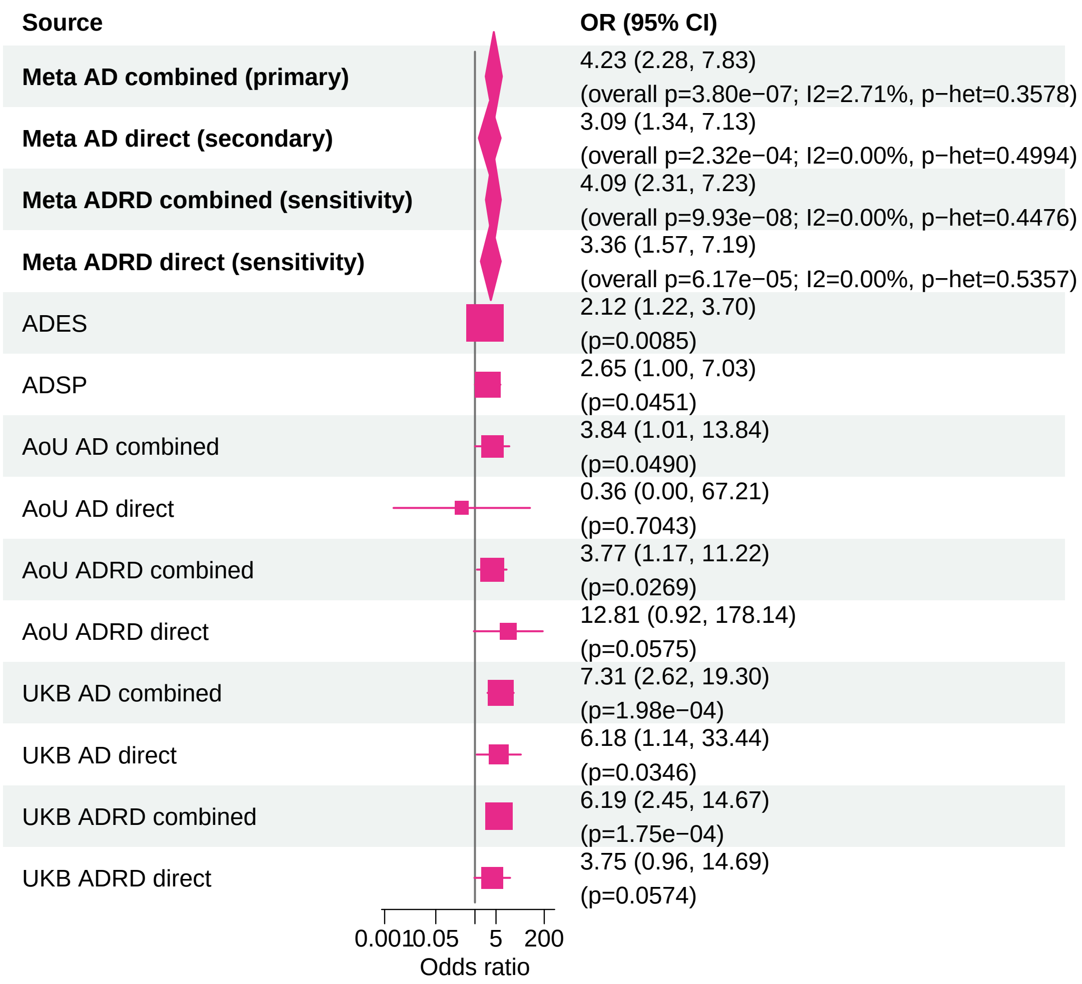


Meta OR/CI values are pooled REGENIE effects. Meta overall P values use ADES+REGENIE signed−Z evidence when the exact ADES mask was available; otherwise they use REGENIE.

#### AD primary/secondary definitions with ADRD sensitivity − TBK1 − LOF+REVEL50 (EUR)

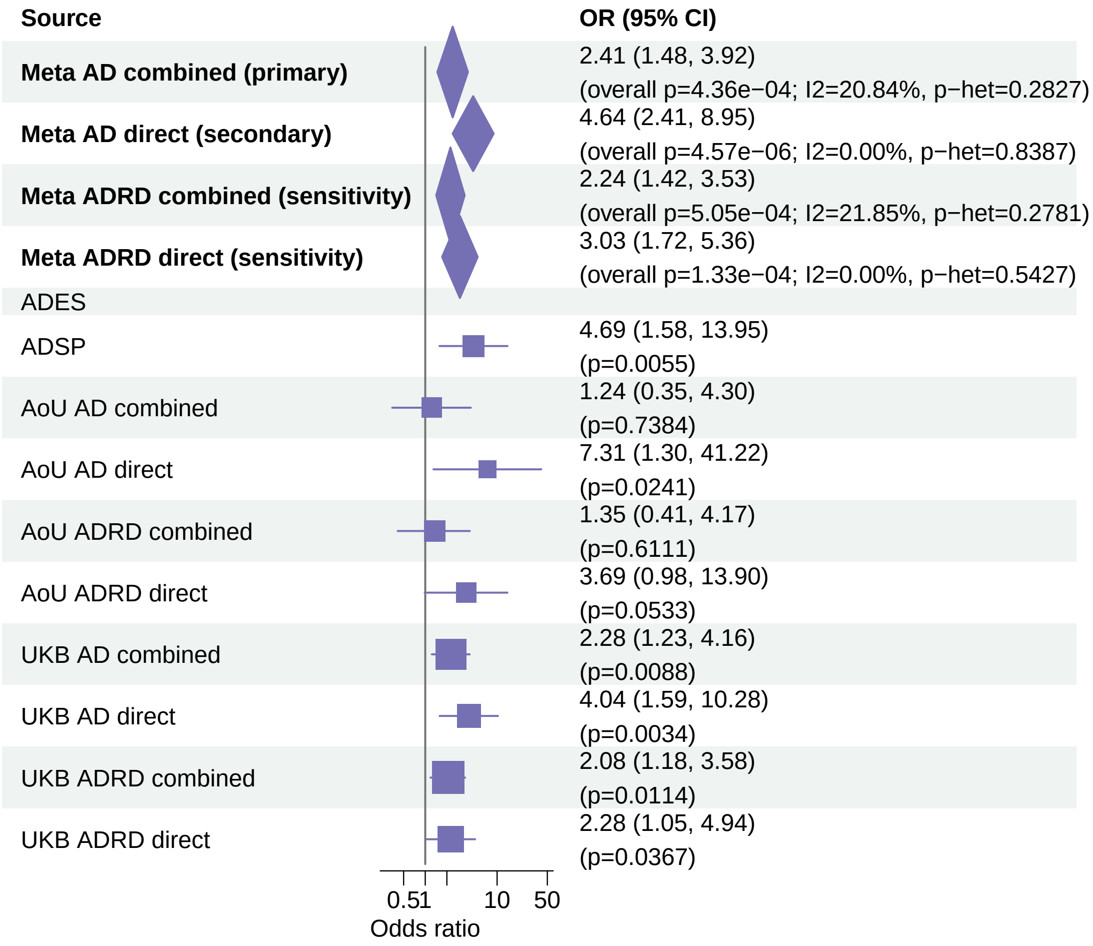


Meta OR/CI values are pooled REGENIE effects. Meta overall P values use ADES+REGENIE signed−Z evidence when the exact ADES mask was available; otherwise they use REGENIE.

#### AD primary/secondary definitions with ADRD sensitivity − ADAM10 − LOF+REVEL75 (EUR)

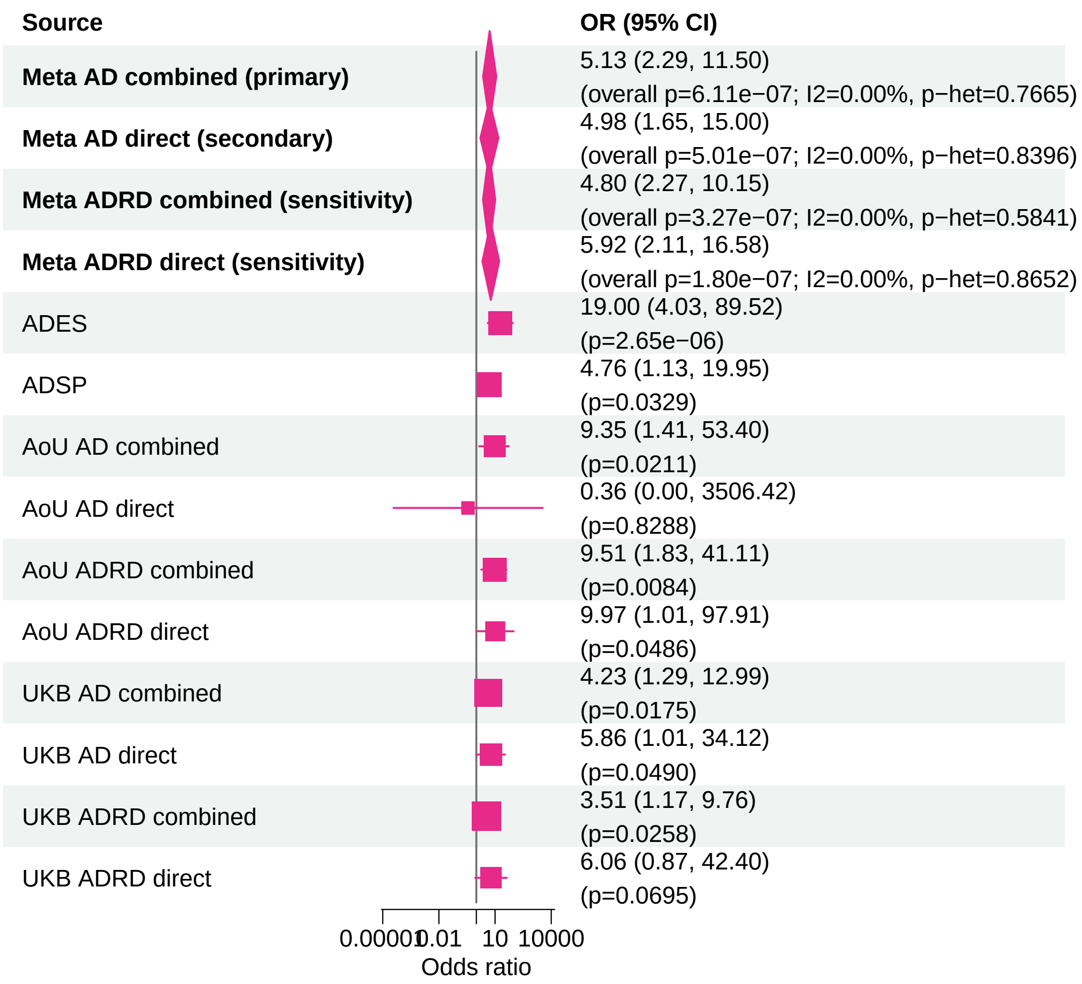


Meta OR/CI values are pooled REGENIE effects. Meta overall P values use ADES+REGENIE signed−Z evidence when the exact ADES mask was available; otherwise they use REGENIE.

#### AD primary/secondary definitions with ADRD sensitivity − IMPA2 − LOF+REVEL75 (EUR)

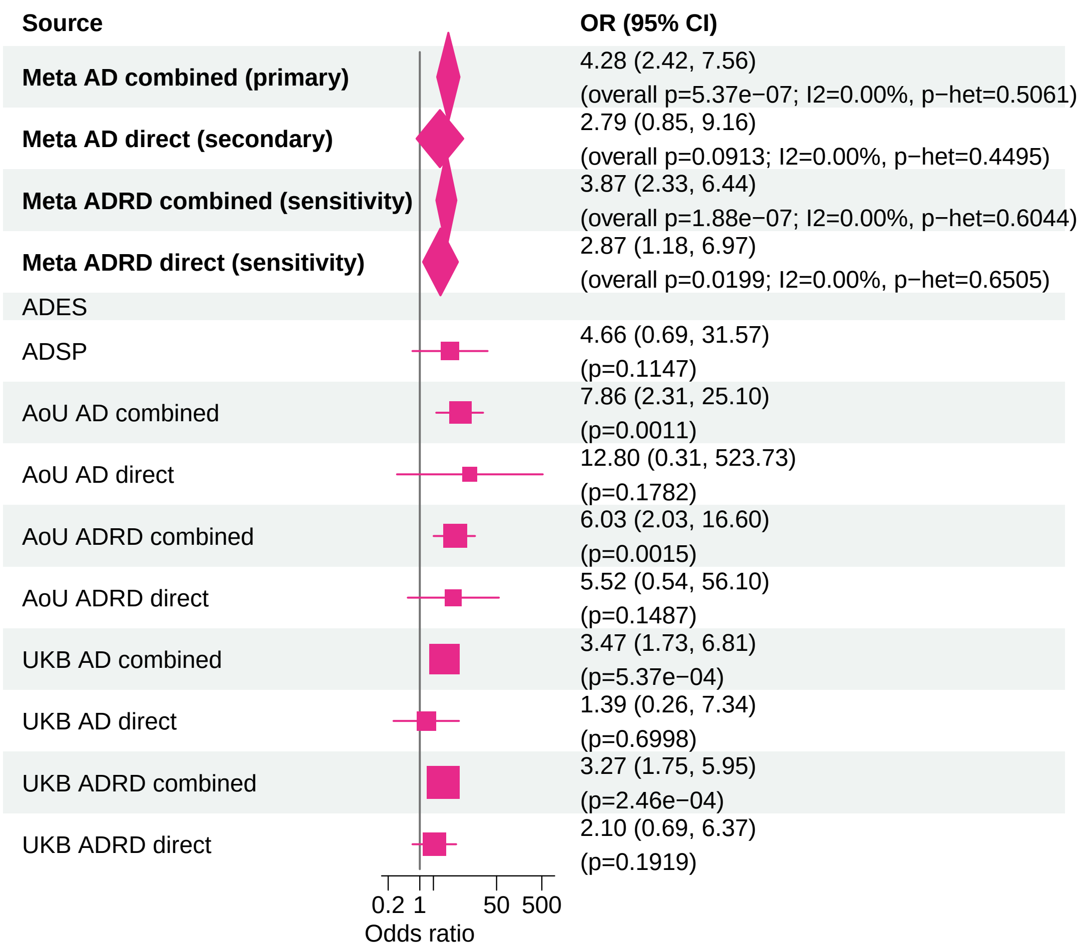


Meta OR/CI values are pooled REGENIE effects. Meta overall P values use ADES+REGENIE signed−Z evidence when the exact ADES mask was available; otherwise they use REGENIE.

**AD primary/secondary definitions with ADRD sensitivity − ADAM10 − LOF+REVEL25 (EUR)**

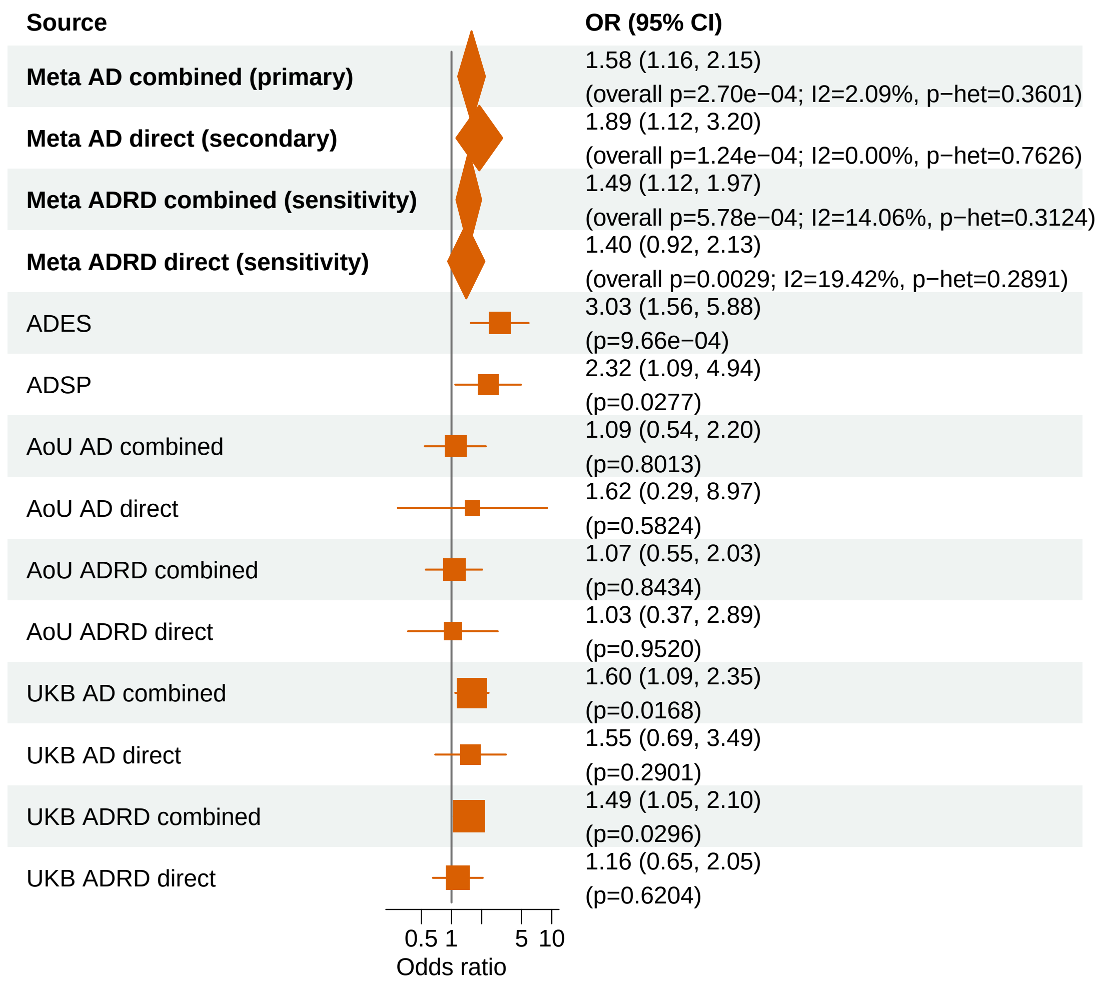


Meta OR/CI values are pooled REGENIE effects. Meta overall P values use ADES+REGENIE signed−Z evidence when the exact ADES mask was available; otherwise they use REGENIE.

### AD primary/secondary definitions with ADRD sensitivity − TBK1 − LOF (EUR)

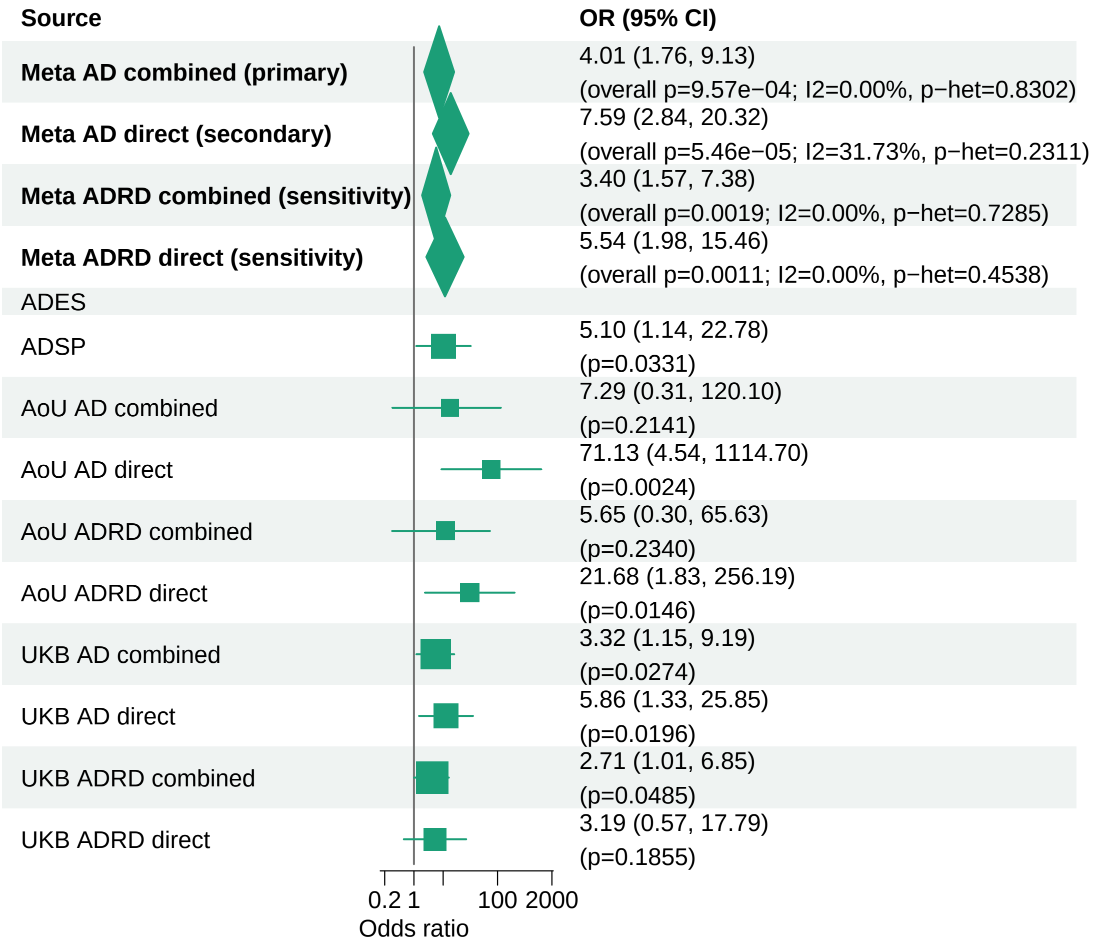


Meta OR/CI values are pooled REGENIE effects. Meta overall P values use ADES+REGENIE signed−Z evidence when the exact ADES mask was available; otherwise they use REGENIE.

#### AD primary/secondary definitions with ADRD sensitivity − TBK1 − LOF+REVEL75 (EUR)

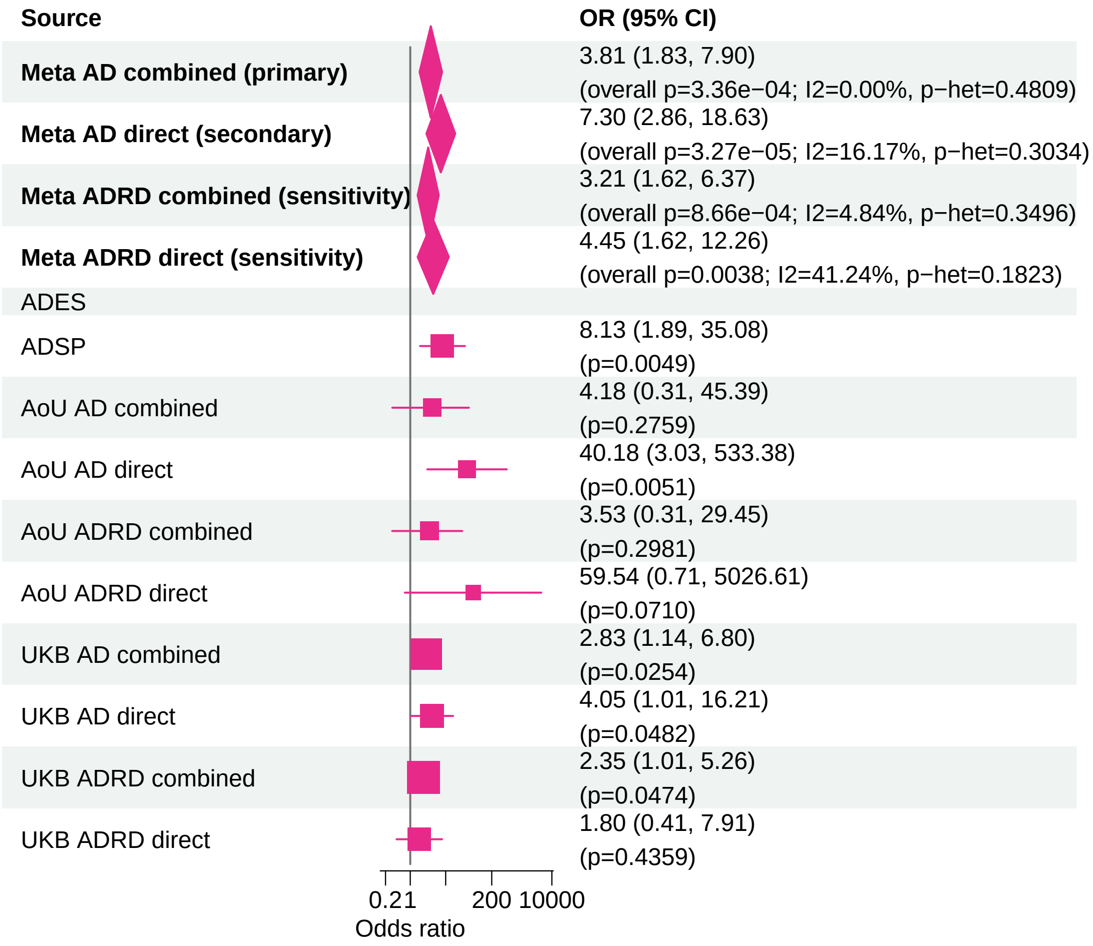


Meta OR/CI values are pooled REGENIE effects. Meta overall P values use ADES+REGENIE signed−Z evidence when the exact ADES mask was available; otherwise they use REGENIE.

### AD primary/secondary definitions with ADRD sensitivity − SHARPIN − LOF+REVEL25 (EUR)

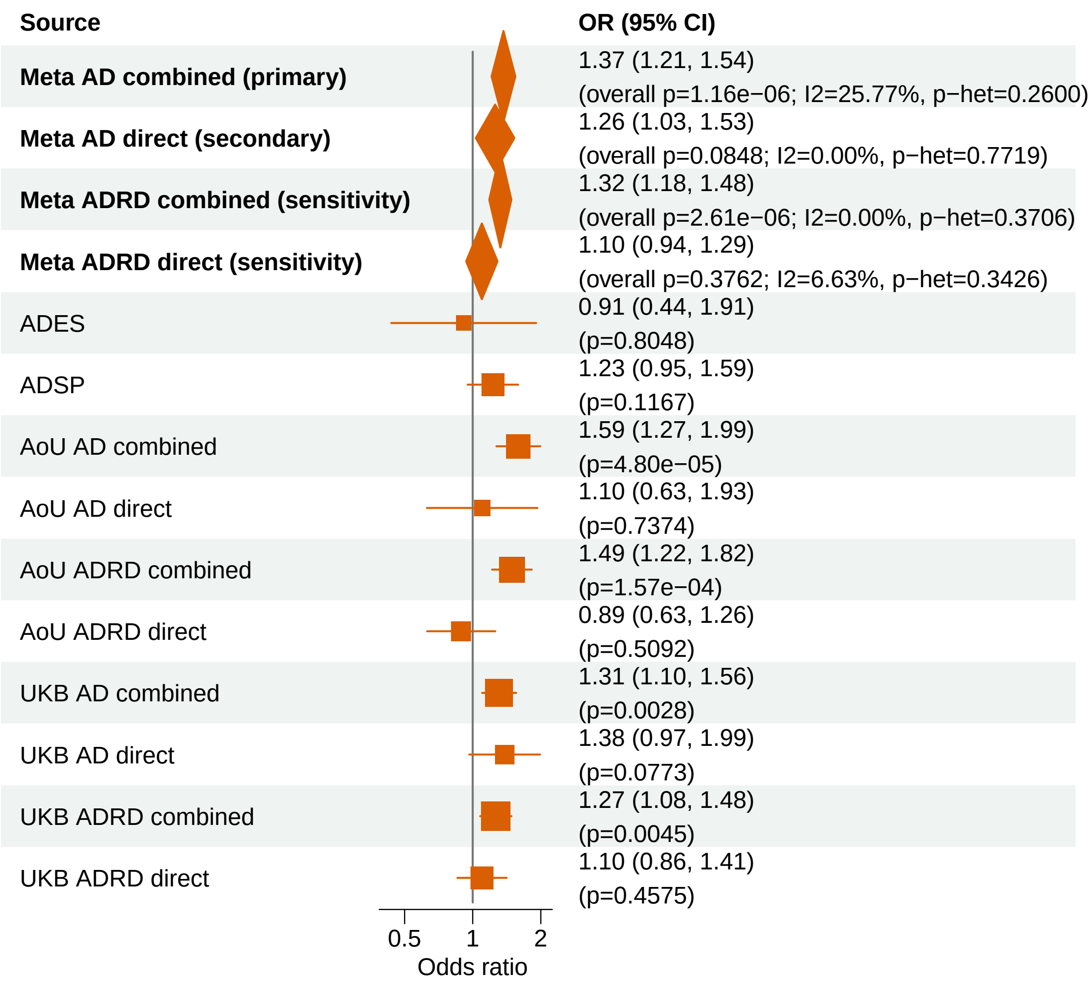


Meta OR/CI values are pooled REGENIE effects. Meta overall P values use ADES+REGENIE signed−Z evidence when the exact ADES mask was available; otherwise they use REGENIE.

#### AD primary/secondary definitions with ADRD sensitivity − SYNE1 − LOF+REVEL50 (EUR)

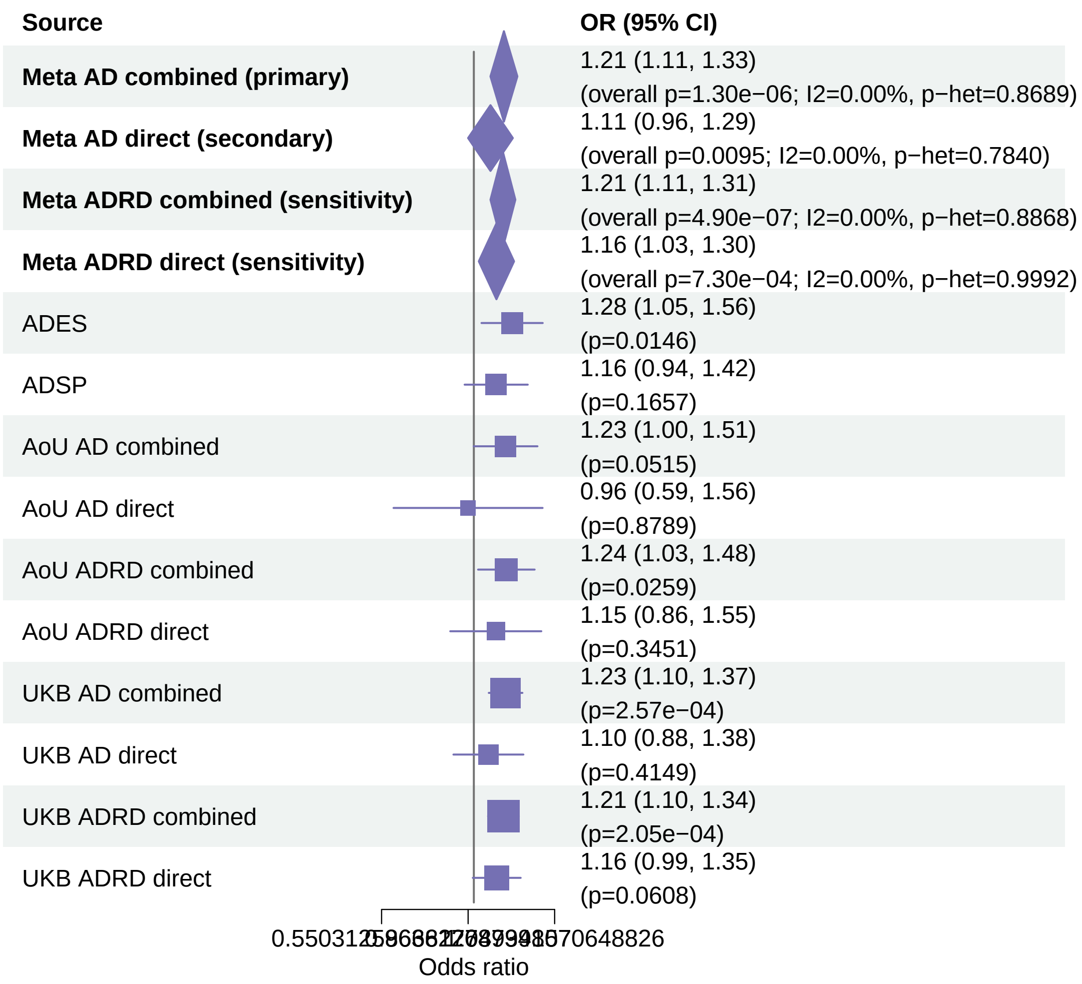


Meta OR/CI values are pooled REGENIE effects. Meta overall P values use ADES+REGENIE signed−Z evidence when the exact ADES mask was available; otherwise they use REGENIE.

#### AD primary/secondary definitions with ADRD sensitivity − APOE − LOF+REVEL25 (EUR)

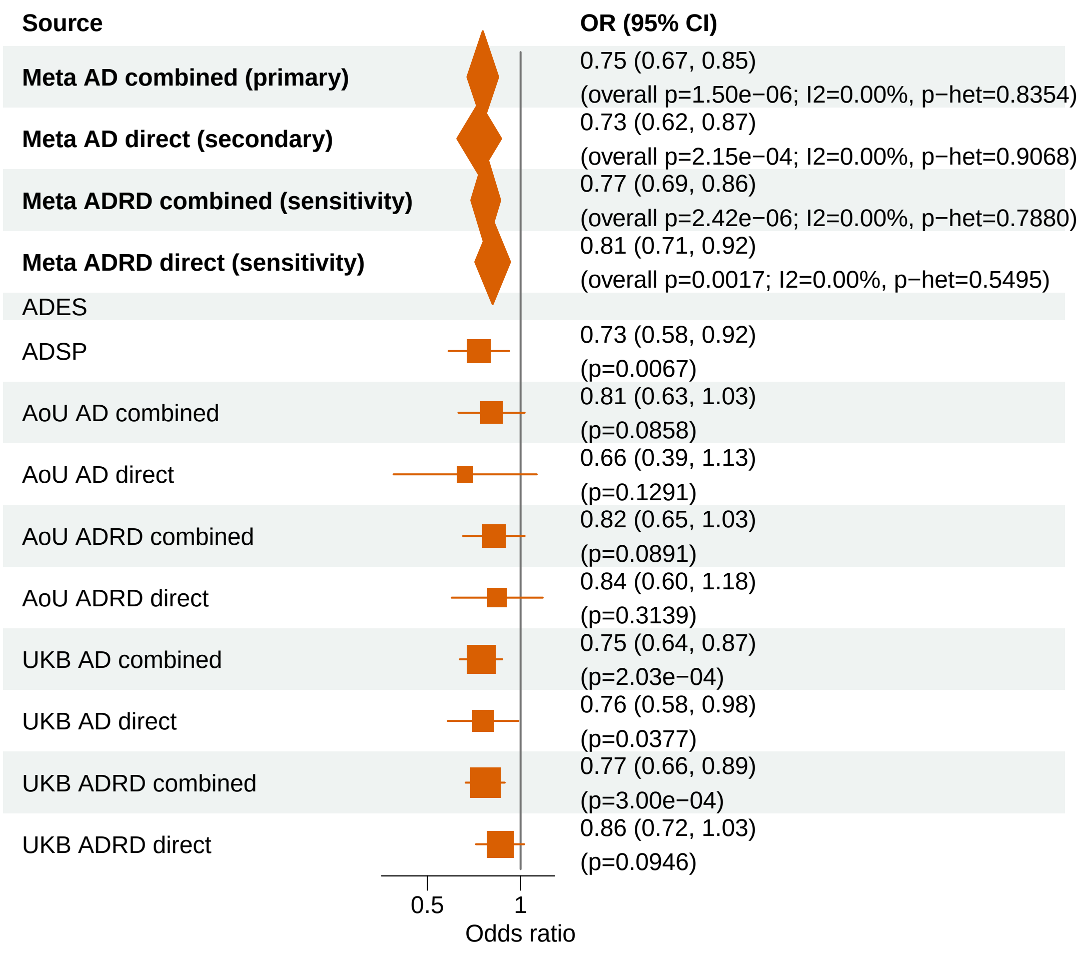


Meta OR/CI values are pooled REGENIE effects. Meta overall P values use ADES+REGENIE signed−Z evidence when the exact ADES mask was available; otherwise they use REGENIE.

#### AD primary/secondary definitions with ADRD sensitivity − GRN − LOF+REVEL50 (EUR)

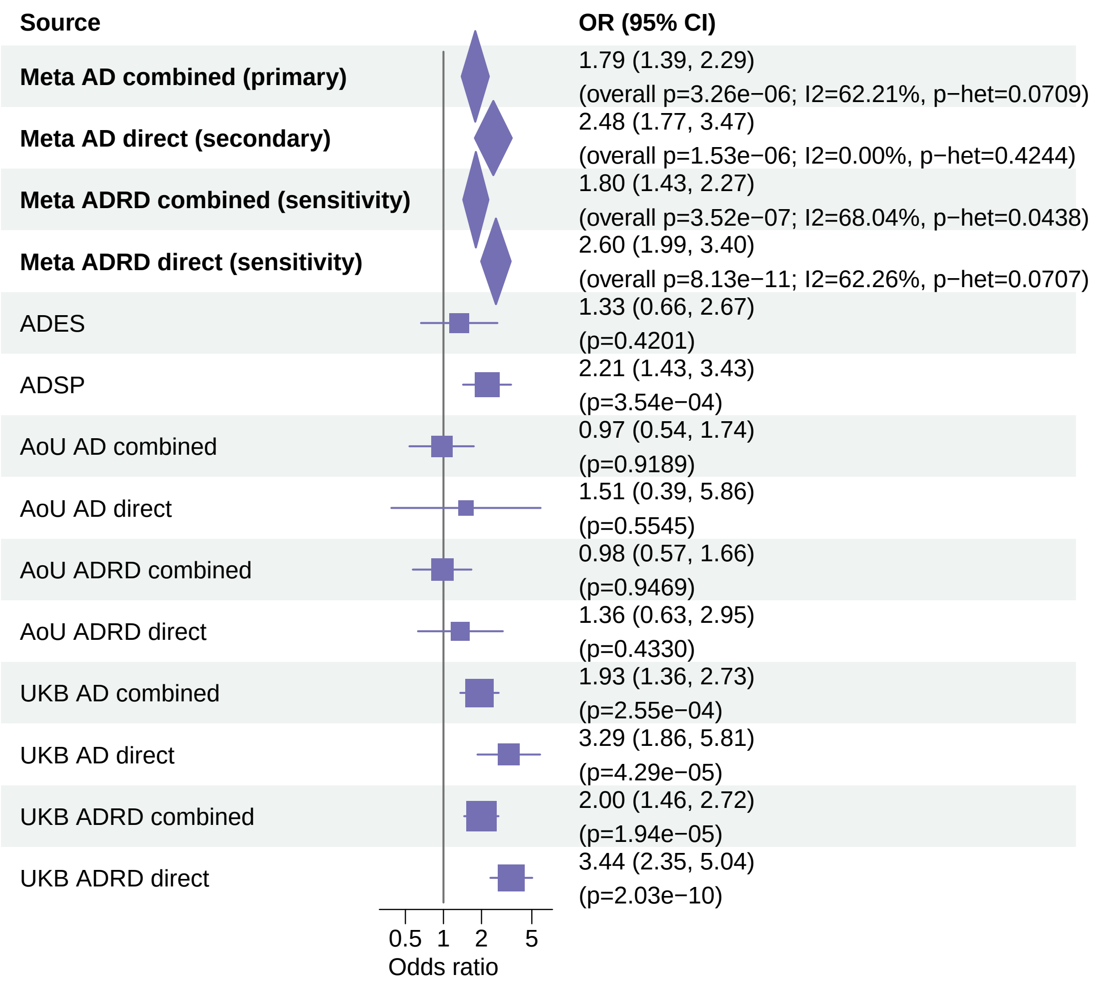


Meta OR/CI values are pooled REGENIE effects. Meta overall P values use ADES+REGENIE signed−Z evidence when the exact ADES mask was available; otherwise they use REGENIE.

#### AD primary/secondary definitions with ADRD sensitivity − RIN3 − LOF+REVEL50 (EUR)

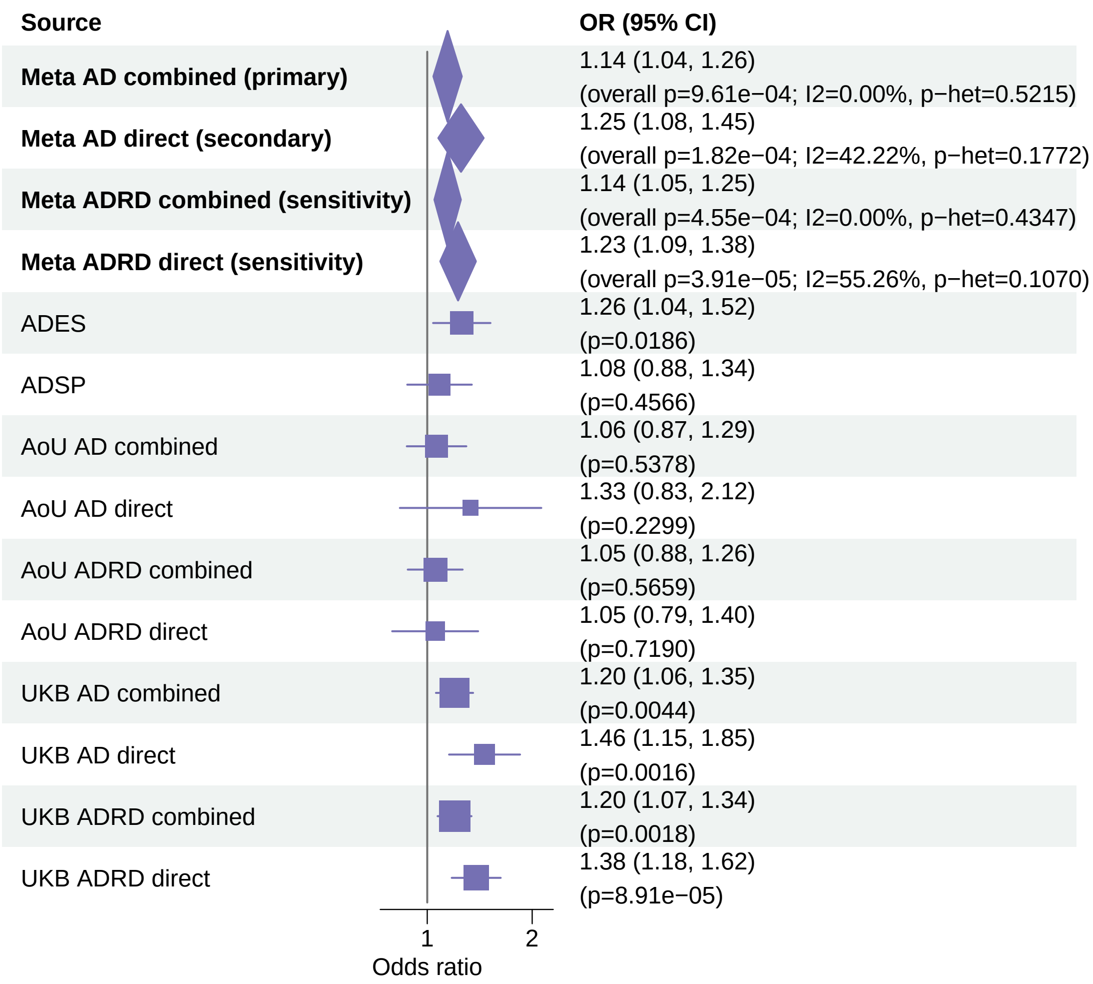


Meta OR/CI values are pooled REGENIE effects. Meta overall P values use ADES+REGENIE signed−Z evidence when the exact ADES mask was available; otherwise they use REGENIE.

#### AD primary/secondary definitions with ADRD sensitivity − CHRNA4 − LOF+REVEL50 (EUR)

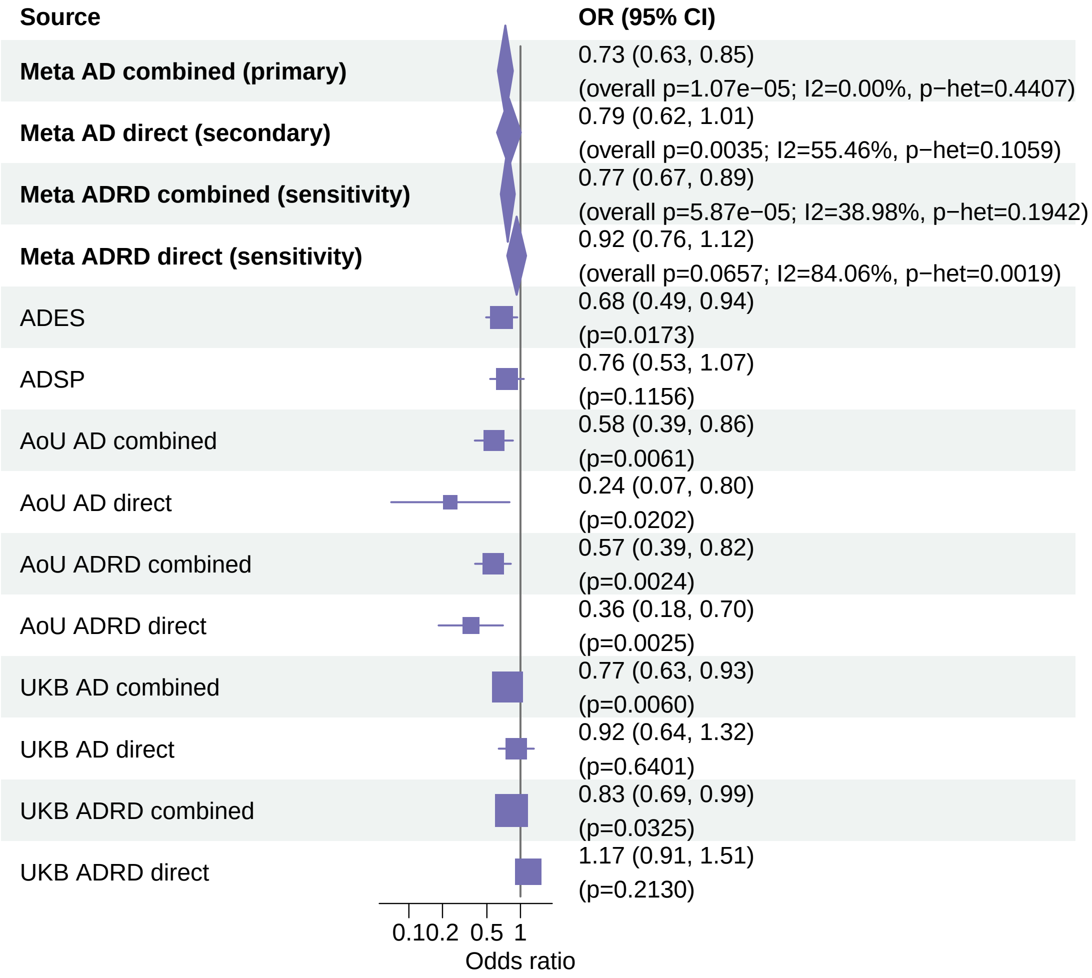


Meta OR/CI values are pooled REGENIE effects. Meta overall P values use ADES+REGENIE signed−Z evidence when the exact ADES mask was available; otherwise they use REGENIE.

#### AD primary/secondary definitions with ADRD sensitivity − CLU − LOF+REVEL50 (EUR)

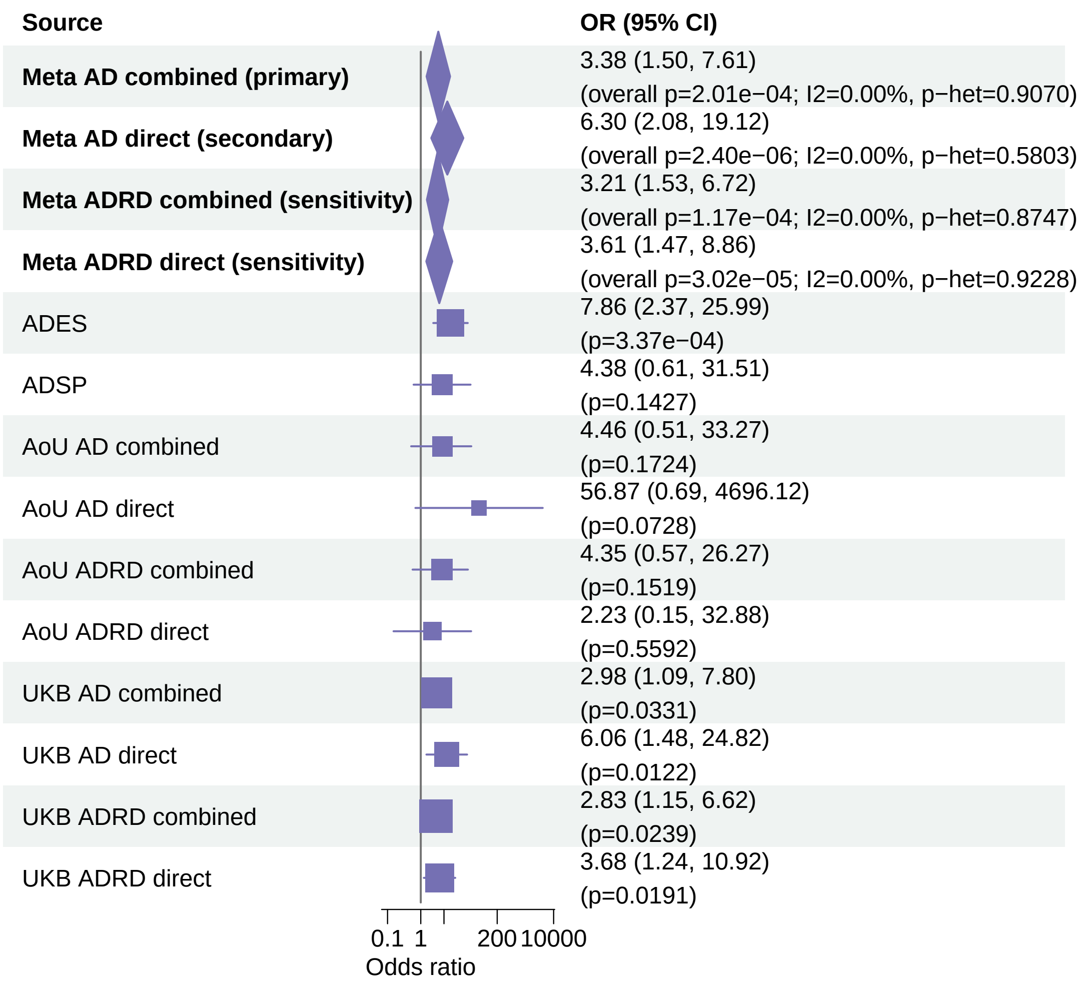


Meta OR/CI values are pooled REGENIE effects. Meta overall P values use ADES+REGENIE signed−Z evidence when the exact ADES mask was available; otherwise they use REGENIE.

#### AD primary/secondary definitions with ADRD sensitivity − ADAM10 − LOF (EUR)

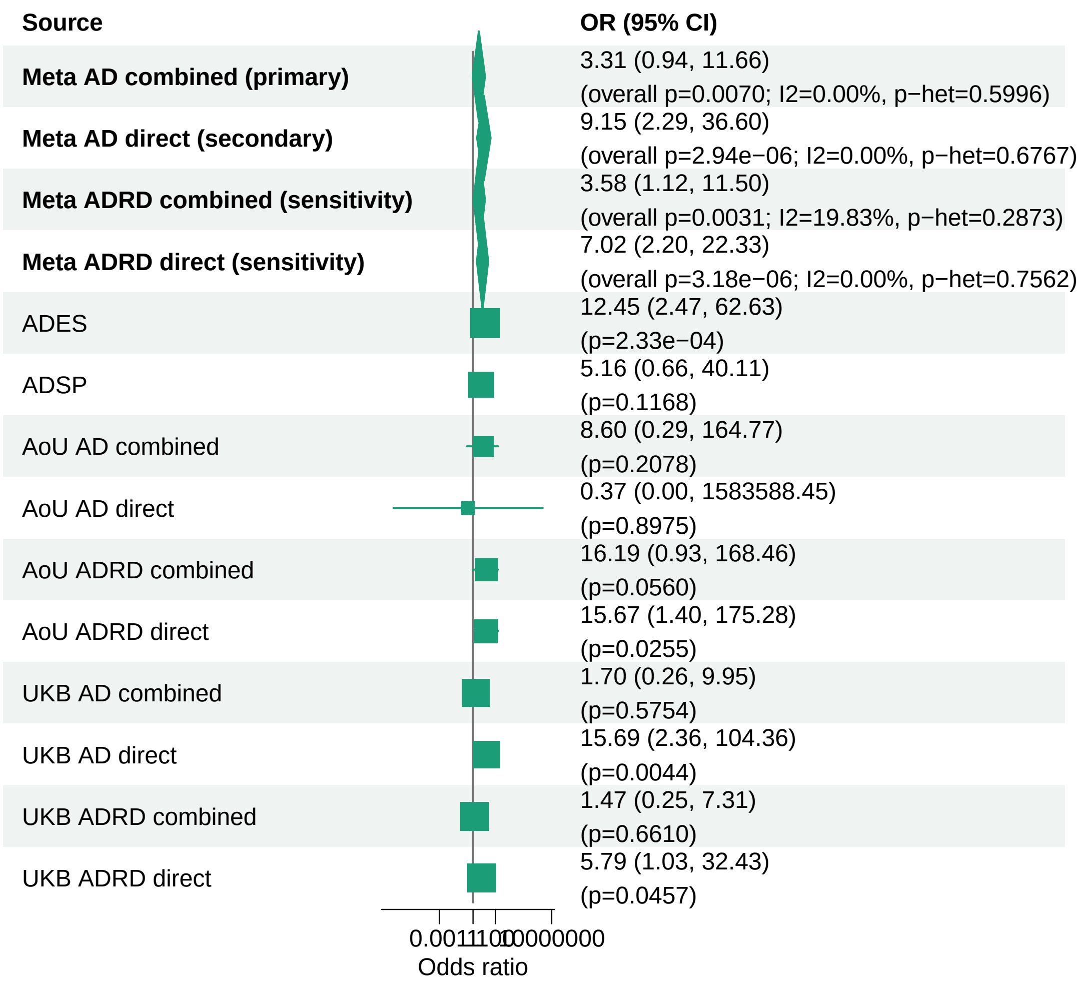


Meta OR/CI values are pooled REGENIE effects. Meta overall P values use ADES+REGENIE signed−Z evidence when the exact ADES mask was available; otherwise they use REGENIE.

#### AD primary/secondary definitions with ADRD sensitivity − RIN3 − LOF+REVEL25 (EUR)

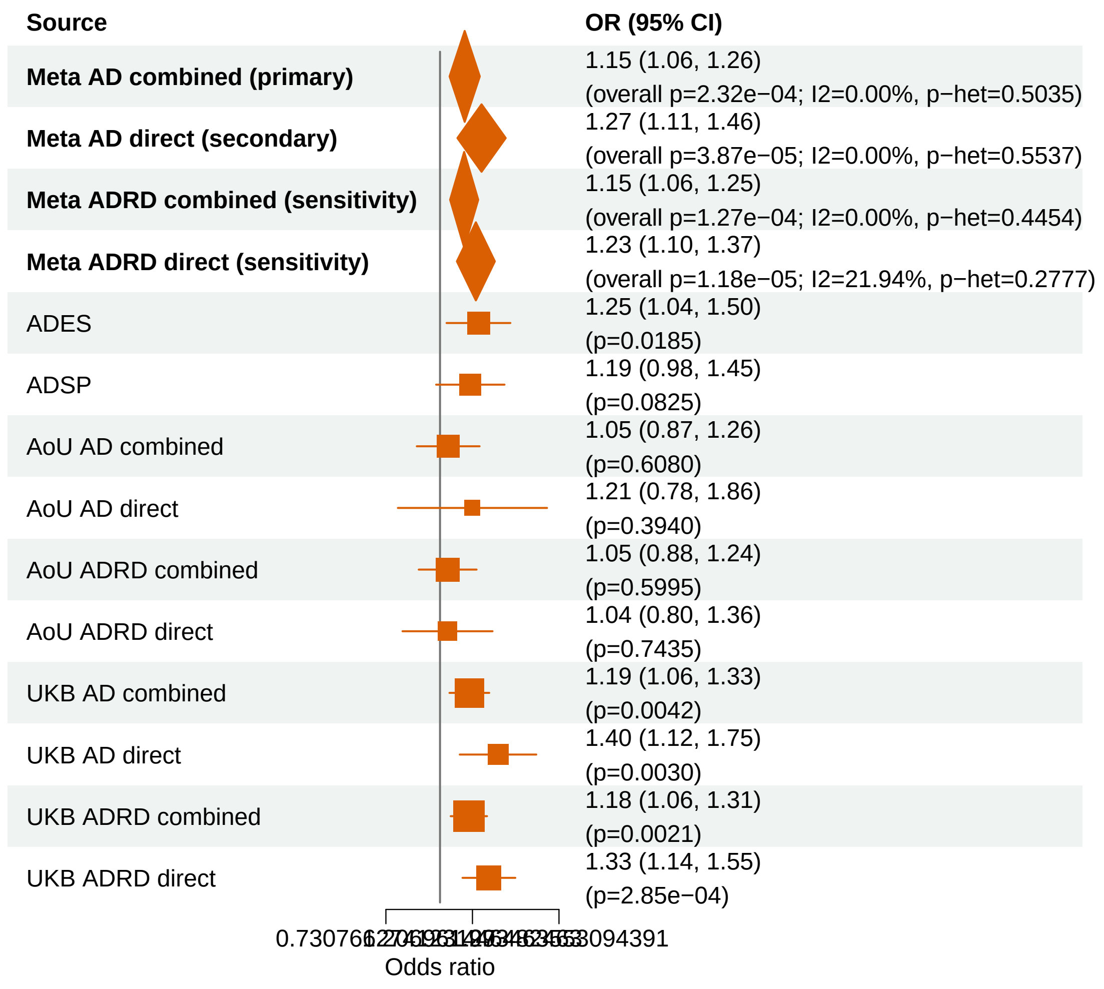


Meta OR/CI values are pooled REGENIE effects. Meta overall P values use ADES+REGENIE signed−Z evidence when the exact ADES mask was available; otherwise they use REGENIE.

#### AD primary/secondary definitions with ADRD sensitivity − SP140L − LOF+REVEL75 (EUR)

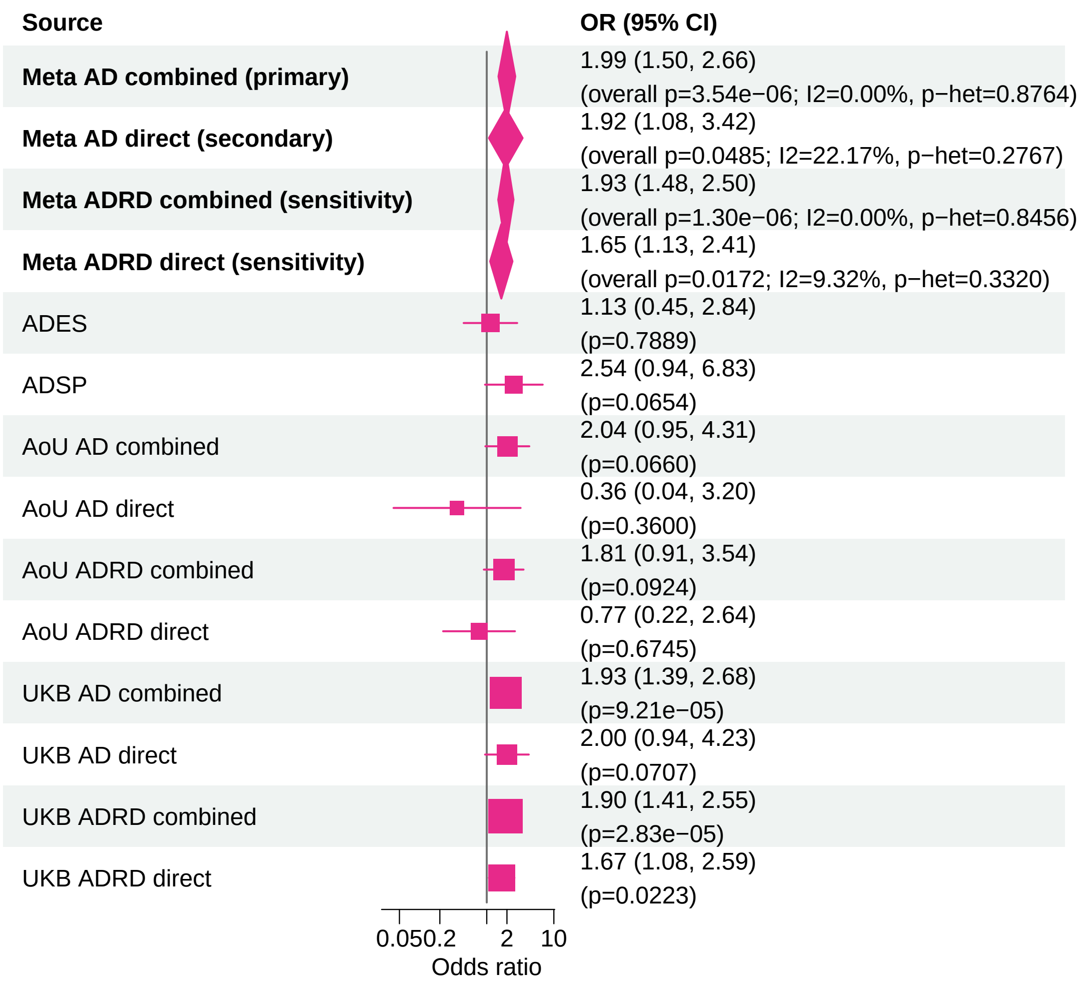


Meta OR/CI values are pooled REGENIE effects. Meta overall P values use ADES+REGENIE signed−Z evidence when the exact ADES mask was available; otherwise they use REGENIE.

#### AD primary/secondary definitions with ADRD sensitivity − PMM2 − LOF+REVEL25 (EUR)

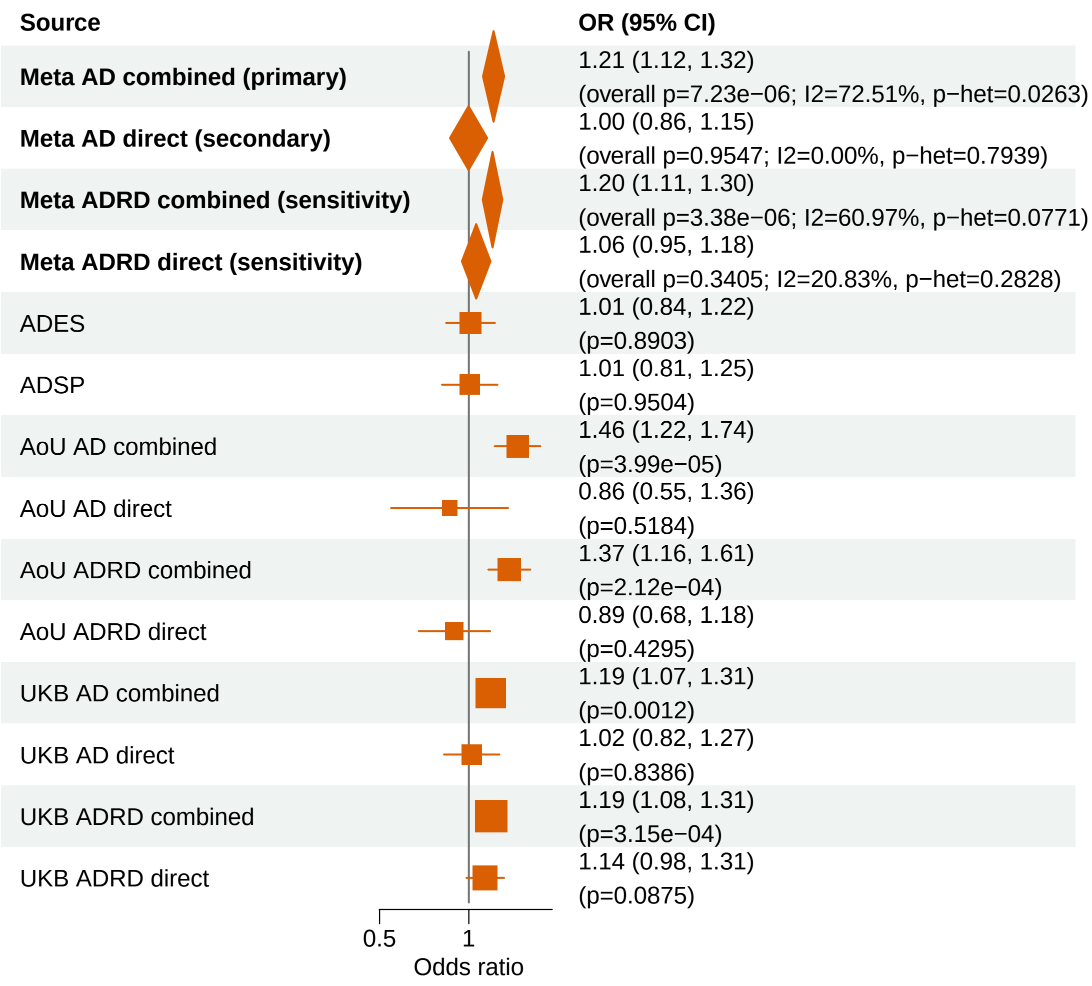


Meta OR/CI values are pooled REGENIE effects. Meta overall P values use ADES+REGENIE signed−Z evidence when the exact ADES mask was available; otherwise they use REGENIE.

#### AD primary/secondary definitions with ADRD sensitivity − TBK1 − LOF+REVEL25 (EUR)

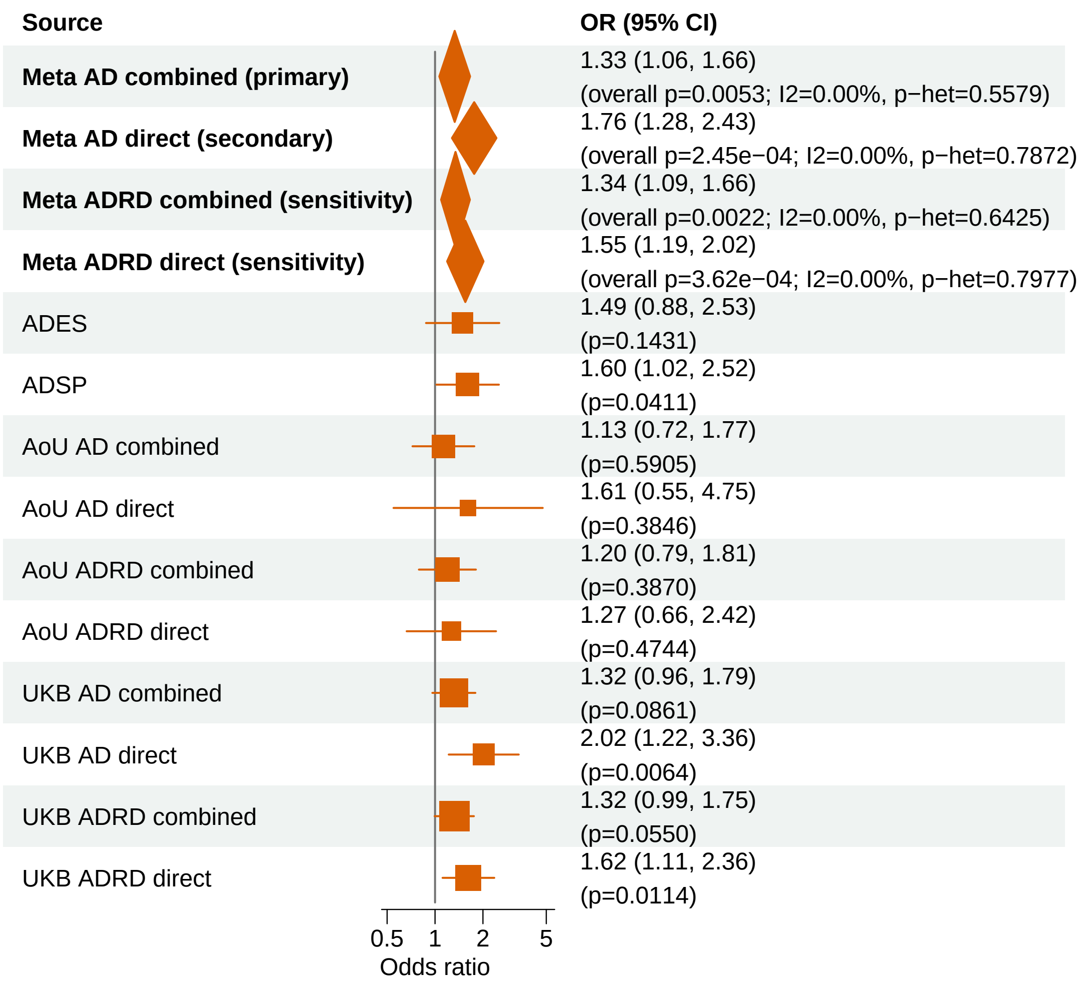


Meta OR/CI values are pooled REGENIE effects. Meta overall P values use ADES+REGENIE signed−Z evidence when the exact ADES mask was available; otherwise they use REGENIE.

#### AD primary/secondary definitions with ADRD sensitivity − ABCA1 − LOF+REVEL50 (EUR)

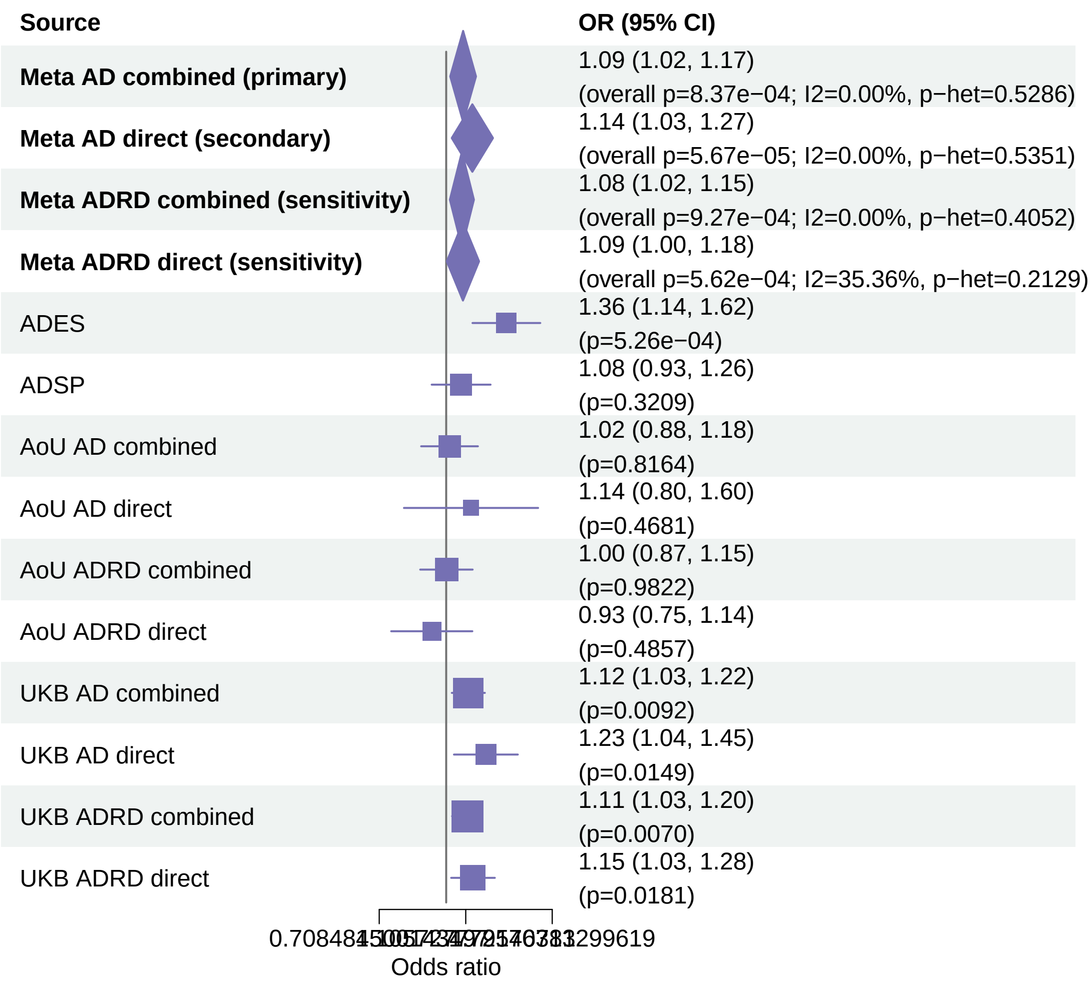


Meta OR/CI values are pooled REGENIE effects. Meta overall P values use ADES+REGENIE signed−Z evidence when the exact ADES mask was available; otherwise they use REGENIE.

#### AD primary/secondary definitions with ADRD sensitivity − RBM12 − LOF (EUR)

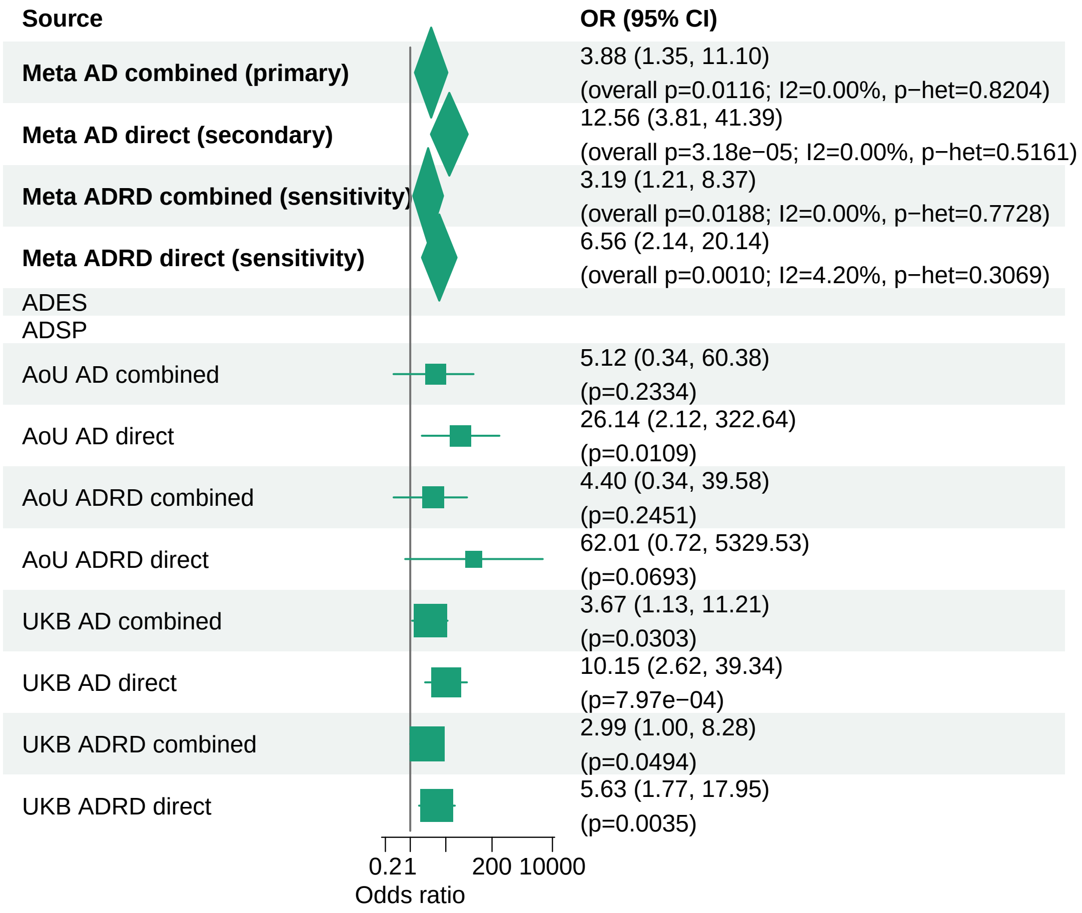


Meta OR/CI values are pooled REGENIE effects. Meta overall P values use ADES+REGENIE signed−Z evidence when the exact ADES mask was available; otherwise they use REGENIE.

#### AD primary/secondary definitions with ADRD sensitivity − ZNF256 − LOF+REVEL25 (EUR)

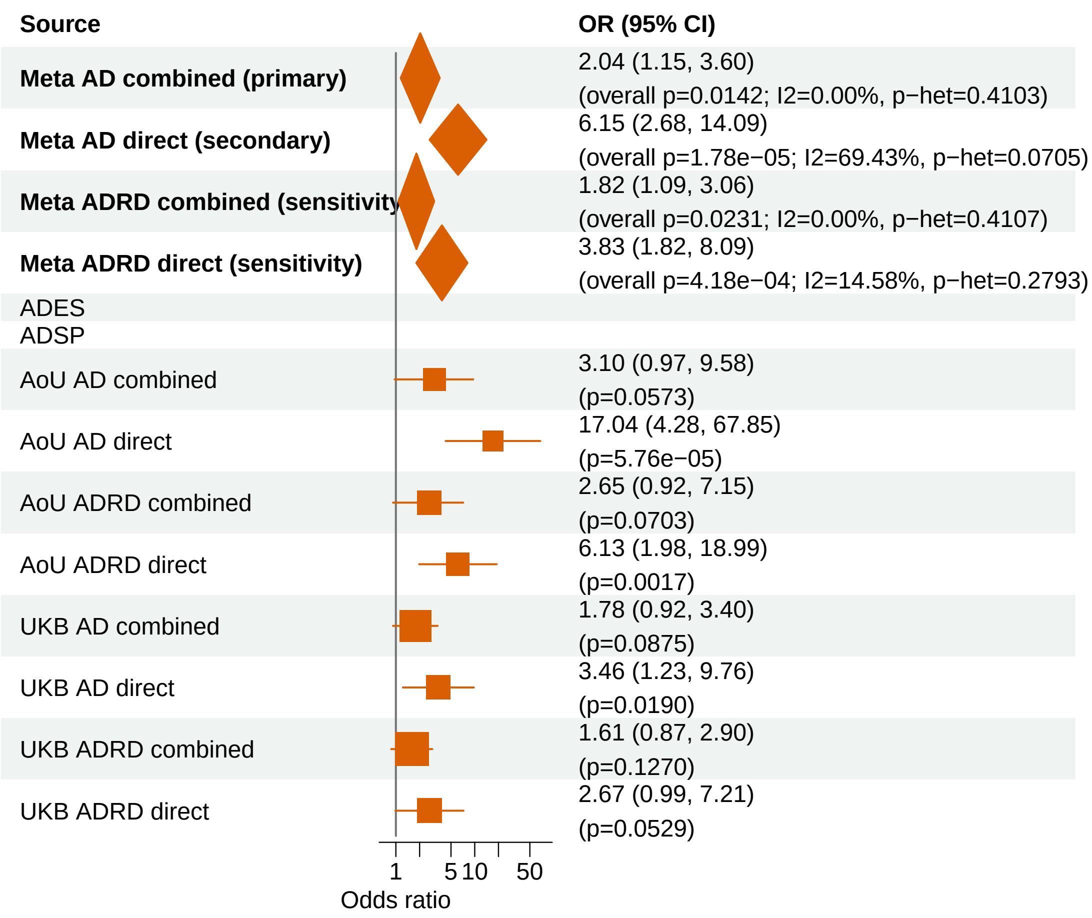


Meta OR/CI values are pooled REGENIE effects. Meta overall P values use ADES+REGENIE signed−Z evidence when the exact ADES mask was available; otherwise they use REGENIE.

### AD primary/secondary definitions with ADRD sensitivity − RBM12 − LOF+REVEL75 (EUR)

| Source | OR (95% CI) |
|---|---|
| **Meta AD combined (primary)** | 3.69 (1.30, 10.53) (overall p=0.0144; I2=0.00%, p−het=0.8578) |
| **Meta AD direct (secondary)** | 12.50 (3.80, 41.15) (overall p=3.25e−05; I2=0.00%, p−het=0.5204) |
| **Meta ADRD combined (sensitivity)** | 3.46 (1.21, 9.89) (overall p=0.0205; I2=0.00%, p−het=0.8844) |
| **Meta ADRD direct (sensitivity)** | 6.50 (2.12, 19.93) (overall p=0.0011; I2=0.24%, p−het=0.3167) |
| ADES | |
| ADSP | |
| AoU AD combined | 4.59 (0.31, 52.94) (p=0.2597) |
| AoU AD direct | 25.78 (2.10, 316.57) (p=0.0111) |
| AoU ADRD combined | 4.05 (0.32, 35.98) (p=0.2671) |
| AoU ADRD direct | 57.85 (0.69, 4823.22) (p=0.0722) |
| UKB AD combined | 3.54 (1.10, 10.77) (p=0.0346) |
| UKB AD direct | 10.12 (2.61, 39.20) (p=8.04e−04) |
| UKB ADRD combined | 3.33 (0.99, 10.23) (p=0.0526) |
| UKB ADRD direct | 5.60 (1.76, 17.82) (p=0.0036) |

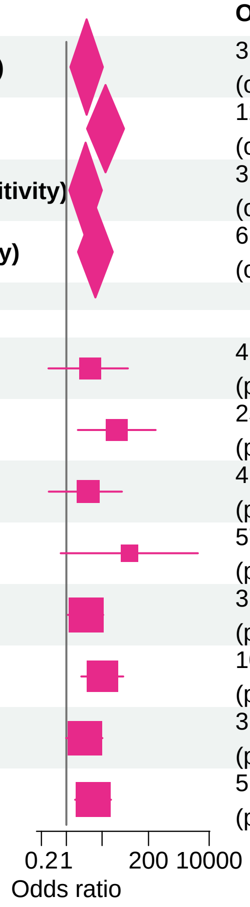


Meta OR/CI values are pooled REGENIE effects. Meta overall P values use ADES+REGENIE signed−Z evidence when the exact ADES mask was available; otherwise they use REGENIE.

#### AD primary/secondary definitions with ADRD sensitivity − CCL7 − LOF (EUR)

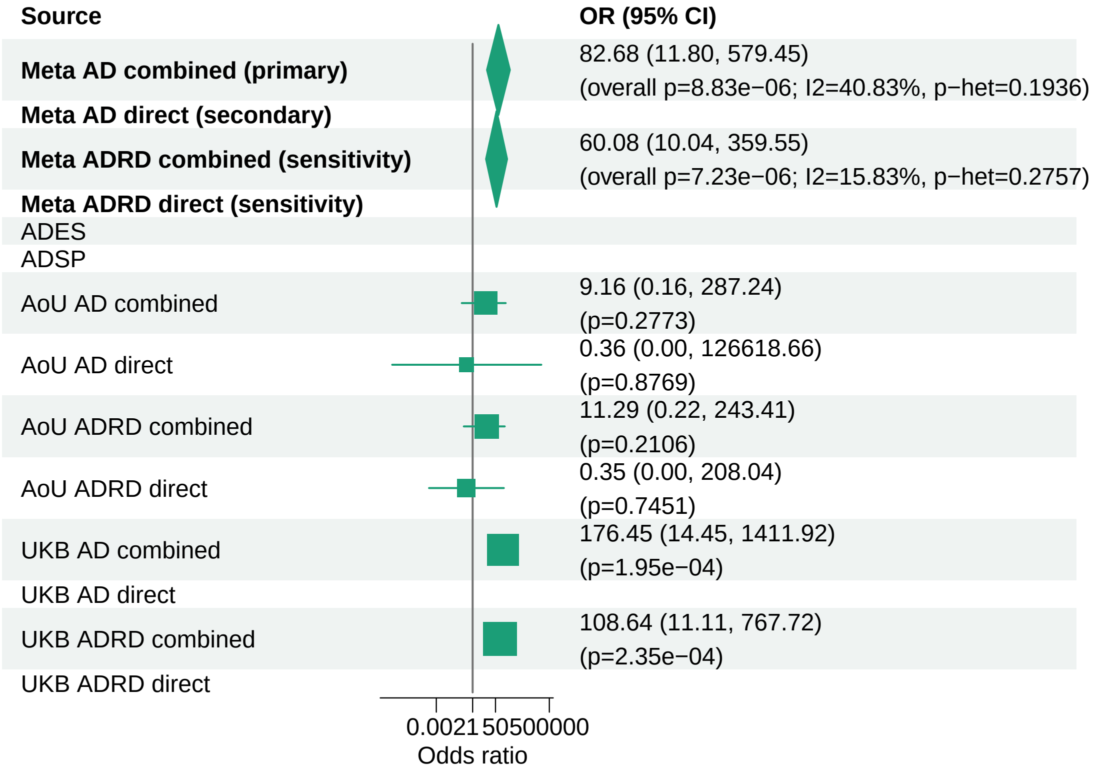


Meta OR/CI values are pooled REGENIE effects. Meta overall P values use ADES+REGENIE signed−Z evidence when the exact ADES mask was available; otherwise they use REGENIE.

#### AD primary/secondary definitions with ADRD sensitivity − MME − LOF+REVEL75 (EUR)

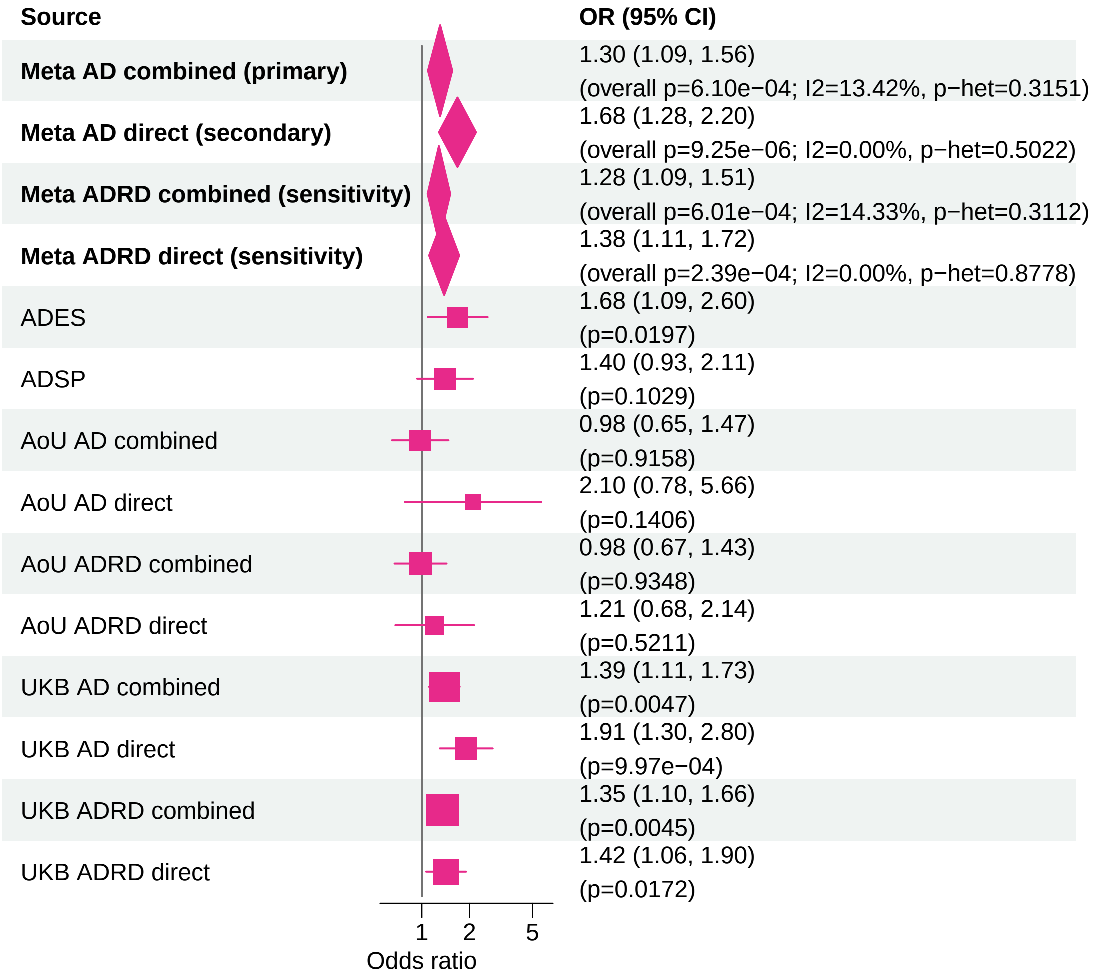


Meta OR/CI values are pooled REGENIE effects. Meta overall P values use ADES+REGENIE signed−Z evidence when the exact ADES mask was available; otherwise they use REGENIE.

### AD primary/secondary definitions with ADRD sensitivity − GRN − LOF+REVEL25 (EUR)

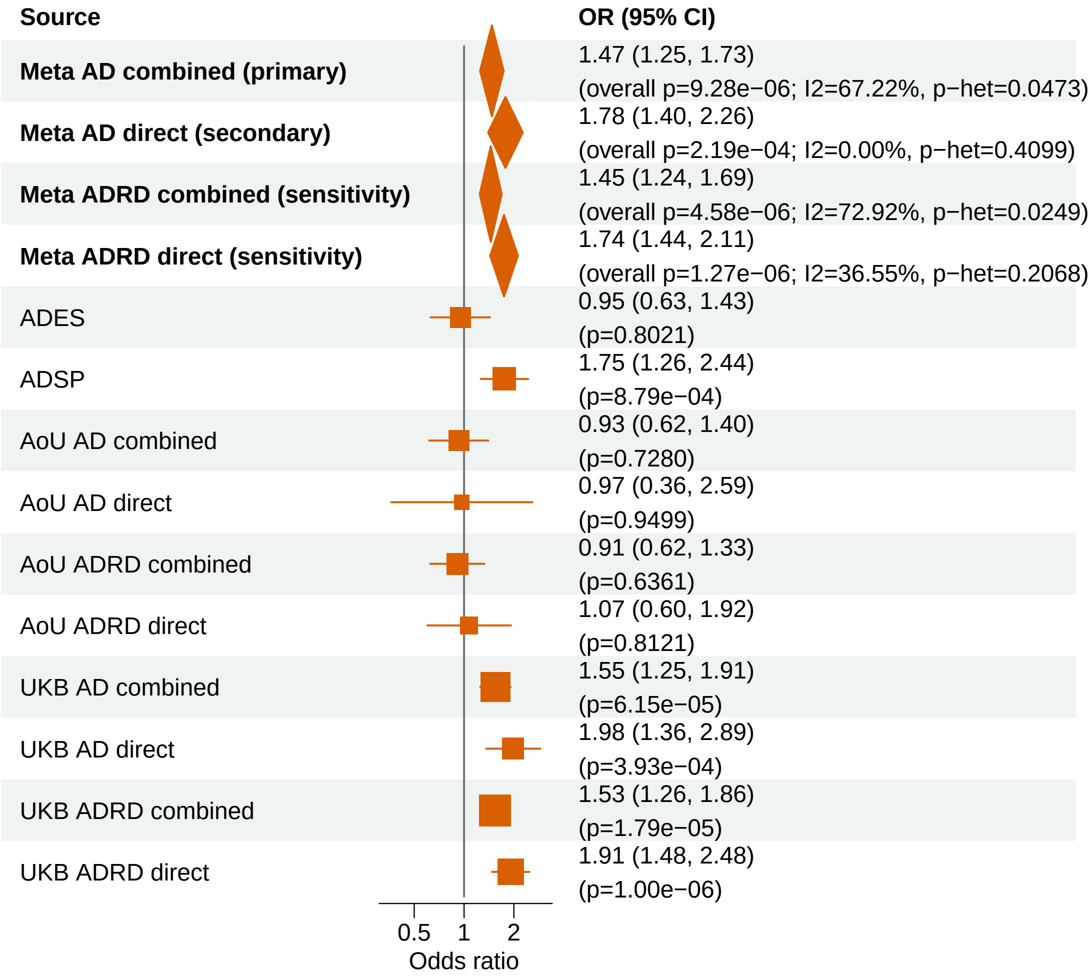


Meta OR/CI values are pooled REGENIE effects. Meta overall P values use ADES+REGENIE signed−Z evidence when the exact ADES mask was available; otherwise they use REGENIE.

#### AD primary/secondary definitions with ADRD sensitivity − ABCA1 − LOF+REVEL25 (EUR)

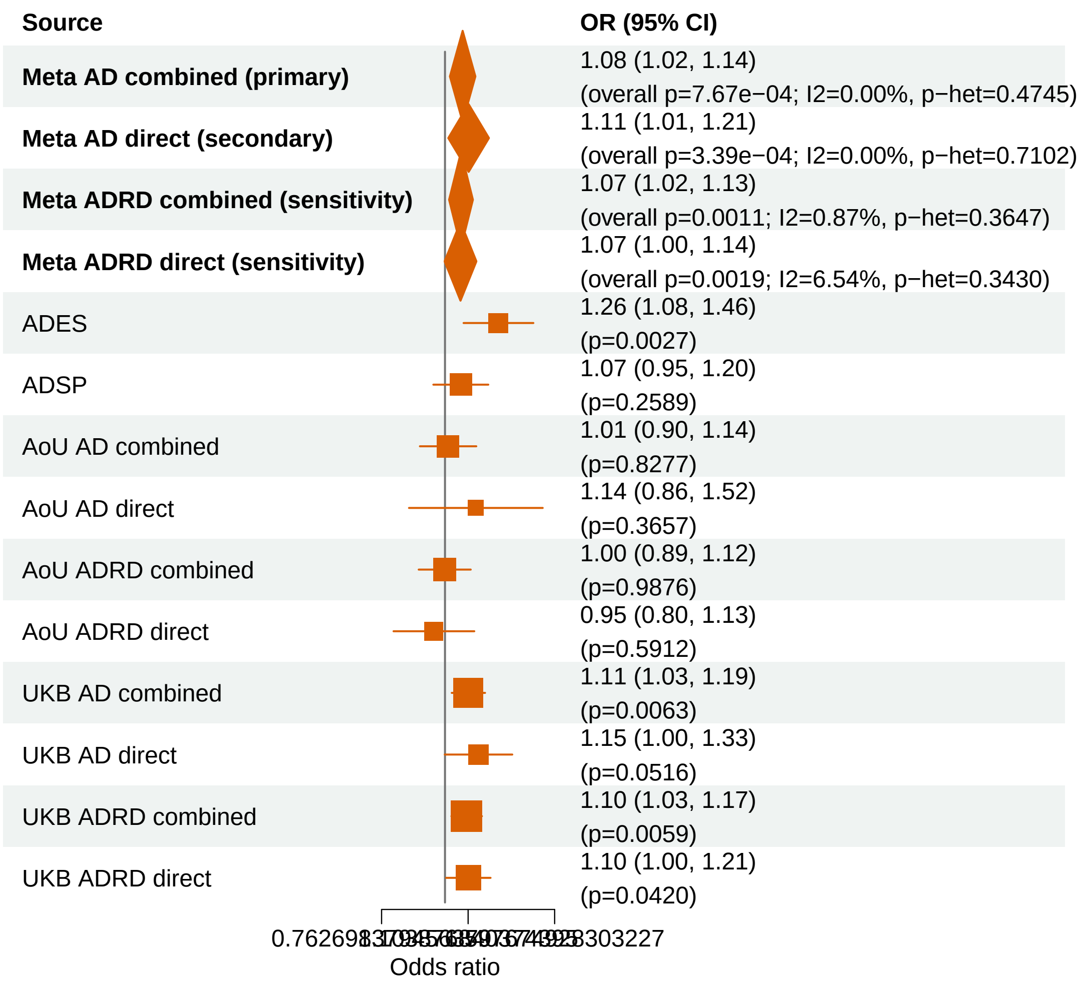


Meta OR/CI values are pooled REGENIE effects. Meta overall P values use ADES+REGENIE signed−Z evidence when the exact ADES mask was available; otherwise they use REGENIE.
