## Supplementary_Fig3 for "Population-scale burden analysis of rare damaging coding variants identifies novel risk genes for Alzheimer’s disease and related dementias and Parkinson’s disease and related disorders"

### AD primary/secondary definitions with ADRD sensitivity − PSEN1 − LOF+REVEL75 (ALL)

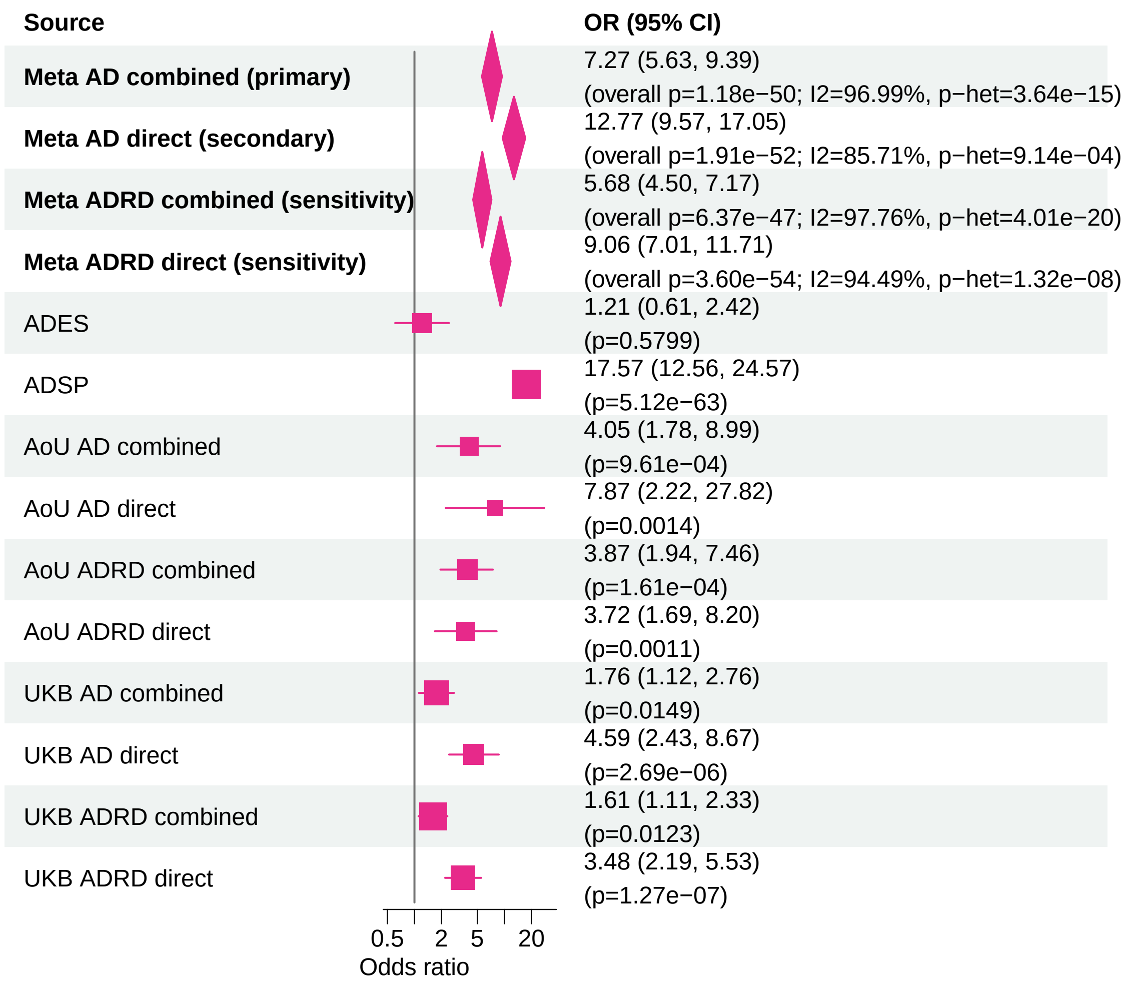


Meta OR/CI values are pooled REGENIE effects. Meta overall P values use ADES+REGENIE signed−Z evidence when the exact ADES mask was available; otherwise they use REGENIE.

#### AD primary/secondary definitions with ADRD sensitivity − TREM2 − LOF+REVEL25 (ALL)

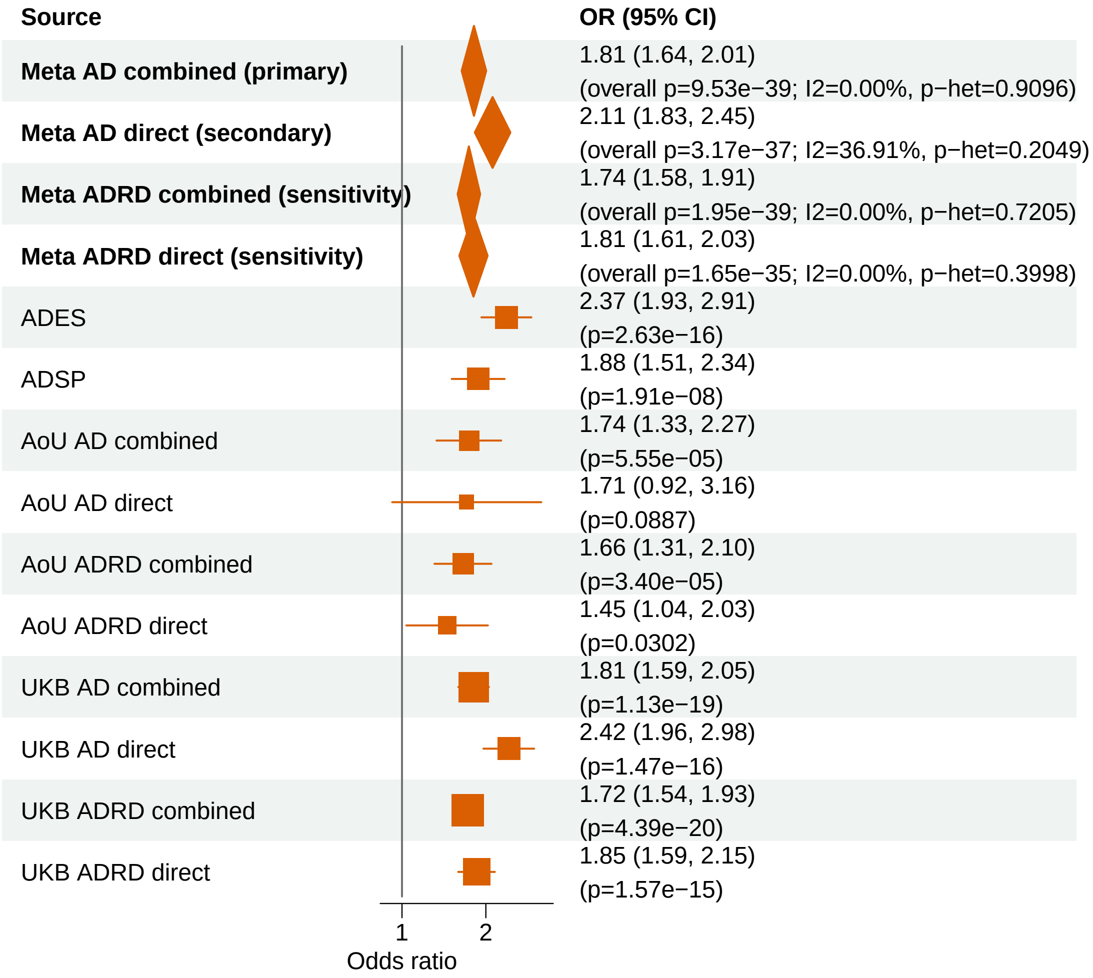


Meta OR/CI values are pooled REGENIE effects. Meta overall P values use ADES+REGENIE signed−Z evidence when the exact ADES mask was available; otherwise they use REGENIE.

#### AD primary/secondary definitions with ADRD sensitivity − ABCA7 − LOF+REVEL25 (ALL)

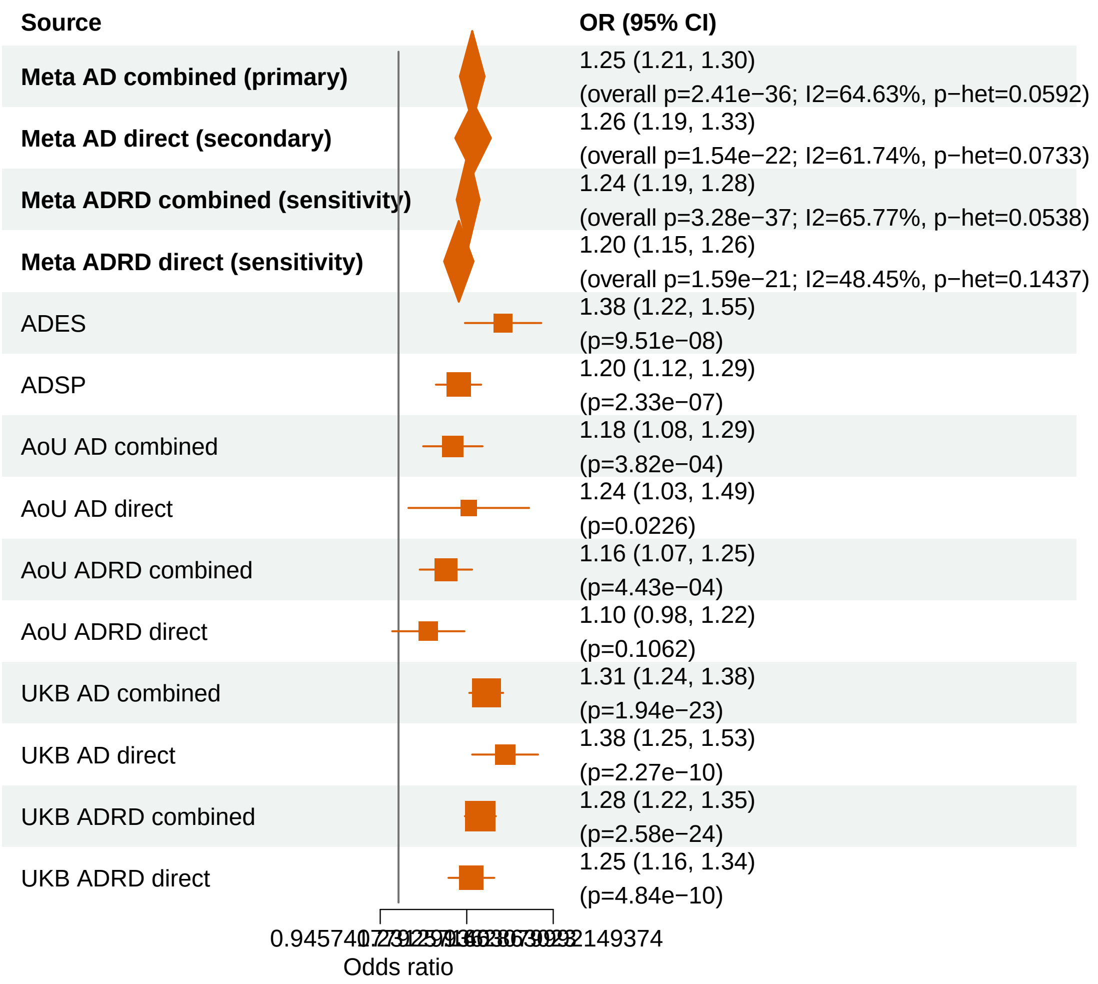


Meta OR/CI values are pooled REGENIE effects. Meta overall P values use ADES+REGENIE signed−Z evidence when the exact ADES mask was available; otherwise they use REGENIE.

#### AD primary/secondary definitions with ADRD sensitivity − PSEN1 − LOF+REVEL50 (ALL)

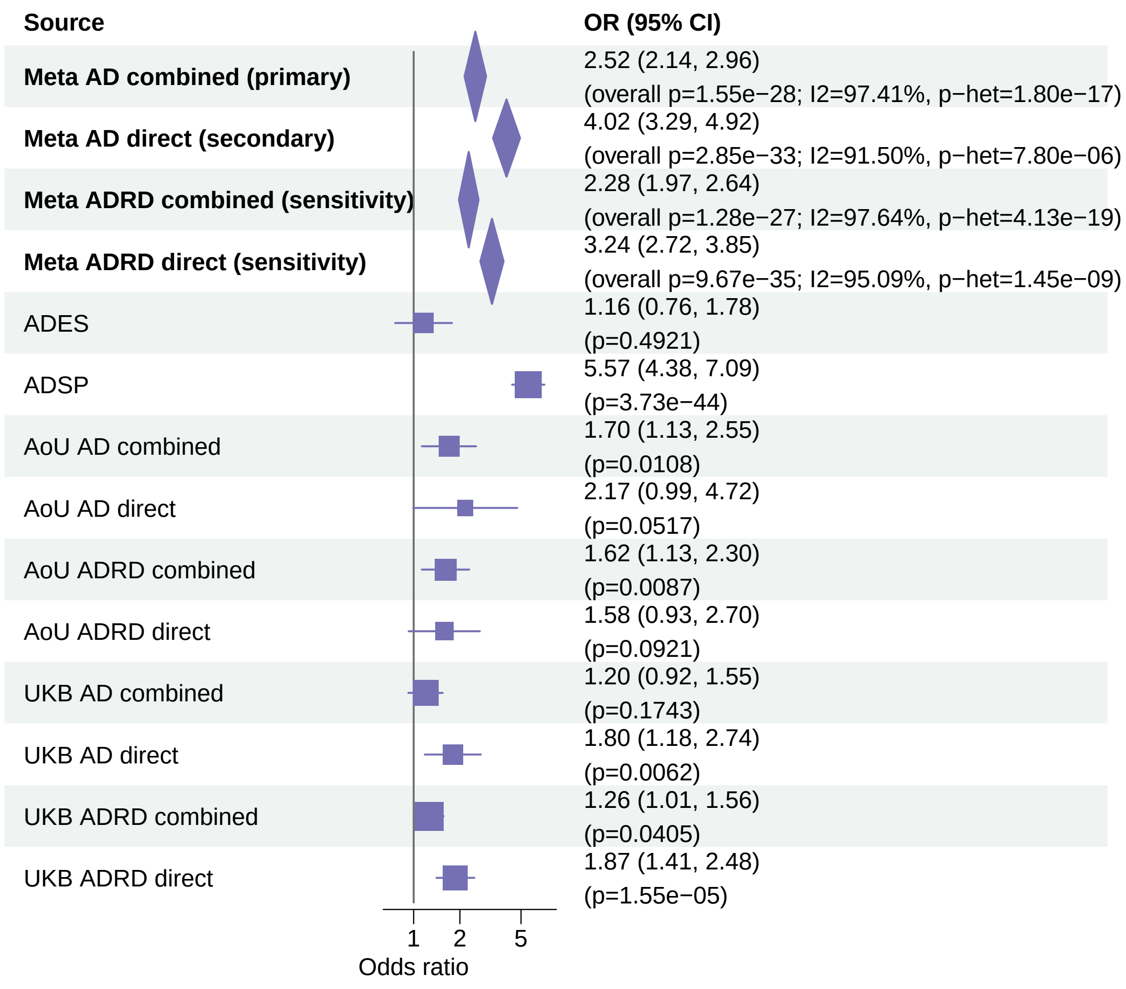


Meta OR/CI values are pooled REGENIE effects. Meta overall P values use ADES+REGENIE signed−Z evidence when the exact ADES mask was available; otherwise they use REGENIE.

#### AD primary/secondary definitions with ADRD sensitivity − ABCA7 − LOF+REVEL75 (ALL)

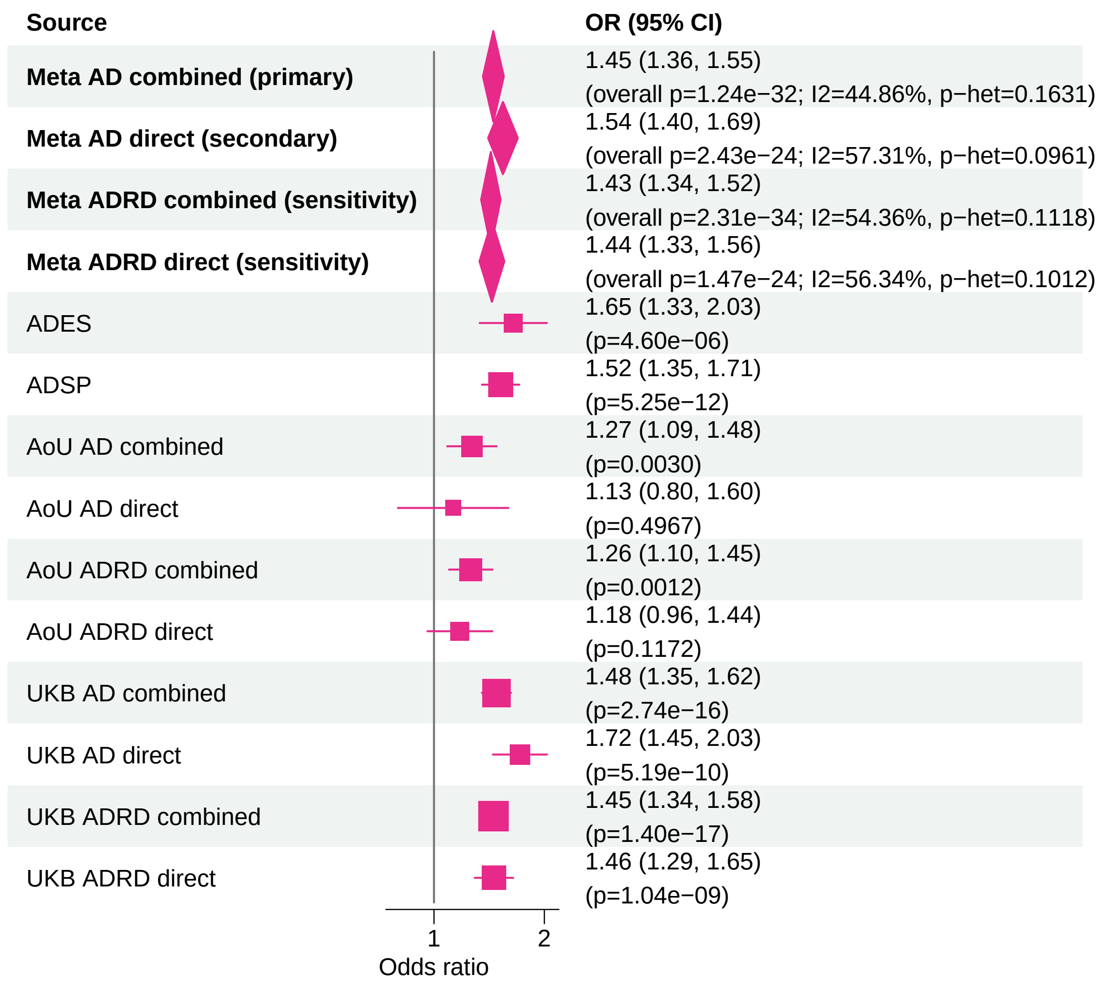


Meta OR/CI values are pooled REGENIE effects. Meta overall P values use ADES+REGENIE signed−Z evidence when the exact ADES mask was available; otherwise they use REGENIE.

### AD primary/secondary definitions with ADRD sensitivity − PSEN1 − LOF+REVEL25 (ALL)

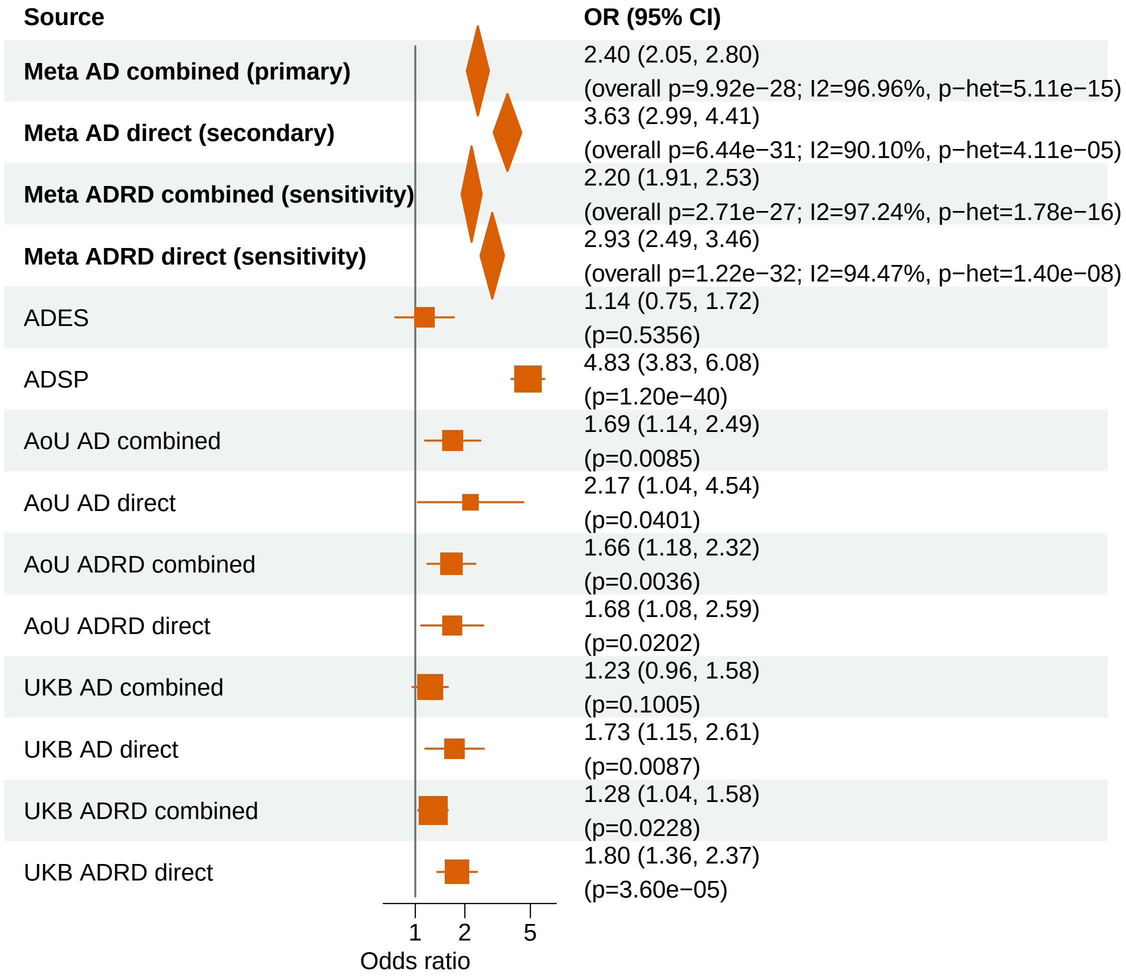


Meta OR/CI values are pooled REGENIE effects. Meta overall P values use ADES+REGENIE signed−Z evidence when the exact ADES mask was available; otherwise they use REGENIE.

#### AD primary/secondary definitions with ADRD sensitivity − ABCA7 − LOF+REVEL50 (ALL)

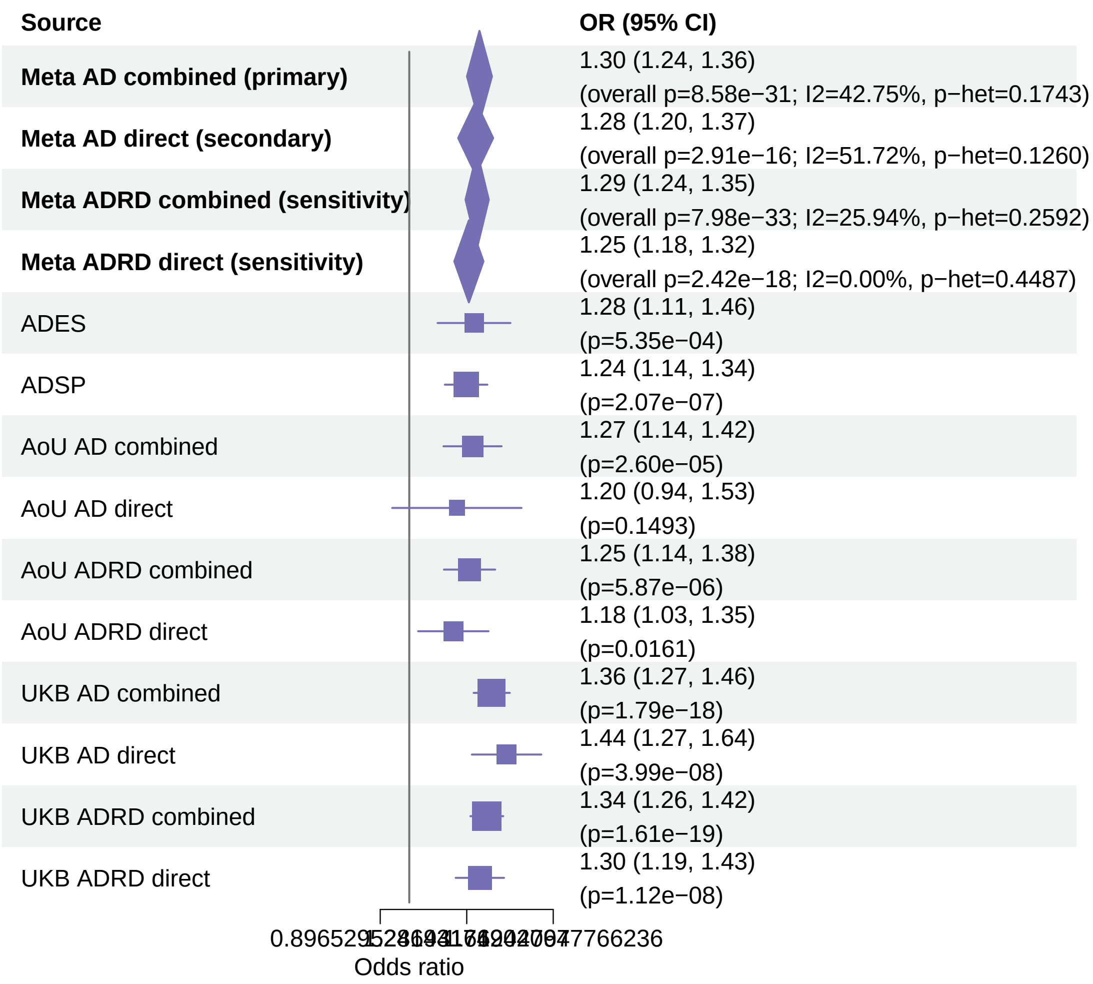


Meta OR/CI values are pooled REGENIE effects. Meta overall P values use ADES+REGENIE signed−Z evidence when the exact ADES mask was available; otherwise they use REGENIE.

### AD primary/secondary definitions with ADRD sensitivity − SORL1 − LOF+REVEL50 (ALL)

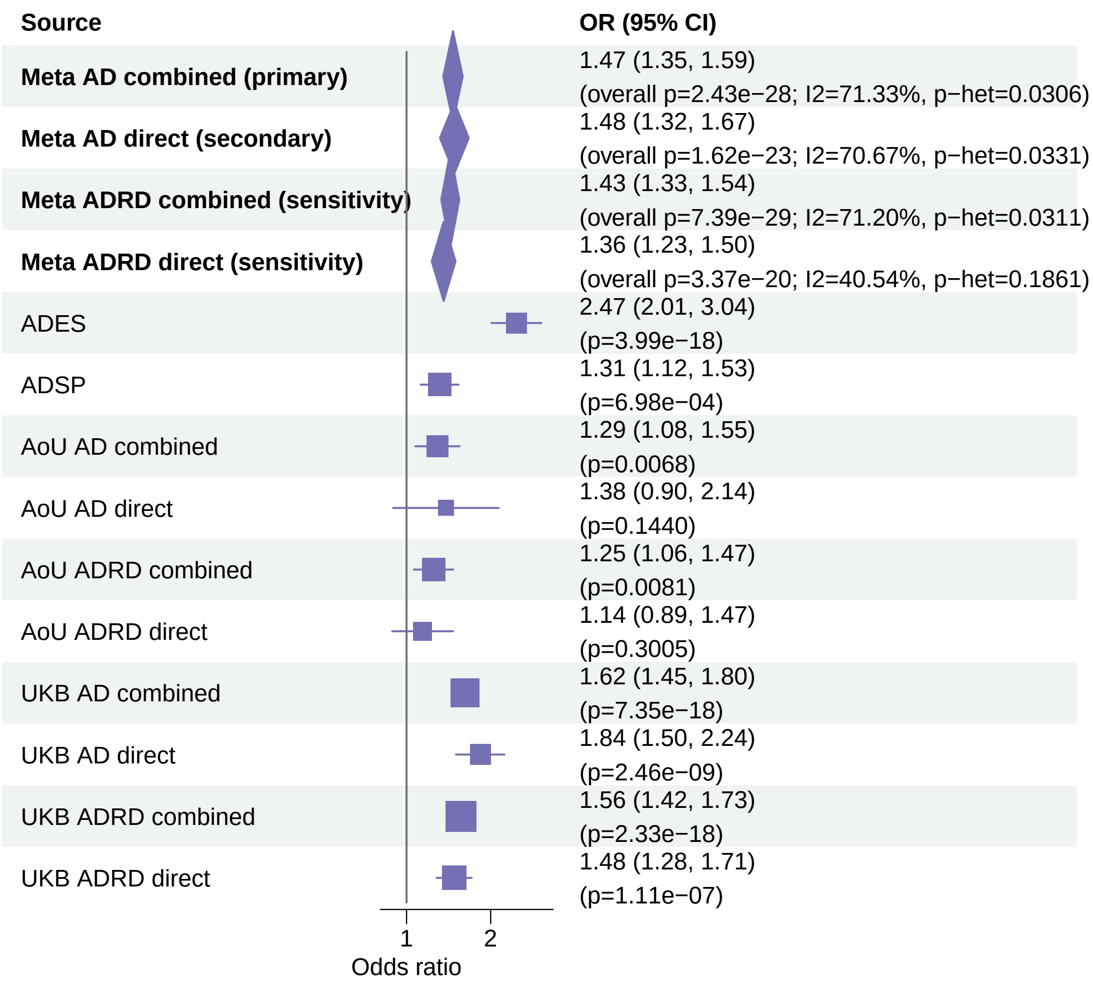


Meta OR/CI values are pooled REGENIE effects. Meta overall P values use ADES+REGENIE signed−Z evidence when the exact ADES mask was available; otherwise they use REGENIE.

### AD primary/secondary definitions with ADRD sensitivity − SORL1 − LOF+REVEL75 (ALL)

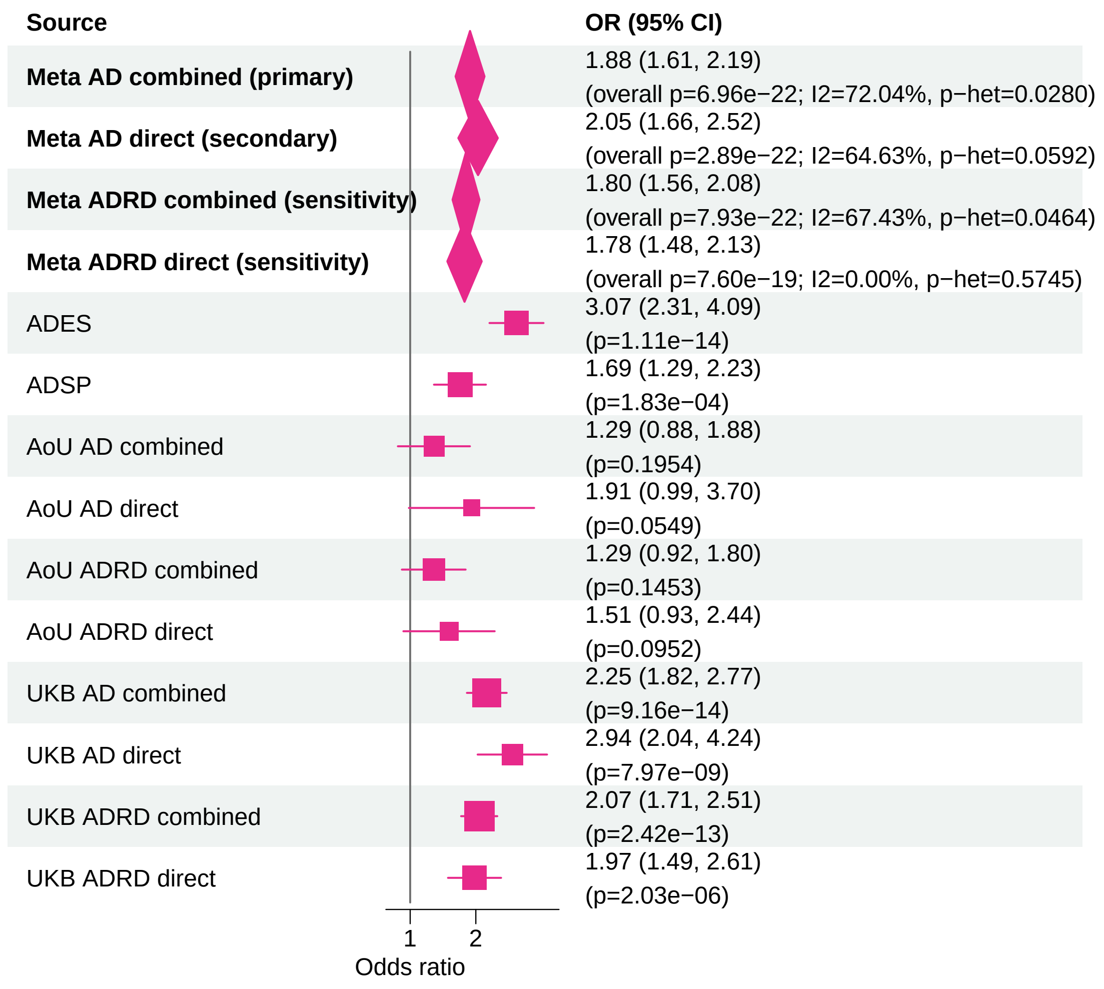


Meta OR/CI values are pooled REGENIE effects. Meta overall P values use ADES+REGENIE signed−Z evidence when the exact ADES mask was available; otherwise they use REGENIE.

#### AD primary/secondary definitions with ADRD sensitivity − SORL1 − LOF (ALL)

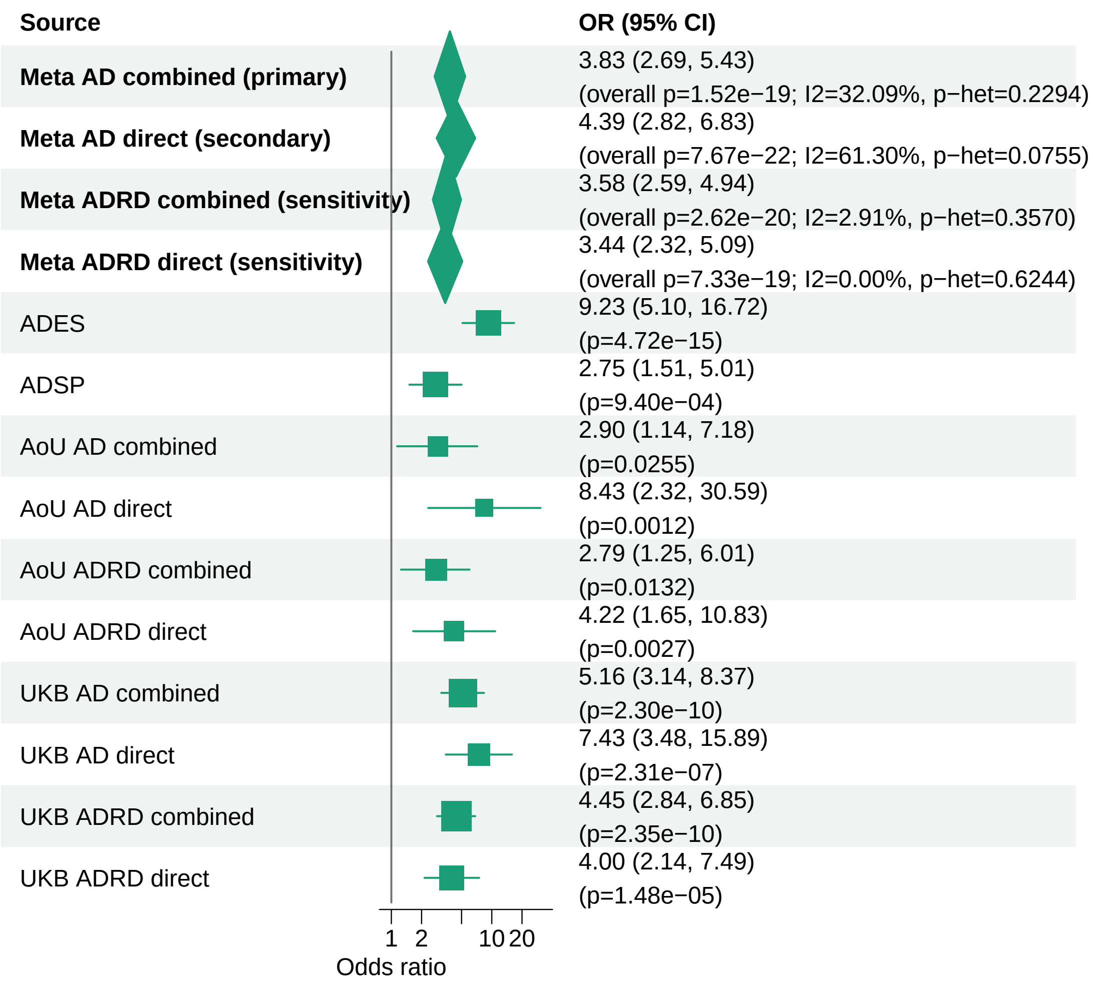


Meta OR/CI values are pooled REGENIE effects. Meta overall P values use ADES+REGENIE signed−Z evidence when the exact ADES mask was available; otherwise they use REGENIE.

#### AD primary/secondary definitions with ADRD sensitivity − ABCA7 − LOF (ALL)

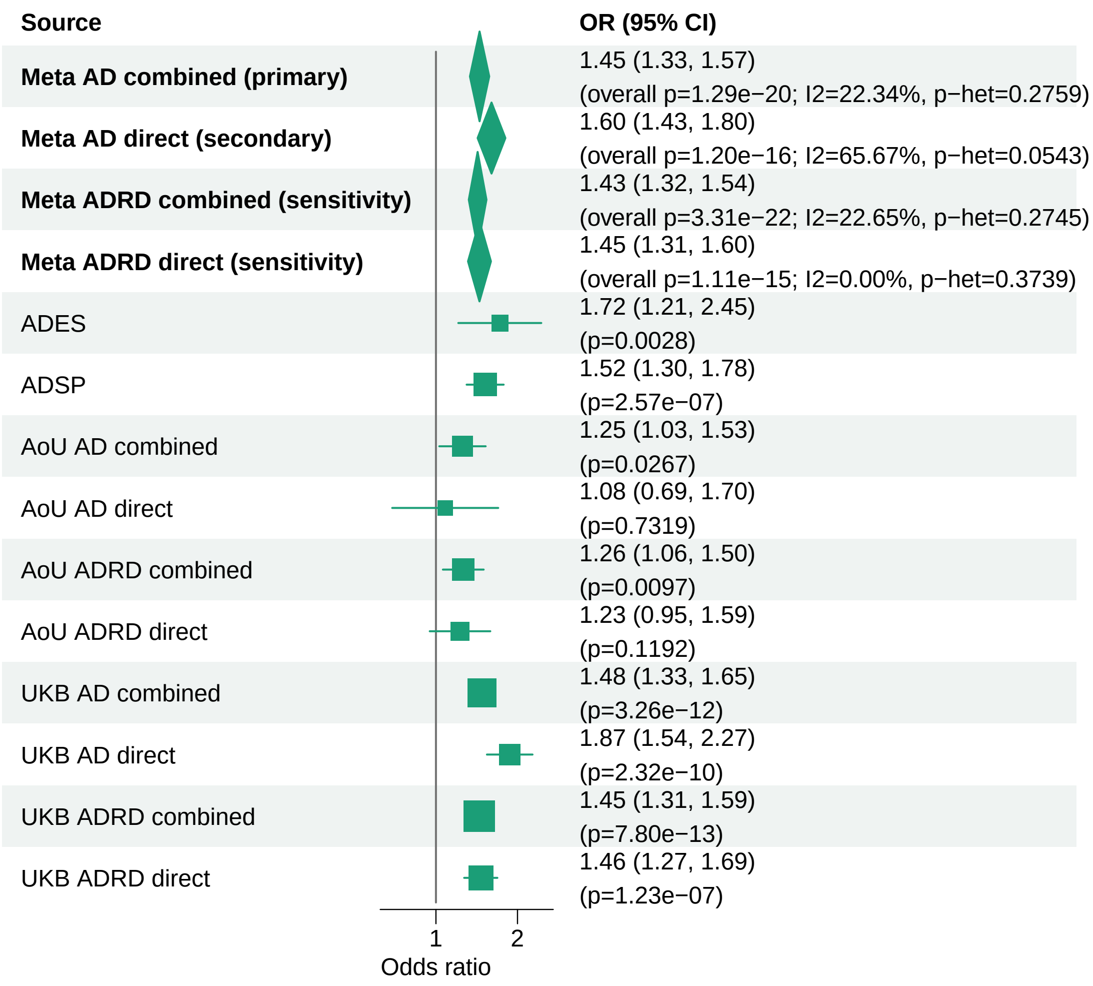


Meta OR/CI values are pooled REGENIE effects. Meta overall P values use ADES+REGENIE signed−Z evidence when the exact ADES mask was available; otherwise they use REGENIE.

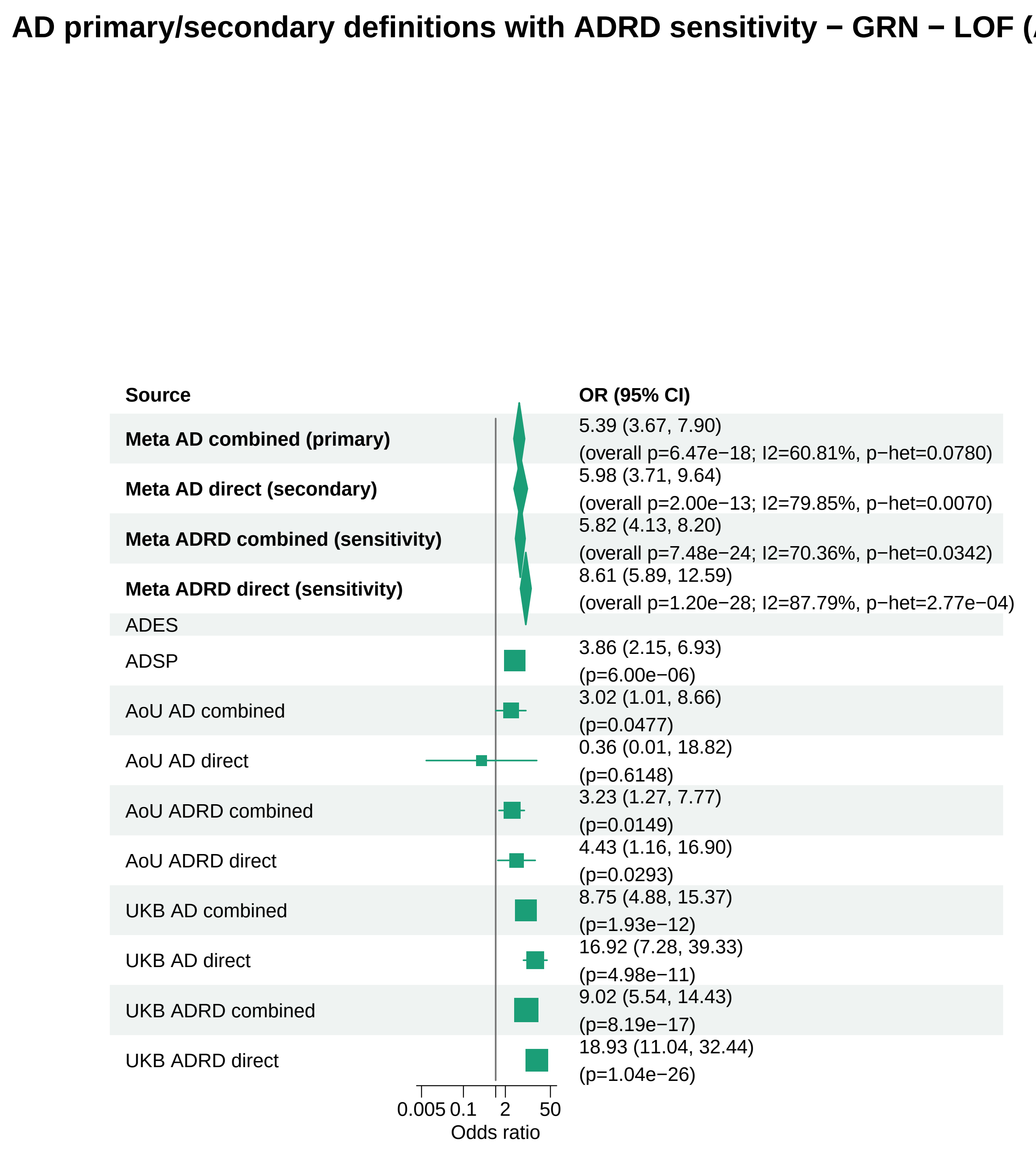


Meta OR/CI values are pooled REGENIE effects. Meta overall P values use ADES+REGENIE signed−Z evidence when the exact ADES mask was available; otherwise they use REGENIE.

#### AD primary/secondary definitions with ADRD sensitivity − SORL1 − LOF+REVEL25 (ALL)

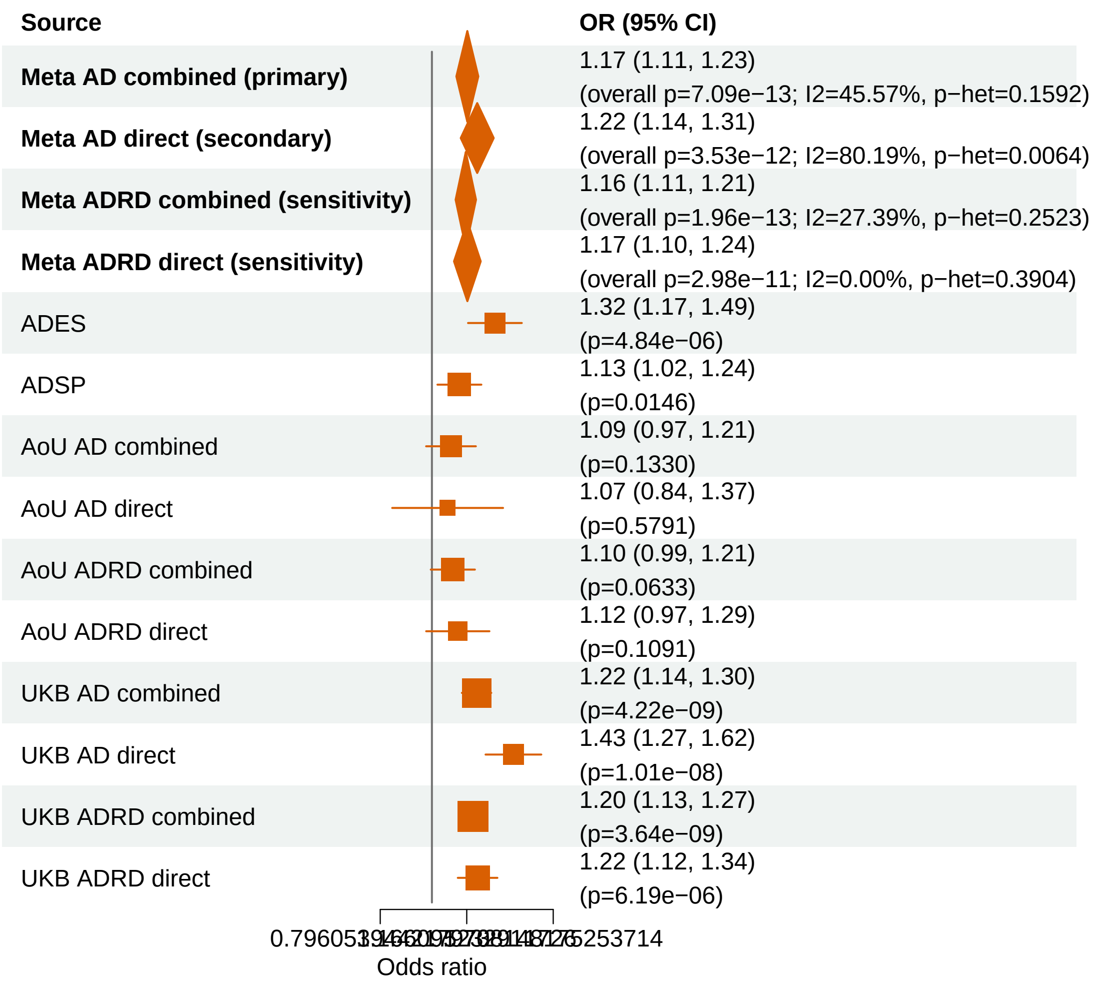


Meta OR/CI values are pooled REGENIE effects. Meta overall P values use ADES+REGENIE signed−Z evidence when the exact ADES mask was available; otherwise they use REGENIE.

#### AD primary/secondary definitions with ADRD sensitivity − GRN − LOF+REVEL75 (ALL)

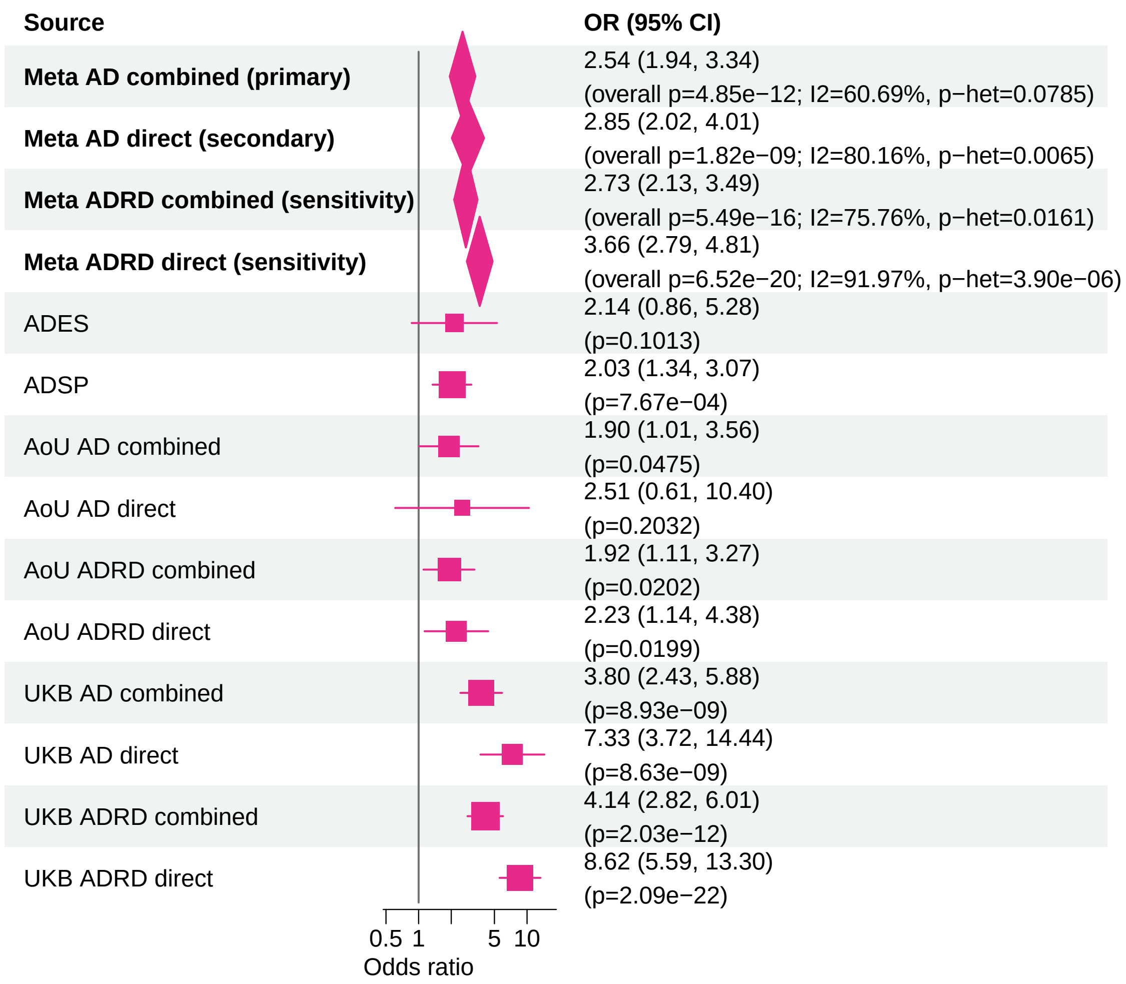


Meta OR/CI values are pooled REGENIE effects. Meta overall P values use ADES+REGENIE signed−Z evidence when the exact ADES mask was available; otherwise they use REGENIE.

#### AD primary/secondary definitions with ADRD sensitivity − ATP8B4 − LOF+REVEL50 (ALL)

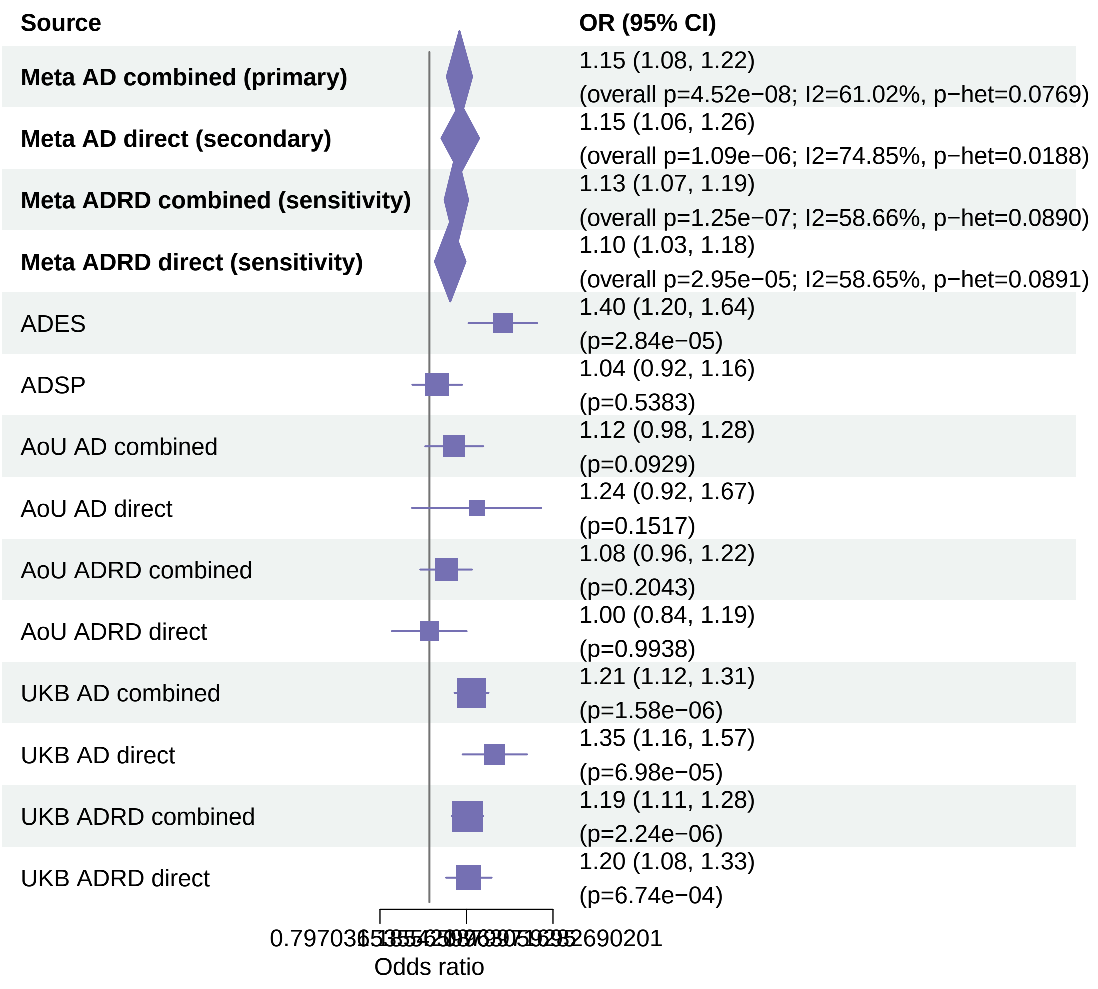


Meta OR/CI values are pooled REGENIE effects. Meta overall P values use ADES+REGENIE signed−Z evidence when the exact ADES mask was available; otherwise they use REGENIE.

#### AD primary/secondary definitions with ADRD sensitivity − TREM2 − LOF+REVEL50 (ALL)

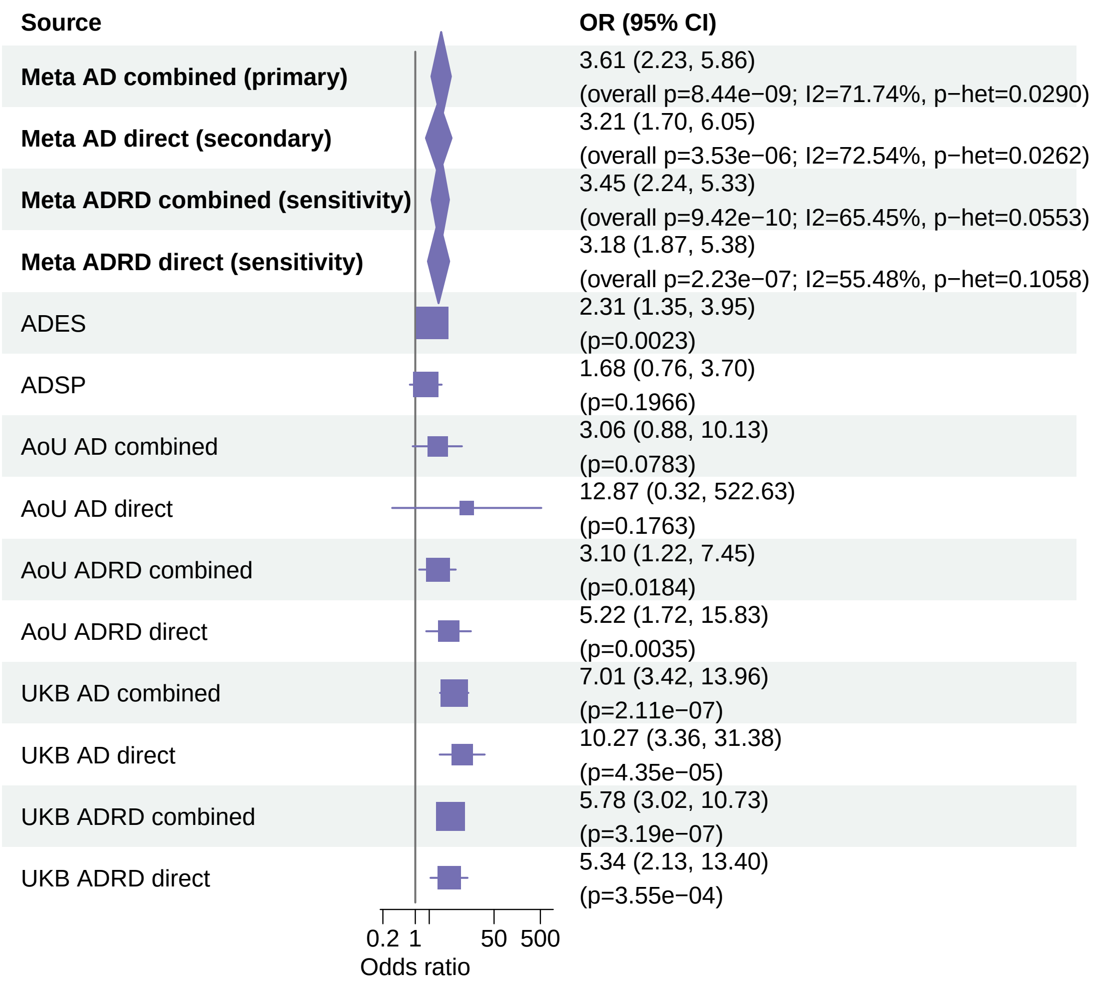


Meta OR/CI values are pooled REGENIE effects. Meta overall P values use ADES+REGENIE signed−Z evidence when the exact ADES mask was available; otherwise they use REGENIE.

**AD primary/secondary definitions with ADRD sensitivity − ATP8B4 − LOF+REVEL75 (ALL)**

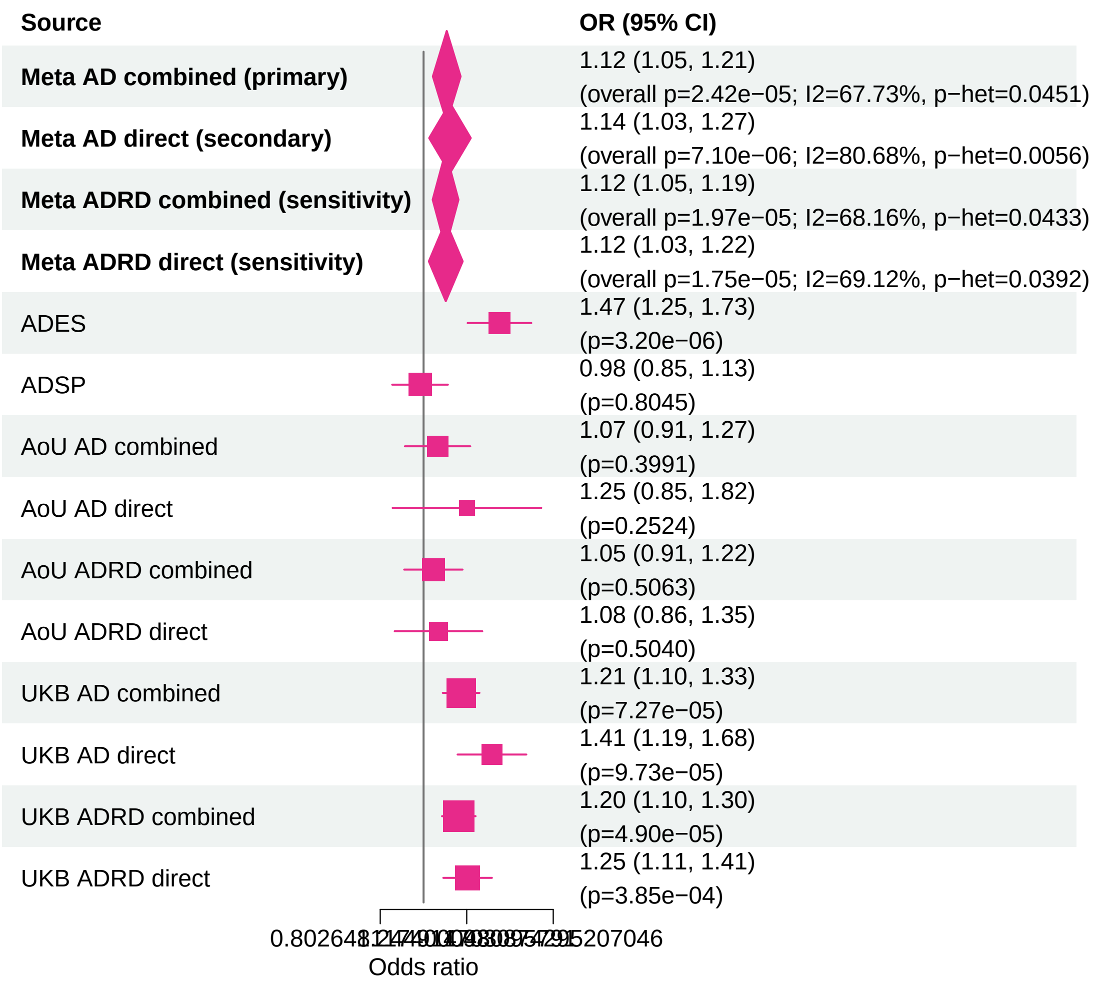


Meta OR/CI values are pooled REGENIE effects. Meta overall P values use ADES+REGENIE signed−Z evidence when the exact ADES mask was available; otherwise they use REGENIE.

#### AD primary/secondary definitions with ADRD sensitivity − ATP8B4 − LOF+REVEL25 (ALL)

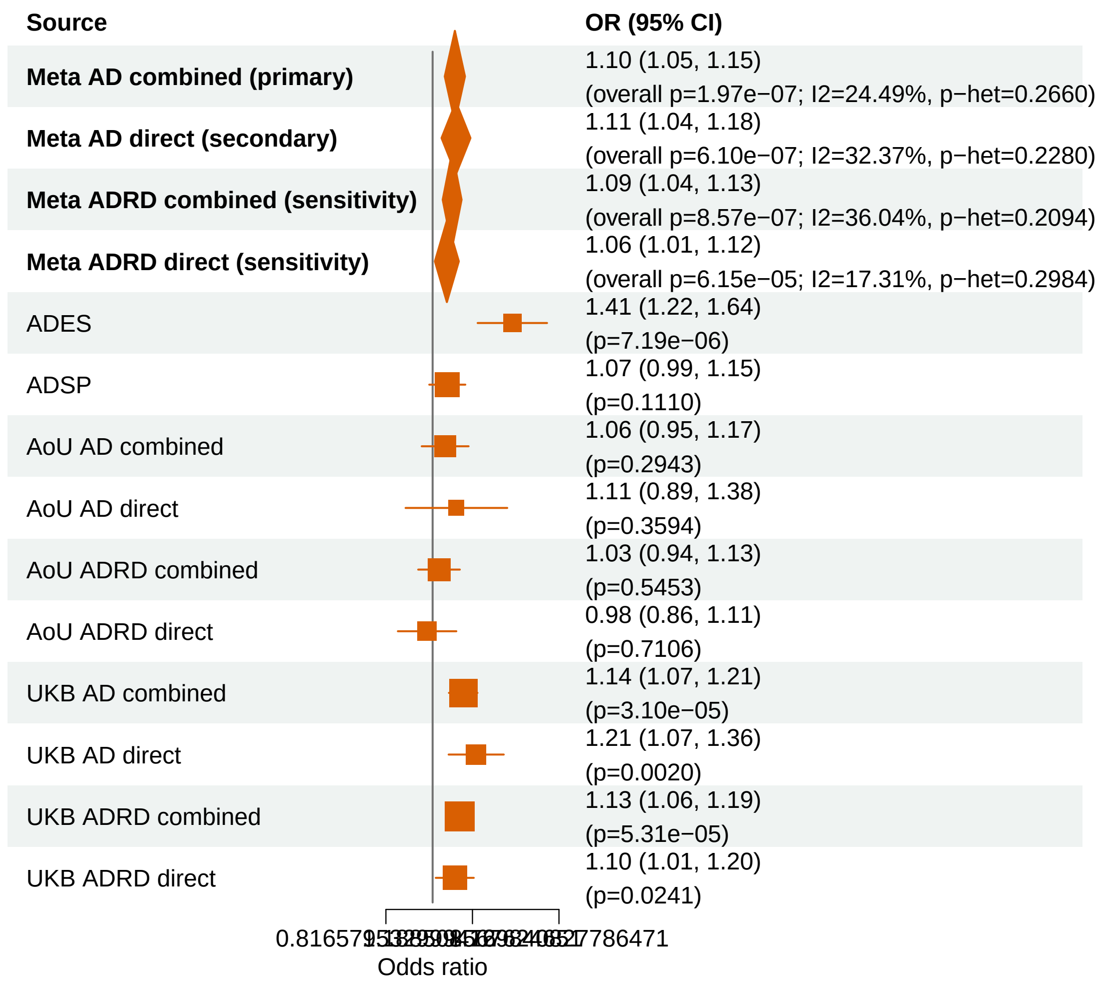


Meta OR/CI values are pooled REGENIE effects. Meta overall P values use ADES+REGENIE signed−Z evidence when the exact ADES mask was available; otherwise they use REGENIE.

#### AD primary/secondary definitions with ADRD sensitivity − TREM2 − LOF (ALL)

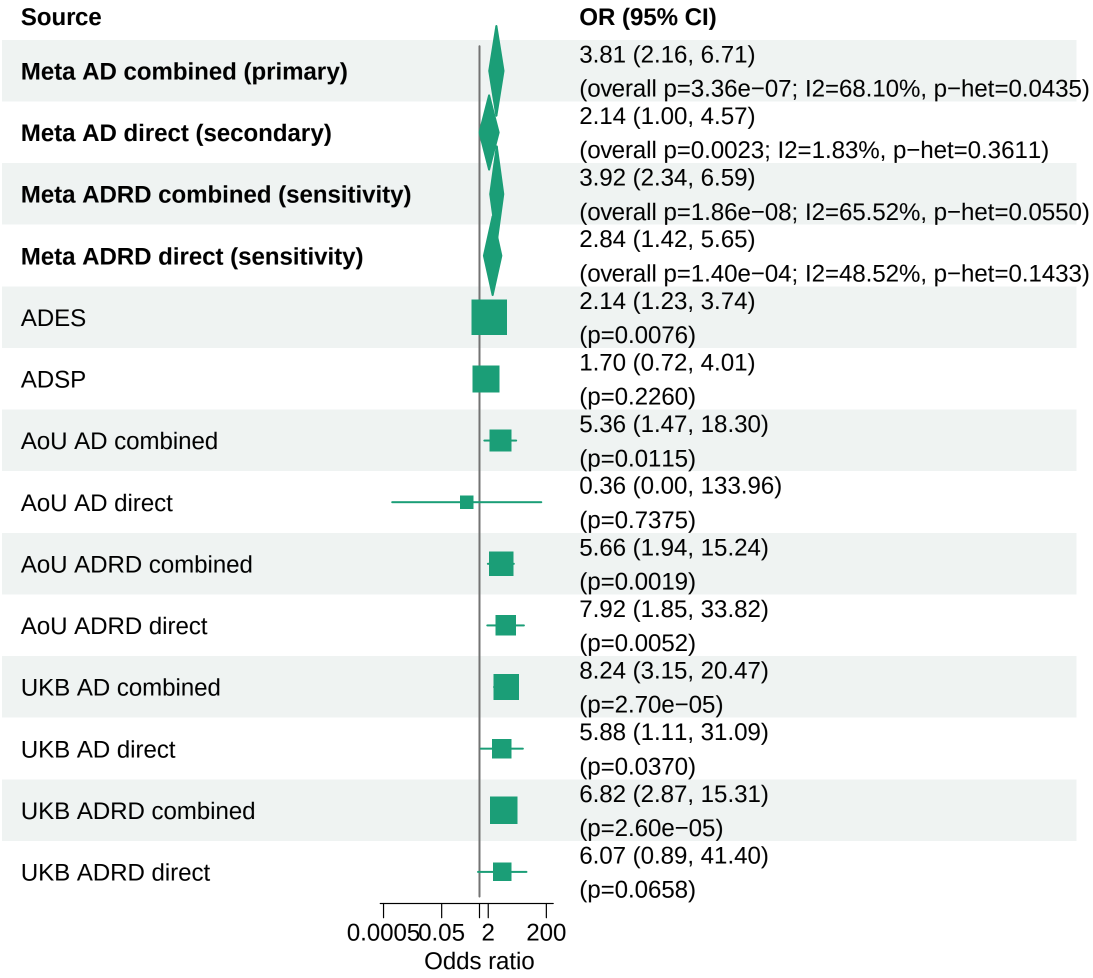


Meta OR/CI values are pooled REGENIE effects. Meta overall P values use ADES+REGENIE signed−Z evidence when the exact ADES mask was available; otherwise they use REGENIE.

#### AD primary/secondary definitions with ADRD sensitivity − ABCA1 − LOF+REVEL75 (ALL)

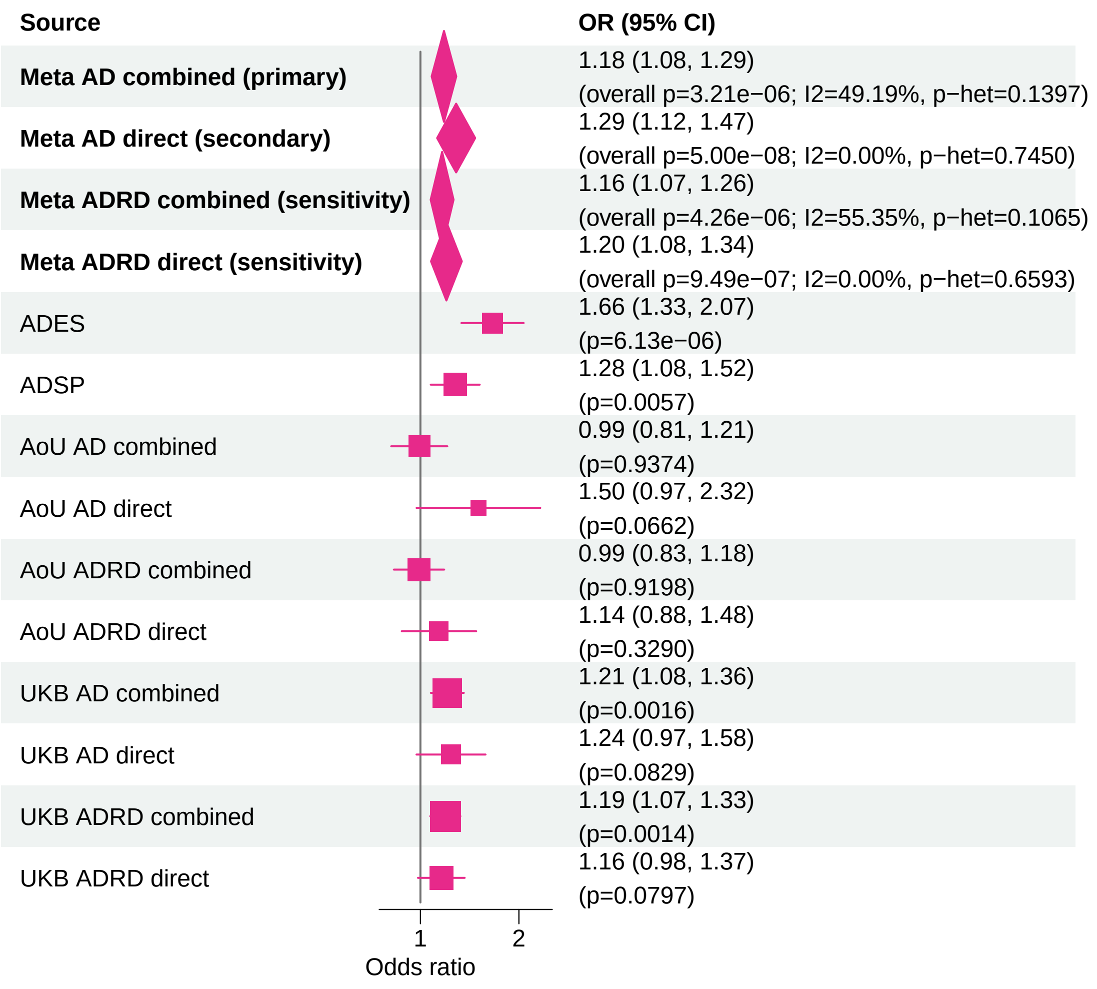


Meta OR/CI values are pooled REGENIE effects. Meta overall P values use ADES+REGENIE signed−Z evidence when the exact ADES mask was available; otherwise they use REGENIE.

#### AD primary/secondary definitions with ADRD sensitivity − ADAM10 − LOF+REVEL50 (ALL)

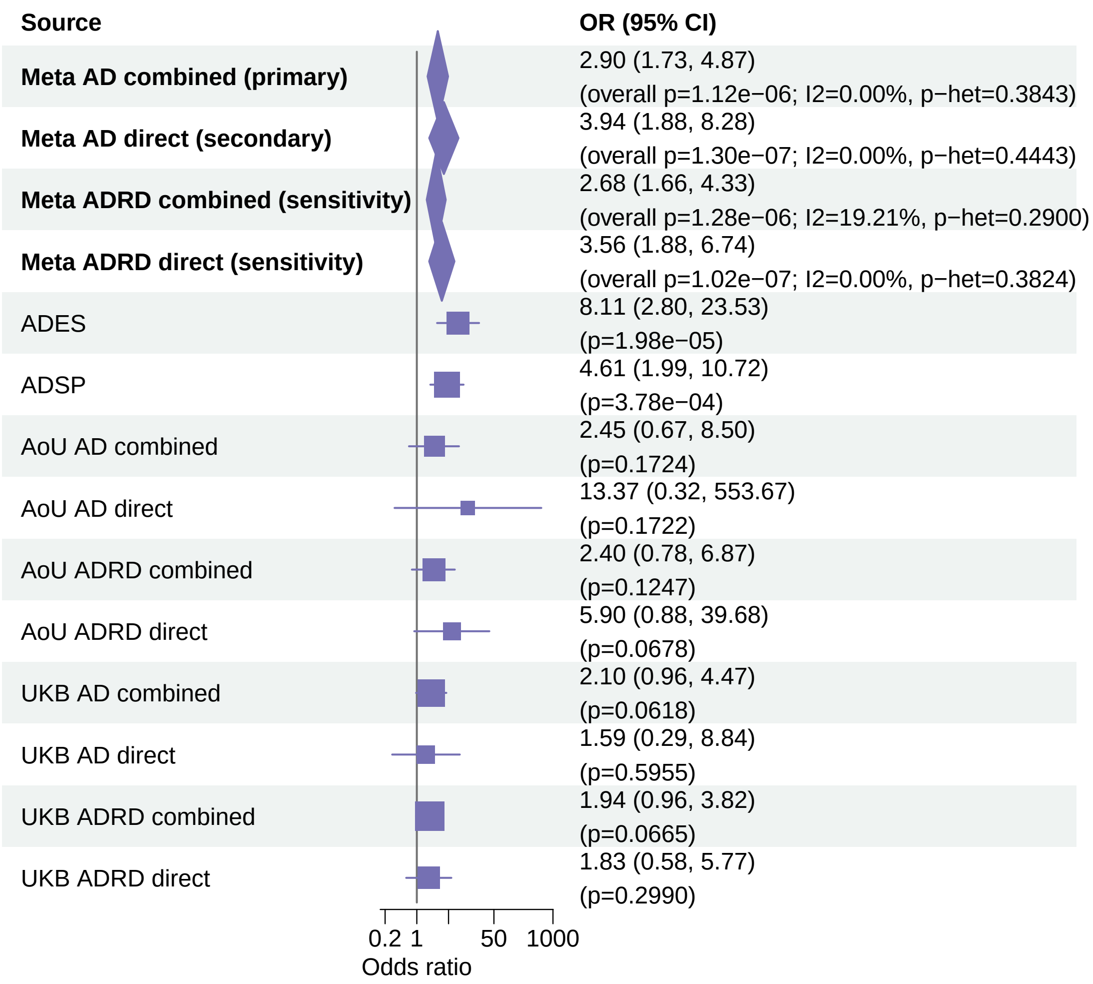


Meta OR/CI values are pooled REGENIE effects. Meta overall P values use ADES+REGENIE signed−Z evidence when the exact ADES mask was available; otherwise they use REGENIE.

#### AD primary/secondary definitions with ADRD sensitivity − TREM2 − LOF+REVEL75 (ALL)

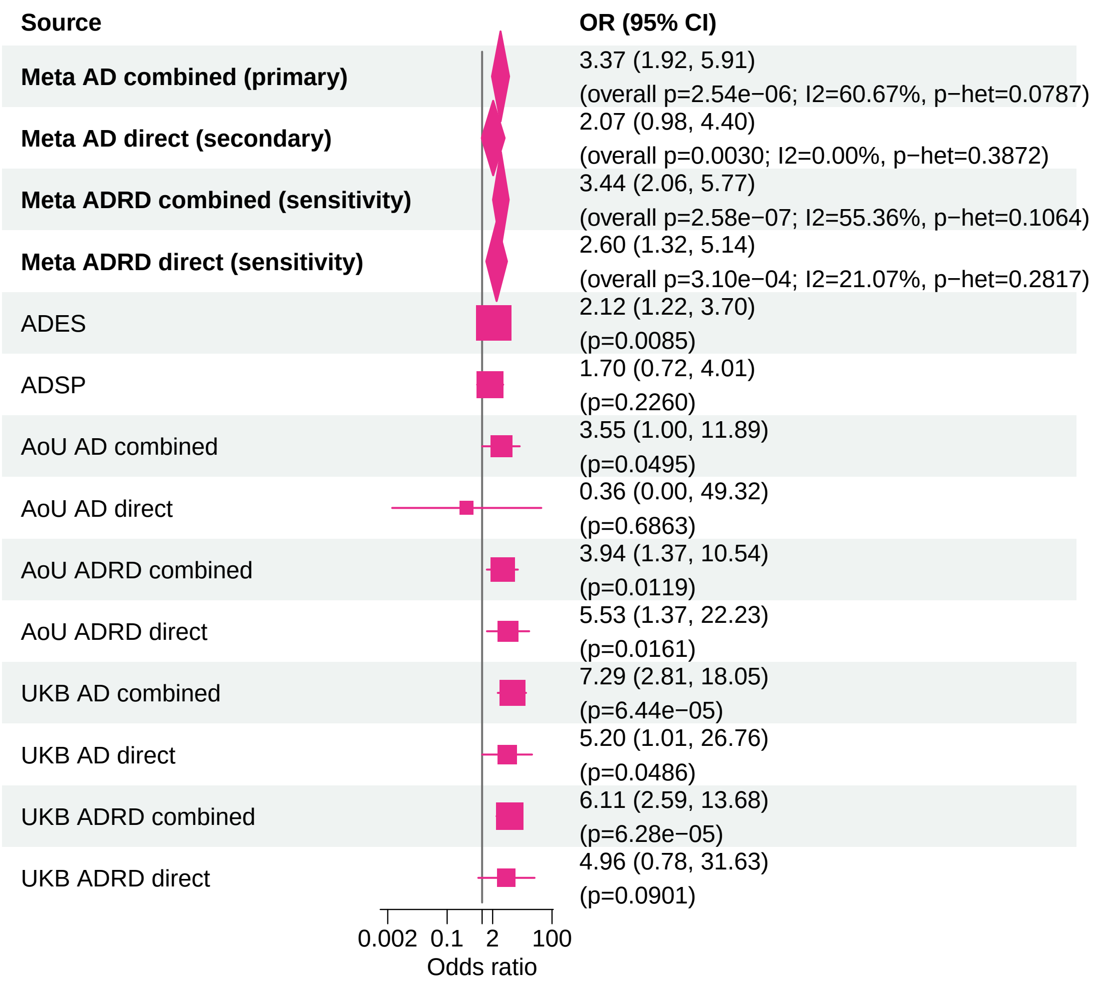


Meta OR/CI values are pooled REGENIE effects. Meta overall P values use ADES+REGENIE signed−Z evidence when the exact ADES mask was available; otherwise they use REGENIE.

#### AD primary/secondary definitions with ADRD sensitivity − TBK1 − LOF+REVEL50 (ALL)

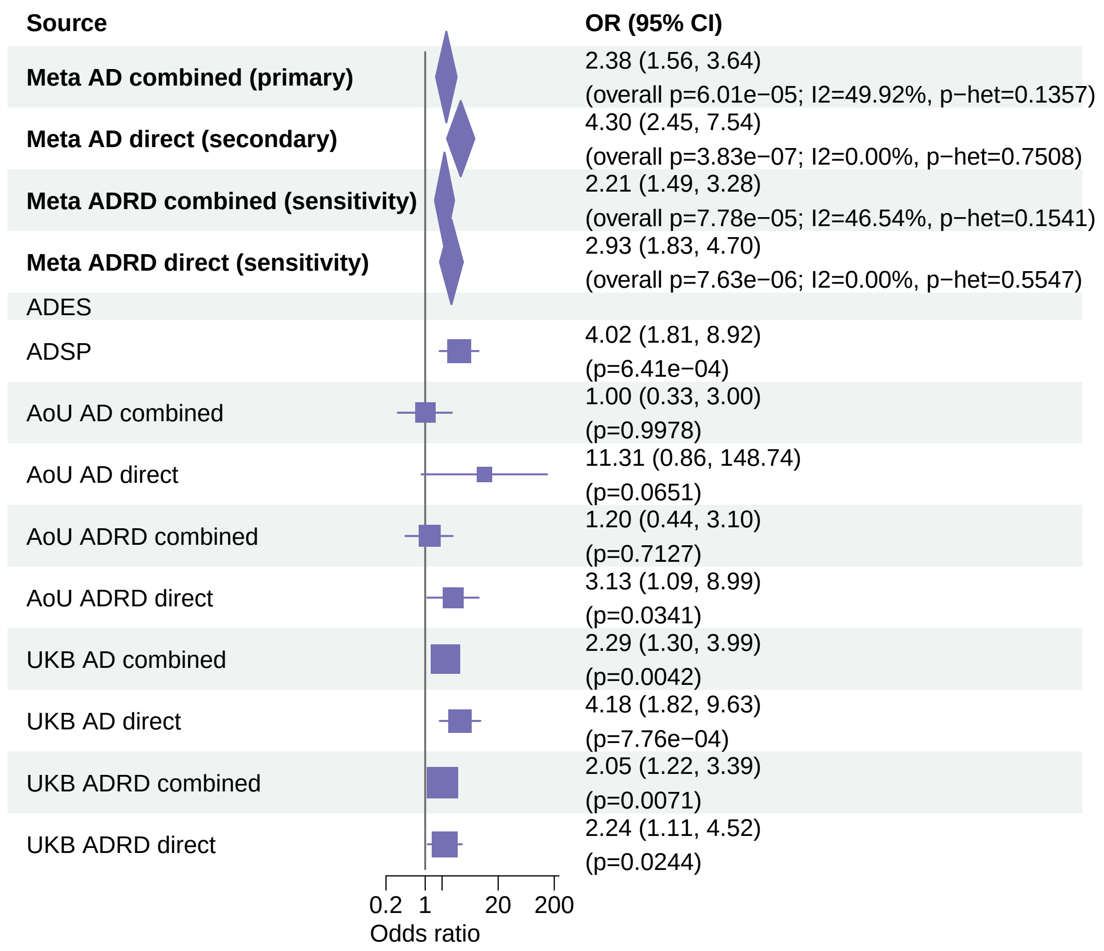


Meta OR/CI values are pooled REGENIE effects. Meta overall P values use ADES+REGENIE signed−Z evidence when the exact ADES mask was available; otherwise they use REGENIE.

#### AD primary/secondary definitions with ADRD sensitivity − ADAM10 − LOF+REVEL75 (ALL)

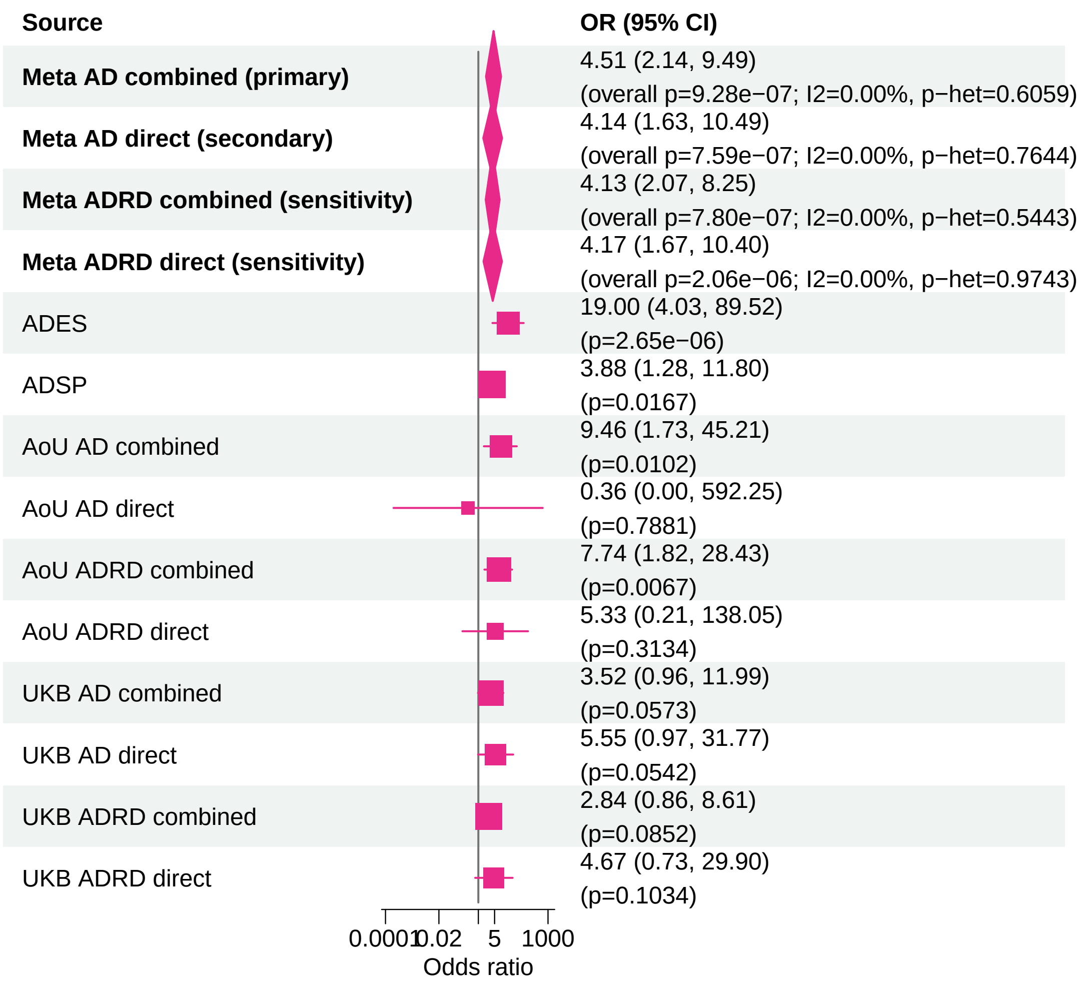


Meta OR/CI values are pooled REGENIE effects. Meta overall P values use ADES+REGENIE signed−Z evidence when the exact ADES mask was available; otherwise they use REGENIE.

#### AD primary/secondary definitions with ADRD sensitivity − IMPA2 − LOF+REVEL75 (ALL)

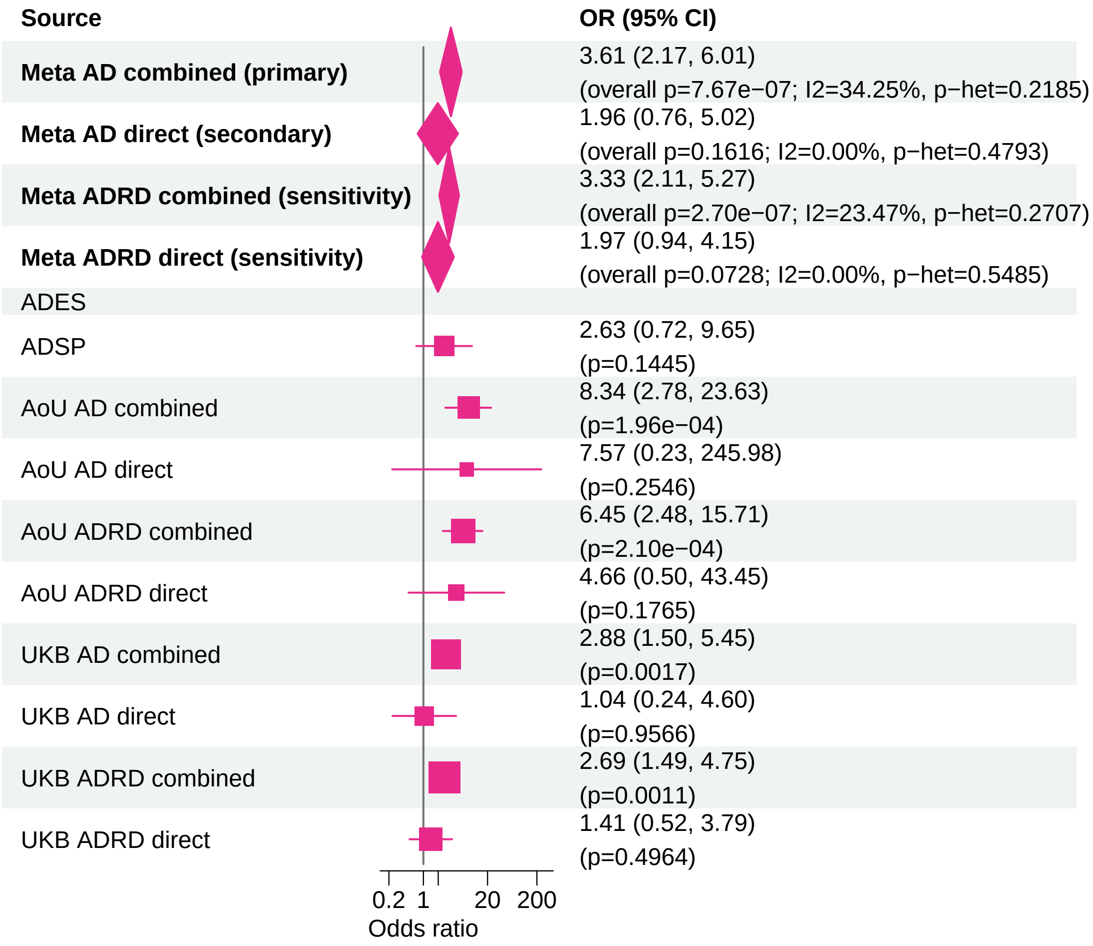


Meta OR/CI values are pooled REGENIE effects. Meta overall P values use ADES+REGENIE signed−Z evidence when the exact ADES mask was available; otherwise they use REGENIE.

#### AD primary/secondary definitions with ADRD sensitivity − ADAM10 − LOF+REVEL25 (ALL)

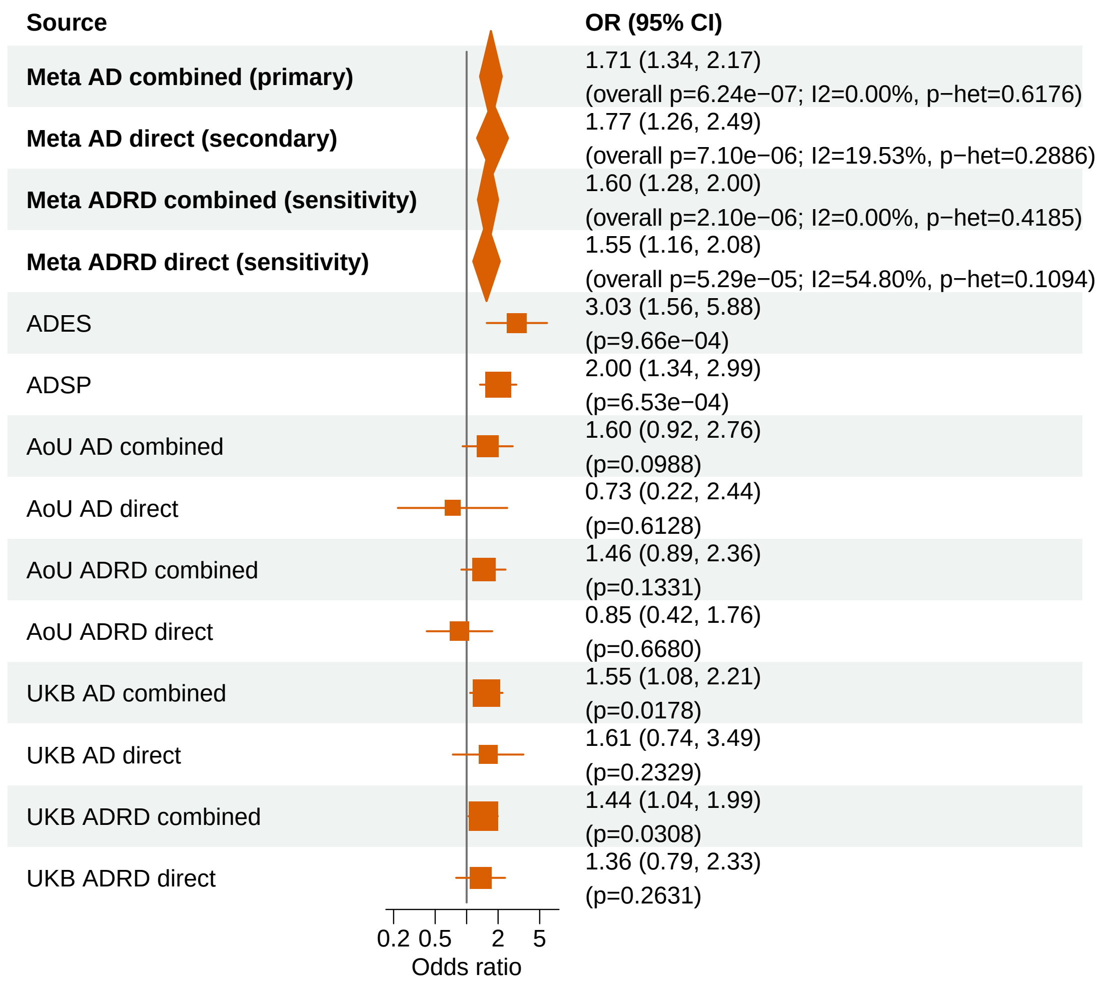


Meta OR/CI values are pooled REGENIE effects. Meta overall P values use ADES+REGENIE signed−Z evidence when the exact ADES mask was available; otherwise they use REGENIE.

### AD primary/secondary definitions with ADRD sensitivity − TBK1 − LOF (ALL)

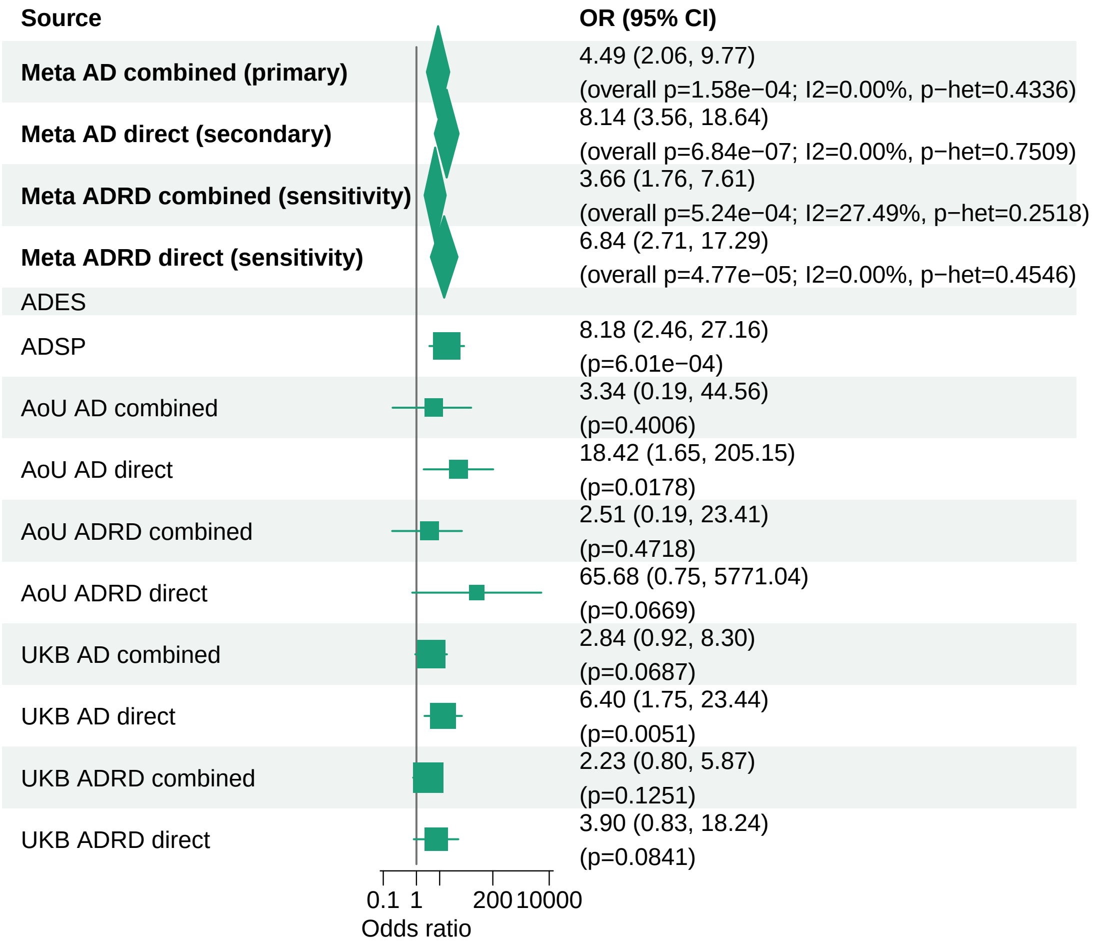


Meta OR/CI values are pooled REGENIE effects. Meta overall P values use ADES+REGENIE signed−Z evidence when the exact ADES mask was available; otherwise they use REGENIE.

#### AD primary/secondary definitions with ADRD sensitivity − TBK1 − LOF+REVEL75 (ALL)

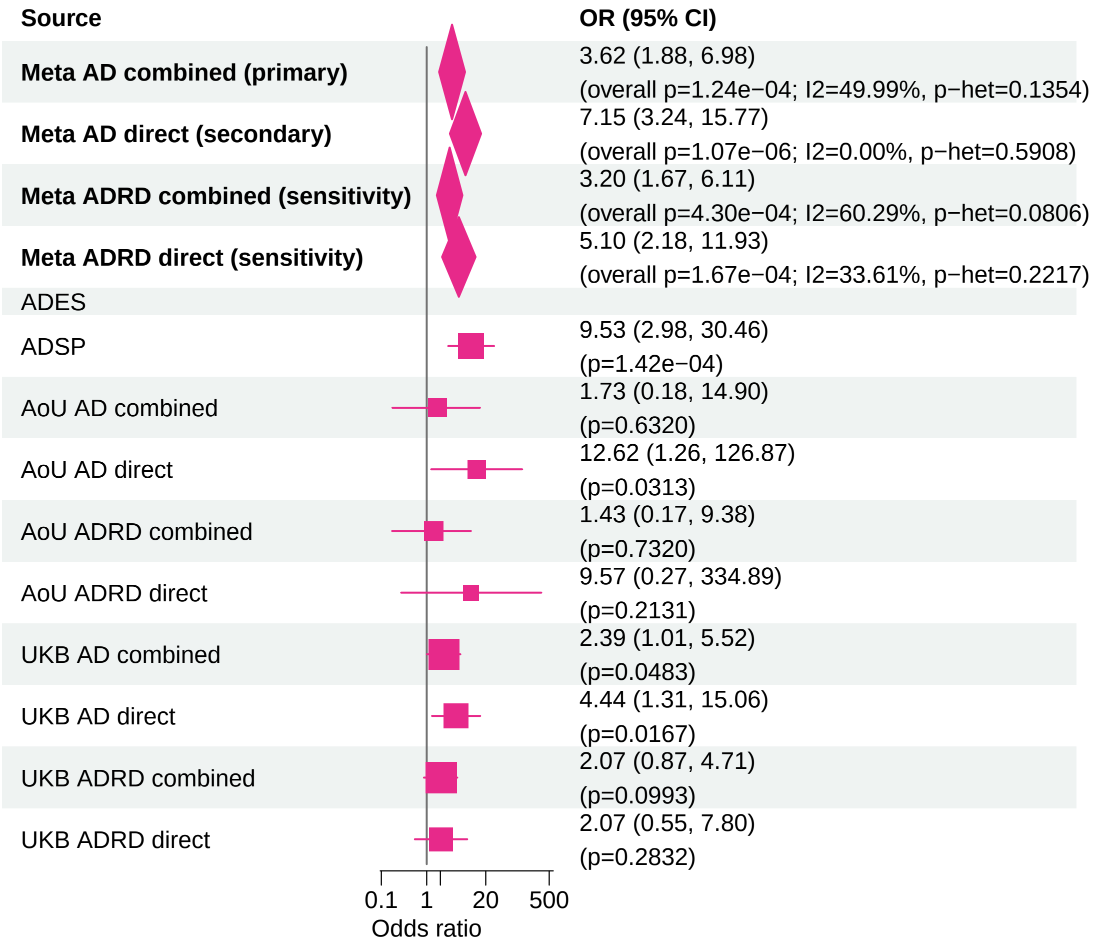


Meta OR/CI values are pooled REGENIE effects. Meta overall P values use ADES+REGENIE signed−Z evidence when the exact ADES mask was available; otherwise they use REGENIE.

#### AD primary/secondary definitions with ADRD sensitivity − SHARPIN − LOF+REVEL25 (ALL)

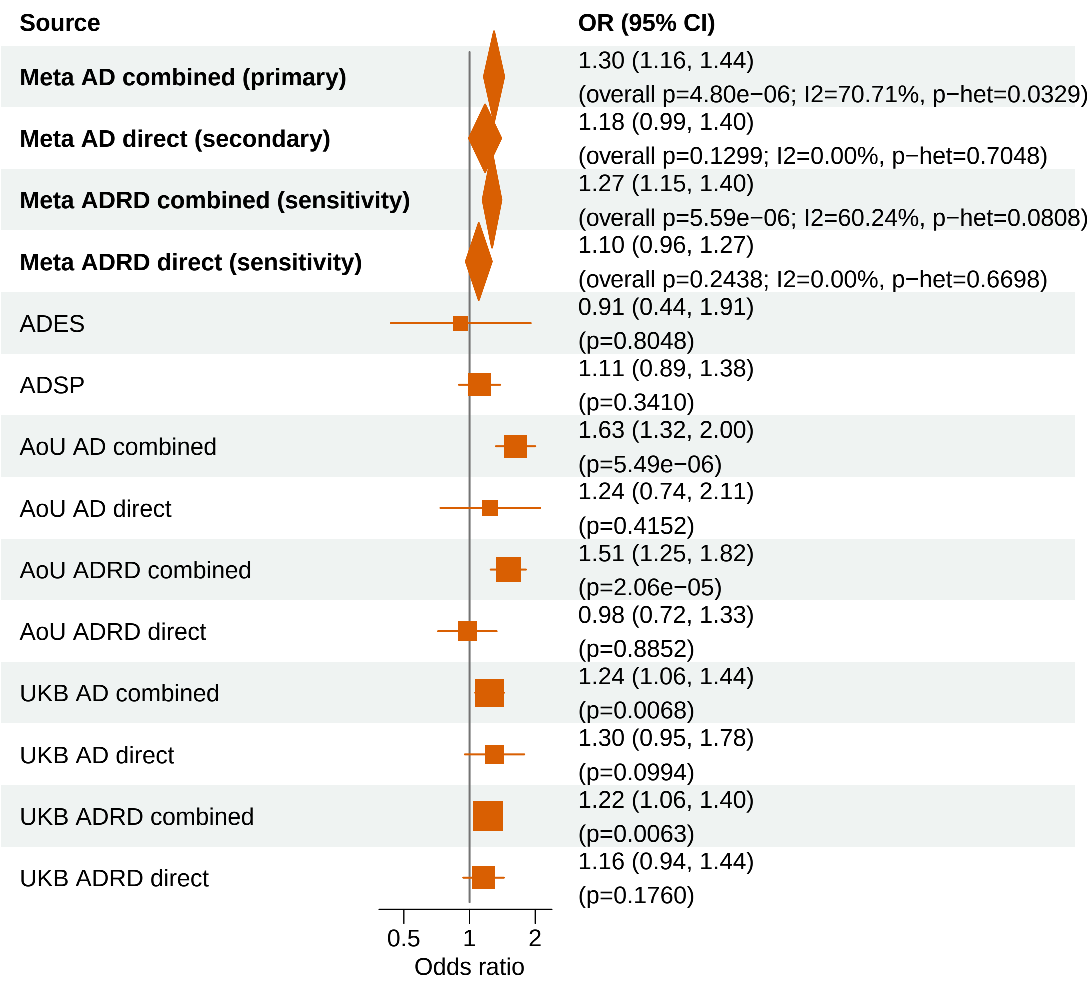


Meta OR/CI values are pooled REGENIE effects. Meta overall P values use ADES+REGENIE signed−Z evidence when the exact ADES mask was available; otherwise they use REGENIE.

### AD primary/secondary definitions with ADRD sensitivity − SYNE1 − LOF+REVEL50 (ALL)

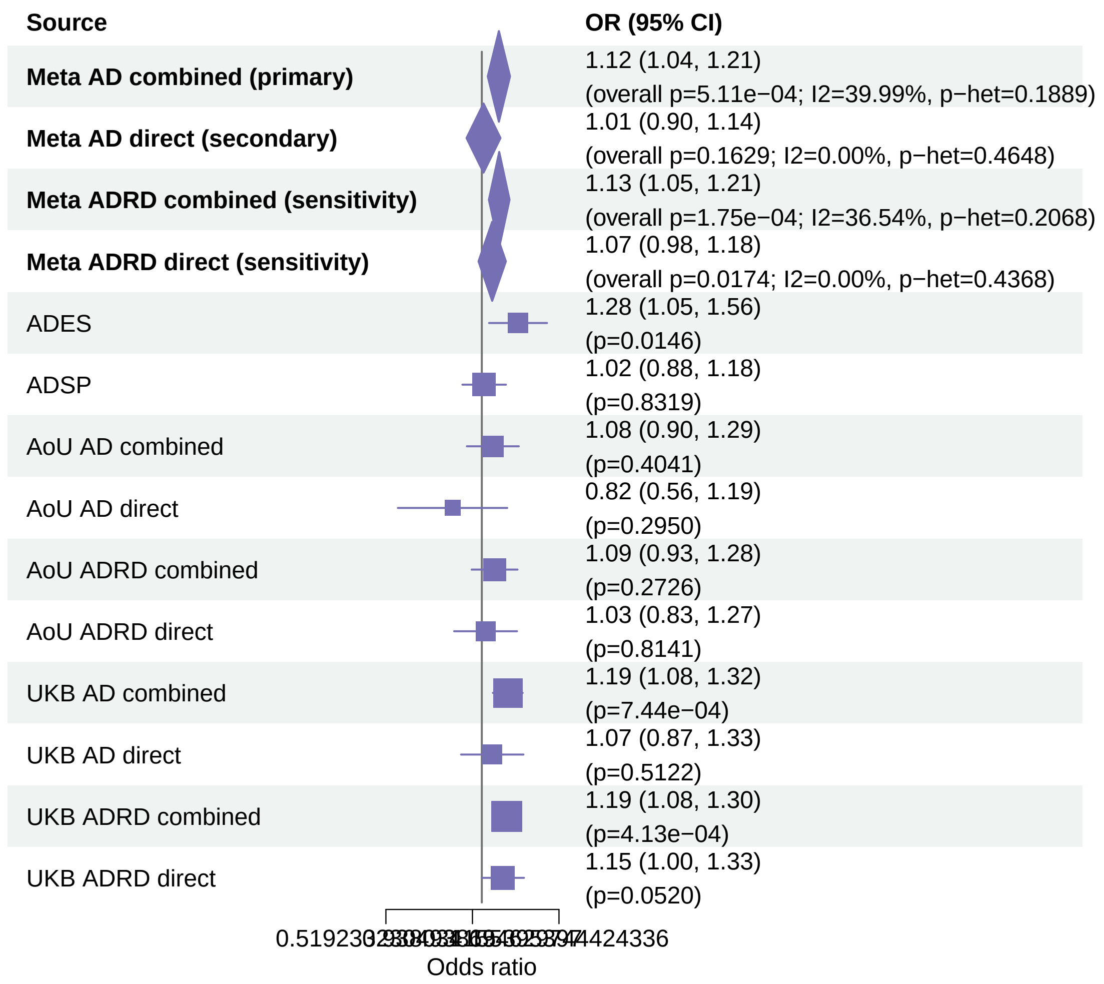


Meta OR/CI values are pooled REGENIE effects. Meta overall P values use ADES+REGENIE signed−Z evidence when the exact ADES mask was available; otherwise they use REGENIE.

#### AD primary/secondary definitions with ADRD sensitivity − APOE − LOF+REVEL25 (ALL)

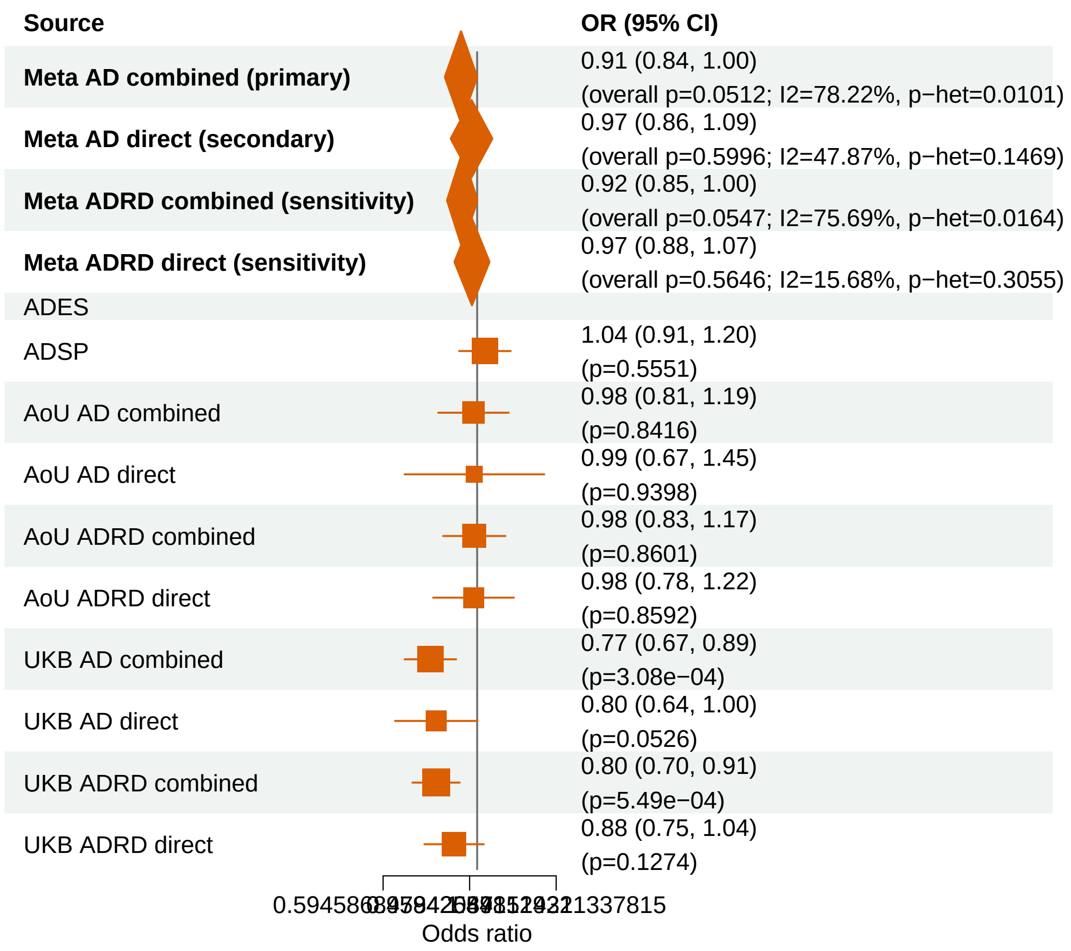


Meta OR/CI values are pooled REGENIE effects. Meta overall P values use ADES+REGENIE signed−Z evidence when the exact ADES mask was available; otherwise they use REGENIE.

#### AD primary/secondary definitions with ADRD sensitivity − GRN − LOF+REVEL50 (ALL)

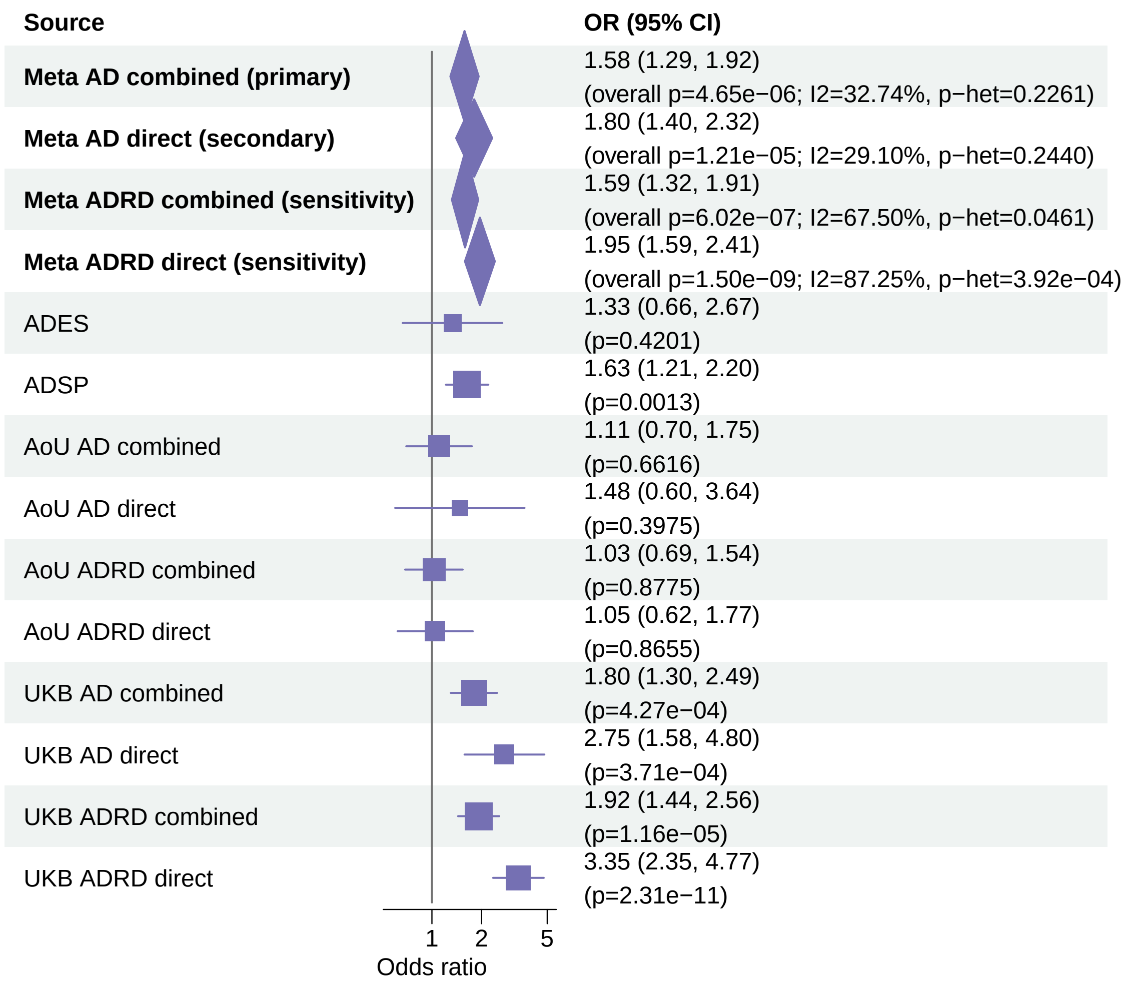


Meta OR/CI values are pooled REGENIE effects. Meta overall P values use ADES+REGENIE signed−Z evidence when the exact ADES mask was available; otherwise they use REGENIE.

### AD primary/secondary definitions with ADRD sensitivity − RIN3 − LOF+REVEL50 (ALL)

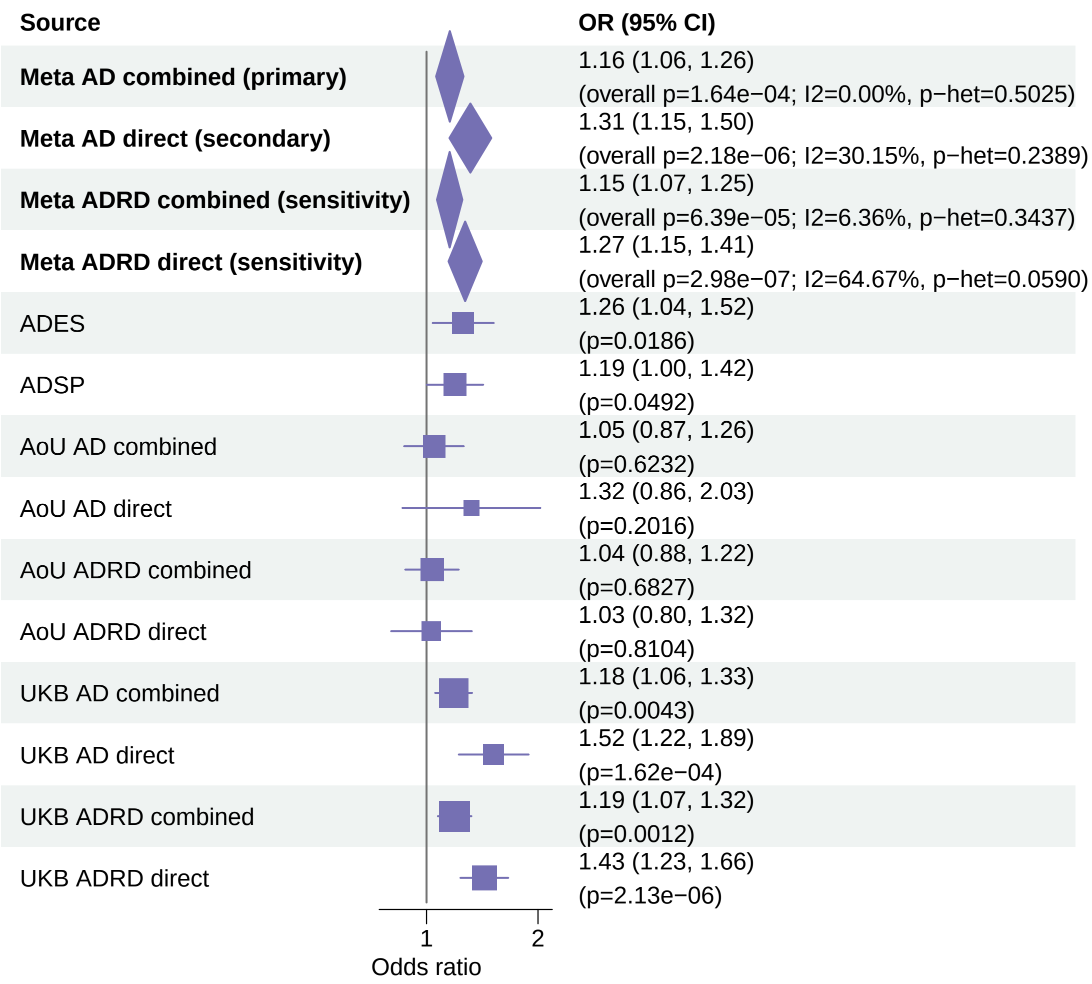


Meta OR/CI values are pooled REGENIE effects. Meta overall P values use ADES+REGENIE signed−Z evidence when the exact ADES mask was available; otherwise they use REGENIE.

#### AD primary/secondary definitions with ADRD sensitivity − CHRNA4 − LOF+REVEL50 (ALL)

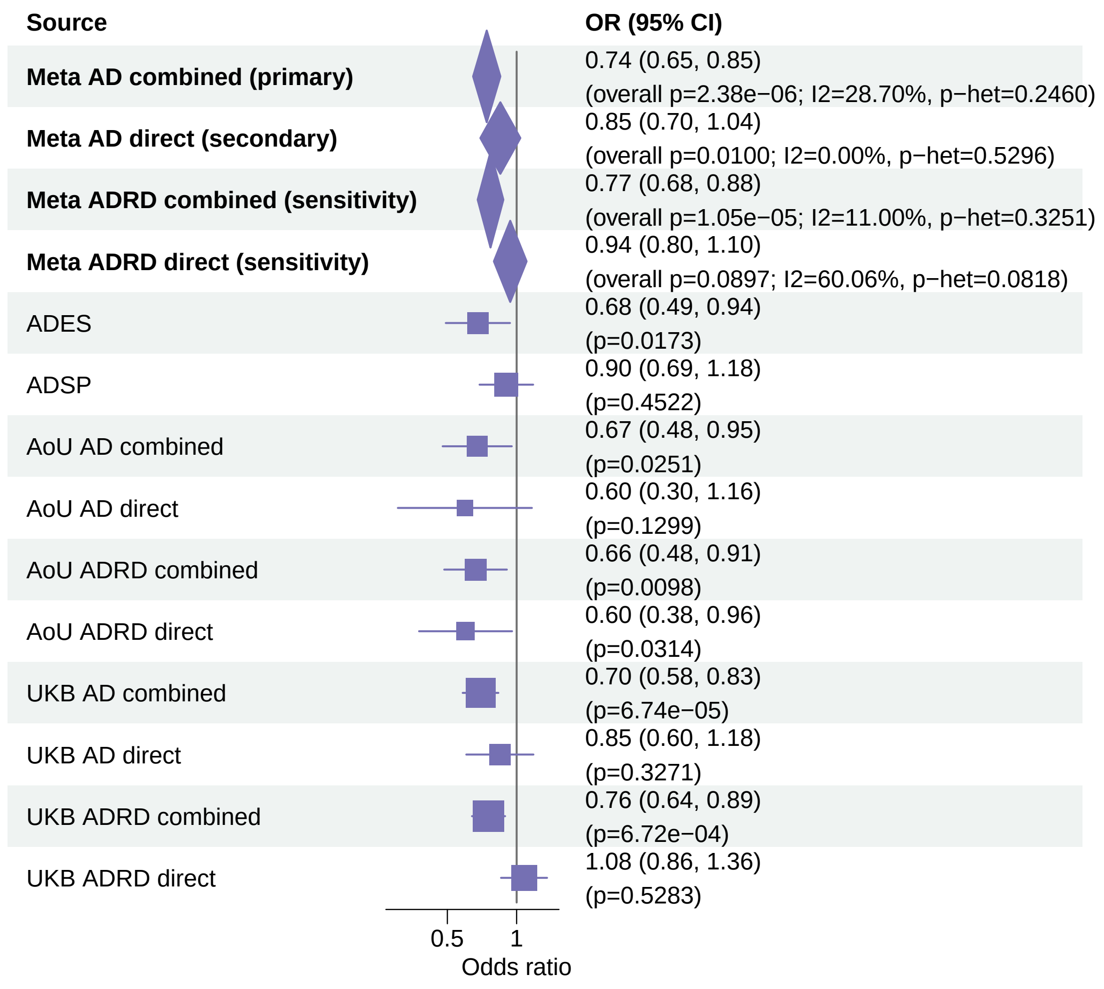


Meta OR/CI values are pooled REGENIE effects. Meta overall P values use ADES+REGENIE signed−Z evidence when the exact ADES mask was available; otherwise they use REGENIE.

### AD primary/secondary definitions with ADRD sensitivity − CLU − LOF+REVEL50 (ALL)

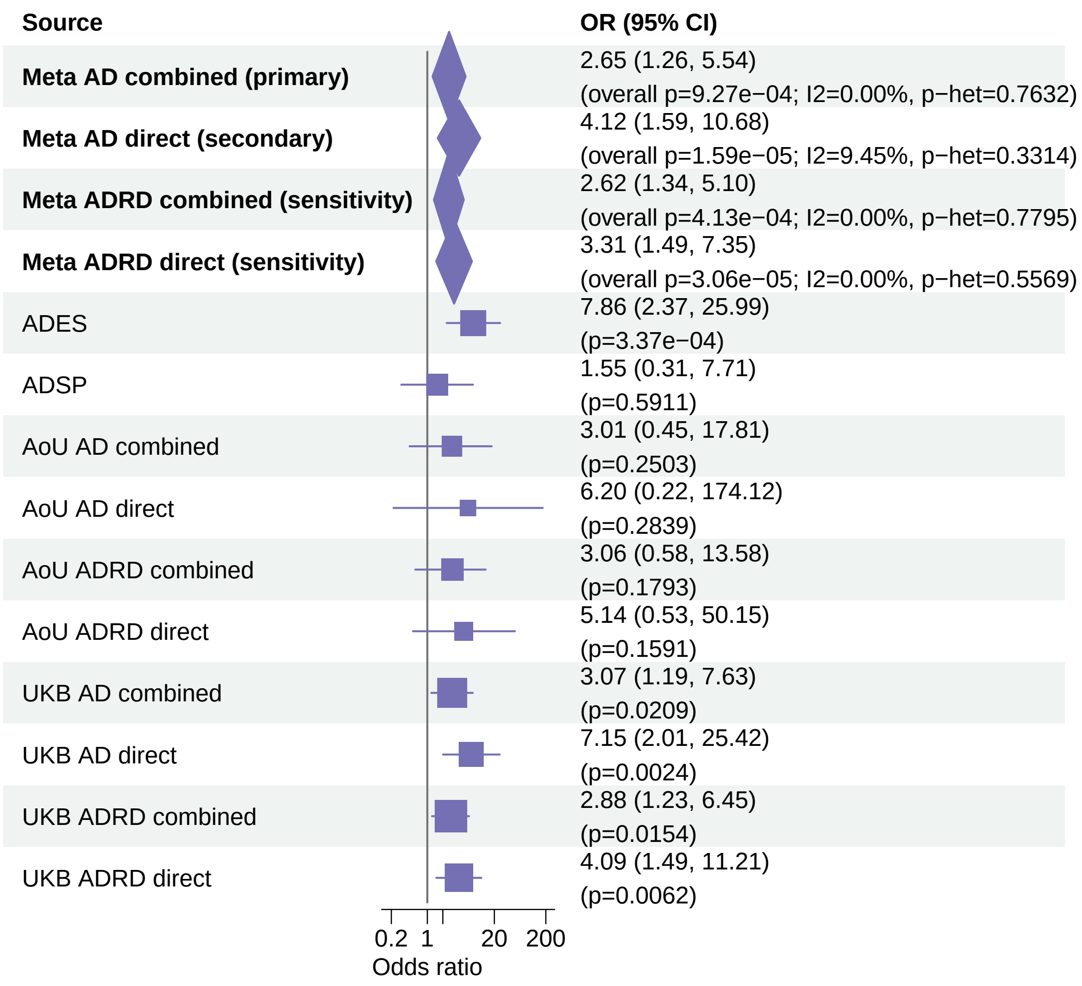


Meta OR/CI values are pooled REGENIE effects. Meta overall P values use ADES+REGENIE signed−Z evidence when the exact ADES mask was available; otherwise they use REGENIE.

#### AD primary/secondary definitions with ADRD sensitivity − ADAM10 − LOF (ALL)

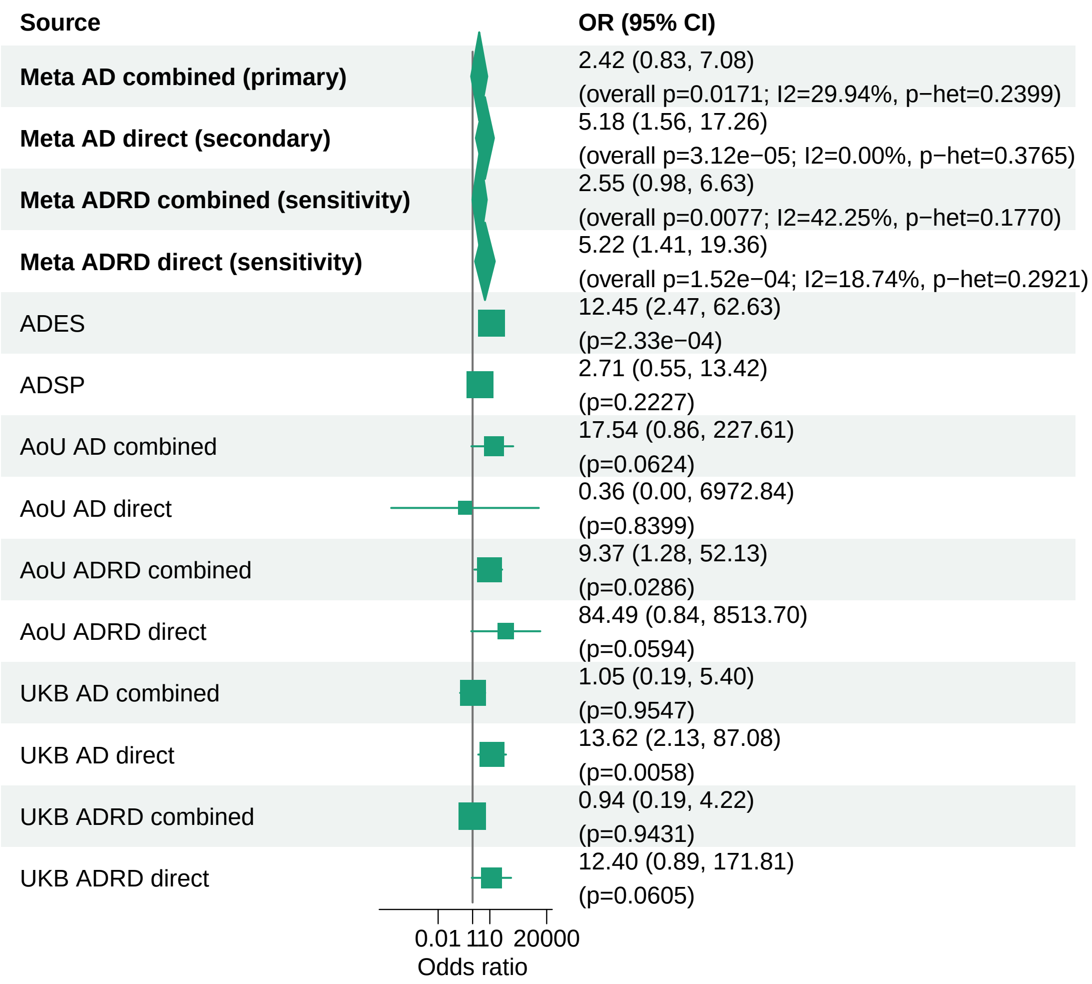


Meta OR/CI values are pooled REGENIE effects. Meta overall P values use ADES+REGENIE signed−Z evidence when the exact ADES mask was available; otherwise they use REGENIE.

#### AD primary/secondary definitions with ADRD sensitivity − RIN3 − LOF+REVEL25 (ALL)

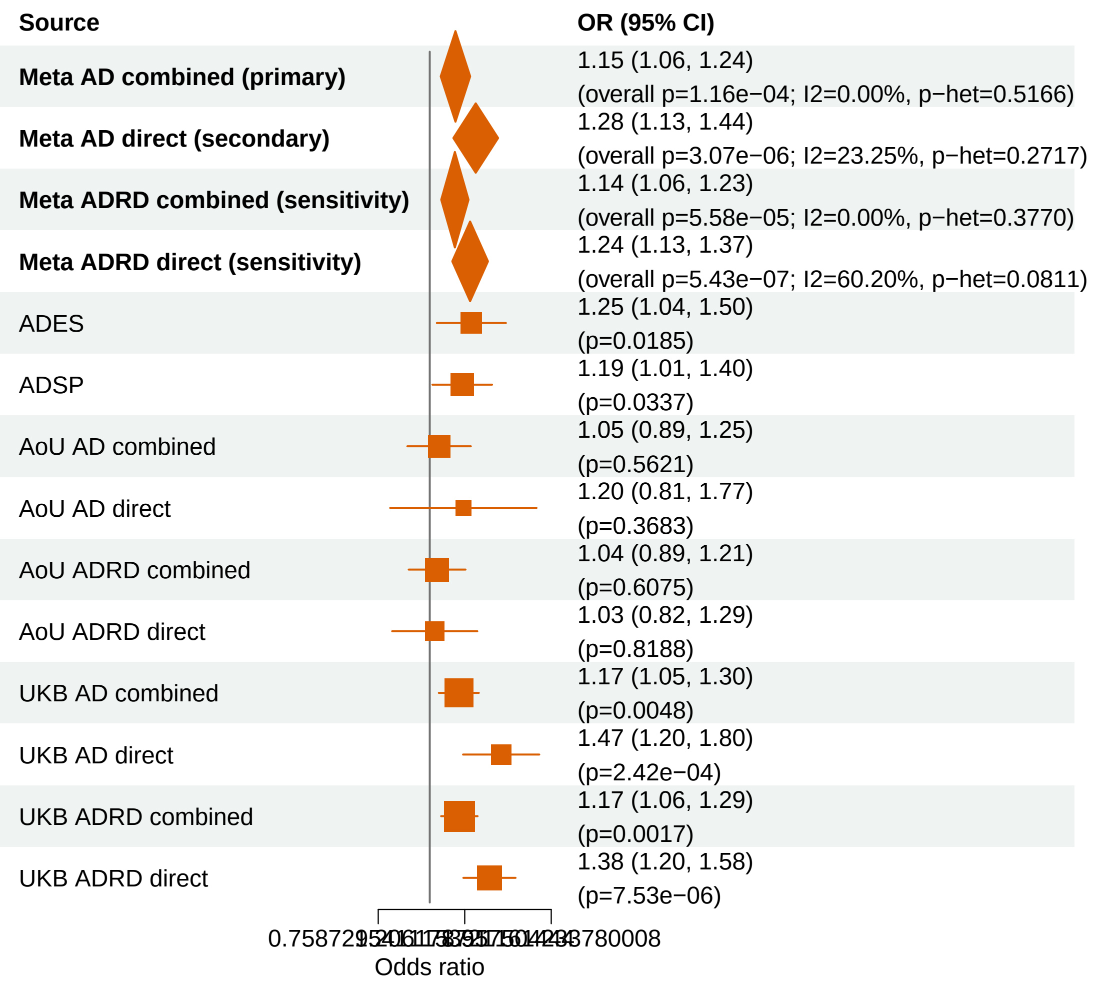


Meta OR/CI values are pooled REGENIE effects. Meta overall P values use ADES+REGENIE signed−Z evidence when the exact ADES mask was available; otherwise they use REGENIE.

#### AD primary/secondary definitions with ADRD sensitivity − SP140L − LOF+REVEL75 (ALL)

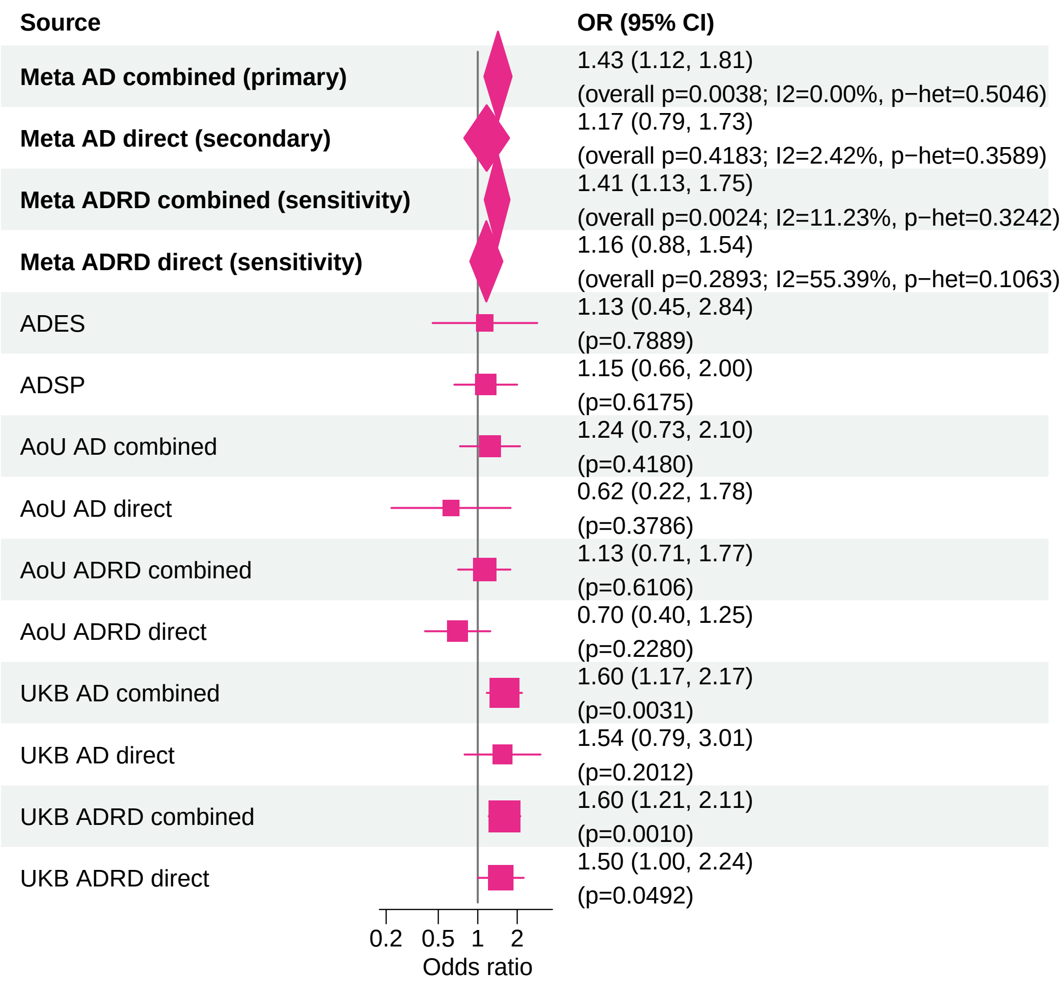


Meta OR/CI values are pooled REGENIE effects. Meta overall P values use ADES+REGENIE signed−Z evidence when the exact ADES mask was available; otherwise they use REGENIE.

#### AD primary/secondary definitions with ADRD sensitivity − PMM2 − LOF+REVEL25 (ALL)

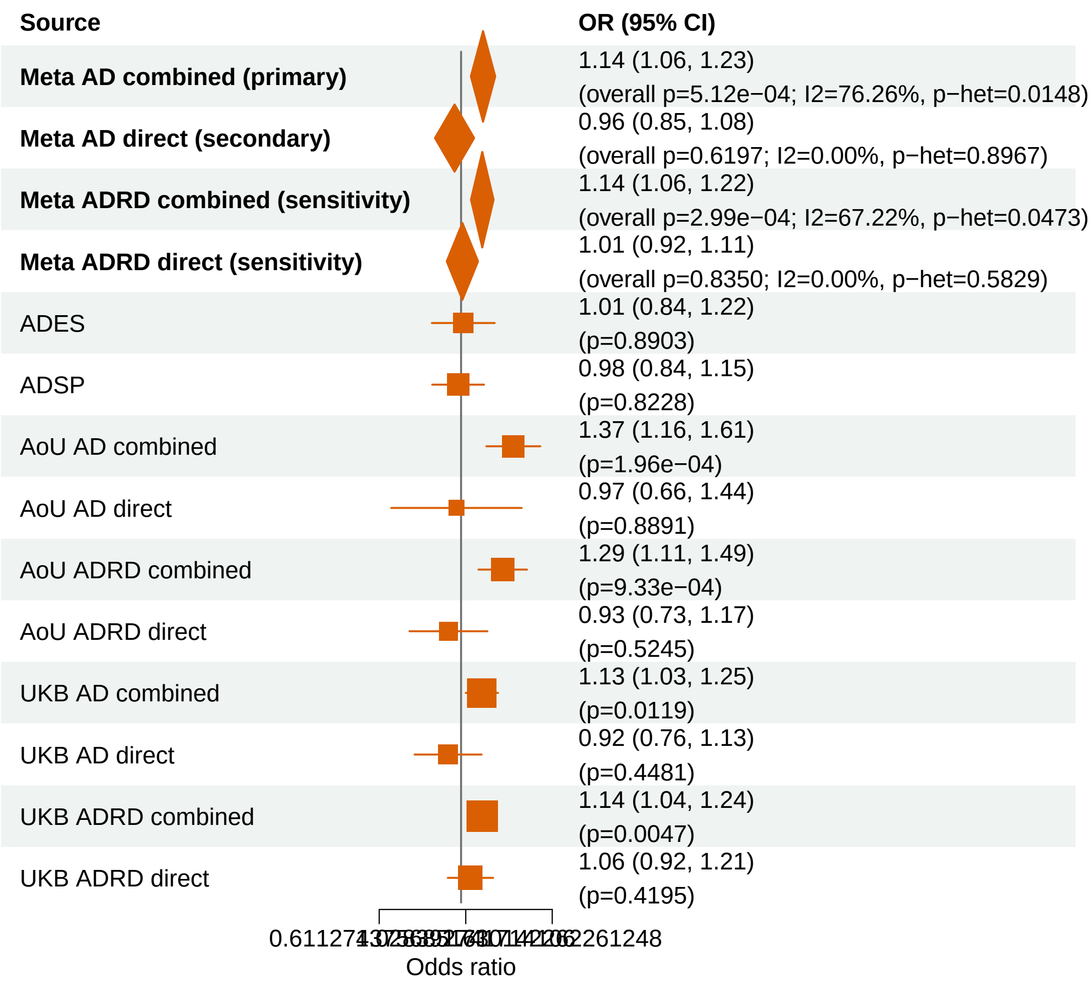


Meta OR/CI values are pooled REGENIE effects. Meta overall P values use ADES+REGENIE signed−Z evidence when the exact ADES mask was available; otherwise they use REGENIE.

### AD primary/secondary definitions with ADRD sensitivity − TBK1 − LOF+REVEL25 (ALL)

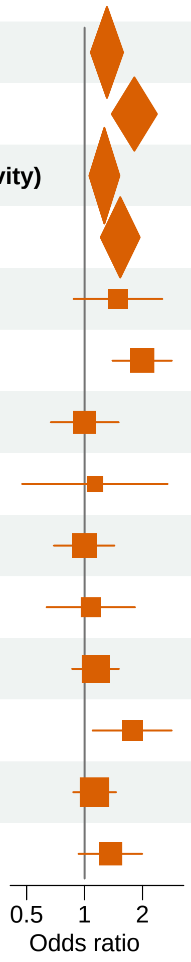


| Source | OR (95% CI) |
| --- | --- |
| **Meta AD combined (primary)** | 1.31 (1.08, 1.59) (overall p=0.0029; I2=74.87%, p−het=0.0187) |
| **Meta AD direct (secondary)** | 1.82 (1.39, 2.38) (overall p=7.40e−06; I2=0.00%, p−het=0.4932) |
| **Meta ADRD combined (sensitivity)** | 1.27 (1.06, 1.52) (overall p=0.0045; I2=77.01%, p−het=0.0129) |
| **Meta ADRD direct (sensitivity)** | 1.53 (1.21, 1.93) (overall p=1.05e−04; I2=52.27%, p−het=0.1231) |
| ADES | 1.49 (0.88, 2.53) (p=0.1431) |
| ADSP | 1.99 (1.40, 2.84) (p=1.54e−04) |
| AoU AD combined | 1.00 (0.67, 1.50) (p=0.9839) |
| AoU AD direct | 1.13 (0.47, 2.70) (p=0.7810) |
| AoU ADRD combined | 1.00 (0.69, 1.43) (p=0.9935) |
| AoU ADRD direct | 1.08 (0.64, 1.83) (p=0.7819) |
| UKB AD combined | 1.14 (0.86, 1.51) (p=0.3526) |
| UKB AD direct | 1.77 (1.10, 2.84) (p=0.0182) |
| UKB ADRD combined | 1.13 (0.87, 1.46) (p=0.3503) |
| UKB ADRD direct | 1.36 (0.93, 1.99) (p=0.1119) |

Meta OR/CI values are pooled REGENIE effects. Meta overall P values use ADES+REGENIE signed−Z evidence when the exact ADES mask was available; otherwise they use REGENIE.

#### AD primary/secondary definitions with ADRD sensitivity − ABCA1 − LOF+REVEL50 (ALL)

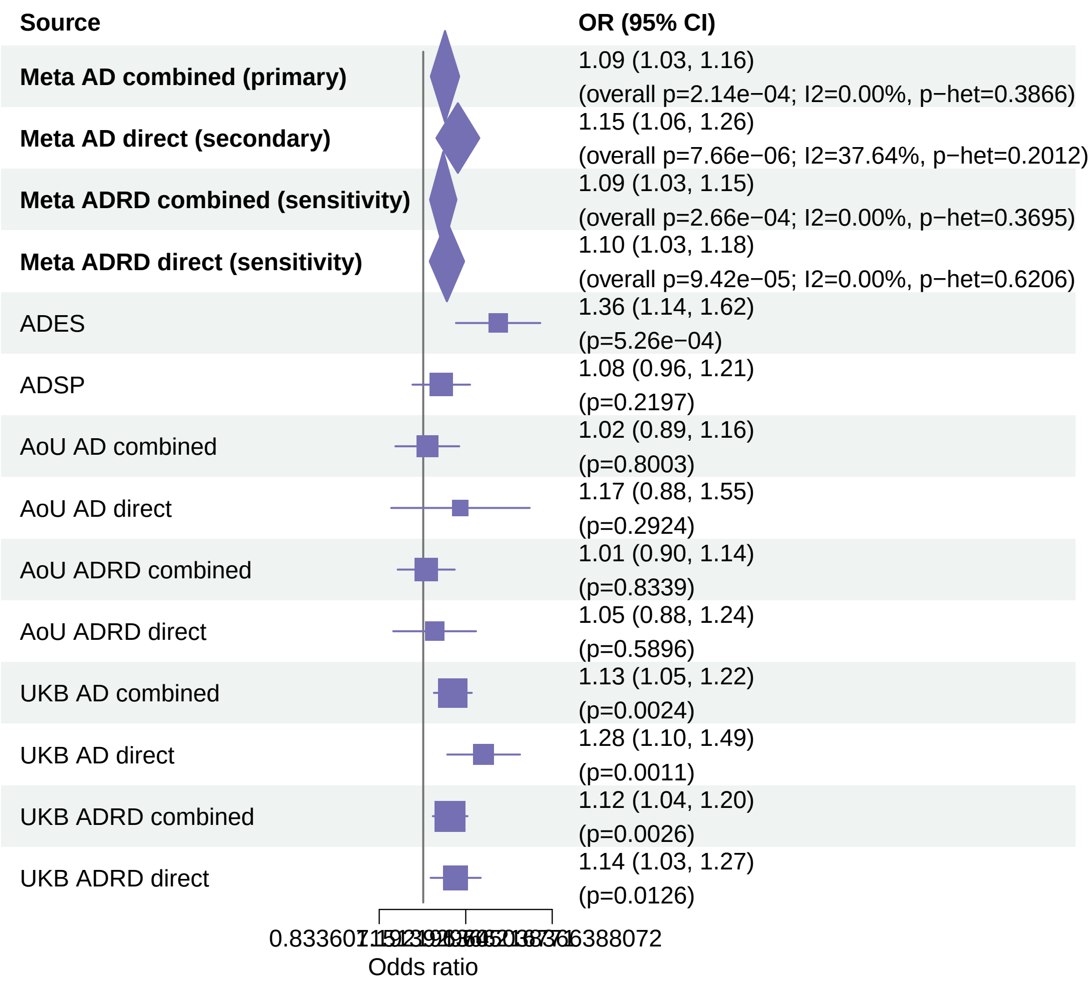


Meta OR/CI values are pooled REGENIE effects. Meta overall P values use ADES+REGENIE signed−Z evidence when the exact ADES mask was available; otherwise they use REGENIE.

### AD primary/secondary definitions with ADRD sensitivity − RBM12 − LOF (ALL)

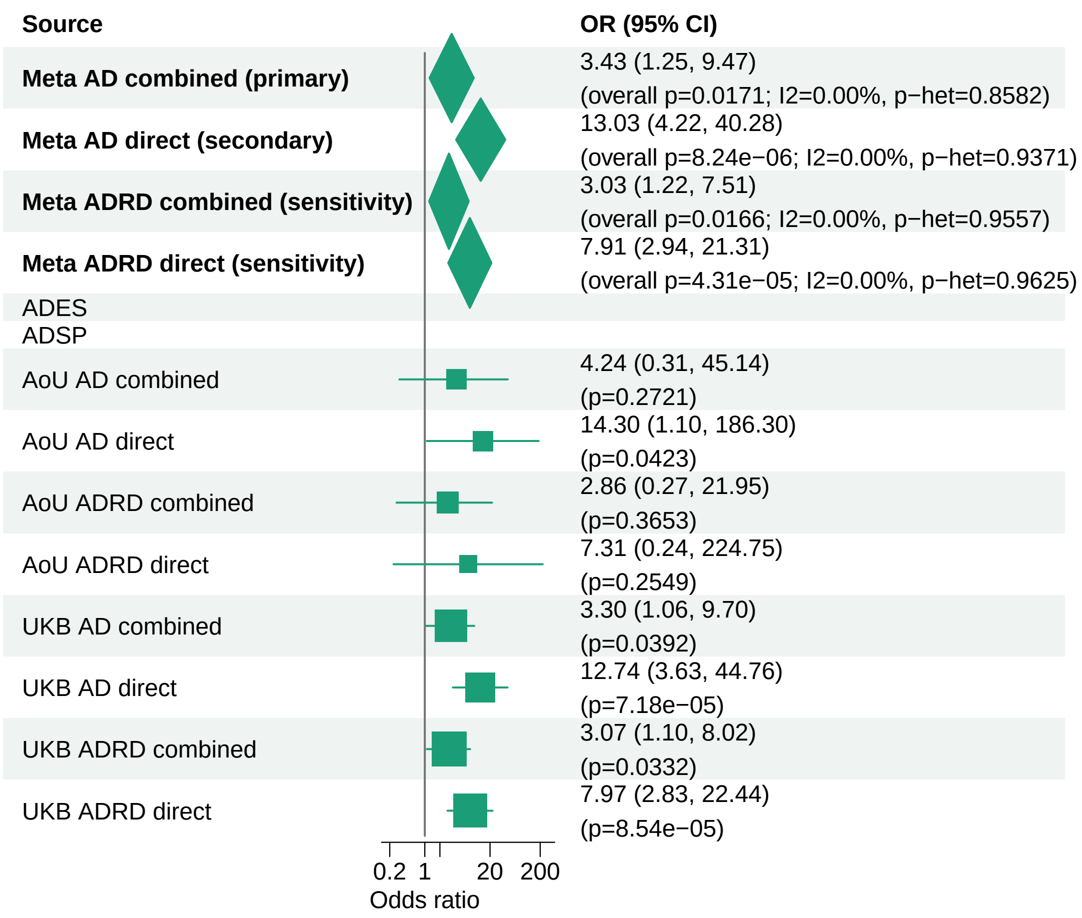


Meta OR/CI values are pooled REGENIE effects. Meta overall P values use ADES+REGENIE signed−Z evidence when the exact ADES mask was available; otherwise they use REGENIE.

#### AD primary/secondary definitions with ADRD sensitivity − ZNF256 − LOF+REVEL25 (ALL)

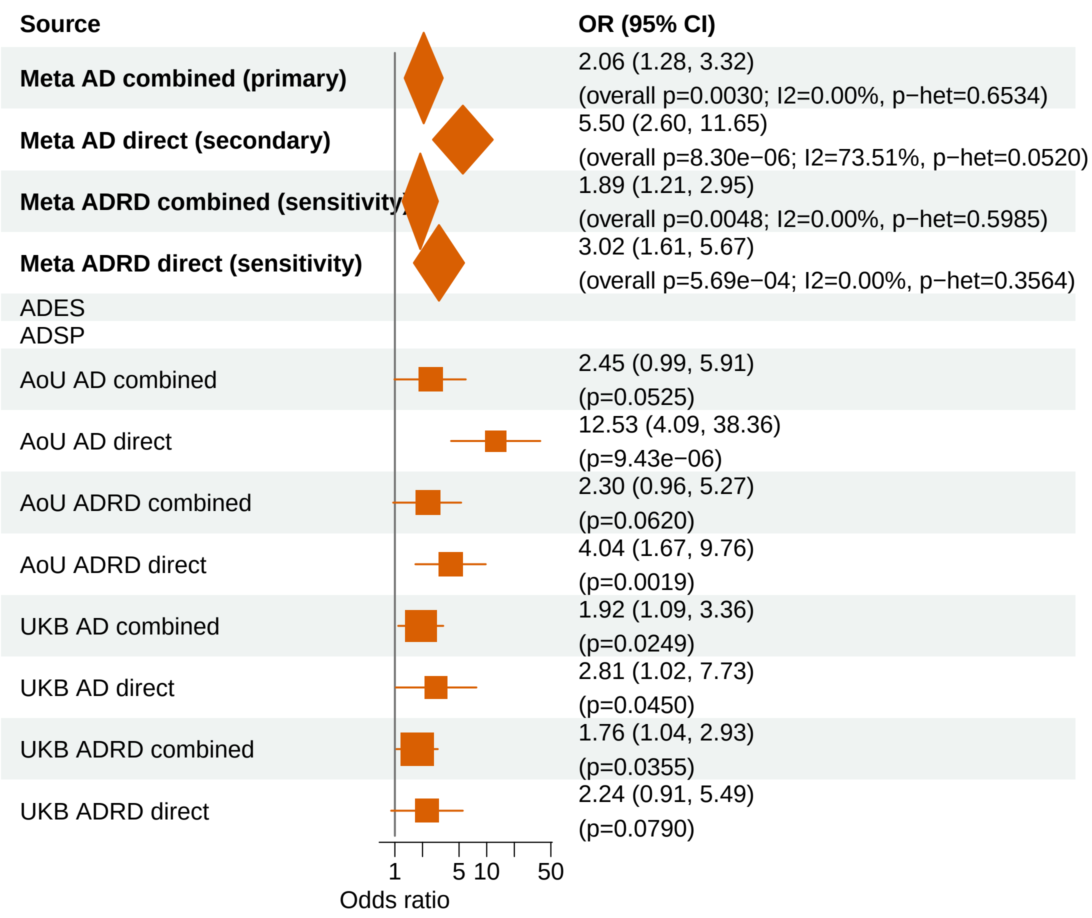


Meta OR/CI values are pooled REGENIE effects. Meta overall P values use ADES+REGENIE signed−Z evidence when the exact ADES mask was available; otherwise they use REGENIE.

#### AD primary/secondary definitions with ADRD sensitivity − RBM12 − LOF+REVEL75 (ALL)

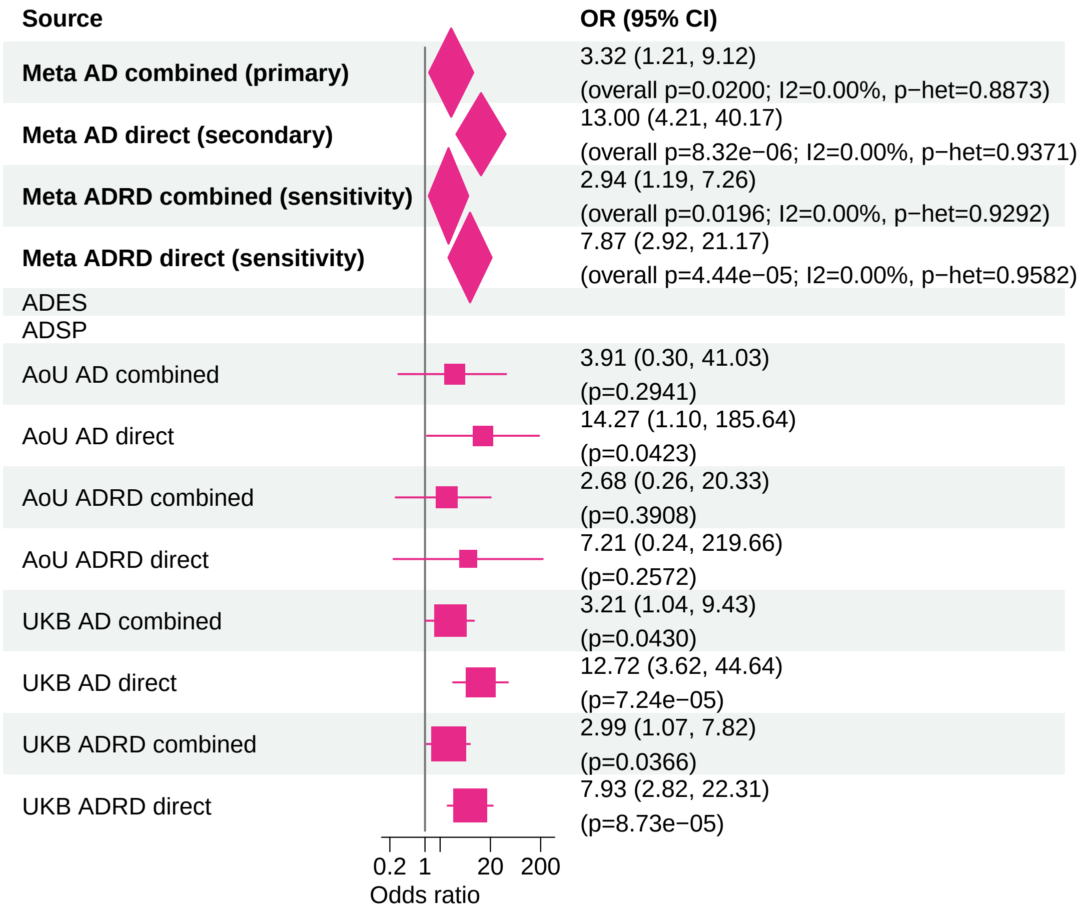


Meta OR/CI values are pooled REGENIE effects. Meta overall P values use ADES+REGENIE signed−Z evidence when the exact ADES mask was available; otherwise they use REGENIE.

#### AD primary/secondary definitions with ADRD sensitivity − CCL7 − LOF (ALL)

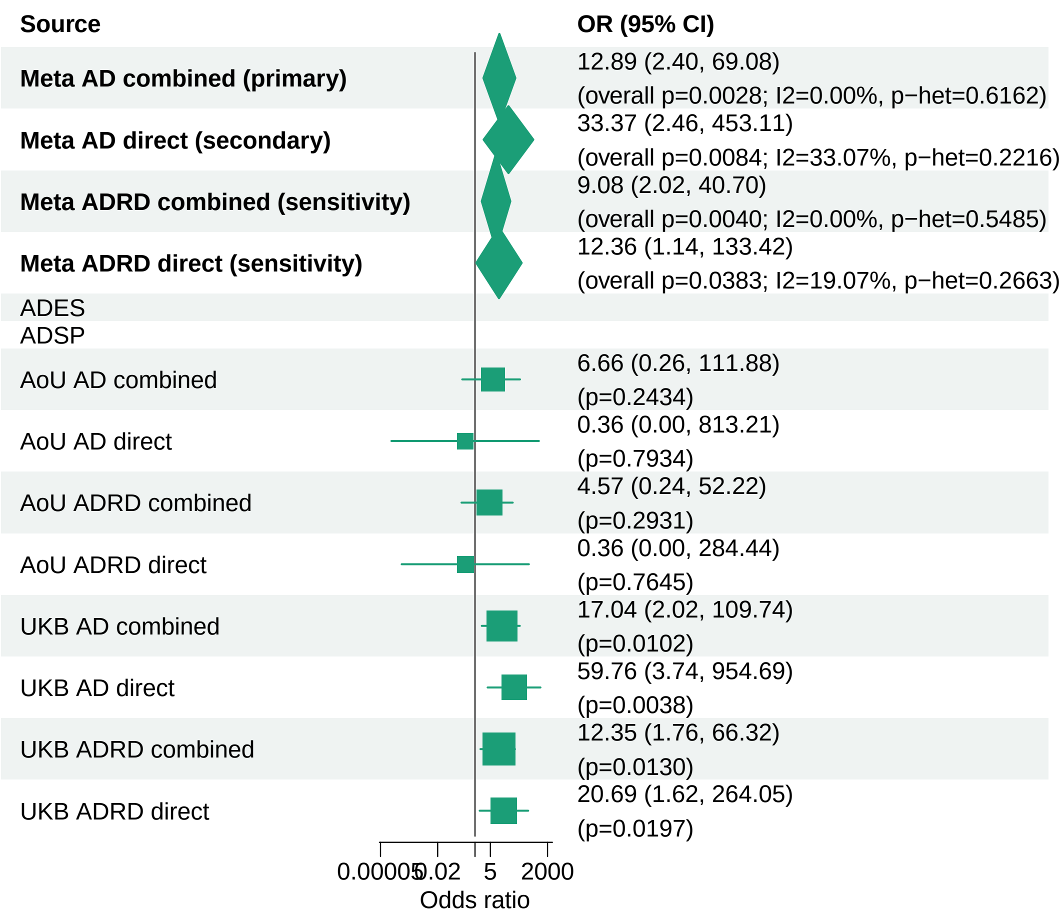


Meta OR/CI values are pooled REGENIE effects. Meta overall P values use ADES+REGENIE signed−Z evidence when the exact ADES mask was available; otherwise they use REGENIE.

#### AD primary/secondary definitions with ADRD sensitivity − MME − LOF+REVEL75 (ALL)

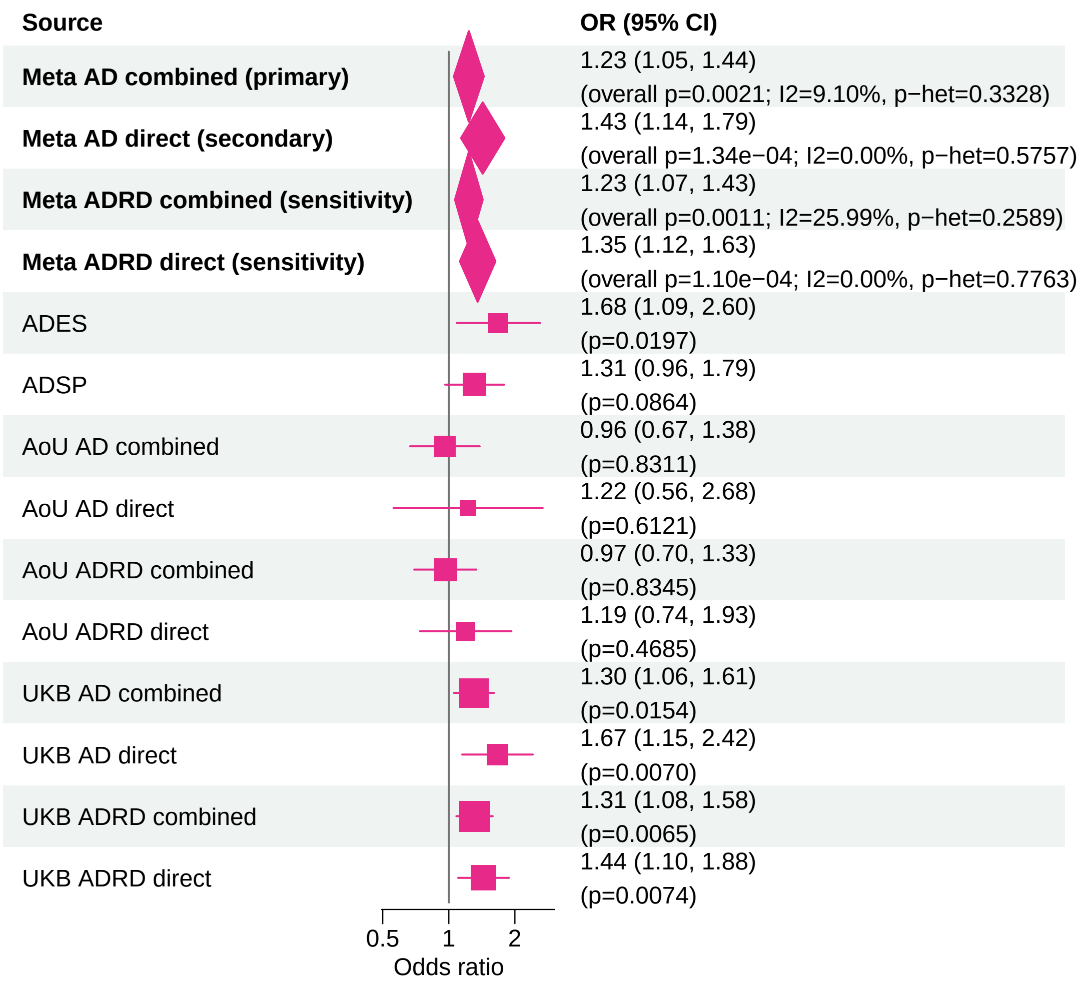


Meta OR/CI values are pooled REGENIE effects. Meta overall P values use ADES+REGENIE signed−Z evidence when the exact ADES mask was available; otherwise they use REGENIE.

#### AD primary/secondary definitions with ADRD sensitivity − GRN − LOF+REVEL25 (ALL)

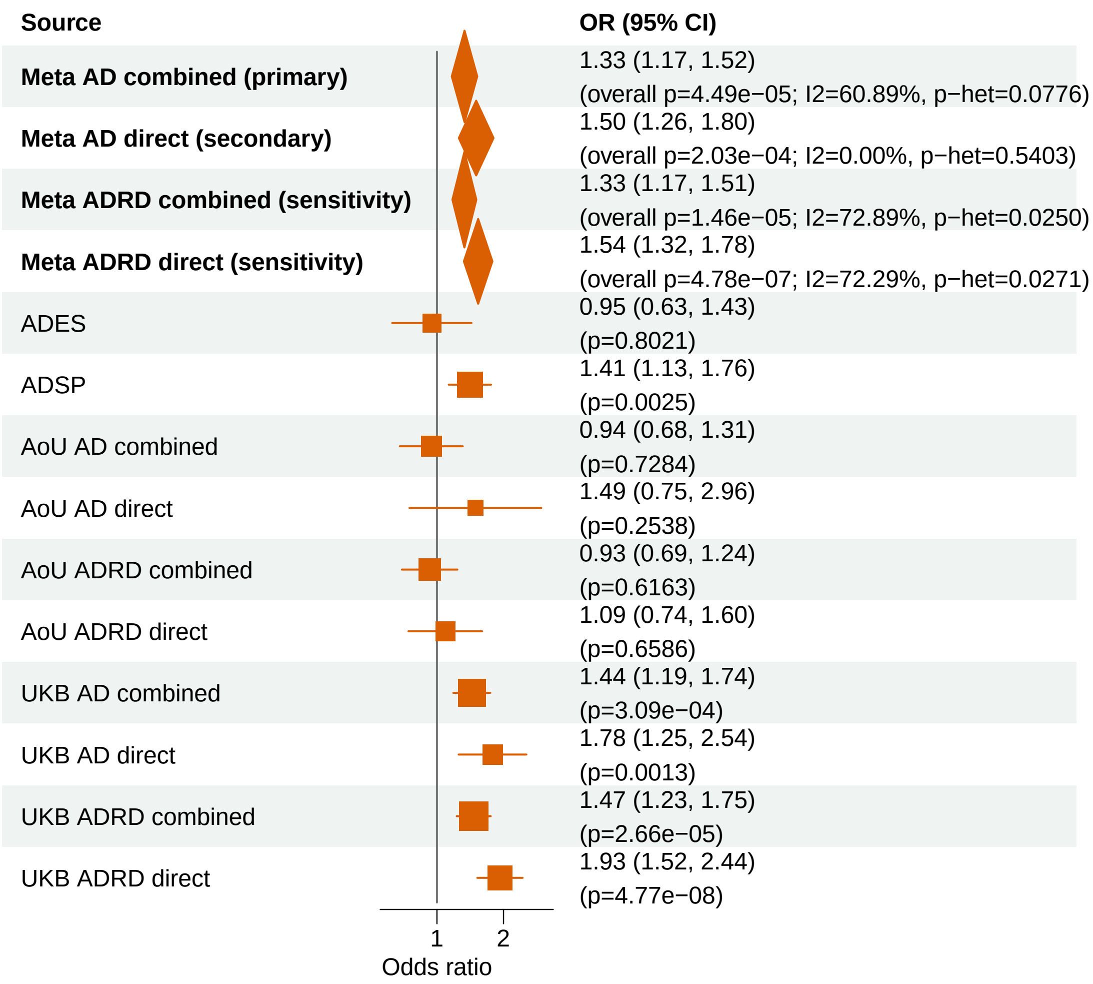


Meta OR/CI values are pooled REGENIE effects. Meta overall P values use ADES+REGENIE signed−Z evidence when the exact ADES mask was available; otherwise they use REGENIE.

### AD primary/secondary definitions with ADRD sensitivity − ABCA1 − LOF+REVEL25 (ALL)

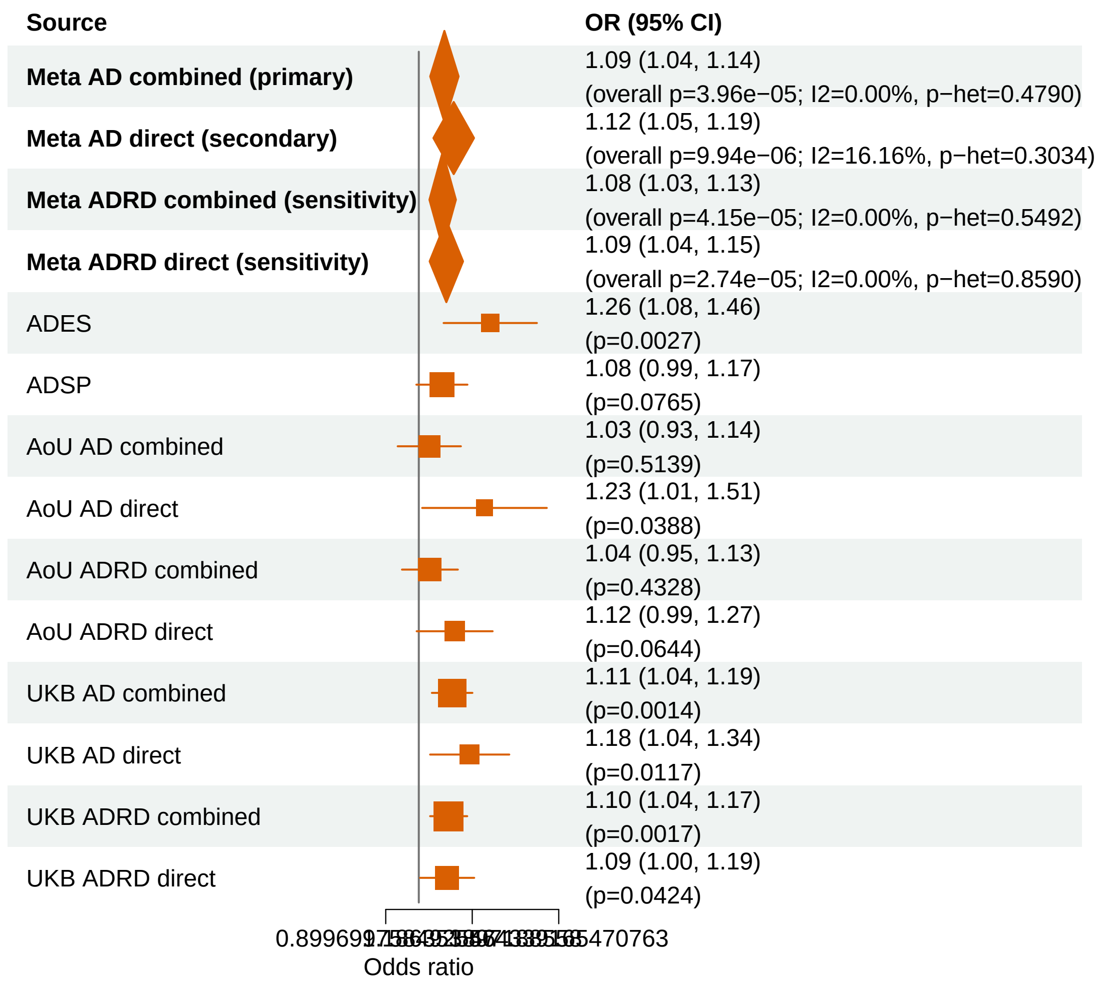


Meta OR/CI values are pooled REGENIE effects. Meta overall P values use ADES+REGENIE signed−Z evidence when the exact ADES mask was available; otherwise they use REGENIE.
