## Supplementary_Fig15 for "Population-scale burden analysis of rare damaging coding variants identifies novel risk genes for Alzheimer’s disease and related dementias and Parkinson’s disease and related disorders"

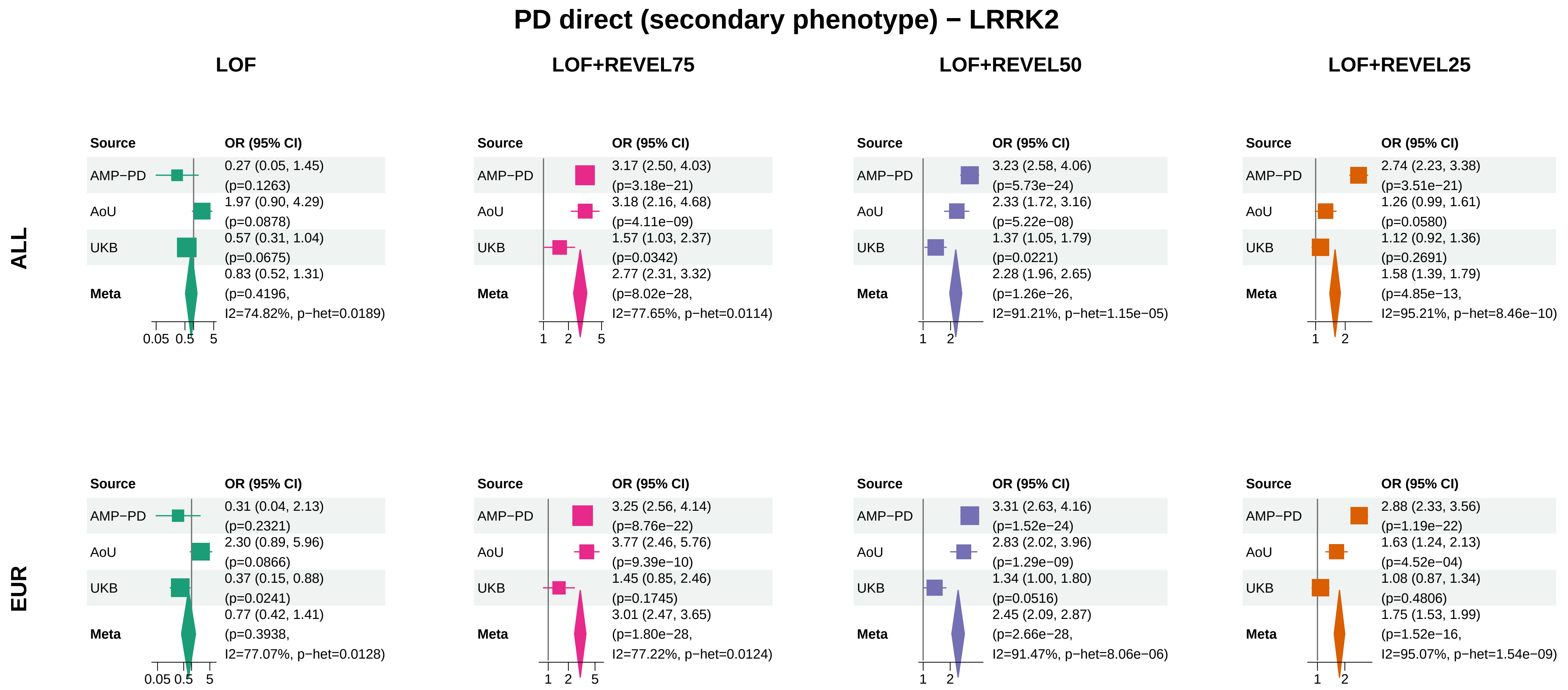


Meta OR/CI and P: exact−kappa=0.5 pooled REGENIE fixed−effects analysis.

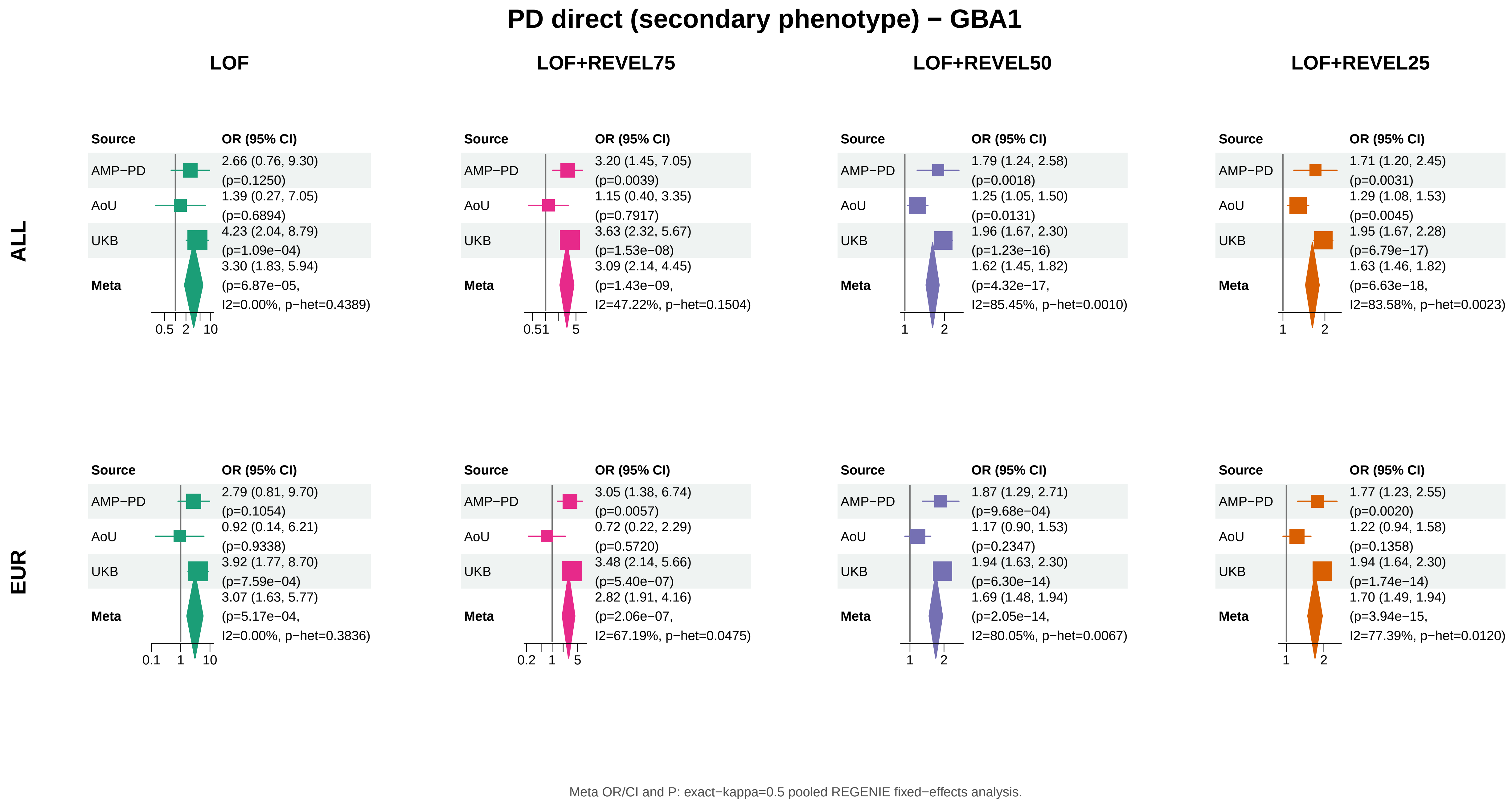

PD direct (secondary phenotype) − GBA1
LOF
LOF+REVEL75
LOF+REVEL50
LOF+REVEL25
ALL
EUR
Source
OR (95% CI)
AMP−PD
AoU
UKB
Meta
ALL – LOF: AMP−PD 2.66 (0.76, 9.30) (p=0.1250); AoU 1.39 (0.27, 7.05) (p=0.6894); UKB 4.23 (2.04, 8.79) (p=1.09e−04); Meta 3.30 (1.83, 5.94) (p=6.87e−05, I2=0.00%, p−het=0.4389)
0.5 2 10
ALL – LOF+REVEL75: AMP−PD 3.20 (1.45, 7.05) (p=0.0039); AoU 1.15 (0.40, 3.35) (p=0.7917); UKB 3.63 (2.32, 5.67) (p=1.53e−08); Meta 3.09 (2.14, 4.45) (p=1.43e−09, I2=47.22%, p−het=0.1504)
0.51 5
ALL – LOF+REVEL50: AMP−PD 1.79 (1.24, 2.58) (p=0.0018); AoU 1.25 (1.05, 1.50) (p=0.0131); UKB 1.96 (1.67, 2.30) (p=1.23e−16); Meta 1.62 (1.45, 1.82) (p=4.32e−17, I2=85.45%, p−het=0.0010)
1 2
ALL – LOF+REVEL25: AMP−PD 1.71 (1.20, 2.45) (p=0.0031); AoU 1.29 (1.08, 1.53) (p=0.0045); UKB 1.95 (1.67, 2.28) (p=6.79e−17); Meta 1.63 (1.46, 1.82) (p=6.63e−18, I2=83.58%, p−het=0.0023)
1 2
EUR – LOF: AMP−PD 2.79 (0.81, 9.70) (p=0.1054); AoU 0.92 (0.14, 6.21) (p=0.9338); UKB 3.92 (1.77, 8.70) (p=7.59e−04); Meta 3.07 (1.63, 5.77) (p=5.17e−04, I2=0.00%, p−het=0.3836)
0.1 1 10
EUR – LOF+REVEL75: AMP−PD 3.05 (1.38, 6.74) (p=0.0057); AoU 0.72 (0.22, 2.29) (p=0.5720); UKB 3.48 (2.14, 5.66) (p=5.40e−07); Meta 2.82 (1.91, 4.16) (p=2.06e−07, I2=67.19%, p−het=0.0475)
0.2 1 5
EUR – LOF+REVEL50: AMP−PD 1.87 (1.29, 2.71) (p=9.68e−04); AoU 1.17 (0.90, 1.53) (p=0.2347); UKB 1.94 (1.63, 2.30) (p=6.30e−14); Meta 1.69 (1.48, 1.94) (p=2.05e−14, I2=80.05%, p−het=0.0067)
1 2
EUR – LOF+REVEL25: AMP−PD 1.77 (1.23, 2.55) (p=0.0020); AoU 1.22 (0.94, 1.58) (p=0.1358); UKB 1.94 (1.64, 2.30) (p=1.74e−14); Meta 1.70 (1.49, 1.94) (p=3.94e−15, I2=77.39%, p−het=0.0120)
1 2
Meta OR/CI and P: exact−kappa=0.5 pooled REGENIE fixed−effects analysis.

### PD direct (secondary phenotype) − ANKRD27

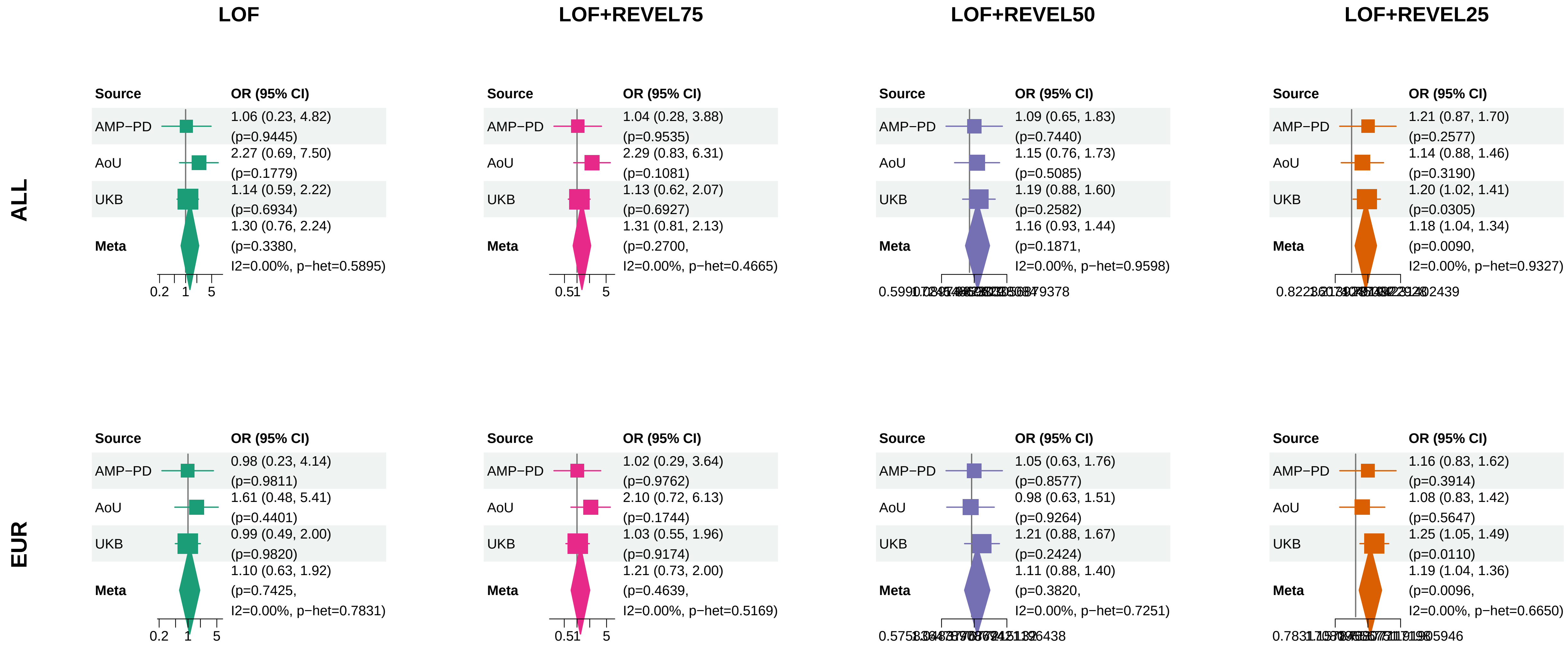


Meta OR/CI and P: exact−kappa=0.5 pooled REGENIE fixed−effects analysis.

### PD direct (secondary phenotype) − CCL7

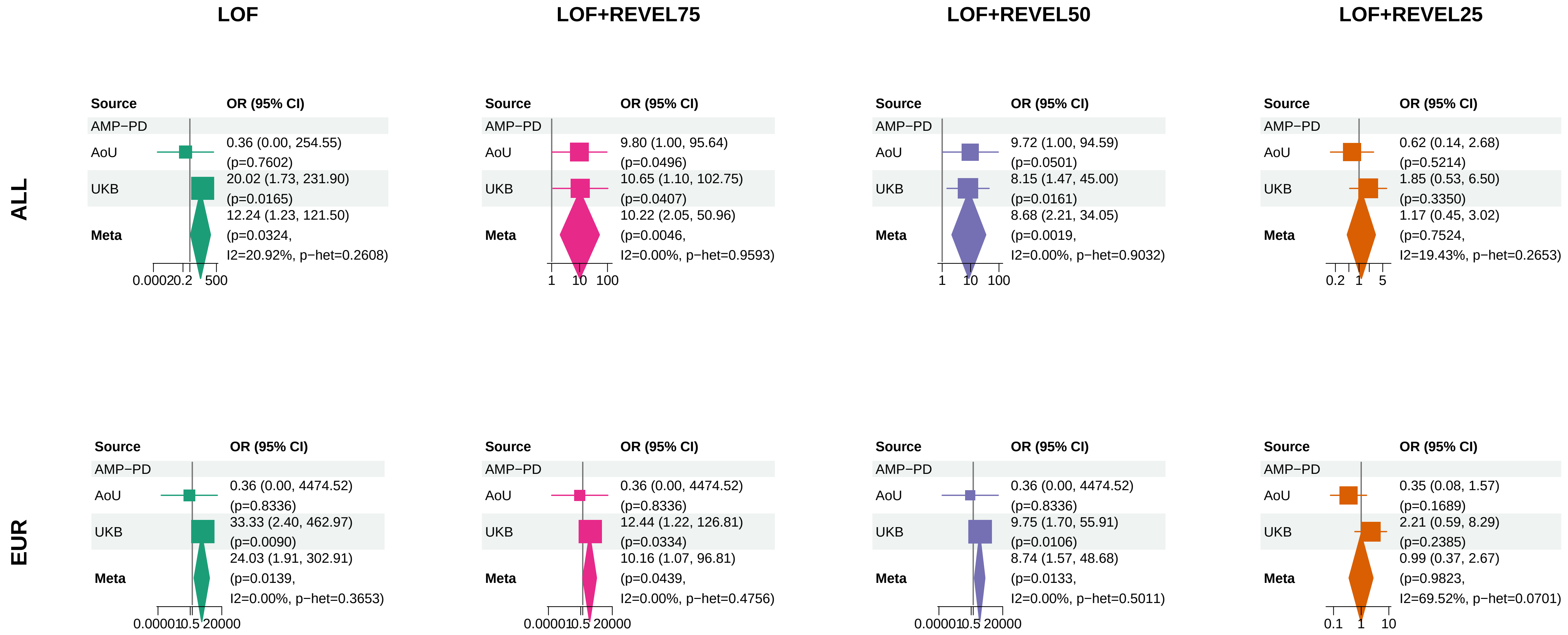


Meta OR/CI and P: exact−kappa=0.5 pooled REGENIE fixed−effects analysis.

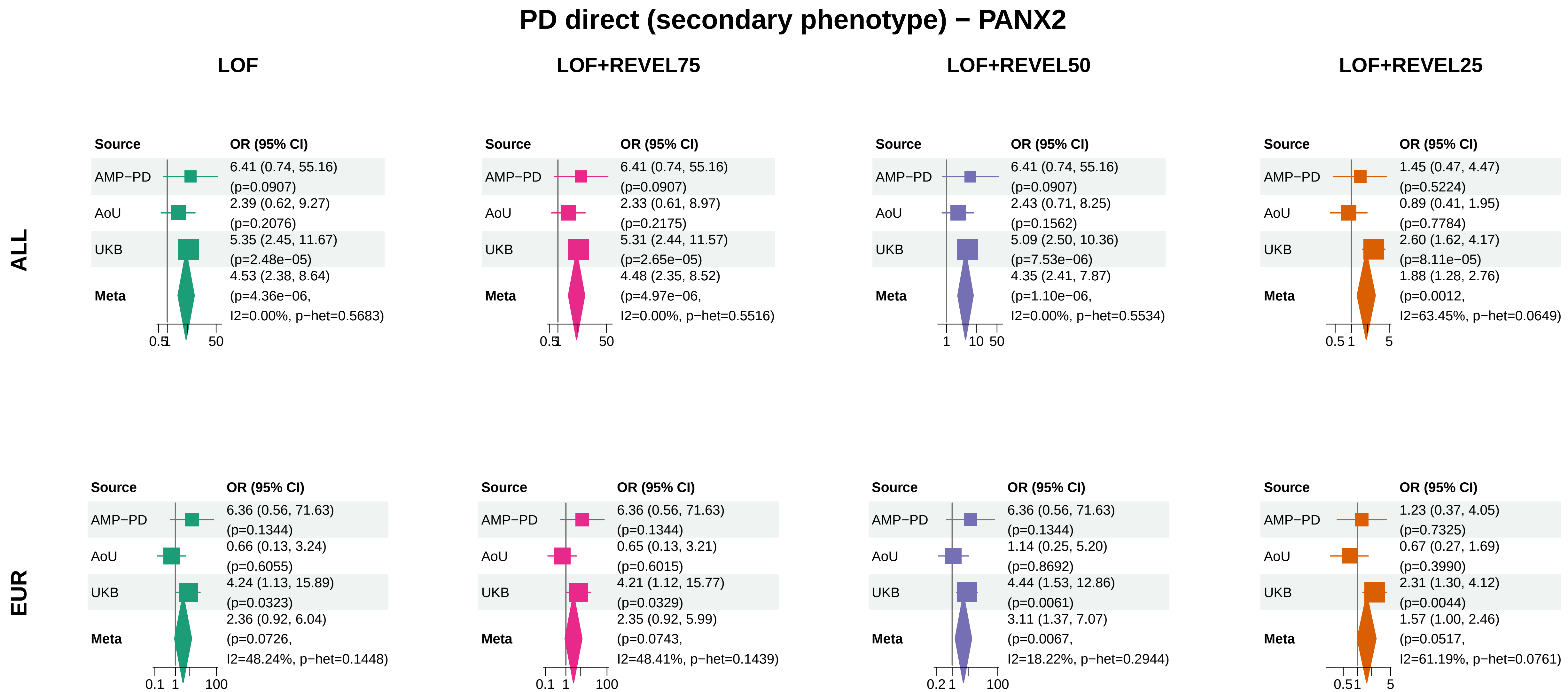


Meta OR/CI and P: exact−kappa=0.5 pooled REGENIE fixed−effects analysis.

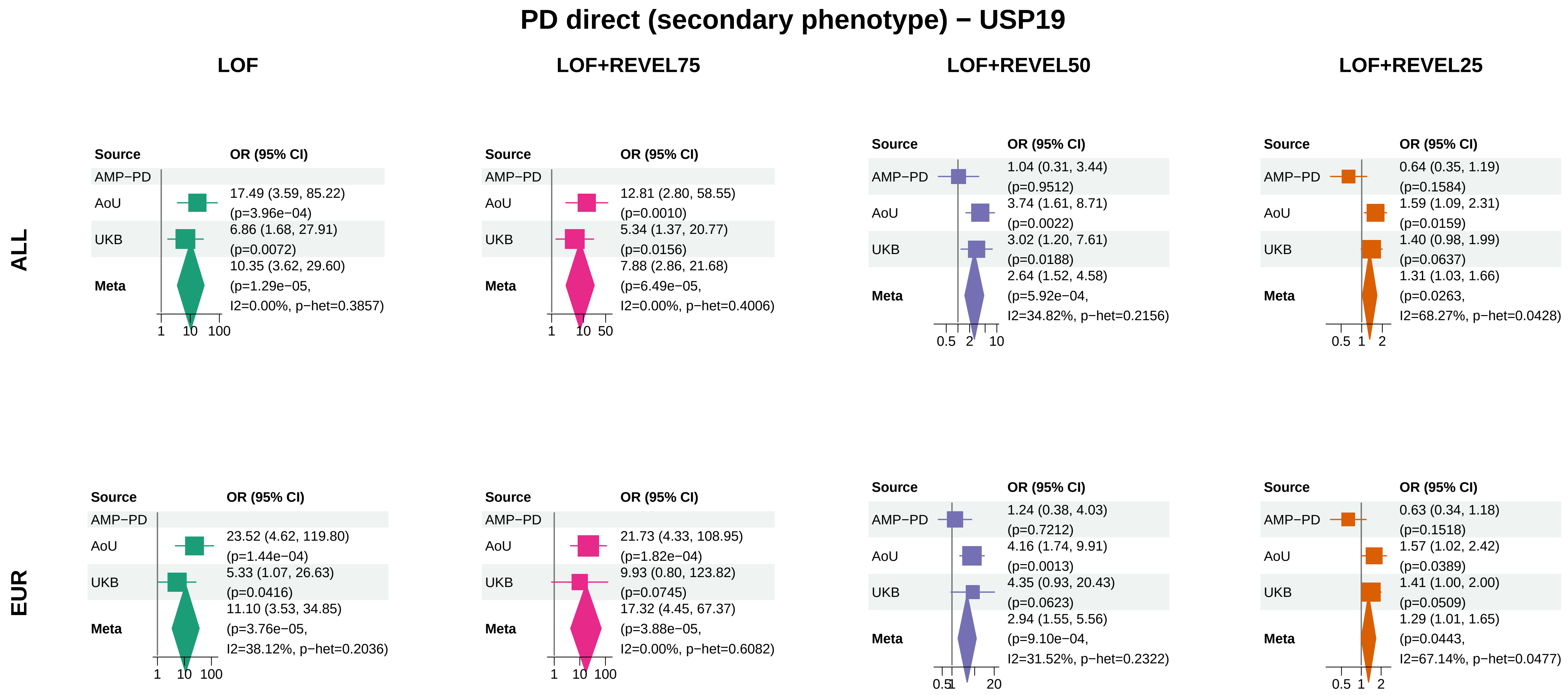

Meta OR/CI and P: exact−kappa=0.5 pooled REGENIE fixed−effects analysis.

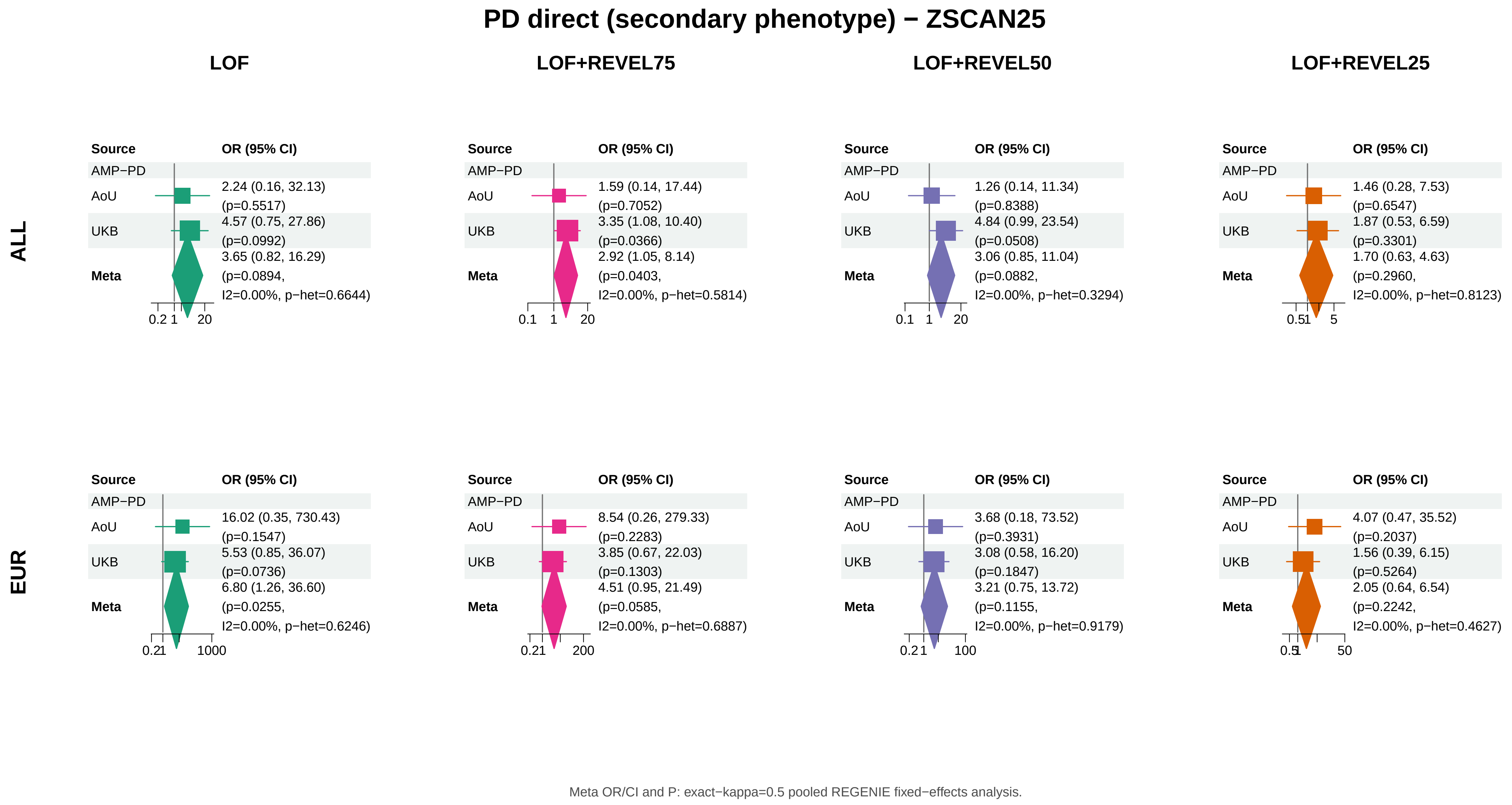
PD direct (secondary phenotype) − ZSCAN25
LOF
LOF+REVEL75
LOF+REVEL50
LOF+REVEL25
ALL
EUR
Source
OR (95% CI)
AMP−PD
AoU
UKB
Meta
ALL, LOF: AoU 2.24 (0.16, 32.13) (p=0.5517); UKB 4.57 (0.75, 27.86) (p=0.0992); Meta 3.65 (0.82, 16.29) (p=0.0894, I2=0.00%, p−het=0.6644)
0.2 1 20
ALL, LOF+REVEL75: AoU 1.59 (0.14, 17.44) (p=0.7052); UKB 3.35 (1.08, 10.40) (p=0.0366); Meta 2.92 (1.05, 8.14) (p=0.0403, I2=0.00%, p−het=0.5814)
0.1 1 20
ALL, LOF+REVEL50: AoU 1.26 (0.14, 11.34) (p=0.8388); UKB 4.84 (0.99, 23.54) (p=0.0508); Meta 3.06 (0.85, 11.04) (p=0.0882, I2=0.00%, p−het=0.3294)
0.1 1 20
ALL, LOF+REVEL25: AoU 1.46 (0.28, 7.53) (p=0.6547); UKB 1.87 (0.53, 6.59) (p=0.3301); Meta 1.70 (0.63, 4.63) (p=0.2960, I2=0.00%, p−het=0.8123)
0.51 5
EUR, LOF: AoU 16.02 (0.35, 730.43) (p=0.1547); UKB 5.53 (0.85, 36.07) (p=0.0736); Meta 6.80 (1.26, 36.60) (p=0.0255, I2=0.00%, p−het=0.6246)
0.21 1000
EUR, LOF+REVEL75: AoU 8.54 (0.26, 279.33) (p=0.2283); UKB 3.85 (0.67, 22.03) (p=0.1303); Meta 4.51 (0.95, 21.49) (p=0.0585, I2=0.00%, p−het=0.6887)
0.21 200
EUR, LOF+REVEL50: AoU 3.68 (0.18, 73.52) (p=0.3931); UKB 3.08 (0.58, 16.20) (p=0.1847); Meta 3.21 (0.75, 13.72) (p=0.1155, I2=0.00%, p−het=0.9179)
0.21 100
EUR, LOF+REVEL25: AoU 4.07 (0.47, 35.52) (p=0.2037); UKB 1.56 (0.39, 6.15) (p=0.5264); Meta 2.05 (0.64, 6.54) (p=0.2242, I2=0.00%, p−het=0.4627)
0.51 50
Meta OR/CI and P: exact−kappa=0.5 pooled REGENIE fixed−effects analysis.

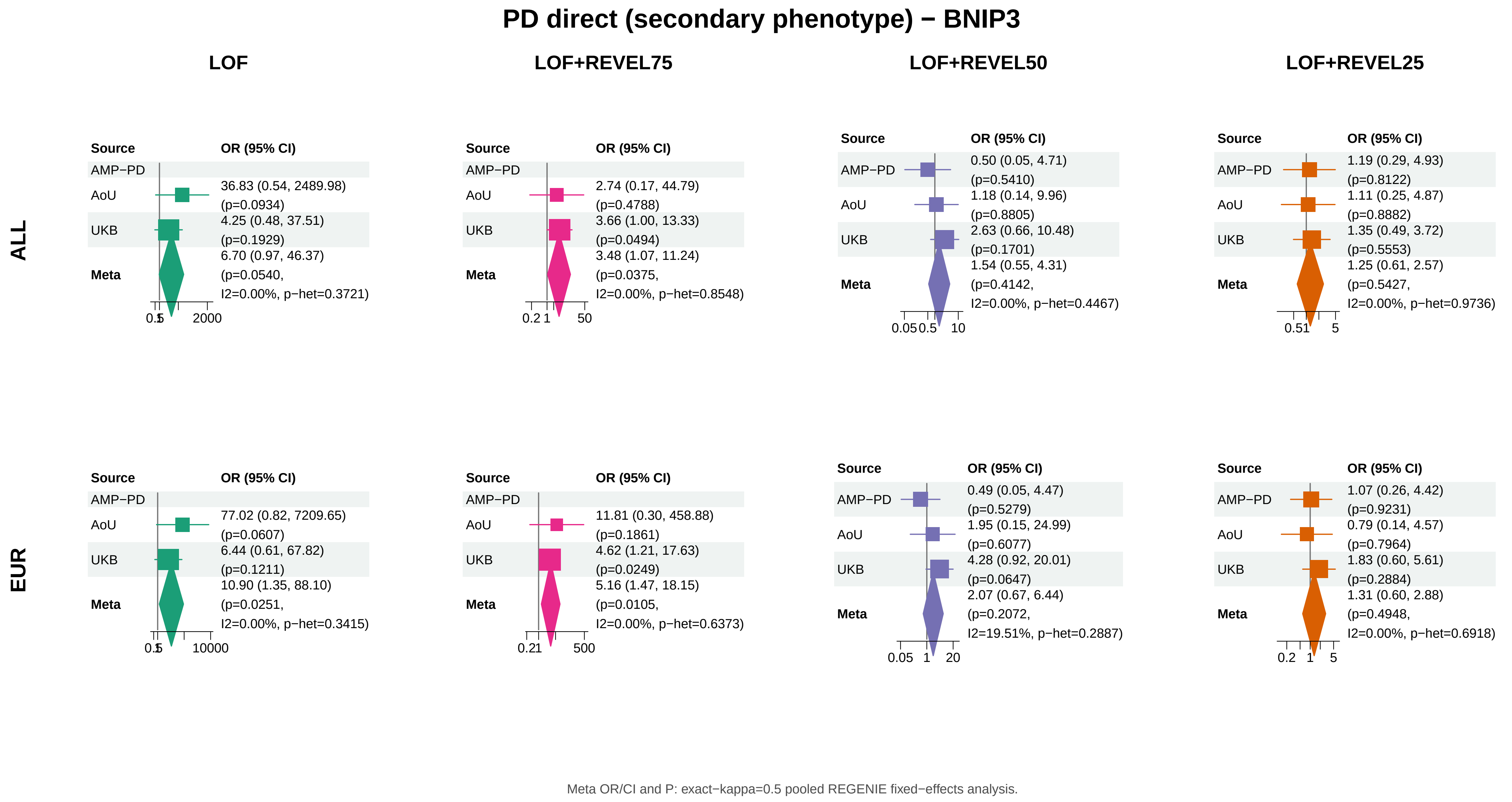
PD direct (secondary phenotype) − BNIP3
LOF
LOF+REVEL75
LOF+REVEL50
LOF+REVEL25
ALL
EUR
Source
OR (95% CI)
AMP−PD
AoU
UKB
Meta
36.83 (0.54, 2489.98) (p=0.0934)
4.25 (0.48, 37.51) (p=0.1929)
6.70 (0.97, 46.37) (p=0.0540, I2=0.00%, p−het=0.3721)
2.74 (0.17, 44.79) (p=0.4788)
3.66 (1.00, 13.33) (p=0.0494)
3.48 (1.07, 11.24) (p=0.0375, I2=0.00%, p−het=0.8548)
0.50 (0.05, 4.71) (p=0.5410)
1.18 (0.14, 9.96) (p=0.8805)
2.63 (0.66, 10.48) (p=0.1701)
1.54 (0.55, 4.31) (p=0.4142, I2=0.00%, p−het=0.4467)
1.19 (0.29, 4.93) (p=0.8122)
1.11 (0.25, 4.87) (p=0.8882)
1.35 (0.49, 3.72) (p=0.5553)
1.25 (0.61, 2.57) (p=0.5427, I2=0.00%, p−het=0.9736)
77.02 (0.82, 7209.65) (p=0.0607)
6.44 (0.61, 67.82) (p=0.1211)
10.90 (1.35, 88.10) (p=0.0251, I2=0.00%, p−het=0.3415)
11.81 (0.30, 458.88) (p=0.1861)
4.62 (1.21, 17.63) (p=0.0249)
5.16 (1.47, 18.15) (p=0.0105, I2=0.00%, p−het=0.6373)
0.49 (0.05, 4.47) (p=0.5279)
1.95 (0.15, 24.99) (p=0.6077)
4.28 (0.92, 20.01) (p=0.0647)
2.07 (0.67, 6.44) (p=0.2072, I2=19.51%, p−het=0.2887)
1.07 (0.26, 4.42) (p=0.9231)
0.79 (0.14, 4.57) (p=0.7964)
1.83 (0.60, 5.61) (p=0.2884)
1.31 (0.60, 2.88) (p=0.4948, I2=0.00%, p−het=0.6918)
Meta OR/CI and P: exact−kappa=0.5 pooled REGENIE fixed−effects analysis.

#### PD direct (secondary phenotype) − SOBP

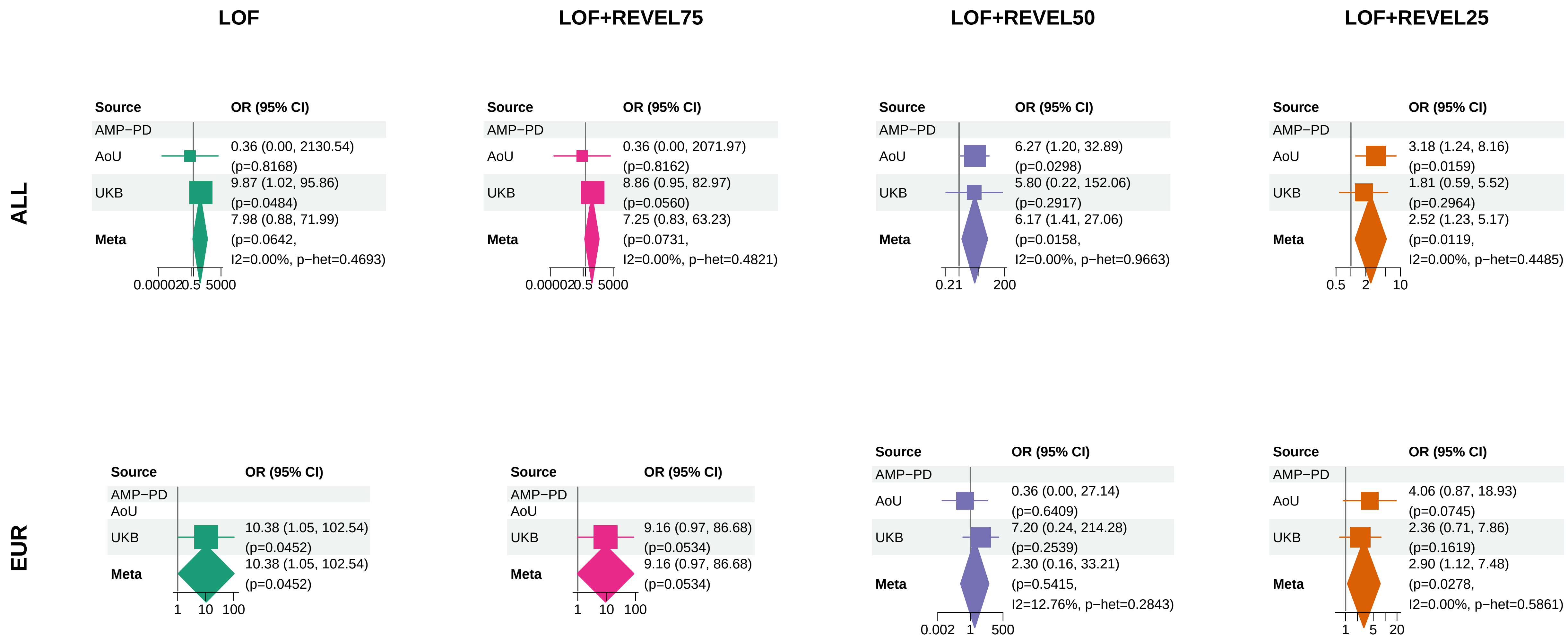


Meta OR/CI and P: exact−kappa=0.5 pooled REGENIE fixed−effects analysis.

### PD direct (secondary phenotype) − KANSL3

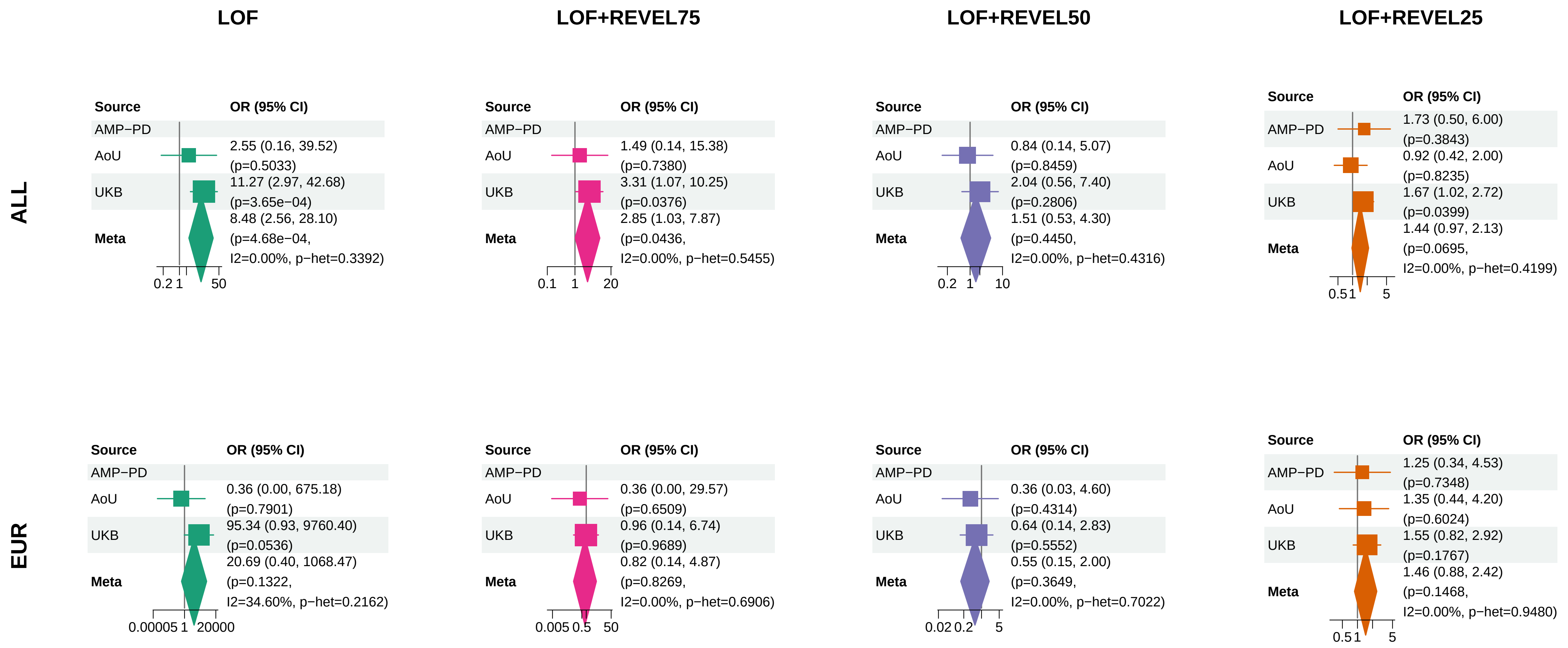


Meta OR/CI and P: exact−kappa=0.5 pooled REGENIE fixed−effects analysis.

#### PD direct (secondary phenotype) − KRBA2

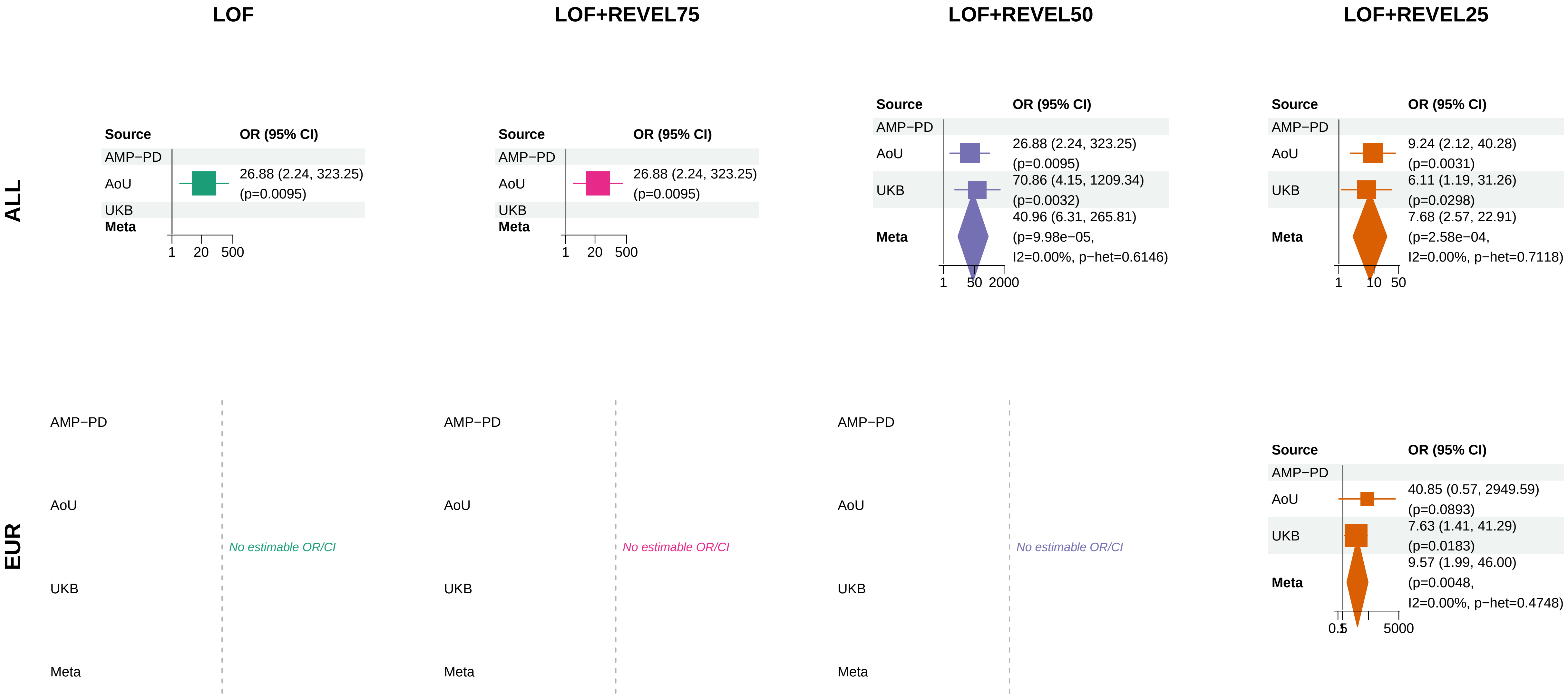


Meta OR/CI and P: exact−kappa=0.5 pooled REGENIE fixed−effects analysis.

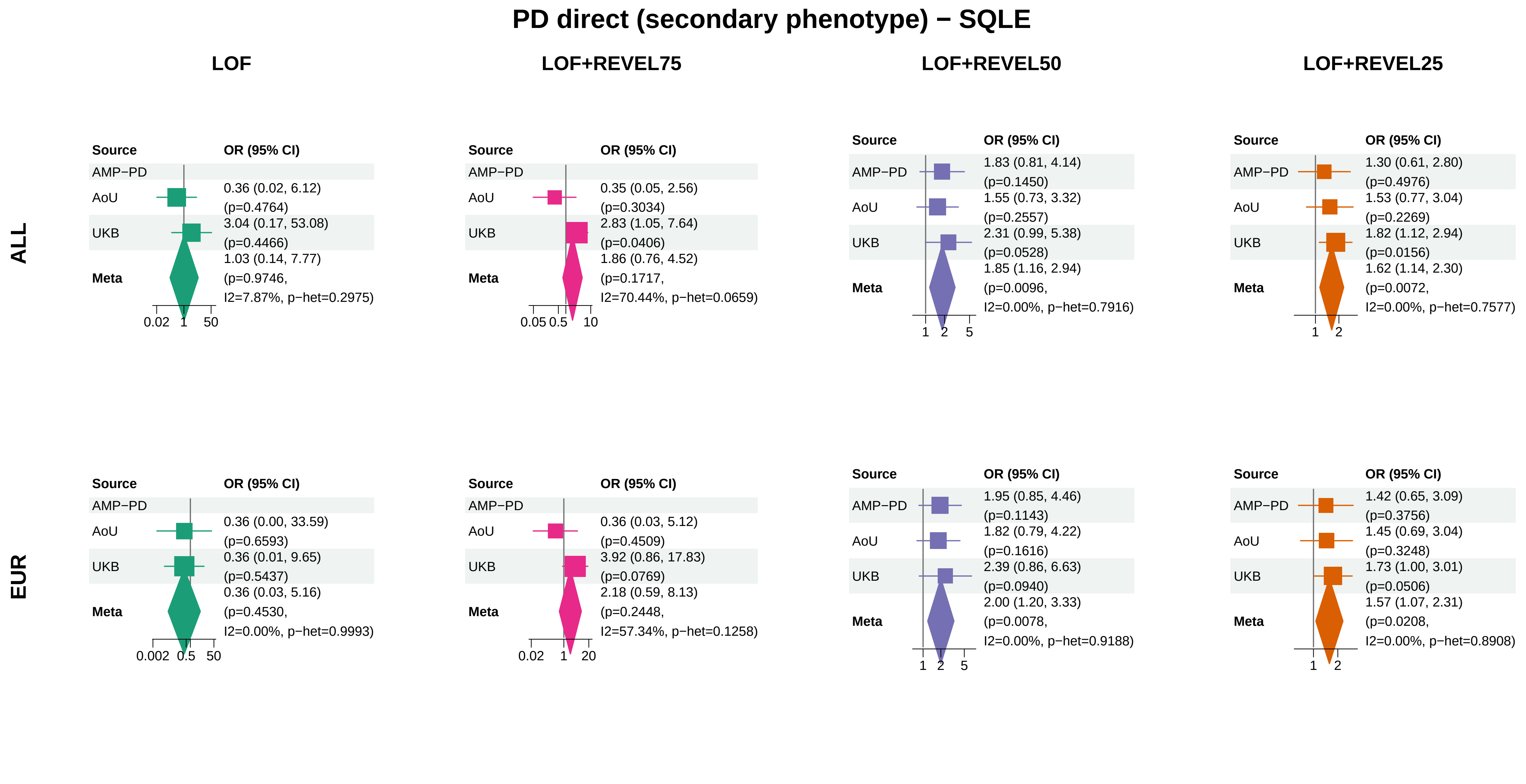


Meta OR/CI and P: exact−kappa=0.5 pooled REGENIE fixed−effects analysis.

### PD direct (secondary phenotype) − SPIRE2

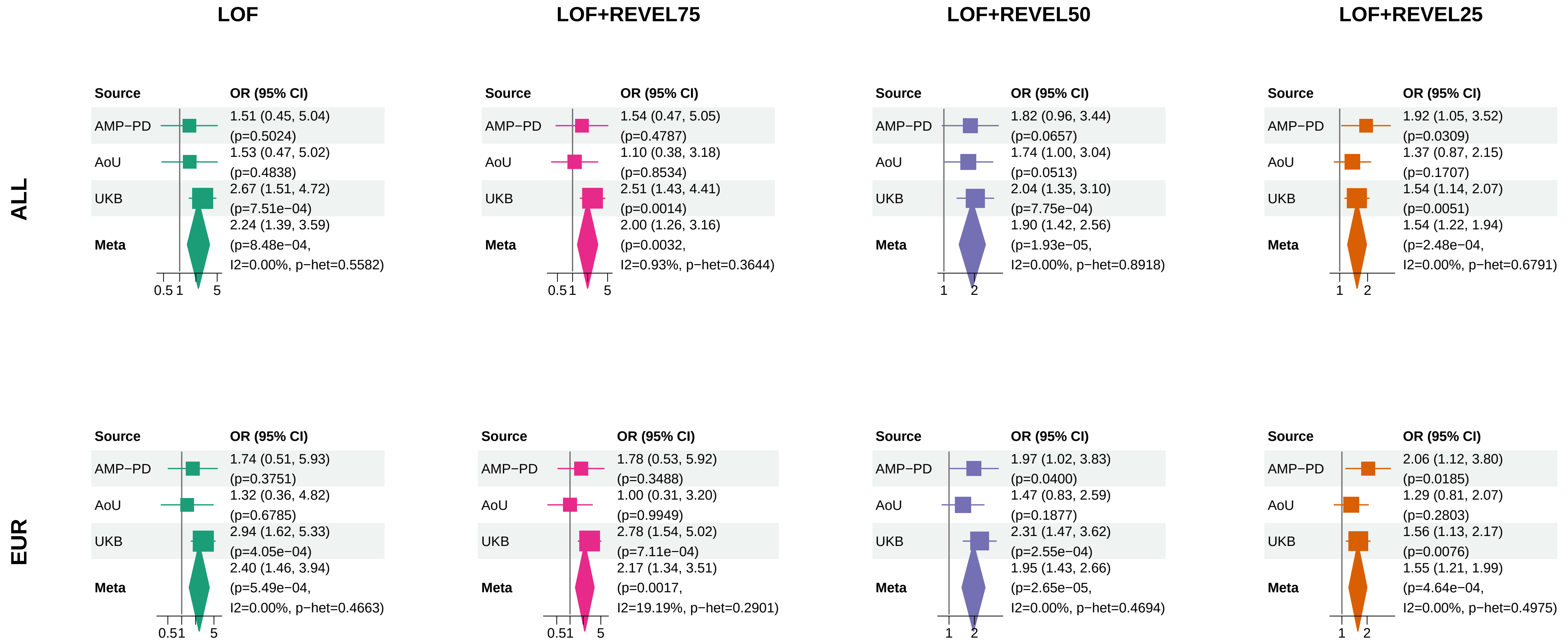


Meta OR/CI and P: exact−kappa=0.5 pooled REGENIE fixed−effects analysis.

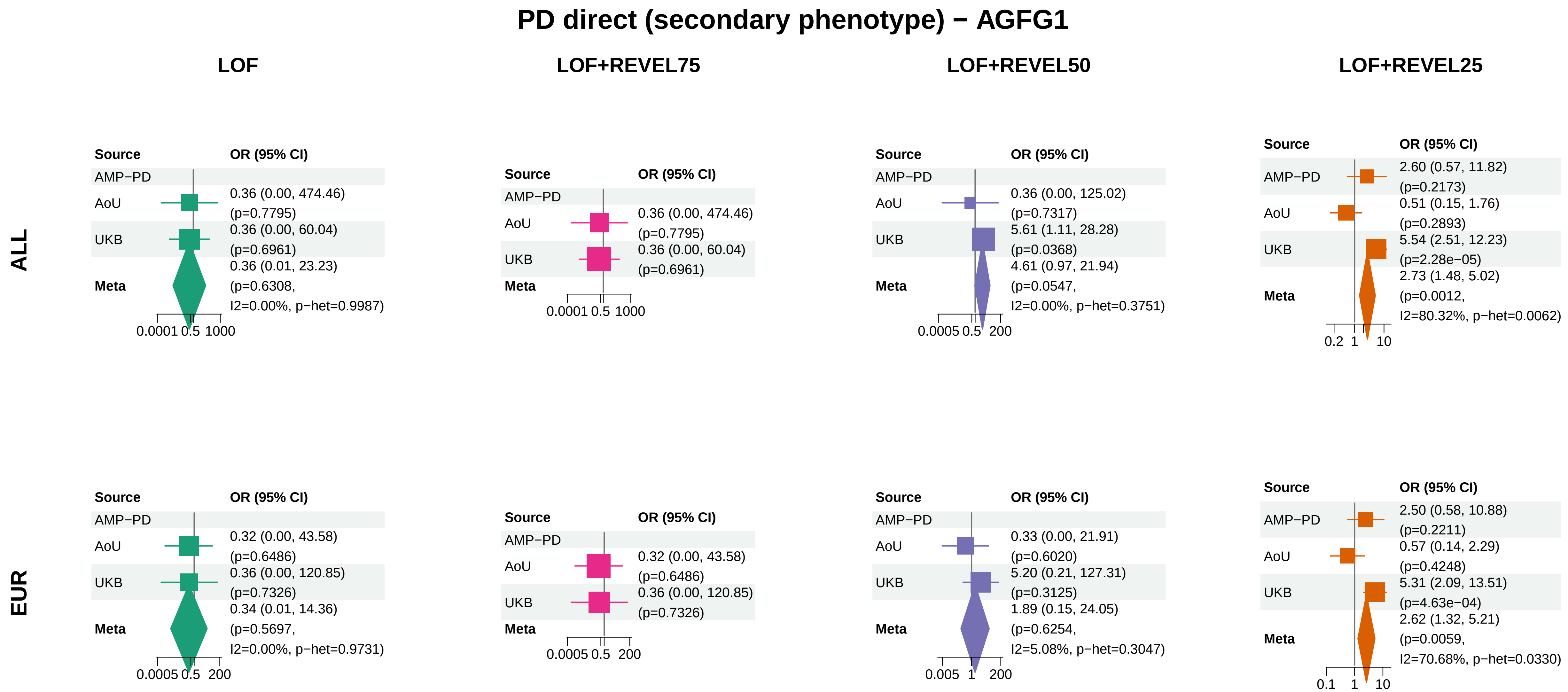


Meta OR/CI and P: exact−kappa=0.5 pooled REGENIE fixed−effects analysis.

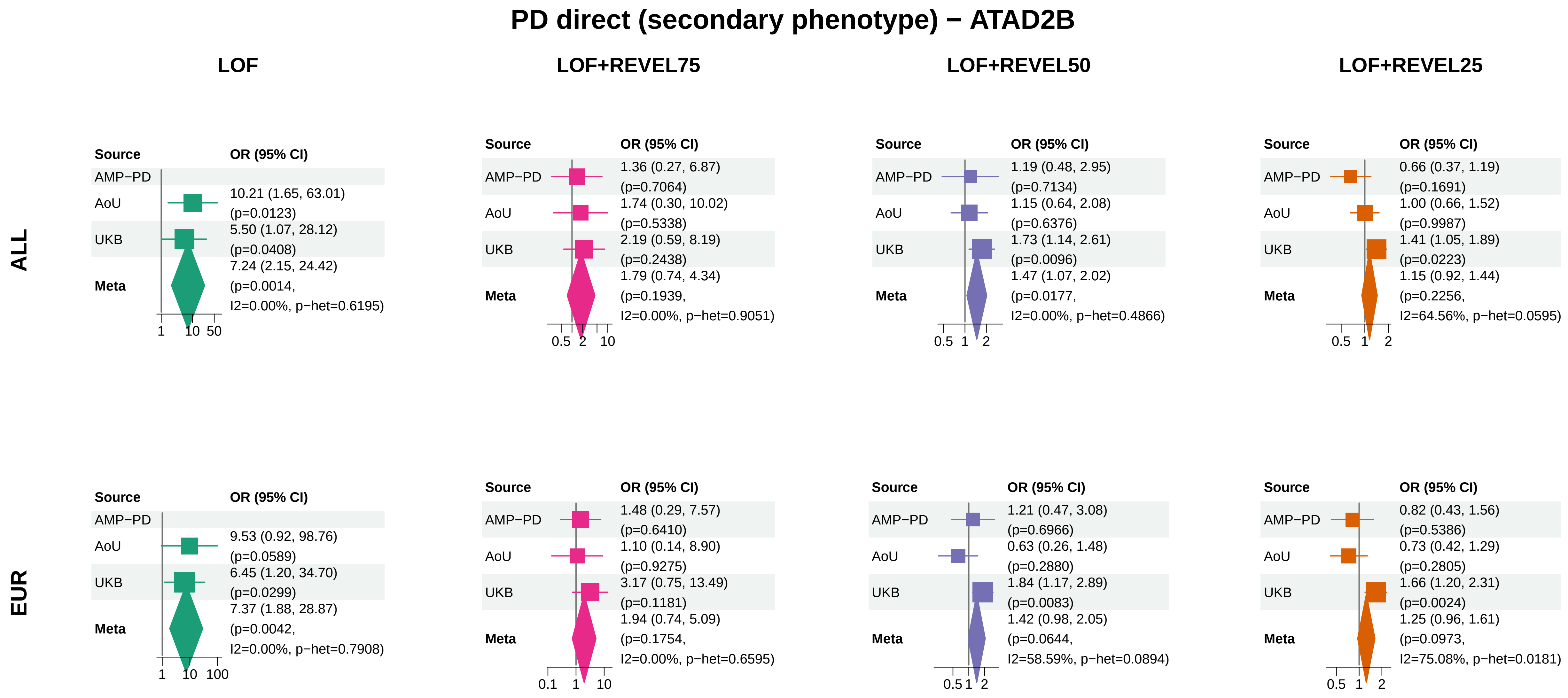


Meta OR/CI and P: exact−kappa=0.5 pooled REGENIE fixed−effects analysis.

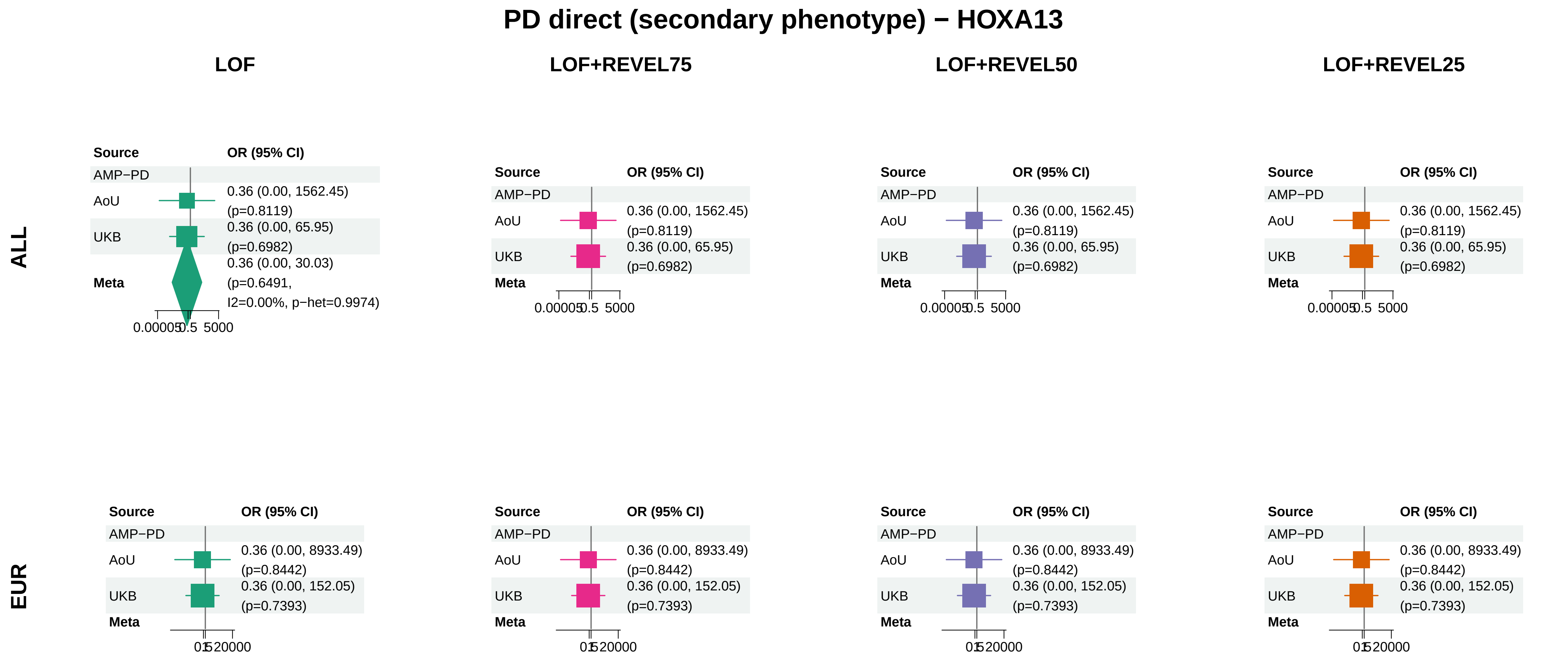


Meta OR/CI and P: exact−kappa=0.5 pooled REGENIE fixed−effects analysis.
