## Supplementary_Fig10 for "Population-scale burden analysis of rare damaging coding variants identifies novel risk genes for Alzheimer’s disease and related dementias and Parkinson’s disease and related disorders"

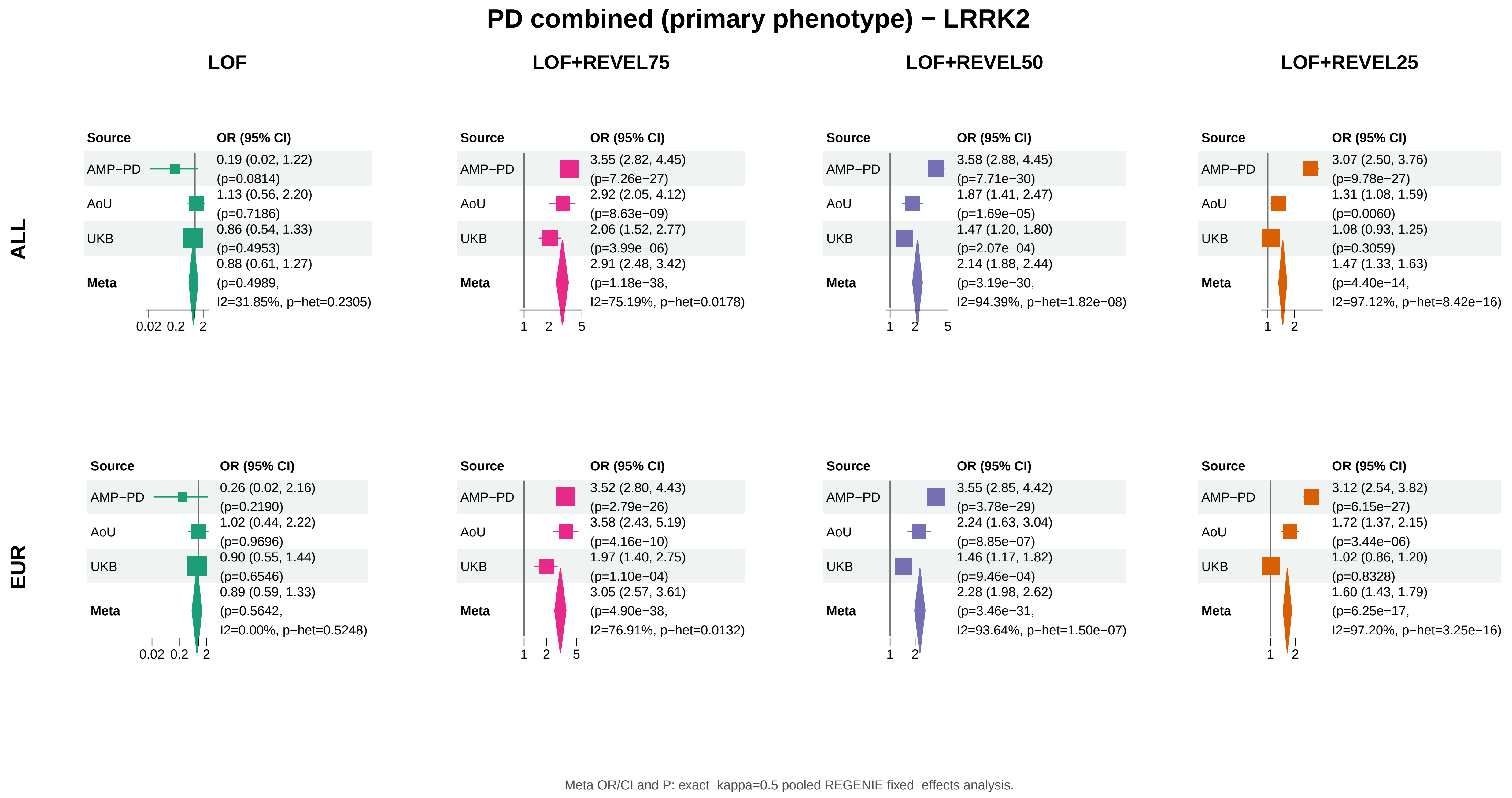
PD combined (primary phenotype) − LRRK2
LOF
LOF+REVEL75
LOF+REVEL50
LOF+REVEL25
ALL
Source
OR (95% CI)
AMP−PD
0.19 (0.02, 1.22) (p=0.0814)
AoU
1.13 (0.56, 2.20) (p=0.7186)
UKB
0.86 (0.54, 1.33) (p=0.4953)
Meta
0.88 (0.61, 1.27) (p=0.4989, I2=31.85%, p−het=0.2305)
0.02 0.2 2
Source
OR (95% CI)
AMP−PD
3.55 (2.82, 4.45) (p=7.26e−27)
AoU
2.92 (2.05, 4.12) (p=8.63e−09)
UKB
2.06 (1.52, 2.77) (p=3.99e−06)
Meta
2.91 (2.48, 3.42) (p=1.18e−38, I2=75.19%, p−het=0.0178)
1 2 5
Source
OR (95% CI)
AMP−PD
3.58 (2.88, 4.45) (p=7.71e−30)
AoU
1.87 (1.41, 2.47) (p=1.69e−05)
UKB
1.47 (1.20, 1.80) (p=2.07e−04)
Meta
2.14 (1.88, 2.44) (p=3.19e−30, I2=94.39%, p−het=1.82e−08)
1 2 5
Source
OR (95% CI)
AMP−PD
3.07 (2.50, 3.76) (p=9.78e−27)
AoU
1.31 (1.08, 1.59) (p=0.0060)
UKB
1.08 (0.93, 1.25) (p=0.3059)
Meta
1.47 (1.33, 1.63) (p=4.40e−14, I2=97.12%, p−het=8.42e−16)
1 2
EUR
Source
OR (95% CI)
AMP−PD
0.26 (0.02, 2.16) (p=0.2190)
AoU
1.02 (0.44, 2.22) (p=0.9696)
UKB
0.90 (0.55, 1.44) (p=0.6546)
Meta
0.89 (0.59, 1.33) (p=0.5642, I2=0.00%, p−het=0.5248)
0.02 0.2 2
Source
OR (95% CI)
AMP−PD
3.52 (2.80, 4.43) (p=2.79e−26)
AoU
3.58 (2.43, 5.19) (p=4.16e−10)
UKB
1.97 (1.40, 2.75) (p=1.10e−04)
Meta
3.05 (2.57, 3.61) (p=4.90e−38, I2=76.91%, p−het=0.0132)
1 2 5
Source
OR (95% CI)
AMP−PD
3.55 (2.85, 4.42) (p=3.78e−29)
AoU
2.24 (1.63, 3.04) (p=8.85e−07)
UKB
1.46 (1.17, 1.82) (p=9.46e−04)
Meta
2.28 (1.98, 2.62) (p=3.46e−31, I2=93.64%, p−het=1.50e−07)
1 2
Source
OR (95% CI)
AMP−PD
3.12 (2.54, 3.82) (p=6.15e−27)
AoU
1.72 (1.37, 2.15) (p=3.44e−06)
UKB
1.02 (0.86, 1.20) (p=0.8328)
Meta
1.60 (1.43, 1.79) (p=6.25e−17, I2=97.20%, p−het=3.25e−16)
1 2
Meta OR/CI and P: exact−kappa=0.5 pooled REGENIE fixed−effects analysis.

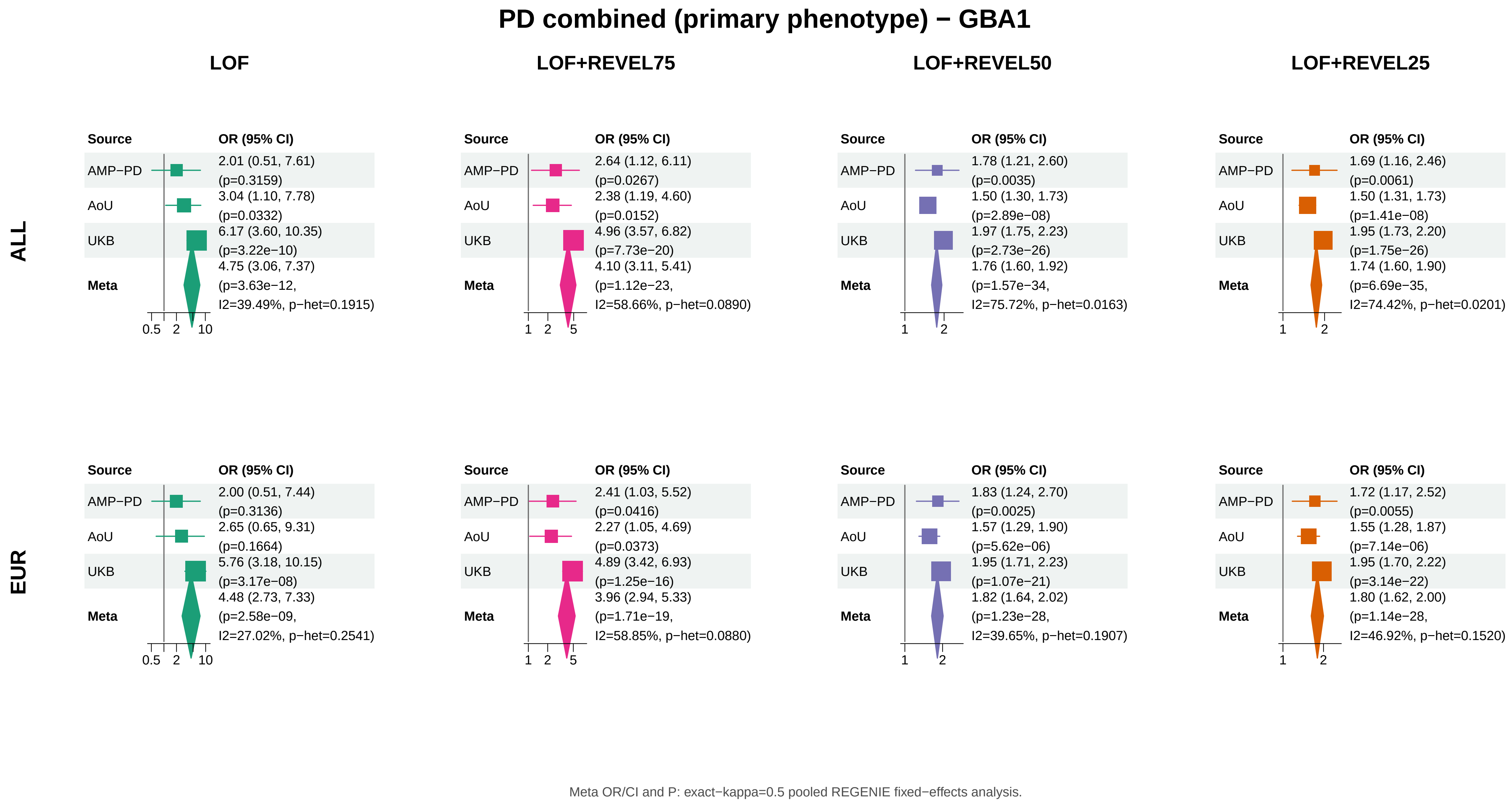
PD combined (primary phenotype) − GBA1
LOF
LOF+REVEL75
LOF+REVEL50
LOF+REVEL25
ALL
EUR
Source
OR (95% CI)
AMP−PD
AoU
UKB
Meta
ALL, LOF: AMP−PD 2.01 (0.51, 7.61) (p=0.3159); AoU 3.04 (1.10, 7.78) (p=0.0332); UKB 6.17 (3.60, 10.35) (p=3.22e−10); Meta 4.75 (3.06, 7.37) (p=3.63e−12, I2=39.49%, p−het=0.1915)
ALL, LOF+REVEL75: AMP−PD 2.64 (1.12, 6.11) (p=0.0267); AoU 2.38 (1.19, 4.60) (p=0.0152); UKB 4.96 (3.57, 6.82) (p=7.73e−20); Meta 4.10 (3.11, 5.41) (p=1.12e−23, I2=58.66%, p−het=0.0890)
ALL, LOF+REVEL50: AMP−PD 1.78 (1.21, 2.60) (p=0.0035); AoU 1.50 (1.30, 1.73) (p=2.89e−08); UKB 1.97 (1.75, 2.23) (p=2.73e−26); Meta 1.76 (1.60, 1.92) (p=1.57e−34, I2=75.72%, p−het=0.0163)
ALL, LOF+REVEL25: AMP−PD 1.69 (1.16, 2.46) (p=0.0061); AoU 1.50 (1.31, 1.73) (p=1.41e−08); UKB 1.95 (1.73, 2.20) (p=1.75e−26); Meta 1.74 (1.60, 1.90) (p=6.69e−35, I2=74.42%, p−het=0.0201)
EUR, LOF: AMP−PD 2.00 (0.51, 7.44) (p=0.3136); AoU 2.65 (0.65, 9.31) (p=0.1664); UKB 5.76 (3.18, 10.15) (p=3.17e−08); Meta 4.48 (2.73, 7.33) (p=2.58e−09, I2=27.02%, p−het=0.2541)
EUR, LOF+REVEL75: AMP−PD 2.41 (1.03, 5.52) (p=0.0416); AoU 2.27 (1.05, 4.69) (p=0.0373); UKB 4.89 (3.42, 6.93) (p=1.25e−16); Meta 3.96 (2.94, 5.33) (p=1.71e−19, I2=58.85%, p−het=0.0880)
EUR, LOF+REVEL50: AMP−PD 1.83 (1.24, 2.70) (p=0.0025); AoU 1.57 (1.29, 1.90) (p=5.62e−06); UKB 1.95 (1.71, 2.23) (p=1.07e−21); Meta 1.82 (1.64, 2.02) (p=1.23e−28, I2=39.65%, p−het=0.1907)
EUR, LOF+REVEL25: AMP−PD 1.72 (1.17, 2.52) (p=0.0055); AoU 1.55 (1.28, 1.87) (p=7.14e−06); UKB 1.95 (1.70, 2.22) (p=3.14e−22); Meta 1.80 (1.62, 2.00) (p=1.14e−28, I2=46.92%, p−het=0.1520)
0.5 2 10
1 2 5
1 2
Meta OR/CI and P: exact−kappa=0.5 pooled REGENIE fixed−effects analysis.

### PD combined (primary phenotype) − ANKRD27

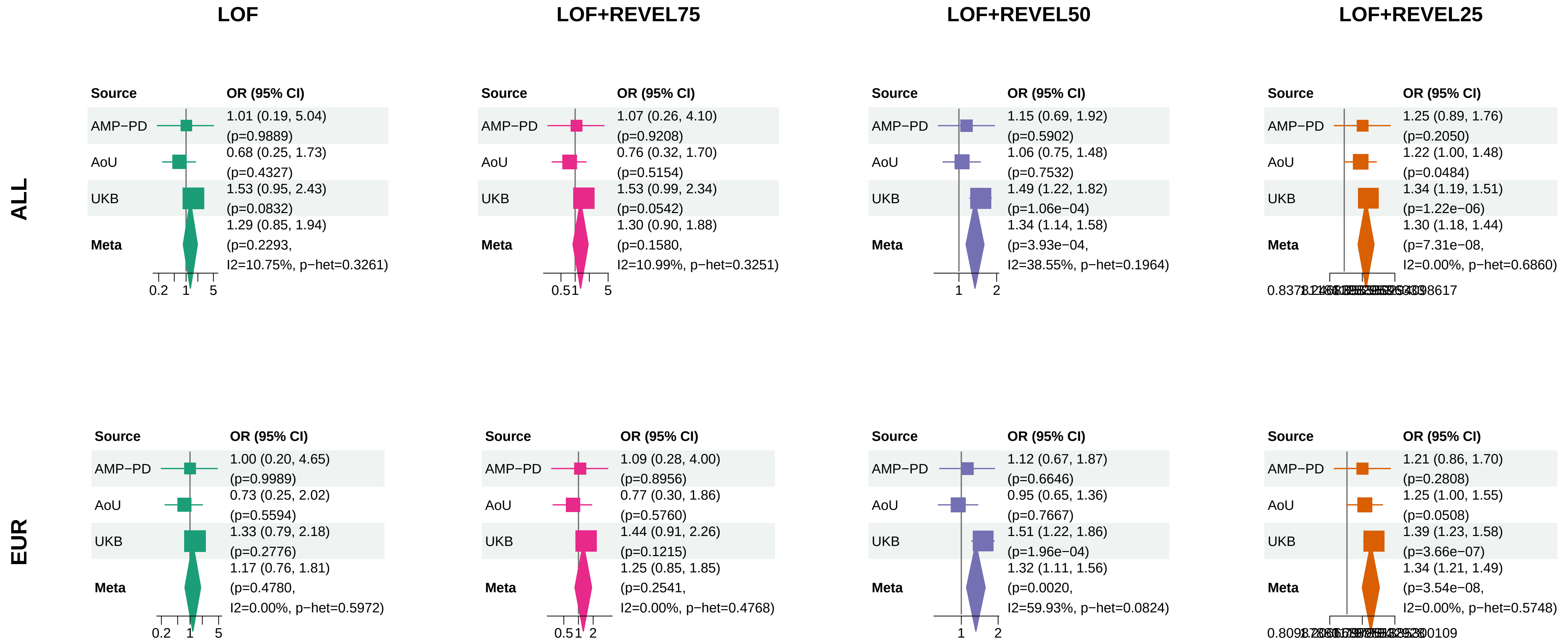


Meta OR/CI and P: exact−kappa=0.5 pooled REGENIE fixed−effects analysis.

### PD combined (primary phenotype) − CCL7

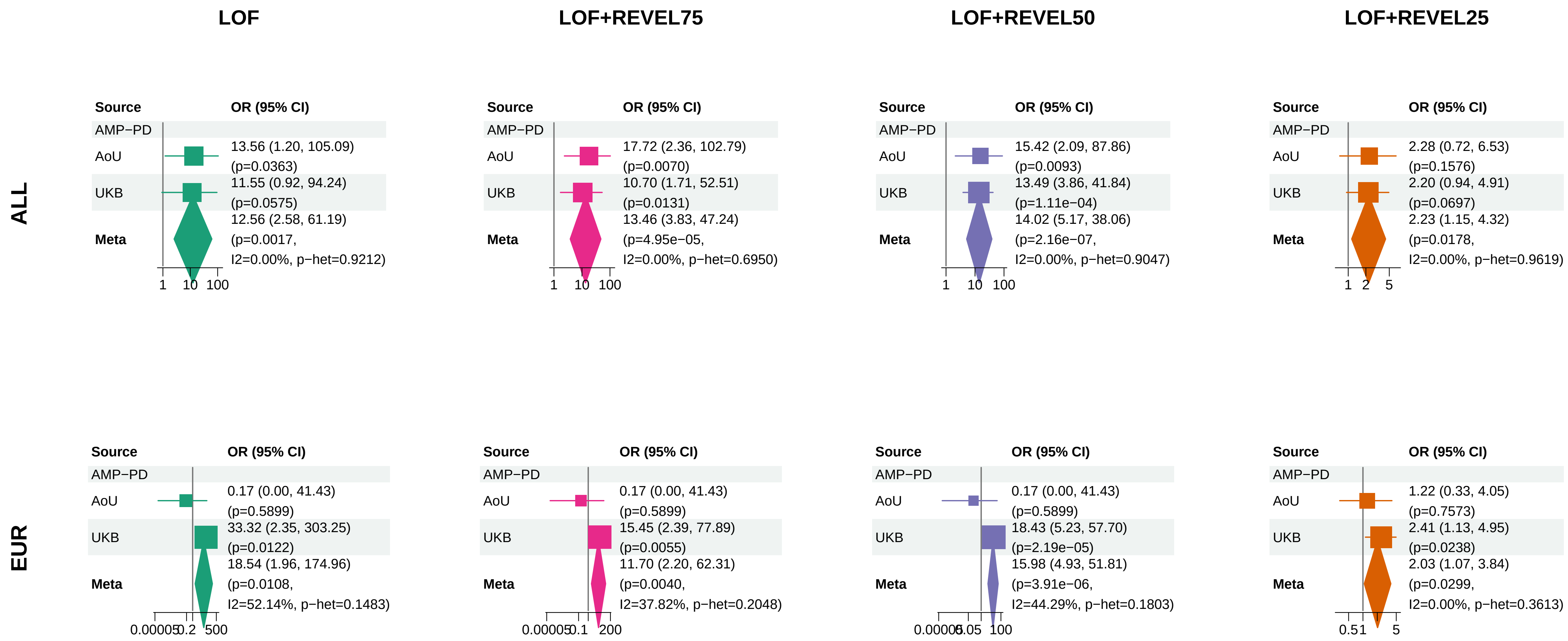


Meta OR/CI and P: exact−kappa=0.5 pooled REGENIE fixed−effects analysis.

### PD combined (primary phenotype) − PANX2

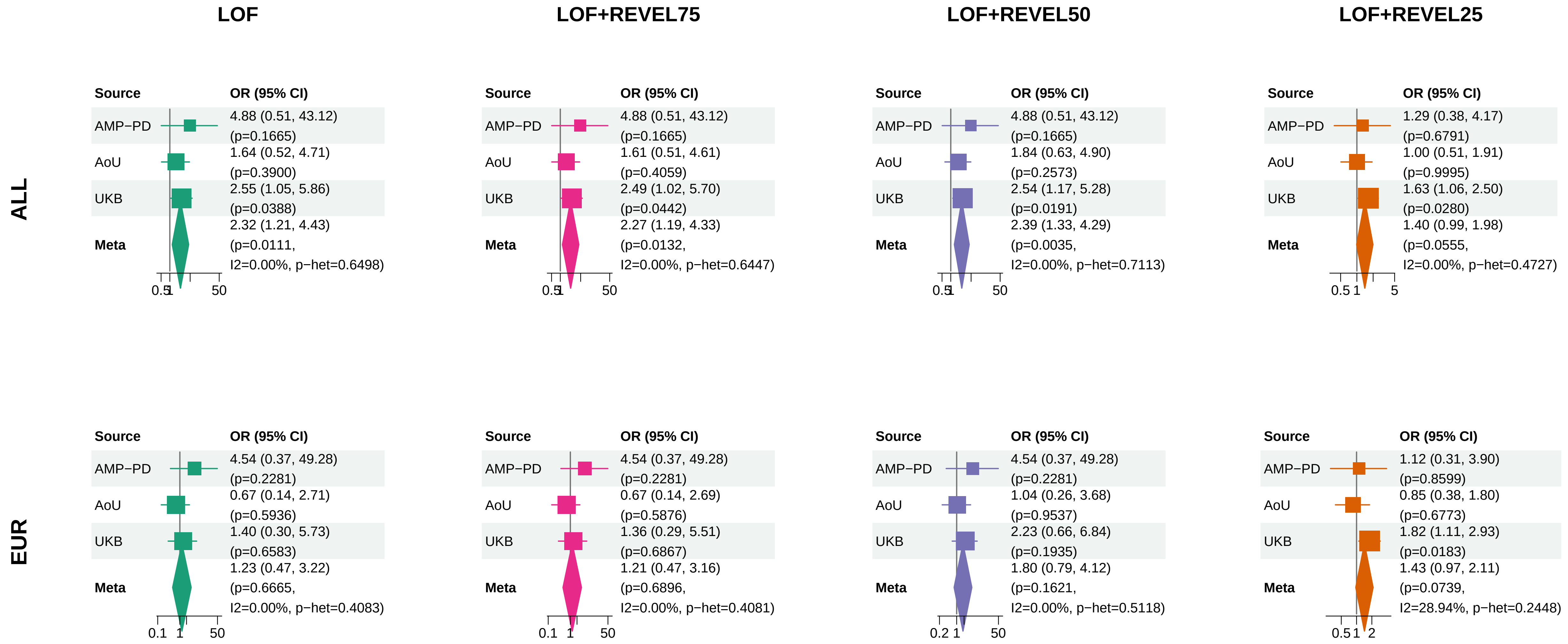


Meta OR/CI and P: exact−kappa=0.5 pooled REGENIE fixed−effects analysis.

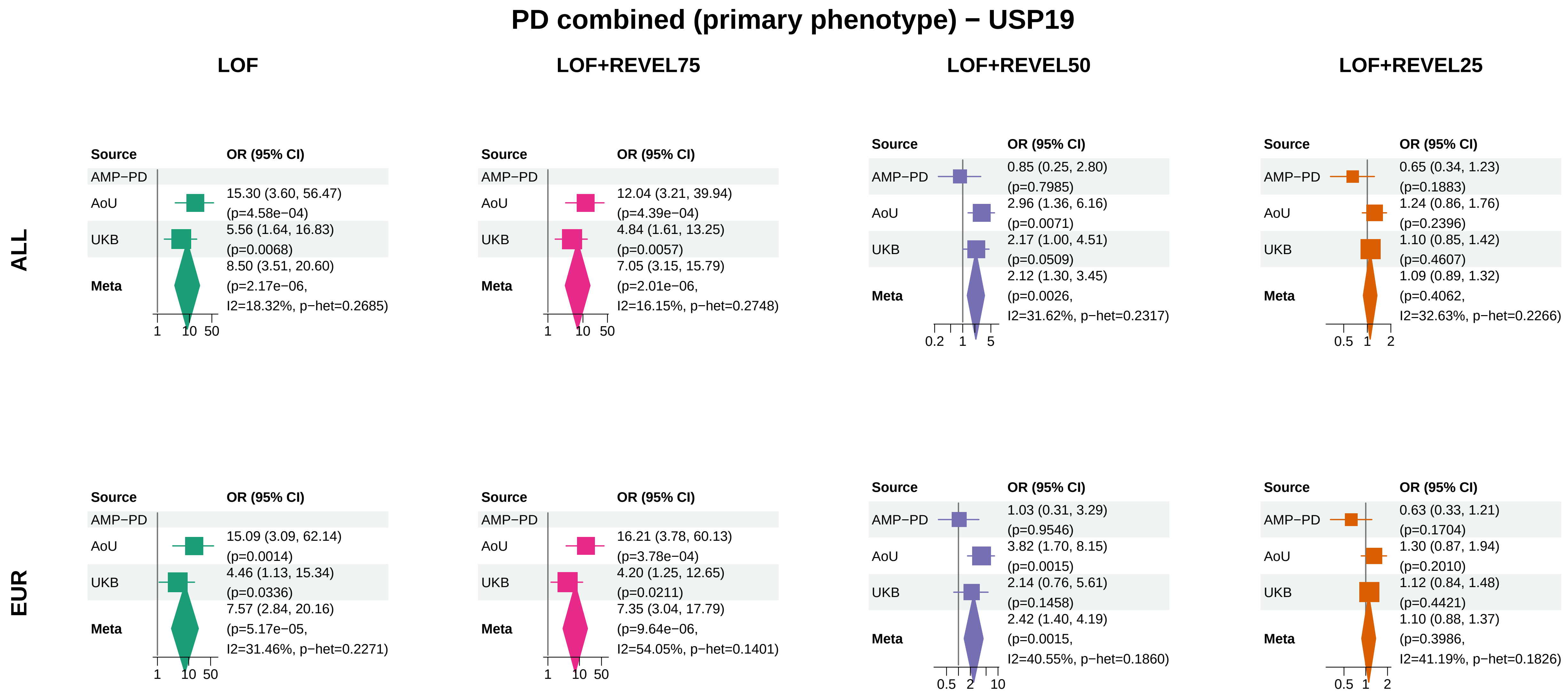


Meta OR/CI and P: exact−kappa=0.5 pooled REGENIE fixed−effects analysis.

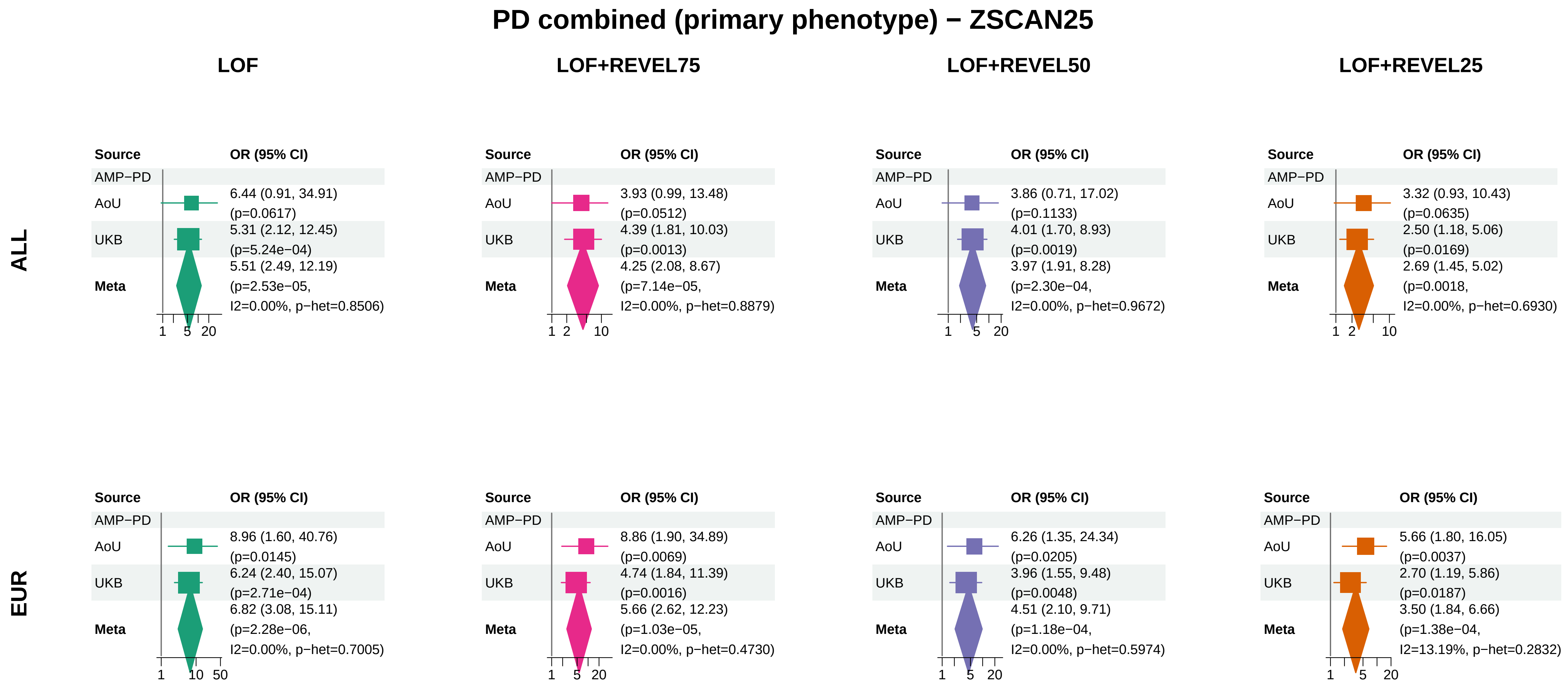


Meta OR/CI and P: exact−kappa=0.5 pooled REGENIE fixed−effects analysis.

### PD combined (primary phenotype) − BNIP3

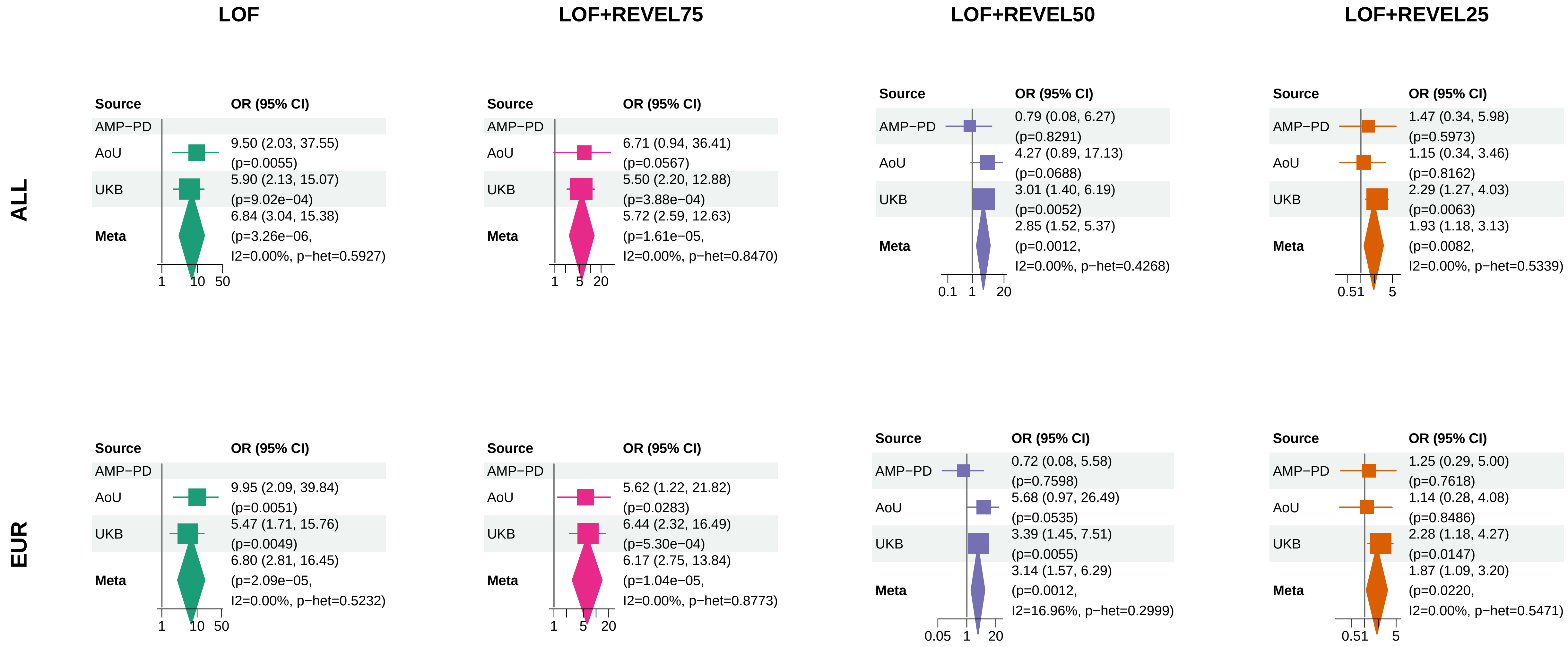


Meta OR/CI and P: exact−kappa=0.5 pooled REGENIE fixed−effects analysis.

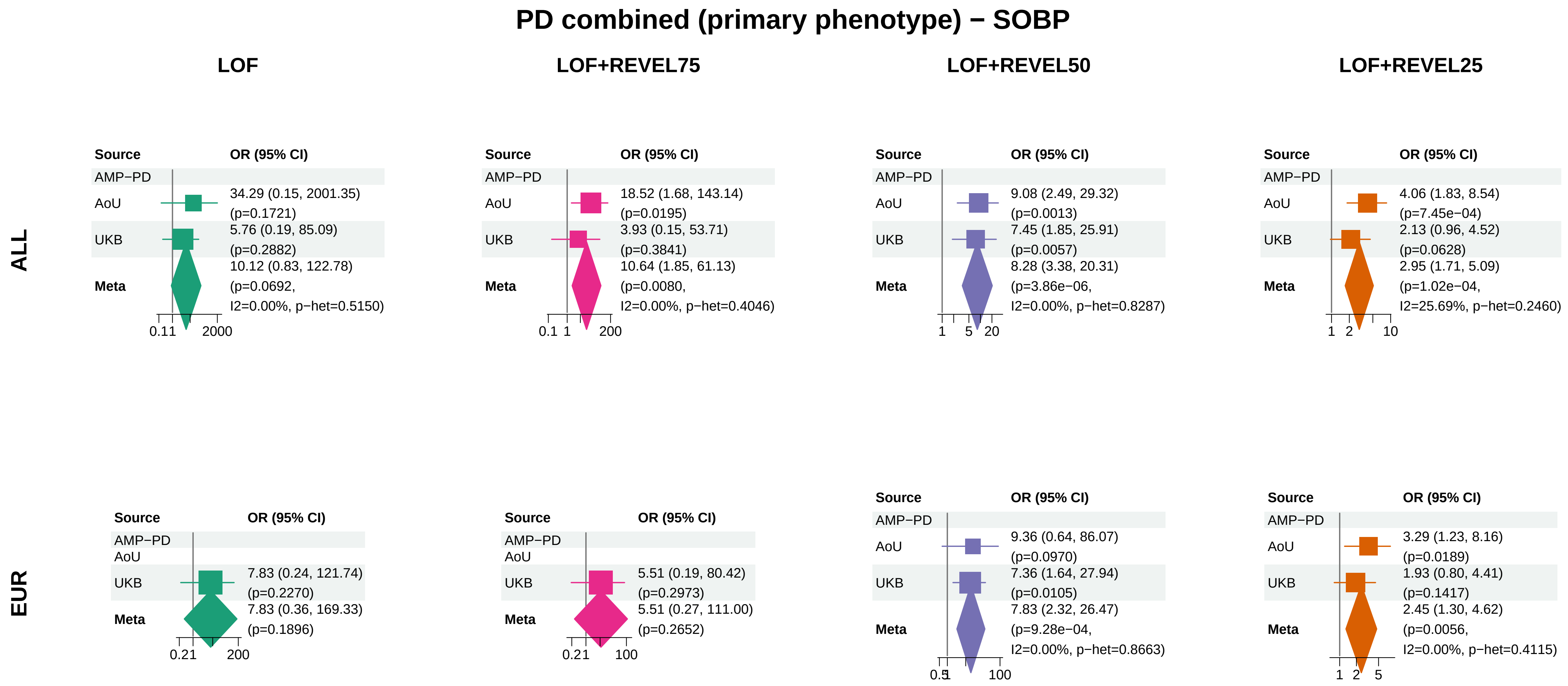


Meta OR/CI and P: exact−kappa=0.5 pooled REGENIE fixed−effects analysis.

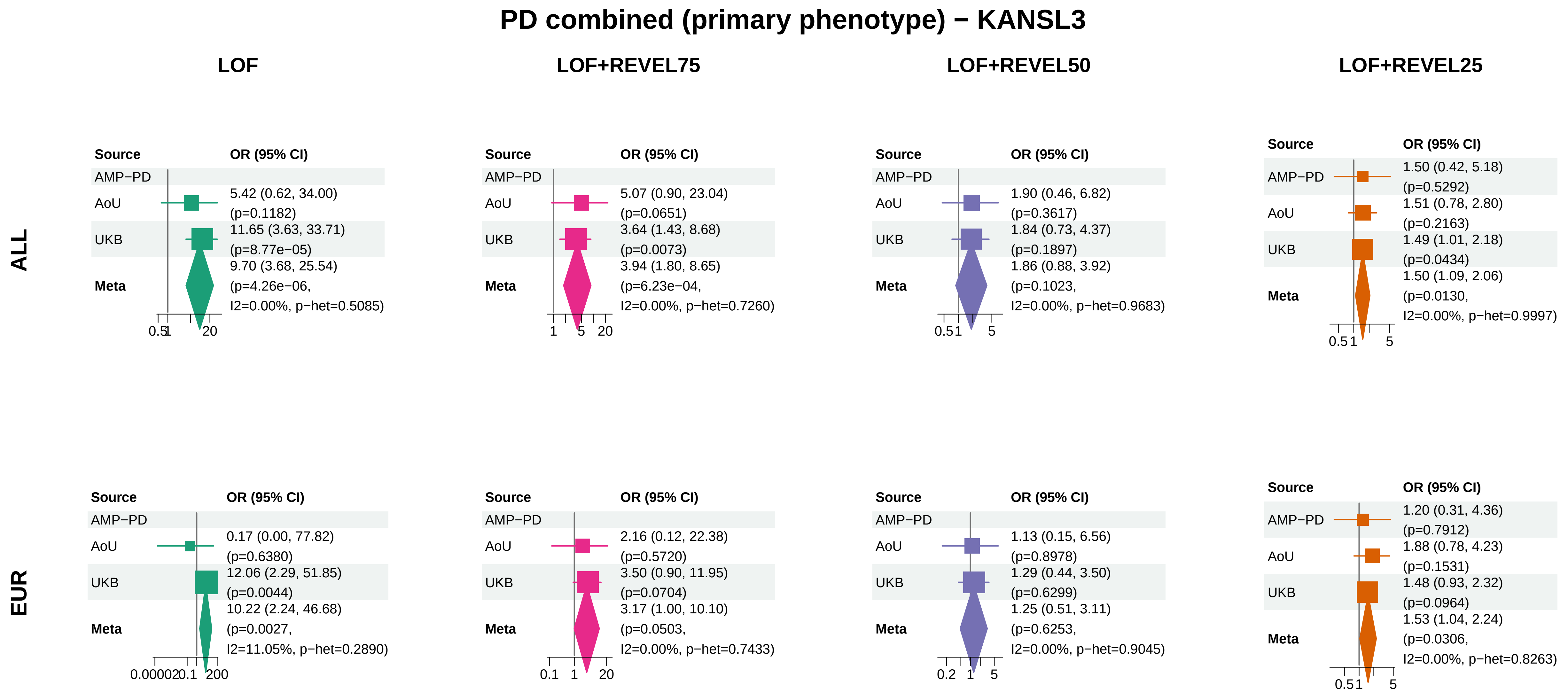


Meta OR/CI and P: exact−kappa=0.5 pooled REGENIE fixed−effects analysis.

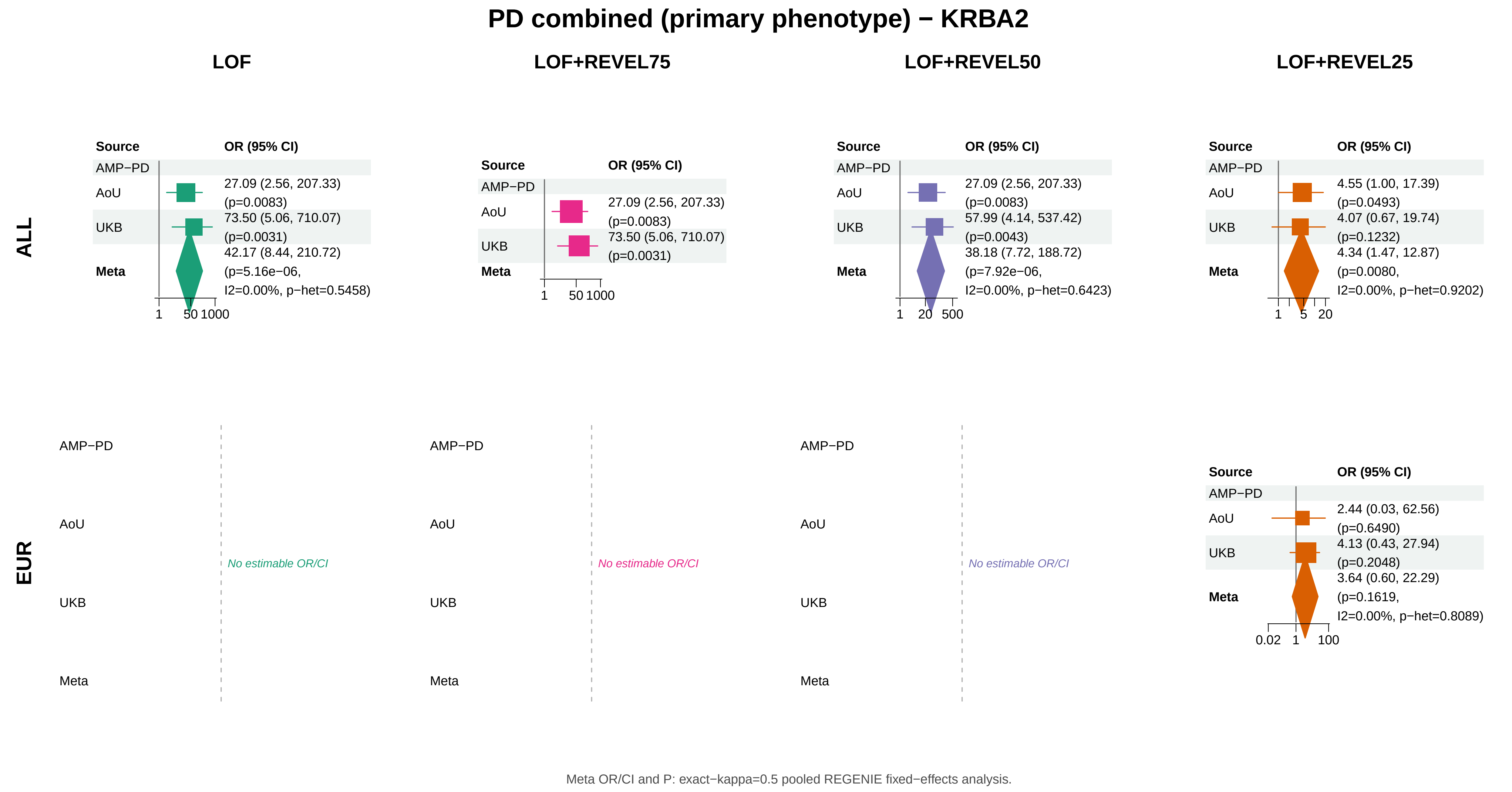
PD combined (primary phenotype) − KRBA2
LOF
LOF+REVEL75
LOF+REVEL50
LOF+REVEL25
ALL
EUR
Source
OR (95% CI)
AMP−PD
AoU
UKB
Meta
27.09 (2.56, 207.33) (p=0.0083)
73.50 (5.06, 710.07) (p=0.0031)
42.17 (8.44, 210.72) (p=5.16e−06, I2=0.00%, p−het=0.5458)
1 50 1000
27.09 (2.56, 207.33) (p=0.0083)
73.50 (5.06, 710.07) (p=0.0031)
1 50 1000
27.09 (2.56, 207.33) (p=0.0083)
57.99 (4.14, 537.42) (p=0.0043)
38.18 (7.72, 188.72) (p=7.92e−06, I2=0.00%, p−het=0.6423)
1 20 500
4.55 (1.00, 17.39) (p=0.0493)
4.07 (0.67, 19.74) (p=0.1232)
4.34 (1.47, 12.87) (p=0.0080, I2=0.00%, p−het=0.9202)
1 5 20
No estimable OR/CI
No estimable OR/CI
No estimable OR/CI
2.44 (0.03, 62.56) (p=0.6490)
4.13 (0.43, 27.94) (p=0.2048)
3.64 (0.60, 22.29) (p=0.1619, I2=0.00%, p−het=0.8089)
0.02 1 100
Meta OR/CI and P: exact−kappa=0.5 pooled REGENIE fixed−effects analysis.

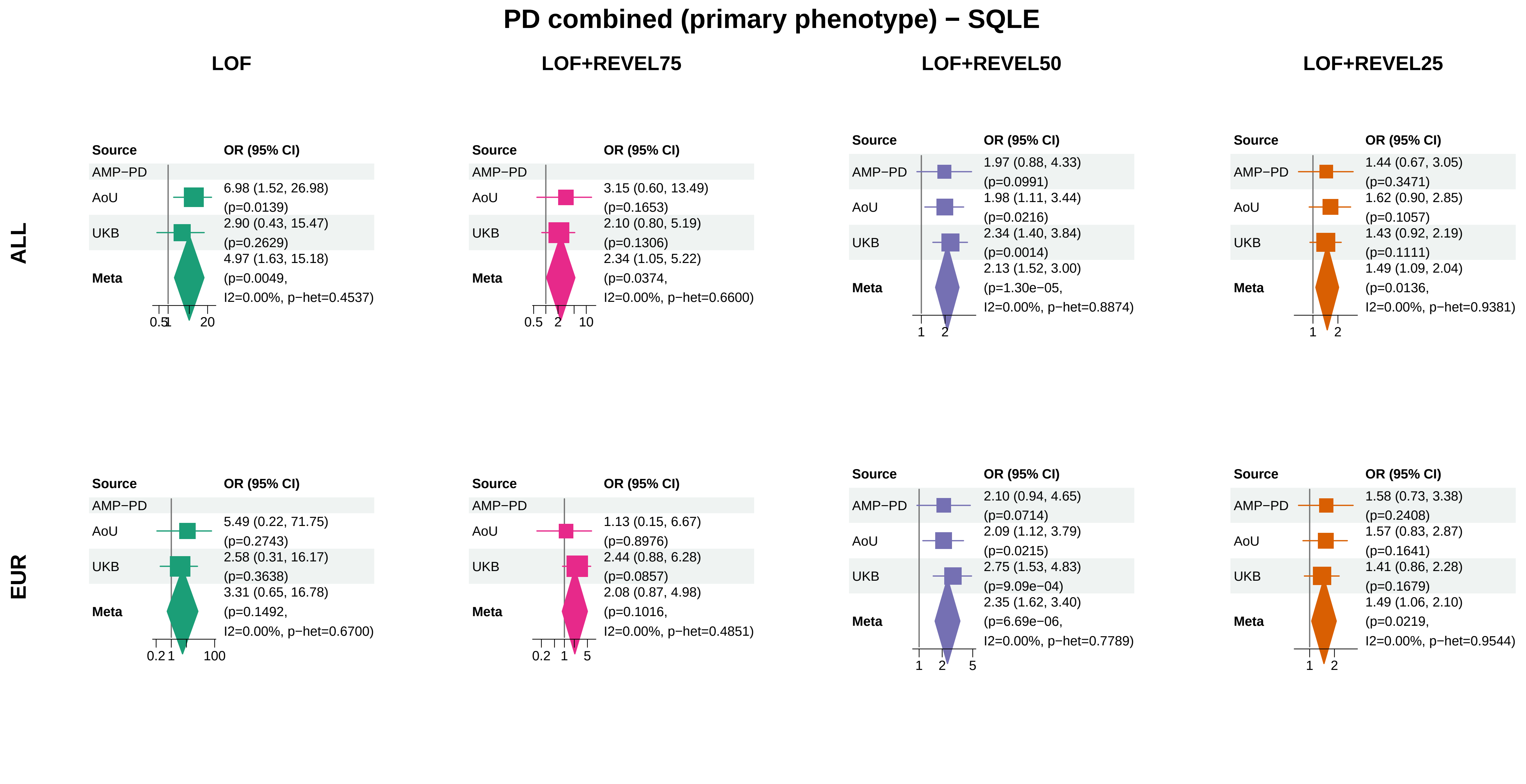


Meta OR/CI and P: exact−kappa=0.5 pooled REGENIE fixed−effects analysis.

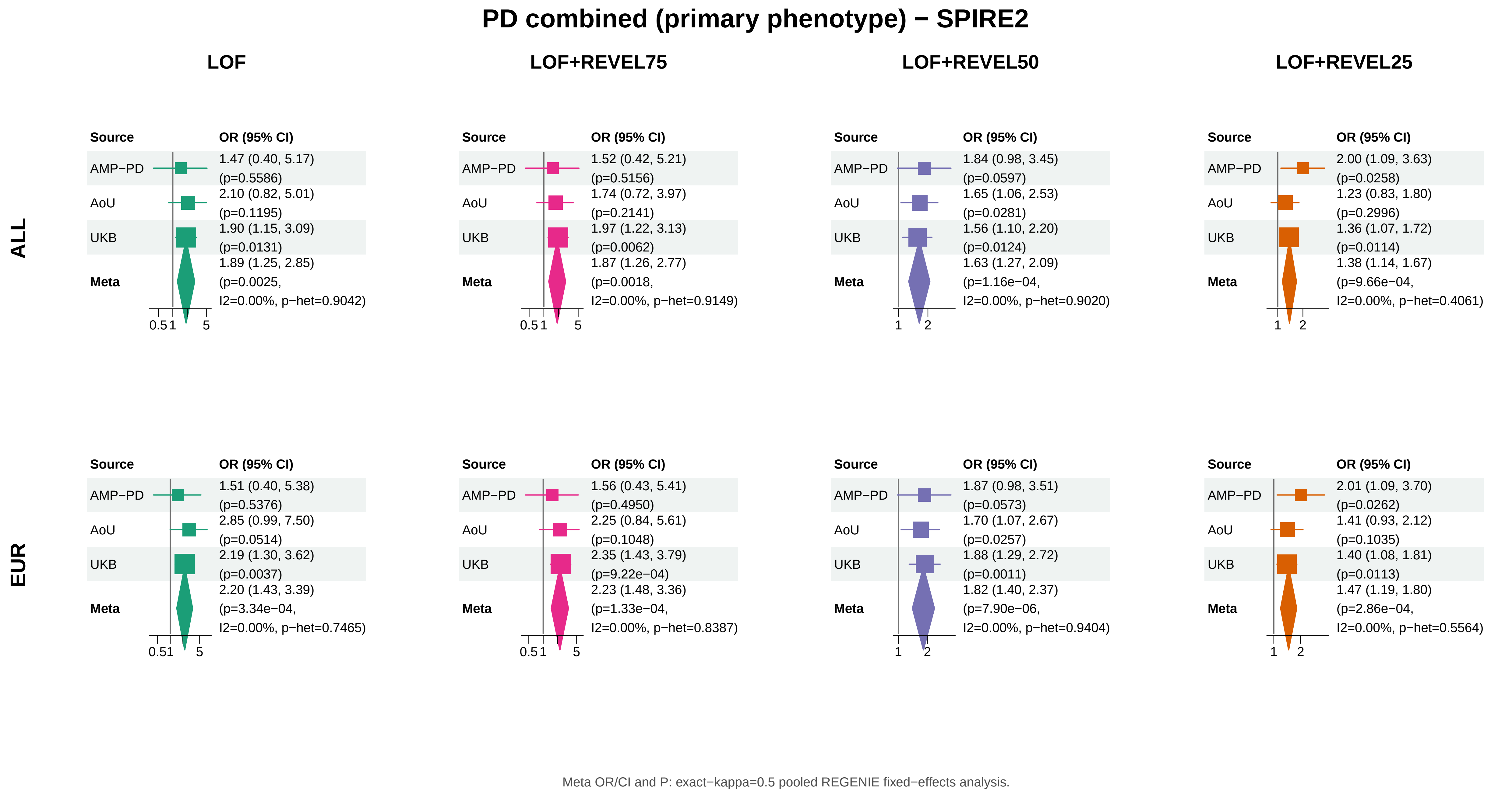
PD combined (primary phenotype) − SPIRE2
LOF
LOF+REVEL75
LOF+REVEL50
LOF+REVEL25
ALL
Source
OR (95% CI)
AMP−PD
1.47 (0.40, 5.17) (p=0.5586)
AoU
2.10 (0.82, 5.01) (p=0.1195)
UKB
1.90 (1.15, 3.09) (p=0.0131)
Meta
1.89 (1.25, 2.85) (p=0.0025, I2=0.00%, p−het=0.9042)
0.5 1 5
AMP−PD
1.52 (0.42, 5.21) (p=0.5156)
AoU
1.74 (0.72, 3.97) (p=0.2141)
UKB
1.97 (1.22, 3.13) (p=0.0062)
Meta
1.87 (1.26, 2.77) (p=0.0018, I2=0.00%, p−het=0.9149)
0.5 1 5
AMP−PD
1.84 (0.98, 3.45) (p=0.0597)
AoU
1.65 (1.06, 2.53) (p=0.0281)
UKB
1.56 (1.10, 2.20) (p=0.0124)
Meta
1.63 (1.27, 2.09) (p=1.16e−04, I2=0.00%, p−het=0.9020)
1 2
AMP−PD
2.00 (1.09, 3.63) (p=0.0258)
AoU
1.23 (0.83, 1.80) (p=0.2996)
UKB
1.36 (1.07, 1.72) (p=0.0114)
Meta
1.38 (1.14, 1.67) (p=9.66e−04, I2=0.00%, p−het=0.4061)
1 2
EUR
Source
OR (95% CI)
AMP−PD
1.51 (0.40, 5.38) (p=0.5376)
AoU
2.85 (0.99, 7.50) (p=0.0514)
UKB
2.19 (1.30, 3.62) (p=0.0037)
Meta
2.20 (1.43, 3.39) (p=3.34e−04, I2=0.00%, p−het=0.7465)
0.51 5
AMP−PD
1.56 (0.43, 5.41) (p=0.4950)
AoU
2.25 (0.84, 5.61) (p=0.1048)
UKB
2.35 (1.43, 3.79) (p=9.22e−04)
Meta
2.23 (1.48, 3.36) (p=1.33e−04, I2=0.00%, p−het=0.8387)
0.5 1 5
AMP−PD
1.87 (0.98, 3.51) (p=0.0573)
AoU
1.70 (1.07, 2.67) (p=0.0257)
UKB
1.88 (1.29, 2.72) (p=0.0011)
Meta
1.82 (1.40, 2.37) (p=7.90e−06, I2=0.00%, p−het=0.9404)
1 2
AMP−PD
2.01 (1.09, 3.70) (p=0.0262)
AoU
1.41 (0.93, 2.12) (p=0.1035)
UKB
1.40 (1.08, 1.81) (p=0.0113)
Meta
1.47 (1.19, 1.80) (p=2.86e−04, I2=0.00%, p−het=0.5564)
1 2
Meta OR/CI and P: exact−kappa=0.5 pooled REGENIE fixed−effects analysis.

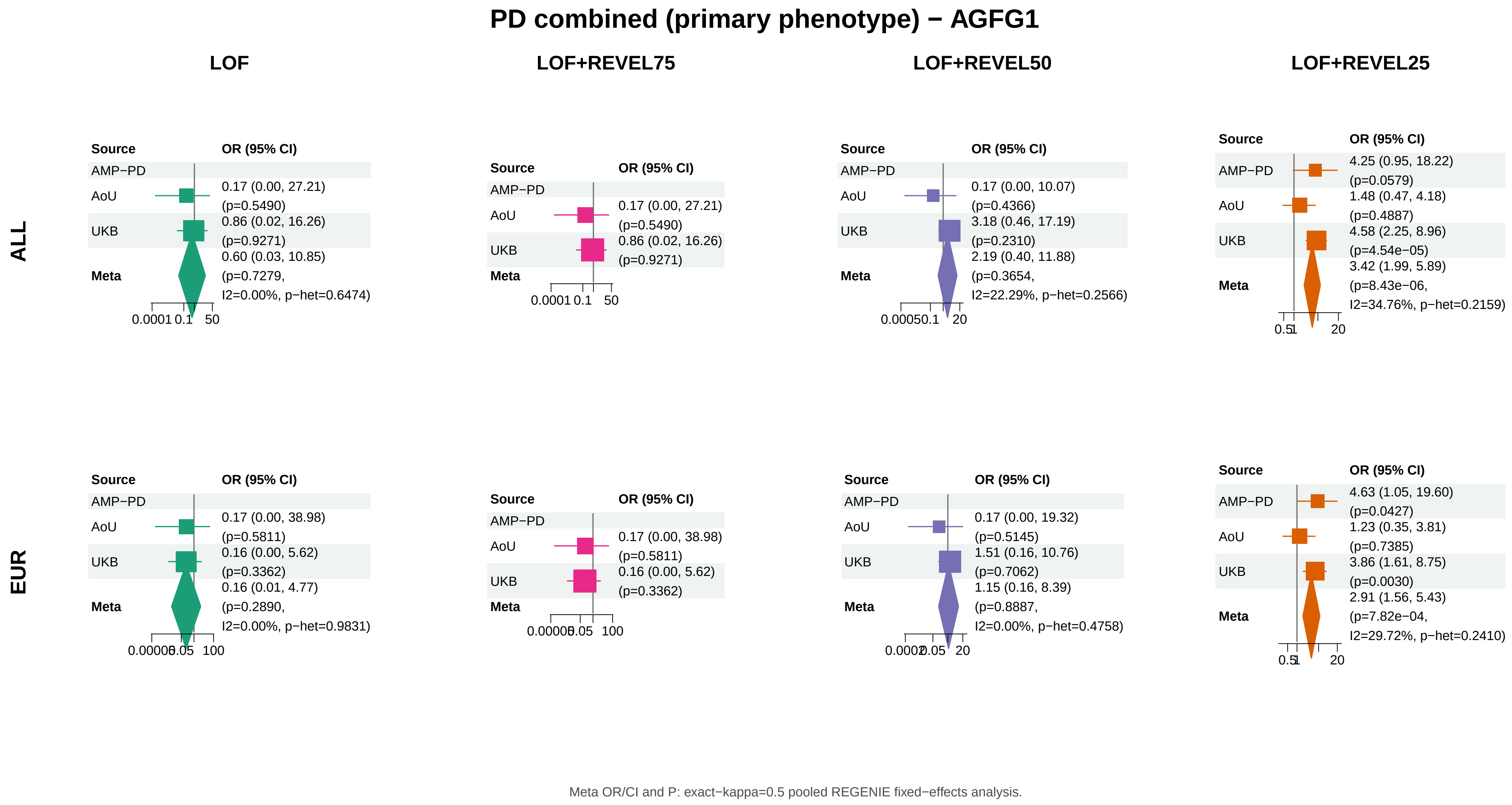
PD combined (primary phenotype) − AGFG1
LOF
LOF+REVEL75
LOF+REVEL50
LOF+REVEL25
ALL
EUR
Source
OR (95% CI)
AMP−PD
AoU
UKB
Meta
0.17 (0.00, 27.21) (p=0.5490)
0.86 (0.02, 16.26) (p=0.9271)
0.60 (0.03, 10.85) (p=0.7279, I2=0.00%, p−het=0.6474)
0.0001 0.1 50
0.17 (0.00, 27.21) (p=0.5490)
0.86 (0.02, 16.26) (p=0.9271)
0.0001 0.1 50
0.17 (0.00, 10.07) (p=0.4366)
3.18 (0.46, 17.19) (p=0.2310)
2.19 (0.40, 11.88) (p=0.3654, I2=22.29%, p−het=0.2566)
0.00050.1 20
4.25 (0.95, 18.22) (p=0.0579)
1.48 (0.47, 4.18) (p=0.4887)
4.58 (2.25, 8.96) (p=4.54e−05)
3.42 (1.99, 5.89) (p=8.43e−06, I2=34.76%, p−het=0.2159)
0.51 20
0.17 (0.00, 38.98) (p=0.5811)
0.16 (0.00, 5.62) (p=0.3362)
0.16 (0.01, 4.77) (p=0.2890, I2=0.00%, p−het=0.9831)
0.00005 0.05 100
0.17 (0.00, 38.98) (p=0.5811)
0.16 (0.00, 5.62) (p=0.3362)
0.00005 0.05 100
0.17 (0.00, 19.32) (p=0.5145)
1.51 (0.16, 10.76) (p=0.7062)
1.15 (0.16, 8.39) (p=0.8887, I2=0.00%, p−het=0.4758)
0.0002 0.05 20
4.63 (1.05, 19.60) (p=0.0427)
1.23 (0.35, 3.81) (p=0.7385)
3.86 (1.61, 8.75) (p=0.0030)
2.91 (1.56, 5.43) (p=7.82e−04, I2=29.72%, p−het=0.2410)
0.51 20
Meta OR/CI and P: exact−kappa=0.5 pooled REGENIE fixed−effects analysis.

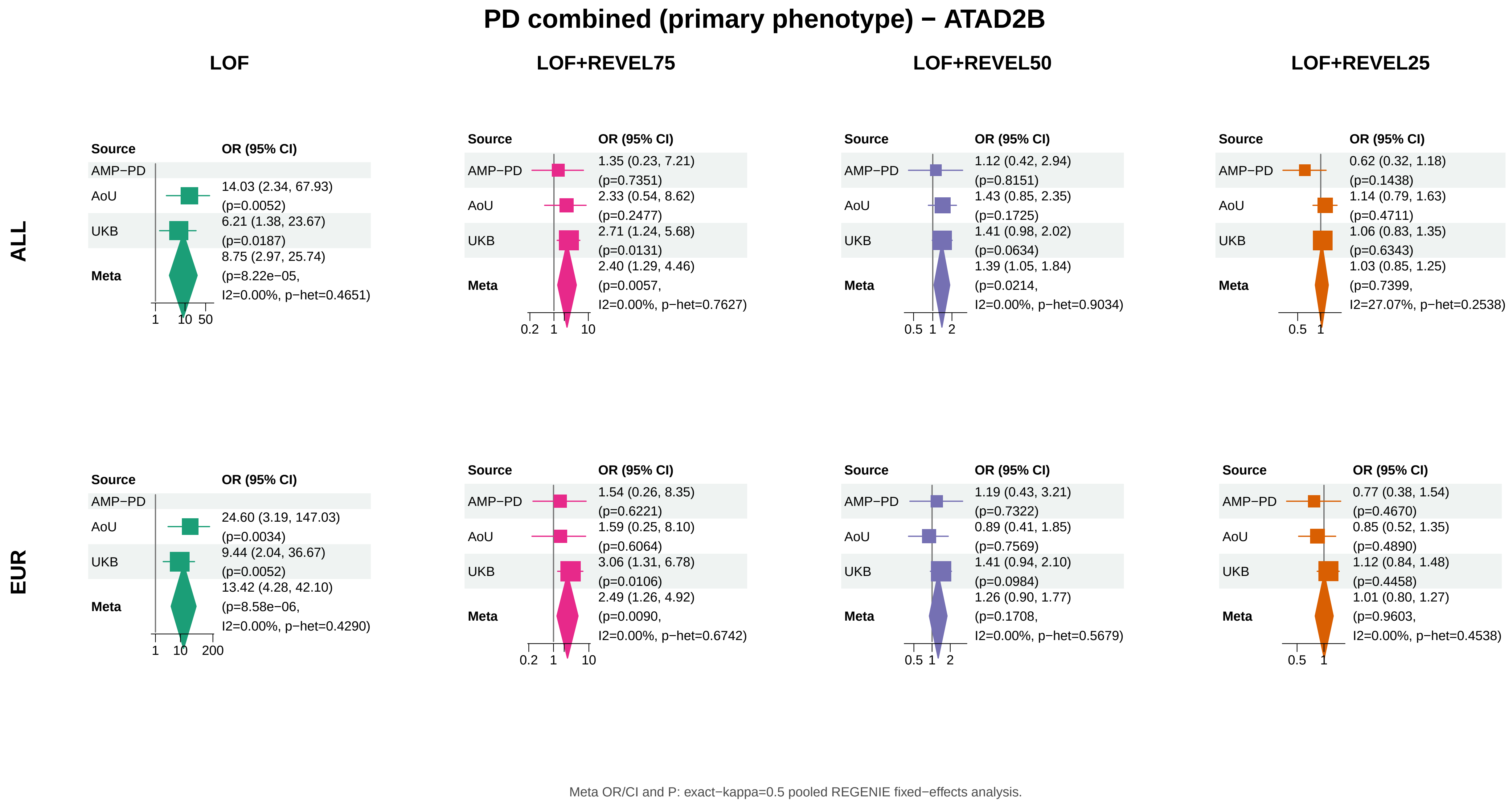
PD combined (primary phenotype) − ATAD2B
LOF
LOF+REVEL75
LOF+REVEL50
LOF+REVEL25
ALL
EUR
Source
OR (95% CI)
AMP−PD
AoU
UKB
Meta
14.03 (2.34, 67.93) (p=0.0052)
6.21 (1.38, 23.67) (p=0.0187)
8.75 (2.97, 25.74) (p=8.22e−05, I2=0.00%, p−het=0.4651)
1 10 50
1.35 (0.23, 7.21) (p=0.7351)
2.33 (0.54, 8.62) (p=0.2477)
2.71 (1.24, 5.68) (p=0.0131)
2.40 (1.29, 4.46) (p=0.0057, I2=0.00%, p−het=0.7627)
0.2 1 10
1.12 (0.42, 2.94) (p=0.8151)
1.43 (0.85, 2.35) (p=0.1725)
1.41 (0.98, 2.02) (p=0.0634)
1.39 (1.05, 1.84) (p=0.0214, I2=0.00%, p−het=0.9034)
0.5 1 2
0.62 (0.32, 1.18) (p=0.1438)
1.14 (0.79, 1.63) (p=0.4711)
1.06 (0.83, 1.35) (p=0.6343)
1.03 (0.85, 1.25) (p=0.7399, I2=27.07%, p−het=0.2538)
0.5 1
24.60 (3.19, 147.03) (p=0.0034)
9.44 (2.04, 36.67) (p=0.0052)
13.42 (4.28, 42.10) (p=8.58e−06, I2=0.00%, p−het=0.4290)
1 10 200
1.54 (0.26, 8.35) (p=0.6221)
1.59 (0.25, 8.10) (p=0.6064)
3.06 (1.31, 6.78) (p=0.0106)
2.49 (1.26, 4.92) (p=0.0090, I2=0.00%, p−het=0.6742)
0.2 1 10
1.19 (0.43, 3.21) (p=0.7322)
0.89 (0.41, 1.85) (p=0.7569)
1.41 (0.94, 2.10) (p=0.0984)
1.26 (0.90, 1.77) (p=0.1708, I2=0.00%, p−het=0.5679)
0.5 1 2
0.77 (0.38, 1.54) (p=0.4670)
0.85 (0.52, 1.35) (p=0.4890)
1.12 (0.84, 1.48) (p=0.4458)
1.01 (0.80, 1.27) (p=0.9603, I2=0.00%, p−het=0.4538)
0.5 1
Meta OR/CI and P: exact−kappa=0.5 pooled REGENIE fixed−effects analysis.

### PD combined (primary phenotype) − HOXA13

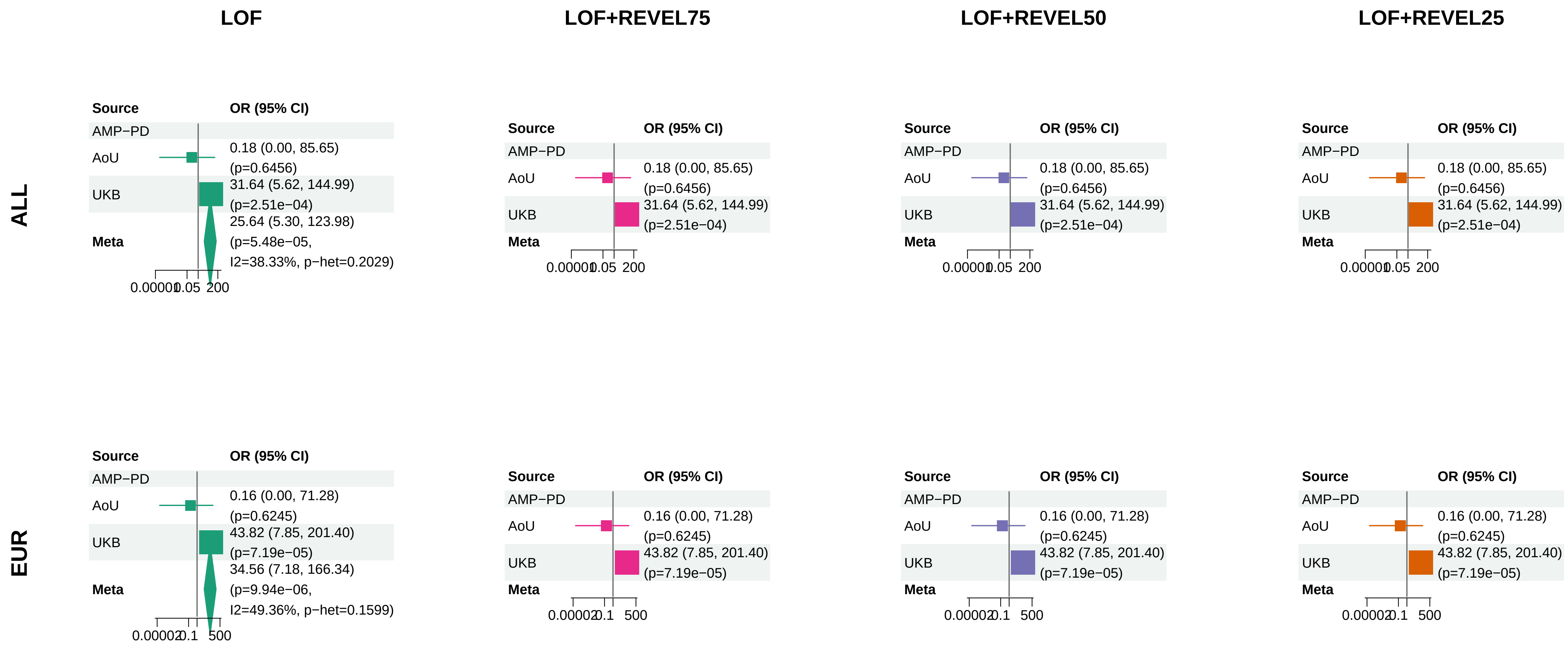


Meta OR/CI and P: exact−kappa=0.5 pooled REGENIE fixed−effects analysis.
