## Supplementary_Fig7 for "Population-scale burden analysis of rare damaging coding variants identifies novel risk genes for Alzheimer’s disease and related dementias and Parkinson’s disease and related disorders"

### AD direct (secondary phenotype) − PSEN1

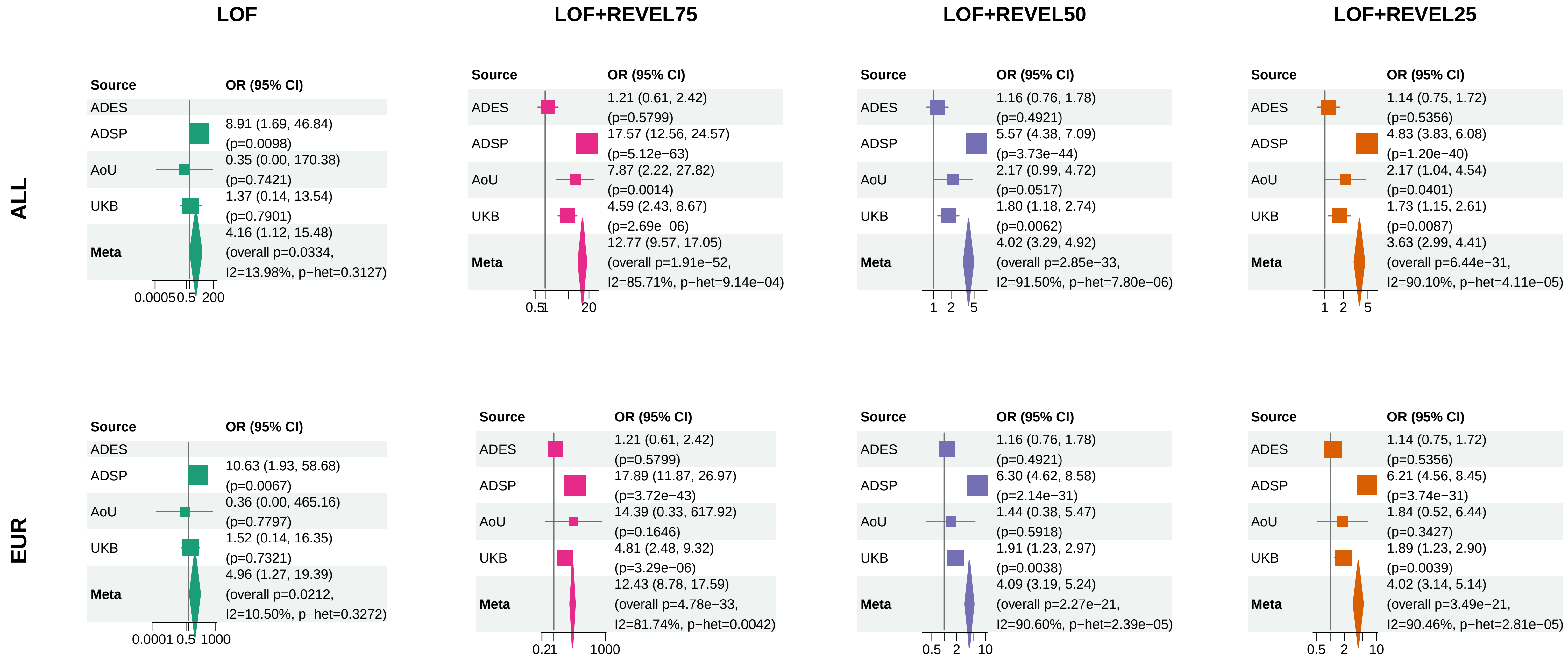


Meta OR/CI: pooled REGENIE effect; overall P: ADES+REGENIE signed−Z when the exact ADES mask was available, otherwise REGENIE.

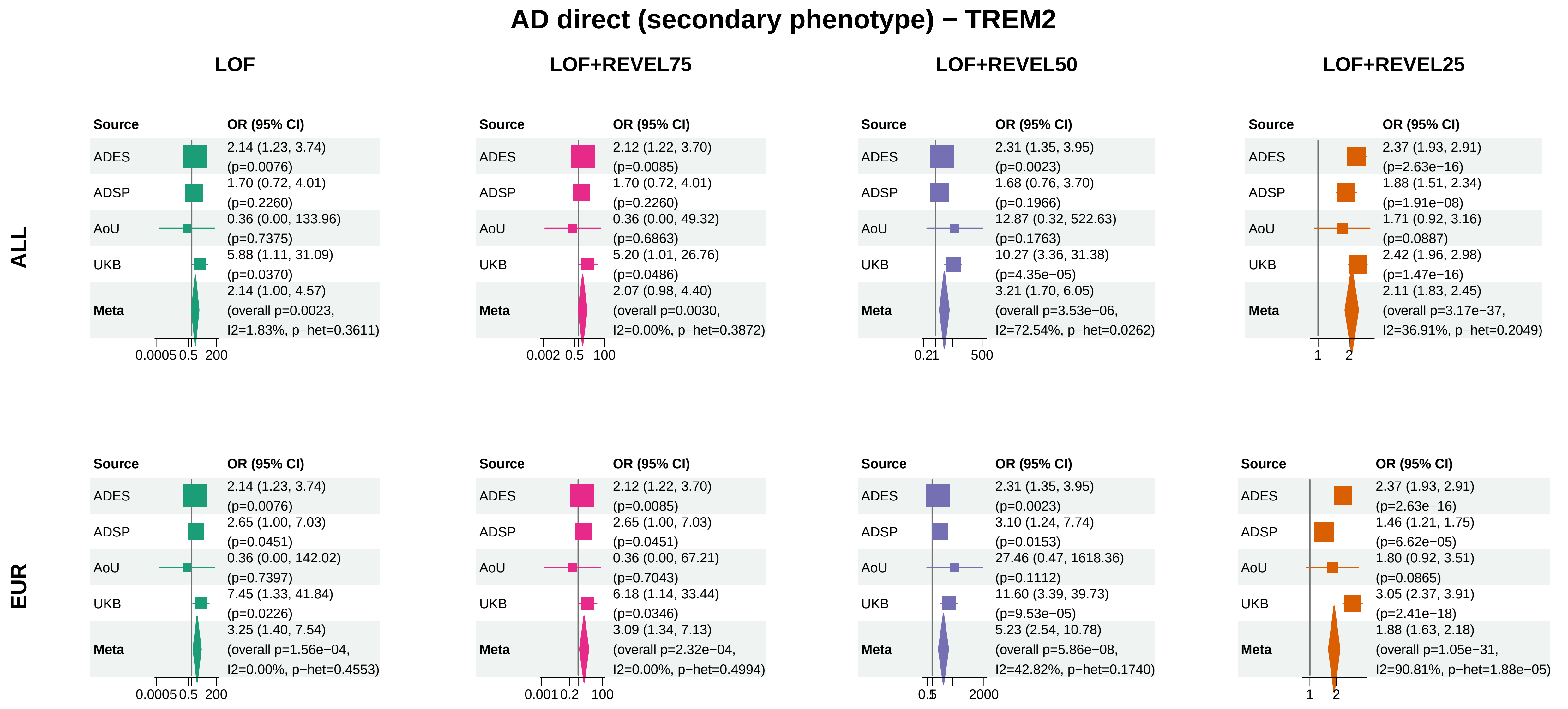


Meta OR/CI: pooled REGENIE effect; overall P: ADES+REGENIE signed−Z when the exact ADES mask was available, otherwise REGENIE.

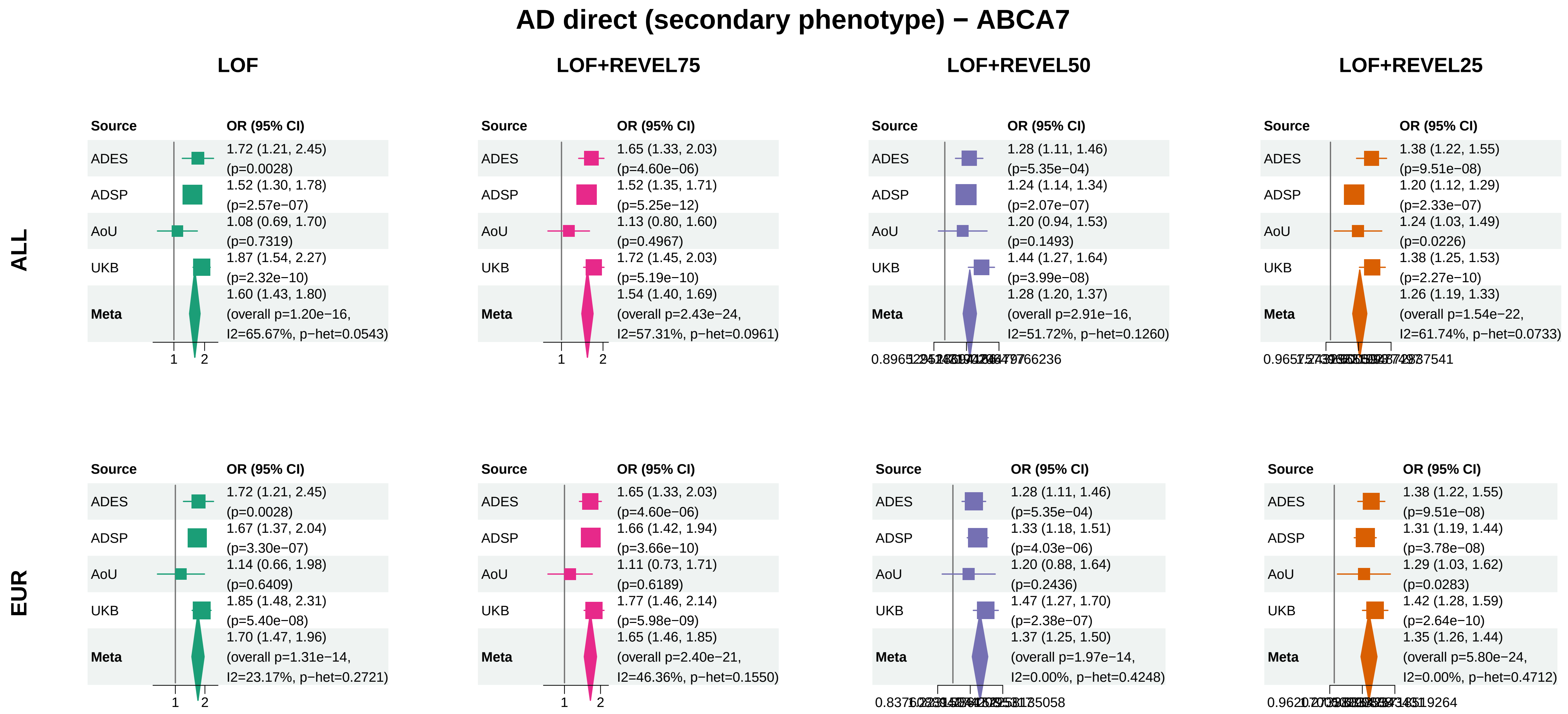


Meta OR/CI: pooled REGENIE effect; overall P: ADES+REGENIE signed−Z when the exact ADES mask was available, otherwise REGENIE.

### AD direct (secondary phenotype) − SORL1

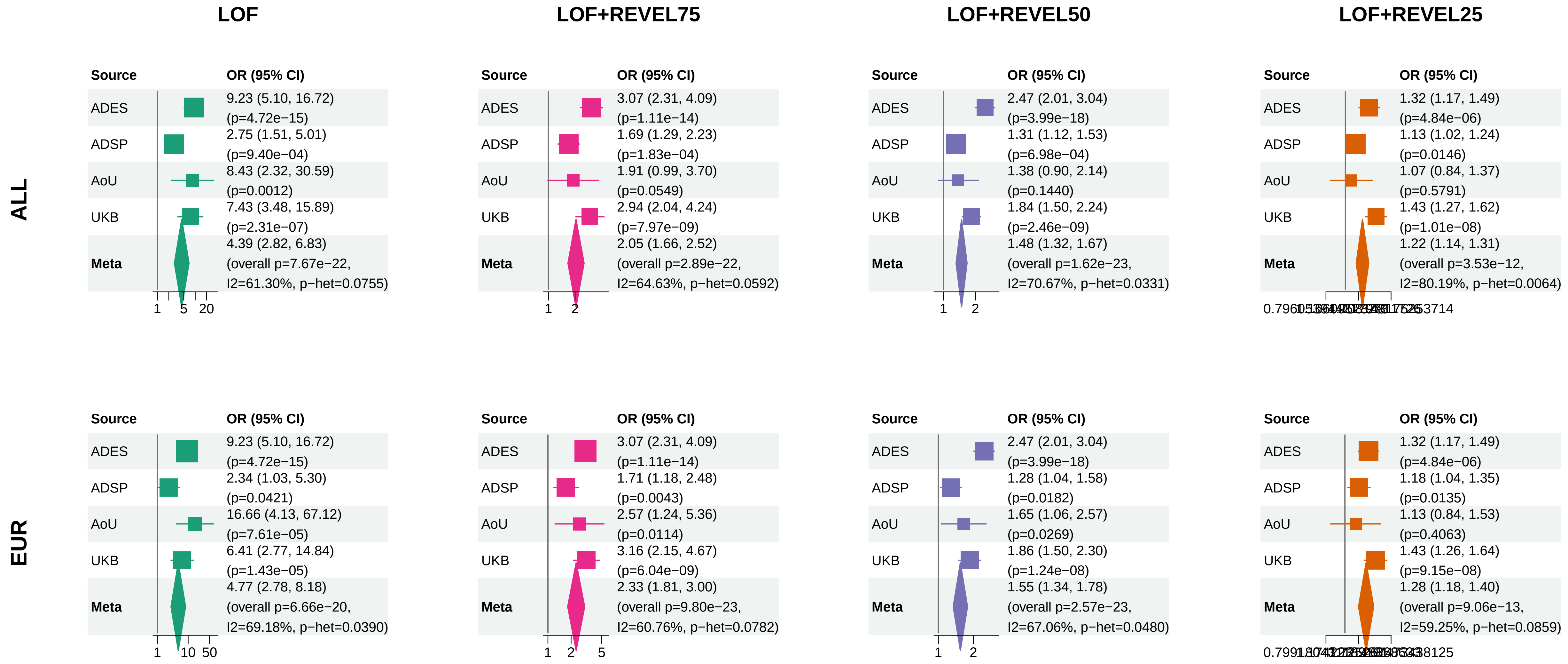


Meta OR/CI: pooled REGENIE effect; overall P: ADES+REGENIE signed−Z when the exact ADES mask was available, otherwise REGENIE.

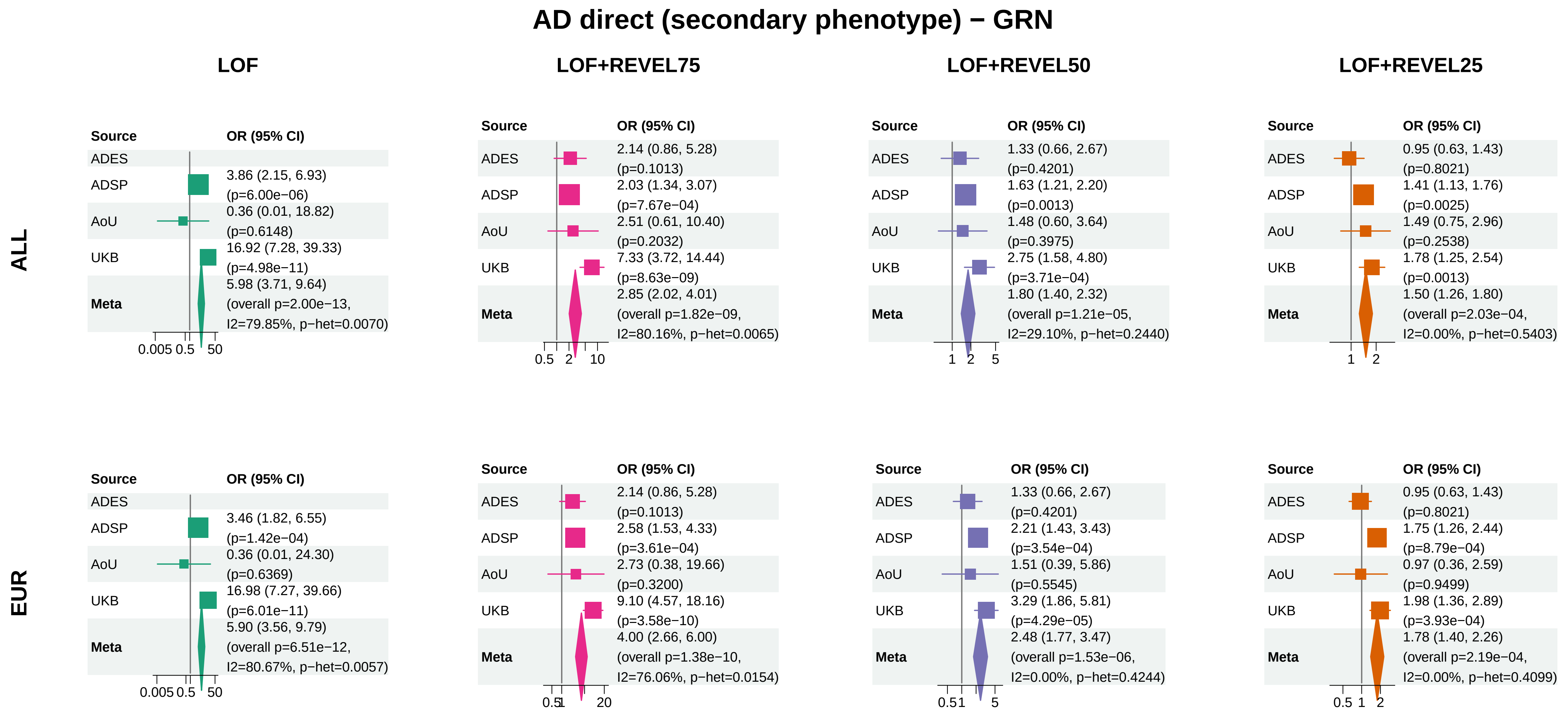


Meta OR/CI: pooled REGENIE effect; overall P: ADES+REGENIE signed−Z when the exact ADES mask was available, otherwise REGENIE.

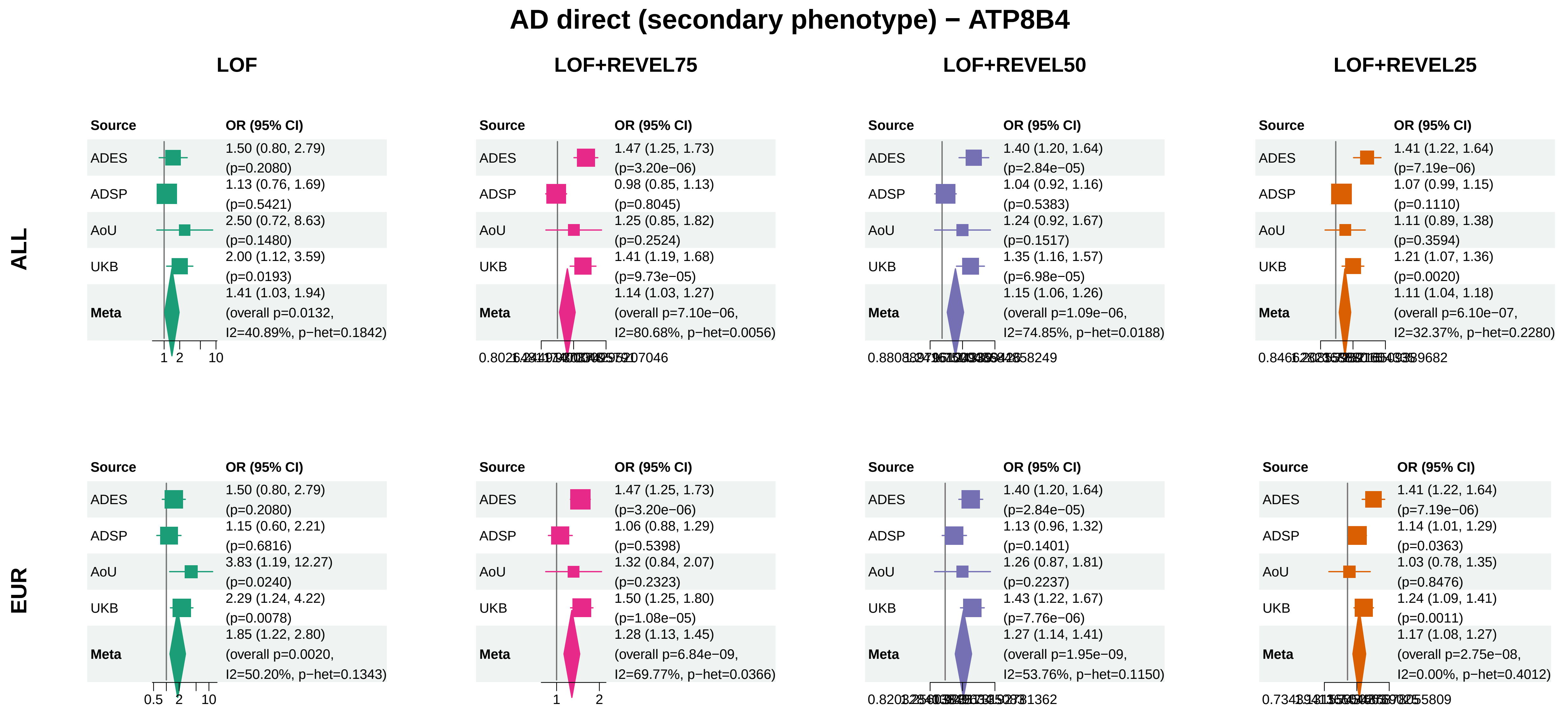


Meta OR/CI: pooled REGENIE effect; overall P: ADES+REGENIE signed−Z when the exact ADES mask was available, otherwise REGENIE.

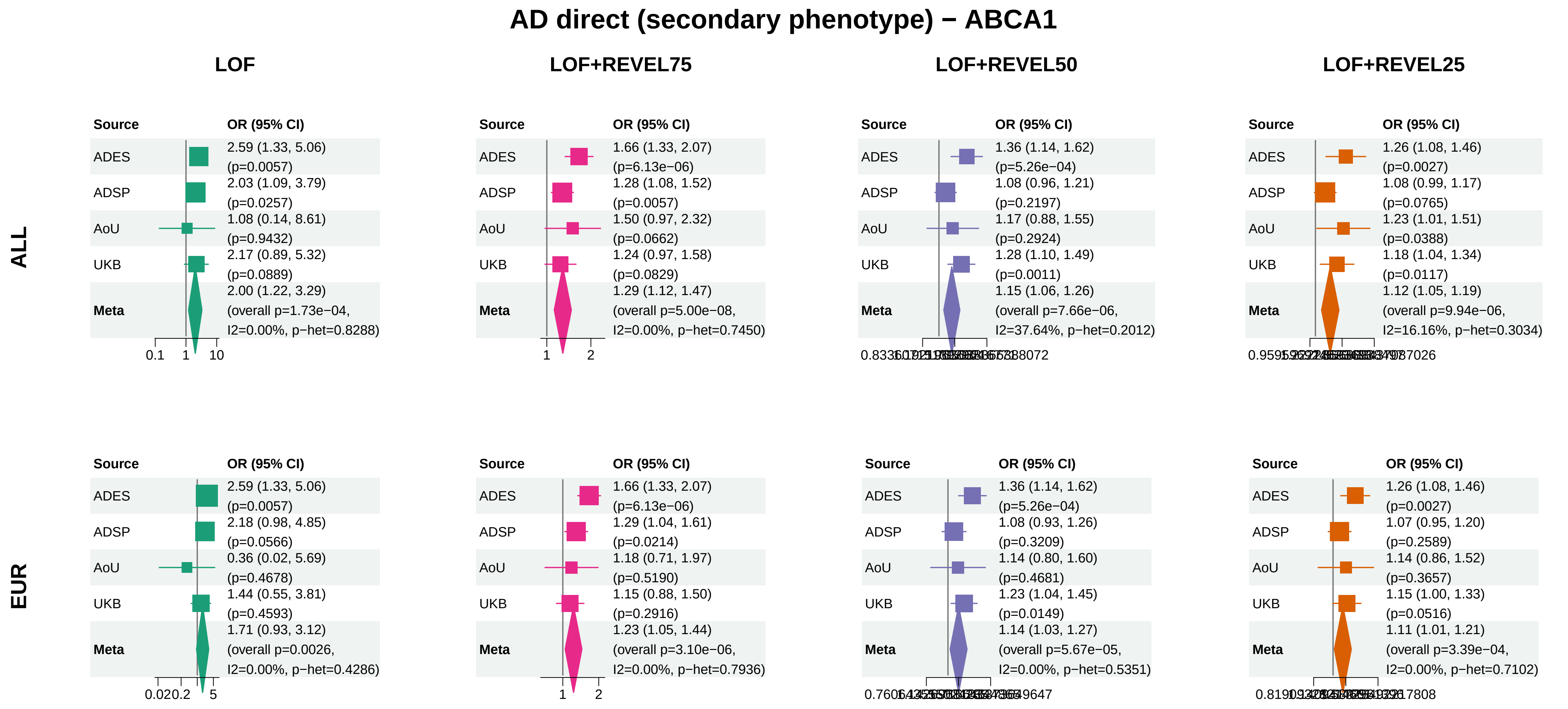


Meta OR/CI: pooled REGENIE effect; overall P: ADES+REGENIE signed−Z when the exact ADES mask was available, otherwise REGENIE.

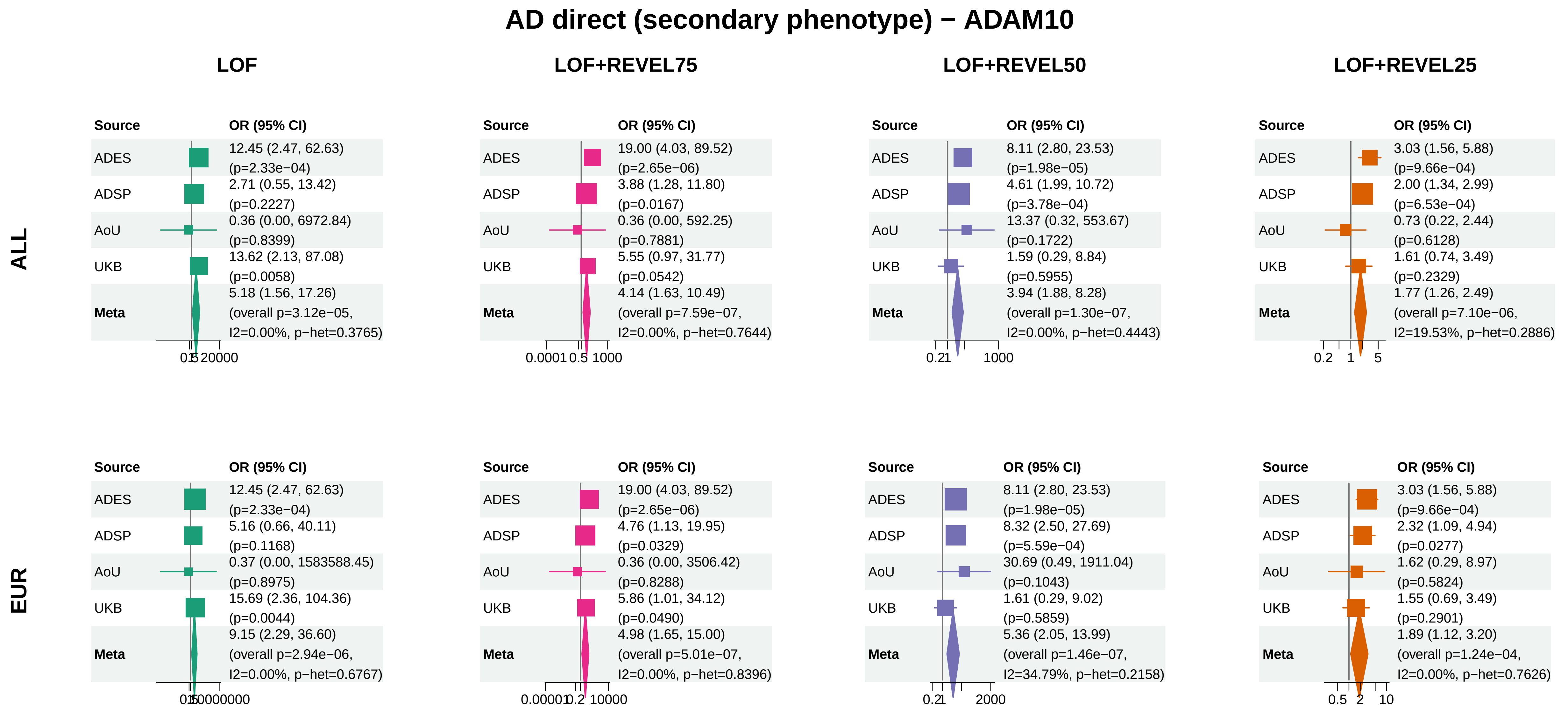


Meta OR/CI: pooled REGENIE effect; overall P: ADES+REGENIE signed−Z when the exact ADES mask was available, otherwise REGENIE.

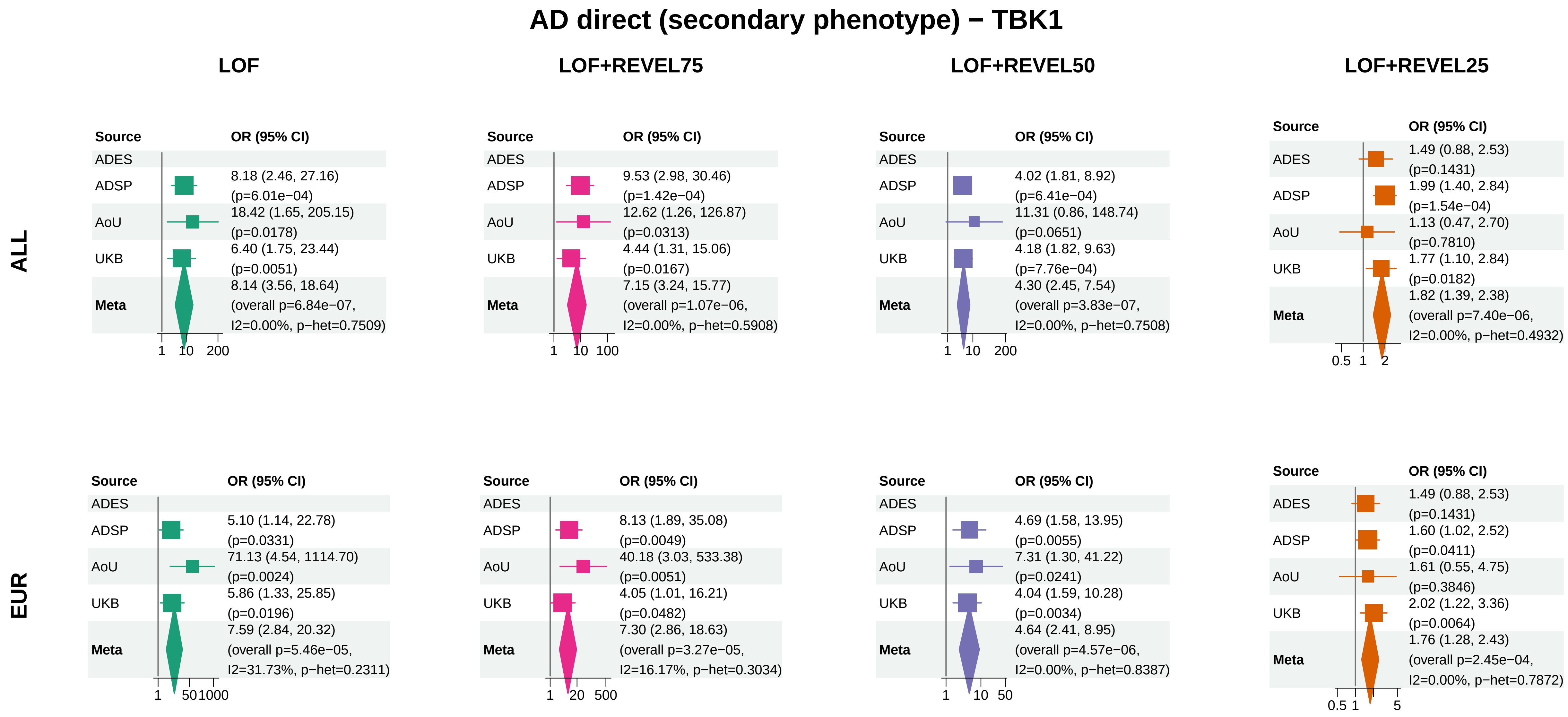


Meta OR/CI: pooled REGENIE effect; overall P: ADES+REGENIE signed−Z when the exact ADES mask was available, otherwise REGENIE.

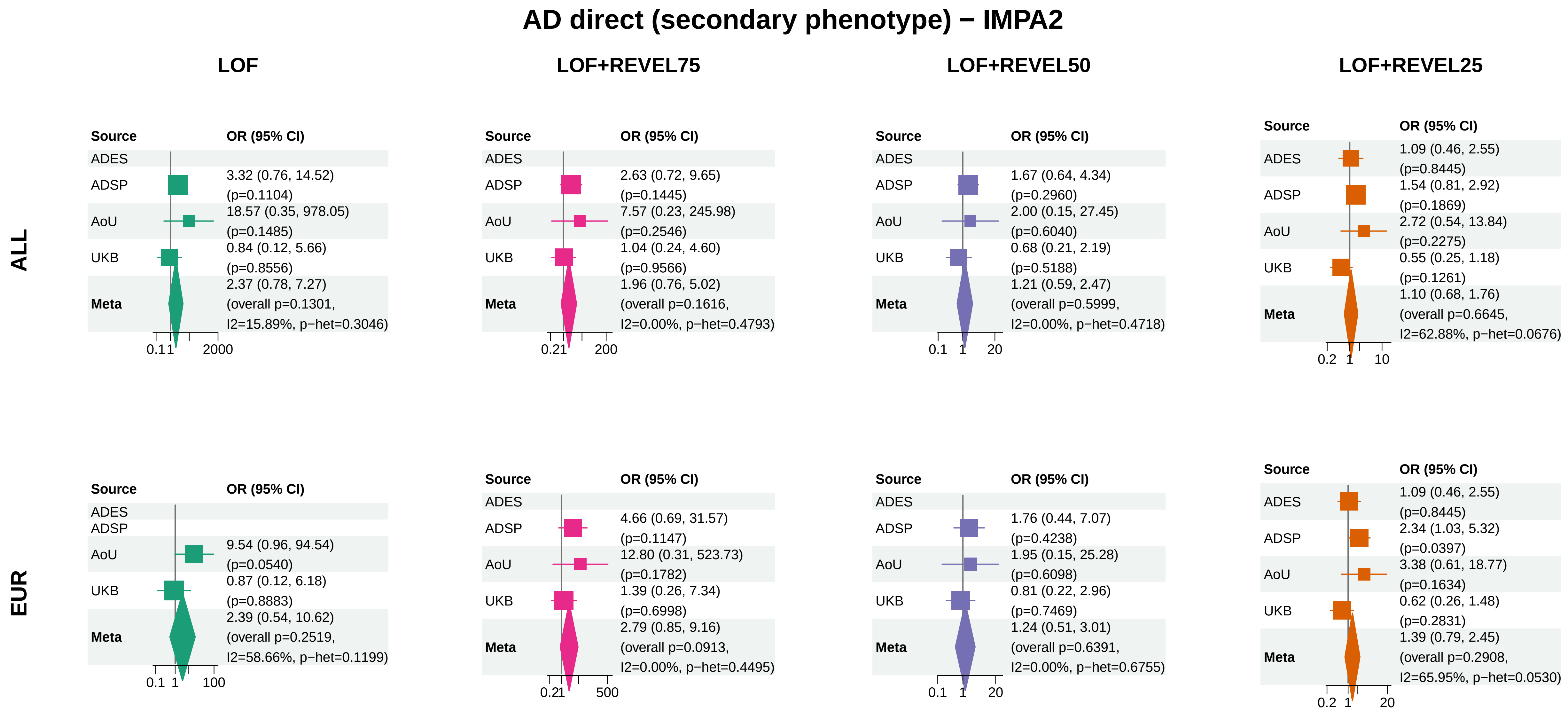


Meta OR/CI: pooled REGENIE effect; overall P: ADES+REGENIE signed−Z when the exact ADES mask was available, otherwise REGENIE.

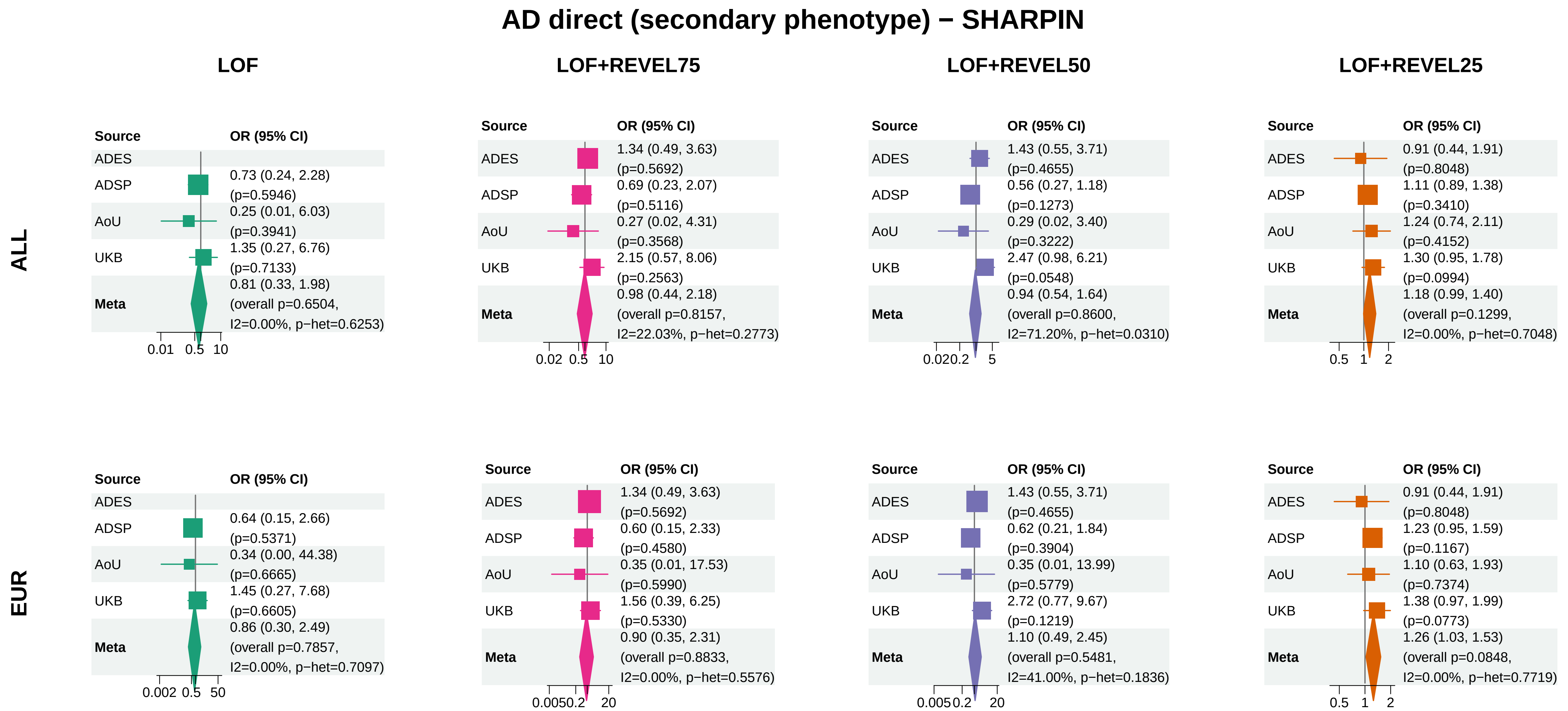


Meta OR/CI: pooled REGENIE effect; overall P: ADES+REGENIE signed−Z when the exact ADES mask was available, otherwise REGENIE.

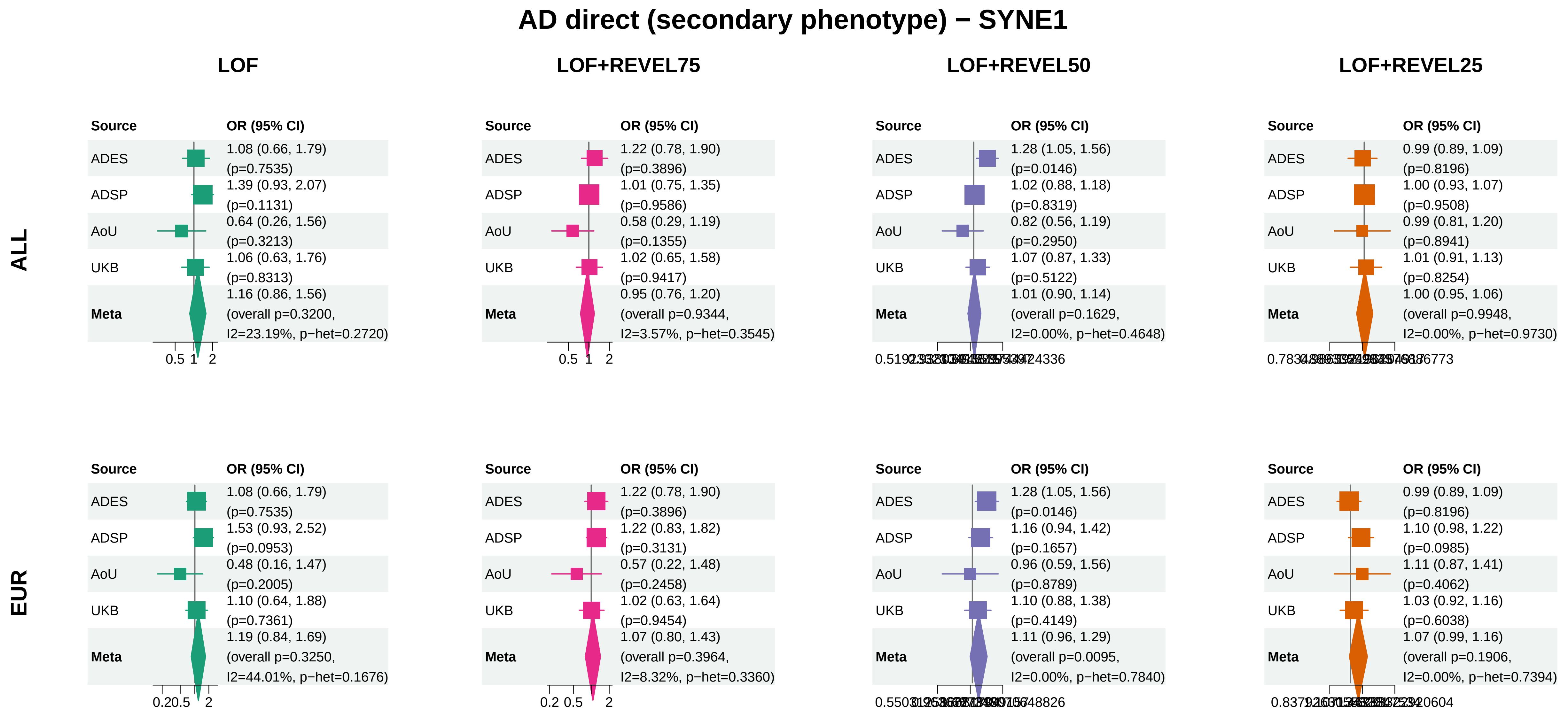


Meta OR/CI: pooled REGENIE effect; overall P: ADES+REGENIE signed−Z when the exact ADES mask was available, otherwise REGENIE.

### AD direct (secondary phenotype) − APOE

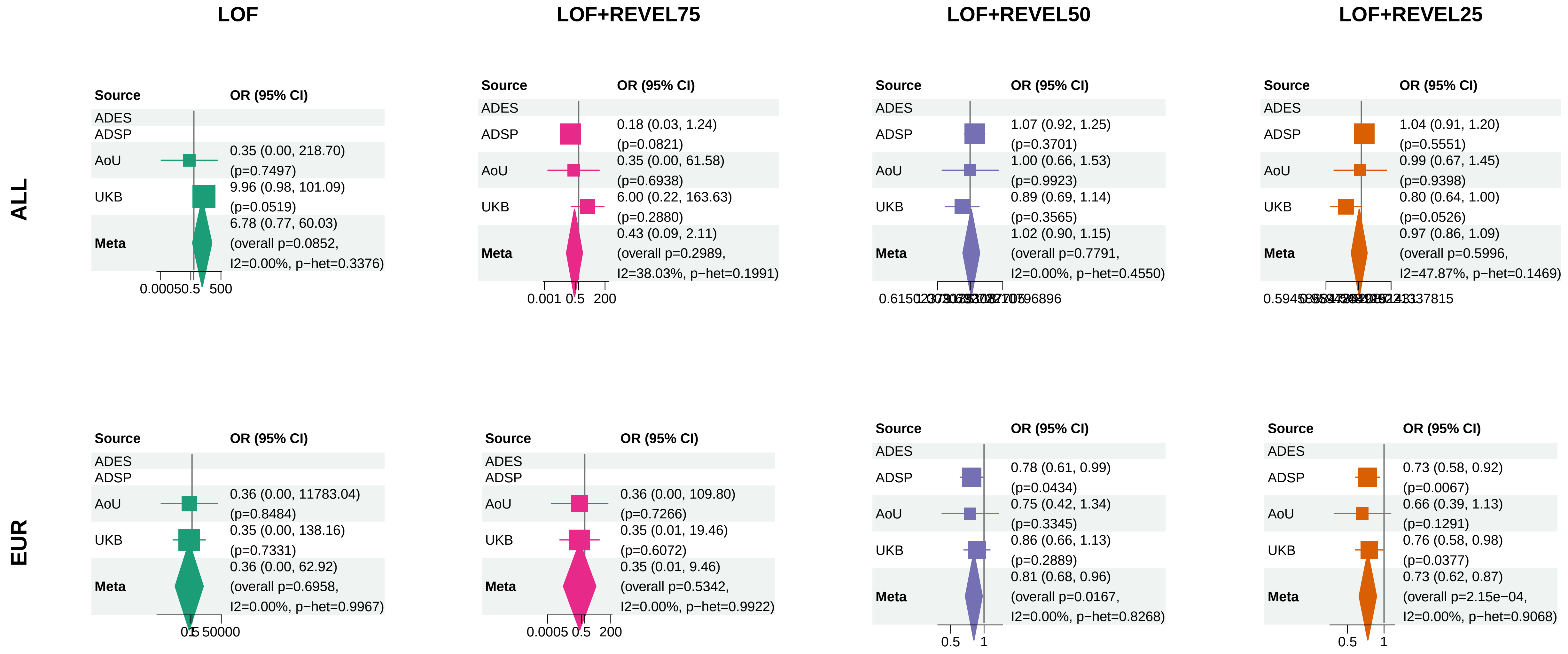


Meta OR/CI: pooled REGENIE effect; overall P: ADES+REGENIE signed−Z when the exact ADES mask was available, otherwise REGENIE.

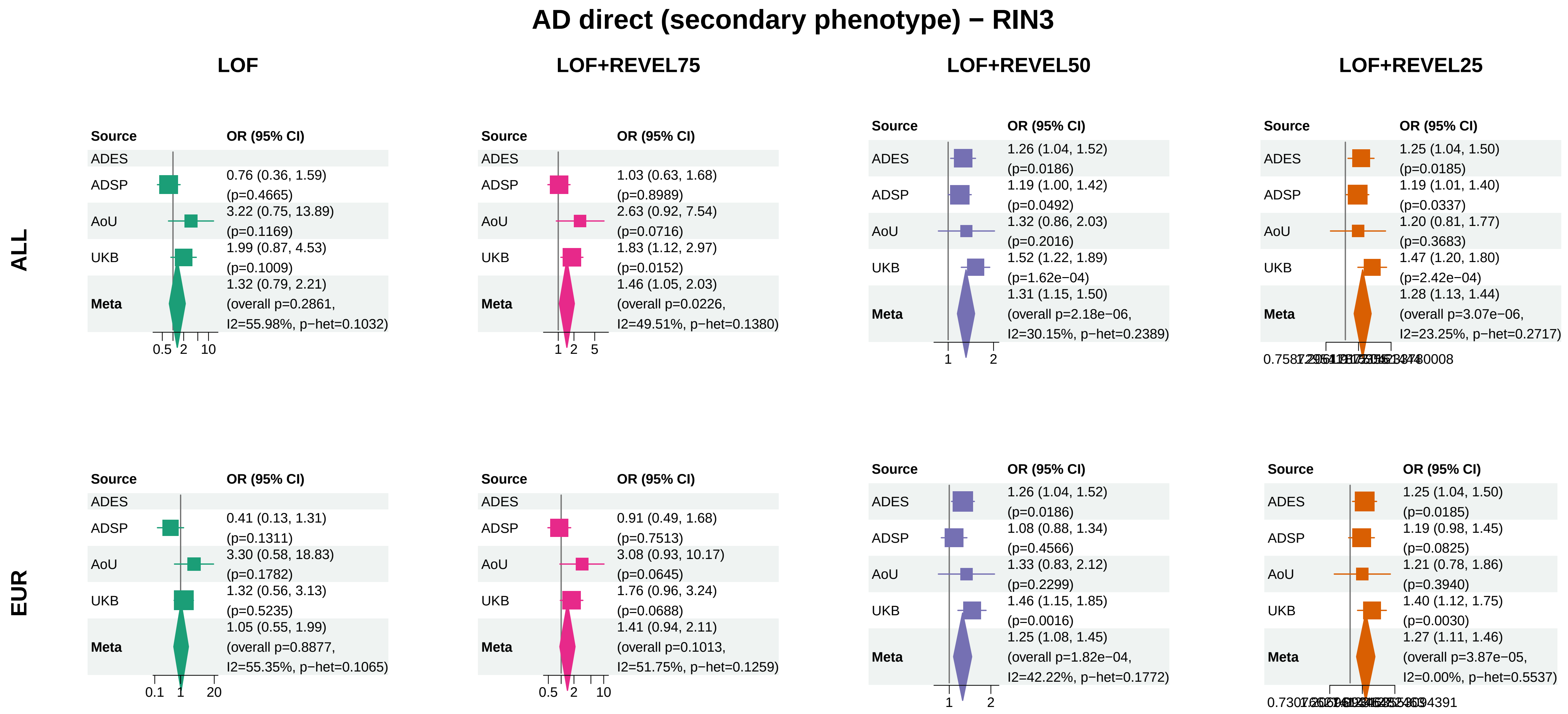


Meta OR/CI: pooled REGENIE effect; overall P: ADES+REGENIE signed−Z when the exact ADES mask was available, otherwise REGENIE.

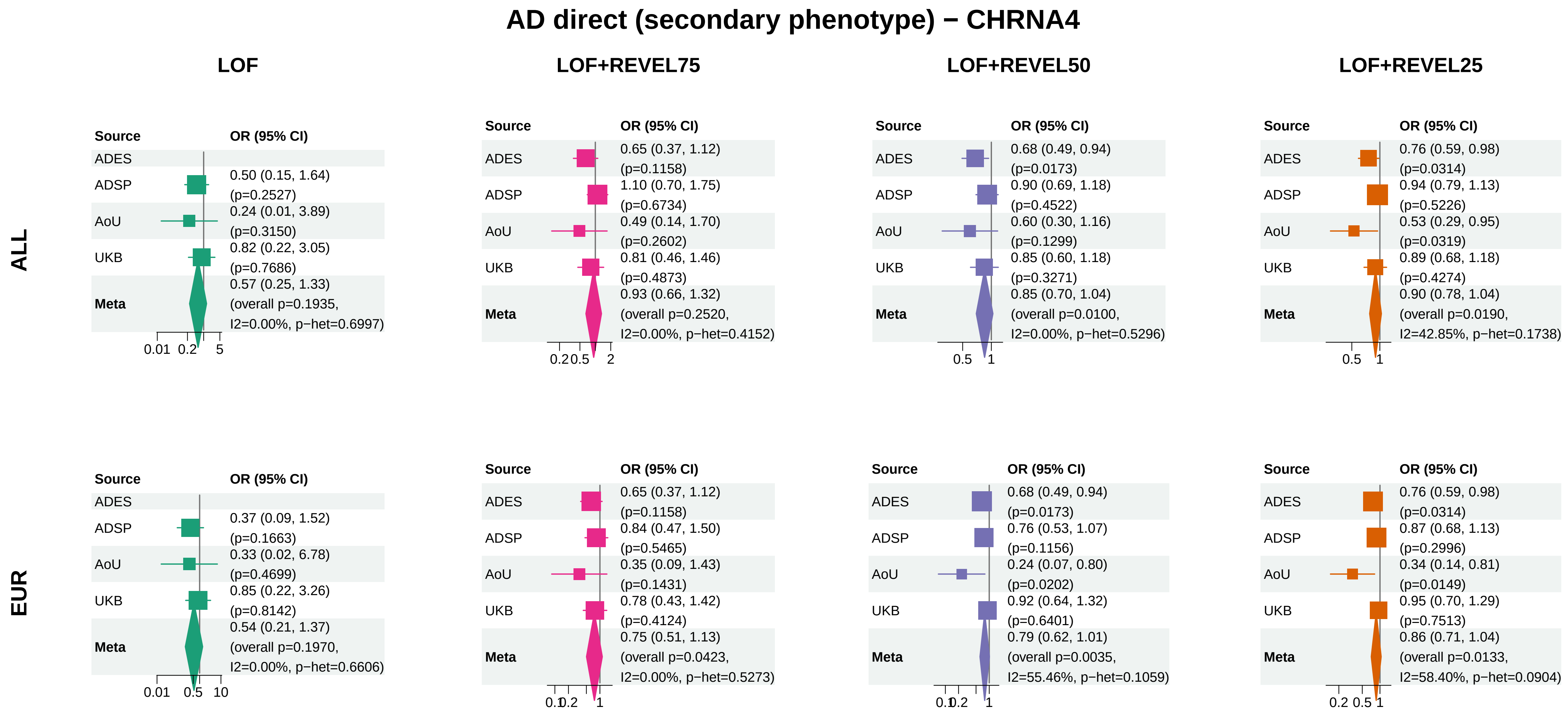


Meta OR/CI: pooled REGENIE effect; overall P: ADES+REGENIE signed−Z when the exact ADES mask was available, otherwise REGENIE.

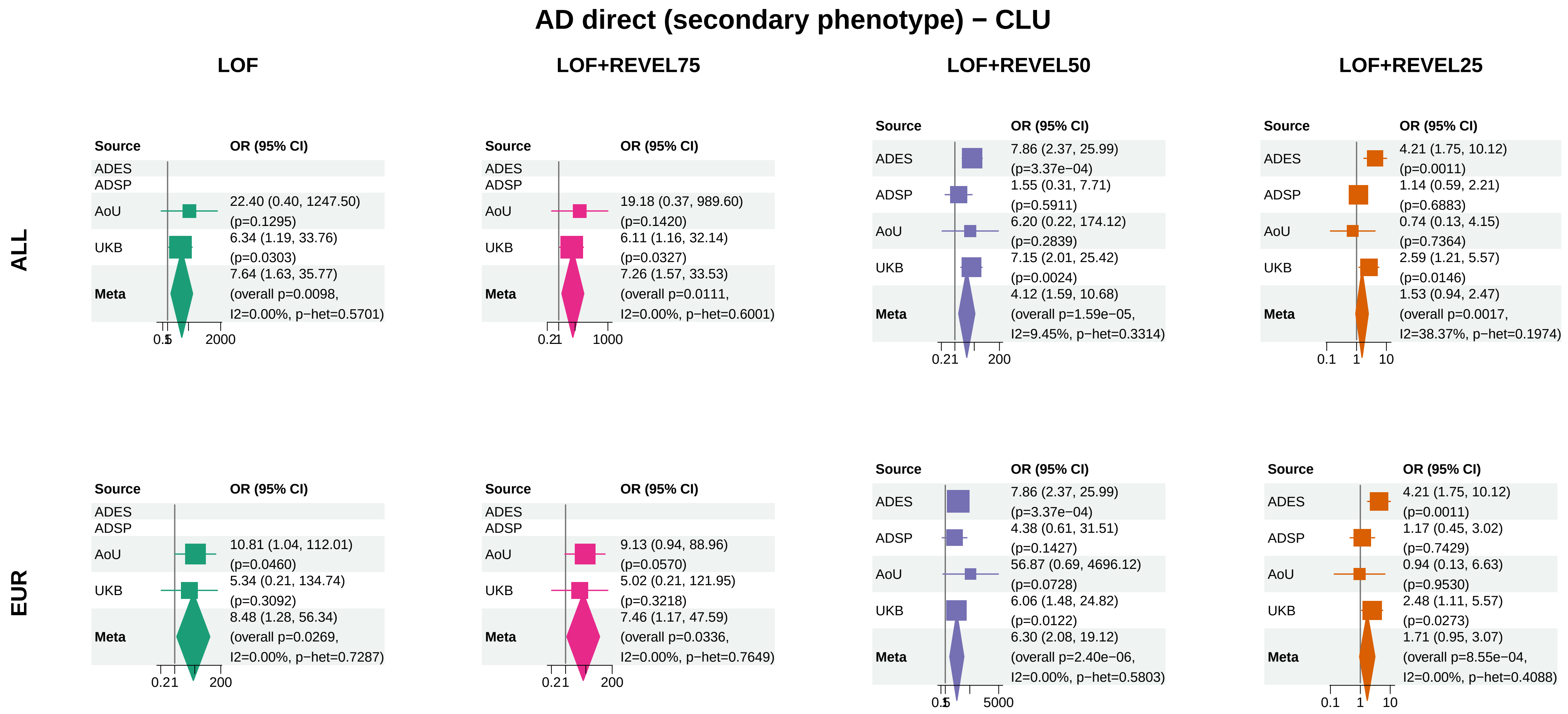


Meta OR/CI: pooled REGENIE effect; overall P: ADES+REGENIE signed−Z when the exact ADES mask was available, otherwise REGENIE.

### AD direct (secondary phenotype) − SP140L

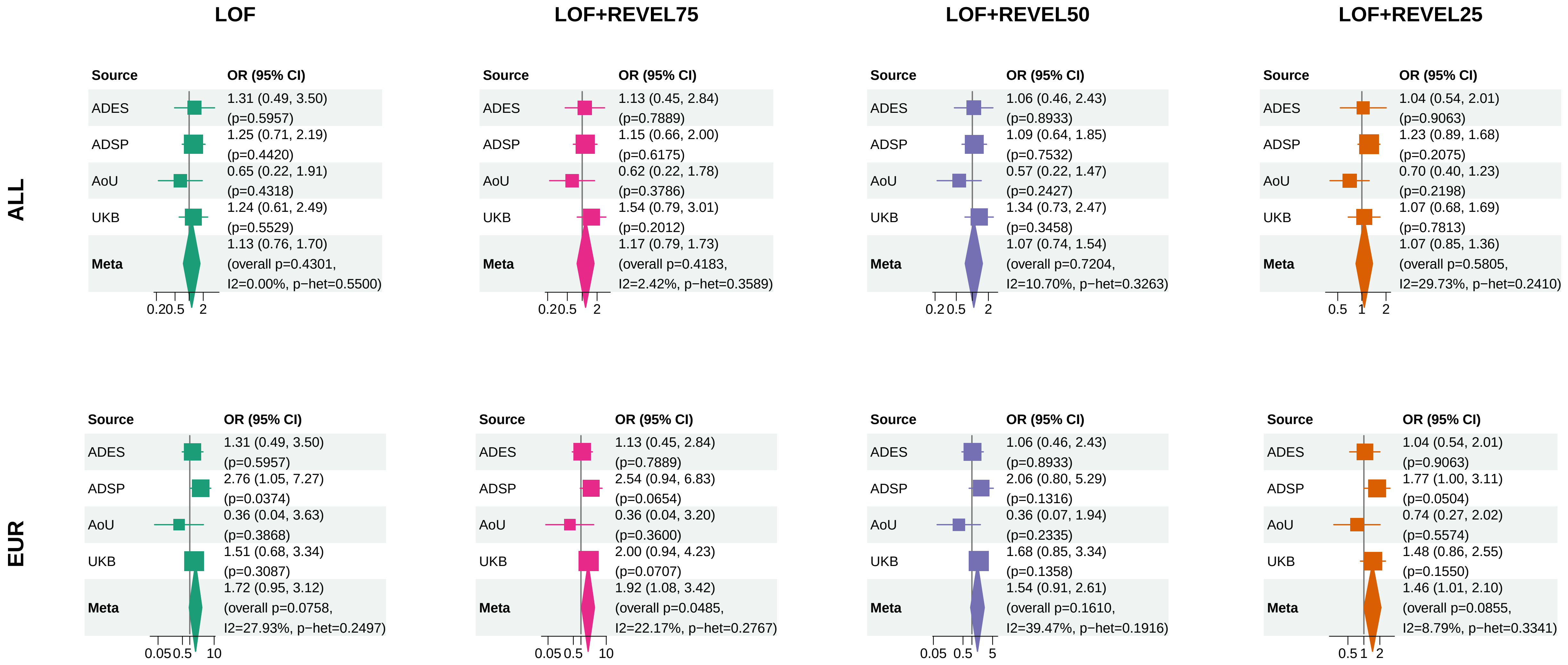


Meta OR/CI: pooled REGENIE effect; overall P: ADES+REGENIE signed−Z when the exact ADES mask was available, otherwise REGENIE.

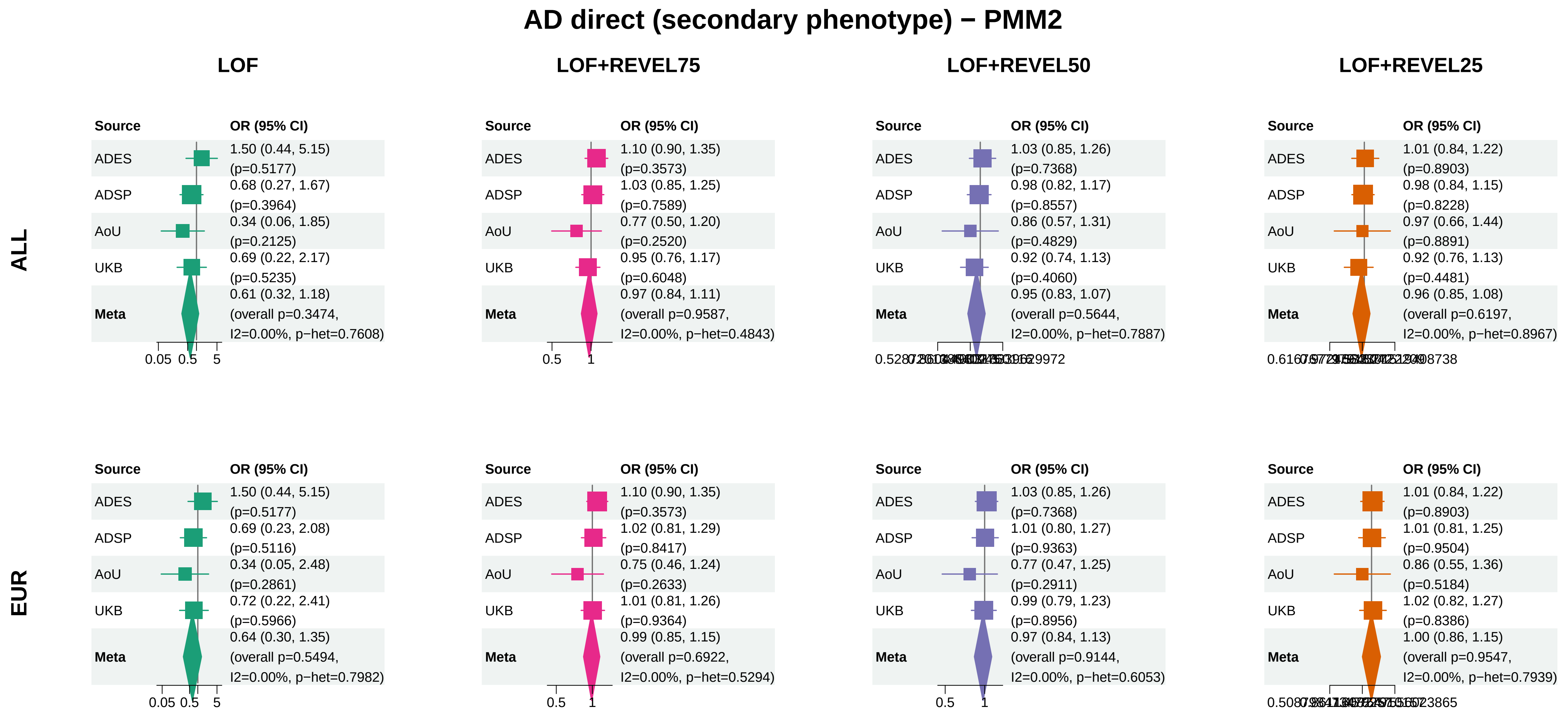


Meta OR/CI: pooled REGENIE effect; overall P: ADES+REGENIE signed−Z when the exact ADES mask was available, otherwise REGENIE.

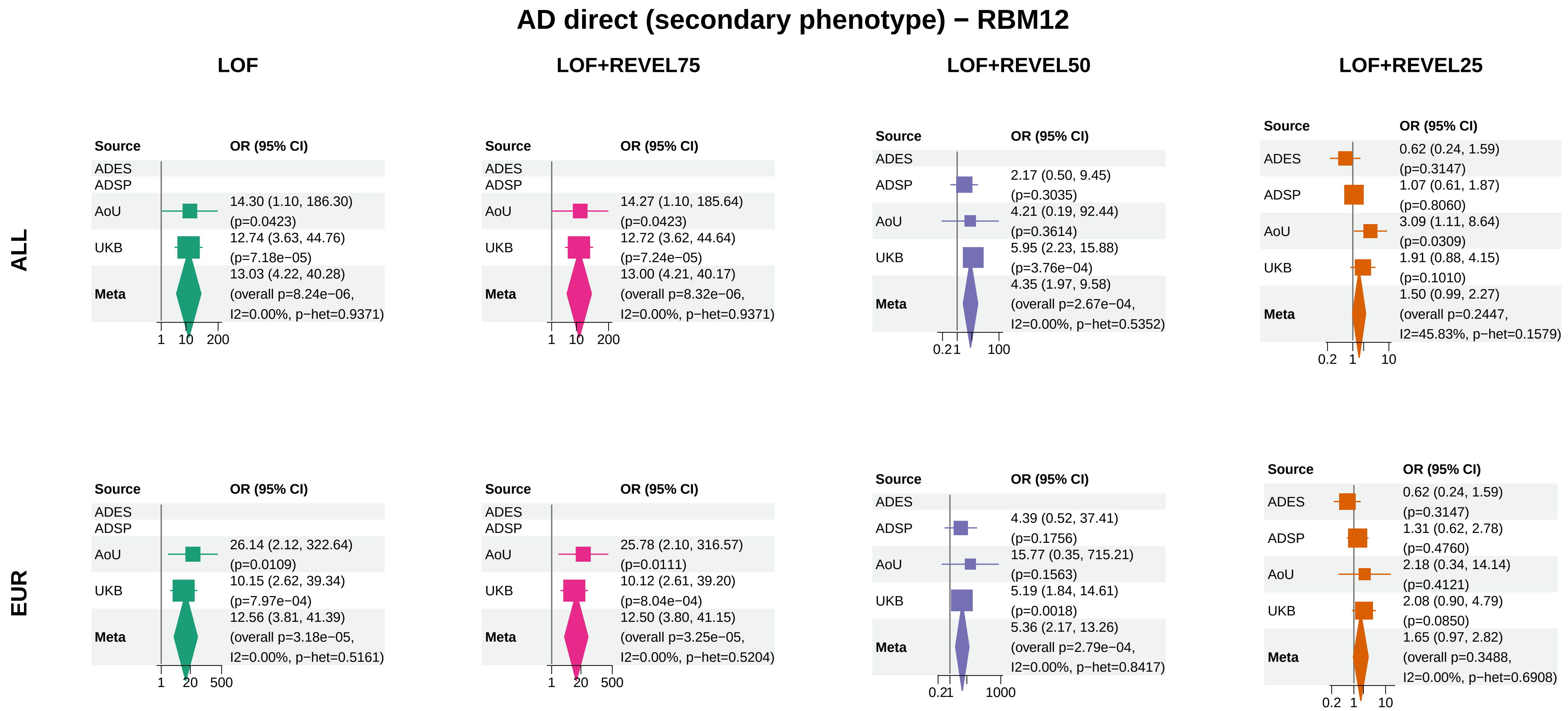


Meta OR/CI: pooled REGENIE effect; overall P: ADES+REGENIE signed−Z when the exact ADES mask was available, otherwise REGENIE.

### AD direct (secondary phenotype) − ZNF256

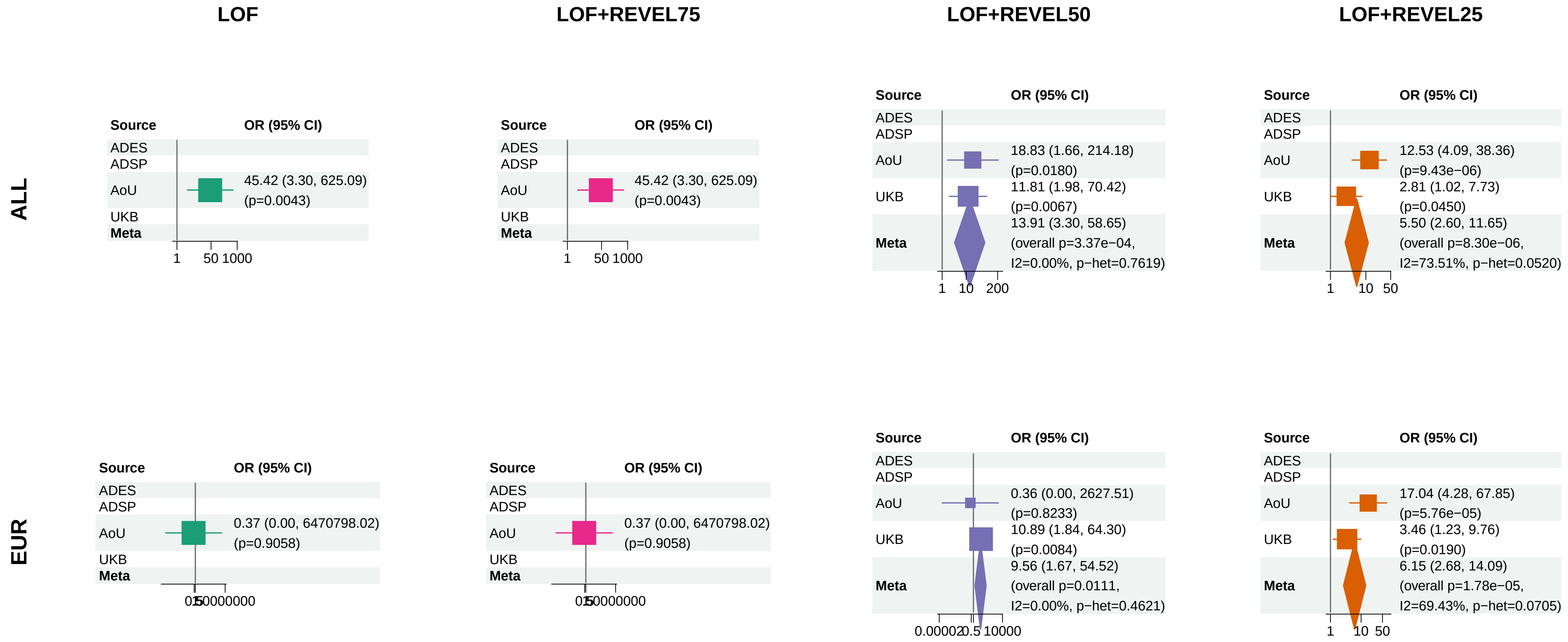


Meta OR/CI: pooled REGENIE effect; overall P: ADES+REGENIE signed−Z when the exact ADES mask was available, otherwise REGENIE.

### AD direct (secondary phenotype) − CCL7

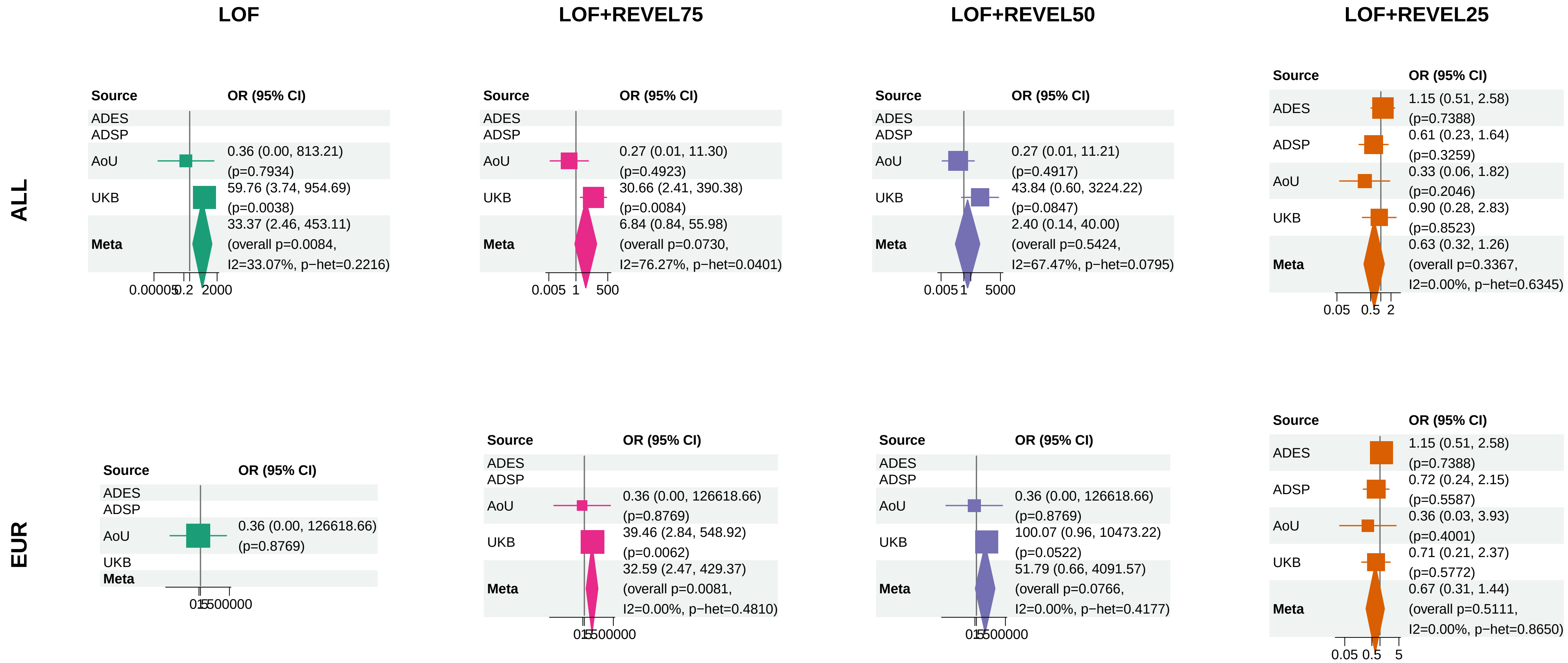


Meta OR/CI: pooled REGENIE effect; overall P: ADES+REGENIE signed−Z when the exact ADES mask was available, otherwise REGENIE.

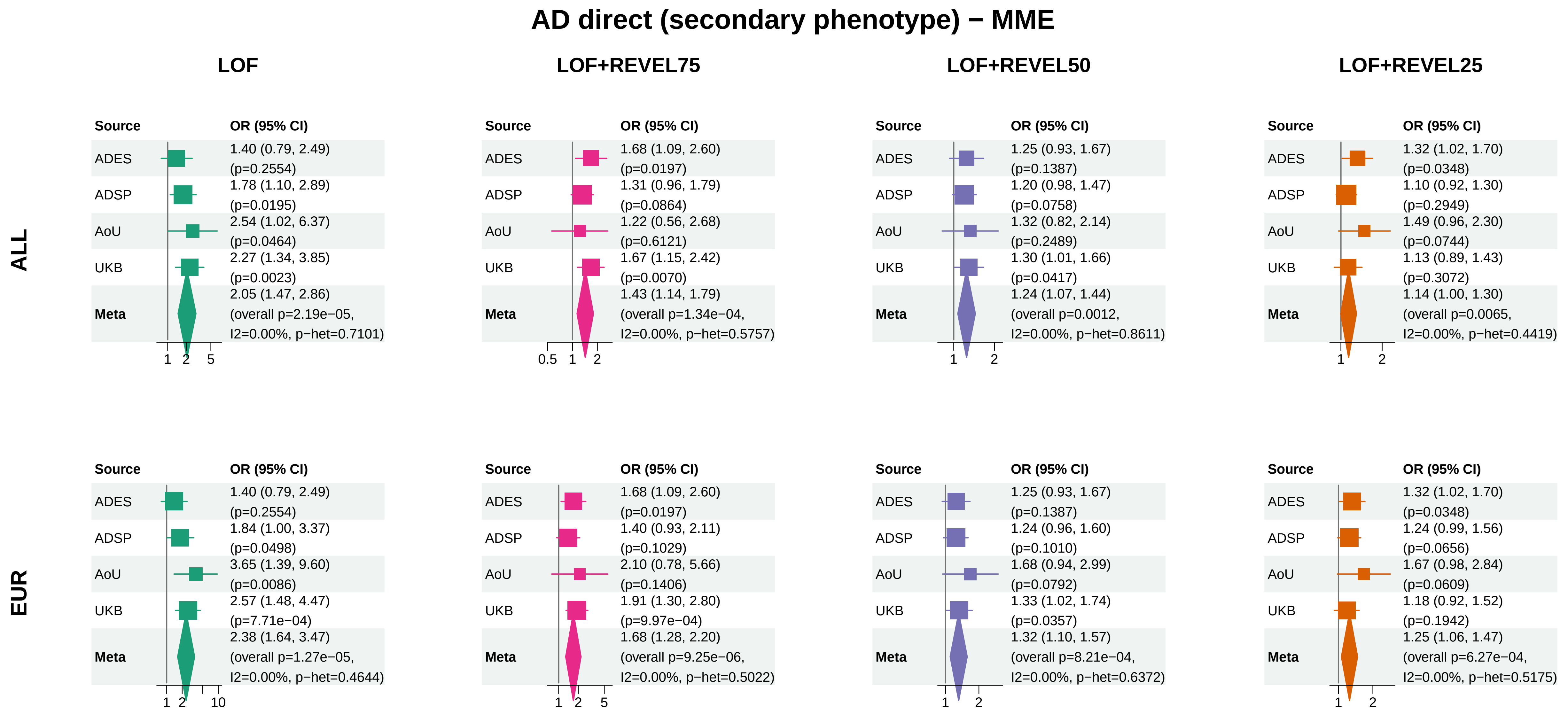


Meta OR/CI: pooled REGENIE effect; overall P: ADES+REGENIE signed−Z when the exact ADES mask was available, otherwise REGENIE.
