## Supplementary_Fig2 for "Population-scale burden analysis of rare damaging coding variants identifies novel risk genes for Alzheimer’s disease and related dementias and Parkinson’s disease and related disorders"

### AD combined (primary phenotype) − PSEN1

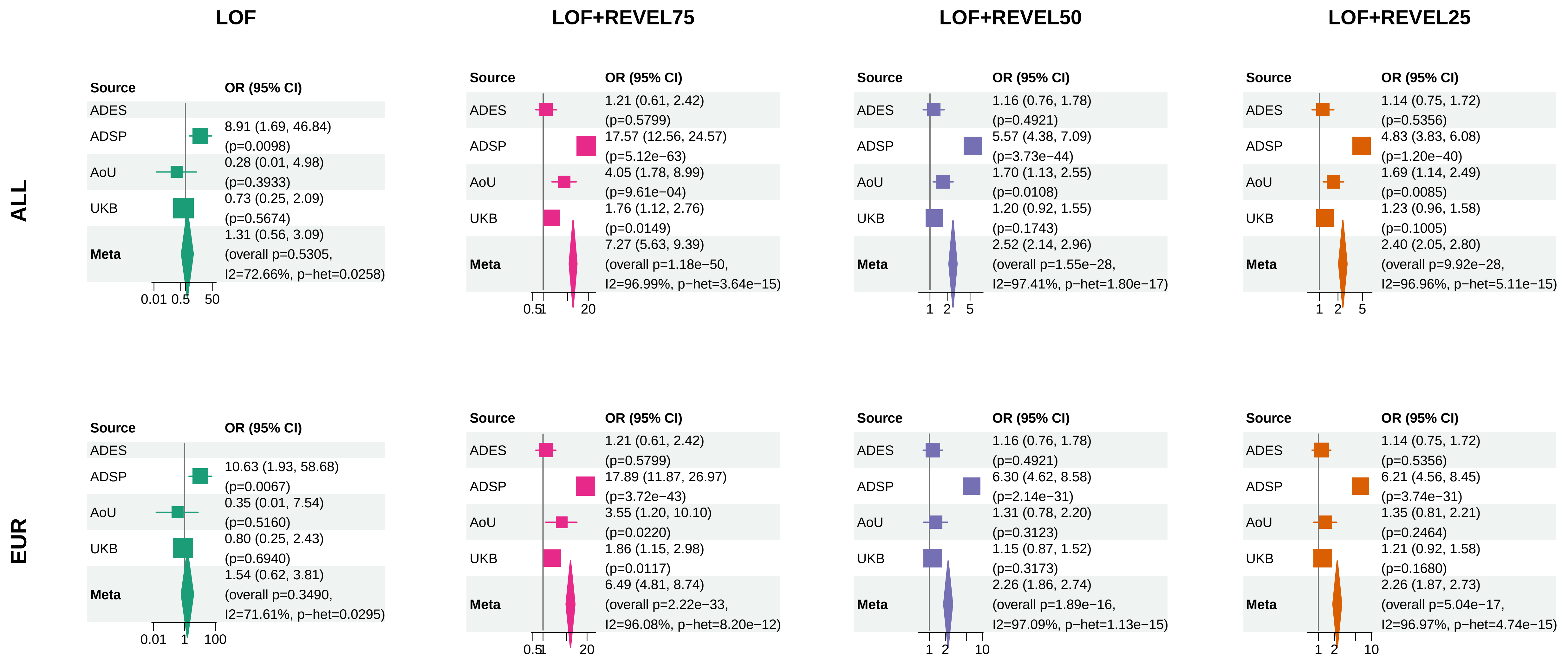


Meta OR/CI: pooled REGENIE effect; overall P: ADES+REGENIE signed−Z when the exact ADES mask was available, otherwise REGENIE.

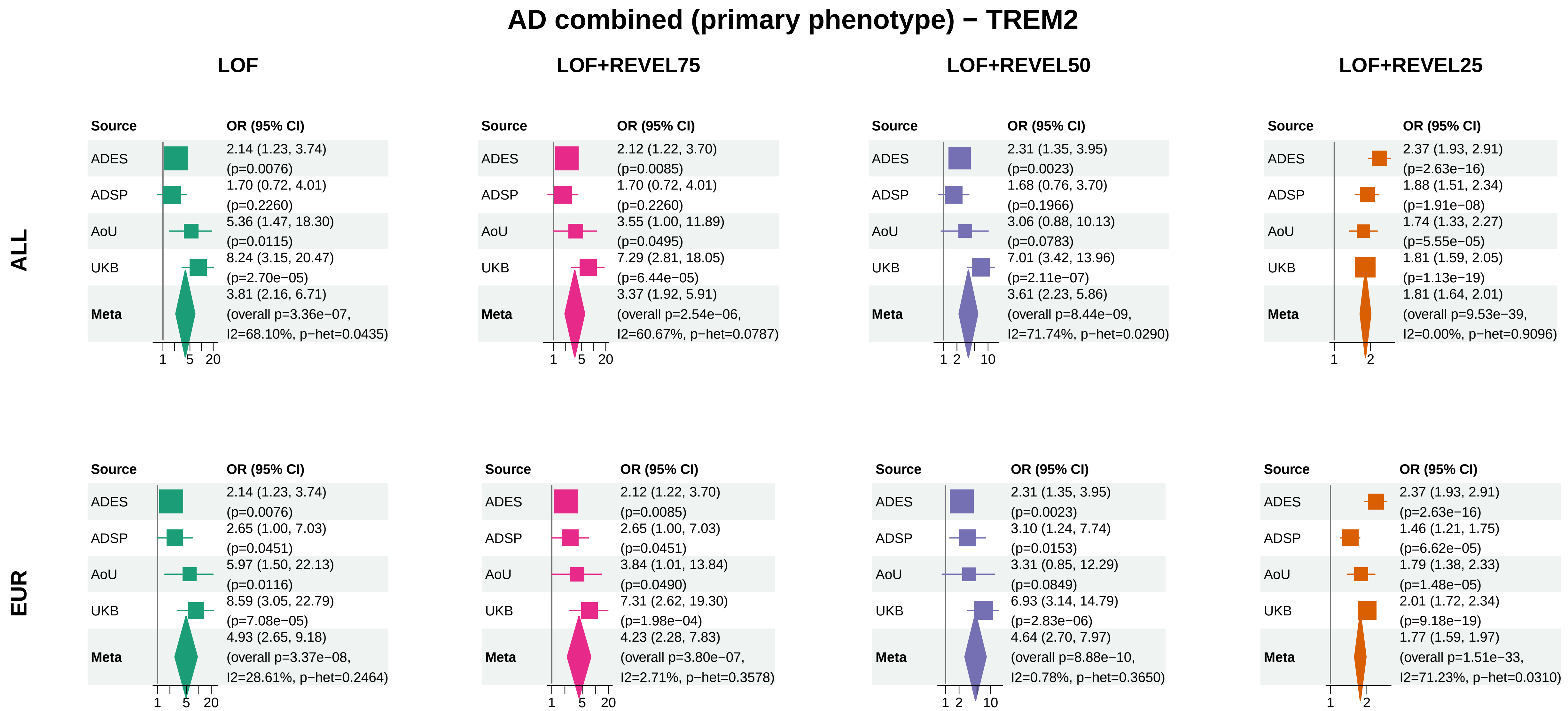


Meta OR/CI: pooled REGENIE effect; overall P: ADES+REGENIE signed−Z when the exact ADES mask was available, otherwise REGENIE.

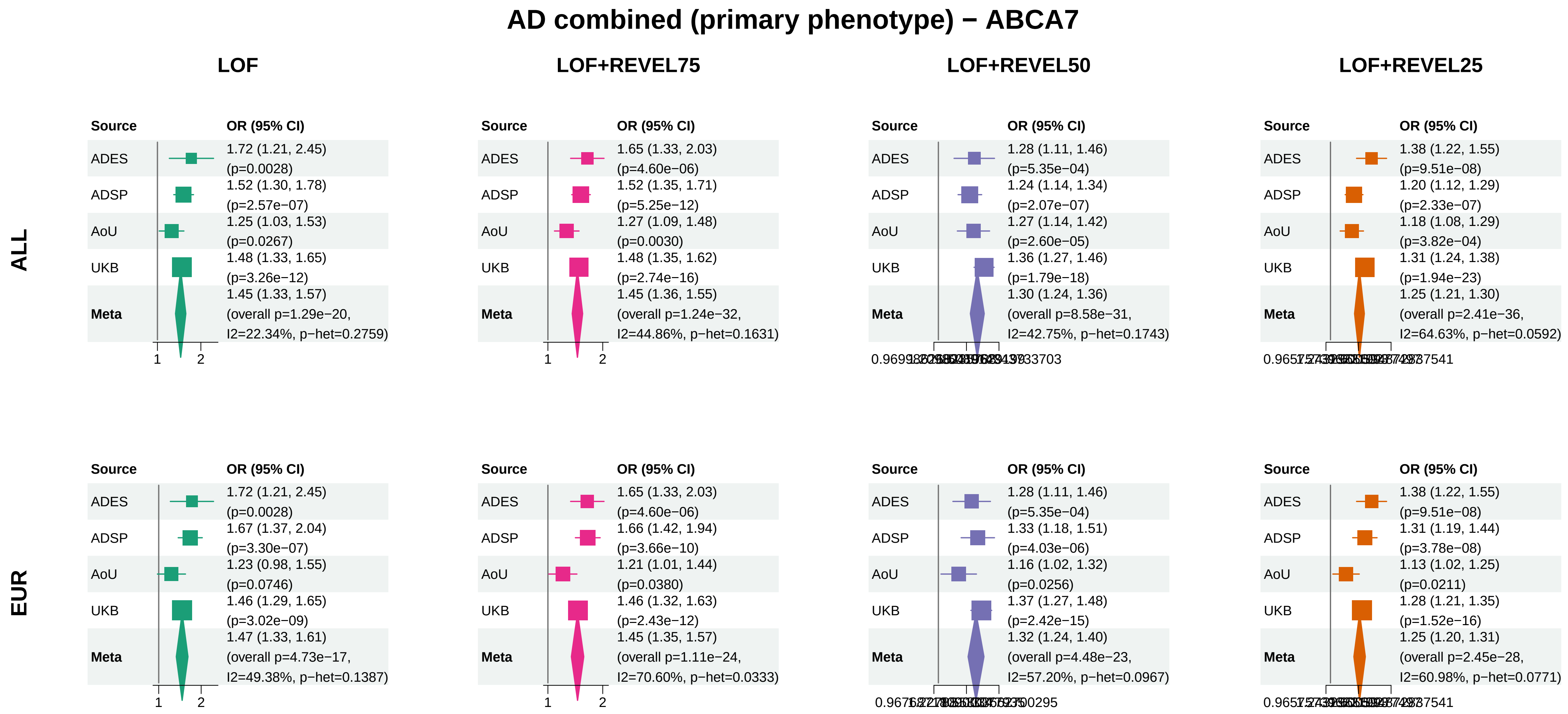


Meta OR/CI: pooled REGENIE effect; overall P: ADES+REGENIE signed−Z when the exact ADES mask was available, otherwise REGENIE.

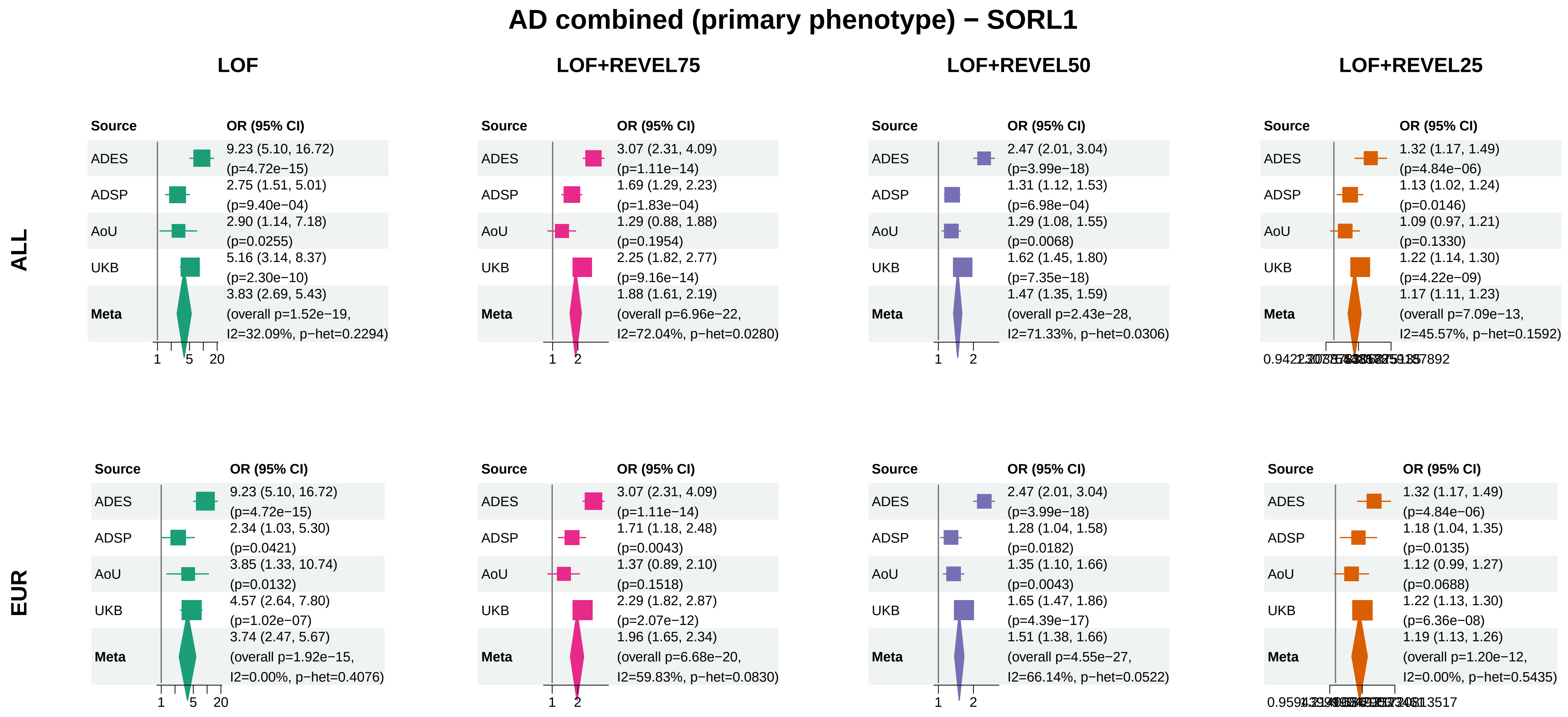


Meta OR/CI: pooled REGENIE effect; overall P: ADES+REGENIE signed−Z when the exact ADES mask was available, otherwise REGENIE.

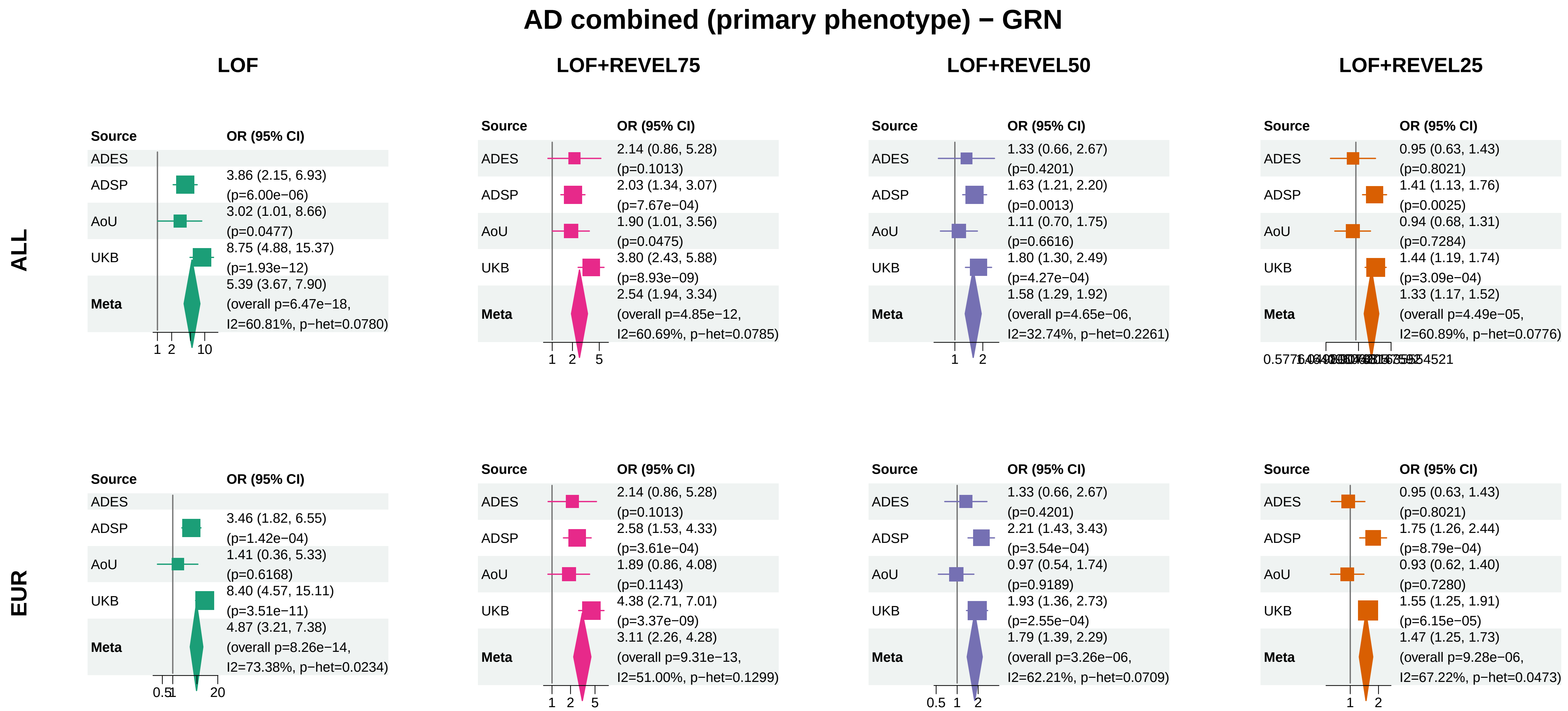


Meta OR/CI: pooled REGENIE effect; overall P: ADES+REGENIE signed−Z when the exact ADES mask was available, otherwise REGENIE.

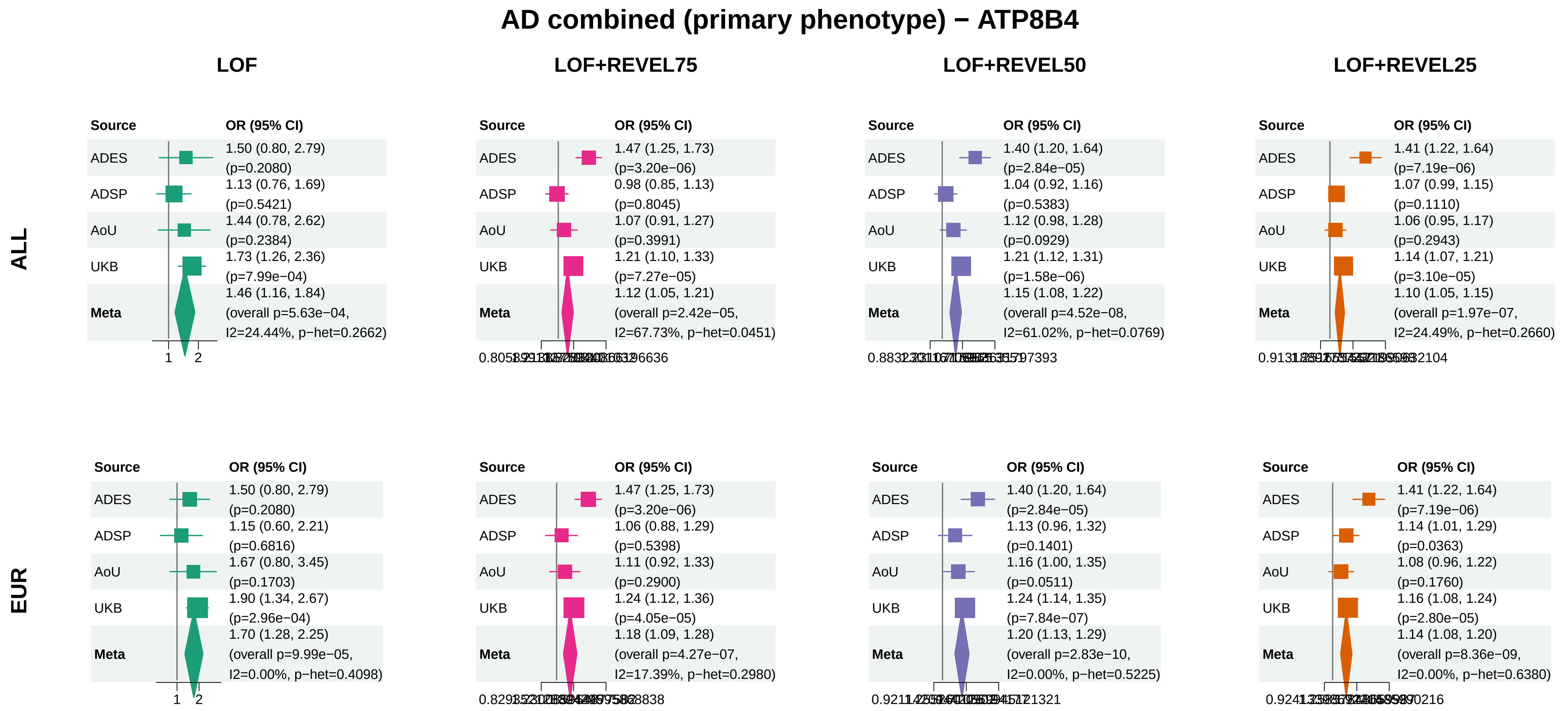


Meta OR/CI: pooled REGENIE effect; overall P: ADES+REGENIE signed−Z when the exact ADES mask was available, otherwise REGENIE.

### AD combined (primary phenotype) − ABCA1

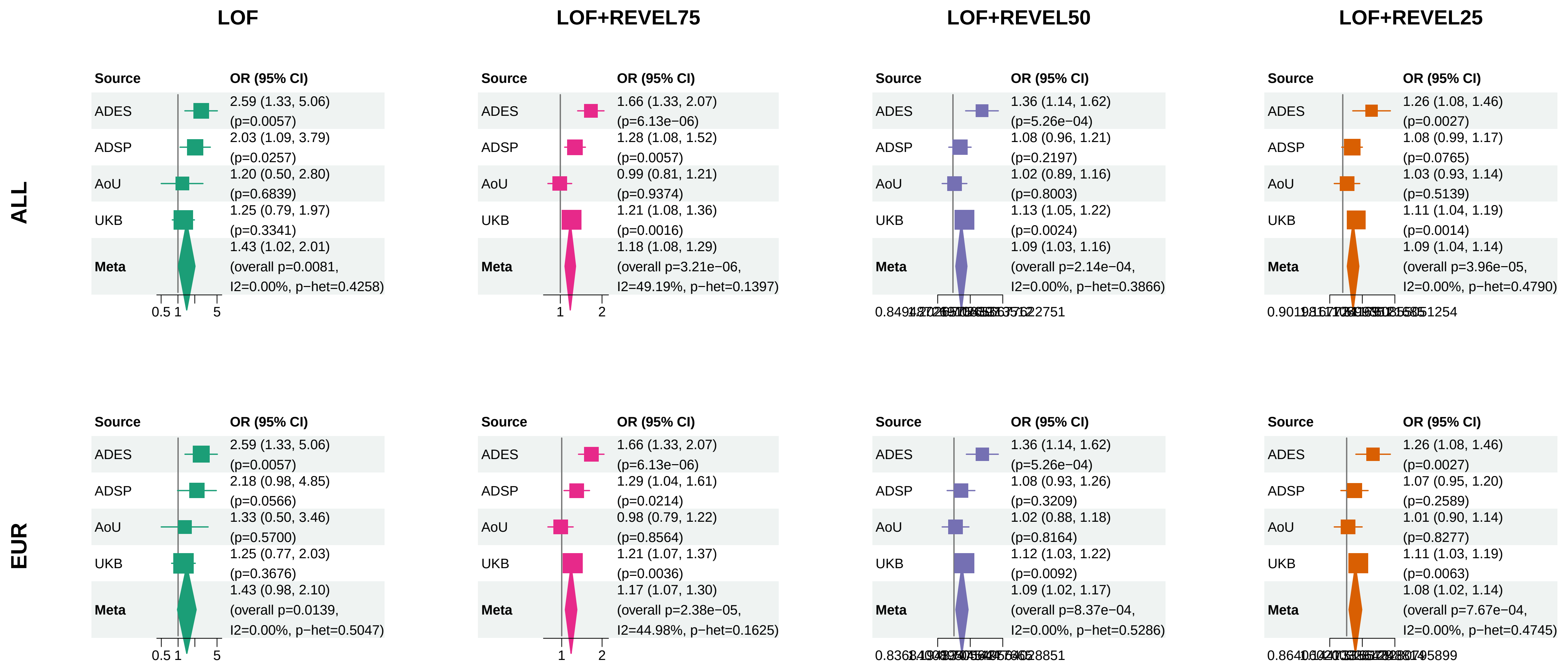


Meta OR/CI: pooled REGENIE effect; overall P: ADES+REGENIE signed−Z when the exact ADES mask was available, otherwise REGENIE.

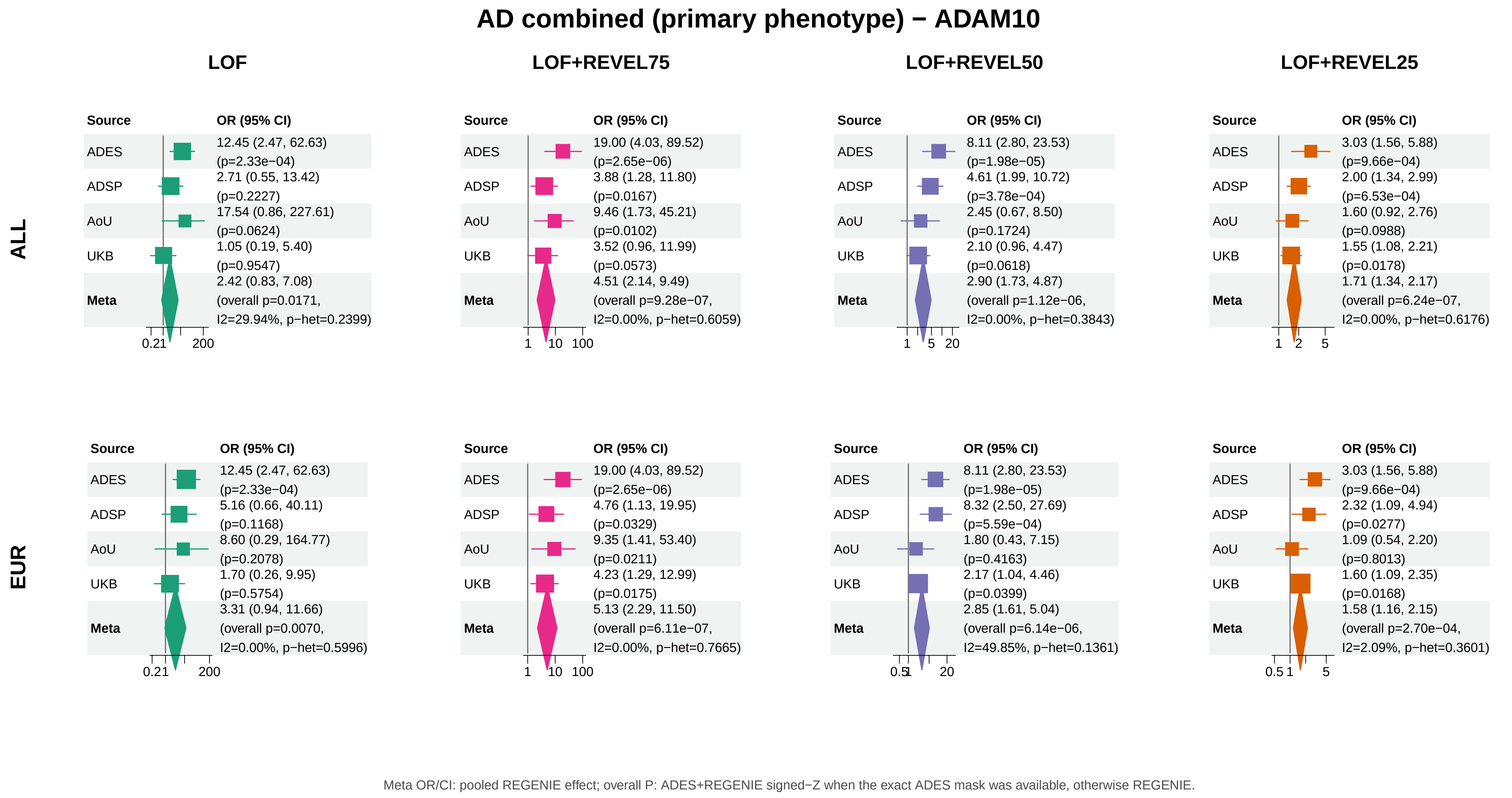
AD combined (primary phenotype) − ADAM10
LOF
LOF+REVEL75
LOF+REVEL50
LOF+REVEL25
ALL
Source
OR (95% CI)
ADES
12.45 (2.47, 62.63)
(p=2.33e−04)
ADSP
2.71 (0.55, 13.42)
(p=0.2227)
AoU
17.54 (0.86, 227.61)
(p=0.0624)
UKB
1.05 (0.19, 5.40)
(p=0.9547)
Meta
2.42 (0.83, 7.08)
(overall p=0.0171,
I2=29.94%, p−het=0.2399)
0.21
200
Source
OR (95% CI)
ADES
19.00 (4.03, 89.52)
(p=2.65e−06)
ADSP
3.88 (1.28, 11.80)
(p=0.0167)
AoU
9.46 (1.73, 45.21)
(p=0.0102)
UKB
3.52 (0.96, 11.99)
(p=0.0573)
Meta
4.51 (2.14, 9.49)
(overall p=9.28e−07,
I2=0.00%, p−het=0.6059)
1
10
100
Source
OR (95% CI)
ADES
8.11 (2.80, 23.53)
(p=1.98e−05)
ADSP
4.61 (1.99, 10.72)
(p=3.78e−04)
AoU
2.45 (0.67, 8.50)
(p=0.1724)
UKB
2.10 (0.96, 4.47)
(p=0.0618)
Meta
2.90 (1.73, 4.87)
(overall p=1.12e−06,
I2=0.00%, p−het=0.3843)
1
5
20
Source
OR (95% CI)
ADES
3.03 (1.56, 5.88)
(p=9.66e−04)
ADSP
2.00 (1.34, 2.99)
(p=6.53e−04)
AoU
1.60 (0.92, 2.76)
(p=0.0988)
UKB
1.55 (1.08, 2.21)
(p=0.0178)
Meta
1.71 (1.34, 2.17)
(overall p=6.24e−07,
I2=0.00%, p−het=0.6176)
1
2
5
EUR
Source
OR (95% CI)
ADES
12.45 (2.47, 62.63)
(p=2.33e−04)
ADSP
5.16 (0.66, 40.11)
(p=0.1168)
AoU
8.60 (0.29, 164.77)
(p=0.2078)
UKB
1.70 (0.26, 9.95)
(p=0.5754)
Meta
3.31 (0.94, 11.66)
(overall p=0.0070,
I2=0.00%, p−het=0.5996)
0.21
200
Source
OR (95% CI)
ADES
19.00 (4.03, 89.52)
(p=2.65e−06)
ADSP
4.76 (1.13, 19.95)
(p=0.0329)
AoU
9.35 (1.41, 53.40)
(p=0.0211)
UKB
4.23 (1.29, 12.99)
(p=0.0175)
Meta
5.13 (2.29, 11.50)
(overall p=6.11e−07,
I2=0.00%, p−het=0.7665)
1
10
100
Source
OR (95% CI)
ADES
8.11 (2.80, 23.53)
(p=1.98e−05)
ADSP
8.32 (2.50, 27.69)
(p=5.59e−04)
AoU
1.80 (0.43, 7.15)
(p=0.4163)
UKB
2.17 (1.04, 4.46)
(p=0.0399)
Meta
2.85 (1.61, 5.04)
(overall p=6.14e−06,
I2=49.85%, p−het=0.1361)
0.5
1
20
Source
OR (95% CI)
ADES
3.03 (1.56, 5.88)
(p=9.66e−04)
ADSP
2.32 (1.09, 4.94)
(p=0.0277)
AoU
1.09 (0.54, 2.20)
(p=0.8013)
UKB
1.60 (1.09, 2.35)
(p=0.0168)
Meta
1.58 (1.16, 2.15)
(overall p=2.70e−04,
I2=2.09%, p−het=0.3601)
0.5
1
5
Meta OR/CI: pooled REGENIE effect; overall P: ADES+REGENIE signed−Z when the exact ADES mask was available, otherwise REGENIE.

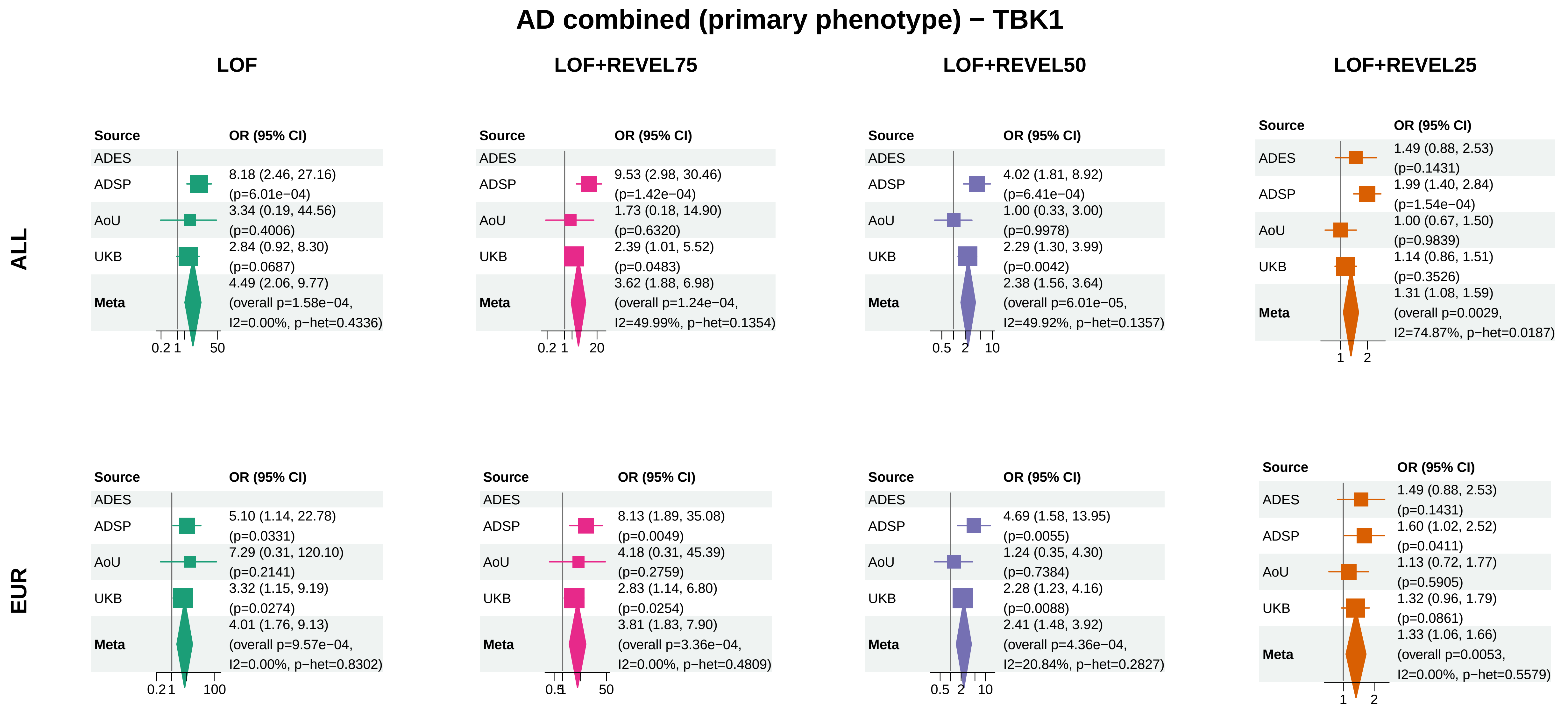


Meta OR/CI: pooled REGENIE effect; overall P: ADES+REGENIE signed−Z when the exact ADES mask was available, otherwise REGENIE.

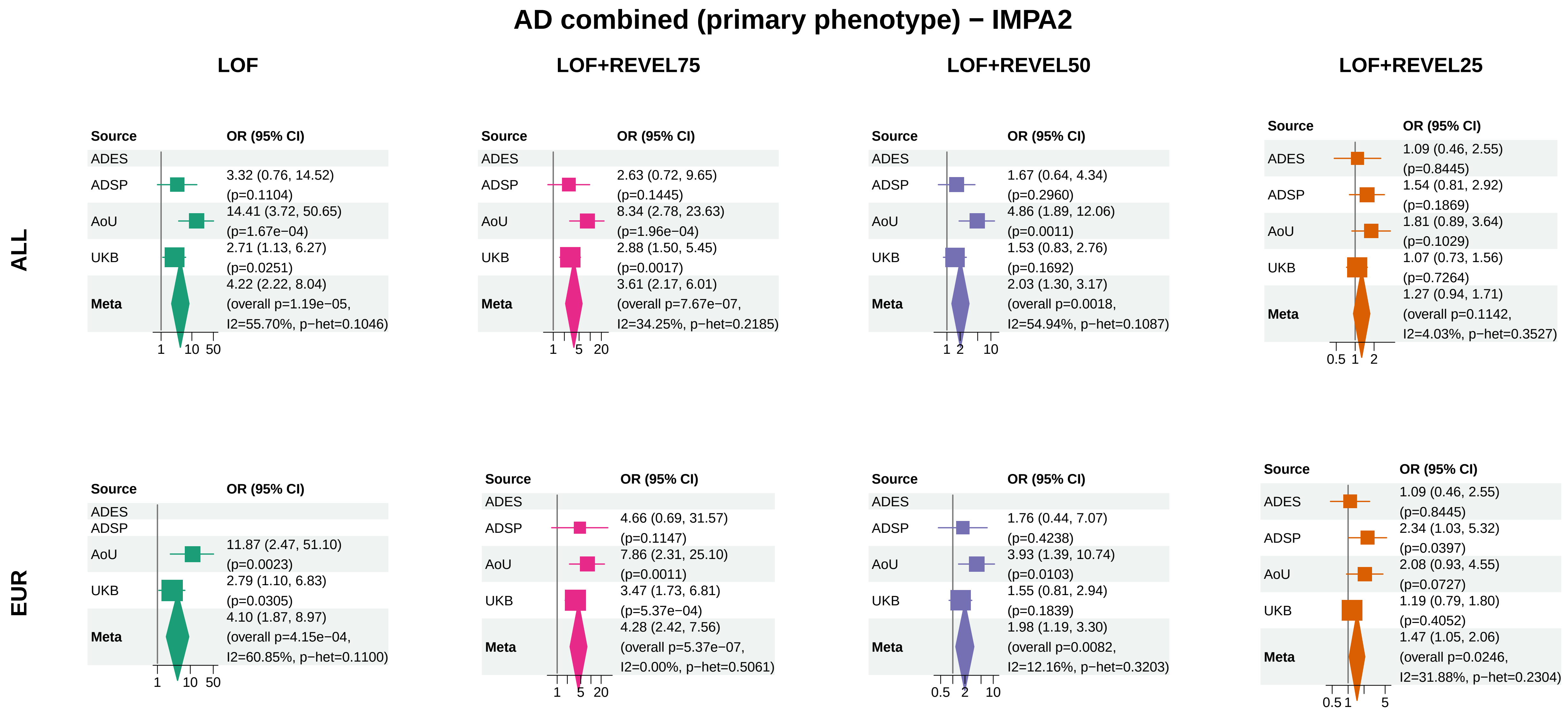


Meta OR/CI: pooled REGENIE effect; overall P: ADES+REGENIE signed−Z when the exact ADES mask was available, otherwise REGENIE.

#### AD combined (primary phenotype) − SHARPIN

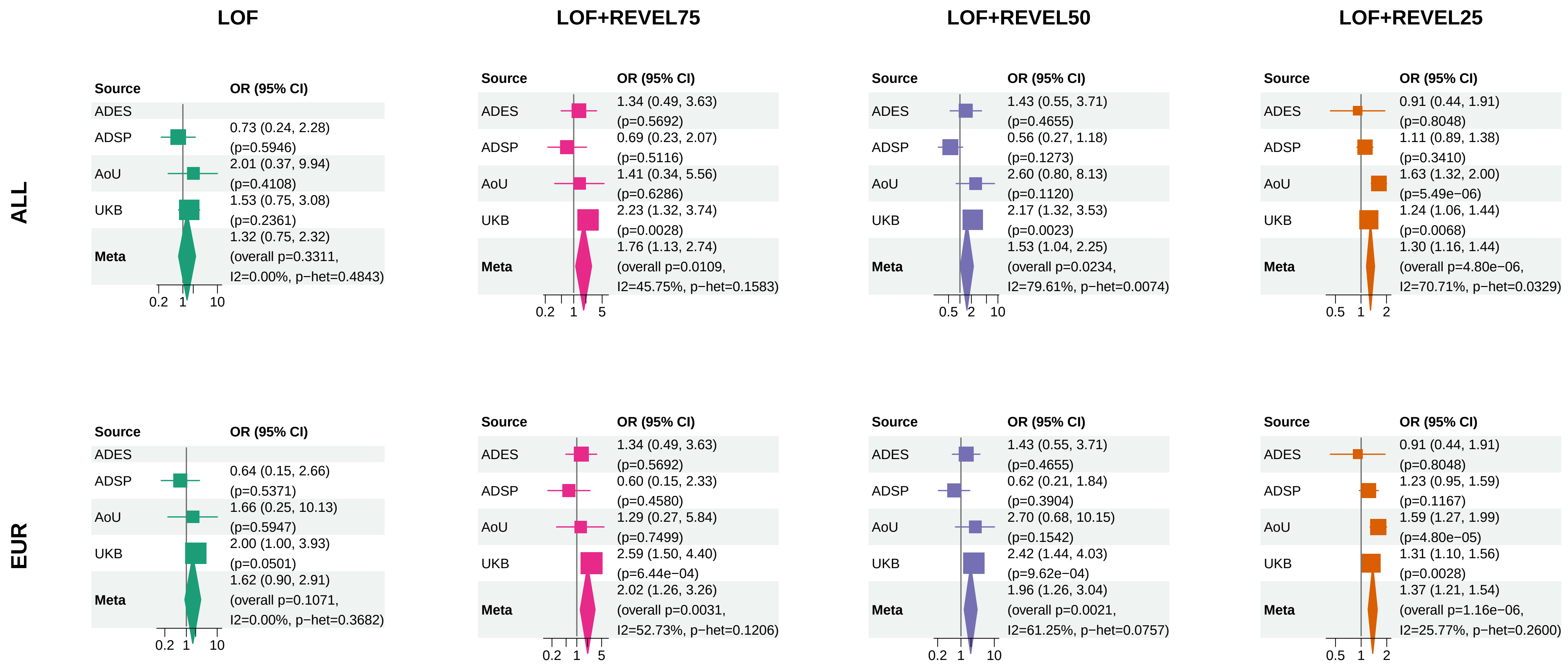


Meta OR/CI: pooled REGENIE effect; overall P: ADES+REGENIE signed−Z when the exact ADES mask was available, otherwise REGENIE.

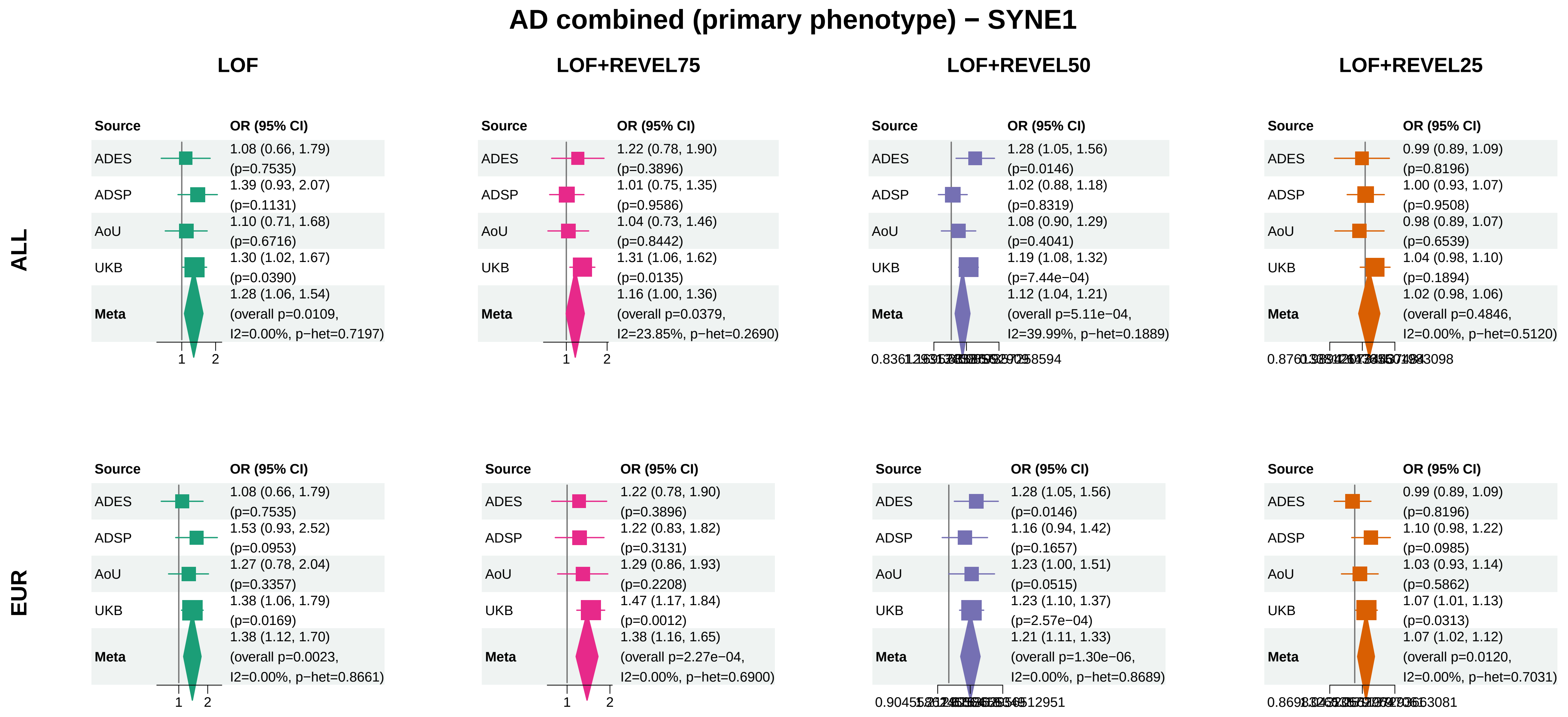


Meta OR/CI: pooled REGENIE effect; overall P: ADES+REGENIE signed−Z when the exact ADES mask was available, otherwise REGENIE.

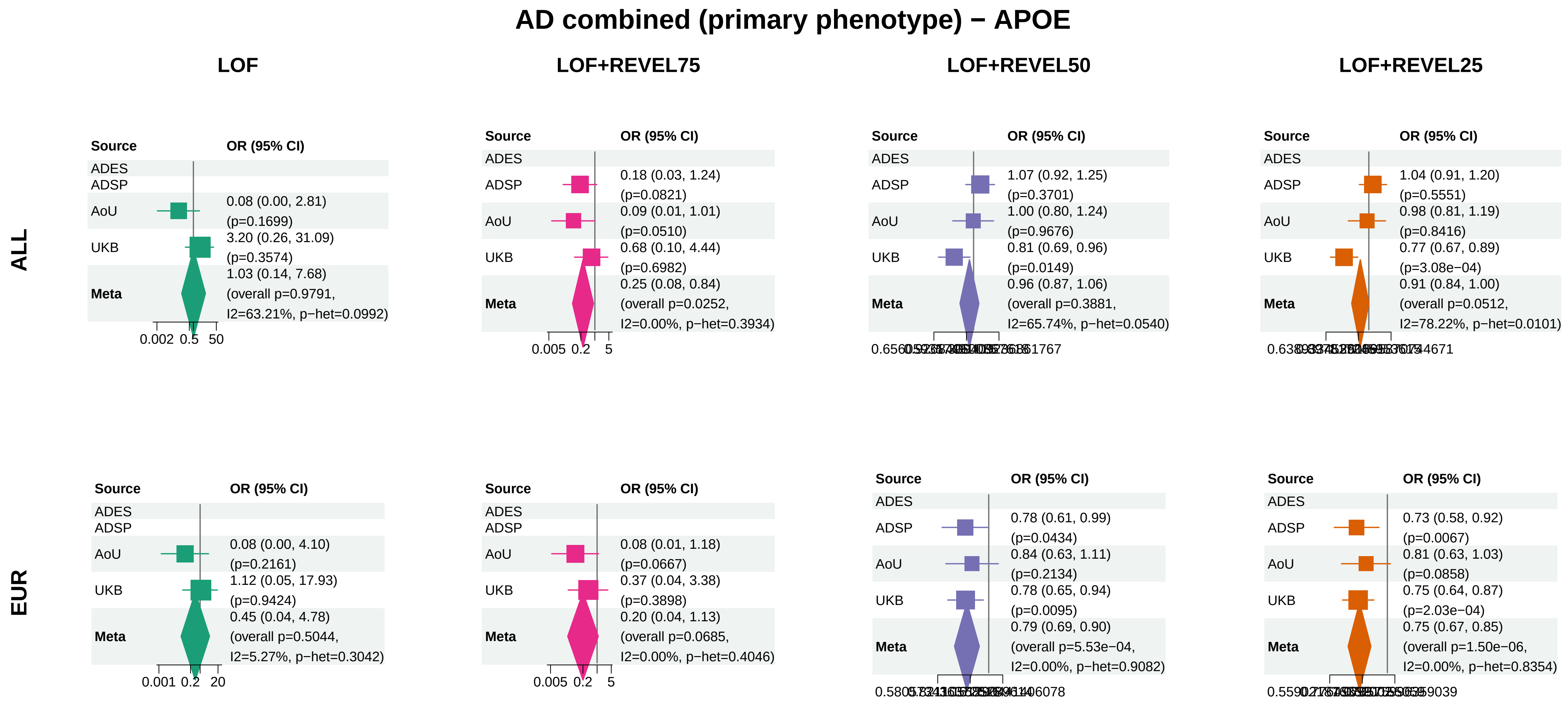


Meta OR/CI: pooled REGENIE effect; overall P: ADES+REGENIE signed−Z when the exact ADES mask was available, otherwise REGENIE.

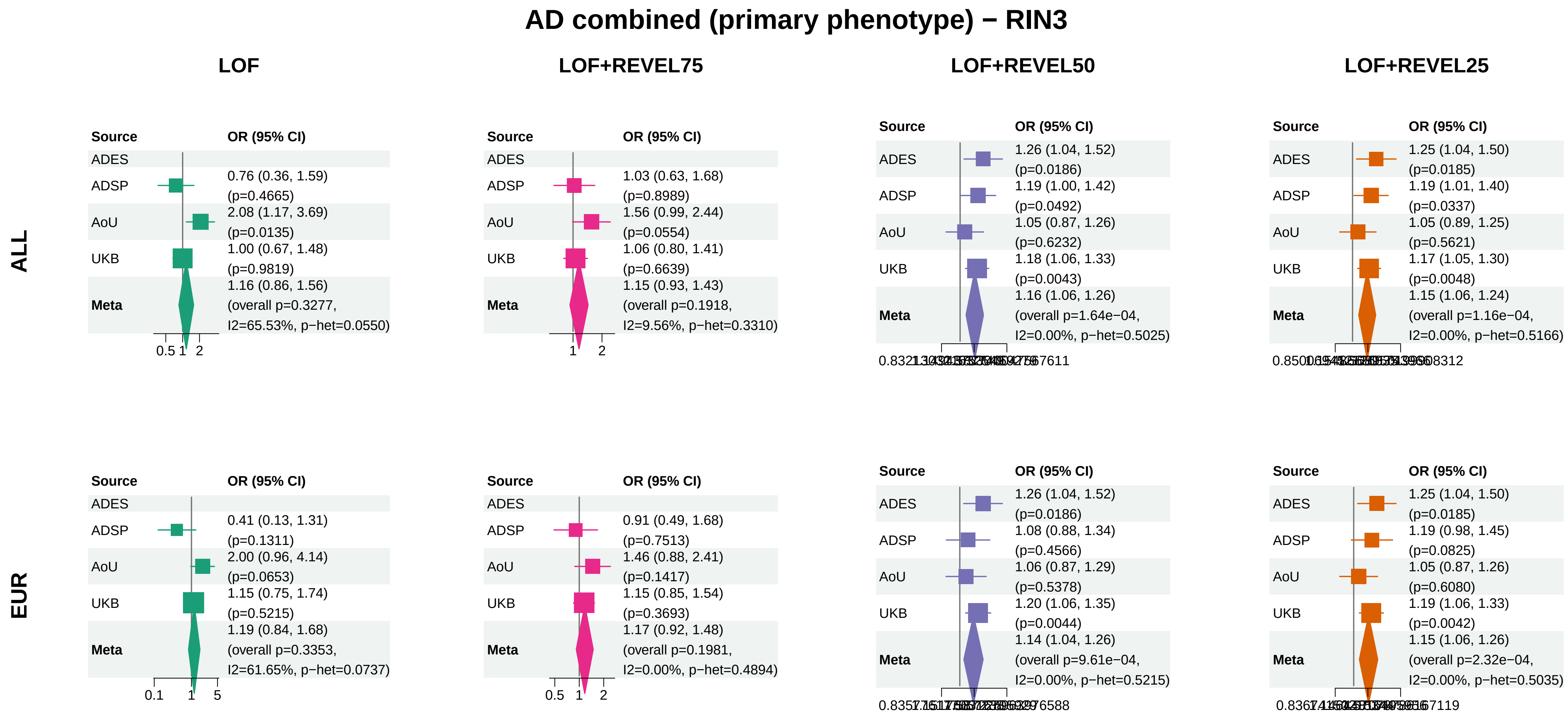


Meta OR/CI: pooled REGENIE effect; overall P: ADES+REGENIE signed−Z when the exact ADES mask was available, otherwise REGENIE.

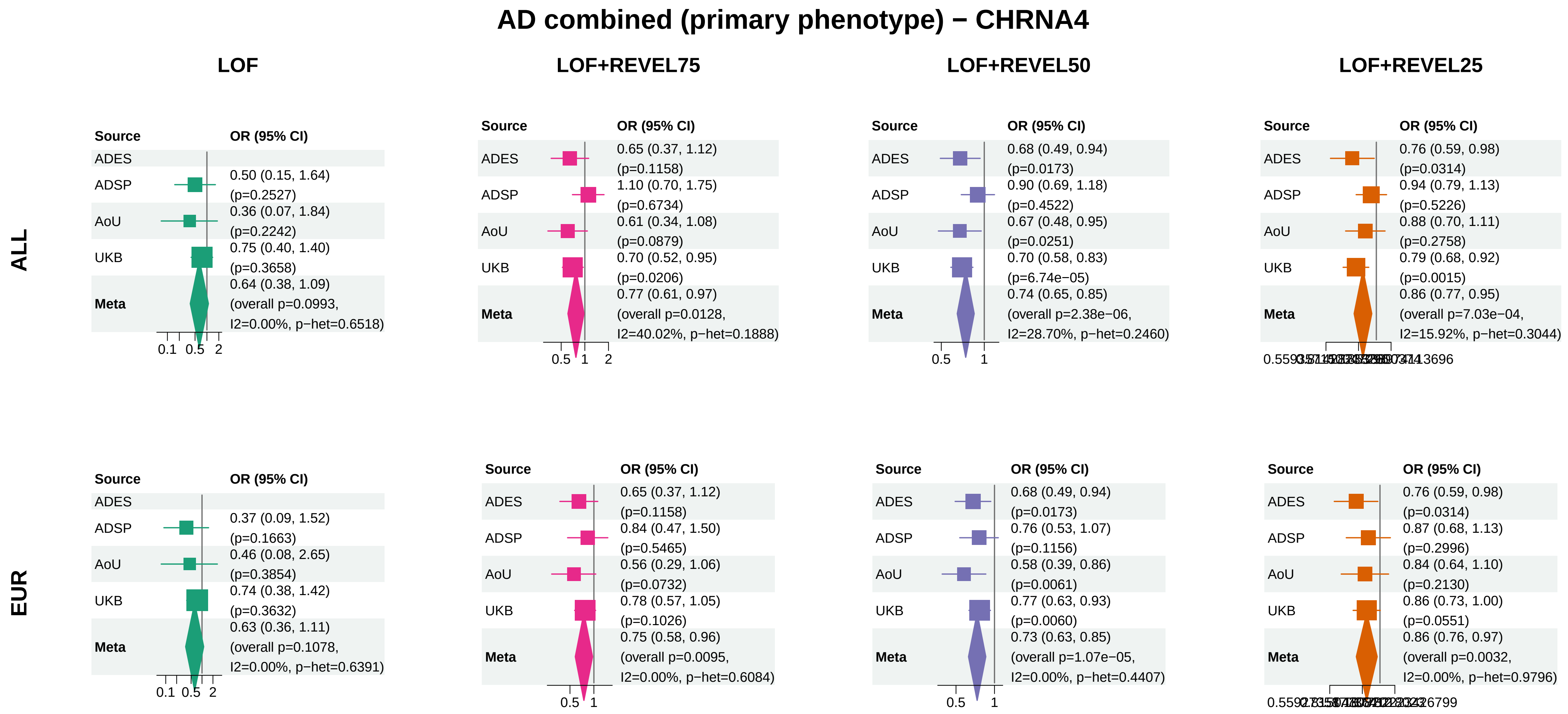


Meta OR/CI: pooled REGENIE effect; overall P: ADES+REGENIE signed−Z when the exact ADES mask was available, otherwise REGENIE.

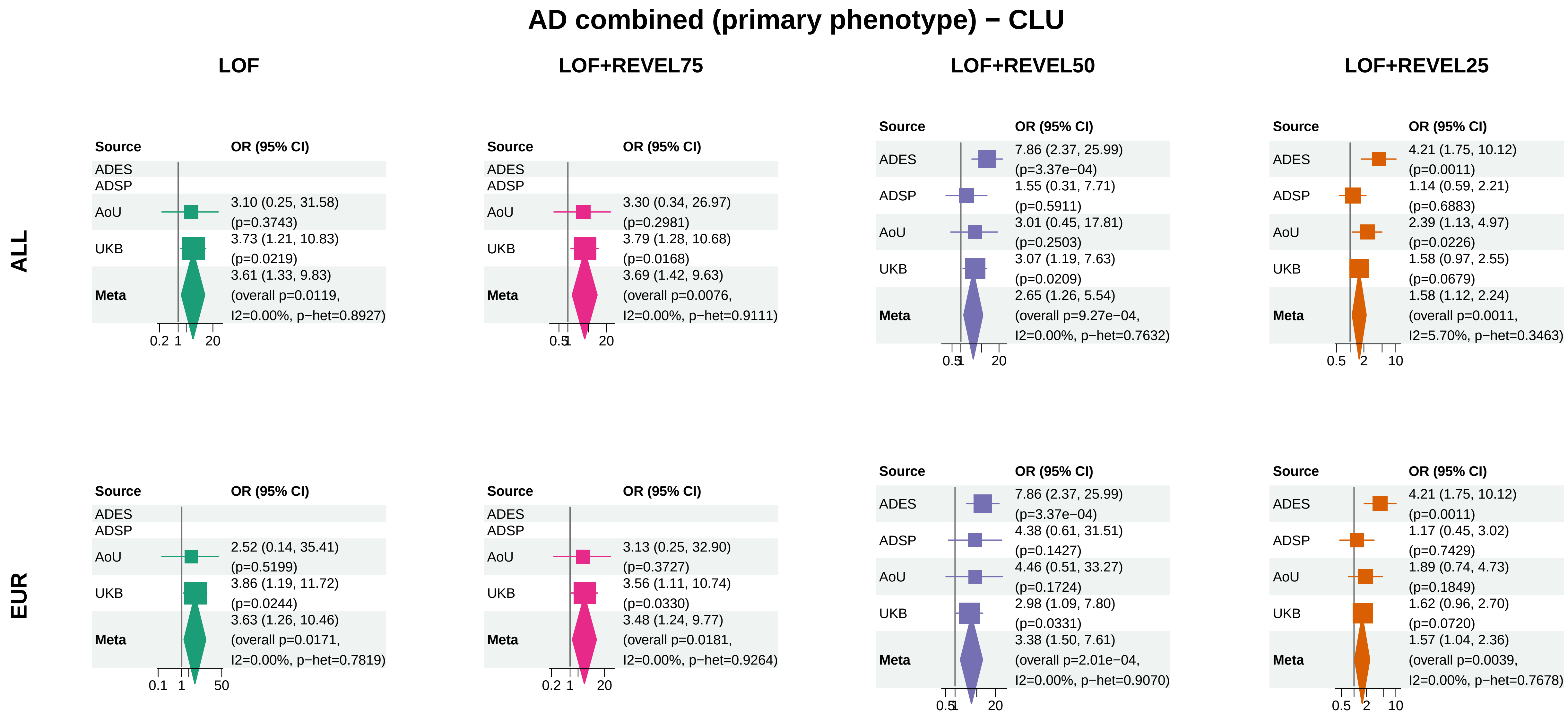


Meta OR/CI: pooled REGENIE effect; overall P: ADES+REGENIE signed−Z when the exact ADES mask was available, otherwise REGENIE.

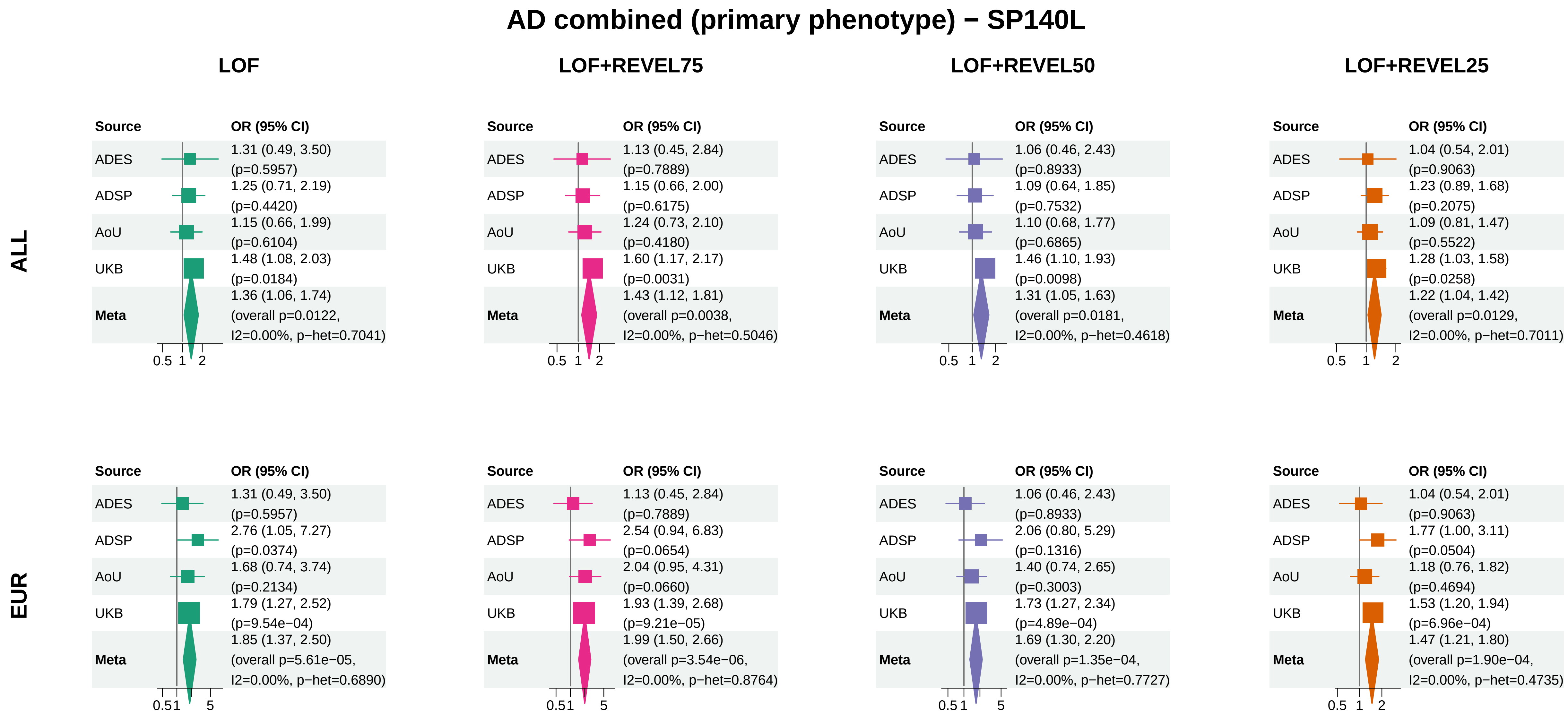


Meta OR/CI: pooled REGENIE effect; overall P: ADES+REGENIE signed−Z when the exact ADES mask was available, otherwise REGENIE.

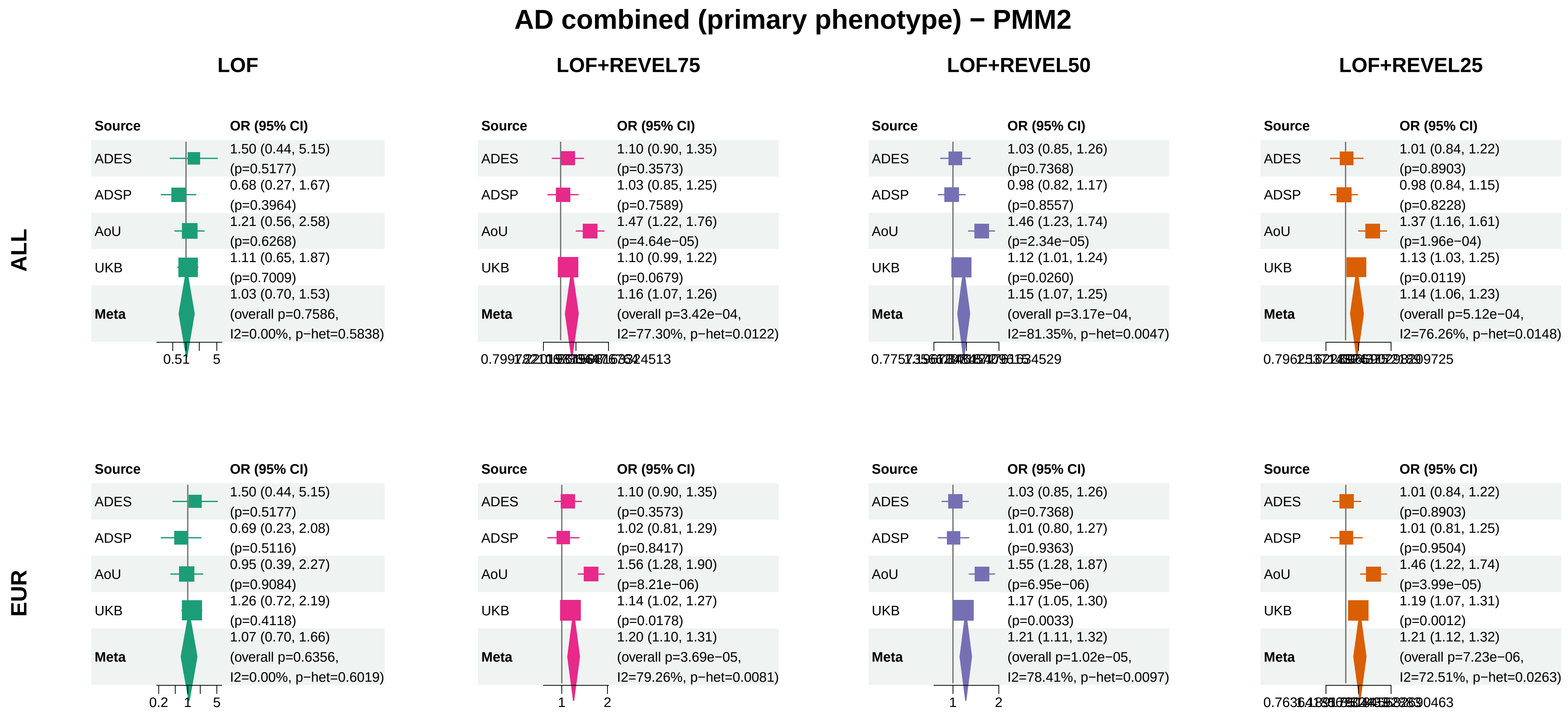


Meta OR/CI: pooled REGENIE effect; overall P: ADES+REGENIE signed−Z when the exact ADES mask was available, otherwise REGENIE.

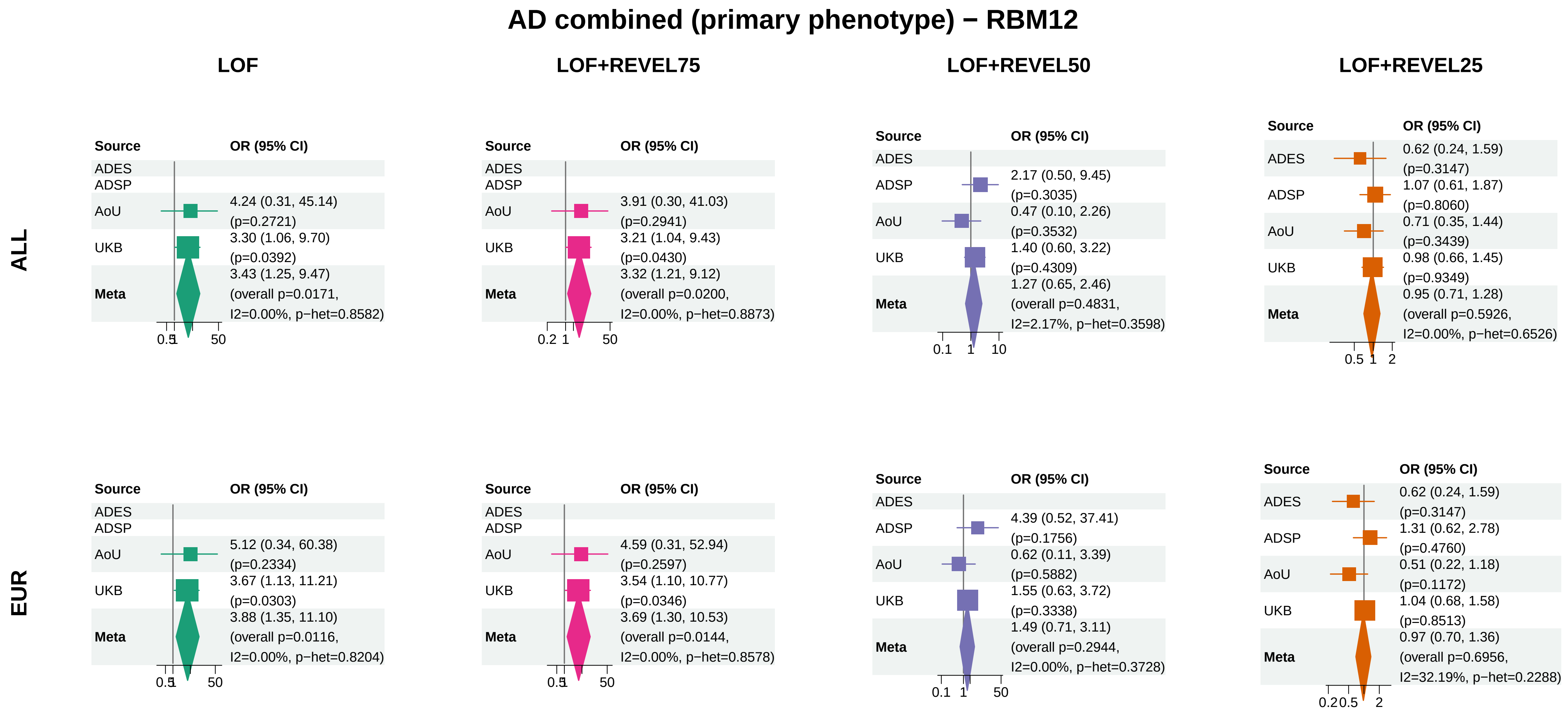


Meta OR/CI: pooled REGENIE effect; overall P: ADES+REGENIE signed−Z when the exact ADES mask was available, otherwise REGENIE.

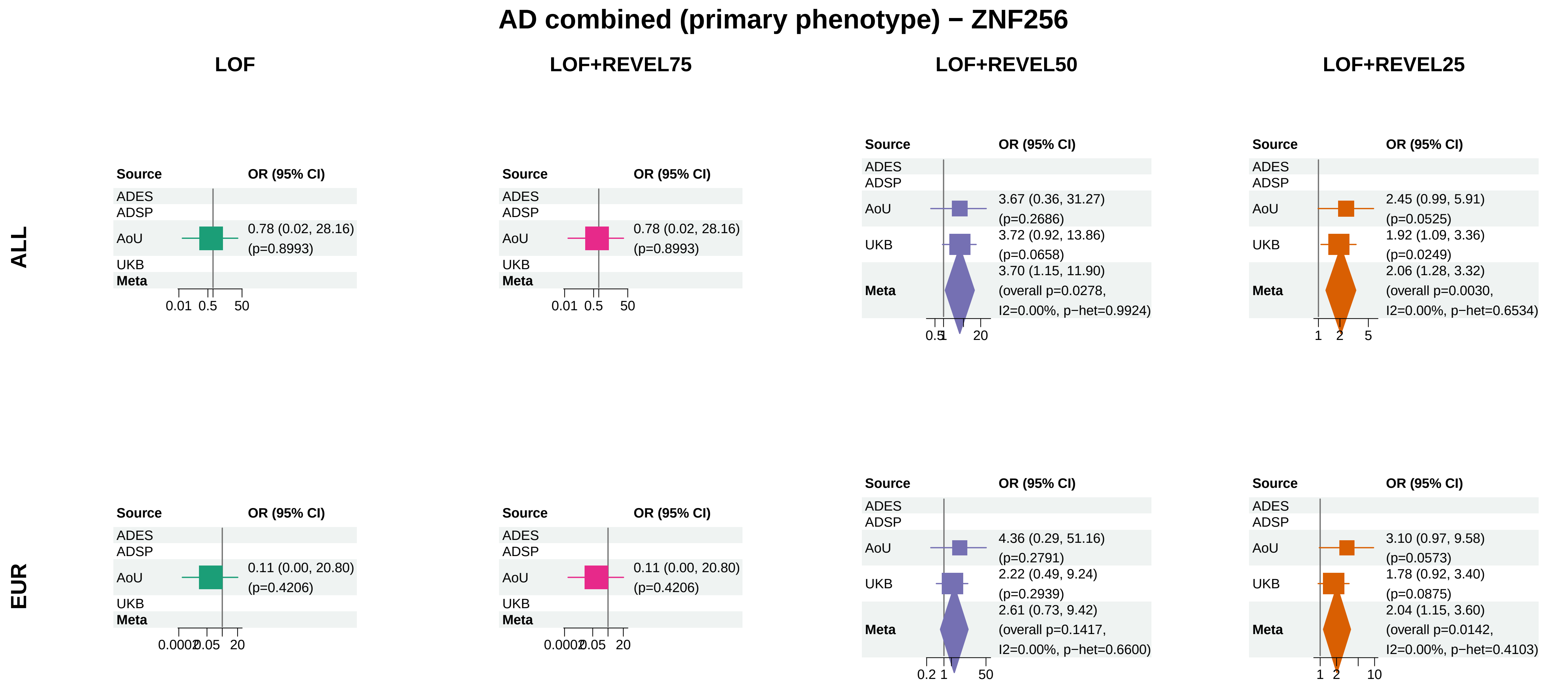


Meta OR/CI: pooled REGENIE effect; overall P: ADES+REGENIE signed−Z when the exact ADES mask was available, otherwise REGENIE.

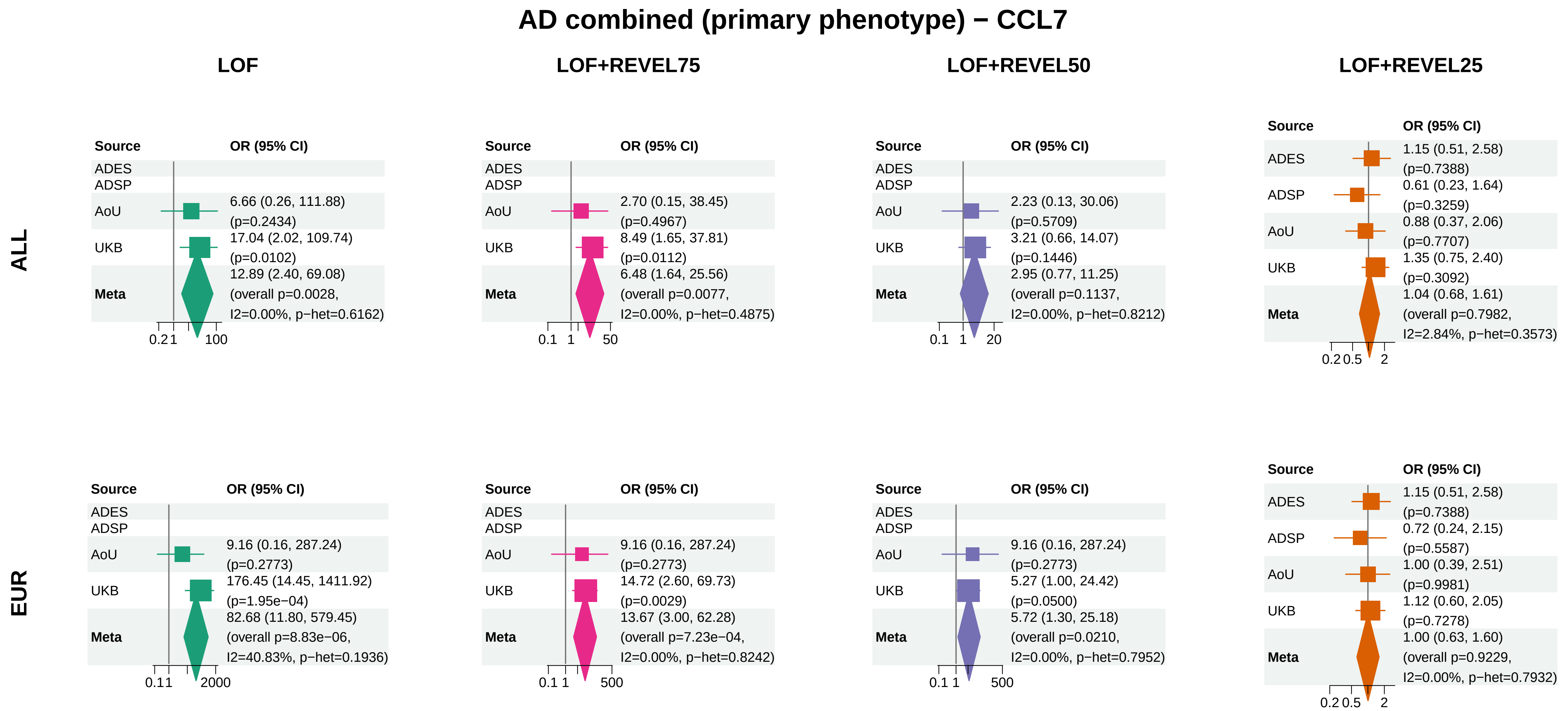


Meta OR/CI: pooled REGENIE effect; overall P: ADES+REGENIE signed−Z when the exact ADES mask was available, otherwise REGENIE.

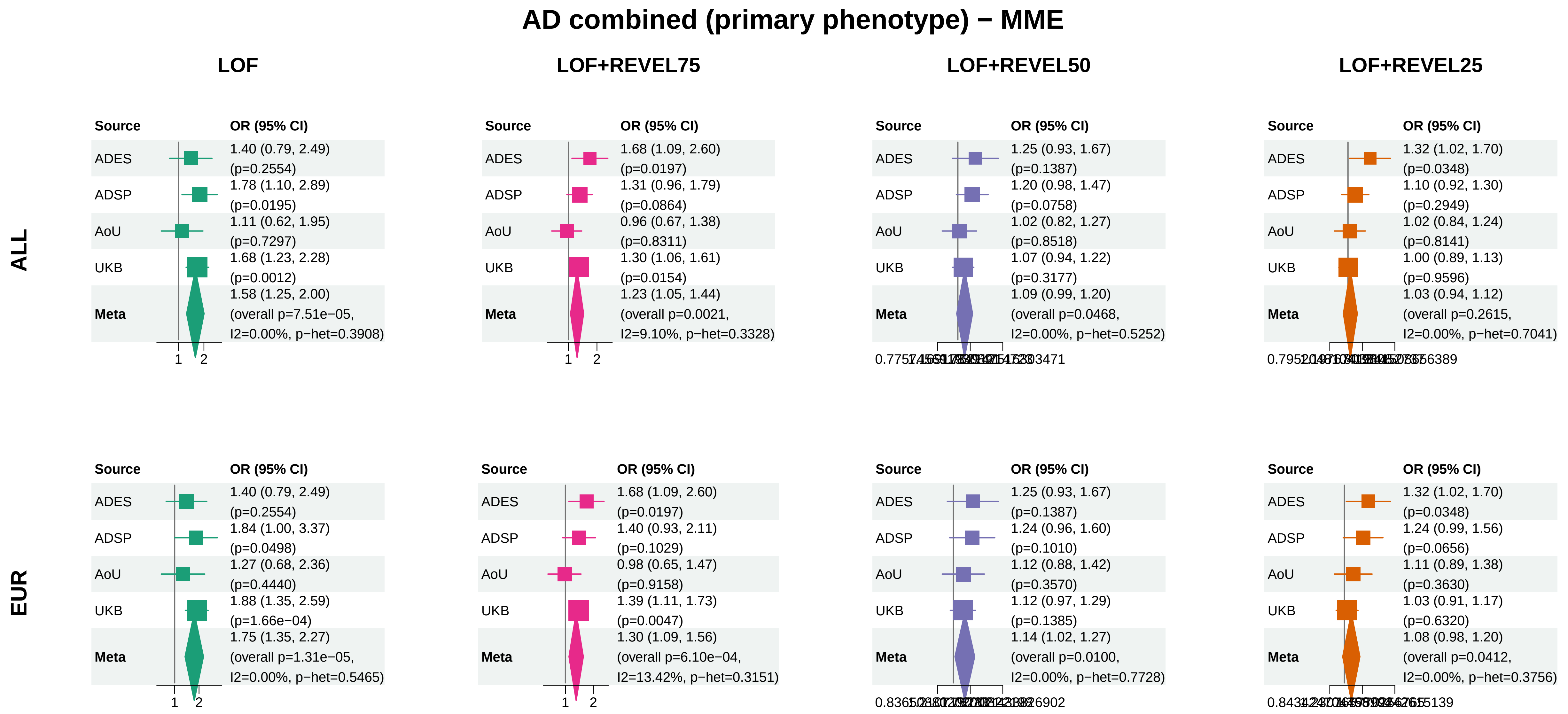


Meta OR/CI: pooled REGENIE effect; overall P: ADES+REGENIE signed−Z when the exact ADES mask was available, otherwise REGENIE.
